# Genome-wide analysis of 944,133 individuals provides insights into the etiology of hemorrhoidal disease

**DOI:** 10.1101/2020.12.03.20242776

**Authors:** Tenghao Zheng, David Ellinghaus, Simonas Juzenas, François Cossais, Greta Burmeister, Gabriele Mayr, Isabella Friis Jørgensen, Maris Teder-Laving, Anne Heidi Skogholt, Karina Banasik, Thomas Becker, Frank Bokelmann, Søren Brunak, Stephan Buch, Hartmut Clausnizer, Christian Datz, DBDS Consortium, Frauke Degenhardt, Marek Doniec, Christian Erikstrup, Tõnu Esko, Michael Forster, Norbert Frey, Lars G. Fritsche, Maiken Elvestad Gabrielsen, Tobias Gräßle, Andrea Gsur, Justus Gross, Jochen Hampe, Alexander Hendricks, Sebastian Hinz, Kristian Hveem, Johannes Jongen, Ralf Junker, Tom Hemming Karlsen, Georg Hemmrich-Stanisak, Wolfgang Kruis, Juozas Kupcinskas, Tilman Laubert, Matthias Laudes, Fabian H. Leendertz, Wolfgang Lieb, Verena Limperger, Nikolaos Margetis, Kerstin Mätz-Rensing, Christopher Georg Németh, Eivind Ness-Jensen, Ulrike Nowak-Göttl, Anita Pandit, Ole Birger Pedersen, Hans Günter Peleikis, Kenneth Peuker, Cristina Leal Rodríguez, Malte Rühlemann, Bodo Schniewind, Martin Schulzky, Jurgita Skieceviciene, Jürgen Tepel, Laurent Thomas, Florian Uellendahl-Werth, Henrik Ullum, Ilka Vogel, Henry Völzke, Lorenzo von Fersen, Witigo von Schoenfels, Brett Vanderwerff, Julia Wilking, Michael Wittig, Sebastian Zeissig, Myrko Zobel, Matthew Zawistowski, Vladimir Vacic, Olga Sazonova, Elizabeth S. Noblin, The 23andMe Research Team, Thilo Wedel, Volker Kahlke, Clemens Schafmayer, Mauro D’Amato, Andre Franke

## Abstract

Hemorrhoidal disease (HEM) affects a large fraction of the population but its etiology including suspected genetic predisposition is poorly understood. We conducted a GWAS meta-analysis of 218,920 HEM patients and 725,213 controls of European ancestry, demonstrating modest heritability and genetic correlation with several other diseases from the gastrointestinal, neuroaffective and cardiovascular domains. HEM polygenic risk scores validated in 180,435 individuals from independent datasets allowed the identification of those at risk and correlated with younger age of onset and recurrent surgery. We identified 102 independent HEM risk loci harboring genes whose expression is enriched in blood vessels and gastrointestinal tissues, and in pathways associated with smooth muscles, epithelial and endothelial development and morphogenesis. Network transcriptomic analyses of affected tissue from HEM patients highlighted HEM gene co-expression modules that are relevant to the development and integrity of the musculoskeletal and epidermal systems, and the organization of the extracellular matrix. We conclude HEM has a genetic component that predisposes to smooth muscle, epithelial and connective tissue dysfunction.

## Introduction

Hemorrhoids are normal anal vascular cushions filled with blood at the junction of the rectum and the anus. It is assumed that their main role in humans is to maintain continence^1^ but other functions such as sensing fullness, pressure and perceiving anal contents have been suggested given the sensory innervation^2^. Hemorrhoidal disease (hereafter referred to as HEM) occurs when hemorrhoids enlarge and become symptomatic (sometimes associated with rectal bleeding and itching/soiling) due to the deterioration or prolapse of the anchoring connective tissue, the dilation of the hemorrhoidal plexus or the formation of blood clots. Severe forms of HEM often require surgical treatment and the removal of abnormally enlarged and/or thrombosed hemorrhoids^1^. HEM prevalence increases with age and shows staggering figures worldwide (up to 86% prevalence in some reports)^3^, whereby a large proportion of cases remain undetected as asymptomatic or mild enough to be self-treated with over-the-counter treatment. After surgical treatment, the average patient cannot go to work for approximately three weeks. Thus, HEM represent a considerable medical and socioeconomic burden with an estimated annual cost of $800 million in the United States alone, mainly related to the large number of hemorrhoidectomies performed every year (which also cause considerable indirect cost due to loss of working hours during recovery)^4^.

A number of HEM risk factors have been suggested, including human erect position. In fact, the tight anal sealing as provided by an elaborated hemorrhoidal plexus may have developed during human evolution co-occurring with permanent bipedalism, as shown by our histology comparison of four different mammals (human, gorilla, baboon, mouse; **Supplementary Figure 1** and **Supplementary Note**). Other suggested risk factors are a sedentary lifestyle, obesity, reduced dietary fiber intake, spending excess time on the toilet, straining during defecation, strenuous lifting, constipation, diarrhea, pelvic floor dysfunction, pregnancy and giving natural birth, with several being controversially reported. The hypothetical model shown in **Supplementary Figure 2** summarizes the contemporary concepts regarding the pathophysiology of HEM development^5^. Until today, HEM etiopathogenesis is poorly investigated, and neither the exact molecular mechanisms nor the reason(s) why only some people develop HEM are known, hence undocumented theories, myths and hypotheses have become manifest even in the scientific literature. Genetic susceptibility may play a role in HEM development, but no large-scale, genome-wide association study (GWAS) for HEM has ever been conducted. To evaluate the contribution of genetic variation to the genetic architecture of HEM, we carried out a GWAS meta-analysis in 218,920 affected individuals and 725,213 population controls of European ancestry.

## Results

### GWAS META-ANALYSIS AND FINE MAPPING GENOMIC REGIONS

To systematically evaluate the contribution of genetic variation to the genetic architecture of HEM, we conducted a GWAS meta-analysis in 944,133 individuals of European ancestry from five large population-based cohorts (23andMe, UK Biobank [UKBB], Estonian Genome Center at the University of Tartu [EGCUT], Michigan Genomics Initiative [MGI], and Genetic Epidemiology Research on Aging [GERA]) including 218,920 HEM cases and 725,213 controls (**Supplementary Table 1**). HEM cases had higher body mass index (BMI), were significantly older and more often female compared to non-HEM controls, hence age, sex, and BMI (if available) and top principal components from principal component analysis (PCA) were included as co-variates in each of the contributing GWAS, **Supplementary Note**).

After data harmonization and quality control, 8,494,288 high-quality, common (MAF>0.01) single nucleotide polymorphisms (SNPs) were included in a fixed-effect inverse variance meta-analysis using the software METAL (see **Methods**). We identified 5,480 genome-wide significant associations (*P*_Meta_<5×10^−8^), which were mapped to 102 independent genomic regions using FUMA (see **Methods)**. A summary of the association results for 102 lead SNPs is provided in **Figure 1** and **Supplementary Table 2**. Although genomic inflation was observed (λ=1.3; **Supplementary Figure 3**), this was likely due to polygenicity rather than population stratification, as determined via linkage disequilibrium score regression analysis (LDSC, intercept=1.06; **Methods**) and based on a normalized λ_1000_ = 1.001. The heritability of HEM was estimated at 5% (SNP-based heritability, *h*^2^_SNP_ computed with LDSC), whereby the newly identified 102 risk variants explained about 0.9% of the variance (see **Methods)**.

**Figure 1:**
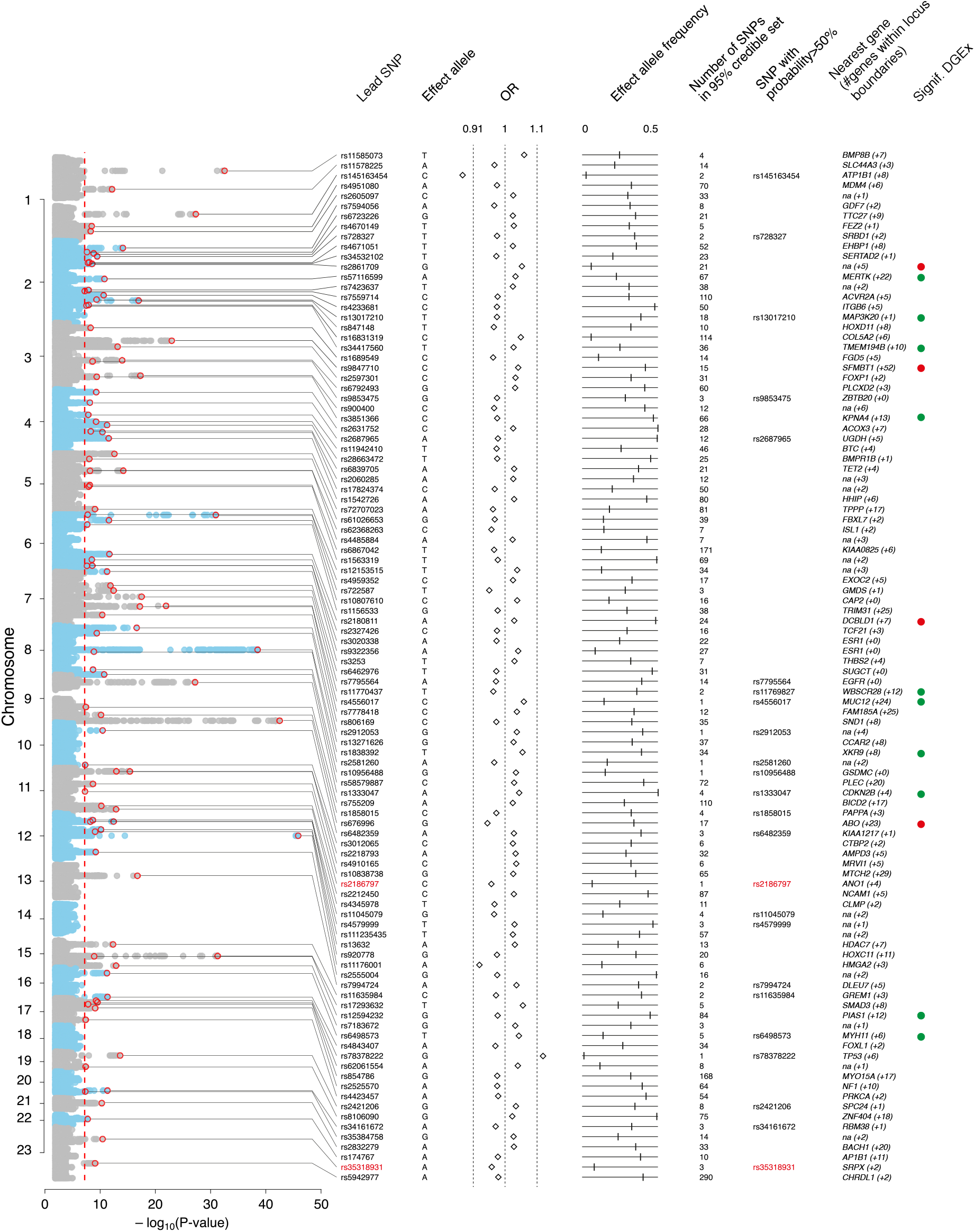
Annotation of 102 HEM GWAS risk loci. From left to right: Manhattan plot of GWAS meta-analysis results, (genome-wide significance level – *P*_Meta_<5×10^−8^ – indicated with vertical dotted red line); Lead SNP – marker associated with the strongest association signal from each locus (also annotated with a red circle in the Manhattan plot); Effect allele – allele associated with reported genetic risk effects (OR), also always the minor allele; OR – odds ratio with respect to the effect allele; Effect allele frequency – frequency of the effect allele in the discovery dataset; Number of SNPs in 95% credible set – the minimum set of variants from Bayesian fine-mapping analysis that is >95% likely to contain the causal variant; SNP with probability >50% – single variant (if detected) with >50% probability of being causal (coding SNPs highlighted in red); Nearest gene (#genes within locus boundaries) – gene closest to the lead SNP (if within 100 kb distance, otherwise “na”) and number of additional genes positionally mapped to the locus using FUMA (**Supplementary Table 2** and **Methods**). Signif. DGEx – locus containing HEM genes differentially expressed in RNA Combo-Seq analysis of HEM affected tissue, detected at higher (red) and/or lower (green) level of expression (see **Methods**).

Bayesian fine-mapping analysis with FINEMAP^5^ identified a total of 3,323 SNPs that belong to the 95% credible sets of variants most likely to be causal at each locus (**Methods, Supplementary Figure 4** and **Supplementary Table 3**). For six loci, we pinpointed the association signal to a single causal variant with greater than 95% certainty, including the missense variants rs2186797 (*ANO1*) and rs35318931 (*SRPX*). For another 24 loci there was evidence that the lead variant is causal with >50% certainty (**Figure 1**).

### CROSS-TRAIT ANALYSES

A lookup of HEM association signals in previous GWAS studies retrieved from GWAS Atlas, GWAS Catalog and via Phenoscanner2 (**Methods**) revealed that 76/102 loci had been previously associated with several diseases and traits across the metabolic, cardiovascular, digestive, psychiatric, environmental and other domains (illustrated in **Figure 2**, full details in **Supplementary Table 4**).

**Figure 2:**
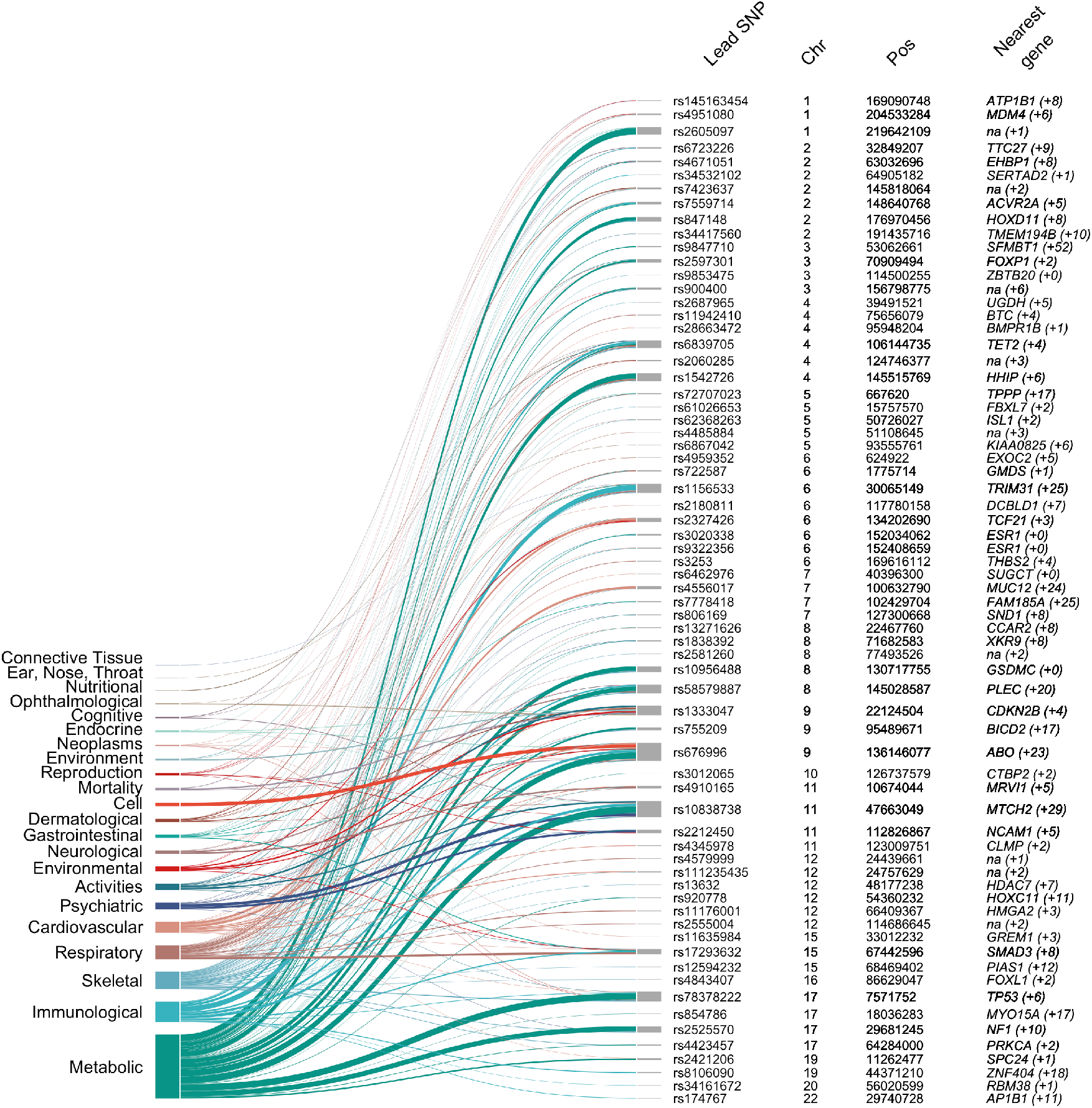
Previously reported associations of HEM risk loci with other traits and diseases, clustered by biological areas. The plot shows associations with other traits, extracted from the GWAS ATLAS for the 102 lead SNPs (and/or their r^2^>0.8 LD proxies) ordered by chromosome and chromosomal position. Associations are grouped by domain and represented with different colors. The ribbon size is proportional to the number of traits associated at the genome-wide significance level (*P*_Meta_<5×10^−8^). Columns from left to right: Lead SNP – marker showing strongest association signal from each locus; Chr – chromosome; Pos – SNP position on chromosome (genome build hg19); Nearest gene (#genes within locus boundaries) – gene closest to the lead SNP (if within 100 kb distance, otherwise “na”) (see **Methods**).

We then investigated genetic correlations with 1,387 other human traits and conditions at the genome-wide level using LDSC as implemented in CTG-VL (see **Methods**). The strongest correlations (r_g_) were observed with diseases and traits from the gastrointestinal (GI) domain, including “other diseases of anus and rectum” (r_g_=0.78, *P*_FDR_=4.94×10^−8^), “fissure and fistula of anal and rectal regions*”* (r_g_=0.58, *P*_FDR_=2.70×10^−12^), “self-reported irritable bowel syndrome*”* (r_g_=0.42, *P*_FDR_= 1.87×10^−7^), use of “laxatives*”* (r_g_=0.42, *P*_FDR_=2.35×10^−14^), and “diverticular disease*”* (r_g_=0.23, *P*_FDR_=6.68×10^−9^) among others (**Figure 3**). Given these similarities, we performed a genome-wide pleiotropy analysis for diverticular disease, irritable bowel syndrome (IBS) and HEM, revealing 44 independent genomic regions shared by at least two phenotypes from the group of diverticular disease, IBS and HEM phenotypes (**Supplementary Table 5**). Other notable correlations were detected for psychiatric and neuroaffective disorders (anxiety, depression and neuroticism), pain-related traits (including abdominal pain and painful gums), diseases of the circulatory system, and diseases of the musculoskeletal system and connective tissue (**Figure 3** and **Supplementary Table 6**).

**Figure 3:**
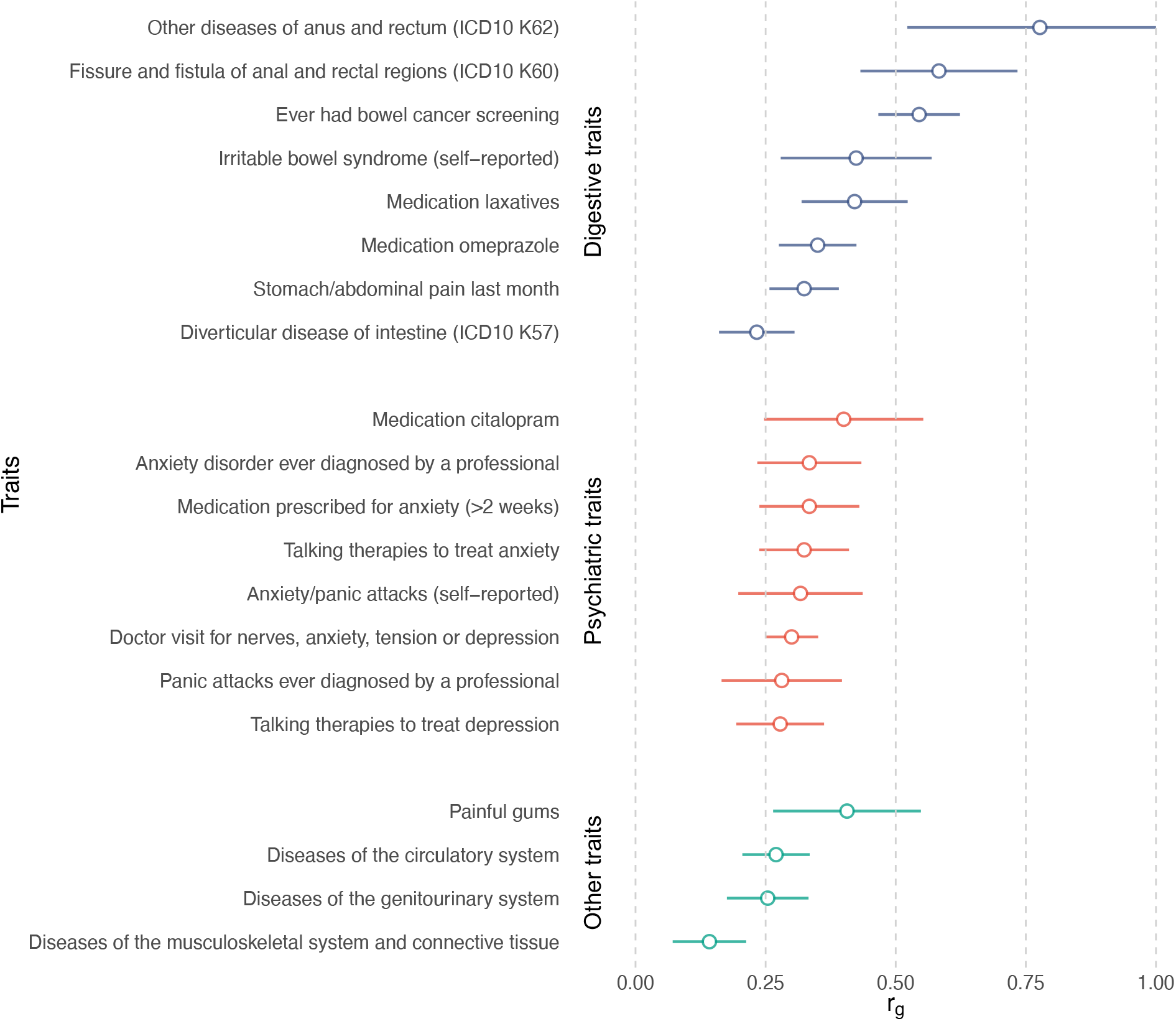
Genetic correlation between HEM and other traits estimated by linkage disequilibrium score regression analysis (LDSC). Genetic correlations (*r*_g_+se) are shown for selected traits, grouped by domain. Only correlations significant after Bonferroni correction were considered (full list available in **Supplementary Table 6)**.

To gain further insight into potential cross-trait overlaps and to validate the LDSC results, we assessed whether genetically correlated traits and conditions also occur more frequently in HEM patients by analyzing data on diagnoses and medications from UKBB. The results were highly consistent with those obtained via LDSC (**Figure 4** and **Supplementary Table 6**). Compared to controls, HEM patients additionally suffered more often from diverticular disease, IBS and other functional gastrointestinal disorders (FGIDs), abdominal pain, hypertension, ischemic heart disease, depression, anxiety, and diseases of the musculoskeletal system and connective tissue among others. To further consolidate these findings, we analyzed an independent population-scale healthcare record dataset comprising 8,172,531 individuals from the Danish National Patient Registry (DNPR; **Methods**), obtaining similar results (see inner circle of **Figure 4**). Therefore, based on these largely overlapping observations at the genetic and epidemiological level, HEM appears to be strongly associated with other diseases of the digestive, neuroaffective and cardiovascular domains.

**Figure 4:**
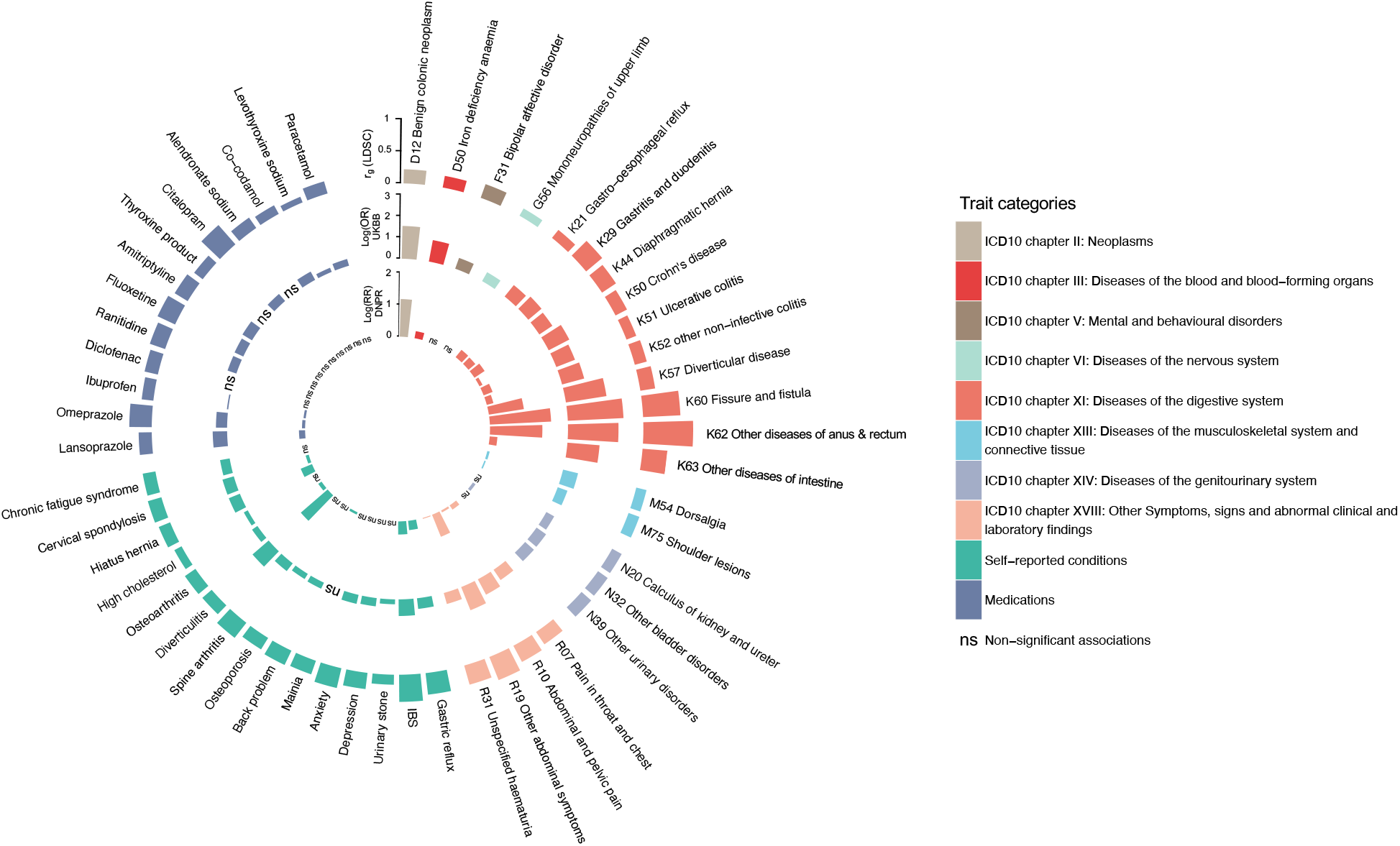
Analysis of HEM genetically correlated traits in UK Biobank (UKBB) and the Danish National Patient Registry (DNPR). Traits and conditions identified in LDSC analyses of genetic correlation with HEM (outer ring in the circos plot, see also **Figure 3** and **Supplementary Table 6**) were studied for their differential prevalence in UKBB and DNPR, based on data extracted from participants’ healthcare records. Significant results are reported, respectively, as odds ratio (log[OR], UKBB, middle ring) and relative risk (log[RR], DNPR, inner ring) or “ns” (for non-significant findings). Diseases and traits are categorized according to ICD10 diagnostic codes or self-reported conditions and use of medications from questionnaire data (see **Methods**). Self-reported traits in UKBB (dark blue color) were manually mapped to ICD10-codes in DNPR.

### POLYGENIC RISK SCORES (PRS)

We next exploited meta-analysis summary statistics to compute HEM PRS and evaluate their relevance and translational potential in independent datasets. HEM PRS were calculated with PRSice2 (**Methods**) and their performance tested in three independent datasets: (i) the Norwegian Trøndelag Health Study (HUNT; n=69,291; 977 cases vs. 68,314 controls), (ii) the Danish Blood Donor Study (DBDS; n=56,397; 1,754 cases vs. 54,643 controls), and (iii) 1,144 HEM cases from gastroenterology clinics compared to 2,740 cross-sectional population controls from Germany (**Supplementary Table 1**). In all three datasets, HEM patients showed significantly higher PRS values compared to controls (OR=1.24, *P*=1.98×10^−11^; OR=1.28, *P*=3.52×10^−22^, and OR=1.36, *P*=7.64×10^−15^, respectively for the two population cohorts HUNT and DBDS, and the German case-control cohort). In HUNT and DBDS, HEM prevalence increased across PRS percentile distributions, with individuals from the top 5% tail being exposed to higher HEM risk as compared to the rest of the population (OR=1.68, *P*=1.55×10^−5^, and OR=1.68, *P*=6.11×10^−8^, respectively for HUNT and DBDS, **Figure 5**). Higher HEM PRS were also associated with a more severe phenotype as defined by the need for recurrent invasive procedures (OR=1.03, *P*=8.63·10^−3^ in German patients) and a younger age of onset (*P*=1.90×10^−3^ in DBDS; *P*=4.01×10^−3^ in German patients).

**Figure 5:**
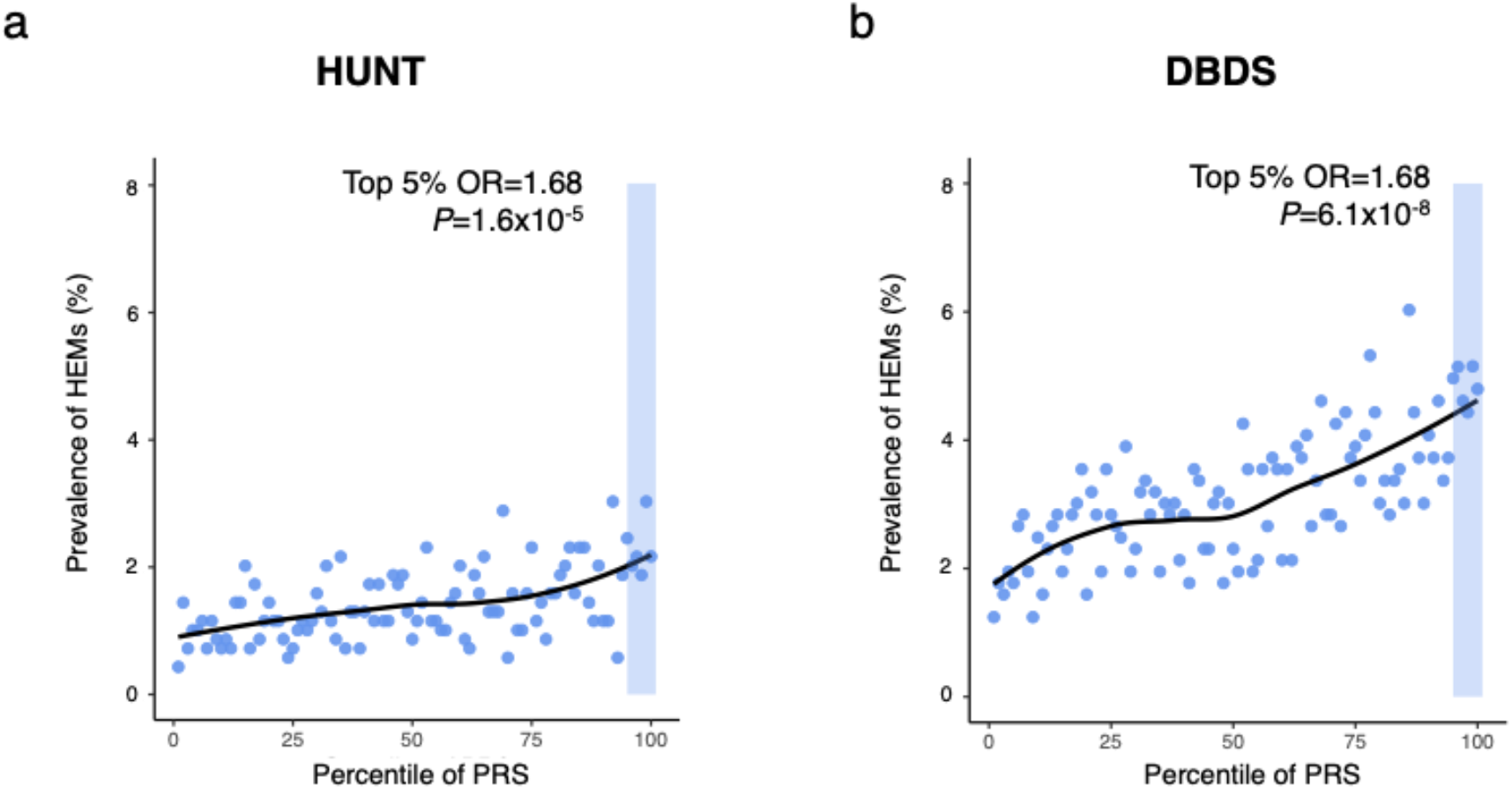
Risk HEM prevalence across polygenic risk score (PRS) percentile distributions. PRS was derived from the results of the association meta-analysis (see **Methods**). HEM prevalence (%, Y axis) is reported on a scatter plot in relation to PRS percentile distribution (X-axis) in the Norwegian Trøndelag Health Study (HUNT) (a) and the Danish Blood Donor Study (DBDS) (b) population cohorts. The top 5% of the distribution is highlighted with a shaded area in both cohorts, and the results of testing HEM prevalence in this group vs the rest of the population is also reported (P-value and OR from logistic regression; **Methods**).

### FUNCTIONAL ANNOTATION OF GWAS LOCI, TISSUE AND PATHWAY ENRICHMENT ANALYSES

In order to identify most likely candidate genes, molecular signaling pathways and potential pathophysiological causes underlying the identified association signals, we used independent computational pipelines for the functional annotation of GWAS results. This yielded a total of 819 non-redundant HEM-associated transcripts (hereafter referred to as HEM genes; **Supplementary Table 7**) derived from alternative positional (N=540 total from FUMA, DEPICT and MAGMA) and expression quantitative trait (eQTL, N=562 from FUMA) mapping efforts (**Methods**). Tissue-specific enrichment analyses (TSEA) of 540 positional candidates led to very similar results for all three approaches, with enrichment of HEM gene expression in blood vessels, colon and other relevant tissues (**Supplementary Figure 5** and **Supplementary Table 8**). Similarly, gene-set enrichment analyses (GSEA) highlighted common pathways important in the development of vasculature and the intestinal tissue including the gene ontology (GO) terms “tube morphogenesis and development”, “artery morphogenesis and development”, “epithelium morphogenesis”, “smooth muscle tissue morphogenesis” and others (**Supplementary Figure 5** & **Supplementary Table 8**). Additionally, DEPICT detected enrichment for a number of traits from the Mammalian Phenotype Ontology including “abnormal intestinal morphology”, “rectal prolapse”, “abnormal blood vessel (and artery) morphology”, and “abnormal smooth muscle morphology and physiology” (**Supplementary Table 8**). TSEA and GSEA analyses performed on all 819 transcripts including eQTL genes from FUMA gave rise to similar results although these did not reach statistical significance (not shown).

### GENE EXPRESSION IN HEMORRHOIDAL DISEASE TISSUE AND GENE-NETWORK ANALYSES

The expression of HEM genes was studied in integrated mRNA and microRNA Combo-Seq analysis of enlarged hemorrhoidal tissue from 20 HEM patients and normal specimens from 18 controls (**Methods** and **Supplementary Table 1**). HEM genes were examined with regard to their expression status, differential expression between cases and controls, and their connectivity and topology in gene co-expression networks. After normalization for cell-type heterogeneity in different tissues (**Methods**), 720 out of 819 candidate genes were found to be expressed in hemorrhoidal tissue, with 287 (39.9%) of these being among the most strongly expressed transcripts (upper quartile) (**Supplementary Table 7**). Compared to normal tissue from controls, 18 HEM candidate genes from 14 independent loci showed differential expression in hemorrhoidal tissue (*P*_FDR_<0.05 and |log_2_ fold change [FC]|>0.5), with 12 genes showing increased and 6 decreased expression (**Figure 6a**). To obtain further biological insight from transcriptomic profiling, HEM candidate genes were further characterized for their membership in co-expressed gene modules identified via weighted gene correlation network analysis (WGCNA) of the normalized whole transcriptome data (**Figure 6b; Methods**). The final network consisted of 36,342 genes partitioned into 41 co-expression modules, and 3 of these (M1, M4 and M7; referred to as HEM modules) were significantly enriched (*P*_FDR_<0.05) for HEM genes (**Figure 6c**). In total, 260 (35.7%) expressed HEM genes were members of modules M1 (n=121), M4 (n=75) or M7 (n=64) (**Supplementary Table 9**). Functional annotation of these modules revealed M1 enrichment for “extracellular matrix (ECM) organization” (*P*_FDR_=5.17×10^−18^) and “muscle contraction” (*P*_FDR_=7.90×10^−17^); M4 for “mitochondrion organization” (*P*_FDR_=2.35×10^−4^) and “glycosylation” (*P*_FDR_=5.70×10^−4^); and M7 for “cornification” (*P*_FDR_=3.97×10^−52^) and “epidermis development” (*P*_FDR_=2.07×10^−41^) (**Figure 6c**). The WGCNA analysis also allowed for the identification of the most interconnected genes, or module hub genes (i.e. central nodes in the scale-free network). Of note, several HEM candidate genes, namely *NEGR1, MRVI1, MYH11, ELN* and *CHRDL1*, were among the top 50 hub genes for module M1 (**Figure 6d**). M1 is the module most significantly enriched for HEM genes (*P*_FDR_=6.40×10^−5^), and also the largest (n=3,975) co-expression module in the hemorrhoidal tissue (**Supplementary Table 9**), thus pointing to the importance of its associated GO terms “ECM organization” and “muscle contraction” in HEM.

**Figure 6:**
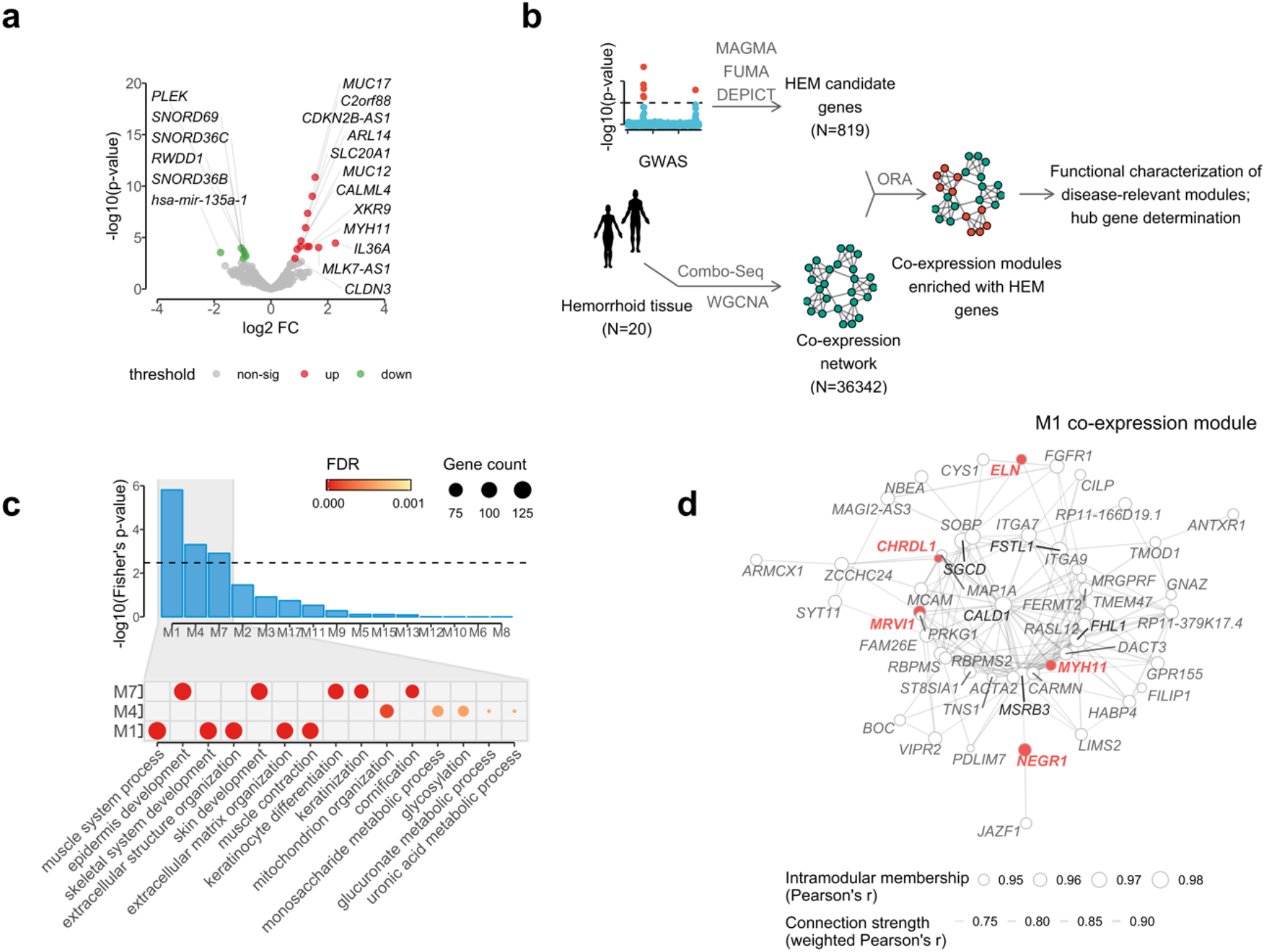
Analysis of mRNA and microRNA (Combo-Seq) data from HEM affected tissue, in relation to HEM genes co-expression networks. **(a)** Volcano plot reporting HEM genes differentially expressed in hemorrhoidal tissue (significantly up-regulated = red, and down-regulated = green); (**b**) schematic representation of the analytical flow for HEM genes co-expression network module identification and characterization; **(c)** upper panel (barplot): Overrepresentation analysis (ORA) of HEM genes in co-expression network modules, with significant enrichment (*P*_FDR_<0.05) in modules M1, M4 and M7; lower panel (dotplot): top 5 GO terms (Biological Process) from gene set enrichment analysis (GSEA) relative to M1, M4 and M7 co-expression modules (gene counts and FDR-adjusted significance level are also reported as indicated); **(d)** Co-expression hub network of module M1. The network represents strength of connections (weighted Pearson’s correlation >0.7) among the top 50 hub genes with highest values of intramodular membership (size of the node). HEM genes and the top 5 hub genes are highlighted in red and black, respectively.

### PRIORITIZED HEM GENES

In order to identify genes most likely to play a causative role in HEM, we selected candidates based on a scoring approach by prioritizing those associated with one or more of the following: (i) linked to a high-confidence fine-mapped variant (posterior probability [PP] >50%), (ii) differentially expressed in enlarged hemorrhoidal tissue, (iii) highlighted by pathway and tissue/cell-type enrichment DEPICT analysis (**Methods**), or (iv) predicted hub of a WGCNA co-expression HEM module (M1, M4, M7). This reduced the number of candidates from 819 to 100 prioritized genes associated with 58 independent HEM loci (priority genes could not be assigned to several loci based on the above selection criteria) (**Supplementary Table 7**). Some notable observations were made in relation to a subset of these prioritized genes, whose associated evidence and known biological function(s) make them remarkably good candidates to play a role in HEM risk.

Two genes, *ANO1* and *SRPX*, were both linked to a single coding variant fine mapped as causal with very high confidence (rs2186797, *ANO1*, p.Phe608Ser with PP=97.0%, and rs35318931, *SRPX*, p.Ser413Phe with PP=87.3%, respectively). *ANO1* encodes the voltage-gated calcium-activated anion channel anoctamin-1 protein, which is highly expressed in the interstitial cells of Cajal throughout the human gastrointestinal tract, where it contributes to the control of intestinal motility and peristalsis^6^. The *ANO1*:p.Phe608Ser variant is predicted to destabilize local protein structure and to disturb ANO1-activating phospholipid interactions (**Supplementary Figure 6**) and may therefore hamper anoctamin-1 function(s) and hence gut motility. The X-linked gene *SRPX* codes for a Sushi repeat-containing protein whose domain composition implies a role in ECM, and the ECM of various tissues including colon and liver express the protein^7^. The *SRPX*:p.Ser413Phe variant (rs35318931) may potentially destabilize the C-terminal domain of unknown function (DUF4174) (**Supplementary Figure 7**), which is conserved in various ECM proteins (**Supplementary Note**, section ***In silico* variant protein analysis**).

**Figure 7:**
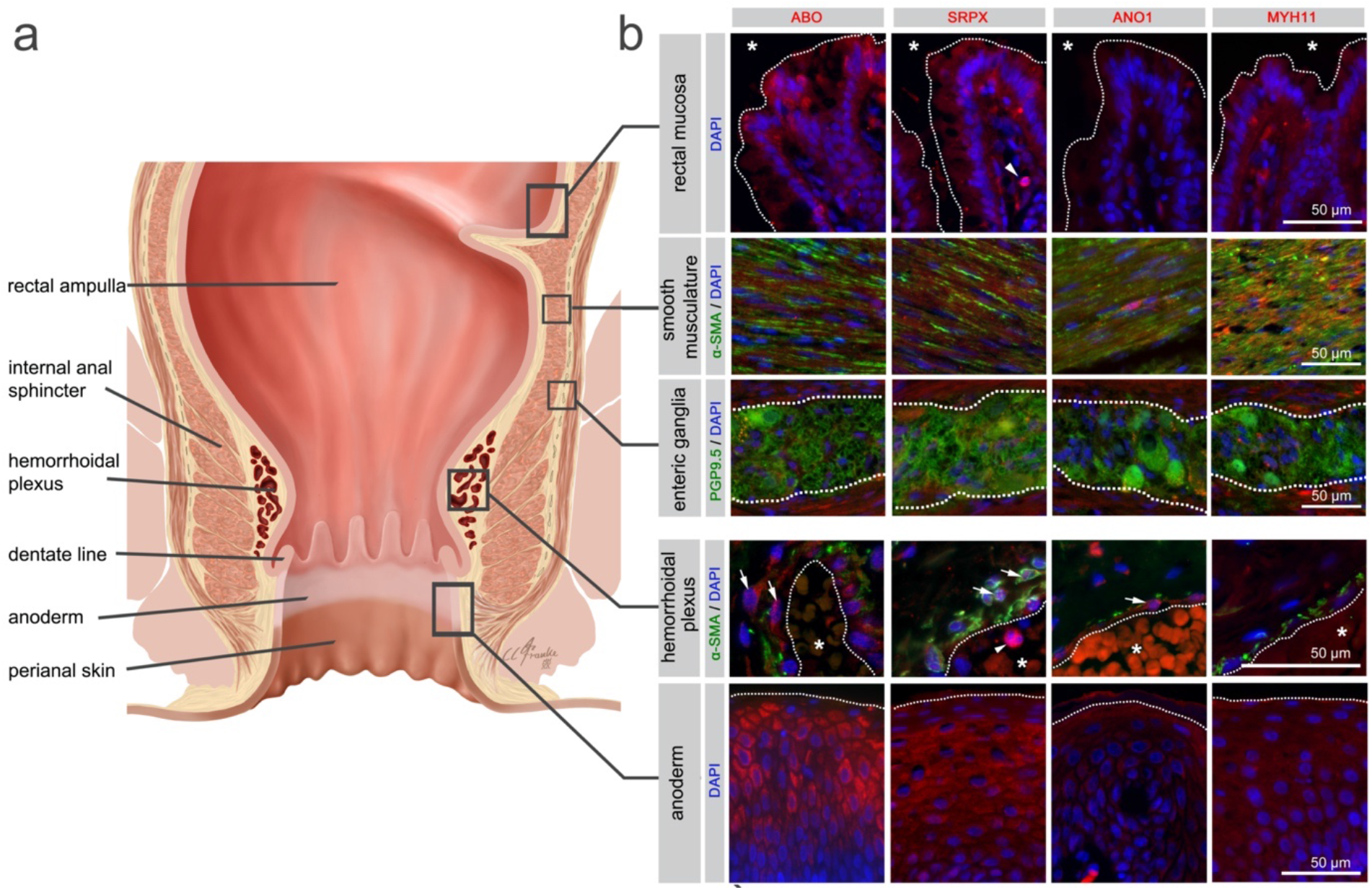
Immunohistochemistry for selected HEM candidate proteins. Illustration of the rectum and anal canal **(a)** indicating the site-specific localization of the immunohistochemical panels analyzed in **(b)**. Results of fluorescence immunohistochemistry are shown for selected HEM candidate proteins encoded by HEM genes *SRPX* (rs35318931), *ANO1* (rs2186797), *MYH11* (rs6498573) and *ABO* (rs676996) (see also **Supplementary Table 11** and **12**). Antibody staining was performed on FFPE anorectal tissue specimens from control individuals. Picture layers correspond to the rectal mucosa (top row, epithelial surface delimited by a white dotted line,*=intestinal lumen), smooth musculature (second row), enteric ganglia (third row, ganglionic boundaries delimited by a white dotted line), hemorrhoidal plexus (fourth row, endothelial surface delimited by a white dotted line, *=vascular lumen), and the anoderm (bottom row, surface of the anoderm delimited by a white dotted line). Blue: DAPI; green: α-SMA (anti-alpha smooth muscle antibody) for row 2 and 4 (smooth musculature/hemorrhoidal plexus) and PGP9.5 (member of the ubiquitin hydrolase family of proteins, neuronal marker) for row 3 (enteric ganglia); red: antibody for the respective candidate protein. Arrows point to respective candidate-positive cells within the vascular wall. Arrowheads point to respective candidate-positive nucleated immune cells.

At locus 7q22.1 the signal was fine-mapped with very high confidence to rs4556017 (PP=96.2%), which exerts eQTL effects on *ACHE* and *SRRT*, both showing high levels of expression in enlarged hemorrhoidal tissue, and found in the M4 and M7 co-expression modules (**Figure 6**), respectively. However, while *SRRT* encodes a poorly characterized capped-RNA binding protein, *ACHE* appears a much better candidate as the gene encodes an enzyme that hydrolyzes the neurotransmitter acetylcholine at neuromuscular junctions, and corresponds to the Cartwright blood group antigen Yt^8,9^. One more locus was fine mapped to single-variant resolution with high confidence, SNP rs10956488 from locus 8q24.21 (PP=0.962), which is linked to an eQTL for *GSDMC*. Gasdermin C (encoded by *GSDMC*) is a poorly characterized member of the gasdermin family of proteins expressed in epithelial cells and in enlarged hemorrhoidal tissue, though the mechanisms of its eventual HEM involvement remain elusive.

Other genes were linked via eQTL to a single variant mapped with posterior probability >50% and were prioritized based on additional experimental evidence. Among these, the fine mapped SNP rs6498573 from locus 16p13.11 (PP=63.2%) is associated with eQTL effects on *MYH11* (encoding muscle myosin heavy chain 11; **Supplementary Table 10**), a gene coding for a smooth muscle myosin heavy chain that shows mRNA upregulation in enlarged hemorrhoidal tissue and constitutes a hub for the M1 co-expression module associated with ECM organization and muscle contraction. Notable prioritized candidates were also observed at loci that were not fine mapped, including *ELN* (encoding elastin, a key component of the ECM found in the connective tissue of many organs, highly expressed in enlarged hemorrhoidal tissue and hub gene for the M1 co-expression module), *COL5A2* (encoding Type V collagen protein; highly expressed in HEM tissue and belong to M1 co-expression module), *PRDM6* (encoding a putative histone methyltransferase regulating vascular smooth muscle cells contractility, expressed in enlarged hemorrhoidal tissue and hub for the M1 module), and others (**Supplementary Table 7**).

Finally, while no candidate genes could be highlighted from the top GWAS hit region on chr12q14.3 (rs11176001), both the second and third strongest GWAS signals were detected at loci linked to genes involved in the determination of blood groups, namely *ABO* (rs676996) and the Kell Blood Group Complex Subunit-Related Family member *XKR9* (rs1838392). In addition, blood group antigens are encoded at additional loci, including *ACHE* (rs4556017) and *XKR6* (identified by MAGMA). Imputation of human ABO blood types from genotype data revealed the O type to be associated with increased and A and B types decreased HEM risk, both in UKBB and GERA datasets (**Supplementary Figure 8**).

### LOCALIZATION OF SELECTED HEM GENE ENCODED PROTEINS

A site-specific analysis of selected candidate proteins for their localization in anorectal tissues underlined the complex multifactorial nature of cellular components potentially involved in the pathogenesis of HEM. Indeed, analyzed candidates displayed a broad spectrum of expression in intestinal mucosal, neuromuscular, immune, and anodermal tissues (**Figure 7, Supplementary Figure 9** and **10, Supplementary Table 11**). They were also directly co-localized with hemorrhoidal blood vessels, suggesting a putative role connected with the hemorrhoidal vasculature itself.

## Discussion

Given the lack of large and systematic epidemiological and molecular studies, HEM can be regarded as an understudied disease in terms of evidenced etiopathogenesis despite of its worldwide distribution. In this study, we report the largest and most detailed genome-wide analysis of HEM, demonstrating for the first time that HEM is a partly inherited condition with a weak but detectable heritability estimated at 5% based on SNP data. We identified 102 independent risk loci, which we functionally annotated based on computational predictions and gene expression analysis of diseased and normal tissue. We provide important novel pathophysiological insight, which we discuss below in relation to individual pathways and mechanisms proposed to contribute to HEM etiology.

### Extracellular matrix, elasticity of the connective tissue and smooth muscle function

The non-vascular components of the anal cushions consist of the transitional epithelium, connective tissue (elastic and collagenous), and the submucosal anal muscle (muscle of Treitz)^10^. Treitz’s muscle tightly maintains the anal cushions in their normal position, and its deterioration is considered one of the most important pathogenetic factors in the formation of enlarged and prolapsed hemorrhoids (**Supplementary Figure 2**). Anal cushion fixation is further facilitated by elastic and collagenous connective tissue, whose degeneration, due, for instance, to abnormalities in collagen composition, has been involved in HEM etiology^11^, although without strong molecular evidence.

The co-expression module M1 identified from HEM tissue is linked to ECM organization and muscle function and is enriched for HEM gene thus providing novel important evidence for a role of these two interconnected processes in HEM pathogenesis. *ELN* (lead SNP rs11770437) is one of the HEM prioritized genes, and also a main hub gene for the M1 module. *ELN* codes for the elastin protein, a key component of elastic fibers that comprise part of the ECM and confer elasticity to organs and tissues including blood vessels. Mutations in the *ELN* gene have been shown to cause *cutis laxa*, a disease in which dysfunctional elastin interferes with the formation of elastic fibers, thus weakening connective tissue in the skin and blood vessels. Of note, *ELN* has been recently implicated also in diverticular disease^12^, a condition characterized by outpouchings of the colonic wall (pseudodiverticula) at sites of relative weakness and/or defective elasticity of the connective and muscle layers. Hence, similar mechanisms may underly HEM risk due to genetic variation in *ELN* and, notably, also the *SRPX* (lead SNP rs35318931) and *COL5A2* (lead SNP rs16831319) genes. The *SRPX* lead SNP is an intriguing candidate that should be further tested for causality as it is a coding variant (SRPX:p.Ser413Phe) that potentially impacts the function of an ECM protein. This marker has also been identified as the second strongest GWAS signal, after *ELN*, in a large study of the common skin condition nonsyndromic *striae distensae* (NSD, also known as stretch marks), whose manifestation is due to lost tissue elasticity at affected skin sites^13^. Although SRPX function is still unknown, its dual predicted role in HEM and NSD points to a similar mechanistic involvement in these conditions. *COL5A2* codes for type V fibril-forming collagen that has regulatory roles during development and growth of type I collagen-positive tissues. Mutations in this gene are known to cause Ehlers-Danlos syndrome (EDS), a rare connective tissue disease that affects the skin, joints and blood vessels, and for which a link to HEM has already been postulated^14^. An additional hub gene for the co-expression module M1, *MYH11* (lead SNP rs6498573) encodes a smooth muscle myosin protein that is important for muscle contraction and relaxation, and whose dysfunction has been linked to vascular diseases^15^ and gastrointestinal dysmotility^16^. Functional studies in smooth muscle cells showed that overexpression of *MYH11* led to a paradoxical decrease of protein levels through increased autophagic degradation followed by disruption of contractile signaling^17^. Our transcriptome analysis also showed *MYH11* RNA up-regulation in HEM tissue, thus suggesting possible alterations in smooth muscle action that may be relevant, for instance, to the Treitz’s muscle function(s). Additional evidence for the involvement of the muscoskeletal system may come from the associations detected for *GSDMC* (lead SNP rs10956488), an uncharacterized gene also shown to be relevant to lumbar disc herniation and back pain^18,19^ and *PRDM6*, a histone methyltransferase that acts as a transcriptional repressor of smooth muscle gene expression. Finally, besides individual association signals, we detected significant genetic correlation with several other complex diseases of shared etiology (hernia, dorsalgia [back pain]), for which connective tissue and/or muscle alterations are described, further supporting the involvement of these pathways in HEM pathophysiology.

### Gut motility

Several lines of evidence link gut motility to the pathophysiology of HEM in this study. In our UK Biobank analyses, HEM patients were found to suffer more often from IBS and other dysmotility syndromes than controls, as evidenced also by the increased use of medications including laxatives (and possibly Omeprazole and Lansoprazole, likely treatment proxies for other FGIDs). These conditions also showed strongest genetic correlation with HEM among all tested traits, indicating similar genetic architecture and predisposing mechanisms. Moreover, given the important role of the gut-brain axis in IBS and other FGIDs^20^, it is possible that the correlations observed for anxiety, depression and other neuroaffective traits may mediate genetic risk effects at least in part via similar mechanisms also involving gut motility. Constipation and prolonged sitting and straining during defecation are associated with delayed gastrointestinal transit and reduced peristalsis and are among the proposed HEM risk factors (summarized in **Supplementary Figure 2)**. Harder stools, due to for instance to infrequent defecations, can cause difficulty in bowel emptying and therefore increase pressure and mechanical friction on the hemorrhoidal cushions, leading to excessive engorgement and stretching or tissue damage. The relationship between HEM and gut motility is probably best evidenced by the association signal detected at the *ANO1* locus: its lead SNP rs2186797 corresponds to the missense variant *ANO1*:p.Phe608Ser that was fine-mapped with very high confidence and predicted to impact anoctamin-1 functional properties. Anoctamin-1 is an ion channel expressed in the interstitial cells of Cajal, the pacemakers of the gastrointestinal tract controlling intestinal peristalsis, has already been implicated in IBS^21^, and hence represents an ideal candidate to also affect HEM risk via genotype-driven modulation of gut motility. An additional interesting candidate is *ACHE*, which shows an eQTL with the lead SNP rs4556017, a good variant to be further tested for causality, from the locus based on fine-mapping results. *ACHE* codes for an enzyme that hydrolyzes the neurotransmitter acetylcholine at neuromuscular junctions and is overexpressed in Hirschsprung’s disease, a condition in which gut motility is compromised due to the absence of nerve cells (aganglionosis) in the distal or entire segments of the large bowel^22^. Of interest, expression in the enteric ganglia next to the hemorrhoidal plexus was observed for several proteins encoded by HEM genes in our immunofluorescence experiments.

### Vasculature and circulatory system

Previous observations showed that HEM are not varicosities and accelerated blood flow velocities were observed in afferent vessels of HEM patients^23^. An impaired drainage or filling of the anal cushion may contribute to cushion slippage and may thus be considered as one of many disease-causing factors, as already previously proposed^23^. Our genetic data support the involvement of the vasculature as an important player in HEM pathophysiology. TSEA and GSEA results downstream of HEM GWAS meta-analysis highlight blood vessels and artery morphogenesis among the HEM gene-enriched tissues and GO pathways, respectively. At the same time, moderate genetic correlation is detected for diseases of the circulatory system in the LDSC analyses. We identified a very strong association signal in correspondence of the ABO locus on chromosome 9, which determines the corresponding ABO blood group type (A, B, AB and O). In addition to red blood cells, ABO antigens are expressed on the surface of many cells and tissues, and have been strongly associated with coronary artery disease, thrombosis, hemorrhage^24^, gastrointestinal bleeding^25^ and other conditions related to the circulatory system. Interestingly, increased risk for coronary artery disease has been reported in HEM patients in at least some studies^26^ and replicated in our UK Biobank cross-disease analyses although amidst other hundreds of associations of similar magnitude of effects. We imputed ABO blood types from genotype data in UK Biobank, and detected increased HEM risk for carriers of the O type. However, O type has been reported to be protective for coronary artery disease in UK Biobank, albeit also predisposing to hypertension^27^. Hence the potential mechanism(s) by which variation at this locus impacts HEM risk remain elusive at this stage. Blood antigens are nevertheless likely relevant, as other genes involved in the determination of specific blood groups are also among the 102 HEM GWAS hits (Kell blood group locus *XKR9*, lead SNP rs1838392).

In summary, our data provide important new insight into HEM pathogenesis. This sets the stage for more detailed genetic and mechanistic follow-up analyses, the search for therapeutically actionable genes and pathways, and the eventual exploitation for the adoption of preventive measures based on computed individual predisposition.

## Data Availability

The data supporting the findings described in this study are available from the corresponding author upon request.

## Acknowledgements

This project was funded by Andre Franke’s and Clemens Schafmayer’s DFG grant “Discovery of risk factors for hemorrhoids” (ID: FR 2821/19-1). The study received infrastructure support from the DFG Cluster of Excellence 2167 “Precision Medicine in Chronic Inflammation (PMI)” (DFG Grant: “EXC2167”). The project was supported by grants from the Swedish Research Council to MD (VR 2017-02403), the Novo Nordisk Foundation (grants NNF17OC0027594 and NNF14CC0001) and BigTempHealth (grant 5153-00002B). We are indebted to the valuable assistance by Tanja Wesse (Genotyping of the German samples) and Petra Röthgen (German DZHK control data set). We thank Clemens Franke (Institute of Anatomy, Christian-Albrechts-University of Kiel, Kiel, Germany) for graphic assistance (Fig. 7, Suppl. Fig. 1). This research has been conducted using the UK Biobank Resource under Application Number 31435.

The work on cross-trait analysis for diverticular disease presented in this manuscript was supported by the German Research Council (DFG, ID: Ha3091/9-1) and the Austrian Science Fund (FWF, ID: I1542-B13). Data access to the UK biobank data for diverticular disease was granted under project numbers 22691.

EGCUT work has also been supported by the European Regional Development Fund and grants SP1GI18045T, No. 2014-2020.4.01.15-0012 GENTRANSMED and 2014-2020.4.01.16-0125 This study was also funded by EU H2020 grant 692145, Estonian Research Council Grant PUT1660. Data analyzes with Estonian datasets were carried out in part in the High-Performance Computing Center of University of Tartu.

We would like to thank the research participants and employees of 23andMe for making this work possible. The following members of the 23andMe Research Team contributed to this study: Michelle Agee, Adam Auton, Robert K. Bell, Katarzyna Bryc, Sarah L. Elson, Pierre Fontanillas, Nicholas A. Furlotte, David A. Hinds, Karen E. Huber, Aaron Kleinman, Nadia K. Litterman, Jennifer C. McCreight, Matthew H. McIntyre, Joanna L. Mountain, Elizabeth S. Noblin, Carrie A.M. Northover, Steven J. Pitts, J. Fah Sathirapongsasuti, Olga V. Sazonova, Janie F. Shelton, Suyash Shringarpure, Chao Tian, Joyce Y. Tung, and Vladimir Vacic.

Vladimir Vacic and Olga V. Sazonova are/were employed by and hold stock or stock options in 23andMe, Inc..

## Author contributions

A.F., C.S. and M.D. were responsible for the concept and the design of the study. C.S. and G.B. coordinated the recruitment of the German clinical cohort. V.K., J.G., I.V., B.S., H.G.P., A.H., J.J., J.W., T.B., M.D. F.B., S.H., J.K., J.S., W.K., M.Z., C.G.N., T.L., J.T., and W.S. recruited and phenotyped patients of the clinical cohorts. V.K. collected and phenotyped the biopsy samples that were used in the immunohistochemistry and transcriptome analyses. Animal samples for histology were taken and provided by K.P., S.Z., L.F., T.G., F.H.L. and K.M.. Immunohistochemistry and histology analyses were performed by F.C. and T.W. with support of J.W.. T.Z., D.E., and F.D. performed genotype and phenotype data collection. T.Z., D.E., and F.D. performed GWAS data quality control and imputation. S.J. performed the transcriptome (RNA-Seq) analysis. F.U.W, S.B., and M.F. helped with bioinformatic analyses. I.F.J., K.B., S.B., C.L.R., C.E., O.B.P., and H.U. conducted analyses in DBDS and DNPR. H.C., V.L, R.J., N.F., W.L., M.L., and U.N.-G. contributed German control data. S.B., C.D., H.V., A.G. and J.H. contributed the GWAS data on diverticular disease. Norwegian HUNT data was contributed by A.H.S., T.H.K., M.E.G., L.T., E.N. and K.H.. The Estonian dataset was provided by T.E. and M.T. Protein models and analyses were performed by G.M.. The analysis of the Michigan Genomics Initiative data set was performed by M.Z., B.V., A.P. and L.G.F.. The 23andMe data set was analyzed and provided by O.S., E.S.N., and V.V.. M.W. implemented ABO blood-group inference. T.Z., D.E., implemented statistical models and performed the (meta-)analysis. T.Z., D.E., and S.J. curated and interpreted results. M.S. designed Supplementary Figure 2 with scientific input from N.M. and A.F.. The GWAS data browser was implemented by M.R. and set up by G.H.. A.F., M.D. and D.E. wrote the manuscript draft with substantial contributions from T.Z. and S.J.. All authors reviewed, edited and approved the final manuscript.

## Methods

### Study cohorts and patients’ material

#### 23andMe

The 23andMe study dataset contains participants drawn from the research participant base of the personal genetics company, 23andMe, Inc^28,29^. Genetic data and comprehensive phenotypic information from health surveys were available for 402,845 unrelated individuals of European ancestry. For details on genotyping, quality control, genotype imputation and GWAS association testing see **Supplementary Note**. Study participants were divided into HEM cases and controls based on their self-completed HEM health questionnaires, resulting in 174,785 HEM cases and 228,060 controls in the current 23andMe GWAS. Demographic data of 23andMe samples are reported in **Supplementary Table 1**. Participants provided informed consent and participated in the research online, under a protocol approved by the external AAHRPP-accredited IRB, Ethical & Independent Review Services (E&I Review). The full GWAS summary statistics for the 23andMe discovery data set will be made available through 23andMe to qualified researchers under an agreement with 23andMe that protects the privacy of the 23andMe participants. Please visit https://research.23andme.com/dataset-access for more information and to apply to access the data.

#### UK Biobank (UKBB)

The UKBB is a large population-based study in the United Kingdom with extensive phenotypic and genotype data from approximately 500,000 participants^30^. Each individual underwent cognitive, physical assessment and sampling for DNA collection when enrolled, and health-related information was collected including data from their electronic health records (EHRs). The diagnoses in the EHRs are coded in the terminology of the International Statistical Classification of Diseases and Related Health Problems (ICD) terminology. For this GWAS study we included 408,592 individuals of European ancestry (self-reported “white” and of genetic Caucasian descent; **Supplementary Note**). Of these, 23,856 samples met our criteria for HEM cases (either ICD10 code I84 or ICD9 code 455 in the medical records). The other part of the cohort (n=384,736) served as study controls. The demographic data of the individuals are reported in **Supplementary Table 1**. UKBB received ethical approval from the competent Research Ethics Committee (REC reference 11/NW/0382) and the project’s Application ID is 31435.

#### Estonian Genome Project of University of Tartu (EGCUT)

The Estonian Biobank is a population-based cohort of the Estonian Genome Center at the University of Tartu (EGCUT), Estonia, with a current size of app. 200,000 participants aged over 18^31^. The whole project is conducted according to the Estonian Gene Research Act and all participants have signed the broad informed consent. Upon recruitment, the biobank participants filled out a detailed questionnaire, covering lifestyle, diet and clinical diagnoses (described by ICD10 codes). In this study, individuals with any entry of the ICD10 code for HEM (I84) were included as HEM cases. Further, we selected 30,441 controls with genome-wide data as study controls, resulting in 6,956 HEM cases and 30,441 population controls (**Supplementary Note)**. The demographic data of the individuals are reported in **Supplementary Table 1**. This study has been reviewed and approved by the Estonian Committee on Bioethics and Human Research.

#### Michigan Genomics Initiative (MGI)

The Michigan Genomics Initiative (MGI) is a longitudinal cohort of participants in Michigan Medicine, USA^32^. MGI participants were recruited primarily through surgical procedures at Michigan Medicine and gave consent for link their EHRs and genetic data for research purposes. We used a current data freeze of 40,000 European individuals for GWAS analysis (**Supplementary Note**). Of these, 4,539 HEM cases were defined based on a review of EHRs (either ICD10 code I84, ICD10-CM code K64 or ICD9 code 455). The rest of the cohort with genome-wide data was defined as study controls (n=35,338). The demographic data of the individuals are reported in **Supplementary Table 1**. This study has been reviewed and approved by the Michigan Institutional Review Board.

#### Genetic Epidemiology Research on Aging

The Genetic Epidemiology Research on Aging (GERA) Cohort comprises more than 100,000 adults who are members of the Kaiser Permanente Medical Care Plan, Northern California Region (KPNC), USA. The health status of participants in the GERA cohort was assessed using EHRs collected at Kaiser Permanente’s facilities in Northern California from January 1, 1995 to March 15, 2013. HEM cases (n=8,813) were those in which at least two ICD9 code diagnoses of HEM (ICD9 code 455) were recorded on separate days. Their genome-wide data were compared with those of the remaining cohort (n=46,780) as controls (**Supplementary Note**). The demographic data of the individuals are reported in **Supplementary Table 1**. The GERA data access was applied for on the dbGaP website (dbGaP Study Accession: phs000674.v3.p3) and the study was approved by the dbGap Access Review Committee.

#### German case-control cohort

Initiated by the Department of General and Thoracic Surgery and the biobank PopGen^33^ of the Medical Faculty of Kiel University, Kiel, Germany, a cohort of HEM patients with symptomatic hemorrhoids and the need of invasive treatment was newly established. Between January 2016 and December 2017, individuals with a prior diagnosis of high-grade hemorrhoids were identified based on the medical records of five hospitals and practices in the North German region using German procedural codes (OPS-301 by German Institute for medical Documentation and Information). The main inclusion criteria were the need for hemorrhoidectomy or invasive treatment (rubber band ligation, sclerotherapy) on high grade hemorrhoidal disease, verified by DRG-code (Diagnosis related Groups). Patients receiving exclusively conservative treatment were not included in this study as the aim was to recruit patients with a strong phenotype of advanced hemorrhoidal disease. The cohort included 1,007 patients undergoing surgical/invasive treatment of a high grade hemorrhoidal disease. In total, 1,144 cases and 2,740 controls were available for PRS analysis (**Supplementary Note** and section **Polygenic risk score (PRS) analysis, Methods**). The demographic data of the individuals are reported in **Supplementary Table 1**. The study protocol was approved by the ethics committee of the Medical Faculty of Kiel University and written informed consent was obtained from all study participants.

#### The Trøndelag Health Study (HUNT)

The Trøndelag Health Study (HUNT) is a large population-based cohort from the county Nord-Trøndelag in Norway. All residents in the county, aged 20 years and older, have been invited to participate. Data was collected through three cross-sectional surveys, HUNT1 (1984-1986), HUNT2 (1995-1997) and HUNT3 (2006-2008), and has been described in detail previously^34^, with the fourth survey recently completed (HUNT4, 2017-2019). All genotyped participants have signed a written informed consent regarding the use of data from questionnaires, biological samples and linkage to other registries for research purposes. Cases were defined as having an ICD10 K64 diagnosis and the reminder of the cohort were used as controls. In total, 977 cases and 68,314 controls were available for PRS analysis (**Supplementary Note** and **Polygenic risk scores (PRS) analysis, Methods**). The demographic data of the individuals are reported in **Supplementary Table 1**.

#### Danish Blood Donor Study

The Danish Blood Donor Study (DBDS) is a large prospective cohort of nation-wide Danish blood donors (n=56,397) and comprises both extensive phenotype data as well as genome-wide genotyping data^35,36^. HEM cases are defined using the ICD-8 code 455 or ICD-10 codes I84 or K64, resulting in 1,754 cases in the DBDS cohort as registered in the National Patient Registry (**Supplementary Note**). In total, 1,754 cases and 54,643 controls were available for genome-wide polygenic risk score (PRS) analysis. The demographic data of the individuals are reported in **Supplementary Table 1**. This study was approved according to the Danish Blood Donor study protocol (ref: 1700407) as a part of “Genetics of healthy ageing and specific diseases among blood donors”.

#### Danish National Patient Registry

The Danish National Patient Registry (DNPR) is a population-wide registry containing all diagnoses made in hospitals in Denmark from 1977 to 2018 and includes more than 8 million patients. The diagnoses in the registry are coded in the terminology of the International Statistical Classification of Diseases and Related Health Problems (ICD) 8th Revision (1997-1993) or 10th Revision (1994-2018) terminology. All patients with a hemorrhoid disease code in the disease registry were identified. In the ICD-8 period patients with ‘Hemorrhoids’ are recognized using the code 455. As the ICD-10 code for hemorrhoids changed in 2013, we combined patients diagnosed with ‘Hemorrhoids’ (ICD-10: I84) from 1994-2012 and patients diagnosed with ‘Hemorrhoids and perianal venous thrombosis’ (K64) from 2013-2018. Information about drugs administered in hospitals is available for more than 1.6 million patients in the period 2006-2016 and is defined using the Anatomical Therapeutic Chemical (ATC) Classification System of the WHO. We integrated data from two different electronic medication modules corresponding to the administrative databases for hospital internal drug consumption from two health regions of Denmark (Capital Region and Region Zealand): OPUS-medicin and Elektronisk patient medicinering^37^. The demographic data of the individuals are reported in **Supplementary Table 1**. This DNPR study has been approved by the Danish Data Protection Agency, Copenhagen (ref: FSEID-00003092, FSEID-00003724 and 3-3013-1731/1).

#### Hemorrhoidal tissue

A group of 38 individuals undergoing surgery for hemorrhoids (n=20; cases) and anal fissures (n=18; controls) was included in this study. Hemorrhoidal tissue samples were obtained either by Milligan Morgan open hemorrhoidectomy or by stapled hemorrhoidopexy for grade (Goligher) 3 and 4 hemorrhoids. Approximately 1cm^3^ of hemorrhoidal tissue was obtained from Milligan-Morgan-specimens just above the dentate line. In hemorrhodopexy patients, approximately 1cm^3^ of biopsies were taken from the “doughnut” tissue at 3 o’clock in the prone position. Healthy hemorrhoid tissue samples were taken from adjacent zones (1-2 cm above the dentate line) of anal fissures. Clinical and demographic data for the sampled individuals are listed in **Supplementary Table 1**. This study was approved by the bioethical committee of medical faculty, University Hospital Schleswig-Holstein Kiel, Germany. All participants provided written informed consent.

### GWAS meta-analysis across discovery cohorts

Prior to GWAS meta-analysis, separate GWAs analyses for discovery cohorts were performed either via logistic regression or mixed linear model association analysis using BOLT-LMM^38^ or SAIGE^39^ including sex, age, BMI (where available), top principle components (PCs) from principal component analysis (PCA; to control for potential residual population stratification) and genotyping array (if relevant) as covariates (**Supplementary Note**). Details on the cohort-specific GWAS pipelines are reported in **Supplementary Note**. File-level QC of the five individual GWAS summary statistics and meta-level QC from discovery cohorts were carried out using the R package “EasyQC” (v9.2)^40^. In short, the QC process verified data integrity and harmonized both SNP marker IDs and allele coding across the datasets. We only included markers with imputation quality metrics (INFO or Rsq)>0.8 and MAF>1% in the meta-analysis. Markers with deviating allele frequency (difference >20% from the Haplotype Reference Consortium (HRC) genome reference panel v1.1 comprising 32,488 reference individuals of European ancestry^41^) were removed along with indels and multi-allelic markers. The resulting summary statistics of the five discovery cohorts (with a total of 218,920 HEM cases and 725,213 controls) were meta-analyses via fixed-effect meta-analysis based on METAL’s inverse-variance weighted approach^42^. We used the generally accepted threshold of 5×10^−8^ for meta-analysis *P*-values to define statistical significance (*P*_Meta_<5×10^−8^). Genome-wide summary statistics of our analyses are publicly available through our web browser (http://hemorrhoids.online) and have been submitted to the European Bioinformatics Institute (www.ebi.ac.uk/gwas).

### Annotation of HEM GWAS risk loci and gene mapping

We used independent computational pipelines for the functional annotation of GWAS meta-analysis results, using FUnctional Mapping and Annotation of Genome-Wide Association Studies (FUMA v1.3.5)^43^, Data-driven Expression-Prioritized Integration for Complex Traits (DEPICT)^44^, and Multi-marker Analysis of GenoMic Annotation (MAGMA, also implemented in FUMA)^45^. The 102 newly identified genome-wide significant risk loci were defined in FUMA (using default parameters and eQTL databases including GTEx v7) as non-overlapping genomic regions that extend a linkage disequilibrium (LD) window (r^2^ = 0.6) around each lead SNP association signal with *P*_Meta_<5×10^−8^. Annotation of these regions with FUMA resulted in 712 transcripts mapped to risk loci (415 positional and 562 eQTL candidates), while 217 genes were identified using DEPICT, and 255 in MAGMA independent gene-based tests, bringing the total of non-redundant HEM candidate genes to 819 (**Supplementary Table 7**). Regional association plots of all 102 risk loci were generated using LocusZoom^46^.

### Bayesian fine-mapping analysis

A Bayesian fine-mapping analysis was carried out using FINEMAP^5^ for the 102 genome-wide significant risk loci in order to calculate the posterior inclusion probability (PIP) for each lead SNP as causal and to determine a credible set for each risk locus, i.e. a minimum set of variants containing all causal variants with certainty ≥0.95%. As input for fine-mapping we extracted all genetic variants located within the 102 risk loci (as defined by FUMA) and calculated the local LD structure using genotypes from UKBB samples (**Supplementary Table 1**) serving as a reference population.

### Heritability analysis via linkage disequilibrium score regression (LDSC)

Narrow-sense heritability (*h*^2^_SNP_) for HEM and the genetic correlation (*r*_g_) between HEM and other traits were estimated using LD score regression, as implemented in the online platform CTG-VL^47^. This platform integrates summary statistics of 1,387 traits from multiple resources such as UKBB, the Psychiatric Genomics Consortium (PGC) and the Genetic Investigation of ANthropometric Traits (GIANT) consortium. Significantly correlated pairs of traits were reported after FDR correction for multiple comparisons at α=0.05.

### Genome-wide pleiotropy analysis

We conducted cross-phenotype association analysis based on subsets (ASSET) methodology^48^ across association summary statistics from diverticular disease^12^, irritable bowel syndrome (IBS)^49^ and HEM to identify shared risk loci. The subset-based meta-analysis (SBM) method maintains similar type-I error rates as for standard meta-analysis and identifies the best subset of non-null studies while in parallel accounting for multiple-hypothesis testing and shared individuals. This method offers a substantial power increase (sometimes approaching between 100-500%)^48^ compared to standard univariate meta-analysis approaches, where the (heterogeneous) effect of a specific SNP is not exclusively restricted to a single phenotype. Under the assumption that association signals from shared risk loci based on positional overlap are tagging same causal variant for different phenotypes, the SBM approach improves power compared to standard fixed-effects meta-analysis methodology.

### Tissue and pathway enrichment analyses

Gene-set and tissue-specific enrichment analyses (respectively GSEA and TSEA) of HEM genes were carried out using integrated default pipelines in FUMA, and DEPICT implemented in the CTG-VL platform^47,44^. HEM gene lists were derived from three alternative approaches including positional and/or eQTL mapping in FUMA, MAGMA gene-based analyses (also implemented n FUMA), and DEPICT functional annotations, and teste against Gene Ontology (GO) terms and 30 GTEx v7 general tissue types. Statistical significance was defined using *P*_Benjamini-Hochberg_<0.05.

### Polygenic risk scores (PRS) analysis

The analysis of polygenic risk scores (PRS) was performed on the basis of a pruning and thresholding approach, using the *P* value and LD-driven clumping procedure as implemented in PRSice-2^50^. Effect estimates and corresponding standard errors from GWAS meta-analysis results were used as the base dataset to generate weights over a range of *P* values (0.5 to 5×10^−8^) and *r*^2^ 0.1 LD thresholds, with the most appropriate thresholds selected as those that include SNPs with the highest Nagelkerke’s R^2^ value. The selected model was then applied to the QCed genetic datasets from the German case-control cohort, HUNT and DBDS, respectively (see **Methods**). Logistic regression was used to test HEM PRS distribution in cases and controls, taking into account sex, age, BMI and the top 10 PCs from PCA. For HUNT and DBDS we also studied HEM prevalence across PRS percentile distributions. PRSs were binned into percentiles and HEM prevalence from the top 5% of PRS distribution was compared to the reminder of the population in a logistic regression adjusting for sex, age, BMI and the top 10 PCs. Additional analyses were performed to evaluate the relationship between HEM PRS and age at diagnosis (Spearman’s correlation test) and need for invasive treatments (number of surgeries and/or rubber-band ligation; tested with linear regression correcting for sex, age, BMI and the top 10 PCs).

### Phenome-wide association studies (PheWAS)

For each of the 102 GWAS risk loci, we queried the lead SNP and its LD proxies (r^2^>0.8, from 1000 Genomes Project samples of European descent) using PhenoScanner^51^, and manually inspecting the GWAS catalog^52^ and GWAS ATLAS^53^. Only genome-wide significant associations (P<5×10^−8^) were taken into account, and those from GWAS ATLAS were collapsed by trait categories and plotted with the R package “ggforce” into an alluvial diagram.

### Cross-trait analyses

Traits genetically correlated with HEM (from LDSC analyses) were tested for their prevalence in HEM patients vs controls in UKBB and DNPR. In UKBB, we derived the ICD10 diagnoses from data-fields “41202” (primary diagnosis) and “41204” (secondary diagnosis), self-reported medical conditions from data-field “20002”, and self-reported medication use from data-field “20003”. Differential prevalence was tested using a logistic regression model adjusted for sex and age, including FDR correction for multiple comparisons. For DNPR, a previously published method^54^ was used to identify diseases that significantly co-occur more often with HEM diagnoses. Each combination of pair-wise disease co-occurrences was compared to a comparison group matched by sex, age, type of hospital encounter and week of discharge. The relative risk (RR) is used to evaluate the strength of the correlation between significant disease pairs (disease A followed by disease B and *vice versa*). Here, we have used this method to evaluate the temporal co-occurrence of selected diseases and medications with the HEM diagnosis in the DNPR, including FDR correction for multiple comparisons.

### RNA library preparation and RNA-sequencing

The RNA-Sequencing (RNA-Seq) libraries were prepared from 20 ng of total RNA from freshly frozen tissue extracted with the mirVana miRNA Isolation Kit according to the manufacturer’s protocol (Ambion). The NEXTFLEX Combo-Seq Kit (Perkin Elmer) was used to generate combined mRNA and microRNA libraries following manufacturer’s instructions. In short, poly-A-tailed RNA species were reverse transcribed to generate DNA:RNA duplexes whose RNA molecules were specifically sheared by RNase H, resulting in RNA fragments containing 5’-monophosphate and a 3’-hydroxyl groups. These mRNA fragments were 3’-polyadenylated together with small RNAs and then 5’ 4N adapters were ligated to their 5’ ends. Finally, first strand synthesis followed by PCR amplification were used to add sequences required for Illumina sequencing. The generated RNA libraries were quality-controlled using the Agilent 2200 TapeStation (Agilent Technologies), randomized and then deep sequenced (5 samples per lane), 1×50bp using the Illumina HiSeq 4000 platform.

### Mapping and quality assessment of RNA-Seq data

The sequenced reads were demultiplexed and obtained as fastq files for each sample. Data pre-processing, quality control, mapping to genome (build hg38) and transcriptome annotation (miRBase v21, Ensembl 83) were performed using the exceRpt^55^ pipeline. More precisely, reads were trimmed for 3’ adapter sequences, 4N nucleotides at 5’ end and low-quality bases (<Q20). The trimmed sequences shorter than 15 bp were discarded and only high-quality reads were then mapped to genome (with minimum sequence match of 15 nucleotides), annotated and quantified. All RNA-Seq libraries were quality controlled for library size (>10M of mapped reads), transcriptome genome ratio (> 0.95) and outliers for number of detected unique genes and microRNAs (< Q1-1.5 IQR). Low abundant gene-level and microRNA arm level counts that were expressed below 0.1 RPM in less than 25% of the samples per trait were removed from downstream analyses. The generated quality-controlled counts and raw sequencing reads have been deposited at NCBI Gene Expression Omnibus (GEO)^56^ under the accession number GSE154650.

### Differential gene expression analysis

The quality-controlled count data were further analyzed using edgeR^57^ workflow for differential expression analysis. Negative binomial generalized log-linear models were fitted to the trimmed mean of M-values (TMM) normalized count data of HEM genes using glmFit() function with trended dispersion estimates and the offsets for GC-content correction generated by EDASeq (default parameters). The glmLRT() function was used to calculate log-likelihood-ratio statistics and *P*-values of differential expression. The models were adjusted for BMI and histological zones of anal canal (see **Supplementary Note**). The nominal *P*-values were corrected for multiple testing according to Benjamini and Hochberg. Transcripts with an FDR corrected *P*-value < 0.05 and a log_2_ fold change > 0.5 (in either direction) were considered to be significantly differentially expressed.

### Identification and characterization of enriched co-expression modules

Weighed gene co-expression network analysis of hemorrhoid-specific tissue was performed using the automated WGCNA^58^ pipeline implemented in the CEMiTool^59^ R package. The quality-controlled and VST normalized data (36,342 genes in 20 samples) was used to calculate signed scale-free topology overlap matrix, which was subsequently used to define gene co-expression modules in an unsupervised manner. More specifically, Pearson correlation coefficients for each gene-gene comparison (including miRNAs) were used to calculate adjacencies defined as following: *a*_*ij*_ = |0.5 + 0.5 × *cor*(*x*_*i*_, *x*_*j*_)|^β^, where *x*_*i*_ and *x*_*j*_ are expression values of *i*^*th*^ and *j*^*th*^ genes and where β is a soft threshold power based on scale-free topology, which was identified by employing pickSoftThreshold() function from the WGCNA R package. The generated adjacencies were then used to compute topological overlap measures (TOM) and their dissimilarity measures (1-TOM) were further used for average linkage hierarchical clustering and dynamic tree cutting (cutoff value of 0.995) to identify gene co-expression modules. Each gene co-expression module contained a minimum of 50 genes and was summarized into eigengene, which is the first principal component of their expression values. Highly similar modules were identified by correlation of their eigengenes (>0.7 Pearson’s r) and merged together. The intramodular connectivity of each gene was measured by Pearson’s correlation of module eigengene and its expression value. The top 10% of genes having the highest connectivity values were considered as being module hub genes (central nodes in the scale-free network). A Fisher’s exact test was used to identify modules with significantly (P_FDR_< 0.05) overrepresented in HEM genes. The ClusterProfiler^60^ R package was used to identify gene ontology (GO) terms “biological process” pathway enrichments for HEM-significant modules.

### ABO blood group analysis

The association between ABO blood types and HEM was tested on 408,592 and 55,593 individuals from UKBB and GERA, respectively. We first imputed ABO blood group information individually based on genotypes at the *ABO locus* on chromosome 9q34.2. We extracted the genotypes of three SNPs: rs8176719, rs41302905 and the adjacent rs8176747 and inferred blood group status based on these SNPs as previously described^61^. Next, the risk of HEM was assessed on samples from each blood groups of the ABO blood group system (“A”, “B”, “AB” and “O”). An association test based on logistic regression was employed to test for significant HEM association for each of the four blood groups, adjusting for sex, age, BMI and the top 10 PCs from PCA. FDR correction was applied for multiple testing.

### Fluorescence Immunohistochemistry

Fluorescence immunohistochemistry was performed as previously described^12,62^. Briefly, anorectal specimens were fixed in 4% paraformaldehyde for 24 hours. Paraffin-embedded tissue sections were pre-treated with citrate buffer and primary antibodies were incubated overnight. Used primary and secondary antibodies are listed in **Supplementary Table 12**. All antibodies were diluted in antibody diluent (ThermoFisher Scientific). Nuclei were counterstained with DAPI (Roche, Mannheim, Germany). Image acquisition was performed on a fluorescence inverted microscope (Axiovert 200 M, Zeiss, Gottingen, Germany) coupled to an AxioCam MR3 camera (Zeiss) using Axiovision software (version 4.7, Zeiss).

## References

1. Jacobs, D. Clinical practice. Hemorrhoids. N Engl J Med 371, 944–51 (2014).

2. Duthie, H.L. & Gairns, F.W. Sensory nerve-endings and sensation in the anal region of man. Br J Surg 47, 585–95 (1960).

3. Haas, P.A., Haas, G.P., Schmaltz, S. & Fox, T.A., Jr. The prevalence of hemorrhoids. Dis Colon Rectum 26, 435–9 (1983).

4. Yang, J.Y., Peery, A.F., Lund, J.L., Pate, V. & Sandler, R.S. Burden and Cost of Outpatient Hemorrhoids in the United States Employer-Insured Population, 2014. Am J Gastroenterol 114, 798–803 (2019).

5. Benner, C. et al. FINEMAP: efficient variable selection using summary data from genome-wide association studies. Bioinformatics 32, 1493–501 (2016).

6. Malysz, J. et al. Conditional genetic deletion of Ano1 in interstitial cells of Cajal impairs Ca(2+) transients and slow waves in adult mouse small intestine. Am J Physiol Gastrointest Liver Physiol 312, G228–G245 (2017).

7. Naba, A. et al. Extracellular matrix signatures of human primary metastatic colon cancers and their metastases to liver. BMC Cancer 14, 518 (2014).

8. Dvir, H., Silman, I., Harel, M., Rosenberry, T.L. & Sussman, J.L. Acetylcholinesterase: from 3D structure to function. Chem Biol Interact 187, 10–22 (2010).

9. Bartels, C.F., Zelinski, T. & Lockridge, O. Mutation at codon 322 in the human acetylcholinesterase (ACHE) gene accounts for YT blood group polymorphism. Am J Hum Genet 52, 928–36 (1993).

10. Margetis, N. Pathophysiology of internal hemorrhoids. Ann Gastroenterol 32, 264–272 (2019).

11. Nasseri, Y.Y. et al. Abnormalities in collagen composition may contribute to the pathogenesis of hemorrhoids: morphometric analysis. Tech Coloproctol 19, 83–7 (2015).

12. Schafmayer, C. et al. Genome-wide association analysis of diverticular disease points towards neuromuscular, connective tissue and epithelial pathomechanisms. Gut 68, 854–865 (2019).

13. Tung, J.Y., Kiefer, A.K., Mullins, M., Francke, U. & Eriksson, N. Genome-wide association analysis implicates elastic microfibrils in the development of nonsyndromic striae distensae. J Invest Dermatol 133, 2628–2631 (2013).

14. Plackett, T.P., Kwon, E., Gagliano, R.A., Jr. & Oh, R.C. Ehlers-danlos syndrome-hypermobility type and hemorrhoids. Case Rep Surg 2014, 171803 (2014).

15. Zhu, L. et al. Mutations in myosin heavy chain 11 cause a syndrome associating thoracic aortic aneurysm/aortic dissection and patent ductus arteriosus. Nat Genet 38, 343–9 (2006).

16. Gilbert, M.A. et al. Protein-elongating mutations in MYH11 are implicated in a dominantly inherited smooth muscle dysmotility syndrome with severe esophageal, gastric, and intestinal disease. Hum Mutat 41, 973–982 (2020).

17. Kwartler, C.S. et al. Overexpression of smooth muscle myosin heavy chain leads to activation of the unfolded protein response and autophagic turnover of thick filament-associated proteins in vascular smooth muscle cells. J Biol Chem 289, 14075–88 (2014).

18. Suri, P. et al. Genome-wide meta-analysis of 158,000 individuals of European ancestry identifies three loci associated with chronic back pain. PLoS Genet 14, e1007601 (2018).

19. Bjornsdottir, G. et al. Sequence variant at 8q24.21 associates with sciatica caused by lumbar disc herniation. Nat Commun 8, 14265 (2017).

20. Holtmann, G., Shah, A. & Morrison, M. Pathophysiology of Functional Gastrointestinal Disorders: A Holistic Overview. Dig Dis 35 Suppl 1, 5–13 (2017).

21. Mazzone, A. et al. Direct repression of anoctamin 1 (ANO1) gene transcription by Gli proteins. FASEB J 33, 6632–6642 (2019).

22. Moore, S.W. & Johnson, G. Acetylcholinesterase in Hirschsprung’s disease. Pediatr Surg Int 21, 255–63 (2005).

23. Aigner, F. et al. Revised morphology and hemodynamics of the anorectal vascular plexus: impact on the course of hemorrhoidal disease. Int J Colorectal Dis 24, 105–13 (2009).

24. Franchini, M. & Lippi, G. Relative Risks of Thrombosis and Bleeding in Different ABO Blood Groups. Semin Thromb Hemost 42, 112–7 (2016).

25. Huang, E., Khalili, H., Strate, L. & Chan, A. ABO Blood Group and the Risk of Gastrointestinal Bleeding. Gastroenterology 142, S–503 (2012).

26. Chang, S.S., Sung, F.C., Lin, C.L. & Hu, W.S. Association between hemorrhoid and risk of coronary heart disease: A nationwide population-based cohort study. Medicine (Baltimore) 96, e7662 (2017).

27. Groot, H.E. et al. Genetically Determined ABO Blood Group and its Associations With Health and Disease. Arterioscler Thromb Vasc Biol 40, 830–838 (2020).

28. Tung, J.Y. et al. Efficient replication of over 180 genetic associations with self-reported medical data. PLoS One 6, e23473 (2011).

29. Do, C.B. et al. Web-based genome-wide association study identifies two novel loci and a substantial genetic component for Parkinson’s disease. PLoS Genet 7, e1002141 (2011).

30. Bycroft, C. et al. The UK Biobank resource with deep phenotyping and genomic data. Nature 562, 203–209 (2018).

31. Leitsalu, L. et al. Cohort Profile: Estonian Biobank of the Estonian Genome Center, University of Tartu. Int J Epidemiol 44, 1137–47 (2015).

32. Fritsche, L.G. et al. Association of Polygenic Risk Scores for Multiple Cancers in a Phenome-wide Study: Results from The Michigan Genomics Initiative. Am J Hum Genet 102, 1048–1061 (2018).

33. Krawczak, M. et al. PopGen: population-based recruitment of patients and controls for the analysis of complex genotype-phenotype relationships. Community Genet 9, 55–61 (2006).

34. Krokstad, S. et al. Cohort Profile: the HUNT Study, Norway. Int J Epidemiol 42, 968–77 (2013).

35. Hansen, T.F. et al. DBDS Genomic Cohort, a prospective and comprehensive resource for integrative and temporal analysis of genetic, environmental and lifestyle factors affecting health of blood donors. BMJ Open 9, e028401 (2019).

36. Burgdorf, K.S. et al. Socio-demographic characteristics of Danish blood donors. PLoS One 12, e0169112 (2017).

37. Jensen, T.B. et al. Content and validation of the Electronic Patient Medication module (EPM)-the administrative in-hospital drug use database in the Capital Region of Denmark. Scand J Public Health 48, 43–48 (2020).

38. Loh, P.R. et al. Efficient Bayesian mixed-model analysis increases association power in large cohorts. Nat Genet 47, 284–90 (2015).

39. Zhou, W. et al. Efficiently controlling for case-control imbalance and sample relatedness in large-scale genetic association studies. Nat Genet 50, 1335–1341 (2018).

40. Winkler, T.W. et al. Quality control and conduct of genome-wide association meta-analyses. Nat Protoc 9, 1192–212 (2014).

41. McCarthy, S. et al. A reference panel of 64,976 haplotypes for genotype imputation. Nat Genet 48, 1279–83 (2016).

42. Willer, C.J., Li, Y. & Abecasis, G.R. METAL: fast and efficient meta-analysis of genomewide association scans. Bioinformatics 26, 2190–1 (2010).

43. Watanabe, K., Taskesen, E., van Bochoven, A. & Posthuma, D. Functional mapping and annotation of genetic associations with FUMA. Nat Commun 8, 1826 (2017).

44. Pers, T.H. et al. Biological interpretation of genome-wide association studies using predicted gene functions. Nat Commun 6, 5890 (2015).

45. de Leeuw, C.A., Mooij, J.M., Heskes, T. & Posthuma, D. MAGMA: generalized gene-set analysis of GWAS data. PLoS Comput Biol 11, e1004219 (2015).

46. Pruim, R.J. et al. LocusZoom: regional visualization of genome-wide association scan results. Bioinformatics 26, 2336–7 (2010).

47. Cuéllar-Partida, G. et al. Complex-Traits Genetics Virtual Lab: A community-driven web platform for post-GWAS analyses. bioRxiv, 518027 (2019).

48. Bhattacharjee, S. et al. A subset-based approach improves power and interpretation for the combined analysis of genetic association studies of heterogeneous traits. Am J Hum Genet 90, 821–35 (2012).

49. Bonfiglio, F. et al. Female-Specific Association Between Variants on Chromosome 9 and Self-Reported Diagnosis of Irritable Bowel Syndrome. Gastroenterology 155, 168–179 (2018).

50. Choi, S.W. & O’Reilly, P.F. PRSice-2: Polygenic Risk Score software for biobank-scale data. Gigascience 8(2019).

51. Staley, J.R. et al. PhenoScanner: a database of human genotype-phenotype associations. Bioinformatics 32, 3207–3209 (2016).

52. Buniello, A. et al. The NHGRI-EBI GWAS Catalog of published genome-wide association studies, targeted arrays and summary statistics 2019. Nucleic Acids Res 47, D1005–D1012 (2019).

53. Watanabe, K. et al. A global overview of pleiotropy and genetic architecture in complex traits. Nat Genet 51, 1339–1348 (2019).

54. Jensen, A.B. et al. Temporal disease trajectories condensed from population-wide registry data covering 6.2 million patients. Nat Commun 5, 4022 (2014).

55. Rozowsky, J. et al. exceRpt: A Comprehensive Analytic Platform for Extracellular RNA Profiling. Cell Syst 8, 352–357 e3 (2019).

56. Edgar, R., Domrachev, M. & Lash, A.E. Gene Expression Omnibus: NCBI gene expression and hybridization array data repository. Nucleic Acids Res 30, 207–10 (2002).

57. McCarthy, D.J., Chen, Y. & Smyth, G.K. Differential expression analysis of multifactor RNA-Seq experiments with respect to biological variation. Nucleic Acids Res 40, 4288–97 (2012).

58. Langfelder, P. & Horvath, S. WGCNA: an R package for weighted correlation network analysis. BMC Bioinformatics 9, 559 (2008).

59. Russo, P.S.T. et al. CEMiTool: a Bioconductor package for performing comprehensive modular co-expression analyses. BMC Bioinformatics 19, 56 (2018).

60. Yu, G., Wang, L.G., Han, Y. & He, Q.Y. clusterProfiler: an R package for comparing biological themes among gene clusters. OMICS 16, 284–7 (2012).

61. Severe Covid, G.G. et al. Genomewide Association Study of Severe Covid-19 with Respiratory Failure. N Engl J Med 383, 1522–1534 (2020).

62. Cossais, F. et al. Altered enteric expression of the homeobox transcription factor Phox2b in patients with diverticular disease. United European Gastroenterol J 7, 349–357 (2019).

