## Supplementary Note for "Genome-wide analysis of 944,133 individuals provides insights into the etiology of hemorrhoidal disease"

### Histology of hemorrhoidal plexus

For histologic examination and phylogenetic comparison of the hemorrhoidal plexus, formalin fixed anorectal specimens were obtained from *Homo sapiens*, *Gorilla gorilla gorilla*, baboon (*Papio anubis*), and mouse (10-week old male C57BL/6JRj mouse).

Human tissue was retrieved from a healthy donor (female, 54 years) who was recruited by the body donation program of the Institute of Anatomy, Kiel University. The donors had previously given written consent to the use of their samples for teaching and research purposes; the donors were free from diseases related to the gastrointestinal tract and the anorectum. The gorilla specimen comes from a 43-year-old female western lowland gorilla from Nuremberg Zoo (Germany). The animal had to be euthanized due to a terminal metastatic adenocarcinoma of the uterus. Rectum and surrounding tissue were removed during post-mortem examination four hours after death, cut and fixed in 10% neutral buffered formalin. Rectum samples from the baboon were taken from a ten year old male olive baboon kept at the German Primate Center Göttingen and included in a study authorized by the governmental veterinary authority, i.e. the Lower Saxony State Office for Consumer Protection (Food Safety Ref. No. 33.19-42502-04-18/3036 according to the regulations of the German Welfare Act and the European Directive 2010/63/EU on the protection of animals used for experimental and other scientific purpose). The rectal specimen was collected during routine necropsy following a standardized necropsy protocol and fixed in 10% buffered formalin.

All tissue samples were taken from the anal canal at the level of the hemorrhoidal plexus, dehydrated, embedded in paraffin wax, cut into sections (6 µm) and processed for hematoxylin-eosin and Azan stainings. The findings were evaluated and documented with a Keyence microscope (BZ-X800) using the integrated stitching tool BZ-X800 Analyser software version 1.1.1.8.

#### Genotyping, quality control and genotype imputation of cohorts included in this study

##### *23andMe*

DNA extraction and genotyping were performed on saliva samples by National Genetics Institute (NGI), a CLIA licensed clinical laboratory and a subsidiary of Laboratory Corporation of America. Samples had been genotyped on one of four genotyping platforms. The V1 and V2 platforms were variants of the Illumina HumanHap550+ BeadChip, including about 25,000 custom SNPs selected by 23andMe, with a total of about 560,000 SNPs. The V3 platform was based on the Illumina OmniExpress+ BeadChip, with custom content to improve the overlap with the V2 array, with a total of about 950,000 SNPs. The V4 platform is a fully custom array, including a lower redundancy subset of V2 and V3 SNPs with additional coverage of lower-frequency coding variation, and about 570,000 SNPs. Samples that failed to reach 98.5% call rate were re-analyzed. Individuals who repeatedly failed analyses were recontacted by the 23andMe customer service to provide additional samples.

For GWAS quality control (QC) analysis, we limited participants to a set of individuals with ≥97% European descent, determined by analysis of local ancestry[1]. In brief, the algorithm initially partitions phased genomic data into short windows of about 100 SNPs. Within each window, a support vector machine (SVM) was used to classify each haplotype into one of 31 reference populations. SVM classifications were translated into a hidden Markov model (HMM) that takes into account switch errors and incorrect assignments and reports probabilities for each reference population in each window. Finally, simulated admixed individuals were used to recalibrate the HMM probabilities so that the reported assignments are consistent with the simulated admixture ratios. Reference population data was derived from public datasets (the Human Genome Diversity Project, HapMap, and 1000 Genomes) and from 23andMe customers who reported having four grandparents from the same country. For each analysis, a maximal set of unrelated individuals was selected using a segmental identity-by-descent (IBD) estimation algorithm[2]. Individuals were identified as related if they shared more than 700 cM IBD, including regions where the two individuals share either one or both genomic segments identical-by-descent. This degree of relatedness (about 20% of the genome) corresponds approximately to the expected minimum proportion between cousins and first-degree cousins in an outbred population. SNPs deviating from Hardy-Weinberg equilibrium ($P<10^{-20}$), having a call rate <95%, or with large discrepancies in allele frequency compared to the European 1000 Genomes reference data were excluded. SNPs with large differences in allele frequency (chi squared $P<10^{-15}$) were identified by computing a 2x2 table of allele counts for European 1000 Genomes samples and 2000 randomly sampled 23andMe customers of European ancestry.

Genotype data were imputed using the September 2013 1000 Genomes Phase1 reference haplotypes[3]. Phasing and imputation was performed separately for the data of each genotyping platform. Phasing was performed with a phasing tool, Finch, developed internally by 23andMe, which implements the Beagle haplotype graph-based phasing algorithm[4] and which was modified to separate the steps of constructing the haplotype graph and phasing. Finch extends the Beagle model to allow genotyping errors and recombination events to handle cases where there are no consistent paths through the haplotype graph for the individual to be phased. From a representative sample of genotyped individuals, haplotype graphs were generated for European and non-European samples for each 23andMe genotyping platform. Subsequently, an out-of-sample phasing of all genotyped individuals against the corresponding graph was performed. In preparation for imputation, the phased chromosomes were divided into segments of no more than 10,000 genotyped SNPs, with overlaps of 200 SNPs. Each phased segment was imputed against all-ethnicity 1000 Genomes haplotypes (excluding monomorphic and singleton sites) using Minimac2[5], using 5 rounds and 200 states for parameter estimation. For the X chromosome, we created separate haplotype graphs for the non-pseudoautosomal region and each pseudoautosomal region, and these regions were separately phased. Then we imputed males and females together using Minimac2, as for the autosomes, and treated males as homozygous pseudo-diploids for the non-pseudoautosomal region. After QC and genotype imputation a total of 7,024,410 SNPs with imputation quality score Rsq>0.8 and minor allele frequency (MAF) >1% in 174,785 cases and 228,060 controls were available for association analysis.

##### *UK Biobank*

DNA samples were genotyped on custom UK Biobank (UKBB) arrays. 408,951 individuals from UKBB were genotyped for 825,927 variants using a custom Affymetrix UK Biobank

Axiom Array, and 49,626 individuals were genotyped for 807,411 variants using a custom Affymetrix UK BiLEVE Axiom Array chip from the UK BiLEVE study[6], which is a subset of UKBB.

All SNPs were subjected to quality control (QC): checks, such as deviations from Hardy-Weinberg equilibrium ($P<10^{-5}$), batch and plate effects, sex effects, and array effects across control replicates. The SNPs that failed call rate <0.95 were set to missing for all individuals. The QC was performed centrally for each sample tested for heterozygosity and missing rates. Samples with excessive relatedness (>10 suspected third-degree relatives) were excluded. Full details of the QC of the genetic data performed centrally by UK Biobank are available in the original publication[7]. To identify sample outliers (i.e. subjects of non-Europeans ancestry), we performed principal component analysis (PCA) with FlashPCA2[8]. PCA revealed no non-European ancestry outliers. Genotypes of 408,592 UKBB participants with European ancestry (self-reported "white" and genetic Caucasian) were used after QC. Of these, 23,856 samples satisfied our criteria for being HEM cases (either ICD10 code I84 or ICD9 code 455 in medical records) and the remainder of the cohort (n=384,736) served as study controls.

Genetic variants were imputed centrally by UKBB using IMPUTE4[7] and a reference panel that merged the UK10K and 1000 Genomes Phase 3 panel as well as the Haplotype Reference Consortium (HRC) panel[7]. After QC and genotype imputation, a total of 9,572,556 SNPs with an imputation quality score INFO>0.8 and MAF >1% in 23,856 cases and 384,736 controls were available for association analysis.

##### *Estonian Genome Project of University of Tartu (EGCUT)*

The Estonian cohort originates from the population-based biobank of the Estonian Genome Project of University of Tartu (EGCUT). The EGCUT project has been conducted according to the Estonian Gene Research Act and all participants have signed the broad informed consent. The current cohort size is about 200,000 aged 18 years and older, which is very close to the age distribution in the adult Estonian population. Subjects were recruited by general practitioners and doctors in hospitals. The persons who visited the general practitioner's practices or hospitals were selected at random. Each participant completed a computer assisted personal interview during 1-2 hours in a doctor's office, which included personal data (place of birth, place(s) of living, nationality etc.), genealogical data (family history, three generations), educational and occupational history and lifestyle data (physical activity, dietary habits, smoking habits, alcohol consumption, women's health, quality of life). Diseases were defined according to the ICD10 coding. Illumina Human CoreExome, OmniExpress, 370CNV BeadChip and Illumina Global Screening Array (GSA) arrays were used for genotyping.

QC included filtering based on sample call rate (<98%), heterozygosity (> mean ± 3SD), genotype and phenotype sex discordance, cryptic relatedness (IBD >20%) and outliers of European ancestry based on a multidimensional scaling (MDS) analysis including 210 HapMap reference samples[9]. SNP QC included testing for call rate (<99%), MAF (<1%) and extreme deviation from Hardy–Weinberg equilibrium ($P<10^{-4}$).

Pre-phasing was performed using SHAPEIT2[10]. Genotype imputation was performed using the Estonian-specific reference panel[11] and IMPUTE2[12] with default parameters. After QC and genotype imputation, a total of 7,462,975 SNPs with imputation quality score INFO>0.8 and minor allele frequency (MAF) >1% in 6,956 cases and 30,441 controls were available for association analysis.

##### ***Michigan Genomics Initiative (MGI)***

DNA samples were genotyped on custom Illumina HumanCoreExome v12.1 bead chips. Samples were excluded if they exhibited (1) a calling rate < 99%, (2) an estimated contamination > 2.5% (BAF Regress)[13] or (3) deviating sex information if the derived sex did not match the self-reported gender. Variants were excluded if they (1) deviated from Hardy-Weinberg equilibrium ($P_{HWE}<10^{-5}$), (2) had a calling rate < 99%. After quality control, 392,323 polymorphic variants were kept in the following analyses. Next, we estimated the pair-wise relationship of the samples using the software KING[14] and we limited the dataset within a subset of individuals without first- or second-degree relationship. The genetic ancestry of the samples were derived by projecting the principal components of the samples onto that of the Human Genome Diversity Project (HGDP) reference panel (938 unrelated individuals)[15]. Principal component analysis was performed using PLINK v1.90[16], including a subset of LD pruned variants ($r^2$ < 0.5) with MAF >1% shared between the HGDP reference and the MGI data. We retained only samples of recent European ancestry (defined as samples that fell into a circle around the center of the reference HGDP populations in the PC1 versus PC2 space).

Genotype imputation was conducted using the Haplotype Reference Consortium (HRC) panel and the Michigan Imputation Server[17]. After QC and genotype imputation, a total of 6,536,218 SNPs with imputation quality score Rsq>0.8 and MAF >1% in 4,539 cases and 35,338 controls were available for association analysis.

***Genetic Epidemiology Research on Aging (GERA)***

DNA samples were collected from participants of the Genetic Epidemiology Research on Aging (GERA) cohort and genotyped on high-density custom designed Affymetrix Axiom arrays. Genetic variants with >5% of missing data, MAF <1% in either disease sets or in controls or deviating from Hardy-Weinberg equilibrium ($P<10^{-5}$) were excluded. Samples with >2% missing data and overall increased/decreased heterozygosity rates were removed. For robust duplicate/relatedness testing (IBS/IBD estimation) and population structure analysis, a pruned subset of 144,799 independent SNPs was used. Pair-wise percentage IBD values were computed using PLINK. By definition, Z0: P(IBD=0), Z1: P(IBD=1), Z2: P(IBD=2), Z0+Z1+Z2=1, and PI_HAT: P(IBD=2) + 0.5 * P(IBD=1) (proportion IBD). One individual (the one showing greater missingness) from each pair with PI_HAT>0.1875 was removed. To identify sample outliers (i.e. subjects of non-Europeans ancestry), we performed principal component analysis (PCA) using the smartpca program[18], based on a set of 144,799 “high-performing” markers after exclusion of SNPs that had an $r^2$ value greater than 0.5, were within 5 MB of each other, within the MHC region, had a call rates lower than 99.5% and that were located in regions with inversions on chromosomes 8p23 and 17q21.

Genotype data were pre-phased with SHAPE-IT v2.5[10], and then imputed with IMPUTE2 v2.3.1[19] using the 1000 Genomes Phase 3 data as a reference panel. After QC and genotype imputation, a total of 6,897,996 SNPs with imputation quality score INFO>0.8 and MAF >1% in 8,813 cases and 46,780 controls were available for association analysis.

***German case-control cohort***

DNA samples were genotyped using Illumina's Global Screening Array version 1.0. Patients with a reported "migration background" were excluded. 3,505 eligible patients were contacted by their treating physician by mail. The initial submission rate was 40%. After consent to participate, the Popgen Biobank sent a study kit with a questionnaire on clinical and socio-demographic characteristics and a set of blood tubes, so that a blood sample could be collected at the family doctor's office and returned to the study center. In addition, a subset of study participants were asked to complete a comprehensive questionnaire on their dietary habits and usual physical activity. Patients were excluded from the study in the absence of informed consent/blood sample or after withdrawal of consent.

Variants that had >2% missing data, a minor allele frequency <0.1% in either of the different disease sets or in controls, had different missing genotype rates in affected and unaffected individuals ($P_{Fisher}<10^{-5}$) or deviated from Hardy-Weinberg equilibrium ($P_{HWE}<10^{-5}$) were excluded. Samples that had >2% missing data and overall increased/decreased heterozygosity rates (with an average marker heterozygosity of ±5 s.d. away from the sample mean) were removed. For robust duplicate/relatedness testing (IBS/IBD estimation) and population structure analysis, we used a pruned subset of 100,596 independent SNPs (MAF>0.05) SNPs excluding X- and Y-chromosomes, SNPs in LD (leaving no pairs with $r^2>0.2$), and 11 high-LD regions as described by Price *et al.*[20]. Pair-wise percentage IBD values were computed using PLINK2. By definition, Z0: P(IBD=0), Z1: P(IBD=1), Z2: P(IBD=2), Z0+Z1+Z2=1, and PI_HAT: P(IBD=2) + 0.5 * P(IBD=1) (proportion IBD). One individual (the one showing greater missingness) from each pair with PI_HAT>0.1875 was removed. To identify sample outliers (i.e.subjects of non-Europeans ancestry), we performed principal component analysis (PCA) with FlashPCA2[8], on the basis of a set of 100,596 independent markers (described above).

Genotype imputation was conducted using the Haplotype Reference Consortium (HRC) panel and the Sanger Imputation Service[17]. After QC and genotype imputation, a total of 7,117,385 SNPs with imputation quality score INFO>0.8 and MAF >1% in 1,144 cases and 2,740 population controls were available for association analysis.

***The Trøndelag Health Study (HUNT)***

DNA was extracted from whole blood from HUNT2 and HUNT3. Genotyping was a research collaboration between researchers from the Norwegian University of Science and Technology (NTNU) and the University of Michigan. Each individual with a DNA sample of an appropriate DNA concentration was selected for genotyping. Samples were taken at random and genotyped in batches. All genotyping was performed at the Genomics-Core Facility (GCF) at NTNU.

Genotype quality control and genotype imputation were conducted by the K.G. Jebsen Center for Genetic Epidemiology, Department of Public health and Nursing, Faculty of Medicine and Health Sciences, NTNU. In total, DNA from 71,860 HUNT samples was genotyped using one of three different Illumina HumanCoreExome arrays: HumanCoreExome12 v1.0, HumanCoreExome12 v1.1 and UM HUNT Biobank v1.0. Samples were excluded if they did not achieve a 99% call rate, had a contamination >2.5% as estimated with BAF Regress[21], had large chromosomal copy number variants, a lower call rate of a technical duplicate pair and a twin pair, gonosomal constellations other than XX and XY, or whose derived sex was inconsistent with the reported sex. Samples that passed quality control were analysed in a second round of genotype calling following the Genome Studio quality control protocol described elsewhere[22]. Genomic position, strand orientation and the reference allele of genotyped variants were determined by aligning their probe sequences against the human genome (Genome Reference Consortium Human genome build 37 and revised Cambridge Reference Sequence of the human mitochondrial DNA; http://genome.ucsc.edu) using BLAT. Variants were excluded if their probe sequences could not be perfectly mapped to the reference genome, cluster separation was <0.3, Gentrain score was <0.15, showed deviations from Hardy-Weinberg equilibrium in unrelated samples of European ancestry with *P*-value <0.0001), their call rate was <99%, or another assay with higher call rate genotyped the same variant. Ancestry of all samples was inferred by projecting all genotyped samples onto top principal components of the Human Genome Diversity Project (HGDP) reference panel (938 unrelated individuals; downloaded from http://csg.sph.umich.edu/chaolong/LASER/)[15,23], using PLINK v1.90. Recent European ancestry was defined for samples that fell into an ellipsoid spans European populations of the HGDP panel. The different arrays were harmonized by reducing them to a set of overlapping variants and excluding variants that had frequency differences >15% between data sets, or that were monomorphic in one data set and had a MAF >1% in another data set. The resulting genotype data were phased using Eagle2 v2.3[24].

Imputation was performed on the 69,716 samples of recent European ancestry using Minimac3 (v2.0.1, http://genome.sph.umich.edu/wiki/Minimac3)[25] with default settings (2.5 Mb reference based chunking with 500kb windows) and a customized Haplotype Reference consortium release 1.1 (HRC v1.1) for autosomal variants and HRC v1.1 for chromosome X variants[17]. The customized reference panel represented the merged panel of two reciprocally imputed reference panels: (1) 2,201 low-coverage whole-genome sequences samples from the HUNT study and (2) HRC v1.1 with 1,023 HUNT WGS samples removed before merging. After QC and genotype imputation, over 24.9 million SNPs with imputation quality score R^2 >=0.3 in 977 cases and 68,314 population controls were available for association analysis.

***Danish Blood Donor Study***

DNA samples were genotyped at deCode genetics, Iceland, using Illumina's Global Screening Array as described elsewhere[26]. Details on genotype quality control and imputation are available in Hansen et al., 2019[26]. First- and second-degree relatives were excluded from the analysis. The phenotypic data used in this project includes sex, age, self-reported BMI and selected diagnoses from the Danish National Patient Registry. Participant where classified as having HEM using the ICD-8 code 455 or ICD-10 codes I84 or K64 from the National Patient Registry, resulting in the identification of 1,754 HEM cases in the DBDS cohorts.

The DBDS Genomic Consortium is represented by the following scientists: Andersen Steffen, Department of Finance, Copenhagen Business School, Copenhagen, Denmark; Banasik Karina, Novo Nordisk Foundation Center for Protein Research, Faculty of Health and Medical Sciences, University of Copenhagen, Copenhagen, Denmark; Brunak Søren, Novo Nordisk Foundation Center for Protein Research, Faculty of Health and Medical Sciences, University of Copenhagen, Copenhagen, Denmark; Burgdorf Kristoffer, Department of Clinical Immunology, Copenhagen University Hospital, Copenhagen, Denmark; Erikstrup Christian, Department of Clinical Immunology, Aarhus University Hospital, Aarhus, Denmark; Hansen Thomas Folkmann, Danish Headache Center, department of Neurology Rigshospitalet, Glostrup, Denmark; Hjalgrim Henrik, Department of Epidemiology Research, Statens Serum Institut, Copenhagen, Denmark; Jemec Gregor, Department of Clinical Medicine, Sealand University hospital, Roskilde, Denmark; Jennum Poul, Department of clinical neurophysiology at University of Copenhagen, Copenhagen, Denmark; Johansson Per Ingemar, Department of Clinical Immunology, Copenhagen University Hospital, Copenhagen, Denmark; Nielsen Kasper Rene, Department of Clinical Immunology, Aalborg University Hospital, Aalborg, Denmark; Nyegaard Mette, Department of Biomedicine, Aarhus University, Denmark; Mie Topholm Bruun, Department of Clinical Immunology, Odense University Hospital, Odense, Denmark; Pedersen Ole Birger, Department of Clinical Immunology, Naestved Hospital, Naestved, Denmark; Petersen Mikkel, Department of Clinical Immunology, Aarhus University Hospital, Aarhus, Denmark; Sørensen Erik, Department of Clinical Immunology, Copenhagen University Hospital Copenhagen, Denmark; Ullum Henrik, Department of Clinical Immunology, Copenhagen University Hospital, Copenhagen, Denmark; Werge Thomas, Institute of Biological Psychiatry, Mental Health Centre Sct. Hans, Copenhagen University Hospital, Roskilde, Denmark; Gudbjartsson Daniel, deCODE genetics, Reykjavik, Iceland; Stefansson Kari, deCODE genetics, Reykjavik, Iceland; Stefánsson Hreinn, deCODE genetics, Reykjavik, Iceland; Þorsteinsdóttir Unnur, deCODE genetics, Reykjavik, Iceland.

#### GWAS association analysis for discovery cohorts

##### *23andMe*

For comparisons between cases and controls, association test results were performed by logistic regression analysis assuming additive allelic effects. For tests using imputed data, imputed allelic dosages were used rather than best-guess genotypes. Age, biological sex, BMI, the top five principal components from principal component analysis (to account for potential residual population structure) as well as indicators for genotype platforms (to account for genotype batch effects) were included as covariates in the regression analysis. The association test *P*-value was computed using a likelihood ratio test. Results for the X chromosome are computed similarly, with male genotypes coded as if they were homozygous diploid for the observed allele. For chromosome X association analysis, haplotypic allele calls in males outside pseudoautosomal regions (PAR) are converted to homozygous calls by doubling the haplotypic allele (assuming inactivation of large parts of one of the two female X chromosomes[27] and sex was used as a covariate for association testing. Association summary statistics were adjusted for an estimated genomic control inflation factor $\lambda_{GC}$=1.200.

***UK Biobank (UKBB), Estonian Genome Project of University of Tartu (EGCUT), Michigan Genomics Initiative (MGI), Genetic Epidemiology Research on Aging (GERA)***

For each individual case-control data set, association testing was performed using a linear mixed model (LMM) under an additive genetic model for all measured and imputed genetic variants in dosage format using BOLT-LMM[28] (UKBB, GERA) or SAIGE[29] (MGI). Within association analysis, we adjusted for the following covariates: sex, age, BMI (available for UKBB and GERA), the top ten principal components from principal component analysis and a binary indicator variable for genotyping platform (e.g. UKBB Axiom Array vs. UK BiLEVE Axiom Array) to account for the different genotyping chips. For GWAS data set from EGCUT, association testing was carried out with EPACTS [https://github.com/statgen/EPACTS], adjusting for age, sex, binary indicator variable for genotyping platform and top four principal components from principal component analysis. For chromosome X association analysis, see text above. The genomic control inflation factors for UKBB, EGCUT, MGI and GERA were $\lambda_{GC}$=1.0966, 1.0263, 0.9822 and 0.9541, respectively. For GWAS meta-analysis across discovery cohorts (23andme, UKBB, EGCUT, MGI and GERA), see **Methods**.

#### Gene signature-based determination of anal canal zones

Histologically, the anal canal can be divided into three zones according to the epithelial lining. The upper part is of the mucosal type (intestinal) and the lower part is of the squamous keratinized (anoderm), while the middle part, where the epithelium varies, is called the anal transitional zone[30,31]. Due to the gradient nature of the anal canal epithelium, keratinocyte and sebocyte marker gene signatures from the xCell catalog[32] were used to discriminate the different histological zones. More specifically, the quality-controlled gene counts were normalized using the variance stabilizing transformation (VST) implemented in the DESeq2 R package[33]. The normalized gene counts were then ranked according to their expression level using the rank() function from the base R package and submitted to single sample gene set enrichment analysis (ssGSEA)[34] implemented in the GSVA R package[35]. The obtained normalized enrichment scores (NES) of keratinocytes and sebocytes were used to cluster samples into 3 groups (in accordance to the number of histological zones) by employing the base R function kmeans() with k=3 and nstart=20 as parameters. The obtained clusters were assigned to histological zones by the relative abundance of keratinocytes and sebocytes (i.e. sebum-producing epithelial cells), and further confirmed by the expression levels of previously defined marker genes, including *KRT4*, *KRT8*, *KRT13* and *KRT20*[31,36] genes (**Supplementary Figure 9**). Multidimensional scaling (MDS) analysis using Spearman's rank correlation distance (1-correlation coefficient) was performed on VST normalized expression data and was used to explore the results.

#### *In silico* variant protein analysis

To construct a first hypothetical model of whether *SRPX* and *ANO1* missense lead variants (shown in red in **Figure 1**) are likely to interfere with functionally active domains at the protein level, we conducted protein domain analyses for SRPX and ANO1.

ANO1 (also TMEM16A) is an anion channel protein that enables the passive flow of Cl anions through the membrane as a result of increased intracellular Ca2+ levels. The decrease in an anion flow occurs over time after prolonged stimulation eventually leads to to complete desensitization to saturated Ca2+. In addition to elevated Ca-levels, ANO1 function is regulated by the PIP2 (Phosphatidylinositol(4,5)-bisphosphate) signal lipid which binds at the cytoplasmic membrane interface[37]. The Interaction with PIP2 has been shown to slow down the ANO1 regulatory process, probably by hindering the gradual collapse of the ion conduction pore[38].

The ANO1 protein functions as a homodimer, with each subunit consisting of ten transmembrane helices and its own anion conduction pore (**Supplementary Figure 6**) which is composed of helices 3-7 and contains a conserved Ca2+ binding site[39,40]. Ion flow through the pore is made possible by local structural rearrangements that open the channel in response to Ca2+ binding[39].

The variant F608S is located at the beginning of the transmembrane helix 5, i.e. near the cytoplasmic interface (**Supplementary Figure 6**). Although helix 5 is part of the ion conduction pore, the sidechain of F608 points in the opposite direction to the dimer interface and is located near the predicted PIP2 binding residues. Adjacent K609 forms a

stabilizing salt bridge with E594 in the TM4-TM5 linker which is conserved in all members of the TMEM16 protein family. Mutation of this salt bridge results in a rapid Ca2+ desensitization, similar to a direct mutation of the predicted PIP2 binding residues[38].
F608 and its sequential and structural neighbors are conserved among ANO1 orthologs (**Supplementary Figure 6**). The variant causes a change from the aromatic and very hydrophobic phenylalanine to the smaller and polar/hydrophilic serine. All members of the TMEM16 superfamily conserved a non-polar residue at this position, suggesting that the polar sidechain of the serine may cause a structural conflict in the region. The variant could interfere with the K609-E594 salt bridge which stabilizes the PIP2 binding. F608S may thus interfere with the PIP2 binding and consequently accelerates ANO1 degradation, similar to the rapid desensitization that was demonstrated by mutational analyses of basic amino acids in the vincitiy and the salt bridge[38].

The SRPX (also DRS, ETX1, SRPX1) variant rs35318931 causes a Ser413Phe exchange at the C-terminal domain of unknown function (**Supplementary Figure 7**). The protein is further composed of three Sushi domains, and one HYR domain. Sushi domains are components involved in extracellular protein-protein interactions and are often found in complement control proteins[41]. The HYR (hyalin repeat) domain is predicted to contribute to cell adhesion since the domain enables the hyalin protein to bind to the receptor[42]. The SRPX C-terminal domain is a phylogenetically widespread protein domain that is well-conserved in vertebrates (**Supplementary Figure 7**) and also in many bacteria, and has been named the DUDES domain (DRO1-URB-DRS-Equarin-SRPX)[43]. Protein structural analyses assign a thioredoxin-like fold to this domain, although the location of potential functional cysteines seem unique for SRPX and SRPX2 proteins[44]. Therefore, the conserved structural core allows fold recognition, but the lack of suitable structural templates including loops and termini complicates in-silico functional prediction for SRPX (**Supplementary Figure 7**). SRPX was originally identified as a tumor suppressor[45] and, in this context, to the induction of apoptosis[46] and downregulation of glucose metabolism via Lactate dehydrogenase-B[47]. Proteomics studies found SRPX expression in the extracellular matrix (ECM) of different tissues (lung[48], cartilage[49] and colon and liver[50]) and is upregulated in the ECM during cardiac remodeling[51]. Further, SRPX was also shown to interact with PELO at the actin cytoskeleton[52].
Other members of the DUDES protein family were shown to localize in the extracellular matrix, e,g, SRPX2 in brain[53], equarin in chick lens[54]. CCDC80 is a remote homologous

that binds activated JAK2 and is consequently more abundant in the extracellular matrix. JAK2-binding was also detected by the paralog SRPX2, and interaction is therefore also predicted for SRPX[55]. CCDC80 is composed of three DUDES domains, that are independently able to bind JAK2, assuming the SRPX DUDES domain is responsible for protein association with the ECM. The ECM provides structural integrity for tissues, and involves in cell differentiation, activation and migration. HEM tissue is less stable and show abnormalities in the ECM collagen composition (compared to healthy tissue[56]).

The variant Ser413Phe locates at the beginning of strand 3 in the central beta sheet. The preceding loop is highly variable among homologs[44] but the following strand is one of the best conserved regions within the protein family, including an invariant F414. The change from the polar and small amino acid serine to the larger, aromatic and hydrophobic phenylalanine potentially destabilizes the domain structure due to its location adjacent the conserved hydrophobic core of the protein fold.

The SRPX domain structure was derived from the UniProt database and by search against the NCBI Conserved Domains Database (CDD). SRPX and SRPX2 protein sequences were derived from UniProt, Ensembl and the consensus sequence of pfam13778 from the CDD. Sequence alignments were conducted using Muscle. The sequence alignment was visualized using JalView applying the Clustal coloring scheme. Protein sequence identifiers UniProt or Ensembl: SRPX: human, P78539; mouse, Q9R0M3; cow, F1MQX1; zebrafish, Q58ED3; xenopus tropicalis, ENSXETT00000018780.4. SRPX2: human, O60687; mouse, Q8R054; cow, Q5EA25; zebrafish, E7F8X0, xenopus tropicalis, ENSXETT00000014699.4.

The structure-based alignment for modeling the SRPX C-terminal domain of unknown function (DUF4174/pfam13778, 332-451) is based on secondary structure predictions, structural alignments of two templates (PDBs 3drn/chain A, 3cmi/chain A) and multiple sequence alignment including the consensus sequence of pfam13778. Structural models of SRPX and ANO1 were visualized using PyMOL.

#### Supplementary Figures

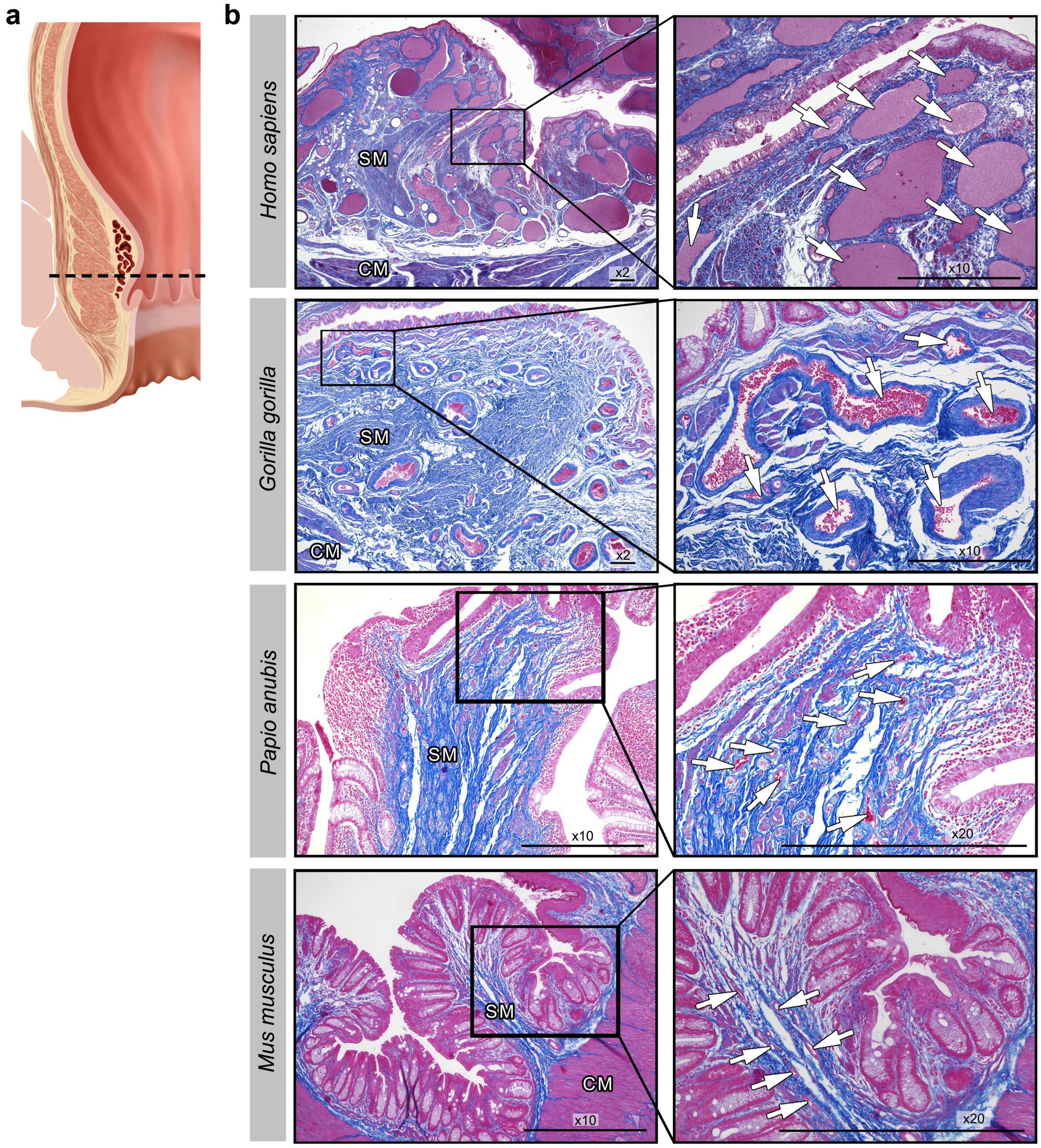

**Supplementary Figure 1.** *Histological analysis of the anorectum in four different species.* The left panel **(a)** shows the section plane of the anal canal at the level of the hemorrhoidal plexus. Panel **(b)** shows the hemorrhoidal plexus of 4 different species: *Homo sapiens* (top row), *Gorilla gorilla* (second row), baboon (*Papio anubis; third row*), and mouse (10-week old male C57BL/6JRj mouse; bottom row). While the human anorectum shows a well-developed hemorrhoidal plexus with densely packed blood vessels of large diameters, the gorilla sample displays a rudimentary hemorrhoidal plexus with fewer and smaller blood vessels. Both baboon and mouse samples exhibit only small-sized and scattered blood vessels which resemble normal vascularization patterns of the regular rectal mucosa. Azan staining with visualization of connective tissue (blue) as well as cell nuclei, erythrocytes and smooth muscle (all purple red). Magnifications for human and gorilla (left 2x, right 10x), for baboon and mouse (left 10x, right 20x). Scale bars: 500 µm. White arrows: hemorrhoidal/submucosal blood vessels, SM = submucosa, CM = circular muscle layer/internal anal sphincter.

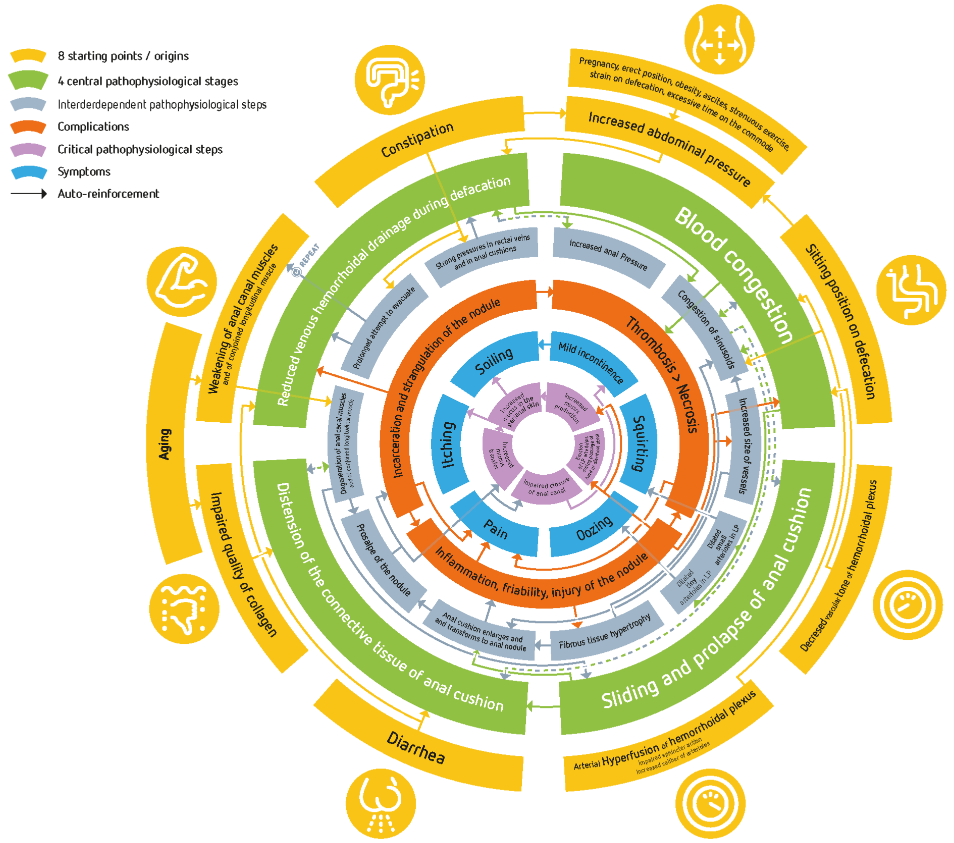

**Supplementary Figure 2.** *Suggested integrated model that summarizes the contemporary thinking on the pathophysiology of HEM (figure and legend are mainly taken from Figure 2 in Nikolaos Margetis' review[57]).*

HEM is a complex and multifactorial disease, most likely resulting from separate origins and combinations thereof. Different origins and causes have been suggested (orange), which force the hemorrhoidal plexus in different abnormal directions and probably converge in four central pathophysiological events (green). Different consecutive pathophysiological stages (grey) connect the primary causes and the 4 central events. These pathophysiological stages are interconnected, interdependent, and mutually reinforcing, creating a vicious cycle. This multidirectional network is continuously auto-reinforced, as shown by the arrows, and over time provides only one outcome, with the hemorrhoids deteriorating. Ultimately, symptoms (blue) and complications (red) occur. For further details we refer to Margetis' review[57].

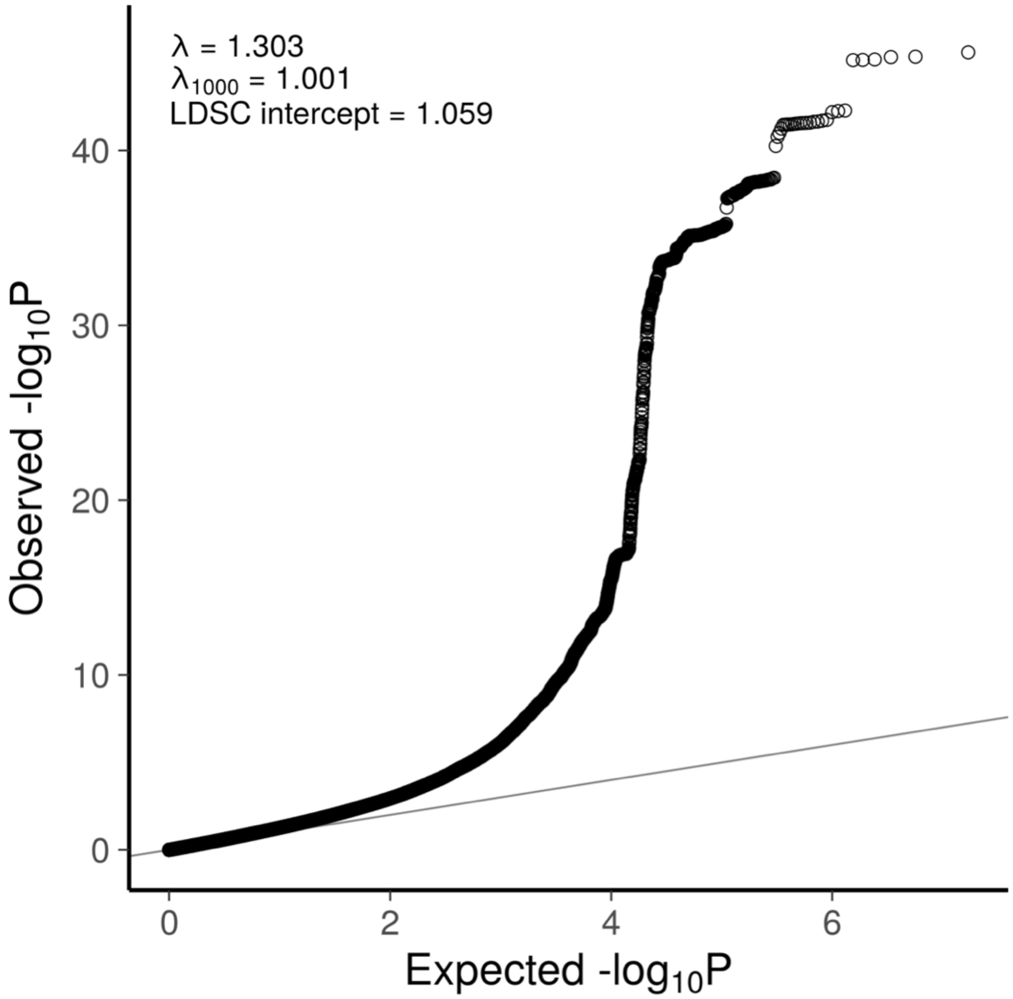

**Supplementary Figure 3.** *Quantile-quantile (QQ) plot of GWAS meta-analysis results.* Only markers that passed the imputation quality score $R^2$>0.8 and MAF>1% were used for the plot. The genomic inflation factor lambda (λ) is defined as the ratio of the medians of the sample χ2 test statistics and the 1-d.f. χ2 distribution (0.455)[58]. Lambda inflation statistics are influenced by the sample size. To facilitate comparison with other studies, $\lambda_{1000}$ converts a given lambda from n cases and m controls so that the value corresponds to an analysis with 1000 cases and 1000 controls. Although genomic inflation was observed (λ=1.303) this was probably due to polygenicity rather than population stratification as determined by linkage disequilibrium score regression analysis (LDSC, intercept=1.059)[59].

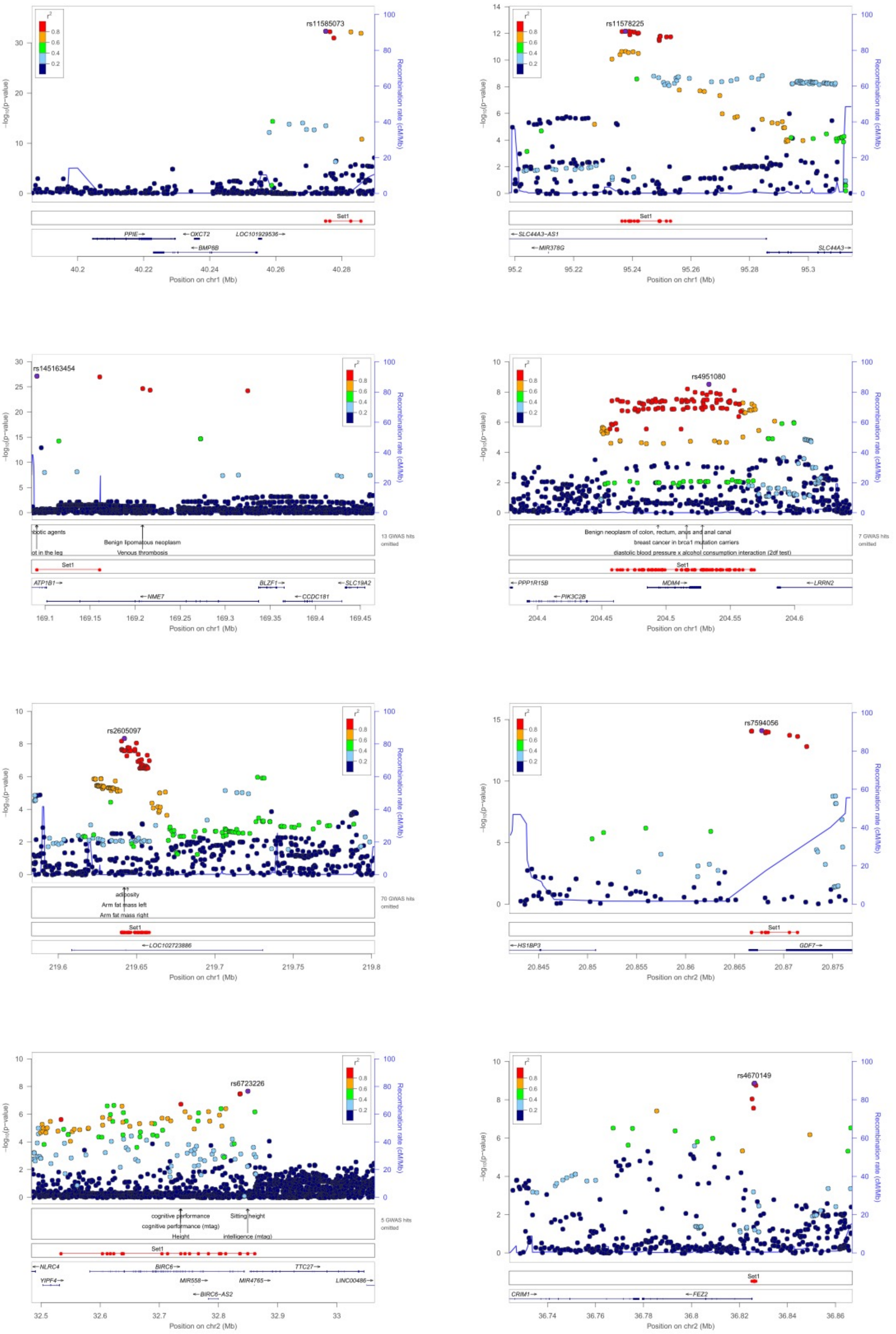
rs11585073
Position on chr1 (Mb)
PPIE
OXCT2
LOC101929536
BMP8B
rs11578225
SLC44A3-AS1
MIR378G
SLC44A3
rs145163454
Benign lipomatous neoplasm
Venous thrombosis
ATP1B1
NME7
BLZF1
CCDC181
SLC19A2
13 GWAS hits omitted
rs4951080
Benign neoplasm of colon, rectum, anus and anal canal
breast cancer in brca1 mutation carriers
diastolic blood pressure x alcohol consumption interaction (2df test)
7 GWAS hits omitted
PPP1R15B
PIK3C2B
MDM4
LRRN2
rs2605097
adiposity
Arm fat mass left
Arm fat mass right
70 GWAS hits omitted
LOC102723886
rs7594056
Position on chr2 (Mb)
HS1BP3
GDF7
rs6723226
cognitive performance
cognitive performance (mtag)
Height
Sitting height
intelligence (mtag)
5 GWAS hits omitted
NLRC4
YIPF4
BIRC6
MIR558
MIR4765
BIRC6-AS2
TTC27
LINC00486
rs4670149
CRIM1
FEZ2
Set1
Recombination rate (cM/Mb)

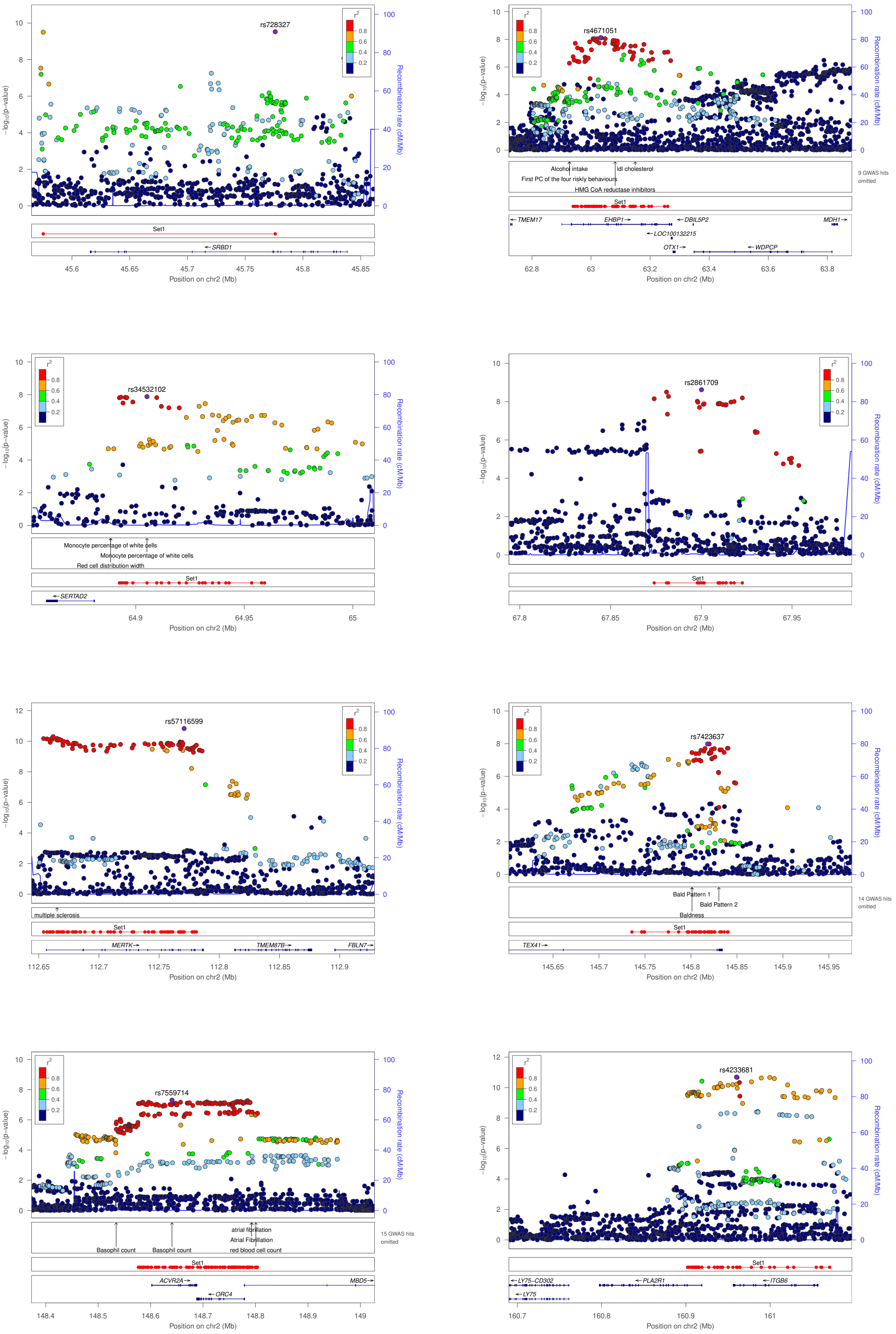

rs728327
SRBD1
Set1
Position on chr2 (Mb)
−log10(p−value)
Recombination rate (cM/Mb)
rs4671051
Alcohol intake
ldl cholesterol
First PC of the four riskly behaviours
HMG CoA reductase inhibitors
9 GWAS hits omitted
TMEM17
EHBP1
DBIL5P2
LOC100132215
OTX1
WDPCP
MDH1
rs34532102
Monocyte percentage of white cells
Red cell distribution width
SERTAD2
rs2861709
rs57116599
multiple sclerosis
MERTK
TMEM87B
FBLN7
rs7423637
Bald Pattern 1
Bald Pattern 2
Baldness
14 GWAS hits omitted
TEX41
rs7559714
Basophil count
atrial fibrillation
Atrial Fibrillation
red blood cell count
15 GWAS hits omitted
ACVR2A
ORC4
MBD5
rs4233681
LY75−CD302
LY75
PLA2R1
ITGB6

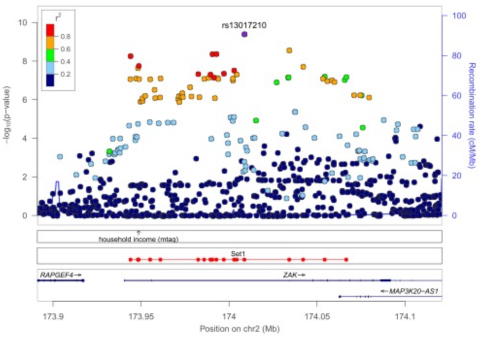
rs13017210
household income (mtag)
Set1
RAPGEF4
ZAK
MAP3K20-AS1
Position on chr2 (Mb)
Recombination rate (cM/Mb)

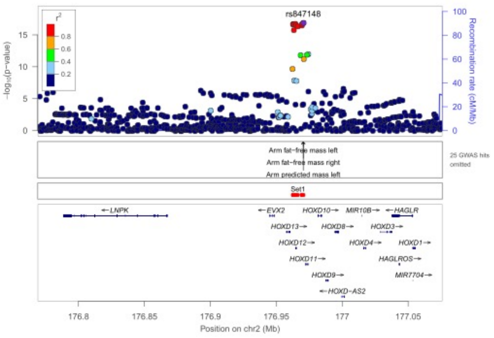
rs847148
Arm fat-free mass left
Arm fat-free mass right
Arm predicted mass left
25 GWAS hits omitted
Set1
LNPK
EVX2
HOXD13
HOXD12
HOXD11
HOXD10
HOXD8
HOXD9
HOXD-AS2
MIR10B
HOXD4
HOXD3
HAGLR
HOXD1
HAGLROS
MIR7704
Position on chr2 (Mb)
Recombination rate (cM/Mb)

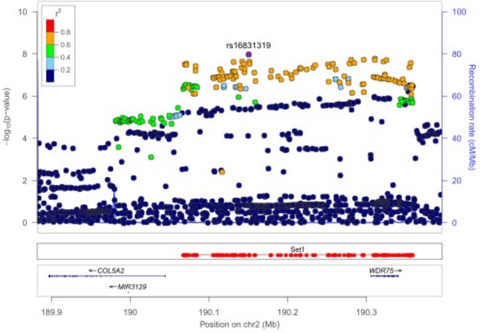
rs16831319
Set1
COL5A2
MIR3129
WDR75
Position on chr2 (Mb)
Recombination rate (cM/Mb)

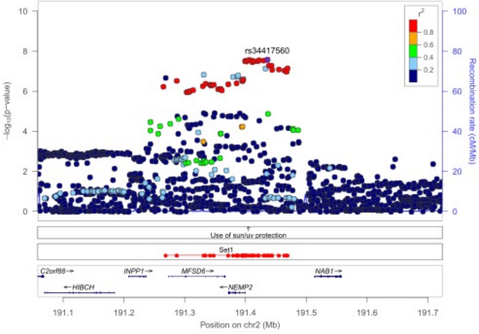
rs34417560
Use of sun/uv protection
Set1
C2orf88
HIBCH
INPP1
MFSD6
NEMP2
NAB1
Position on chr2 (Mb)
Recombination rate (cM/Mb)

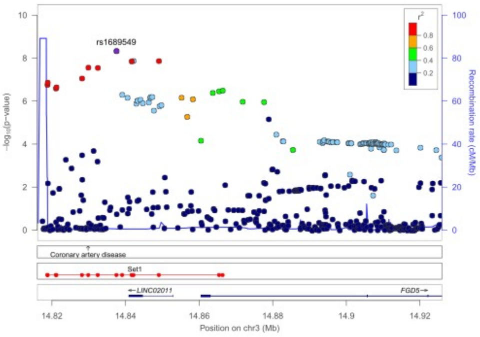
rs1689549
Coronary artery disease
Set1
LINC02011
FGD5
Position on chr3 (Mb)
Recombination rate (cM/Mb)

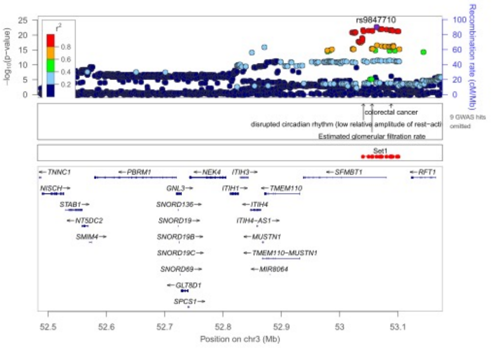
rs9847710
colorectal cancer
disrupted circadian rhythm (low relative amplitude of rest-acti
Estimated glomerular filtration rate
9 GWAS hits omitted
Set1
TNNC1
NISCH
STAB1
NT5DC2
SMIM4
PBRM1
GNL3
SNORD136
SNORD19
SNORD19B
SNORD19C
SNORD69
GLT8D1
SPCS1
NEK4
ITIH3
ITIH1
ITIH4
ITIH4-AS1
MUSTN1
TMEM110
TMEM110-MUSTN1
MIR8064
SFMBT1
RFT1
Position on chr3 (Mb)
Recombination rate (cM/Mb)

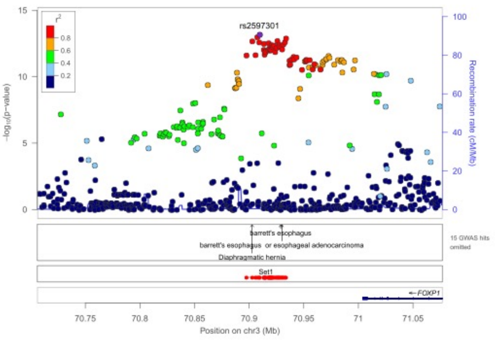
rs2597301
barrett's esophagus
barrett's esophagus or esophageal adenocarcinoma
Diaphragmatic hernia
15 GWAS hits omitted
Set1
FOXP1
Position on chr3 (Mb)
Recombination rate (cM/Mb)

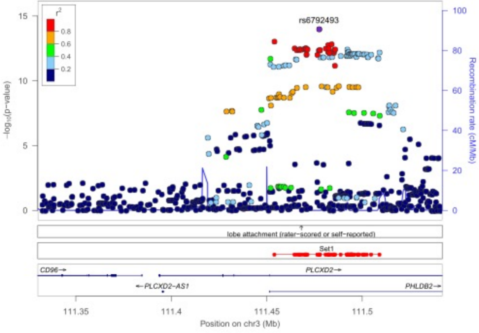
rs6792493
lobe attachment (rater-scored or self-reported)
Set1
CD96
PLCXD2-AS1
PLCXD2
PHLDB2
Position on chr3 (Mb)
Recombination rate (cM/Mb)

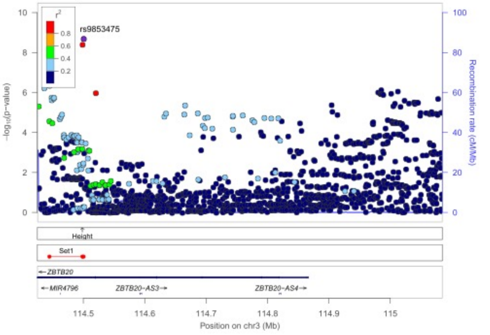
rs9853475
Height
Set1
ZBTB20
MIR4796
ZBTB20-AS3
ZBTB20-AS4
Position on chr3 (Mb)
Recombination rate (cM/Mb)

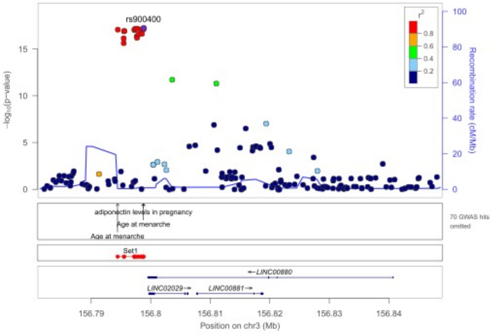
rs900400
adiponectin levels in pregnancy
Age at menarche
Age at menarche
70 GWAS hits omitted
Set1
LINC00880
LINC02029
LINC00881
Position on chr3 (Mb)
Recombination rate (cM/Mb)

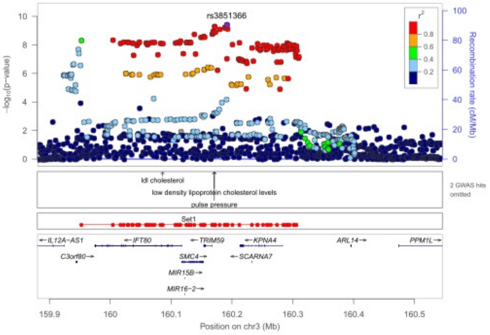
rs3851366
ldl cholesterol
low density lipoprotein cholesterol levels
pulse pressure
2 GWAS hits omitted
Set1
IL12A-AS1
IFT80
TRIM59
KPNA4
ARL14
PPM1L
C3orf80
SMC4
SCARNA7
MIR15B
MIR16-2
Position on chr3 (Mb)
Recombination rate (cM/Mb)

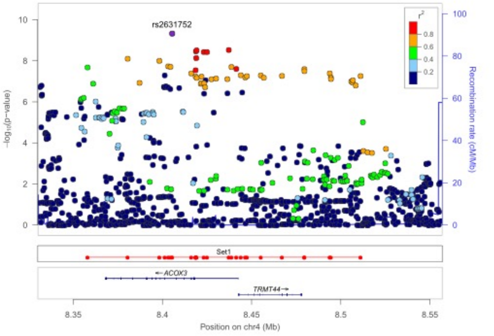
rs2631752
Set1
ACOX3
TRMT44
Position on chr4 (Mb)
Recombination rate (cM/Mb)

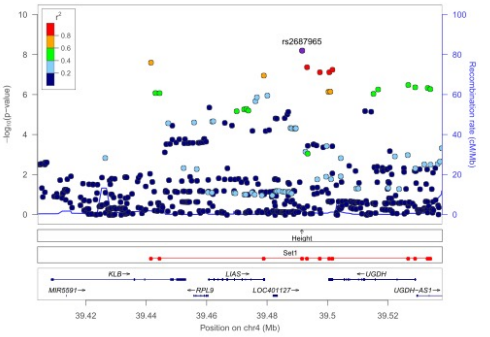
rs2687965
Height
Set1
KLB
LIAS
UGDH
MIR5591
RPL9
LOC401127
UGDH-AS1
Position on chr4 (Mb)
Recombination rate (cM/Mb)

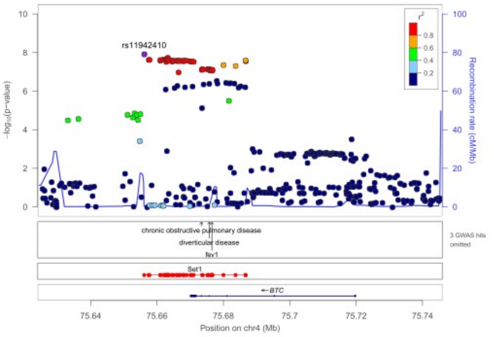
rs11942410
chronic obstructive pulmonary disease
diverticular disease
fev1
3 GWAS hits omitted
Set1
BTC
Position on chr4 (Mb)
Recombination rate (cM/Mb)

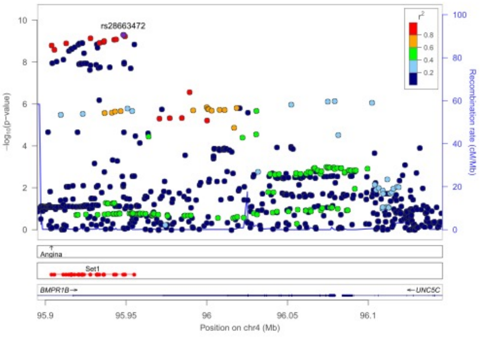
rs28663472
Angina
Set1
BMPR1B
UNC5C
Position on chr4 (Mb)
Recombination rate (cM/Mb)

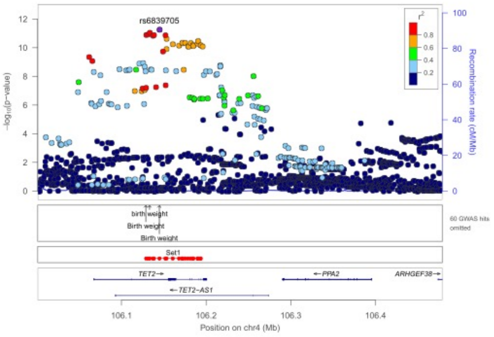
rs6839705
birth weight
Birth weight
Birth weight
60 GWAS hits omitted
Set1
TET2
PPA2
ARHGEF38
TET2-AS1
Position on chr4 (Mb)
Recombination rate (cM/Mb)

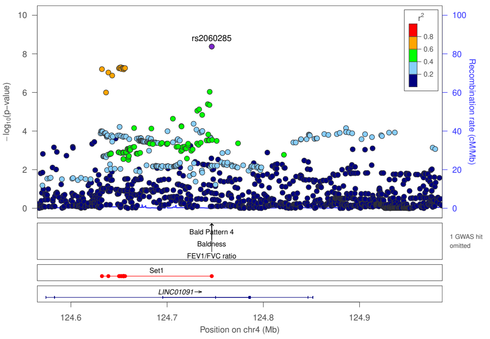
rs2060285
−log10(p−value)
Recombination rate (cM/Mb)
Bald Pattern 4
Baldness
FEV1/FVC ratio
1 GWAS hit omitted
Set1
LINC01091→
Position on chr4 (Mb)

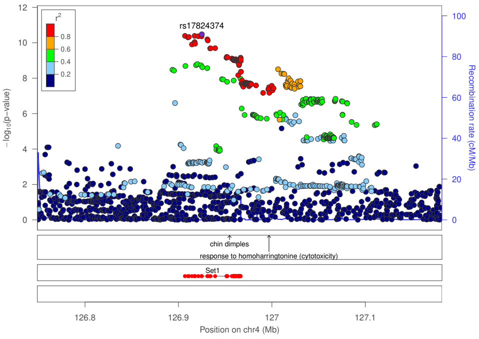
rs17824374
−log10(p−value)
Recombination rate (cM/Mb)
chin dimples
response to homoharringtonine (cytotoxicity)
Set1
Position on chr4 (Mb)

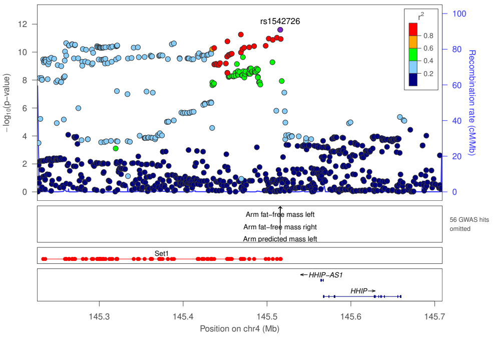
rs1542726
−log10(p−value)
Recombination rate (cM/Mb)
Arm fat−free mass left
Arm fat−free mass right
Arm predicted mass left
56 GWAS hits omitted
Set1
←HHIP−AS1
HHIP→
Position on chr4 (Mb)

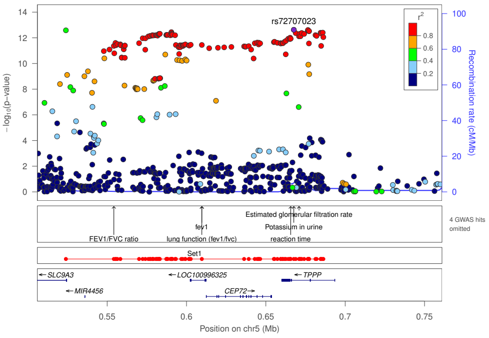
rs72707023
−log10(p−value)
Recombination rate (cM/Mb)
Estimated glomerular filtration rate
Potassium in urine
reaction time
fev1
FEV1/FVC ratio
lung function (fev1/fvc)
4 GWAS hits omitted
Set1
←SLC9A3
←LOC100996325
←TPPP
←MIR4456
CEP72→
Position on chr5 (Mb)

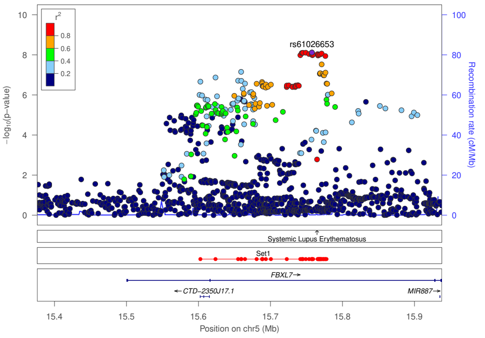
rs61026653
−log10(p−value)
Recombination rate (cM/Mb)
Systemic Lupus Erythematosus
Set1
FBXL7→
←CTD−2350J17.1
MIR887→
Position on chr5 (Mb)

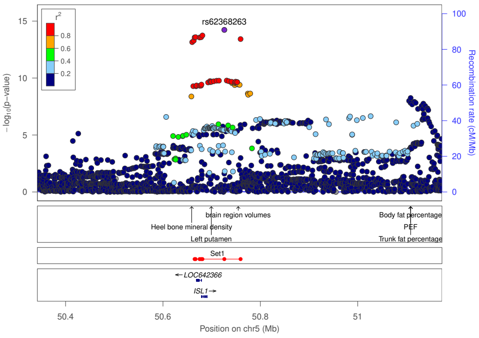
rs62368263
−log10(p−value)
Recombination rate (cM/Mb)
brain region volumes
Heel bone mineral density
Left putamen
Body fat percentage
PEF
Trunk fat percentage
Set1
←LOC642366
ISL1→
Position on chr5 (Mb)

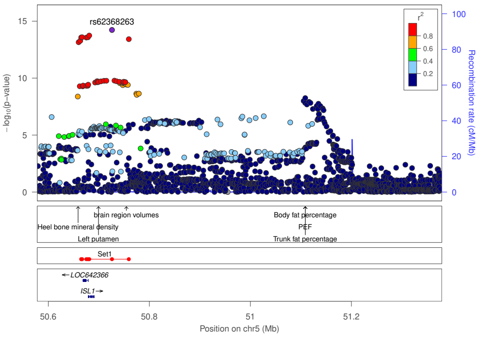
rs62368263
−log10(p−value)
Recombination rate (cM/Mb)
brain region volumes
Heel bone mineral density
Left putamen
Body fat percentage
PEF
Trunk fat percentage
Set1
←LOC642366
ISL1→
Position on chr5 (Mb)

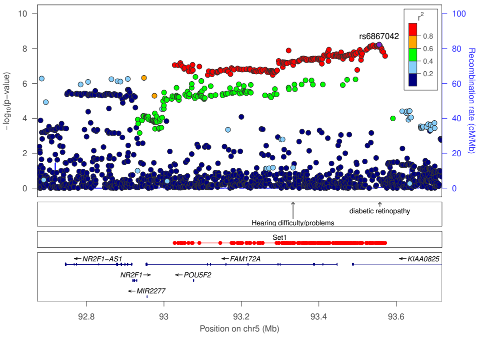
rs6867042
−log10(p−value)
Recombination rate (cM/Mb)
diabetic retinopathy
Hearing difficulty/problems
Set1
←NR2F1−AS1
←FAM172A
←KIAA0825
NR2F1→
←POU5F2
←MIR2277
Position on chr5 (Mb)

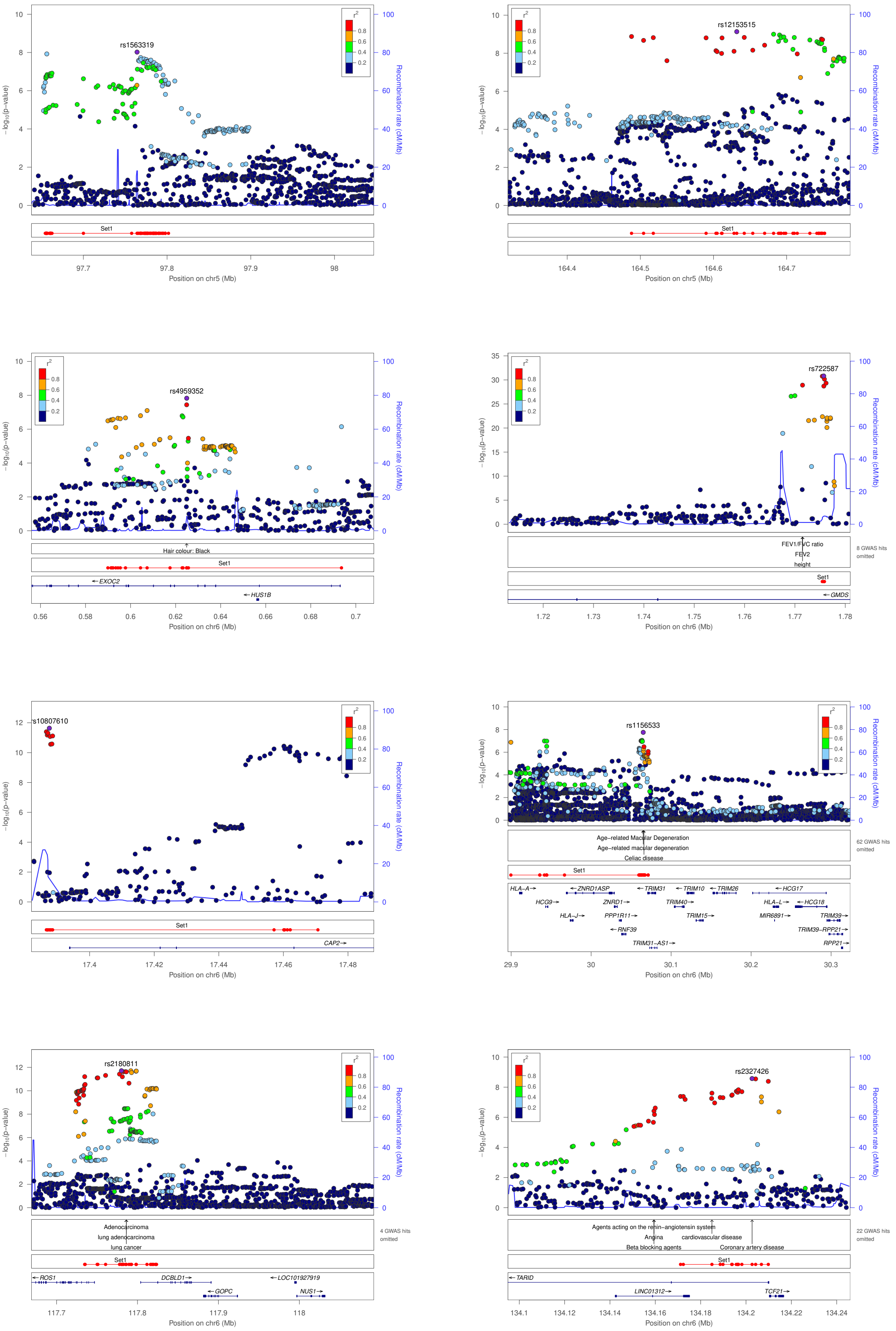

rs1563319
Position on chr5 (Mb)
rs12153515
Position on chr5 (Mb)
rs4959352
Hair colour: Black
EXOC2
HUS1B
Position on chr6 (Mb)
rs722587
FEV1/FVC ratio
FEV2
height
8 GWAS hits omitted
GMDS
Position on chr6 (Mb)
rs10807610
CAP2
Position on chr6 (Mb)
rs1156533
Age−related Macular Degeneration
Age−related macular degeneration
Celiac disease
62 GWAS hits omitted
HLA−A
ZNRD1ASP
TRIM31
TRIM10
TRIM26
HCG17
HCG9
ZNRD1
TRIM40
HLA−L
HCG18
HLA−J
PPP1R11
TRIM15
MIR6891
TRIM39
RNF39
TRIM39−RPP21
TRIM31−AS1
RPP21
Position on chr6 (Mb)
rs2180811
Adenocarcinoma
lung adenocarcinoma
lung cancer
4 GWAS hits omitted
ROS1
DCBLD1
LOC101927919
GOPC
NUS1
Position on chr6 (Mb)
rs2327426
Agents acting on the renin−angiotensin system
Angina
cardiovascular disease
Beta blocking agents
Coronary artery disease
22 GWAS hits omitted
TARID
LINC01312
TCF21
Position on chr6 (Mb)
−log10(p−value)
Recombination rate (cM/Mb)
Set1

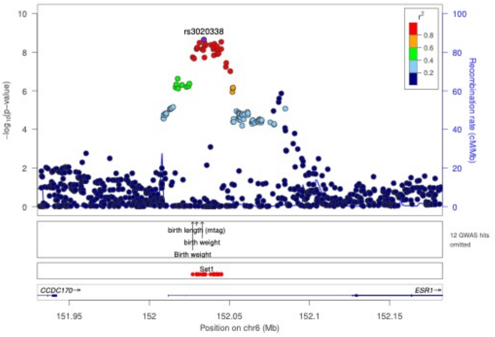
rs3020338
birth length (mtag)
birth weight
Birth weight
12 GWAS hits omitted
Set1
CCDC170
ESR1
Position on chr6 (Mb)
Recombination rate (cM/Mb)
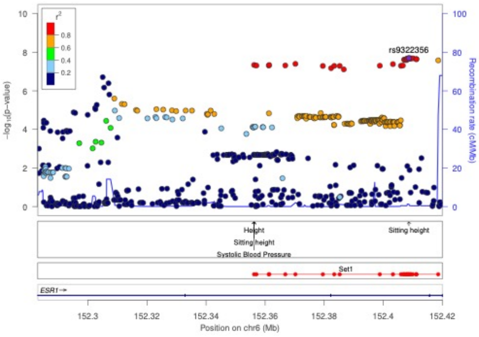
rs9322356
Height
Sitting height
Systolic Blood Pressure
Sitting height
Set1
ESR1
Position on chr6 (Mb)
Recombination rate (cM/Mb)
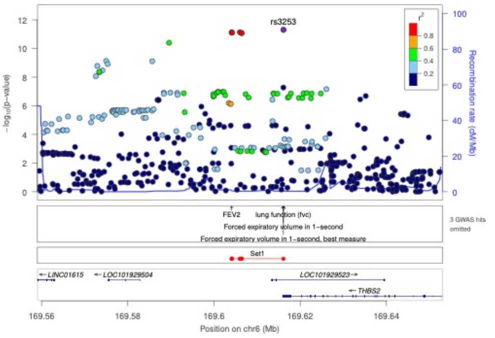
rs3253
FEV2
lung function (fvc)
Forced expiratory volume in 1−second
Forced expiratory volume in 1−second, best measure
3 GWAS hits omitted
Set1
LINC01615
LOC101929504
LOC101929523
THBS2
Position on chr6 (Mb)
Recombination rate (cM/Mb)
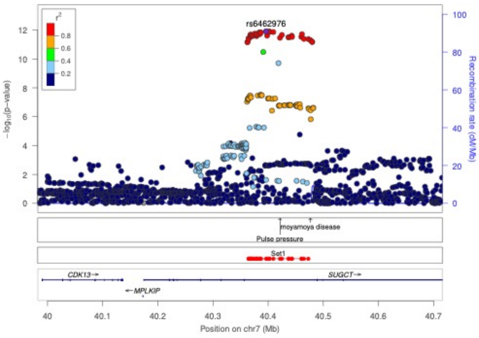
rs6462976
moyamoya disease
Pulse pressure
Set1
CDK13
SUGCT
MPLKIP
Position on chr7 (Mb)
Recombination rate (cM/Mb)
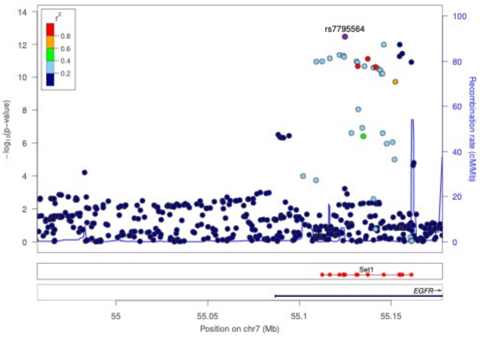
rs7795564
Set1
EGFR
Position on chr7 (Mb)
Recombination rate (cM/Mb)
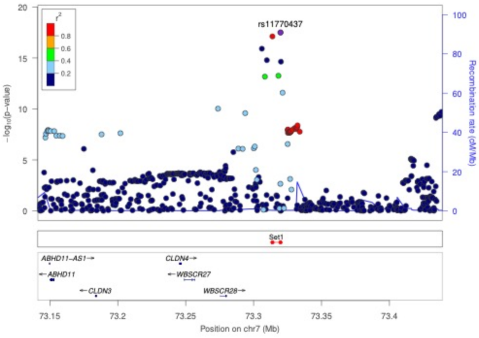
rs11770437
Set1
ABHD11−AS1
ABHD11
CLDN3
CLDN4
WBSCR27
WBSCR28
Position on chr7 (Mb)
Recombination rate (cM/Mb)
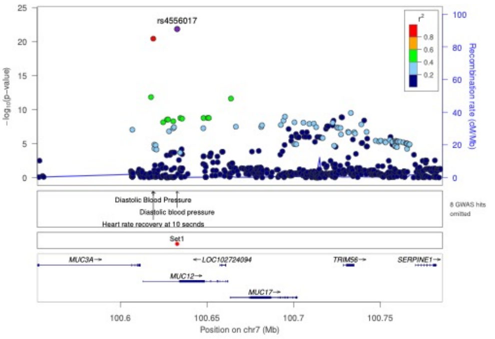
rs4556017
Diastolic Blood Pressure
Diastolic blood pressure
Heart rate recovery at 10 seconds
8 GWAS hits omitted
Set1
MUC3A
LOC102724094
TRIM56
SERPINE1
MUC12
MUC17
Position on chr7 (Mb)
Recombination rate (cM/Mb)
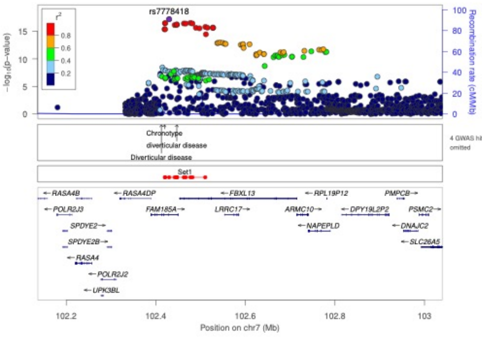
rs7778418
Chronotype
diverticular disease
Diverticular disease
4 GWAS hits omitted
Set1
RASA4B
RASA4DP
FBXL13
RPL19P12
PMPCB
POLR2J3
FAM185A
LRRC17
ARMC10
DPY19L2P2
PSMC2
SPDYE2
NAPEPLD
DNAJC2
SPDYE2B
SLC26A5
RASA4
POLR2J2
UPK3BL
Position on chr7 (Mb)
Recombination rate (cM/Mb)

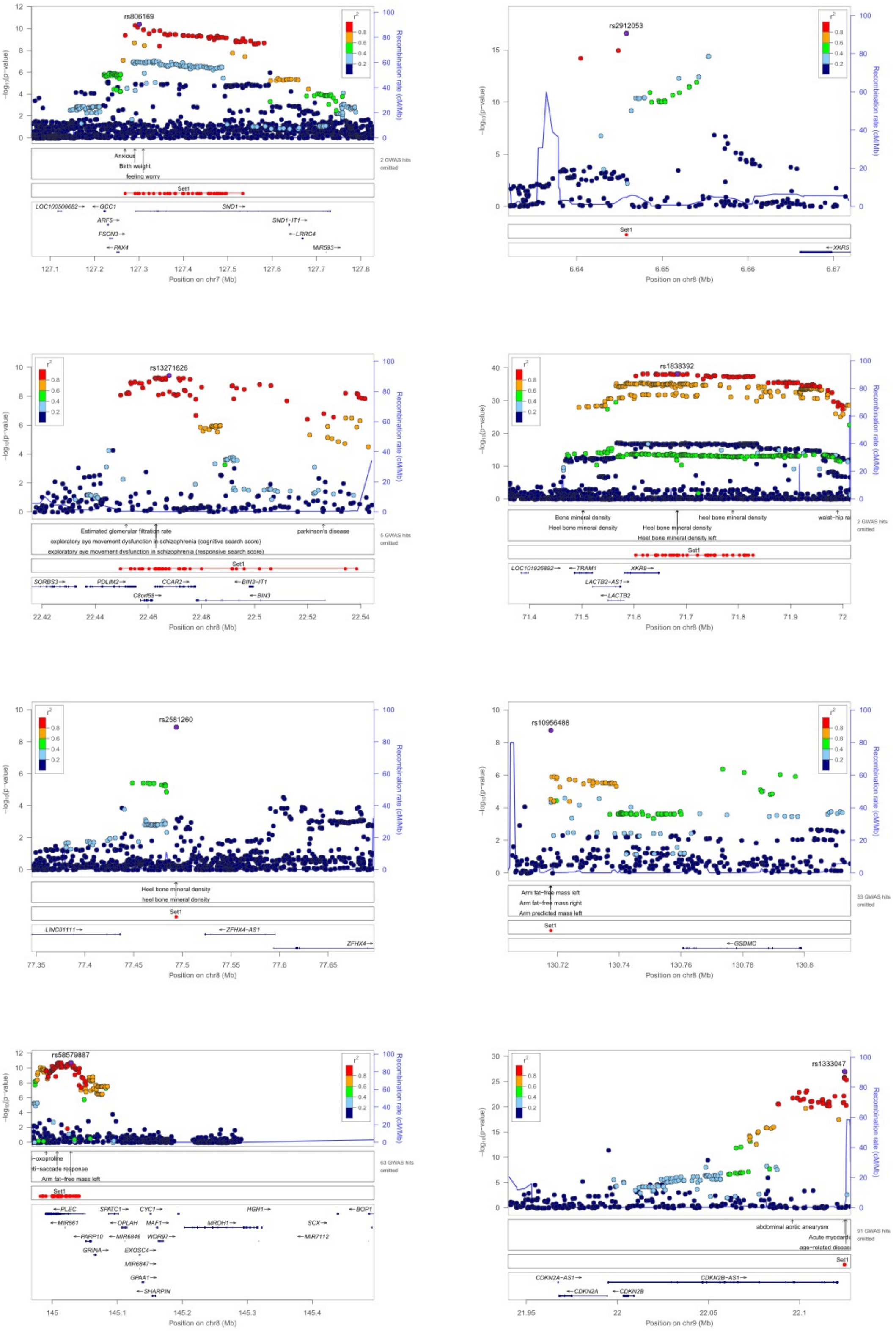
rs806169
Position on chr7 (Mb)
rs2912053
Position on chr8 (Mb)
rs13271626
Position on chr8 (Mb)
rs1838392
Position on chr8 (Mb)
rs2581260
Position on chr8 (Mb)
rs10956488
Position on chr8 (Mb)
rs58579887
Position on chr8 (Mb)
rs1333047
Position on chr9 (Mb)
Recombination rate (cM/Mb)
Set1

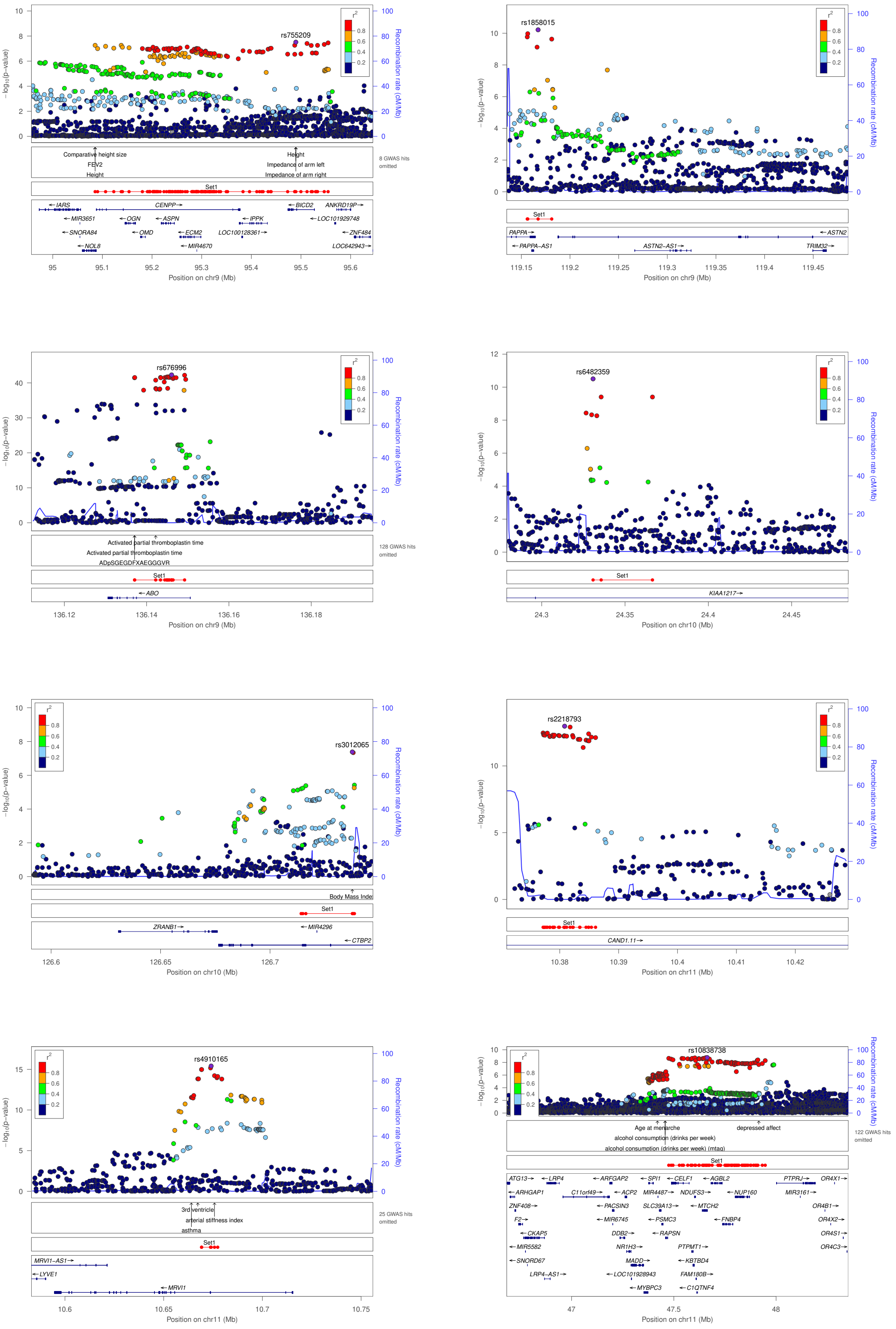

rs755209
Comparative height size
FEV2
Height
Impedance of arm left
Impedance of arm right
8 GWAS hits omitted
Set1
Position on chr9 (Mb)
rs1858015
Position on chr9 (Mb)
rs676996
Activated partial thromboplastin time
ADpSGEGDFXAEGGGVR
128 GWAS hits omitted
Position on chr9 (Mb)
rs6482359
Position on chr10 (Mb)
rs3012065
Body Mass Index
Position on chr10 (Mb)
rs2218793
Position on chr11 (Mb)
rs4910165
3rd ventricle
arterial stiffness index
asthma
25 GWAS hits omitted
Position on chr11 (Mb)
rs10838738
Age at menarche
alcohol consumption (drinks per week)
alcohol consumption (drinks per week) (mtag)
depressed affect
122 GWAS hits omitted
Position on chr11 (Mb)
−log10(p−value)
Recombination rate (cM/Mb)

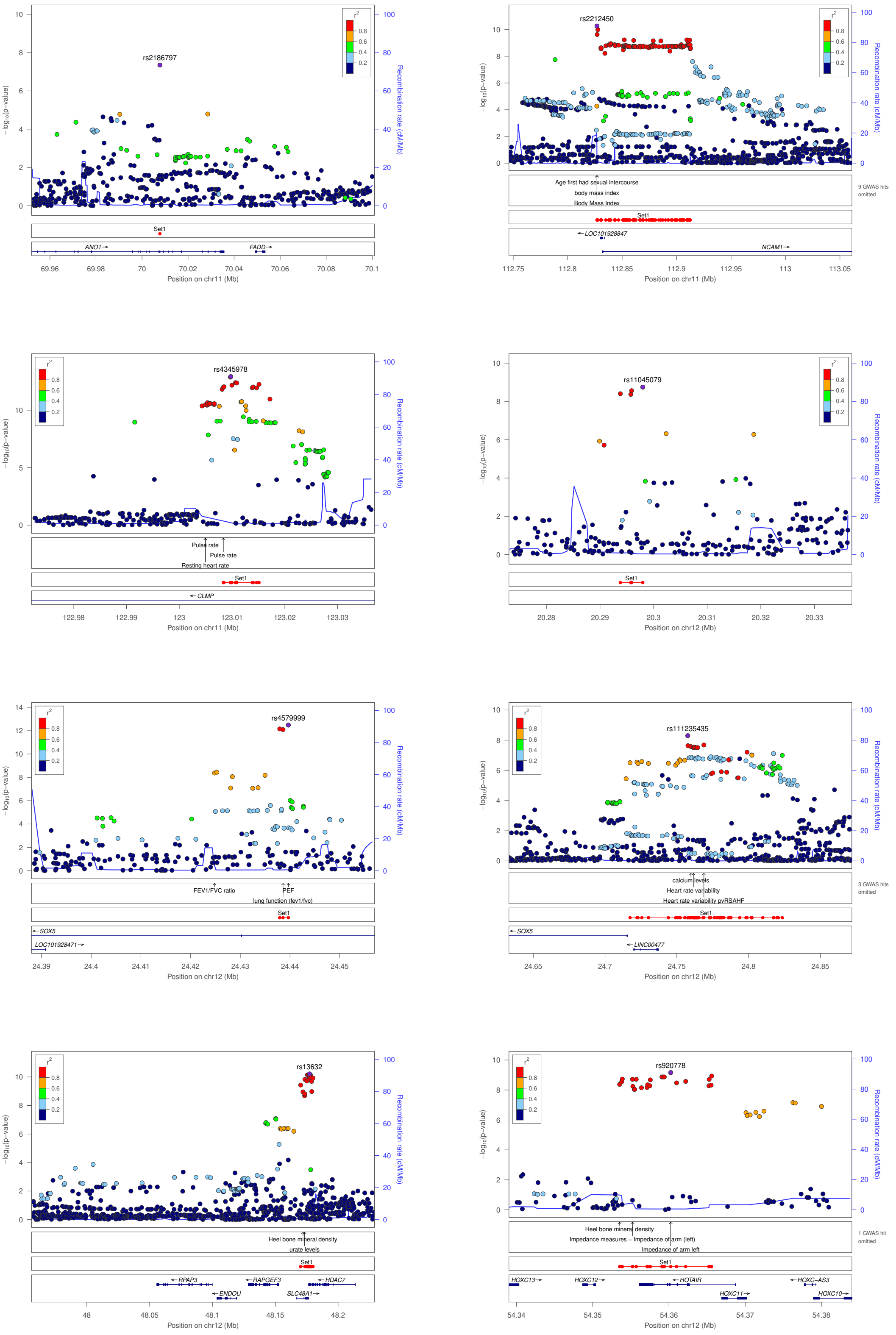

rs2186797
ANO1→
FADD→
Position on chr11 (Mb)
rs2212450
Age first had sexual intercourse
body mass index
Body Mass Index
9 GWAS hits omitted
←LOC101928847
NCAM1→
Position on chr11 (Mb)
rs4345978
Pulse rate
Pulse rate
Resting heart rate
←CLMP
Position on chr11 (Mb)
rs11045079
Position on chr12 (Mb)
rs4579999
FEV1/FVC ratio
PEF
lung function (fev1/fvc)
←SOX5
LOC101928471→
Position on chr12 (Mb)
rs111235435
calcium levels
Heart rate variability
Heart rate variability pvRSAHF
3 GWAS hits omitted
←SOX5
←LINC00477
Position on chr12 (Mb)
rs13632
Heel bone mineral density
urate levels
←RPAP3
←RAPGEF3
←HDAC7
←ENDOU
SLC48A1→
Position on chr12 (Mb)
rs920778
Heel bone mineral density
Impedance measures ~ Impedance of arm (left)
Impedance of arm left
1 GWAS hit omitted
HOXC13→
HOXC12→
←HOTAIR
←HOXC-AS3
HOXC11→
HOXC10→
Position on chr12 (Mb)
−log10(p−value)
Recombination rate (cM/Mb)
Set1

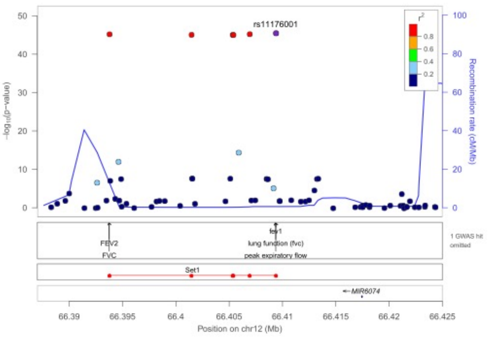
rs11176001
-log10(p-value)
Recombination rate (cM/Mb)
FEV2
FVC
fev1
lung function (fvc)
peak expiratory flow
1 GWAS hit omitted
Set1
MIR6074
Position on chr12 (Mb)

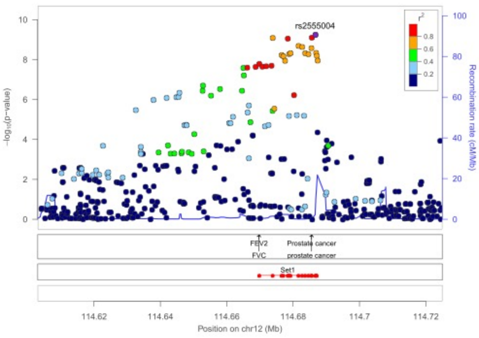
rs2555004
-log10(p-value)
Recombination rate (cM/Mb)
FEV2
FVC
Prostate cancer
prostate cancer
Set1
Position on chr12 (Mb)

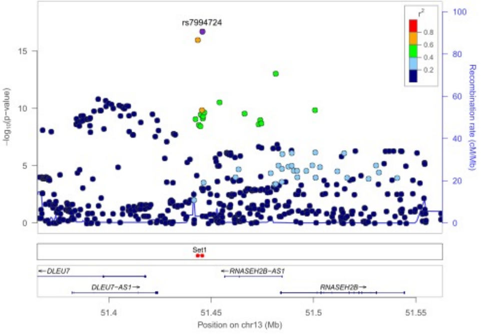
rs7994724
-log10(p-value)
Recombination rate (cM/Mb)
Set1
DLEU7
RNASEH2B-AS1
DLEU7-AS1
RNASEH2B
Position on chr13 (Mb)

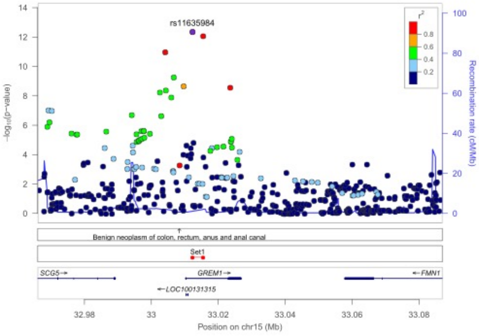
rs11635984
-log10(p-value)
Recombination rate (cM/Mb)
Benign neoplasm of colon, rectum, anus and anal canal
Set1
SCG5
GREM1
FMN1
LOC100131315
Position on chr15 (Mb)

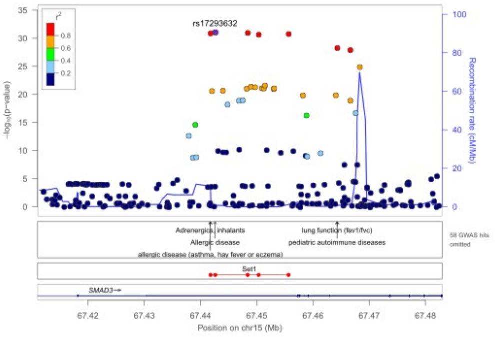
rs17293632
-log10(p-value)
Recombination rate (cM/Mb)
Adrenergics, inhalants
Allergic disease
allergic disease (asthma, hay fever or eczema)
lung function (fev1/fvc)
pediatric autoimmune diseases
58 GWAS hits omitted
Set1
SMAD3
Position on chr15 (Mb)

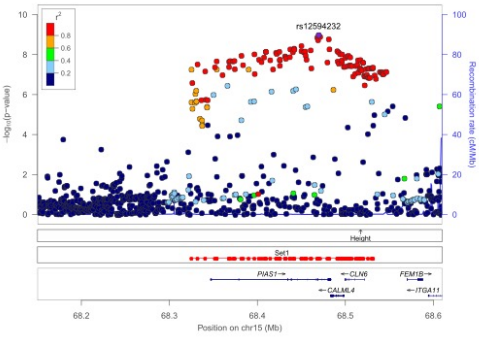
rs12594232
-log10(p-value)
Recombination rate (cM/Mb)
Height
Set1
PIAS1
CLN6
FEM1B
CALML4
ITGA11
Position on chr15 (Mb)

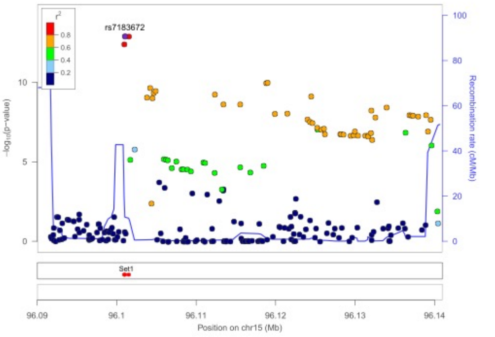
rs7183672
-log10(p-value)
Recombination rate (cM/Mb)
Set1
Position on chr15 (Mb)

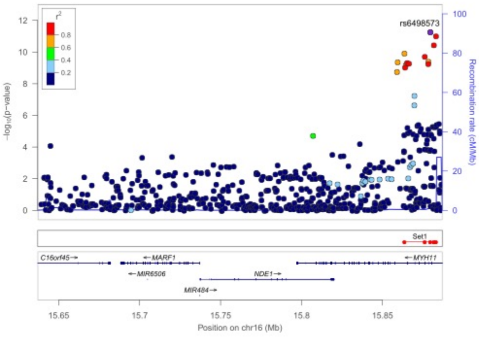
rs6498573
-log10(p-value)
Recombination rate (cM/Mb)
Set1
C16orf45
MARF1
MYH11
MIR6506
NDE1
MIR484
Position on chr16 (Mb)

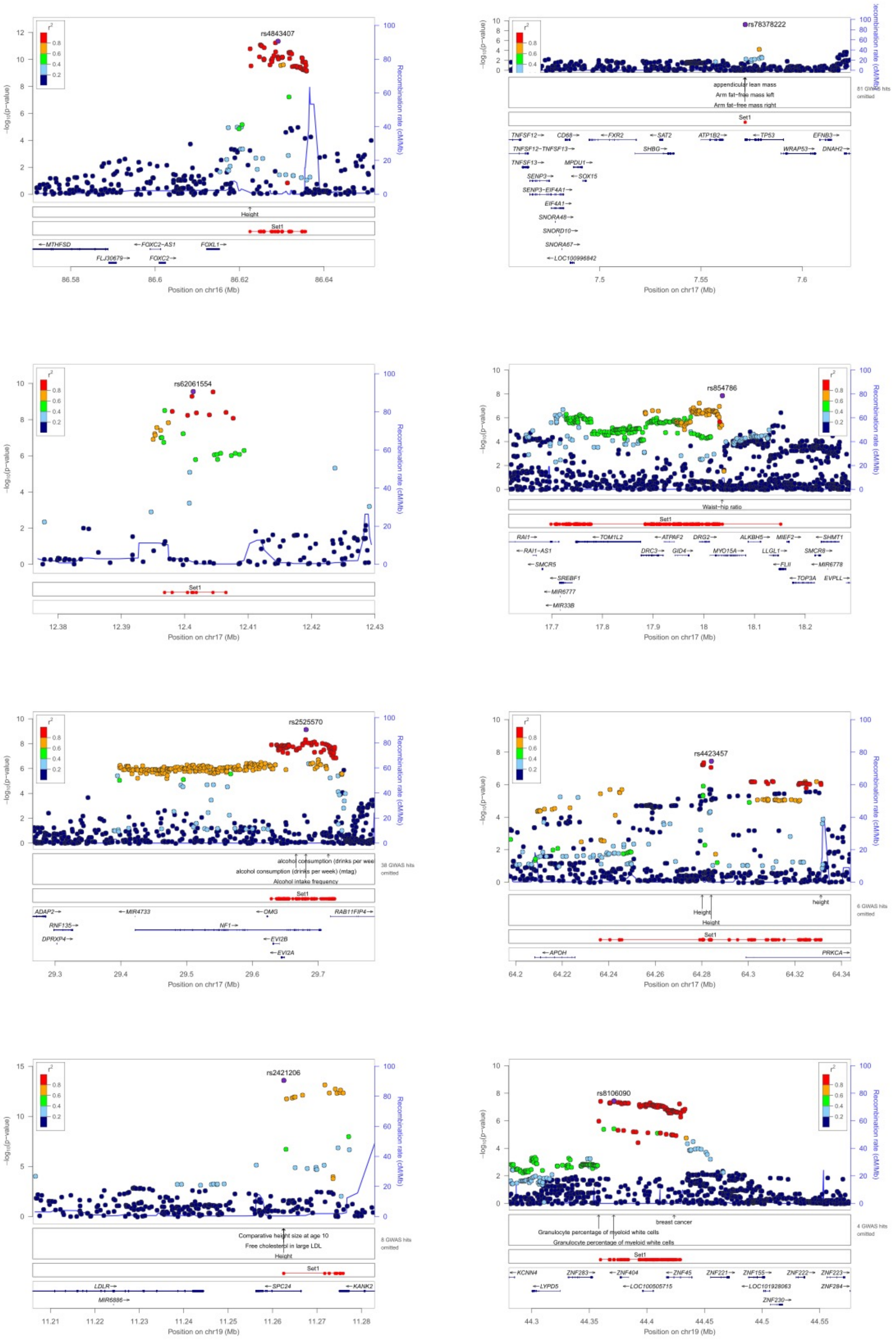
rs4843407
Position on chr16 (Mb)
rs78378222
Position on chr17 (Mb)
rs62061554
Position on chr17 (Mb)
rs854786
Waist−hip ratio
Position on chr17 (Mb)
rs2525570
Alcohol intake frequency
Position on chr17 (Mb)
rs4423457
Height
Position on chr17 (Mb)
rs2421206
Height
Position on chr19 (Mb)
rs8106090
breast cancer
Position on chr19 (Mb)
Recombination rate (cM/Mb)
Set1

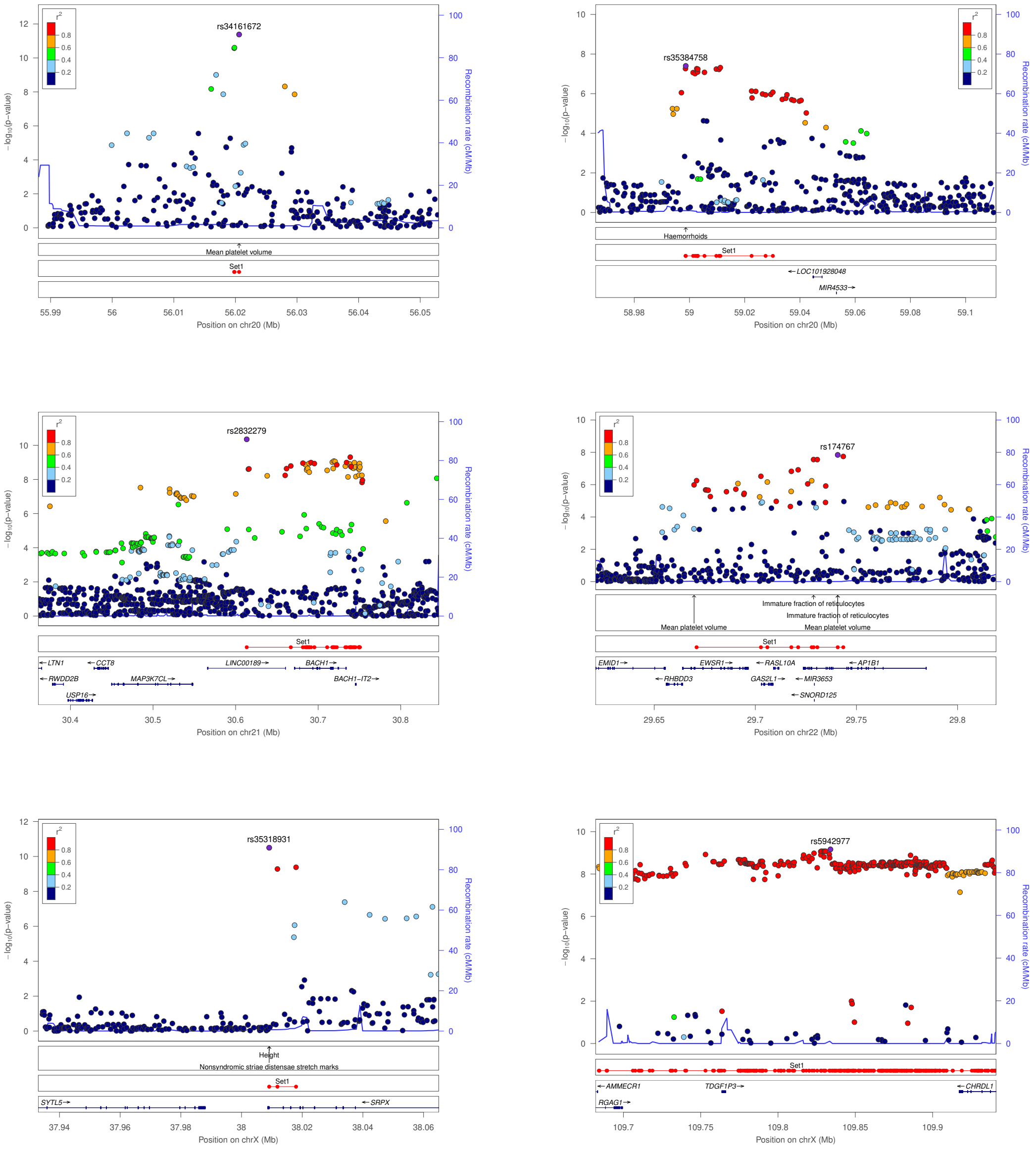

**Supplementary Figure 4.** *Regional association plots of HEM GWAS risk loci.* Shown are the -$\log_{10}$ *P*-values from meta-analysis with regard to the physical location of markers and the degree of linkage disequilibrium ($r^2$). Purple circle: lead SNP; line: recombination intensity (cM/Mb). Positions and gene annotations are according to NCBI's build 37 (hg19). Plots were generate using LocusZoom[60], also reporting the 95%-fine mapped credible sets at each locus.

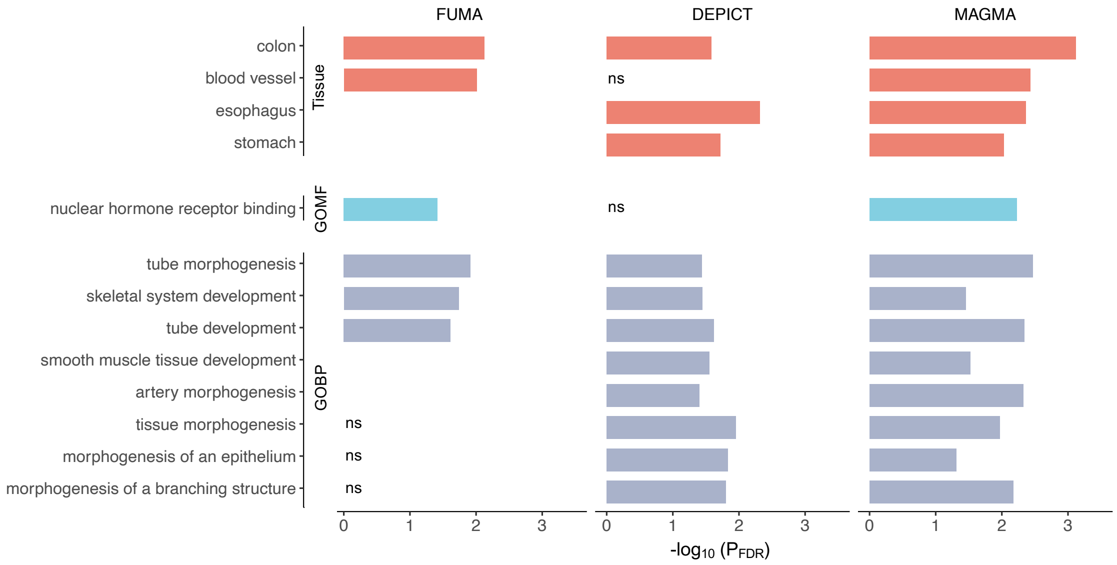

**Supplementary Figure 5.** *Gene set enrichment analyses of HEM genes.*

Tissues and pathways are shown, which resulted significantly enriched in at least 2/3 analyses (using FUMA, MAGMA or DEPICT generate HEM gene lists; **Methods**). Gene Ontology Biological Processes (GOBP) and Molecular Function (GOMF) categories are reported; ns=non-significant; some tissues/pathways were not available in all analyses (missing bars).

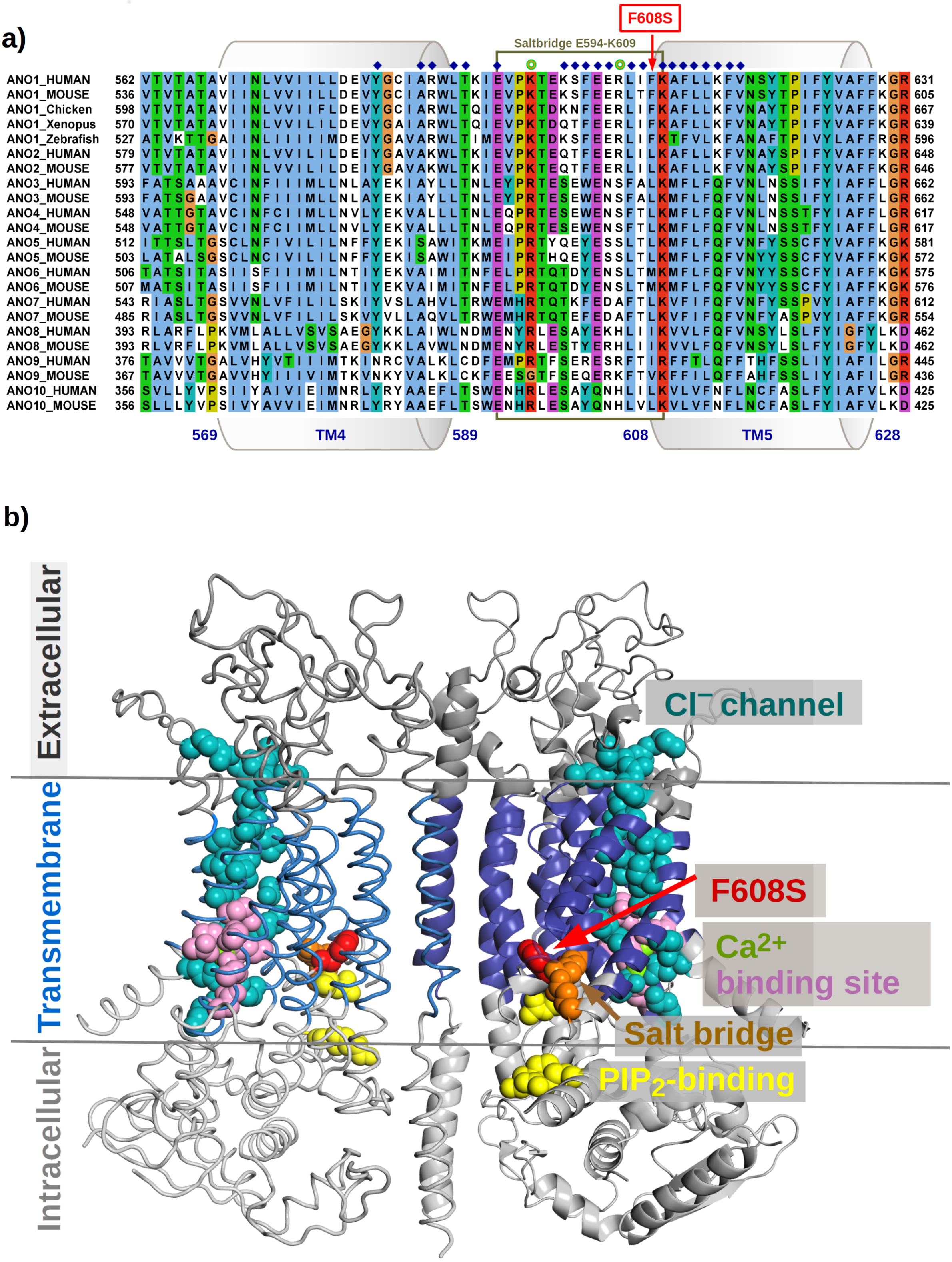
a)
F608S
Saltbridge E594-K609
ANO1_HUMAN
ANO1_MOUSE
ANO1_Chicken
ANO1_Xenopus
ANO1_Zebrafish
ANO2_HUMAN
ANO2_MOUSE
ANO3_HUMAN
ANO3_MOUSE
ANO4_HUMAN
ANO4_MOUSE
ANO5_HUMAN
ANO5_MOUSE
ANO6_HUMAN
ANO6_MOUSE
ANO7_HUMAN
ANO7_MOUSE
ANO8_HUMAN
ANO8_MOUSE
ANO9_HUMAN
ANO9_MOUSE
ANO10_HUMAN
ANO10_MOUSE
569
TM4
589
608
TM5
628
b)
Extracellular
Transmembrane
Intracellular
Cl⁻ channel
F608S
Ca2+ binding site
Salt bridge
PIP2-binding

**Supplementary Figure 6.** *ANO1 Alignment of TM4-5 and ANO1 structure.*

**(a)** Protein sequence alignment of ANO1 transmembrane helices TM4 and TM5 including the intracellular linker connected to TM5 via a salt bridge. Blue diamonds mark conserved amino acid positions next to and in close proximity to the F608S variant. Green spheres mark PIP2-binding positions K597 and R605, whose mutation has bee shown to lead to rapid channel inactivation through increased desensitization to Ca2+. The same effect was observed with the mutation of E594 or K609 which form a stabilizing salt bridge [38]. Only hydrophobic amino acids (blue) are conserved at the site of the F608S variant. It is therefore predicted that the mutation to the polar serine destabilizes the local protein structure and affects the integrity of this salt bridge. Accelerated desensitization of the anion channel may result from conformational changes of the putative PIP2 binding site due to a disruption of the salt bridge[38]. **Consequently, F608S may be able to down-regulate ANO1 activity.**

**(b)** Structural model of the ANO1 dimer and localization of the F608S variant. The F608S variant (red spheres) is located at the beginning of transmembrane helix 5 and thus at the membrane-cytosolic interface and a predicted PIiP2 interaction site[38] (yellow spheres). The exchange of the hydrophobic sidechain of phenylalanine (F) to a polar serine (S) within a conserved hydrophobic region is expected to destabilize the structure by disrupting the stability conducted by the salt bridge of K609 and E594 (orange spheres), which could accelerate the down-regulation of ANO1 by a faster channel inactivation by desensitization to Ca2+. This effect was shown by an alanine mutation of the salt bridge[38]. The ANO1 structural model is based on cryo-electron microscopy of the murine homolog (PDB ID 5oyb[39]) The two monomers are distinguished by representation as ribbons and cartoons, respectively.

Extracellular and intracellular domains are colored dark and light grey, the transmembrane domain is blue. The Cl anion channel is highlighted as teal spheres. The Ca2+-binding site is colored pink, the calcium atoms are shown as yellow-green spheres. Predicted PIP2-interacting residues (R481, K597 and R605[38]) are depicted as yellow spheres. Protein sequences were derived from the UniProt sequence database and visualized using JalView[61] with the Clustal X color scheme.

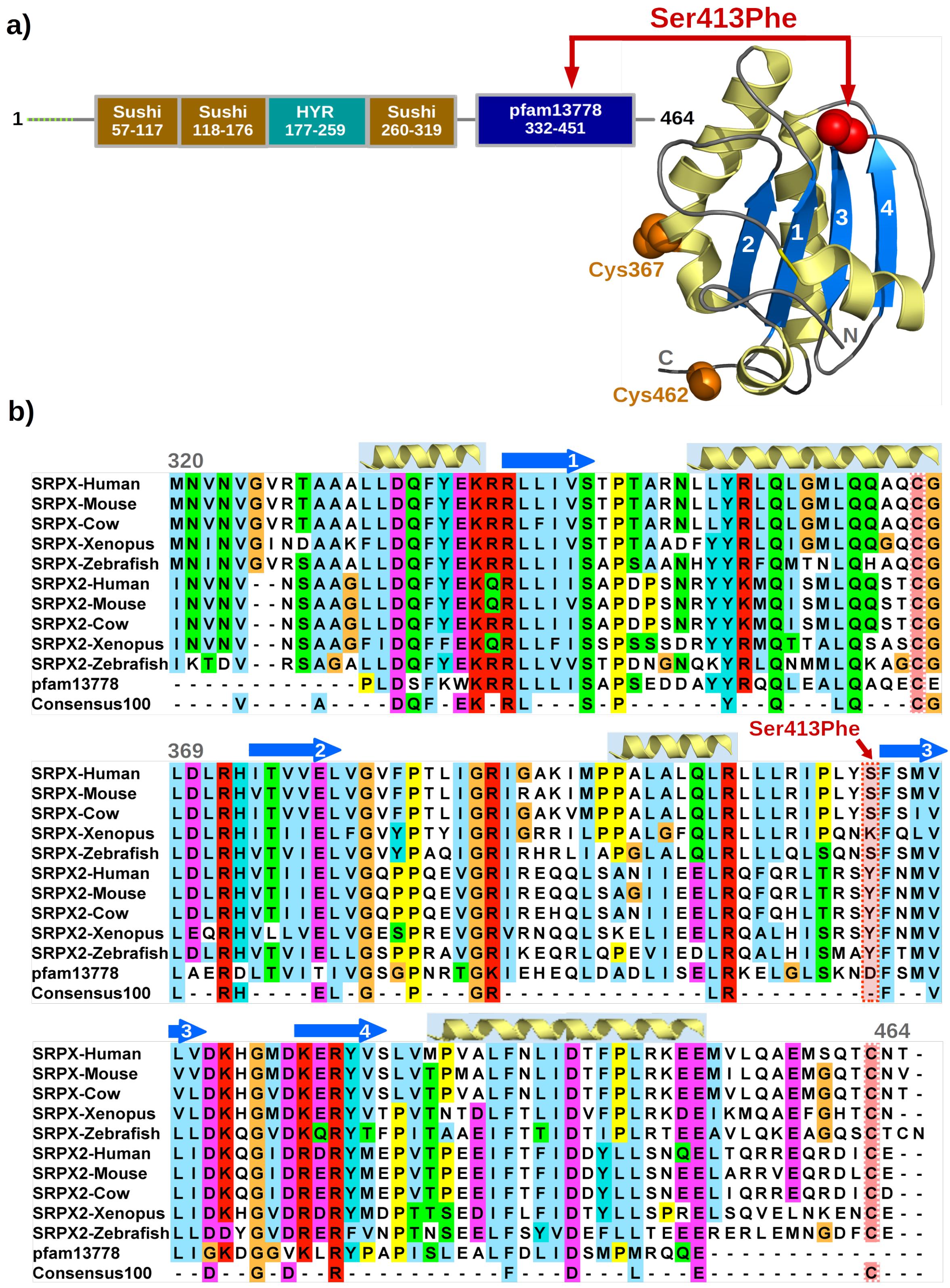

a)
Ser413Phe
1
Sushi 57-117
Sushi 118-176
HYR 177-259
Sushi 260-319
pfam13778 332-451
464
1
2
3
4
Cys367
N
C
Cys462
b)
320
SRPX-Human MNVNVGVRTAAALLDQFYEKRRLLIVSTPTARNLLYRLQLGMLQQAQCG
SRPX-Mouse MNVNVGVRTAAALLDQFYEKRRLLIVSTPTARNLLYRLQLGMLQQAQCG
SRPX-Cow MNVNVGVRTAAALLDQFYEKRRLFIVSTPTARNLLYRLQLGMLQQAQCG
SRPX-Xenopus MNINVGINDAAKFLDQFYEKRRLLIVSTPTAADFYYRLQIGMLQQGQCG
SRPX-Zebrafish MNINVGVRSAAALLDQFYEKRRLLIISAPSAANHYYRFQMTNLQHAQCG
SRPX2-Human INVNV--NSAAGLLDQFYEKQRLLIISAPDPSNRYYKMQISMLQQSTCG
SRPX2-Mouse INVNV--NSAAGLLDQFYEKQRLLIVSAPDPSNRYYKMQISMLQQSTCG
SRPX2-Cow INVNV--NSAAGLLDQFYEKRRLLIISAPDPSNRYYKMQISMLQQSTCG
SRPX2-Xenopus INVNV--NSAAGFIDQFFEKQRLLFISSPSSSDRYYRMQTTALQSASCG
SRPX2-Zebrafish IKTDV--RSAGALLDQFYEKRRLLVVSTPDNGNQKYRLQNMMLQKAGCG
pfam13778 ------------PLDSFKWKRRLLLISAPSEDDAYYRQQLEALQAQECE
Consensus100 ----V----A----DQF-EK-RL---S-P------Y--Q---LQ---CG
369
Ser413Phe
SRPX-Human LDLRHITVVELVGVFPTLIGRIGAKIMPPALALQLRLLLRIPLYSFSMV
SRPX-Mouse LDLRHVTVVELVGVFPTLIGRIRAKIMPPALALQLRLLLRIPLYSFSMV
SRPX-Cow LDLRHITVVELVGVFPTLIGRIGAKVMPPALALQLRLLLRIPLYSFSIV
SRPX-Xenopus LDLRHITIIELFGVYPTYIGRIGRRILPPALGFQLRLLLRIPQNKFQLV
SRPX-Zebrafish LDLRHVTVIELVGVYPAQIGRIRHRLIAPGLALQLRLLLQLSQNSFSMV
SRPX2-Human LDLRHVTIIELVGQPPQEVGRIREQQLSANIIEELRQFQRLTRSYFNMV
SRPX2-Mouse LDLRHVTIIELVGQPPQEVGRIREQQLSAGIIEELRQFQRLTRSYFNMV
SRPX2-Cow LDLRHVTIIELVGQPPQEVGRIREHQLSANIIEELRQFQHLTRSYFNMV
SRPX2-Xenopus LEQRHVLLVELVGESPREVGRVRNQQLSKELIEELRQALHISRSYFNMV
SRPX2-Zebrafish LDLRHVTVIELLGSPPRAVGRIKEQRLQPEVIEDLRQALHISMAYFTMV
pfam13778 LAERDLTVITIVGSGPNRTGKIEHEQLDADLISELRKELGLSKNDFSMV
Consensus100 L--RH----EL-G--P---GR-------------LR---------F--V
464
SRPX-Human LVDKHGMDKERYVSLVMPVALFNLIDTFPLRKEEMVLQAEMSQTCNT-
SRPX-Mouse VVDKHGMDKERYVSLVTPMALFNLIDTFPLRKEEMILQAEMGQTCNV-
SRPX-Cow VLDKHGVDKERYVSLVTPVALFNLIDTFPLRKEEMVLQAEMGQTCNT-
SRPX-Xenopus VLDKHGMDKERYVTPVTNTDLFTLIDVFPLRKDEIKMQAEFGHTCN--
SRPX-Zebrafish LLDKQGVDKQRYTFPITAAEIFTTIDTIPLRTEEAVLQKEAGQSCTCN
SRPX2-Human LIDKQGIDRDRYMEPVTPEEIFTFIDDYLLSNQELTQRREQRDICE--
SRPX2-Mouse LIDKQGIDRERYMEPVTPEEIFTFIDDYLLSNEELARRVEQRDLCE--
SRPX2-Cow LIDKQGIDRERYMEPVTPEEIFTFIDDYLLSNEELIQRREQRDICD--
SRPX2-Xenopus LIDKHGVDRDRYMDPTTSEDIFLFIDTYLLSPRELSQVELNKENCE--
SRPX2-Zebrafish LLDDYGVDRERFVNPTNSEELFSYVDEFLLTEEERERLEMNGDFCE--
pfam13778 LIGKDGGVKLRYPAPISLEALFDLIDSMPMRQQE--------------
Consensus100 --D--G-D--R-----------F---D---L---E-----------C---

**Supplementary Figure 7.** *Sushi repeat-containing protein (SRPX) structure und alignment.*

**(a)** SRPX domain structure and the predicted protein fold of the C-terminal domain. The N-terminal signal peptide is shown as a green dashed line. Predicting the 3D location of the Ser413Phe variant is based on a model with lower confidence, with loop and helical structures being less reliable than the central beta sheet. In this model it is predicted that the polar Ser413 stabilizes loops originating from strands 1, 3 and 4, and a mutation to a hydrophobic phenylalanine could interfere with this function **(b)** Multiple sequence alignment with predicted secondary structures. Conserved sequence positions are largely consistent with the pfam13778 family, in particular with the central beta sheet, which enhances the confidence of the core regions in the above structural model. Ser413Phe is located adjacent to the conserved beta strand 3 and the invariant Phe414 which supports an important structural role of the variant. For further details see **Supplementary Note**, section ***In silico* variant protein analysis**.

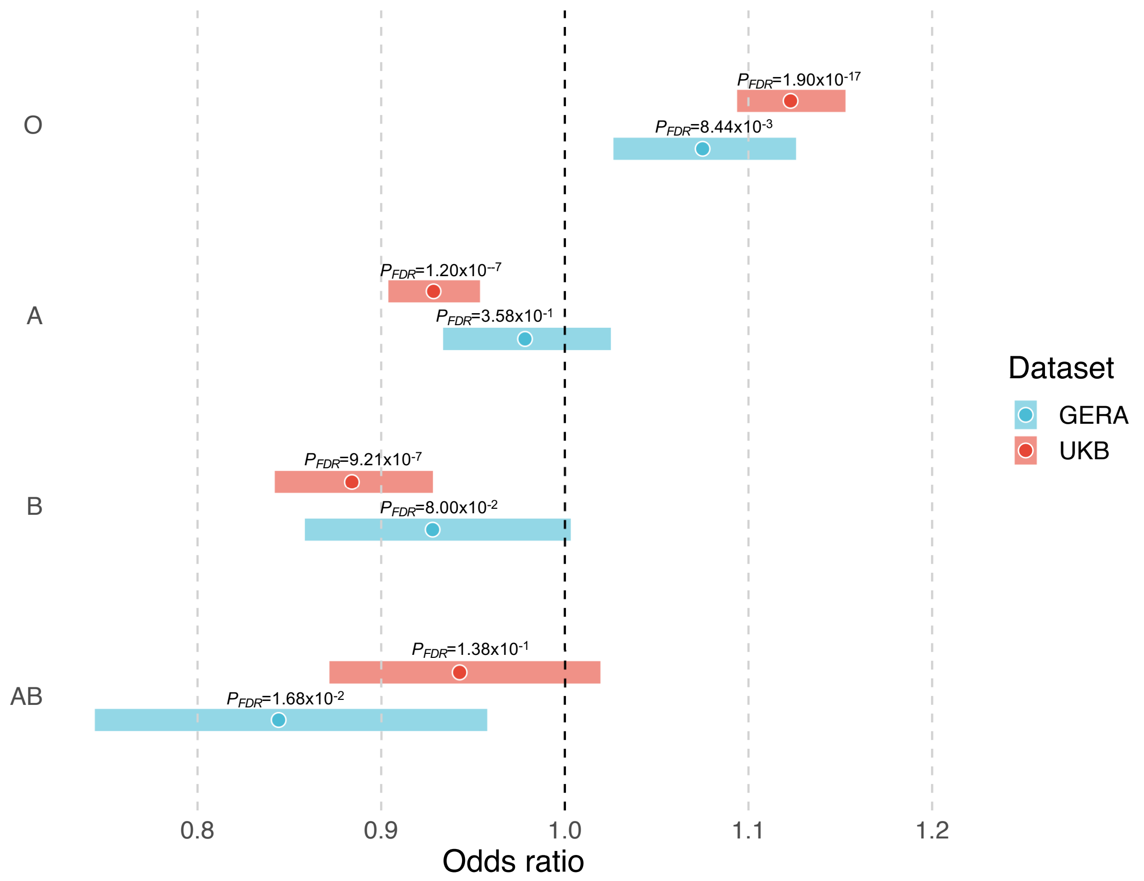

**Supplementary Figure 8.** *ABO blood groups and HEM risk in UKBB and GERA.*
The plot shows odds ratios (and 95% confidence intervals) form testing ABO blood groups vs HEM risk in UKBB and GERA (see **Methods**). An association test based on logistic regression is used to test for a significant HEM association for each of the four blood groups, taking into account sex, age, BMI and the top 10 PCs from PCA. FDR correction was applied to correct for multiple testing. FDR: false discovery rate.

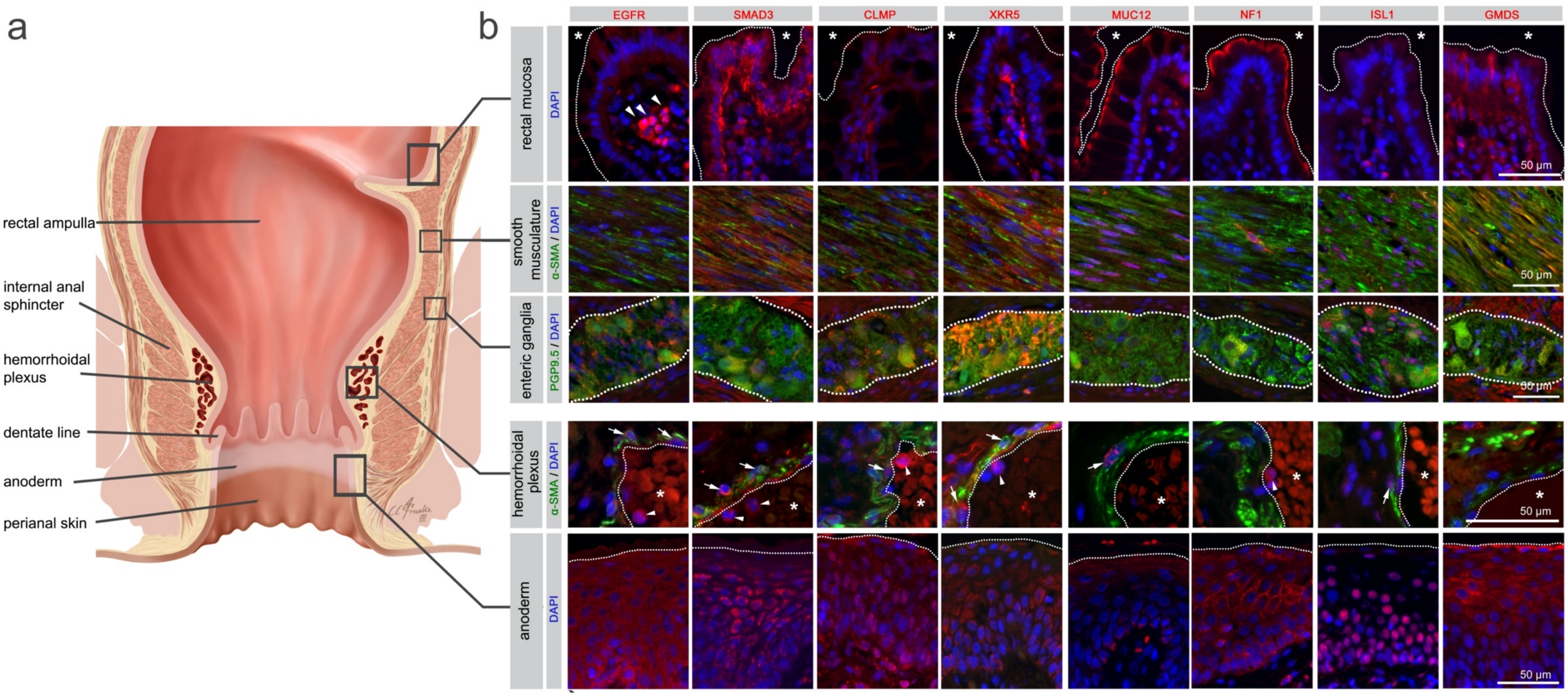
a
b
rectal ampulla
internal anal sphincter
hemorrhoidal plexus
dentate line
anoderm
perianal skin
EGFR
SMAD3
CLMP
XKR5
MUC12
NF1
ISL1
GMDS
rectal mucosa
DAPI
smooth musculature
α-SMA / DAPI
enteric ganglia
PGP9.5 / DAPI
hemorrhoidal plexus
α-SMA / DAPI
anoderm
DAPI
50 µm

**Supplementary Figure 9.** *Immunohistochemistry for selected HEM candidate proteins.* Illustration of the rectum and anal canal **(a)** with indication the site-specific localization of the immunohistochemical panels analyzed in **(b).** Fluorescence immunohistochemistry (B) for selected HEM candidate proteins (see also **Supplementary Table 11**), encoded by candidate genes within our 102 identified genome-wide significant loci, are shown. *SRPX* (rs35318931), *ANO1* (rs2186797) and *MYH11* (rs6498573) were determined as prioritized HEM genes in our study. *ANO1* and *SRPX* are interesting HEM candidate genes since the lead SNPs at these loci are (missense) coding variants. *MYH11* is also a main hub gene within the M1 co-expression module of our transcriptome analysis. Given the ABO blood group association observed in our study in HEM patients (Supplementary Figure 9), we have included ABO as further target for immunohistochemistry.

Antibody staining was performed on anorectal FFPE tissue specimens from control individuals. The rows correspond to the rectal mucosa (top row, epithelial surface delimited by dashed line,*: intestinal lumen), smooth musculature (second row), enteric ganglia (third row, ganglionic boundaries delimited by dashed line), hemorrhoidal plexus (fourth row, endothelial surface delimited by dashed line, *: vascular lumen), and the anoderm (bottom row, border of the anoderm delimited by dashed line). Blue: DAPI; green: α-SMA (anti-alpha smooth muscle antibody) for row 2 and 4 (smooth musculature/hemorrhoidal plexus) and PGP9.5 (member of the ubiquitin hydrolase family of proteins, neuronal marker) for row 3 (enteric ganglia); red: antibody for the respective candidate protein. Arrows point to corresponding candidate-positive cells within the vascular wall. Arrowheads point to corresponding candidate-positive nucleated immune cells.

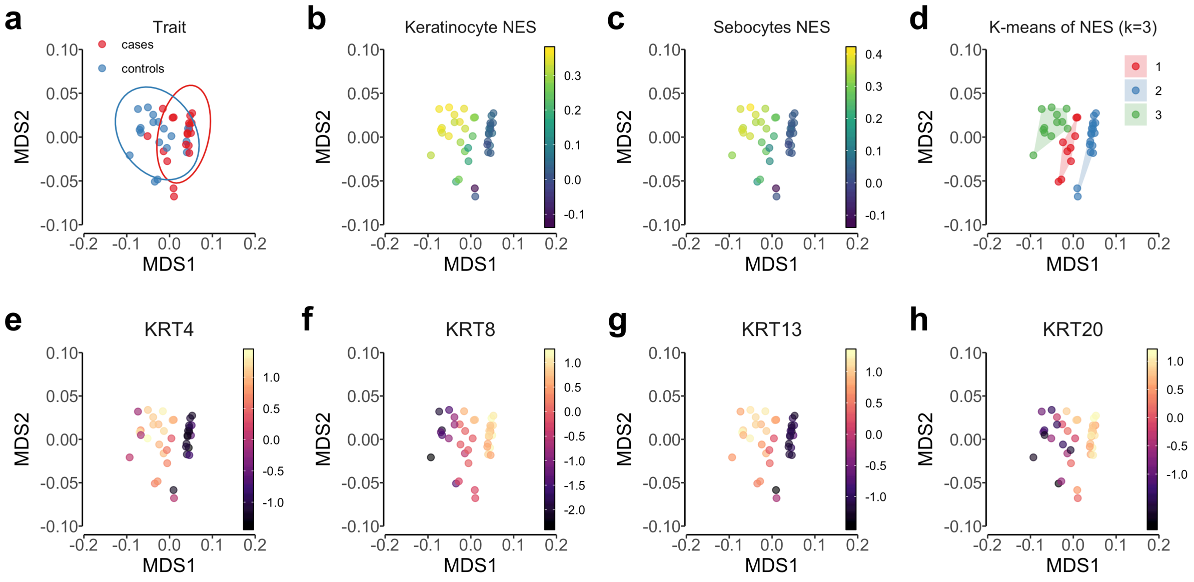

**Supplementary Figure 10.** *Gene signature-based determination of anal canal zones.* Multidimensional scaling (MDS) analysis of the transcriptome data using Spearman's correlation distance (1 – correlation coefficient); **(a)** Colored by trait status, where the cases are enlarged hemorrhoidal tissue samples and controls are healthy hemorrhoidal tissue; **(b)** Colored by normalized enrichment score (NES) of keratinocyte cells; **(c)** Colored by NES of sebocytes; **(d)** Colored by clusters obtained by applying the k-means algorithm; **(e-h)** Colored by normalized expression values of anal canal marker genes, including *KRT4*, *KRT8*, *KRT13* and *KRT20*.

#### Supplementary Tables

**Supplementary Table 1.** *Study cohorts included the individual GWAS, meta-analyses, and follow up.*

SEE EXCEL FILE

For cohort description, see **Methods**. For details on quality control, see **Supplementary Note**. **Cohort:** shorthand of the case-control panel; **Phenotype:** HEM, Hemorrhoidal disease cases, **CTRL**, control; **P**: *P*-value from student's t-test comparing age and BMI, and from chi-square test comparing sex between hemorrhoidal disease cases and controls in each individual cohort; **Age**, mean (±SD): mean age of onset/age of sampling and standard deviation; **Sex**, female %: percentage of females; **BMI**, mean (±SD): mean of body mass index. SD: standard deviation. UKBB: UK Biobank, EGCUT: Estonian Genome Center at the University of Tartu, MGI: Michigan Genomics Initiative, GERA: Genetic Epidemiology Research on Aging, HUNT: The Trøndelag Health Study, DBDS: Danish Blood Donor Study, DNPR: Danish National Patient Registry. * Age is calculated from birthday to follow-up date (either death, emigration or end of follow-up, which ever one came first).

**Supplementary Table 2.** *HEM GWAS risk loci.*

SEE EXCEL FILE

102 newly identified genetic susceptibility loci associated with HEM at genome-wide significance ($P_{Meta}<5\times10^{-8}$). **Loci number**: number of susceptibility loci; **CHR**: chromosome; **start-end**: left/right association boundaries for each lead SNP defined by FUMA (see **Supplementary Note**). Genomic positions were retrieved from NCBI's dbSNP build v150 (genome build hg19); **Lead SNP (rsID)**: rs ID retrieved from NCBI's dbSNP build v150; **BP**: base pair position; **A1**: minor allele; **A2:** major allele; **EAF:** effect allele (i.e. minor allele) frequency; **OR (95% CI):** odds ratio (OR) and 95% confidence interval (CI 95%) with respect to A1; **P**: *P*-value; **Nearest Gene**: Nearest gene refers to the nearest protein-coding mapped gene within 100kb of the lead SNP, the number of additional mapped gene within each locus is given in brackets (see also **Supplementary Table 7**). "na" means that there was no protein-coding mapped gene within 100Kb of the lead SNP; **Association directions in individual cohorts**: Directions of the effect allele in five individual discovery datasets (being shown in order from 23andMe, UKBB, EGCUT, MGI to GERA). "+" represent risk effects, "-" represents protective effect and "?" represent that the tested lead SNP is not available in the association results of the individual cohort.

**Supplementary Table 3.** *Number of fine-mapped variants in 95% credible sets.*

SEE EXCEL FILE

Variants in 95% fine-mapped credible sets for 102 susceptibility loci associated with HEM at genome-wide significance ($P_{Meta}$<5×10-8). For 6 loci, the 95% credible set consisted of a single variant ('single variant credible sets'), and for 96 others the credible set consisted of multiple variant. **Locus**: number of susceptibility locus according to **Supplementary Table 2**. **SNP**: variants in the 95% credible sets (see **Methods**). Variants were sorted by the posterior probability of association, and variants were added to the 'credible set' of associated variants until the sum of their posterior probability exceeded 95%. Rs-numbers retrieved from NCBI's dbSNP build v150 (genome build hg19); **CHR**: chromosome; **BP**: base pair position; **Posterior probability**: posterior probability of causality to each SNP variant.

**Supplementary Table 4.** *Association of HEM risk variants with other traits and diseases.*

SEE EXCEL FILE

This table contains other reported GWAS associations of 102 lead SNPs (and/or their $r^2>0.8$ LD proxies) of HEM for other traits, extracted from three PheWAS databases: PhenoScanner[62], GWAS catalog[63] and GWAS ATLAS[64]. **Lead SNP (rsID)**: rs ID retrieved from NCBI's dbSNP build v150; **Lead SNP BP**: base pair position of the lead SNP (in "chromosome:base pair" format), genomic positions were retrieved from NCBI's dbSNP build v150 (genome build hg19); **Proxy SNP (rsID):** rs ID of a SNP in LD ($r^2>0.8$) with the lead SNP and has a reported GWAS association with other traits; **Proxy SNP BP:** base pair position of the proxy SNP (in "chromosome:base pair" format); **$r^2$**: the square of the correlation coefficient**; Associated trait:** the name of thar trait that has a reported GWAS association with the lead SNP or its proxy SNP**; Domain:** the assigned trait category of the trait (only available for traits from GWAS ATLAS); **PheWAS database:** the database where the reported association is extracted from; **Data source or PUBMEDID:** the publication reference or data source (UKBB, http://www.nealelab.is/uk-biobank) of the reported GWAS association.

**Supplementary Table 5.** *Results of subset-based pleiotropy meta-analysis (SBM).*

SEE EXCEL FILE

Summary statistics of 44 independent genomic regions shared by at least two phenotypes from subset-based pleiotropy meta-analysis (SBM) of 7,251,618 SNPs on the phenotypes diverticular disease (DIV), irritable bowel syndrome (IBS) and hemorrhoidal disease (HEM) with a maximum of 1,741,260 samples and adjusted for sample overlap. Lines in bold type are novel loci not overlapping the 102 HEM risk loci of **Supplementary Table 2**. **Lead SNP (rsID):** rs ID retrieved from NCBI's dbSNP build v150; **CHR:** chromosome; **BP:** base pair position; **start-end**: left/right association boundaries for each lead SNP defined by FUMA (see **Supplementary Note**); **Phenotype(s):** subset of phenotypes from SBM with genome-wide significance ($P_{SBM}$<5×10$^{-8}$); **P**: Final *P*-value ($P_{SBM}$) from two-sided statistic for the detection of potential effects in opposite directions (combined from two-sided statistics *P* subset1 and *P* subset 2). i.e. adjusted (disease-combined) *P*-value ($P_{\mathrm{SBM}}$) from SBM (see **Methods**); **Phenotype(s) subset 1:** risk (OR>1) disease subset from SBM; **P subset 1:** *P*-value from two-sided statistic across phenotype(s) of subset 1; **OR (95% CI) subset 1:** odds ratio and corresponding 95% confidence interval of phenotype(s) from subset 1; **Phenotype(s) subset 2:** protective (OR<1) disease subset from SBM; **P subset 2:** *P*-value from two-sided statistic across phenotype(s) of subset 2; **OR (95% CI) subset 2:** odds ratio and corresponding 95% confidence interval of phenotype(s) from subset 2.

**Supplementary Table 6.** *Significant genetic correlations between HEM and other traits estimated by genome-wide LD Score Regression (LDSC).*

SEE EXCEL FILE

**Trait:** the name of the complex trait or disease being compared to HEM for genetic correlation; **rg:** genetic correlation between HEM and each trait, ranging from -1 to 1. rg>0 represents a positive and rg<0 represents a negative genetic correlation; **se:** standard error of rg; **z score:** the z-score from statistical test of genetic correlation; **P FDR:** the z-score from statistical test of genetic correlation after FDR correction for multiple testing at $\alpha$=0.05; **N total:** the sample size being included in the genetic analysis of each trait; **Cohort:** the data source from which the summary statistic of the trait were extracted.

**Supplementary Table 7.** *Summary of HEM genes mapping and prioritization.*

SEE EXCEL FILE

This table contains detailed information of all 819 HEM mapped genes and the criteria for gene prioritization (see **Results** and **Methods). Gene symbol:** gene ID extracted from HUGO Gene Nomenclature Committee[65]; **EntrezID:** gene ID extracted from Entrez Gene database[66]; **EnsemblID:** gene ID extracted from ensembl database GRCh38.p13[67]; **Gene type:** the biotype of the gene annotated by ensembl database GRCh38.p13[67]; **Locus number:** the sequential number of the HEM HWAS loci; **Lead SNP (rsID):** rs ID retrieved from NCBI's dbSNP build v150**; Prioritized HEM genes**: a binary variable indicating whether the gene is a prioritized HEM gene (1=yes, 0=no); **Gene mapping**: specifying the gene mapping method for each gene among four mapping categories (FUMA_positional, FUMA_eQTL, MAGMA, DEPICT, 1=yes, 0=no, see **Methods**); **DEPICT prioritized:** indicating whether a gene is prioritized by DEPICT gene prioritization algorithm[68] (1=yes, 0=no); **Linked to fine mapped variant (probabliity>50%):** Genes that are linked to a fine mapped variant (posterior probability > 50%) by either significant eQTL associations only (labelled "eQTL") or significant eQTL associations plus physically containing the fine mapped coding variant (labelled "coding variant & eQTL"). The rs ID of the linked fine mapped variant and its posterior probability are shown in the brackets. **Expression in HEM tissue:** Genes that were expressed (labelled "E") or not expressed (labelled "NE") from RNAseq results of HEM tissues. Differentially expressed genes were highlighted with either "E+" (for overexpressed genes) or "E-" (for underexpressed genes) according to the RNAseq analysis results comparing tissue expression profiles between HEM patients and healthy controls. Genes were labelled with "(UQ)" if were highly expressed (in upper quartile). **WGCNA module**: indicating whether a gene is included in any of the significant gene co-expression modules of HEM tissue (M1, M4 or M7, see **Methods**), a suffix "_hub" after the module names (e.g. M1_hub) represents a hub gene within the gene co-expression module. **Mouse phenotypes from MGI**: type of mammalian phenotype categories that are associated with the gene mutations from Mouse Genome Informatics Database (MGI, http://www.informatics.jax.org/)[69]. **OMIM Phenotypes:** a list of genetic disorders that are associated with the gene mutations, information extracted from Online Mendelian Inheritance in Man (OMIM)[70]. **"-"**: data is not available.

**Supplementary Table 8.** *Gene set and tissue enrichment analyses of HEM genes.*

SEE EXCEL FILE

This table contains all tissues and pathways if they are significantly enriched in at least one out of three analytic tools (FUMA[71], MAGMA[72] or DEPICT[68]; **Methods**). **Genesets**: The enriched pathway or tissue. Gene-sets were only reported if their FDR corrected *P*-values were lower than 0.05 in at least one of the three methods. **Category**: GTEx tissue: 30 GTEx general tissue types using data from GTEx release v7 database)[73], GO_BP: gene ontology biological process, GP_MF: gene ontology molecular function and GO_CC: gene ontology cellular component from Molecular Signature Database[74]; **P.FDR.FUMA:** the *P*-value from the statistical test of enrichment in FUMA after FDR correction for multiple comparisons at $\alpha$=0.05; **P.FDR.MAGMA:** the *P*-value from the statistical test of enrichment in MAGMA after FDR correction for multiple comparisons at $\alpha$=0.05; **P.FDR.DEPICT:** the *P*-value from the statistical test of enrichment in DEPICT after FDR correction for multiple comparisons at $\alpha$=0.05. **"-"**: data is not available.

**Supplementary Table 9.** *HEM gene overrepresentation analysis in gene co-expression network modules of hemorrhoidal tissue.*

SEE EXCEL FILE

The overrepresentation analysis was performed using Fisher's exact test with the alternative hypothesis that the true odds ratio is greater than one. **Module:** network module of co-expressed genes in hemorrhoids tissue; **N HEM genes:** number of HEM candidate genes in module; **N genes:** total number of genes in module; **HEM gene frac, %:** percentage of HEM candidate genes in a given module; **OR:** odds ratio; **P-value:** nominal *P*-value of Fisher's exact test; **FDR:** *P*-value after FDR correction.

**Supplementary Table 10.** *Significant eQTL associations of fine-mapped variants and HEM genes.*

SEE EXCEL FILE

This table contains a list of significant eQTL associations (FDR<0.05) with details for fine-mapped variants (posterior probability>50%, see **Methods** and **Supplementary Table 3**). **Fine mapped SNP (rsID):** rs ID of the fine-mapped variants retrieved from NCBI's dbSNP build v150**; Position:** base pair position of the lead SNP (in "chromosome:base pair" format), genomic positions were retrieved from NCBI's dbSNP build v150 (genome build hg19); **Posterior probability:** posterior probability of causality to each SNP variant; **SNP Type:** functional consequence of the SNP obtained from ANNOVAR[75]; **eQTL database:** Data source of eQTLs from the 10 repositories (see **Methods**)**; tissue:** tissue type of the eQTL association**; EnsemblID:** gene ID extracted from ensembl database GRCh38.p13 (www.ensembl.org/); **Gene symbol:** gene ID extracted from HUGO Gene Nomenclature Committee[65]; **P.FDR:** *P*-value of eQTLs after FDR correction for multiple comparisons at $\alpha$=0.05.

**Supplementary Table 11.** *Summary of protein expression shown in **Figure 7** and **Supplementary Figure 9.***

Immunohistochemistry was performed on three independent patients and three control samples for all 12 selected candidate proteins. No obvious differences in immunofluorescent signal intensity were observed between cases and controls for any of the candidate proteins (data not shown), however, the small sample size must be considered. A semi-quantitative grading system was used to evaluate the intensity of immunofluorescent signals: ++: strong signal; +: moderate signal; ± low signal; -: no signal. ICC, interstitial cells of Cajal.

| | **anoderm** | **intestinal mucosa** | **hemorrhoidal plexus** | **immune cells** | **smooth musculature** | **enteric ganglia** |
|---|---|---|---|---|---|---|
| ***EGFR*** | ++ | ± | ± | + | ± | ± |
| ***SMAD3*** | + | ++ | + | + | + | + |
| ***CLMP*** | + | - | + | ++ | + | ± |
| ***XKR5*** | ++ | + | + | + | + | ++ |
| ***ABO*** | ++ | + | + | - | + | ± |
| ***SRPX*** | + | + | + | ++ | + | ± |
| ***MUC12*** | - | ++ | + | - | + | - |
| ***ANO1*** | - | ± | + | - | + (also ICC) | ± |
| ***NF1*** | + | ++ | - | + | + (also ICC) | - |
| ***ISL1*** | ++ | - | + | - | + | + |
| ***GMDS*** | + | + | ± | - | + | ± |
| ***MYH11*** | ± | - | ± | ± | + | - |

**Supplementary Table 12:** *List of antibodies used in the immunohistochemistry analyses shown in* ***Figure 7*** *and* ***Supplementary Figure 9.***

| Antibody Name | Dilution | Species | Vendor | Order Number/ID | Lot |
|---|---|---|---|---|---|
| EGFr | 1/200 | rabbit | Abcam | ab52894 | GR3214138-6 |
| Smad3 | 1/200 | rabbit | Abcam | Ab40854 | GR3190602-17 |
| CLMP | 1/200 | rabbit | sigma | HPA002385 | A35819 |
| XKR5 | 1/200 | rabbit | Thermoscientif. | PA5-71286 | UJ2852378 |
| ABO | 1/200 | rabbit | Thermoscientif. | PA5-37352 | TB2515446 |
| SRPX | 1/200 | rabbit | Abcam | ab206836 | GR241480-6 |
| MUC12 | 1/200 | rabbit | sigma | HPA023835 | 000003770 |
| ANO1 | 1/200 | rabbit | Abcam | Ab53212 | GR3295356-1 |
| NF1 | 1/200 | rabbit | Abcam | ab128054 | GR86480-17 |
| ISL1 | 1/200 | rabbit | Abcam | ab109517 | GR290344-19 |
| GMDS | 1/200 | mouse | Abcam | Ab128046 | GR3334809-1 |
| MYh11 | 1/200 | mouse | Abcam | ab683 | GR3317192-1 |
| PGP9.5 | 1/500 | rabbit | Ultraclone | RA95101 | J1412 |
| PGP9.5 | 1/500 | mouse | OriGene | UM870136 | F001 |
| a-SMA | 1/1000 | rabbit | Abcam | Ab5694 | 825197 |
| a-SMA | 1/1000 | mouse | Dako | M0851 clone 1a4 | 32 |
| Alexafluor 555 Donkey-anti-rabbit | 1/700 | Donkey | Thermoscientif. | A31572 | 1454443 |
| Alexafluor 488 Donkey anti-mouse | 1/700 | Donkey | Thermoscientif. | A21202 | 1305303 |
| Alexafluor 488 Goat anti-rabbit | 1/700 | goat | Thermoscientif. | A31628 | 899848 |
| Alexafluor 555 Donkey anti-mouse | 1/700 | Donkey | Thermoscientif. | A31570 | 1048568 |
| XKR9 | did not work | rabbit | Sigma | HPA044430 | R40675 |
| GSDMC | did not work | rabbit | Abcam | Ab238613 | GR3294276-2 |
| AChE | did not work on paraffin sections | mouse | Abcam | Ab2803 | GR3277114-6 |

#### References

1. Durand, E.Y., Do, C.B., Mountain, J.L. & Macpherson, J.M. Ancestry Composition: A Novel, Efficient Pipeline for Ancestry Deconvolution. *bioRxiv*, 010512 (2014).
2. Henn, B.M. *et al.* Cryptic distant relatives are common in both isolated and cosmopolitan genetic samples. *PLoS One* **7**, e34267 (2012).
3. Genomes Project, C. *et al.* A map of human genome variation from population-scale sequencing. *Nature* **467**, 1061-73 (2010).
4. Browning, S.R. & Browning, B.L. Rapid and accurate haplotype phasing and missing-data inference for whole-genome association studies by use of localized haplotype clustering. *Am J Hum Genet* **81**, 1084-97 (2007).
5. Fuchsberger, C., Abecasis, G.R. & Hinds, D.A. minimac2: faster genotype imputation. *Bioinformatics* **31**, 782-4 (2015).
6. Wain, L.V. *et al.* Novel insights into the genetics of smoking behaviour, lung function, and chronic obstructive pulmonary disease (UK BiLEVE): a genetic association study in UK Biobank. *Lancet Respir Med* **3**, 769-81 (2015).
7. Bycroft, C. *et al.* The UK Biobank resource with deep phenotyping and genomic data. *Nature* **562**, 203-209 (2018).
8. Abraham, G. & Inouye, M. Fast principal component analysis of large-scale genome-wide data. *PLoS One* **9**, e93766 (2014).
9. International HapMap, C. *et al.* A second generation human haplotype map of over 3.1 million SNPs. *Nature* **449**, 851-61 (2007).
10. Delaneau, O., Marchini, J. & Zagury, J.F. A linear complexity phasing method for thousands of genomes. *Nat Methods* **9**, 179-81 (2011).
11. Mitt, M. *et al.* Improved imputation accuracy of rare and low-frequency variants using population-specific high-coverage WGS-based imputation reference panel. *Eur J Hum Genet* **25**, 869-876 (2017).
12. Howie, B.N., Donnelly, P. & Marchini, J. A flexible and accurate genotype imputation method for the next generation of genome-wide association studies. *PLoS Genet* **5**, e1000529 (2009).
13. Jun, G. *et al.* Detecting and estimating contamination of human DNA samples in sequencing and array-based genotype data. *Am J Hum Genet* **91**, 839-48 (2012).
14. Manichaikul, A. *et al.* Robust relationship inference in genome-wide association studies. *Bioinformatics* **26**, 2867-73 (2010).
15. Li, J.Z. *et al.* Worldwide human relationships inferred from genome-wide patterns of variation. *Science* **319**, 1100-4 (2008).
16. Chang, C.C. *et al.* Second-generation PLINK: rising to the challenge of larger and richer datasets. *Gigascience* **4**, 7 (2015).
17. McCarthy, S. *et al.* A reference panel of 64,976 haplotypes for genotype imputation. *Nat Genet* **48**, 1279-83 (2016).
18. Patterson, N., Price, A.L. & Reich, D. Population structure and eigenanalysis. *PLoS Genet* **2**, e190 (2006).
19. Howie, B., Fuchsberger, C., Stephens, M., Marchini, J. & Abecasis, G.R. Fast and accurate genotype imputation in genome-wide association studies through pre-phasing. *Nat Genet* **44**, 955-9 (2012).
20. Price, A.L. *et al.* Long-range LD can confound genome scans in admixed populations. *Am J Hum Genet* **83**, 132-5; author reply 135-9 (2008).
21. Jun, T.H., Rouf Mian, M.A. & Michel, A.P. Genetic mapping revealed two loci for soybean aphid resistance in PI 567301B. *Theor Appl Genet* **124**, 13-22 (2012).

22. Guo, Y. *et al.* Illumina human exome genotyping array clustering and quality control. *Nat Protoc* **9**, 2643-62 (2014).
23. Wang, C., Zhan, X., Liang, L., Abecasis, G.R. & Lin, X. Improved ancestry estimation for both genotyping and sequencing data using projection procrustes analysis and genotype imputation. *Am J Hum Genet* **96**, 926-37 (2015).
24. Loh, P.R., Palamara, P.F. & Price, A.L. Fast and accurate long-range phasing in a UK Biobank cohort. *Nat Genet* **48**, 811-6 (2016).
25. Das, S. *et al.* Next-generation genotype imputation service and methods. *Nat Genet* **48**, 1284-1287 (2016).
26. Hansen, T.F. *et al.* DBDS Genomic Cohort, a prospective and comprehensive resource for integrative and temporal analysis of genetic, environmental and lifestyle factors affecting health of blood donors. *BMJ Open* **9**, e028401 (2019).
27. Konig, I.R., Loley, C., Erdmann, J. & Ziegler, A. How to include chromosome X in your genome-wide association study. *Genet Epidemiol* **38**, 97-103 (2014).
28. Loh, P.R. *et al.* Efficient Bayesian mixed-model analysis increases association power in large cohorts. *Nat Genet* **47**, 284-90 (2015).
29. Zhou, W. *et al.* Efficiently controlling for case-control imbalance and sample relatedness in large-scale genetic association studies. *Nat Genet* **50**, 1335-1341 (2018).
30. Fenger, C. The anal transitional zone. *Acta Pathol Microbiol Immunol Scand Suppl* **289**, 1-42 (1987).
31. Iacobuzio-Donahue, C.A. Inflammatory and Neoplastic Disorders of the Anal Canal. in *Surgical Pathology of the GI Tract, Liver, Biliary Tract, and Pancreas* (ed. Robert Odze, J.G.) 733-761 (Elsevier Inc., 2009).
32. Aran, D., Hu, Z. & Butte, A.J. xCell: digitally portraying the tissue cellular heterogeneity landscape. *Genome Biol* **18**, 220 (2017).
33. Love, M.I., Huber, W. & Anders, S. Moderated estimation of fold change and dispersion for RNA-seq data with DESeq2. *Genome Biol* **15**, 550 (2014).
34. Barbie, D.A. *et al.* Systematic RNA interference reveals that oncogenic KRAS-driven cancers require TBK1. *Nature* **462**, 108-12 (2009).
35. Hanzelmann, S., Castelo, R. & Guinney, J. GSVA: gene set variation analysis for microarray and RNA-seq data. *BMC Bioinformatics* **14**, 7 (2013).
36. Williams, G.R., Talbot, I.C., Northover, J.M. & Leigh, I.M. Keratin expression in the normal anal canal. *Histopathology* **26**, 39-44 (1995).
37. De Jesus-Perez, J.J. *et al.* Phosphatidylinositol 4,5-bisphosphate, cholesterol, and fatty acids modulate the calcium-activated chloride channel TMEM16A (ANO1). *Biochim Biophys Acta Mol Cell Biol Lipids* **1863**, 299-312 (2018).
38. Le, S.C., Jia, Z., Chen, J. & Yang, H. Molecular basis of PIP2-dependent regulation of the Ca(2+)-activated chloride channel TMEM16A. *Nat Commun* **10**, 3769 (2019).
39. Paulino, C., Kalienkova, V., Lam, A.K.M., Neldner, Y. & Dutzler, R. Activation mechanism of the calcium-activated chloride channel TMEM16A revealed by cryo-EM. *Nature* **552**, 421-425 (2017).
40. Dang, S. *et al.* Cryo-EM structures of the TMEM16A calcium-activated chloride channel. *Nature* **552**, 426-429 (2017).
41. Kirkitadze, M.D. & Barlow, P.N. Structure and flexibility of the multiple domain proteins that regulate complement activation. *Immunol Rev* **180**, 146-61 (2001).
42. Callebaut, I., Gilges, D., Vigon, I. & Mornon, J.P. HYR, an extracellular module involved in cellular adhesion and related to the immunoglobulin-like fold. *Protein Sci* **9**, 1382-90 (2000).
43. Bommer, G.T. *et al.* DRO1, a gene down-regulated by oncogenes, mediates growth inhibition in colon and pancreatic cancer cells. *J Biol Chem* **280**, 7962-75 (2005).

44. Pawlowski, K. *et al.* A widespread peroxiredoxin-like domain present in tumor suppression- and progression-implicated proteins. *BMC Genomics* **11**, 590 (2010).
45. Inoue, H., Pan, J. & Hakura, A. Suppression of v-src transformation by the drs gene. *J Virol* **72**, 2532-7 (1998).
46. Tambe, Y. *et al.* Tumor prone phenotype of mice deficient in a novel apoptosis-inducing gene, drs. *Carcinogenesis* **28**, 777-84 (2007).
47. Tambe, Y., Hasebe, M., Kim, C.J., Yamamoto, A. & Inoue, H. The drs tumor suppressor regulates glucose metabolism via lactate dehydrogenase-B. *Mol Carcinog* **55**, 52-63 (2016).
48. Burgstaller, G. *et al.* The instructive extracellular matrix of the lung: basic composition and alterations in chronic lung disease. *Eur Respir J* **50**(2017).
49. Wilson, R. *et al.* Changes in the chondrocyte and extracellular matrix proteome during post-natal mouse cartilage development. *Mol Cell Proteomics* **11**, M111 014159 (2012).
50. Naba, A. *et al.* Extracellular matrix signatures of human primary metastatic colon cancers and their metastases to liver. *BMC Cancer* **14**, 518 (2014).
51. Perea-Gil, I. *et al.* In vitro comparative study of two decellularization protocols in search of an optimal myocardial scaffold for recellularization. *Am J Transl Res* **7**, 558-73 (2015).
52. Burnicka-Turek, O. *et al.* Pelota interacts with HAX1, EIF3G and SRPX and the resulting protein complexes are associated with the actin cytoskeleton. *BMC Cell Biol* **11**, 28 (2010).
53. Royer-Zemmour, B. *et al.* Epileptic and developmental disorders of the speech cortex: ligand/receptor interaction of wild-type and mutant SRPX2 with the plasminogen activator receptor uPAR. *Hum Mol Genet* **17**, 3617-30 (2008).
54. Song, X., Tanaka, H. & Ohta, K. Multiple roles of Equarin during lens development. *Dev Growth Differ* **56**, 199-205 (2014).
55. O'Leary, E.E. *et al.* Identification of steroid-sensitive gene-1/Ccdc80 as a JAK2-binding protein. *Mol Endocrinol* **27**, 619-34 (2013).
56. Nasseri, Y.Y. *et al.* Abnormalities in collagen composition may contribute to the pathogenesis of hemorrhoids: morphometric analysis. *Tech Coloproctol* **19**, 83-7 (2015).
57. Margetis, N. Pathophysiology of internal hemorrhoids. *Ann Gastroenterol* **32**, 264-272 (2019).
58. Devlin, B. & Roeder, K. Genomic control for association studies. *Biometrics* **55**, 997-1004 (1999).
59. Finucane, H.K. *et al.* Partitioning heritability by functional annotation using genome-wide association summary statistics. *Nat Genet* **47**, 1228-35 (2015).
60. Pruim, R.J. *et al.* LocusZoom: regional visualization of genome-wide association scan results. *Bioinformatics* **26**, 2336-7 (2010).
61. Waterhouse, A.M., Procter, J.B., Martin, D.M., Clamp, M. & Barton, G.J. Jalview Version 2--a multiple sequence alignment editor and analysis workbench. *Bioinformatics* **25**, 1189-91 (2009).
62. Staley, J.R. *et al.* PhenoScanner: a database of human genotype-phenotype associations. *Bioinformatics* **32**, 3207-3209 (2016).
63. Buniello, A. *et al.* The NHGRI-EBI GWAS Catalog of published genome-wide association studies, targeted arrays and summary statistics 2019. *Nucleic Acids Res* **47**, D1005-D1012 (2019).
64. Watanabe, K. *et al.* A global overview of pleiotropy and genetic architecture in complex traits. *Nat Genet* **51**, 1339-1348 (2019).
65. Braschi, B. *et al.* Genenames.org: the HGNC and VGNC resources in 2019. *Nucleic Acids Res* **47**, D786-D792 (2019).

66. Maglott, D., Ostell, J., Pruitt, K.D. & Tatusova, T. Entrez Gene: gene-centered information at NCBI. *Nucleic Acids Res* **39**, D52-7 (2011).
67. Aken, B.L. *et al.* The Ensembl gene annotation system. *Database (Oxford)* **2016**(2016).
68. Pers, T.H. *et al.* Biological interpretation of genome-wide association studies using predicted gene functions. *Nat Commun* **6**, 5890 (2015).
69. Smith, C.L. & Eppig, J.T. The mammalian phenotype ontology: enabling robust annotation and comparative analysis. *Wiley Interdiscip Rev Syst Biol Med* **1**, 390-399 (2009).
70. Amberger, J.S., Bocchini, C.A., Schiettecatte, F., Scott, A.F. & Hamosh, A. OMIM.org: Online Mendelian Inheritance in Man (OMIM(R)), an online catalog of human genes and genetic disorders. *Nucleic Acids Res* **43**, D789-98 (2015).
71. Watanabe, K., Taskesen, E., van Bochoven, A. & Posthuma, D. Functional mapping and annotation of genetic associations with FUMA. *Nat Commun* **8**, 1826 (2017).
72. de Leeuw, C.A., Mooij, J.M., Heskes, T. & Posthuma, D. MAGMA: generalized gene-set analysis of GWAS data. *PLoS Comput Biol* **11**, e1004219 (2015).
73. Consortium, G.T. *et al.* Genetic effects on gene expression across human tissues. *Nature* **550**, 204-213 (2017).
74. Liberzon, A. *et al.* The Molecular Signatures Database (MSigDB) hallmark gene set collection. *Cell Syst* **1**, 417-425 (2015).
75. Wang, K., Li, M. & Hakonarson, H. ANNOVAR: functional annotation of genetic variants from high-throughput sequencing data. *Nucleic Acids Res* **38**, e164 (2010).
