## Supplementary Table 1 for "Genome-wide analysis of 944,133 individuals provides insights into the etiology of hemorrhoidal disease"

**Supplementary Table 1. Study cohorts included the individual GWAS, meta-analyses, and follow up**

Case-control panels (after GWAS quality control) included in this study. For cohort description, see Online Methods. For details on quality control, see Supplementary Note.

**Cohort:** shorthand of the case-control panel; **Phenotype:** HEM, Hemorrhoidal disease cases, **CTRL**, control; **P**: *P*-value from student's t-test comparing age and BMI, and from chi-square test comparing sex between hemorrhoidal disease cases and controls in each individual cohort; **Age**, mean (±SD): mean age of onset/age of sampling and standard deviation; **Sex**, female %: percentage of females; **BMI**, mean (±SD): mean of body mass index. SD: standard deviation.

BMI: body mass index, UKBB:UK Biobank, EGCUT: Estonian Genome Center at the University of Tartu, MGI: Michigan Genomics Initiative, GERA:Genetic Epidemiology Research on Aging, HUNT: The Trøndelag Health Study, DBDS:Danish Blood Donor Study, DNPR:Danish National Patient Registry. * Age is calculated from birthday to follow-up date (either death, emigration or end of follow-up, which ever one came first).

| Cohort | Phenotype | N | Age, mean (±SD) | Sex, Female% | BMI, mean (±SD) |
|---|---|---|---|---|---|
| 23andMe | HEM | 174,785 | | 58,9% | |
| | CTRL | 228,060 | | 54,7% | |
| | P | | < 2.2E-16 | < 2.2E-16 | < 2.2E-16 |
| UKBB | HEM | 23,856 | 58.15 (±7.63) | 51,0% | 27.79 (±5.48) |
| | CTRL | 384,736 | 56.84 (±8.02) | 54,3% | 27.27 (±5.15) |
| | P | | <1.0E-04 | <1.0E-04 | <1.0E-04 |
| EGCUT | HEM | 6,927 | 57.84(16.21) | 70.0% | - |
| | CTRL | 30,299 | 53.56(17.03) | 65.0% | - |
| | P | | <1.0E-04 | <1.0E-04 | - |
| MGI | HEM | 4,539 | 60.87 (13.01) | 57,5% | - |
| | CTRL | 35,338 | 54.86 (16.51) | 64,5% | - |
| | P | | < 2.2E-16 | 7,6E-10 | - |
| GERA | HEM | 8,813 | 77.80 (±10.33) | 54.4% | 26.98 (±5.34) |
| | CTRL | 46,780 | 74.51 (±12.89) | 61.6% | 26.85 (±5.53) |
| | P | | <1.0E-04 | <1.0E-04 | 1.6E-02 |
| German haemorrhids case-control dataset | HEM | 1,144 | 61.29 (±11.39) | 46.4% | 26.64 (±4.51) |
| | CTRL | 2,740 | 48.38 (±15.77) | 50.9% | 26.02 (±4.73) |
| | P | | <1.0E-04 | 1.2E-02 | 1.2E-04 |
| HUNT | HEM | 977 | 60.22 (±14.88) | 59.0% | 27.00 (±4.37) |
| | CTRL | 68,314 | 53.64 (±17.37) | 52.9% | 27.03 (±4.41) |
| | P | | 1.8E-04 | <1.0E-04 | 8.4E-01 |
| DBDS | HEM | 1,754 | 47.44 (±11.03) | 49,0% | 25.93 (±3.90) |
| | CTRL | 54,643 | 39.33 (±12.66) | 49,4% | 25.32 (±3.97) |
| | P | | < 2.2E-16 | 0.7674 | 3.15E-10 |
| DNPR | HEM | 248,592 | 63.83 (±16.30)* | 51,0% | - |
| | CTRL | 7,923,939 | 49.93 (±26.41)* | 50,2% | - |
| | P | | < 2.2E-16 | 3.8E-15 | - |
| German anal canal tissue samples | HEM | 20 | 49.15 (±11.06 ) | 40.0% | 27.54 (±6.8) |
| | CTRL | 18 | 51.17 (±14.38) | 50.0% | 28.93 (±5.08) |
| | P | | 6.3E-01 | 7.7E-01 | 4.8E-01 |
