## Supplementary Table 2 for "Genome-wide analysis of 944,133 individuals provides insights into the etiology of hemorrhoidal disease"

**Supplementary Table 2. HEM GWAS risk loci**

102 newly identified genetic susceptibility loci associated with HEM at genome-wide significance ($P_{Meta}$<5×10$^{-8}$).

**Loci number**: number of susceptibility loci; **CHR**: chromosome; **start-end**: left/right association boundaries for each lead SNP defined by FUMA (see **Supplementary Note**). Genomic positions were retrieved from NCBI's dbSNP build v150 (genome build hg19); **Lead SNP (rsID)**: rs ID retrieved from NCBI's dbSNP build v150; **BP**: base pair position; **A1**: minor allele; **A2:** major allele; **EAF:** effect allele (i.e. minor allele) frequency; **OR (95% CI):** odds ratio (OR) and 95% confidence interval (CI 95%) with respect to A1; **P**: *P*-value; **Nearest Gene**: Nearest gene refers to the nearest protein-coding mapped gene within 100kb of the lead SNP, the number of additional mapped gene within each locus is given in brackets (see also **Supplementary Table 7**). "na" means that there was no protein-coding mapped gene within 100Kb of the lead SNP; **Association directions in individual cohorts**: Directions of the effect allele in five individual discovery datasets (being shown in order from 23andMe, UKBB, EGCUT, MGI to GERA). "+" represent risk effects, "-" represents protective effect and "?" represent that the tested lead SNP is not available in the association results of the individual cohort.

| Loci number | CHR | start-end | Lead SNP (rsID) | BP | A1 | A2 | EAF | OR (95% CI) | P | Nearest gene | Association directions in individual cohorts |
|---|---|---|---|---|---|---|---|---|---|---|---|
| 1 | 1 | 40257964-40301897 | rs11585073 | 40275089 | T | A | 0,25 | 1.06 (1.05-1.07) | 3,9E-33 | *BMP8B (+7)* | ++++? |
| 2 | 1 | 95203992-95309134 | rs11578225 | 95237528 | A | G | 0,21 | 0.97 (0.96-0.98) | 6,4E-13 | *SLC44A3 (+3)* | ----+ |
| 3 | 1 | 169081792-169521553 | rs145163454 | 169090748 | C | T | 0,03 | 0.87 (0.85-0.89) | 5,9E-28 | *ATP1B1 (+8)* | ----- |
| 4 | 1 | 204449952-204599461 | rs4951080 | 204533284 | A | G | 0,32 | 0.97 (0.97-0.98) | 2,8E-09 | *MDM4 (+6)* | --?-- |
| 5 | 1 | 219622596-219669226 | rs2605097 | 219642109 | C | A | 0,30 | 1.03 (1.02-1.03) | 4,3E-09 | *na (+1)* | +++++ |
| 6 | 2 | 20866699-20881840 | rs7594056 | 20867744 | A | G | 0,31 | 0.97 (0.96-0.97) | 7,2E-15 | *GDF7 (+2)* | ----? |
| 7 | 2 | 32499150-32849224 | rs6723226 | 32849207 | G | A | 0,35 | 1.02 (1.02-1.03) | 2,0E-08 | *TTC27 (+9)* | ++-+- |
| 8 | 2 | 36767257-36849220 | rs4670149 | 36826170 | T | A | 0,31 | 1.03 (1.02-1.04) | 1,3E-09 | *FEZ2 (+1)* | +++-+ |
| 9 | 2 | 45573050-45842064 | rs728327 | 45775995 | T | C | 0,34 | 0.97 (0.97-0.98) | 3,0E-10 | *SRBD1 (+2)* | ----- |
| 10 | 2 | 62926859-63298150 | rs4671051 | 63032696 | T | A | 0,36 | 1.02 (1.02-1.03) | 7,5E-09 | *EHBP1 (+8)* | +++++ |
| 11 | 2 | 64887382-65004346 | rs34532102 | 64905182 | T | C | 0,20 | 0.97 (0.96-0.98) | 1,3E-08 | *SERTAD2 (+1)* | ----- |
| 12 | 2 | 67874054-67954046 | rs2861709 | 67900116 | G | A | 0,06 | 1.05 (1.03-1.07) | 2,4E-09 | *na (+5)* | ++?-+ |
| 13 | 2 | 112654009-113830959 | rs57116599 | 112770799 | A | G | 0,23 | 1.03 (1.02-1.04) | 1,5E-11 | *MERTK (+22)* | +++++ |
| 14 | 2 | 145756260-145904907 | rs7423637 | 145818064 | T | A | 0,31 | 1.02 (1.02-1.03) | 1,0E-08 | *na (+2)* | +++++ |
| 15 | 2 | 148457312-148956584 | rs7559714 | 148640768 | C | T | 0,31 | 0.98 (0.97-0.99) | 5,0E-08 | *ACVR2A (+5)* | --?+- |
| 16 | 2 | 160899369-161340963 | rs4233681 | 160960058 | C | T | 0,48 | 0.97 (0.97-0.98) | 2,0E-11 | *ITGB6 (+5)* | ----+ |
| 17 | 2 | 173943763-174074375 | rs13017210 | 174008623 | T | A | 0,39 | 0.97 (0.97-0.98) | 3,8E-10 | *MAP3K20 (+1)* | --?-- |
| 18 | 2 | 176962102-176974104 | rs847148 | 176970456 | T | A | 0,32 | 0.96 (0.96-0.97) | 1,1E-17 | *HOXD11 (+8)* | ---+- |
| 19 | 2 | 189998020-190359085 | rs16831319 | 190150205 | C | T | 0,06 | 1.05 (1.03-1.07) | 1,0E-08 | *COL5A2 (+6)* | ++++- |
| 20 | 2 | 191259714-191478211 | rs34417560 | 191435716 | T | A | 0,25 | 1.03 (1.02-1.04) | 2,5E-08 | *TMEM194B (+10)* | ++++- |
| 21 | 3 | 14818716-14858226 | rs1689549 | 14837473 | C | T | 0,11 | 0.96 (0.95-0.98) | 4,4E-09 | *FGD5 (+5)* | ----- |
| 22 | 3 | 52214640-53171555 | rs9847710 | 53062661 | C | T | 0,41 | 1.04 (1.03-1.05) | 1,2E-23 | *SFMBT1 (+52)* | +++++ |
| 23 | 3 | 70779444-71074194 | rs2597301 | 70909494 | C | G | 0,32 | 1.03 (1.02-1.04) | 6,3E-14 | *FOXP1 (+2)* | +++++ |
| 24 | 3 | 111419457-111521268 | rs6792493 | 111477348 | G | A | 0,41 | 1.03 (1.02-1.04) | 9,5E-15 | *PLCXD2 (+3)* | +++-+ |
| 25 | 3 | 114498351-114520038 | rs9853475 | 114500255 | G | A | 0,28 | 0.97 (0.97-0.98) | 2,0E-09 | *ZBTB20 (+0)* | --+-+ |
| 26 | 3 | 156791268-156813672 | rs900400 | 156798775 | C | T | 0,41 | 0.97 (0.96-0.97) | 5,0E-18 | *na (+6)* | ----+ |
| 27 | 3 | 159922979-160309529 | rs3851366 | 160191374 | C | T | 0,47 | 0.98 (0.97-0.98) | 3,5E-10 | *KPNA4 (+13)* | ---+- |
| 28 | 4 | 8354149-8525162 | rs2631752 | 8405314 | C | G | 0,50 | 1.02 (1.02-1.03) | 4,4E-10 | *ACOX3 (+7)* | +++++ |
| 29 | 4 | 39441490-39501647 | rs2687965 | 39491521 | A | G | 0,49 | 0.98 (0.97-0.98) | 5,9E-09 | *UGDH (+5)* | ----- |
| 30 | 4 | 75656079-75686640 | rs11942410 | 75656079 | T | C | 0,26 | 0.97 (0.96-0.98) | 1,1E-08 | *BTC (+4)* | ---+- |
| 31 | 4 | 95903741-96099025 | rs28663472 | 95948204 | T | C | 0,45 | 0.98 (0.97-0.98) | 4,7E-10 | *BMPR1B (+1)* | ----- |
| 32 | 4 | 106048360-106276493 | rs6839705 | 106144735 | A | C | 0,37 | 1.03 (1.02-1.04) | 5,0E-12 | *TET2 (+4)* | +++++ |
| 33 | 4 | 124632243-124746377 | rs2060285 | 124746377 | A | C | 0,34 | 1.02 (1.02-1.03) | 4,2E-09 | *na (+3)* | +++++ |
| 34 | 4 | 126893588-127030561 | rs17824374 | 126924999 | C | T | 0,20 | 0.97 (0.96-0.98) | 3,3E-11 | *na (+2)* | ----- |
| 35 | 4 | 145227879-145517578 | rs1542726 | 145515769 | A | C | 0,42 | 1.03 (1.02-1.04) | 2,5E-12 | *HHIP (+6)* | ++-++ |
| 36 | 5 | 507307-686244 | rs72707023 | 667620 | A | G | 0,18 | 0.96 (0.95-0.97) | 2,4E-13 | *TPPP (+17)* | ---+- |
| 37 | 5 | 15670234-15785038 | rs61026653 | 15757570 | G | A | 0,14 | 0.97 (0.96-0.98) | 7,6E-09 | *FBXL7 (+2)* | ----- |
| 38 | 5 | 50658334-50779225 | rs62368263 | 50726027 | C | T | 0,14 | 0.96 (0.95-0.97) | 5,9E-15 | *ISL1 (+2)* | ----- |
| 39 | 5 | 51105360-51158351 | rs4485884 | 51108645 | A | T | 0,43 | 1.02 (1.02-1.03) | 5,6E-09 | *na (+3)* | ++-++ |
| 40 | 5 | 92948485-93571190 | rs6867042 | 93555761 | T | C | 0,13 | 0.97 (0.95-0.98) | 6,0E-09 | *KIAA0825 (+6)* | ----- |
| 41 | 5 | 97652398-97841726 | rs1563319 | 97764143 | T | A | 0,49 | 0.98 (0.97-0.98) | 9,5E-09 | *na (+2)* | ----- |
| 42 | 5 | 164487955-164786699 | rs12153515 | 164631794 | T | C | 0,13 | 1.04 (1.03-1.05) | 7,3E-10 | *na (+3)* | ++++- |
| 43 | 6 | 589924-646508 | rs4959352 | 624922 | C | A | 0,33 | 1.02 (1.02-1.03) | 1,5E-08 | *EXOC2 (+5)* | +++++ |
| 44 | 6 | 1767134-1777854 | rs722587 | 1775714 | T | A | 0,28 | 0.95 (0.94-0.96) | 1,5E-31 | *GMDS (+1)* | ----- |
| 45 | 6 | 17386607-17479588 | rs10807610 | 17387538 | C | A | 0,18 | 1.04 (1.03-1.05) | 2,3E-12 | *CAP2 (+0)* | +++-+ |

| | | | | | | | | | | | |
|---|---|---|---|---|---|---|---|---|---|---|---|
| 46 | 6 | 30062345-30071330 | rs1156533 | 30065149 | G | A | 0,30 | 0.98 (0.97-0.98) | 1,8E-08 | *TRIM31 (+25)* | ---++ |
| 47 | 6 | 117723640-117823508 | rs2180811 | 117780158 | A | T | 0,48 | 1.03 (1.02-1.04) | 1,9E-12 | *DCBLD1 (+7)* | +++++ |
| 48 | 6 | 134150561-134209837 | rs2327426 | 134202690 | C | T | 0,29 | 0.97 (0.97-0.98) | 2,7E-09 | *TCF21 (+3)* | ---+- |
| 49 | 6 | 152027074-152052215 | rs3020338 | 152034062 | A | G | 0,25 | 0.97 (0.96-0.98) | 2,2E-09 | *ESR1 (+0)* | ----+ |
| 50 | 6 | 152308973-152418575 | rs9322356 | 152408659 | A | G | 0,08 | 1.04 (1.03-1.06) | 2,0E-08 | *ESR1 (+0)* | +-+++ |
| 51 | 6 | 169569045-169618454 | rs3253 | 169616112 | T | C | 0,32 | 1.03 (1.02-1.04) | 5,0E-12 | *THBS2 (+4)* | +++++ |
| 52 | 7 | 40361834-40481812 | rs6462976 | 40396300 | T | C | 0,46 | 0.97 (0.96-0.98) | 1,2E-12 | *SUGCT (+0)* | ----- |
| 53 | 7 | 55103203-55161043 | rs7795564 | 55124829 | A | G | 0,39 | 0.97 (0.96-0.98) | 3,4E-13 | *EGFR (+0)* | ----- |
| 54 | 7 | 73146742-73461430 | rs11770437 | 73319809 | T | C | 0,36 | 0.96 (0.95-0.97) | 3,2E-18 | *WBSCR28 (+12)* | ----- |
| 55 | 7 | 100528223-100753880 | rs4556017 | 100632790 | C | T | 0,14 | 1.06 (1.05-1.07) | 1,3E-22 | *MUC12 (+24)* | +++++ |
| 56 | 7 | 102407236-103186271 | rs7778418 | 102429704 | C | T | 0,34 | 1.04 (1.03-1.05) | 6,4E-18 | *FAM185A (+25)* | +++-+ |
| 57 | 7 | 127264150-127757563 | rs806169 | 127300668 | C | G | 0,33 | 0.97 (0.96-0.98) | 3,6E-11 | *SND1 (+8)* | ---+- |
| 58 | 8 | 6361229-6655391 | rs2912053 | 6645793 | G | C | 0,40 | 1.04 (1.03-1.05) | 2,3E-17 | *na (+4)* | +++++ |
| 59 | 8 | 22447426-22542962 | rs13271626 | 22467760 | G | C | 0,33 | 1.03 (1.02-1.04) | 3,4E-10 | *CCAR2 (+8)* | ++-++ |
| 60 | 8 | 71042430-72277688 | rs1838392 | 71682583 | T | G | 0,39 | 1.05 (1.05-1.06) | 4,6E-39 | *XKR9 (+8)* | +++++ |
| 61 | 8 | 77493526-77493526 | rs2581260 | 77493526 | A | T | 0,16 | 0.97 (0.96-0.98) | 1,1E-09 | *na (+2)* | -+-+- |
| 62 | 8 | 130717716-130738972 | rs10956488 | 130717755 | G | A | 0,15 | 1.03 (1.02-1.04) | 1,7E-09 | *GSDMC (+0)* | +++-+ |
| 63 | 8 | 144973183-145086428 | rs58579887 | 145028587 | C | T | 0,40 | 1.03 (1.02-1.04) | 1,7E-11 | *PLEC (+20)* | +++-? |
| 64 | 9 | 21995044-22125503 | rs1333047 | 22124504 | A | T | 0,50 | 1.04 (1.04-1.05) | 7,8E-28 | *CDKN2B (+4)* | +++++ |
| 65 | 9 | 94940133-95555488 | rs755209 | 95489671 | A | G | 0,28 | 1.02 (1.02-1.03) | 3,1E-08 | *BICD2 (+17)* | +++++ |
| 66 | 9 | 119155568-119238367 | rs1858015 | 119167220 | C | T | 0,32 | 0.97 (0.96-0.98) | 6,1E-11 | *PAPPA (+3)* | ----- |
| 67 | 9 | 136065526-136356448 | rs676996 | 136146077 | G | T | 0,34 | 0.94 (0.94-0.95) | 5,3E-43 | *ABO (+23)* | ----- |
| 68 | 10 | 24326623-24366458 | rs6482359 | 24330805 | A | G | 0,39 | 1.03 (1.02-1.04) | 3,1E-11 | *KIAA1217 (+1)* | ++-++ |
| 69 | 10 | 126688200-126738634 | rs3012065 | 126737579 | C | T | 0,32 | 1.02 (1.02-1.03) | 4,1E-08 | *CTBP2 (+2)* | +++++ |
| 70 | 11 | 10377175-10386083 | rs2218793 | 10380828 | A | C | 0,29 | 1.03 (1.02-1.04) | 1,1E-13 | *AMPD3 (+5)* | +++++ |
| 71 | 11 | 10655021-10701556 | rs4910165 | 10674044 | C | G | 0,32 | 1.03 (1.03-1.04) | 3,9E-16 | *MRVI1 (+5)* | +++++ |
| 72 | 11 | 47372377-47989003 | rs10838738 | 47663049 | G | A | 0,35 | 1.03 (1.02-1.03) | 1,7E-09 | *MTCH2 (+29)* | +++++ |
| 73 | 11 | 69971277-70028543 | rs2186797 | 70007770 | C | T | 0,06 | 0.96 (0.94-0.97) | 4,5E-08 | *ANO1 (+4)* | +++-+ |
| 74 | 11 | 112788472-113034787 | rs2212450 | 112826867 | C | T | 0,43 | 1.03 (1.02-1.04) | 5,4E-11 | *NCAM1 (+5)* | ++?+- |
| 75 | 11 | 122991556-123026823 | rs4345978 | 123009751 | T | G | 0,25 | 0.97 (0.96-0.98) | 1,2E-13 | *CLMP (+2)* | ---++ |
| 76 | 12 | 20289914-20318700 | rs11045079 | 20297976 | G | A | 0,14 | 0.97 (0.95-0.98) | 1,8E-09 | *na (+2)* | ----- |
| 77 | 12 | 24424791-24442214 | rs4579999 | 24439661 | T | C | 0,47 | 1.03 (1.02-1.04) | 3,3E-13 | *na (+1)* | +++++ |
| 78 | 12 | 24714667-24802324 | rs111235435 | 24757629 | T | C | 0,38 | 1.02 (1.02-1.03) | 5,0E-09 | *na (+2)* | +++++ |
| 79 | 12 | 48164758-48181028 | rs13632 | 48177238 | A | G | 0,24 | 1.03 (1.02-1.04) | 6,1E-11 | *HDAC7 (+7)* | +++-- |
| 80 | 12 | 54353523-54376264 | rs920778 | 54360232 | G | A | 0,35 | 0.97 (0.97-0.98) | 6,9E-10 | *HOXC11 (+11)* | ---+- |
| 81 | 12 | 66326943-66413327 | rs11176001 | 66409367 | A | C | 0,13 | 0.92 (0.91-0.93) | 2,5E-46 | *HMGA2 (+3)* | ----- |
| 82 | 12 | 114632506-114687311 | rs2555004 | 114686645 | G | A | 0,49 | 0.98 (0.97-0.98) | 5,4E-10 | *na (+2)* | ----+ |
| 83 | 13 | 51064533-51531815 | rs7994724 | 51445560 | A | G | 0,37 | 1.04 (1.03-1.04) | 1,7E-17 | *DLEU7 (+5)* | +++-+ |
| 84 | 15 | 32994056-33023486 | rs11635984 | 33012232 | C | T | 0,39 | 0.97 (0.96-0.98) | 4,1E-13 | *GREM1 (+3)* | ----- |
| 85 | 15 | 67437863-67468285 | rs17293632 | 67442596 | T | C | 0,24 | 1.06 (1.05-1.07) | 6,9E-32 | *SMAD3 (+8)* | +++++ |
| 86 | 15 | 68324830-68546275 | rs12594232 | 68469402 | G | A | 0,45 | 0.98 (0.97-0.98) | 1,0E-09 | *PIAS1 (+12)* | ----- |
| 87 | 15 | 96100923-96139843 | rs7183672 | 96101018 | G | A | 0,32 | 1.03 (1.02-1.04) | 1,2E-13 | *na (+1)* | +++-+ |
| 88 | 16 | 15858791-15882857 | rs6498573 | 15879373 | T | C | 0,14 | 1.04 (1.03-1.05) | 5,2E-12 | *MYH11 (+6)* | +++++ |
| 89 | 16 | 86621902-86635787 | rs4843407 | 86629047 | A | G | 0,27 | 0.97 (0.96-0.98) | 4,0E-12 | *FOXL1 (+2)* | ---+- |
| 90 | 17 | 7417640-7571752 | rs78378222 | 7571752 | G | T | 0,01 | 1.12 (1.08-1.16) | 4,7E-10 | *TP53 (+6)* | +++-+ |
| 91 | 17 | 12394980-12410649 | rs62061554 | 12401309 | A | G | 0,12 | 1.04 (1.03-1.05) | 2,6E-10 | *na (+1)* | +++++ |
| 92 | 17 | 17883848-18036283 | rs854786 | 18036283 | G | A | 0,32 | 0.98 (0.97-0.98) | 1,3E-08 | *MYO15A (+17)* | ---+- |
| 93 | 17 | 29400158-29735829 | rs2525570 | 29681245 | A | G | 0,39 | 0.98 (0.97-0.98) | 6,8E-10 | *NF1 (+10)* | ----+ |
| 94 | 17 | 64236318-64331156 | rs4423457 | 64284000 | A | G | 0,42 | 0.98 (0.97-0.99) | 3,4E-08 | *PRKCA (+2)* | ----- |
| 95 | 19 | 11262477-11277074 | rs2421206 | 11262477 | G | T | 0,35 | 1.03 (1.02-1.04) | 2,3E-14 | *SPC24 (+1)* | +++++ |
| 96 | 19 | 44358042-44432840 | rs8106090 | 44371210 | G | A | 0,49 | 1.02 (1.01-1.03) | 3,3E-08 | *ZNF404 (+18)* | ++--+ |
| 97 | 20 | 56016072-56029604 | rs34161672 | 56020599 | A | G | 0,32 | 0.97 (0.96-0.98) | 4,2E-12 | *RBM38 (+1)* | ---+? |
| 98 | 20 | 58993857-59049432 | rs35384758 | 58998651 | G | A | 0,24 | 1.03 (1.02-1.04) | 4,0E-08 | *na (+2)* | +++++ |
| 99 | 21 | 30309586-30843684 | rs2832279 | 30613466 | A | C | 0,35 | 1.03 (1.02-1.04) | 4,5E-11 | *BACH1 (+20)* | ++++- |
| 100 | 22 | 29669693-29793525 | rs174767 | 29740728 | A | G | 0,38 | 0.98 (0.97-0.99) | 1,4E-08 | *AP1B1 (+11)* | ----- |
| 101 | X | 37933437-38064927 | rs35318931 | 38009121 | A | G | 0,08 | 0.96 (0.95-0.97) | 3,2E-11 | *SRPX (+2)* | --??- |
| 102 | X | 109682057-109940577 | rs5942977 | 109833905 | A | G | 0,40 | 0.98 (0.97-0.99) | 6,9E-10 | *CHRDL1 (+2)* | --??- |
