## Supplementary Table 3 for "Genome-wide analysis of 944,133 individuals provides insights into the etiology of hemorrhoidal disease"

**Supplementary Table 3. Number of fine-mapped variants in 95% credible sets.**

Variants in 95% fine-mapped credible sets for 102 susceptibility loci associated with HEM at genome-wide significance (PMeta<5×10-8). For 6 loci, the 95% credible set consisted of a single variant ('single variant credible sets'), and for 96 others the credible set consisted of multiple variant.

| Size of 95% credible set | 1 | 2-5 | 6-10 | 11-20 | 21-50 | >50 |
|---|---|---|---|---|---|---|
| Number of loci | 6 | 19 | 11 | 18 | 26 | 22 |

**Locus:** number of susceptibility locus according to **Supplementary Table 2. SNP**: variants in the 95% credible sets (see **Online Methods**). Variants were sorted by the posterior probability of association, and variants were added to the 'credible set' of associated variants until the sum of their posterior probability exceeded 95%. Rs-numbers retrieved from NCBI's dbSNP build v150 (genome build hg19); **CHR**: chromosome; **BP**: base pair position; **Posterior probability**: posterior probability of causality to each SNP variant.

| Locus | SNP | CHR | BP | Posterior_probability |
|---|---|---|---|---|
| 1 | rs11585073 | 1 | 40275089 | 0.243938 |
| 1 | rs12084017 | 1 | 40276348 | 0.243938 |
| 1 | rs59766044 | 1 | 40282708 | 0.401525 |
| 1 | rs11578451 | 1 | 40285773 | 0.0932837 |
| 2 | rs17414785 | 1 | 95236421 | 0.0953714 |
| 2 | rs11578225 | 1 | 95237528 | 0.0953714 |
| 2 | rs11577484 | 1 | 95238755 | 0.0953714 |
| 2 | rs17414905 | 1 | 95239080 | 0.0520791 |
| 2 | rs17414947 | 1 | 95239533 | 0.0953714 |
| 2 | rs11578548 | 1 | 95240643 | 0.0953714 |
| 2 | rs11580446 | 1 | 95241697 | 0.0821776 |
| 2 | rs61772753 | 1 | 95242022 | 0.0953714 |
| 2 | rs72733523 | 1 | 95249105 | 0.0220702 |
| 2 | rs72733524 | 1 | 95249106 | 0.0293339 |
| 2 | rs11581918 | 1 | 95249408 | 0.0527094 |
| 2 | rs11581919 | 1 | 95249432 | 0.0527094 |
| 2 | rs61772759 | 1 | 95251579 | 0.0527094 |
| 2 | rs61774560 | 1 | 95252976 | 0.0527094 |
| 3 | rs145163454 | 1 | 169090748 | 0.617945 |
| 3 | rs144737447 | 1 | 169160458 | 0.378568 |
| 4 | rs10494852 | 1 | 204457786 | 0.00958976 |
| 4 | rs11240751 | 1 | 204462050 | 0.0123881 |
| 4 | rs4951075 | 1 | 204464446 | 0.0132927 |
| 4 | rs4951076 | 1 | 204466176 | 0.0074396 |
| 4 | rs10900593 | 1 | 204470096 | 0.0116532 |
| 4 | rs11240753 | 1 | 204475513 | 0.00414886 |
| 4 | rs4951389 | 1 | 204475834 | 0.00471121 |
| 4 | rs7367519 | 1 | 204479176 | 0.00655801 |
| 4 | rs12032733 | 1 | 204481209 | 0.00578402 |
| 4 | rs12738124 | 1 | 204481368 | 0.00510412 |
| 4 | rs4951077 | 1 | 204482262 | 0.00311196 |
| 4 | rs11240756 | 1 | 204482594 | 0.00578402 |
| 4 | rs4245736 | 1 | 204486205 | 0.00311196 |
| 4 | rs4245737 | 1 | 204486242 | 0.0132927 |
| 4 | rs1380576 | 1 | 204488278 | 0.00655801 |
| 4 | rs12136299 | 1 | 204489143 | 0.00655801 |
| 4 | rs4951393 | 1 | 204489557 | 0.0151716 |
| 4 | rs61817958 | 1 | 204491612 | 0.0108965 |
| 4 | rs12041075 | 1 | 204492153 | 0.0074396 |
| 4 | rs7556655 | 1 | 204493844 | 0.00351888 |
| 4 | rs4252677 | 1 | 204495253 | 0.0074396 |

| | | | | |
|---|---|---|---|---|
| 4 | rs4252685 | 1 | 204496856 | 0.00844425 |
| 4 | rs4252686 | 1 | 204496895 | 0.0074396 |
| 4 | rs4252687 | 1 | 204497335 | 0.00351888 |
| 4 | rs4252694 | 1 | 204499056 | 0.0074396 |
| 4 | rs4252708 | 1 | 204508159 | 0.0108965 |
| 4 | rs4252717 | 1 | 204512100 | 0.00655801 |
| 4 | rs4252718 | 1 | 204512195 | 0.0132927 |
| 4 | rs2369244 | 1 | 204515299 | 0.0132927 |
| 4 | rs2290855 | 1 | 204515863 | 0.00655801 |
| 4 | rs2290854 | 1 | 204516025 | 0.00351888 |
| 4 | rs1563828 | 1 | 204516577 | 0.0526447 |
| 4 | rs4252734 | 1 | 204516947 | 0.00351888 |
| 4 | rs4252736 | 1 | 204517097 | 0.0074396 |
| 4 | rs4951396 | 1 | 204520209 | 0.0151716 |
| 4 | rs12119098 | 1 | 204521577 | 0.00958976 |
| 4 | rs10900596 | 1 | 204522457 | 0.00311196 |
| 4 | rs10900597 | 1 | 204522489 | 0.0445876 |
| 4 | rs1460036 | 1 | 204527379 | 0.00311196 |
| 4 | rs12741351 | 1 | 204528256 | 0.0388893 |
| 4 | rs11240758 | 1 | 204528651 | 0.00655801 |
| 4 | rs11801289 | 1 | 204528969 | 0.00450659 |
| 4 | rs12125533 | 1 | 204529302 | 0.0132927 |
| 4 | rs12740680 | 1 | 204530505 | 0.00351888 |
| 4 | rs4951397 | 1 | 204531567 | 0.0296348 |
| 4 | rs4951398 | 1 | 204531876 | 0.0132927 |
| 4 | rs4951080 | 1 | 204533284 | 0.119139 |
| 4 | rs4951400 | 1 | 204534571 | 0.0445876 |
| 4 | rs6681905 | 1 | 204535789 | 0.00398115 |
| 4 | rs6679336 | 1 | 204536197 | 0.00311196 |
| 4 | rs4951402 | 1 | 204539122 | 0.0388893 |
| 4 | rs4951403 | 1 | 204539241 | 0.0074396 |
| 4 | rs6594016 | 1 | 204539462 | 0.0182624 |
| 4 | rs10900599 | 1 | 204541943 | 0.0339385 |
| 4 | rs7541589 | 1 | 204542521 | 0.00351888 |
| 4 | rs12039454 | 1 | 204542580 | 0.0151716 |
| 4 | rs885012 | 1 | 204544001 | 0.00351888 |
| 4 | rs11240760 | 1 | 204547268 | 0.00311196 |
| 4 | rs35270244 | 1 | 204548416 | 0.0116532 |
| 4 | rs10793765 | 1 | 204549375 | 0.0074396 |
| 4 | rs10793766 | 1 | 204549496 | 0.00958976 |
| 4 | rs10793767 | 1 | 204549564 | 0.0116532 |
| 4 | rs12139477 | 1 | 204553104 | 0.0674278 |
| 4 | rs12130686 | 1 | 204554057 | 0.00535282 |
| 4 | rs12133735 | 1 | 204556836 | 0.00955999 |
| 4 | rs12566957 | 1 | 204556958 | 0.00955999 |
| 4 | rs10900600 | 1 | 204559707 | 0.0108965 |
| 4 | rs6682208 | 1 | 204566183 | 0.0036557 |
| 4 | rs11240761 | 1 | 204566700 | 0.00897098 |
| 4 | rs6594019 | 1 | 204568610 | 0.00655801 |
| 5 | rs2605091 | 1 | 219640305 | 0.100364 |
| 5 | rs2605092 | 1 | 219640389 | 0.0330308 |
| 5 | rs2605093 | 1 | 219640521 | 0.0289239 |

| | | | | |
|---|---|---|---|---|
| 5 | rs2820436 | 1 | 219640680 | 0.0330308 |
| 5 | rs2605094 | 1 | 219640763 | 0.0330308 |
| 5 | rs2605095 | 1 | 219641450 | 0.0289239 |
| 5 | rs2605096 | 1 | 219641900 | 0.0253419 |
| 5 | rs2605097 | 1 | 219642109 | 0.131485 |
| 5 | rs1982499 | 1 | 219642518 | 0.0289239 |
| 5 | rs2605098 | 1 | 219643649 | 0.0150187 |
| 5 | rs2605100 | 1 | 219644224 | 0.0452194 |
| 5 | rs2605101 | 1 | 219644496 | 0.0289239 |
| 5 | rs2605102 | 1 | 219644993 | 0.0253419 |
| 5 | rs2605103 | 1 | 219645192 | 0.0377422 |
| 5 | rs2605104 | 1 | 219645197 | 0.04315 |
| 5 | rs2605105 | 1 | 219646223 | 0.0289239 |
| 5 | rs2605106 | 1 | 219648800 | 0.0377422 |
| 5 | rs2791546 | 1 | 219648808 | 0.0493607 |
| 5 | rs1340098 | 1 | 219649771 | 0.0767756 |
| 5 | rs2605107 | 1 | 219650741 | 0.00973076 |
| 5 | rs748273 | 1 | 219650950 | 0.0236173 |
| 5 | rs2820430 | 1 | 219651779 | 0.0110269 |
| 5 | rs1563352 | 1 | 219652795 | 0.00542023 |
| 5 | rs1563353 | 1 | 219653026 | 0.0110269 |
| 5 | rs1563355 | 1 | 219653101 | 0.00330074 |
| 5 | rs2445127 | 1 | 219653685 | 0.00542023 |
| 5 | rs2486895 | 1 | 219653704 | 0.00422496 |
| 5 | rs2791551 | 1 | 219655031 | 0.00330074 |
| 5 | rs1531093 | 1 | 219655838 | 0.00478407 |
| 5 | rs1531094 | 1 | 219655876 | 0.00373331 |
| 5 | rs1531096 | 1 | 219656701 | 0.0207737 |
| 5 | rs1531097 | 1 | 219656760 | 0.00373331 |
| 5 | rs2791549 | 1 | 219658003 | 0.00973076 |
| 6 | rs77525005 | 2 | 20866699 | 0.101926 |
| 6 | rs10205228 | 2 | 20866720 | 0.0859151 |
| 6 | rs7594056 | 2 | 20867744 | 0.279283 |
| 6 | rs7580188 | 2 | 20868125 | 0.11545 |
| 6 | rs7589372 | 2 | 20868179 | 0.11545 |
| 6 | rs35148785 | 2 | 20868414 | 0.137618 |
| 6 | rs13013489 | 2 | 20870579 | 0.0813767 |
| 6 | rs56353595 | 2 | 20871398 | 0.0683738 |
| 7 | rs1153122 | 2 | 32533457 | 0.00428172 |
| 7 | rs4019436 | 2 | 32603640 | 0.00490245 |
| 7 | rs4952211 | 2 | 32611512 | 0.0290416 |
| 7 | rs3769600 | 2 | 32616879 | 0.00658711 |
| 7 | rs116617324 | 2 | 32622709 | 0.0256121 |
| 7 | rs176401 | 2 | 32636511 | 0.0364014 |
| 7 | rs176404 | 2 | 32638846 | 0.0138728 |
| 7 | rs10182170 | 2 | 32704186 | 0.00371816 |
| 7 | rs2366894 | 2 | 32713706 | 0.00371816 |
| 7 | rs17428810 | 2 | 32736043 | 0.0314355 |
| 7 | rs2069213 | 2 | 32743084 | 0.0155856 |
| 7 | rs2249109 | 2 | 32750879 | 0.00527769 |
| 7 | rs2710629 | 2 | 32765844 | 0.00841334 |
| 7 | rs2754513 | 2 | 32782763 | 0.0108054 |

| | | | | |
|---|---|---|---|---|
| 7 | rs2754522 | 2 | 32801257 | 0.00538223 |
| 7 | rs62136333 | 2 | 32803874 | 0.0176306 |
| 7 | rs2710606 | 2 | 32811909 | 0.0174757 |
| 7 | rs1901355 | 2 | 32835664 | 0.157517 |
| 7 | rs13035097 | 2 | 32836362 | 0.218341 |
| 7 | rs6723226 | 2 | 32849207 | 0.325179 |
| 7 | rs35890136 | 2 | 32861027 | 0.00995975 |
| 8 | rs1544655 | 2 | 36825137 | 0.0350794 |
| 8 | rs4670570 | 2 | 36825932 | 0.237847 |
| 8 | rs4670148 | 2 | 36826126 | 0.237847 |
| 8 | rs4670149 | 2 | 36826170 | 0.272761 |
| 8 | rs11124521 | 2 | 36826743 | 0.181135 |
| 9 | rs10460504 | 2 | 45575110 | 0.299284 |
| 9 | rs728327 | 2 | 45775995 | 0.668817 |
| 10 | rs360804 | 2 | 62939397 | 0.00276768 |
| 10 | rs360797 | 2 | 62942840 | 0.00190954 |
| 10 | rs1906198 | 2 | 62946658 | 0.0218346 |
| 10 | rs360800 | 2 | 62954326 | 0.00989426 |
| 10 | rs11125942 | 2 | 62966758 | 0.00820527 |
| 10 | rs2136737 | 2 | 62969310 | 0.00671153 |
| 10 | rs9678329 | 2 | 62979759 | 0.00215979 |
| 10 | rs4671450 | 2 | 62981869 | 0.00215979 |
| 10 | rs4671050 | 2 | 62988169 | 0.00244422 |
| 10 | rs7573756 | 2 | 62990637 | 0.00215979 |
| 10 | rs34104251 | 2 | 62995183 | 0.0249631 |
| 10 | rs3927201 | 2 | 63001335 | 0.00763419 |
| 10 | rs10153716 | 2 | 63007059 | 0.0374314 |
| 10 | rs7562149 | 2 | 63008414 | 0.0326847 |
| 10 | rs13013218 | 2 | 63011671 | 0.0374314 |
| 10 | rs1553832 | 2 | 63013515 | 0.0285561 |
| 10 | rs9973612 | 2 | 63019815 | 0.0285561 |
| 10 | rs12989754 | 2 | 63020970 | 0.0374314 |
| 10 | rs4461246 | 2 | 63027671 | 0.016733 |
| 10 | rs4671051 | 2 | 63032696 | 0.0837335 |
| 10 | rs1828398 | 2 | 63034870 | 0.0726274 |
| 10 | rs1022110 | 2 | 63046039 | 0.016733 |
| 10 | rs7562734 | 2 | 63047973 | 0.0428917 |
| 10 | rs11682530 | 2 | 63073194 | 0.016733 |
| 10 | rs6545971 | 2 | 63074986 | 0.0413508 |
| 10 | rs2421816 | 2 | 63075033 | 0.0249631 |
| 10 | rs11893131 | 2 | 63079528 | 0.00590372 |
| 10 | rs13418889 | 2 | 63079775 | 0.0237681 |
| 10 | rs17027429 | 2 | 63079983 | 0.0272728 |
| 10 | rs2080227 | 2 | 63081759 | 0.0444162 |
| 10 | rs6545972 | 2 | 63084682 | 0.0237681 |
| 10 | rs7567923 | 2 | 63085848 | 0.0191089 |
| 10 | rs7557501 | 2 | 63086639 | 0.0272728 |
| 10 | rs6545973 | 2 | 63089434 | 0.0218346 |
| 10 | rs9789661 | 2 | 63089644 | 0.0313129 |
| 10 | rs2058567 | 2 | 63091426 | 0.00671153 |
| 10 | rs2430388 | 2 | 63106784 | 0.0086886 |
| 10 | rs2539982 | 2 | 63111004 | 0.00457585 |

| | | | | |
|---|---|---|---|---|
| 10 | rs1420018 | 2 | 63112500 | 0.0146608 |
| 10 | rs2215870 | 2 | 63132342 | 0.00403194 |
| 10 | rs2539985 | 2 | 63134507 | 0.00989426 |
| 10 | rs2710639 | 2 | 63138963 | 0.00171976 |
| 10 | rs2539987 | 2 | 63142864 | 0.00190954 |
| 10 | rs2710640 | 2 | 63144305 | 0.00355469 |
| 10 | rs2710642 | 2 | 63149557 | 0.0207261 |
| 10 | rs2539990 | 2 | 63170185 | 0.00178744 |
| 10 | rs2710637 | 2 | 63173450 | 0.00519608 |
| 10 | rs2058566 | 2 | 63179076 | 0.0128525 |
| 10 | rs2539978 | 2 | 63195179 | 0.00171976 |
| 10 | rs1468748 | 2 | 63204611 | 0.00707924 |
| 10 | rs2710644 | 2 | 63248968 | 0.00590372 |
| 10 | rs6705798 | 2 | 63259881 | 0.00403194 |
| 11 | rs34338966 | 2 | 64892466 | 0.0822522 |
| 11 | rs12614829 | 2 | 64893183 | 0.0922782 |
| 11 | rs7596561 | 2 | 64894148 | 0.0372 |
| 11 | rs71424129 | 2 | 64895167 | 0.0922782 |
| 11 | rs34642907 | 2 | 64895731 | 0.0822522 |
| 11 | rs34218958 | 2 | 64895903 | 0.0922782 |
| 11 | rs11894737 | 2 | 64898662 | 0.0465691 |
| 11 | rs34532102 | 2 | 64905182 | 0.103569 |
| 11 | rs6759291 | 2 | 64909691 | 0.073346 |
| 11 | rs6754136 | 2 | 64911801 | 0.0349224 |
| 11 | rs13007087 | 2 | 64915276 | 0.0310133 |
| 11 | rs6750285 | 2 | 64920032 | 0.0310133 |
| 11 | rs13013483 | 2 | 64923311 | 0.00774235 |
| 11 | rs1968179 | 2 | 64929212 | 0.0349224 |
| 11 | rs13026184 | 2 | 64930786 | 0.00866899 |
| 11 | rs6735239 | 2 | 64931994 | 0.0527686 |
| 11 | rs72814070 | 2 | 64935357 | 0.00971112 |
| 11 | rs1558703 | 2 | 64938289 | 0.00866899 |
| 11 | rs13016603 | 2 | 64941506 | 0.00618439 |
| 11 | rs35636928 | 2 | 64942953 | 0.00774235 |
| 11 | rs35862374 | 2 | 64953393 | 0.00618439 |
| 11 | rs13031521 | 2 | 64957938 | 0.00691803 |
| 11 | rs12994639 | 2 | 64959331 | 0.00691803 |
| 12 | rs74445493 | 2 | 67874054 | 0.0516043 |
| 12 | rs58909924 | 2 | 67880862 | 0.11175 |
| 12 | rs114233333 | 2 | 67881931 | 0.0842064 |
| 12 | rs75984732 | 2 | 67897842 | 0.0419267 |
| 12 | rs76316246 | 2 | 67898111 | 0.0365321 |
| 12 | rs72905801 | 2 | 67898999 | 0.0192683 |
| 12 | rs2861709 | 2 | 67900116 | 0.139554 |
| 12 | rs2163363 | 2 | 67901174 | 0.0277843 |
| 12 | rs2163364 | 2 | 67901540 | 0.0297457 |
| 12 | rs79994788 | 2 | 67909327 | 0.0297457 |
| 12 | rs76794380 | 2 | 67909332 | 0.0297457 |
| 12 | rs78133950 | 2 | 67909406 | 0.0318501 |
| 12 | rs76290730 | 2 | 67910151 | 0.0297457 |
| 12 | rs76307505 | 2 | 67910238 | 0.0318501 |
| 12 | rs77274689 | 2 | 67910334 | 0.0297457 |

| | | | | |
|---|---|---|---|---|
| 12 | rs80194914 | 2 | 67911856 | 0.0277843 |
| 12 | rs77481855 | 2 | 67913448 | 0.025956 |
| 12 | rs77516753 | 2 | 67913713 | 0.0277843 |
| 12 | rs79131920 | 2 | 67916332 | 0.0391337 |
| 12 | rs80250779 | 2 | 67918089 | 0.0419267 |
| 12 | rs79059272 | 2 | 67922634 | 0.0682068 |
| 13 | rs12466812 | 2 | 112654009 | 0.0336981 |
| 13 | rs6729037 | 2 | 112656777 | 0.0253633 |
| 13 | rs1516629 | 2 | 112658779 | 0.0220198 |
| 13 | rs2001444 | 2 | 112659538 | 0.0125688 |
| 13 | rs17835605 | 2 | 112661106 | 0.0220198 |
| 13 | rs55785997 | 2 | 112661447 | 0.0292283 |
| 13 | rs113645544 | 2 | 112662040 | 0.0336981 |
| 13 | rs79559916 | 2 | 112665080 | 0.0191261 |
| 13 | rs17174870 | 2 | 112665201 | 0.0220198 |
| 13 | rs55728465 | 2 | 112666096 | 0.0191261 |
| 13 | rs55708724 | 2 | 112666922 | 0.0220198 |
| 13 | rs12477716 | 2 | 112668644 | 0.0191261 |
| 13 | rs7568632 | 2 | 112669950 | 0.0191261 |
| 13 | rs1400321 | 2 | 112670046 | 0.00829476 |
| 13 | rs58641606 | 2 | 112670732 | 0.0166205 |
| 13 | rs7575552 | 2 | 112673634 | 0.0144499 |
| 13 | rs80088736 | 2 | 112674805 | 0.0109377 |
| 13 | rs867311 | 2 | 112677970 | 0.00829476 |
| 13 | rs2001201 | 2 | 112678350 | 0.00952274 |
| 13 | rs59650987 | 2 | 112678976 | 0.0109377 |
| 13 | rs55972521 | 2 | 112680519 | 0.0109377 |
| 13 | rs1400325 | 2 | 112680934 | 0.0109377 |
| 13 | rs1400324 | 2 | 112681220 | 0.0109377 |
| 13 | rs57640566 | 2 | 112684173 | 0.00722854 |
| 13 | rs74562473 | 2 | 112684680 | 0.00952274 |
| 13 | rs75508922 | 2 | 112688397 | 0.00722854 |
| 13 | rs79433506 | 2 | 112695056 | 0.00829476 |
| 13 | rs113415773 | 2 | 112695716 | 0.00630235 |
| 13 | rs76852916 | 2 | 112701291 | 0.00829476 |
| 13 | rs1113419 | 2 | 112707483 | 0.00722854 |
| 13 | rs12470338 | 2 | 112709756 | 0.00952274 |
| 13 | rs17175275 | 2 | 112712329 | 0.00722854 |
| 13 | rs17175331 | 2 | 112712870 | 0.00722854 |
| 13 | rs113494688 | 2 | 112717927 | 0.00952274 |
| 13 | rs56272510 | 2 | 112727604 | 0.00691869 |
| 13 | rs4434006 | 2 | 112728811 | 0.00829476 |
| 13 | rs113629868 | 2 | 112738246 | 0.0109377 |
| 13 | rs77290039 | 2 | 112741106 | 0.00829476 |
| 13 | rs10207726 | 2 | 112744260 | 0.00622263 |
| 13 | rs76944504 | 2 | 112745774 | 0.00952274 |
| 13 | rs76514293 | 2 | 112751516 | 0.0109377 |
| 13 | rs77226570 | 2 | 112751534 | 0.00952274 |
| 13 | rs77702371 | 2 | 112753020 | 0.00952274 |
| 13 | rs3811632 | 2 | 112754828 | 0.00622263 |
| 13 | rs58065683 | 2 | 112756581 | 0.00952274 |
| 13 | rs79464516 | 2 | 112757446 | 0.0109377 |

| | | | | |
|---|---|---|---|---|
| 13 | rs4521048 | 2 | 112757746 | 0.00952274 |
| 13 | rs79058693 | 2 | 112758559 | 0.00722854 |
| 13 | rs4528767 | 2 | 112759864 | 0.0109377 |
| 13 | rs4278932 | 2 | 112761310 | 0.00829476 |
| 13 | rs55766181 | 2 | 112766304 | 0.00952274 |
| 13 | rs55799805 | 2 | 112766319 | 0.00952274 |
| 13 | rs11884641 | 2 | 112767406 | 0.0144499 |
| 13 | rs11884693 | 2 | 112767483 | 0.0144499 |
| 13 | rs75313362 | 2 | 112767739 | 0.00722854 |
| 13 | rs77781750 | 2 | 112767824 | 0.0125688 |
| 13 | rs77306155 | 2 | 112767941 | 0.0125688 |
| 13 | rs7563113 | 2 | 112768177 | 0.0125688 |
| 13 | rs12469210 | 2 | 112768991 | 0.0125688 |
| 13 | rs11887652 | 2 | 112769339 | 0.0090584 |
| 13 | rs28377357 | 2 | 112769721 | 0.00622263 |
| 13 | rs78116208 | 2 | 112769801 | 0.0109377 |
| 13 | rs57116599 | 2 | 112770799 | 0.148086 |
| 13 | rs6712080 | 2 | 112777441 | 0.00630235 |
| 13 | rs55792453 | 2 | 112778389 | 0.00549743 |
| 13 | rs6737989 | 2 | 112779449 | 0.00829476 |
| 13 | rs55812028 | 2 | 112780969 | 0.0109377 |
| 14 | rs1918878 | 2 | 145735762 | 0.00555562 |
| 14 | rs1980383 | 2 | 145746350 | 0.00633181 |
| 14 | rs1980382 | 2 | 145746593 | 0.00487761 |
| 14 | rs1528189 | 2 | 145749279 | 0.00487761 |
| 14 | rs1528188 | 2 | 145749357 | 0.00487761 |
| 14 | rs1006923 | 2 | 145775399 | 0.0144263 |
| 14 | rs12477111 | 2 | 145786540 | 0.00613268 |
| 14 | rs12992411 | 2 | 145796032 | 0.00902083 |
| 14 | rs6430076 | 2 | 145797185 | 0.0069704 |
| 14 | rs13389190 | 2 | 145797389 | 0.00792726 |
| 14 | rs13428423 | 2 | 145797575 | 0.00902083 |
| 14 | rs2252654 | 2 | 145801113 | 0.0315131 |
| 14 | rs2252383 | 2 | 145803003 | 0.0102714 |
| 14 | rs12476764 | 2 | 145805218 | 0.00983571 |
| 14 | rs78809153 | 2 | 145805251 | 0.0276261 |
| 14 | rs13384504 | 2 | 145808513 | 0.0276261 |
| 14 | rs1852686 | 2 | 145809250 | 0.00983571 |
| 14 | rs12474485 | 2 | 145810833 | 0.00943812 |
| 14 | rs1819055 | 2 | 145810894 | 0.0410743 |
| 14 | rs12995180 | 2 | 145814198 | 0.0410743 |
| 14 | rs7607110 | 2 | 145815262 | 0.0613808 |
| 14 | rs7423637 | 2 | 145818064 | 0.0641356 |
| 14 | rs1830318 | 2 | 145819803 | 0.029646 |
| 14 | rs6747171 | 2 | 145819888 | 0.121261 |
| 14 | rs6747275 | 2 | 145819922 | 0.0339367 |
| 14 | rs35462154 | 2 | 145821133 | 0.0339367 |
| 14 | rs1881409 | 2 | 145821520 | 0.0388715 |
| 14 | rs1881410 | 2 | 145821717 | 0.0339367 |
| 14 | rs1830320 | 2 | 145822380 | 0.0293684 |
| 14 | rs13015800 | 2 | 145822655 | 0.00866658 |
| 14 | rs12691704 | 2 | 145822778 | 0.00866658 |

| | | | | |
|---|---|---|---|---|
| 14 | rs6749506 | 2 | 145825390 | 0.0334065 |
| 14 | rs6721988 | 2 | 145825928 | 0.0111689 |
| 14 | rs10928241 | 2 | 145831428 | 0.0293684 |
| 14 | rs957293 | 2 | 145833187 | 0.0293684 |
| 14 | rs10206585 | 2 | 145836055 | 0.0227344 |
| 14 | rs7604735 | 2 | 145839655 | 0.0380203 |
| 14 | rs34372836 | 2 | 145840281 | 0.0380203 |
| 15 | rs12464617 | 2 | 148576646 | 0.00753912 |
| 15 | rs1424944 | 2 | 148578930 | 0.00753912 |
| 15 | rs11889608 | 2 | 148579233 | 0.00753912 |
| 15 | rs6760703 | 2 | 148583330 | 0.00852953 |
| 15 | rs13013867 | 2 | 148584812 | 0.00965527 |
| 15 | rs13025589 | 2 | 148586061 | 0.00852953 |
| 15 | rs13026220 | 2 | 148586459 | 0.0020098 |
| 15 | rs13025219 | 2 | 148589984 | 0.0114683 |
| 15 | rs13004451 | 2 | 148593593 | 0.0114683 |
| 15 | rs12469504 | 2 | 148596102 | 0.0109355 |
| 15 | rs1424954 | 2 | 148600794 | 0.00753912 |
| 15 | rs12053113 | 2 | 148601660 | 0.0130375 |
| 15 | rs13028411 | 2 | 148609135 | 0.0114683 |
| 15 | rs1014064 | 2 | 148612154 | 0.00666731 |
| 15 | rs10803523 | 2 | 148615831 | 0.0100937 |
| 15 | rs2113793 | 2 | 148623453 | 0.0058995 |
| 15 | rs13000597 | 2 | 148626140 | 0.0100937 |
| 15 | rs929939 | 2 | 148627528 | 0.00888885 |
| 15 | rs1895693 | 2 | 148636993 | 0.0114683 |
| 15 | rs11904758 | 2 | 148640108 | 0.00666731 |
| 15 | rs7559714 | 2 | 148640768 | 0.0118069 |
| 15 | rs7559925 | 2 | 148640930 | 0.0020098 |
| 15 | rs6711673 | 2 | 148643259 | 0.00753912 |
| 15 | rs13033696 | 2 | 148645327 | 0.00753912 |
| 15 | rs1469210 | 2 | 148646742 | 0.00753912 |
| 15 | rs2161984 | 2 | 148648486 | 0.00753912 |
| 15 | rs2161983 | 2 | 148649386 | 0.0114683 |
| 15 | rs13019809 | 2 | 148650473 | 0.0114683 |
| 15 | rs1128919 | 2 | 148657117 | 0.00965527 |
| 15 | rs723681 | 2 | 148668509 | 0.0020098 |
| 15 | rs3768689 | 2 | 148669774 | 0.00753912 |
| 15 | rs3768687 | 2 | 148672020 | 0.00753912 |
| 15 | rs13026650 | 2 | 148674201 | 0.00852953 |
| 15 | rs3764955 | 2 | 148674797 | 0.0020098 |
| 15 | rs2303392 | 2 | 148680427 | 0.0100937 |
| 15 | rs2059422 | 2 | 148690055 | 0.0148298 |
| 15 | rs1345994 | 2 | 148695126 | 0.00753912 |
| 15 | rs13033589 | 2 | 148702999 | 0.0130375 |
| 15 | rs1531032 | 2 | 148703310 | 0.00852953 |
| 15 | rs10460259 | 2 | 148704721 | 0.00852953 |
| 15 | rs1030317 | 2 | 148704932 | 0.0130375 |
| 15 | rs13006184 | 2 | 148706370 | 0.00226035 |
| 15 | rs13012311 | 2 | 148707032 | 0.0168781 |
| 15 | rs13027200 | 2 | 148709653 | 0.0020098 |
| 15 | rs12104822 | 2 | 148710361 | 0.0148298 |

| | | | | |
|---|---|---|---|---|
| 15 | rs13034793 | 2 | 148710806 | 0.0130375 |
| 15 | rs11888152 | 2 | 148715038 | 0.00753912 |
| 15 | rs12463798 | 2 | 148716099 | 0.0130375 |
| 15 | rs2307394 | 2 | 148716428 | 0.0148298 |
| 15 | rs4972317 | 2 | 148718920 | 0.0130375 |
| 15 | rs34539489 | 2 | 148722877 | 0.00852953 |
| 15 | rs6729465 | 2 | 148723126 | 0.00852953 |
| 15 | rs11901963 | 2 | 148726771 | 0.0130375 |
| 15 | rs12469939 | 2 | 148732099 | 0.00226035 |
| 15 | rs1020653 | 2 | 148732410 | 0.0130375 |
| 15 | rs13027706 | 2 | 148732703 | 0.00852953 |
| 15 | rs57679331 | 2 | 148737325 | 0.00753912 |
| 15 | rs12990262 | 2 | 148737829 | 0.0148298 |
| 15 | rs7594075 | 2 | 148739823 | 0.00852953 |
| 15 | rs4972318 | 2 | 148740587 | 0.0130375 |
| 15 | rs13004041 | 2 | 148741621 | 0.00226035 |
| 15 | rs13031064 | 2 | 148741978 | 0.00852953 |
| 15 | rs13003171 | 2 | 148748577 | 0.0020098 |
| 15 | rs72855232 | 2 | 148748967 | 0.00254352 |
| 15 | rs9789434 | 2 | 148749393 | 0.0114683 |
| 15 | rs12463554 | 2 | 148751612 | 0.00226035 |
| 15 | rs9789640 | 2 | 148753501 | 0.0114683 |
| 15 | rs13008838 | 2 | 148754825 | 0.0130375 |
| 15 | rs13035475 | 2 | 148754862 | 0.0130375 |
| 15 | rs13014936 | 2 | 148755237 | 0.0130375 |
| 15 | rs13022962 | 2 | 148756624 | 0.00783227 |
| 15 | rs12053401 | 2 | 148756668 | 0.0130375 |
| 15 | rs13028348 | 2 | 148757144 | 0.0114683 |
| 15 | rs1598207 | 2 | 148757870 | 0.00852953 |
| 15 | rs12476827 | 2 | 148759348 | 0.00254352 |
| 15 | rs7425436 | 2 | 148759656 | 0.00462645 |
| 15 | rs6729803 | 2 | 148760626 | 0.00249061 |
| 15 | rs6729804 | 2 | 148760627 | 0.00249061 |
| 15 | rs6430281 | 2 | 148762568 | 0.00965527 |
| 15 | rs6430282 | 2 | 148762793 | 0.0168781 |
| 15 | rs6724719 | 2 | 148764890 | 0.00965527 |
| 15 | rs2382201 | 2 | 148765496 | 0.00254352 |
| 15 | rs6732463 | 2 | 148766475 | 0.00965527 |
| 15 | rs12997165 | 2 | 148768873 | 0.0168781 |
| 15 | rs13032810 | 2 | 148770054 | 0.0148298 |
| 15 | rs13033664 | 2 | 148770274 | 0.00322594 |
| 15 | rs6746415 | 2 | 148771691 | 0.00965527 |
| 15 | rs2168145 | 2 | 148773395 | 0.0168781 |
| 15 | rs12989250 | 2 | 148776438 | 0.00753912 |
| 15 | rs1975748 | 2 | 148776859 | 0.0148298 |
| 15 | rs897172 | 2 | 148778272 | 0.0148298 |
| 15 | rs1015096 | 2 | 148782358 | 0.0168781 |
| 15 | rs11886876 | 2 | 148783807 | 0.0109355 |
| 15 | rs13007770 | 2 | 148784324 | 0.0148298 |
| 15 | rs4972320 | 2 | 148784570 | 0.00226035 |
| 15 | rs4972321 | 2 | 148784984 | 0.0109355 |
| 15 | rs7560425 | 2 | 148785312 | 0.00852953 |

| | | | | |
|---|---|---|---|---|
| 15 | rs13021605 | 2 | 148786336 | 0.00852953 |
| 15 | rs13029038 | 2 | 148787308 | 0.00254352 |
| 15 | rs13012455 | 2 | 148788236 | 0.00254352 |
| 15 | rs4417662 | 2 | 148789386 | 0.00852953 |
| 15 | rs2890915 | 2 | 148789543 | 0.00852953 |
| 15 | rs11896010 | 2 | 148789730 | 0.00852953 |
| 15 | rs12992412 | 2 | 148792665 | 0.00179818 |
| 15 | rs12473564 | 2 | 148793730 | 0.00254352 |
| 15 | rs12992231 | 2 | 148799710 | 0.00179818 |
| 15 | rs12105411 | 2 | 148800730 | 0.00203292 |
| 15 | rs12995413 | 2 | 148801652 | 0.00203292 |
| 15 | rs10186643 | 2 | 148803587 | 0.00290452 |
| 15 | rs13032786 | 2 | 148803672 | 0.00226035 |
| 16 | rs56297666 | 2 | 160902023 | 0.00551868 |
| 16 | rs34248629 | 2 | 160902497 | 0.00551868 |
| 16 | rs877200 | 2 | 160903723 | 0.00646887 |
| 16 | rs2175416 | 2 | 160904021 | 0.00646887 |
| 16 | rs1511218 | 2 | 160906390 | 0.00646887 |
| 16 | rs10929965 | 2 | 160906931 | 0.00758738 |
| 16 | rs6719686 | 2 | 160908292 | 0.00758738 |
| 16 | rs10929966 | 2 | 160908341 | 0.00758738 |
| 16 | rs17831161 | 2 | 160909360 | 0.00646887 |
| 16 | rs62175515 | 2 | 160909524 | 0.00758738 |
| 16 | rs7563607 | 2 | 160910215 | 0.00758738 |
| 16 | rs56293836 | 2 | 160910958 | 0.00646887 |
| 16 | rs17831191 | 2 | 160911332 | 0.00646887 |
| 16 | rs62175517 | 2 | 160911575 | 0.00646887 |
| 16 | rs925410 | 2 | 160912652 | 0.00646887 |
| 16 | rs6759924 | 2 | 160912747 | 0.004711 |
| 16 | rs6759836 | 2 | 160912802 | 0.00551868 |
| 16 | rs17831251 | 2 | 160914156 | 0.00646887 |
| 16 | rs6707458 | 2 | 160914540 | 0.00646887 |
| 16 | rs16844715 | 2 | 160915106 | 0.00758738 |
| 16 | rs17241792 | 2 | 160915205 | 0.004711 |
| 16 | rs17831329 | 2 | 160915819 | 0.004711 |
| 16 | rs3749119 | 2 | 160919020 | 0.0432286 |
| 16 | rs58354887 | 2 | 160927112 | 0.00551868 |
| 16 | rs6744567 | 2 | 160933781 | 0.0170011 |
| 16 | rs7593593 | 2 | 160933811 | 0.0170011 |
| 16 | rs62175552 | 2 | 160937220 | 0.0144497 |
| 16 | rs4520993 | 2 | 160937673 | 0.0200155 |
| 16 | rs4520994 | 2 | 160937750 | 0.0170011 |
| 16 | rs4292050 | 2 | 160940899 | 0.0327842 |
| 16 | rs6739867 | 2 | 160942818 | 0.0200155 |
| 16 | rs4363984 | 2 | 160943570 | 0.0200155 |
| 16 | rs4637057 | 2 | 160950161 | 0.0235792 |
| 16 | rs62175559 | 2 | 160955833 | 0.0235792 |
| 16 | rs4233681 | 2 | 160960058 | 0.0860257 |
| 16 | rs62177860 | 2 | 160960571 | 0.0277946 |
| 16 | rs12987602 | 2 | 160960788 | 0.0860257 |
| 16 | rs17216980 | 2 | 160960932 | 0.0144497 |
| 16 | rs10497213 | 2 | 160963603 | 0.0364561 |

| | | | | |
|---|---|---|---|---|
| 16 | rs13009231 | 2 | 160964057 | 0.00551868 |
| 16 | rs6731260 | 2 | 160977769 | 0.0386935 |
| 16 | rs2124969 | 2 | 160989486 | 0.0755519 |
| 16 | rs7567781 | 2 | 160999537 | 0.0755519 |
| 16 | rs60908067 | 2 | 161004983 | 0.0638538 |
| 16 | rs13001911 | 2 | 161020703 | 0.0104576 |
| 16 | rs7589820 | 2 | 161027884 | 0.00551868 |
| 16 | rs17218566 | 2 | 161054748 | 0.00890486 |
| 16 | rs4665162 | 2 | 161060184 | 0.00646887 |
| 16 | rs59019419 | 2 | 161061287 | 0.00890486 |
| 16 | rs1870102 | 2 | 161070582 | 0.00646887 |
| 17 | rs6433395 | 2 | 173943880 | 0.0407086 |
| 17 | rs67719248 | 2 | 173948094 | 0.00831088 |
| 17 | rs72905046 | 2 | 173948571 | 0.0112133 |
| 17 | rs3769203 | 2 | 173955062 | 0.00324889 |
| 17 | rs16861272 | 2 | 173960905 | 0.00324889 |
| 17 | rs34681024 | 2 | 173982366 | 0.00371902 |
| 17 | rs36103648 | 2 | 173985747 | 0.00351608 |
| 17 | rs34014656 | 2 | 173989306 | 0.00371902 |
| 17 | rs13020796 | 2 | 173990421 | 0.0471257 |
| 17 | rs16861278 | 2 | 173992693 | 0.0471257 |
| 17 | rs2278883 | 2 | 173996917 | 0.00425984 |
| 17 | rs67683956 | 2 | 174002739 | 0.00642558 |
| 17 | rs3769187 | 2 | 174004202 | 0.00401303 |
| 17 | rs13017210 | 2 | 174008623 | 0.6953 |
| 17 | rs17761169 | 2 | 174034099 | 0.0471257 |
| 17 | rs34658268 | 2 | 174041982 | 0.0141831 |
| 17 | rs55913048 | 2 | 174054206 | 0.00371902 |
| 17 | rs3769161 | 2 | 174066475 | 0.00425984 |
| 18 | rs847157 | 2 | 176962501 | 0.0637883 |
| 18 | rs847156 | 2 | 176962561 | 0.0637883 |
| 18 | rs847154 | 2 | 176962958 | 0.0637883 |
| 18 | rs847153 | 2 | 176962989 | 0.0637883 |
| 18 | rs711812 | 2 | 176964304 | 0.0779652 |
| 18 | rs847151 | 2 | 176964904 | 0.0522188 |
| 18 | rs711813 | 2 | 176966190 | 0.0637883 |
| 18 | rs741610 | 2 | 176969206 | 0.0779652 |
| 18 | rs711814 | 2 | 176969341 | 0.142842 |
| 18 | rs847148 | 2 | 176970456 | 0.322776 |
| 19 | rs1515866 | 2 | 190067064 | 0.0015611 |
| 19 | rs2351631 | 2 | 190068470 | 0.00185544 |
| 19 | rs142571446 | 2 | 190068870 | 0.0180132 |
| 19 | rs10755002 | 2 | 190069176 | 0.0022078 |
| 19 | rs17270980 | 2 | 190070890 | 0.0190507 |
| 19 | rs13015183 | 2 | 190070927 | 0.0022078 |
| 19 | rs16831217 | 2 | 190070983 | 0.017034 |
| 19 | rs7599222 | 2 | 190073658 | 0.00171252 |
| 19 | rs939160 | 2 | 190074622 | 0.0022078 |
| 19 | rs6732534 | 2 | 190079156 | 0.00185544 |
| 19 | rs1913892 | 2 | 190080984 | 0.00185544 |
| 19 | rs114826780 | 2 | 190082597 | 0.0103475 |
| 19 | rs16841601 | 2 | 190082747 | 0.0144131 |

| | | | | |
|---|---|---|---|---|
| 19 | rs1356161 | 2 | 190083141 | 0.00185544 |
| 19 | rs2351633 | 2 | 190084791 | 0.0015611 |
| 19 | rs10198505 | 2 | 190104983 | 0.0045214 |
| 19 | rs4031918 | 2 | 190106007 | 0.00406882 |
| 19 | rs10164448 | 2 | 190108891 | 0.00178539 |
| 19 | rs2102874 | 2 | 190109425 | 0.0045214 |
| 19 | rs1515867 | 2 | 190111412 | 0.0045214 |
| 19 | rs2030293 | 2 | 190112458 | 0.00428893 |
| 19 | rs870922 | 2 | 190113708 | 0.00329896 |
| 19 | rs870921 | 2 | 190113745 | 0.00956683 |
| 19 | rs10204942 | 2 | 190115961 | 0.00804497 |
| 19 | rs7596033 | 2 | 190118235 | 0.00366299 |
| 19 | rs7599676 | 2 | 190119066 | 0.00174536 |
| 19 | rs7591652 | 2 | 190122574 | 0.00476696 |
| 19 | rs4666773 | 2 | 190123256 | 0.0045214 |
| 19 | rs1464262 | 2 | 190127123 | 0.00476696 |
| 19 | rs9973614 | 2 | 190129604 | 0.00858516 |
| 19 | rs1606931 | 2 | 190130694 | 0.00366299 |
| 19 | rs1850122 | 2 | 190130976 | 0.0209685 |
| 19 | rs6434335 | 2 | 190131182 | 0.00813399 |
| 19 | rs1473999 | 2 | 190132854 | 0.00813399 |
| 19 | rs10755003 | 2 | 190133932 | 0.00476696 |
| 19 | rs10931409 | 2 | 190134275 | 0.0045214 |
| 19 | rs4667275 | 2 | 190138684 | 0.0021464 |
| 19 | rs76287931 | 2 | 190141143 | 0.0129005 |
| 19 | rs78156759 | 2 | 190141718 | 0.0225493 |
| 19 | rs2351634 | 2 | 190144159 | 0.0220241 |
| 19 | rs75897057 | 2 | 190144880 | 0.0085004 |
| 19 | rs73982009 | 2 | 190145173 | 0.0125345 |
| 19 | rs16831319 | 2 | 190150205 | 0.0434912 |
| 19 | rs939157 | 2 | 190151140 | 0.0225493 |
| 19 | rs939159 | 2 | 190151221 | 0.0220241 |
| 19 | rs4494782 | 2 | 190157357 | 0.00804497 |
| 19 | rs148113645 | 2 | 190158743 | 0.00316772 |
| 19 | rs58288282 | 2 | 190178034 | 0.00492407 |
| 19 | rs2176649 | 2 | 190178839 | 0.0106059 |
| 19 | rs2680630 | 2 | 190187457 | 0.00442021 |
| 19 | rs2683019 | 2 | 190191290 | 0.00320541 |
| 19 | rs2683011 | 2 | 190197517 | 0.00646126 |
| 19 | rs56920210 | 2 | 190203950 | 0.00579408 |
| 19 | rs77321529 | 2 | 190204800 | 0.0043534 |
| 19 | rs73978612 | 2 | 190210152 | 0.00720826 |
| 19 | rs79118748 | 2 | 190215468 | 0.00979496 |
| 19 | rs78861577 | 2 | 190215681 | 0.0125345 |
| 19 | rs77448694 | 2 | 190216205 | 0.0109323 |
| 19 | rs114022205 | 2 | 190223717 | 0.00979496 |
| 19 | rs73978617 | 2 | 190231883 | 0.0118543 |
| 19 | rs78631812 | 2 | 190234621 | 0.0175566 |
| 19 | rs79903972 | 2 | 190250925 | 0.015237 |
| 19 | rs7556928 | 2 | 190251709 | 0.00290932 |
| 19 | rs58796942 | 2 | 190252289 | 0.0140186 |
| 19 | rs16831356 | 2 | 190257103 | 0.0208073 |

| | | | | |
|---|---|---|---|---|
| 19 | rs13405433 | 2 | 190260299 | 0.00324204 |
| 19 | rs149780113 | 2 | 190260412 | 0.0103475 |
| 19 | rs28545536 | 2 | 190260537 | 0.00324204 |
| 19 | rs13035381 | 2 | 190265596 | 0.00269544 |
| 19 | rs1520858 | 2 | 190271533 | 0.00442021 |
| 19 | rs77945618 | 2 | 190276153 | 0.00407461 |
| 19 | rs75399423 | 2 | 190276958 | 0.00407461 |
| 19 | rs1520856 | 2 | 190276973 | 0.00269544 |
| 19 | rs73978620 | 2 | 190278087 | 0.0208073 |
| 19 | rs74704182 | 2 | 190280320 | 0.0267149 |
| 19 | rs75971232 | 2 | 190284274 | 0.0220241 |
| 19 | rs1851104 | 2 | 190294549 | 0.0021464 |
| 19 | rs7563184 | 2 | 190296131 | 0.00231521 |
| 19 | rs115233151 | 2 | 190296153 | 0.0282737 |
| 19 | rs74542138 | 2 | 190308506 | 0.00502634 |
| 19 | rs115256769 | 2 | 190310326 | 0.0355091 |
| 19 | rs73978626 | 2 | 190310485 | 0.0252446 |
| 19 | rs1520854 | 2 | 190310982 | 0.00499508 |
| 19 | rs6738193 | 2 | 190313396 | 0.00499508 |
| 19 | rs1992295 | 2 | 190313841 | 0.00462089 |
| 19 | rs12693539 | 2 | 190318009 | 0.00427565 |
| 19 | rs13015207 | 2 | 190320810 | 0.00462089 |
| 19 | rs73978632 | 2 | 190321347 | 0.0238577 |
| 19 | rs7574403 | 2 | 190321663 | 0.00462089 |
| 19 | rs76926223 | 2 | 190322950 | 0.00621914 |
| 19 | rs2351933 | 2 | 190323177 | 0.00514194 |
| 19 | rs73978633 | 2 | 190325150 | 0.0282737 |
| 19 | rs2304702 | 2 | 190327126 | 0.00427565 |
| 19 | rs16831454 | 2 | 190329447 | 0.00395705 |
| 19 | rs6434343 | 2 | 190332997 | 0.00395705 |
| 19 | rs10166420 | 2 | 190336705 | 0.00339152 |
| 19 | rs6747222 | 2 | 190340453 | 0.00314084 |
| 19 | rs734859 | 2 | 190342069 | 0.00290932 |
| 19 | rs114473932 | 2 | 190345807 | 0.00361577 |
| 19 | rs11678414 | 2 | 190347630 | 0.00269544 |
| 19 | rs893408 | 2 | 190350646 | 0.00176708 |
| 19 | rs744556 | 2 | 190351066 | 0.00176708 |
| 19 | rs12623394 | 2 | 190352703 | 0.00193919 |
| 19 | rs1595749 | 2 | 190353770 | 0.00184601 |
| 19 | rs143030753 | 2 | 190354297 | 0.0122401 |
| 19 | rs12988143 | 2 | 190354514 | 0.0021385 |
| 19 | rs2351934 | 2 | 190354798 | 0.00184601 |
| 19 | rs2119069 | 2 | 190354871 | 0.00242485 |
| 19 | rs10497703 | 2 | 190355355 | 0.00188709 |
| 19 | rs80077412 | 2 | 190355887 | 0.0200049 |
| 19 | rs1866605 | 2 | 190356197 | 0.0021464 |
| 19 | rs4309611 | 2 | 190357006 | 0.00158902 |
| 19 | rs10198663 | 2 | 190357442 | 0.00249783 |
| 19 | rs35997606 | 2 | 190358689 | 0.00185927 |
| 20 | rs2043989 | 2 | 191268293 | 0.00941032 |
| 20 | rs6707846 | 2 | 191286516 | 0.00748167 |
| 20 | rs4853509 | 2 | 191330786 | 0.0158579 |

| | | | | |
|---|---|---|---|---|
| 20 | rs4264585 | 2 | 191334031 | 0.00540718 |
| 20 | rs7601745 | 2 | 191341366 | 0.00540718 |
| 20 | rs12613365 | 2 | 191347310 | 0.00483324 |
| 20 | rs6711006 | 2 | 191371217 | 0.00540718 |
| 20 | rs4586658 | 2 | 191379268 | 0.0107117 |
| 20 | rs13019278 | 2 | 191384924 | 0.00540718 |
| 20 | rs7584794 | 2 | 191386158 | 0.0107117 |
| 20 | rs7585173 | 2 | 191386594 | 0.0107117 |
| 20 | rs10165897 | 2 | 191387612 | 0.0122007 |
| 20 | rs4500977 | 2 | 191393787 | 0.00727629 |
| 20 | rs4073873 | 2 | 191394583 | 0.00759365 |
| 20 | rs4146923 | 2 | 191394715 | 0.00727629 |
| 20 | rs12619560 | 2 | 191395646 | 0.00851215 |
| 20 | rs12693578 | 2 | 191396188 | 0.00827222 |
| 20 | rs12622496 | 2 | 191399013 | 0.0532165 |
| 20 | rs12619125 | 2 | 191399201 | 0.0532165 |
| 20 | rs55754136 | 2 | 191401425 | 0.0472374 |
| 20 | rs4853718 | 2 | 191404964 | 0.0599807 |
| 20 | rs55987230 | 2 | 191409274 | 0.0599807 |
| 20 | rs34274882 | 2 | 191410795 | 0.0472374 |
| 20 | rs34825614 | 2 | 191413863 | 0.0532165 |
| 20 | rs13013848 | 2 | 191419442 | 0.0532165 |
| 20 | rs12464923 | 2 | 191421830 | 0.0599807 |
| 20 | rs36055519 | 2 | 191429380 | 0.0599807 |
| 20 | rs12478437 | 2 | 191432415 | 0.0269913 |
| 20 | rs34417560 | 2 | 191435716 | 0.0676366 |
| 20 | rs12615925 | 2 | 191437902 | 0.0331311 |
| 20 | rs6434400 | 2 | 191442468 | 0.0331311 |
| 20 | rs7608180 | 2 | 191443403 | 0.0207828 |
| 20 | rs12987797 | 2 | 191454544 | 0.0207828 |
| 20 | rs4853720 | 2 | 191463602 | 0.0185176 |
| 20 | rs12621493 | 2 | 191467149 | 0.0178934 |
| 20 | rs13028201 | 2 | 191468706 | 0.0233362 |
| 21 | rs829186 | 3 | 14818716 | 0.0111679 |
| 21 | rs829187 | 3 | 14818787 | 0.0128268 |
| 21 | rs829189 | 3 | 14821017 | 0.00763229 |
| 21 | rs829190 | 3 | 14821162 | 0.00894636 |
| 21 | rs1687287 | 3 | 14828111 | 0.0265549 |
| 21 | rs1687289 | 3 | 14829826 | 0.0571806 |
| 21 | rs294601 | 3 | 14832407 | 0.0524582 |
| 21 | rs1689549 | 3 | 14837473 | 0.369695 |
| 21 | rs729157 | 3 | 14839035 | 0.00505489 |
| 21 | rs192676 | 3 | 14841602 | 0.111471 |
| 21 | rs190902 | 3 | 14842095 | 0.139562 |
| 21 | rs1159749 | 3 | 14849002 | 0.132866 |
| 21 | rs10510432 | 3 | 14865367 | 0.00712073 |
| 21 | rs60388387 | 3 | 14866331 | 0.00779308 |
| 22 | rs1529544 | 3 | 53039455 | 0.0264952 |
| 22 | rs4687694 | 3 | 53049047 | 0.0163242 |
| 22 | rs2581778 | 3 | 53055311 | 0.0163242 |
| 22 | rs2244552 | 3 | 53055522 | 0.0163242 |
| 22 | rs9847710 | 3 | 53062661 | 0.309812 |

| | | | | |
|---|---|---|---|---|
| 22 | rs11709427 | 3 | 53068042 | 0.0703217 |
| 22 | rs2564956 | 3 | 53070462 | 0.0898973 |
| 22 | rs2581817 | 3 | 53071797 | 0.0898973 |
| 22 | rs6808387 | 3 | 53079732 | 0.114994 |
| 22 | rs12488768 | 3 | 53086234 | 0.0337864 |
| 22 | rs13088455 | 3 | 53087482 | 0.055043 |
| 22 | rs9831861 | 3 | 53088285 | 0.055043 |
| 22 | rs4687697 | 3 | 53090490 | 0.0264952 |
| 22 | rs6445558 | 3 | 53097660 | 0.0207905 |
| 22 | rs6770152 | 3 | 53100214 | 0.0207905 |
| 23 | rs34440469 | 3 | 70897294 | 0.00521651 |
| 23 | rs1522551 | 3 | 70902653 | 0.0461583 |
| 23 | rs2687195 | 3 | 70905581 | 0.00624632 |
| 23 | rs1522552 | 3 | 70907252 | 0.119338 |
| 23 | rs2699491 | 3 | 70909039 | 0.034725 |
| 23 | rs2597301 | 3 | 70909494 | 0.170677 |
| 23 | rs11926363 | 3 | 70913181 | 0.0103884 |
| 23 | rs1522554 | 3 | 70913293 | 0.0245285 |
| 23 | rs35270590 | 3 | 70914733 | 0.00521651 |
| 23 | rs35109540 | 3 | 70914736 | 0.00442777 |
| 23 | rs34341381 | 3 | 70914738 | 0.00442777 |
| 23 | rs2597302 | 3 | 70917189 | 0.00725225 |
| 23 | rs4676893 | 3 | 70917639 | 0.0492714 |
| 23 | rs1018340 | 3 | 70918127 | 0.012322 |
| 23 | rs1403202 | 3 | 70918938 | 0.012322 |
| 23 | rs12054322 | 3 | 70919162 | 0.0146238 |
| 23 | rs17007949 | 3 | 70920041 | 0.0146238 |
| 23 | rs56254492 | 3 | 70920168 | 0.0119363 |
| 23 | rs4499560 | 3 | 70920485 | 0.0245285 |
| 23 | rs7626449 | 3 | 70921329 | 0.0141082 |
| 23 | rs2687197 | 3 | 70922320 | 0.0119363 |
| 23 | rs6776545 | 3 | 70923584 | 0.00376032 |
| 23 | rs2687199 | 3 | 70924971 | 0.012322 |
| 23 | rs13068175 | 3 | 70925604 | 0.0547934 |
| 23 | rs1533834 | 3 | 70926988 | 0.0233714 |
| 23 | rs34518676 | 3 | 70927382 | 0.0197415 |
| 23 | rs2687201 | 3 | 70928930 | 0.0245285 |
| 23 | rs2687202 | 3 | 70929983 | 0.00855797 |
| 23 | rs34577575 | 3 | 70931081 | 0.0263313 |
| 23 | rs35378923 | 3 | 70932586 | 0.0547934 |
| 23 | rs2687203 | 3 | 70933568 | 0.119338 |
| 24 | rs6438003 | 3 | 111453889 | 0.0489309 |
| 24 | rs10212300 | 3 | 111464449 | 0.0135721 |
| 24 | rs9870823 | 3 | 111465590 | 0.00790721 |
| 24 | rs13069980 | 3 | 111466766 | 0.00790721 |
| 24 | rs13074817 | 3 | 111467155 | 0.00946149 |
| 24 | rs4284954 | 3 | 111468099 | 0.00946149 |
| 24 | rs9834030 | 3 | 111469374 | 0.00946149 |
| 24 | rs12496925 | 3 | 111470108 | 0.0113284 |
| 24 | rs12496971 | 3 | 111470259 | 0.0113284 |
| 24 | rs10934106 | 3 | 111470468 | 0.0113284 |
| 24 | rs12490595 | 3 | 111470696 | 0.00463276 |

| | | | | |
|---|---|---|---|---|
| 24 | rs13079085 | 3 | 111471217 | 0.00946149 |
| 24 | rs6764720 | 3 | 111476395 | 0.00388139 |
| 24 | rs6438008 | 3 | 111477120 | 0.00790721 |
| 24 | rs6792493 | 3 | 111477348 | 0.473398 |
| 24 | rs10934107 | 3 | 111477636 | 0.00790721 |
| 24 | rs6768713 | 3 | 111477774 | 0.00790721 |
| 24 | rs6802995 | 3 | 111480737 | 0.0113284 |
| 24 | rs4431081 | 3 | 111482048 | 0.0195171 |
| 24 | rs7427201 | 3 | 111482067 | 0.00553303 |
| 24 | rs12490166 | 3 | 111482157 | 0.00661238 |
| 24 | rs7614443 | 3 | 111482419 | 0.0113284 |
| 24 | rs9838371 | 3 | 111482802 | 0.00463276 |
| 24 | rs6786386 | 3 | 111483856 | 0.0234265 |
| 24 | rs6790072 | 3 | 111485002 | 0.00553303 |
| 24 | rs6790079 | 3 | 111485020 | 0.00661238 |
| 24 | rs6777540 | 3 | 111485349 | 0.00946149 |
| 24 | rs9863403 | 3 | 111485681 | 0.00946149 |
| 24 | rs6800922 | 3 | 111488596 | 0.00397667 |
| 24 | rs9858228 | 3 | 111491772 | 0.00397667 |
| 24 | rs9862472 | 3 | 111492054 | 0.00397667 |
| 24 | rs66786213 | 3 | 111492085 | 0.0105925 |
| 24 | rs66856712 | 3 | 111492208 | 0.00694465 |
| 24 | rs58124137 | 3 | 111492406 | 0.00694465 |
| 24 | rs73222475 | 3 | 111492710 | 0.00694465 |
| 24 | rs60867472 | 3 | 111492718 | 0.00694465 |
| 24 | rs58663746 | 3 | 111492948 | 0.00397667 |
| 24 | rs6762246 | 3 | 111493336 | 0.00525112 |
| 24 | rs6800047 | 3 | 111493648 | 0.00525112 |
| 24 | rs6800065 | 3 | 111493717 | 0.00525112 |
| 24 | rs6787278 | 3 | 111493739 | 0.0045688 |
| 24 | rs6789977 | 3 | 111493868 | 0.00603765 |
| 24 | rs6802960 | 3 | 111494319 | 0.00525112 |
| 24 | rs6790521 | 3 | 111494363 | 0.00525112 |
| 24 | rs6790355 | 3 | 111494386 | 0.00525112 |
| 24 | rs6790612 | 3 | 111494458 | 0.00525112 |
| 24 | rs6790825 | 3 | 111494663 | 0.00525112 |
| 24 | rs6790977 | 3 | 111494820 | 0.00946149 |
| 24 | rs13321524 | 3 | 111495092 | 0.00525112 |
| 24 | rs13322494 | 3 | 111495918 | 0.00525112 |
| 24 | rs28534728 | 3 | 111497463 | 0.00525112 |
| 24 | rs28648833 | 3 | 111497781 | 0.00525112 |
| 24 | rs4390898 | 3 | 111497948 | 0.00525112 |
| 24 | rs4634085 | 3 | 111498125 | 0.00525112 |
| 24 | rs28675079 | 3 | 111500002 | 0.0045688 |
| 24 | rs6762572 | 3 | 111500801 | 0.00525112 |
| 24 | rs6778818 | 3 | 111500998 | 0.00525112 |
| 24 | rs6782513 | 3 | 111502233 | 0.00525112 |
| 24 | rs9843288 | 3 | 111504556 | 0.0045688 |
| 24 | rs67970120 | 3 | 111508929 | 0.00919848 |
| 25 | rs11711620 | 3 | 114444572 | 0.00377173 |
| 25 | rs9842905 | 3 | 114498351 | 0.349488 |
| 25 | rs9853475 | 3 | 114500255 | 0.598875 |

| | | | | |
|---|---|---|---|---|
| 26 | rs4680338 | 3 | 156794425 | 0.109528 |
| 26 | rs13322435 | 3 | 156795468 | 0.0718359 |
| 26 | rs9854955 | 3 | 156795525 | 0.047227 |
| 26 | rs55730982 | 3 | 156797208 | 0.0582287 |
| 26 | rs56082403 | 3 | 156797225 | 0.0311222 |
| 26 | rs10049210 | 3 | 156797373 | 0.157888 |
| 26 | rs10049090 | 3 | 156797702 | 0.157888 |
| 26 | rs67261871 | 3 | 156797941 | 0.0227986 |
| 26 | rs1482852 | 3 | 156798294 | 0.127023 |
| 26 | rs1482853 | 3 | 156798473 | 0.0227986 |
| 26 | rs900399 | 3 | 156798732 | 0.066391 |
| 26 | rs900400 | 3 | 156798775 | 0.0823688 |
| 27 | rs6441306 | 3 | 159952278 | 0.00780863 |
| 27 | rs7624902 | 3 | 160004026 | 0.00780863 |
| 27 | rs6791261 | 3 | 160016446 | 0.00438612 |
| 27 | rs4680576 | 3 | 160021681 | 0.00506172 |
| 27 | rs4679881 | 3 | 160028014 | 0.0119325 |
| 27 | rs55687857 | 3 | 160029964 | 0.0102632 |
| 27 | rs6441313 | 3 | 160036400 | 0.0102632 |
| 27 | rs1953506 | 3 | 160036448 | 0.0102632 |
| 27 | rs58665246 | 3 | 160043456 | 0.00286455 |
| 27 | rs62272186 | 3 | 160046177 | 0.0102632 |
| 27 | rs869468 | 3 | 160047440 | 0.00506172 |
| 27 | rs6803828 | 3 | 160058562 | 0.0102632 |
| 27 | rs6792612 | 3 | 160060748 | 0.00329959 |
| 27 | rs6441314 | 3 | 160061145 | 0.0102632 |
| 27 | rs60009222 | 3 | 160063356 | 0.0102632 |
| 27 | rs11918749 | 3 | 160067930 | 0.0102632 |
| 27 | rs55731730 | 3 | 160072378 | 0.0119325 |
| 27 | rs9798898 | 3 | 160082154 | 0.00883324 |
| 27 | rs10513551 | 3 | 160086055 | 0.00883324 |
| 27 | rs7611130 | 3 | 160106781 | 0.0102632 |
| 27 | rs4680579 | 3 | 160109657 | 0.0102632 |
| 27 | rs6793560 | 3 | 160113469 | 0.0102632 |
| 27 | rs4680580 | 3 | 160115004 | 0.0121141 |
| 27 | rs7634108 | 3 | 160124695 | 0.0119325 |
| 27 | rs9968151 | 3 | 160126036 | 0.0119325 |
| 27 | rs2305407 | 3 | 160130110 | 0.0102632 |
| 27 | rs1451762 | 3 | 160131576 | 0.00438612 |
| 27 | rs1451761 | 3 | 160131667 | 0.00506172 |
| 27 | rs73154592 | 3 | 160137748 | 0.0164036 |
| 27 | rs7631792 | 3 | 160148756 | 0.010458 |
| 27 | rs13403 | 3 | 160152299 | 0.010458 |
| 27 | rs7629 | 3 | 160153305 | 0.0121141 |
| 27 | rs4679885 | 3 | 160154747 | 0.010458 |
| 27 | rs4680585 | 3 | 160167194 | 0.0188994 |
| 27 | rs2279457 | 3 | 160168292 | 0.021947 |
| 27 | rs60460891 | 3 | 160168769 | 0.0296512 |
| 27 | rs7629202 | 3 | 160169197 | 0.025502 |
| 27 | rs56394279 | 3 | 160171092 | 0.138562 |
| 27 | rs6808083 | 3 | 160171453 | 0.0545295 |
| 27 | rs6441320 | 3 | 160172274 | 0.025502 |

| | | | | |
|---|---|---|---|---|
| 27 | rs7634826 | 3 | 160179503 | 0.0401601 |
| 27 | rs4091703 | 3 | 160187959 | 0.0467819 |
| 27 | rs4680587 | 3 | 160188242 | 0.0545295 |
| 27 | rs3851366 | 3 | 160191374 | 0.0742253 |
| 27 | rs4680588 | 3 | 160192518 | 0.0401601 |
| 27 | rs6798668 | 3 | 160200408 | 0.00380307 |
| 27 | rs4679888 | 3 | 160211871 | 0.00506172 |
| 27 | rs3773372 | 3 | 160232432 | 0.00248842 |
| 27 | rs62272791 | 3 | 160241657 | 0.00438612 |
| 27 | rs62272792 | 3 | 160241674 | 0.00438612 |
| 27 | rs62272794 | 3 | 160250880 | 0.00329959 |
| 27 | rs7640072 | 3 | 160257386 | 0.00329959 |
| 27 | rs56197102 | 3 | 160258281 | 0.00216303 |
| 27 | rs62272800 | 3 | 160263384 | 0.00286455 |
| 27 | rs2366969 | 3 | 160273758 | 0.00329959 |
| 27 | rs1882126 | 3 | 160284194 | 0.00329959 |
| 27 | rs3732707 | 3 | 160286646 | 0.00286455 |
| 27 | rs62272810 | 3 | 160290299 | 0.00506172 |
| 27 | rs4292274 | 3 | 160291468 | 0.00380307 |
| 27 | rs4679896 | 3 | 160291866 | 0.00329959 |
| 27 | rs4679655 | 3 | 160292900 | 0.00286455 |
| 27 | rs17570467 | 3 | 160294561 | 0.00380307 |
| 27 | rs4679899 | 3 | 160301365 | 0.00286455 |
| 27 | rs4679900 | 3 | 160301772 | 0.00506172 |
| 27 | rs1920662 | 3 | 160303591 | 0.00216303 |
| 27 | rs6806505 | 3 | 160305648 | 0.00329959 |
| 28 | rs2279193 | 4 | 8357835 | 0.00693859 |
| 28 | rs10938709 | 4 | 8380329 | 0.0333178 |
| 28 | rs2386223 | 4 | 8398125 | 0.0161764 |
| 28 | rs3796740 | 4 | 8401124 | 0.00414019 |
| 28 | rs12503034 | 4 | 8403089 | 0.00917748 |
| 28 | rs1880025 | 4 | 8404346 | 0.00797741 |
| 28 | rs2631752 | 4 | 8405314 | 0.456529 |
| 28 | rs2245515 | 4 | 8415982 | 0.00399594 |
| 28 | rs2245808 | 4 | 8418430 | 0.00265905 |
| 28 | rs2245809 | 4 | 8418440 | 0.00869527 |
| 28 | rs2049302 | 4 | 8418605 | 0.0596051 |
| 28 | rs2631766 | 4 | 8418779 | 0.0287989 |
| 28 | rs2631765 | 4 | 8418997 | 0.0694363 |
| 28 | rs2631763 | 4 | 8422987 | 0.0596051 |
| 28 | rs2140336 | 4 | 8424831 | 0.0596051 |
| 28 | rs2688244 | 4 | 8437033 | 0.0694363 |
| 28 | rs2631728 | 4 | 8438534 | 0.00735125 |
| 28 | rs2688246 | 4 | 8441264 | 0.0100434 |
| 28 | rs2631727 | 4 | 8443776 | 0.00348645 |
| 28 | rs56343159 | 4 | 8446263 | 0.00265905 |
| 28 | rs2688234 | 4 | 8447205 | 0.00304382 |
| 28 | rs11729265 | 4 | 8455011 | 0.00304382 |
| 28 | rs2631769 | 4 | 8466866 | 0.00304382 |
| 28 | rs2631737 | 4 | 8479418 | 0.00693859 |
| 28 | rs2688221 | 4 | 8479448 | 0.005259 |
| 28 | rs2631757 | 4 | 8493651 | 0.00399594 |

| | | | | |
|---|---|---|---|---|
| 28 | rs4325994 | 4 | 8494292 | 0.00265905 |
| 28 | rs3115396 | 4 | 8510952 | 0.00328446 |
| 29 | rs12507413 | 4 | 39441490 | 0.121655 |
| 29 | rs6835490 | 4 | 39444337 | 0.00592364 |
| 29 | rs1048140 | 4 | 39478832 | 0.0358467 |
| 29 | rs2687965 | 4 | 39491521 | 0.50179 |
| 29 | rs6858752 | 4 | 39493141 | 0.0923214 |
| 29 | rs6531721 | 4 | 39497391 | 0.0468253 |
| 29 | rs1450 | 4 | 39500514 | 0.0468253 |
| 29 | rs58958665 | 4 | 39501549 | 0.0702361 |
| 29 | rs13129975 | 4 | 39526641 | 0.00978489 |
| 29 | rs13146483 | 4 | 39528939 | 0.00760379 |
| 29 | rs62308020 | 4 | 39533080 | 0.00760379 |
| 29 | rs12512204 | 4 | 39533948 | 0.00670925 |
| 30 | rs11942410 | 4 | 75656079 | 0.0433275 |
| 30 | rs7440630 | 4 | 75657434 | 0.0226323 |
| 30 | rs4299672 | 4 | 75657662 | 0.0274761 |
| 30 | rs72659187 | 4 | 75660896 | 0.0242708 |
| 30 | rs62316262 | 4 | 75660946 | 0.0274761 |
| 30 | rs4535401 | 4 | 75661825 | 0.0274761 |
| 30 | rs59178205 | 4 | 75662446 | 0.0274761 |
| 30 | rs62316264 | 4 | 75662524 | 0.0274761 |
| 30 | rs62316265 | 4 | 75662635 | 0.0274761 |
| 30 | rs62316266 | 4 | 75662692 | 0.0214499 |
| 30 | rs62316267 | 4 | 75663035 | 0.0242708 |
| 30 | rs62316268 | 4 | 75663155 | 0.0352648 |
| 30 | rs62316269 | 4 | 75663157 | 0.0352648 |
| 30 | rs62316270 | 4 | 75663307 | 0.0242708 |
| 30 | rs62316271 | 4 | 75663588 | 0.0242708 |
| 30 | rs12503079 | 4 | 75664504 | 0.0189663 |
| 30 | rs12505535 | 4 | 75664544 | 0.0242708 |
| 30 | rs57917278 | 4 | 75664936 | 0.0214499 |
| 30 | rs62316273 | 4 | 75665824 | 0.0214499 |
| 30 | rs72659189 | 4 | 75666083 | 0.0214499 |
| 30 | rs62316274 | 4 | 75666318 | 0.0214499 |
| 30 | rs11946159 | 4 | 75666873 | 0.0214499 |
| 30 | rs11933389 | 4 | 75666880 | 0.0214499 |
| 30 | rs4426860 | 4 | 75667907 | 0.0214499 |
| 30 | rs4345250 | 4 | 75667920 | 0.0214499 |
| 30 | rs62316275 | 4 | 75668103 | 0.0214499 |
| 30 | rs4496650 | 4 | 75668955 | 0.0214499 |
| 30 | rs72659193 | 4 | 75670109 | 0.0214499 |
| 30 | rs72659195 | 4 | 75670183 | 0.0214499 |
| 30 | rs72659197 | 4 | 75670402 | 0.0214499 |
| 30 | rs72659198 | 4 | 75670976 | 0.0214499 |
| 30 | rs62316277 | 4 | 75673533 | 0.00915839 |
| 30 | rs62316278 | 4 | 75673724 | 0.00915839 |
| 30 | rs62316279 | 4 | 75675110 | 0.00915839 |
| 30 | rs62316280 | 4 | 75675218 | 0.0103271 |
| 30 | rs35926103 | 4 | 75675706 | 0.0103271 |
| 30 | rs11938276 | 4 | 75676256 | 0.0103271 |
| 30 | rs62316308 | 4 | 75676337 | 0.00915839 |

| | | | | |
|---|---|---|---|---|
| 30 | rs62316309 | 4 | 75676504 | 0.0103271 |
| 30 | rs62316310 | 4 | 75676529 | 0.00915839 |
| 30 | rs62316311 | 4 | 75676572 | 0.0103271 |
| 30 | rs62316312 | 4 | 75676596 | 0.0103271 |
| 30 | rs62316313 | 4 | 75679954 | 0.0157841 |
| 30 | rs4352548 | 4 | 75683594 | 0.0126806 |
| 30 | rs6840529 | 4 | 75686605 | 0.0343925 |
| 30 | rs6811334 | 4 | 75686640 | 0.0205265 |
| 31 | rs28676957 | 4 | 95903741 | 0.0280904 |
| 31 | rs3775031 | 4 | 95905449 | 0.017871 |
| 31 | rs6532522 | 4 | 95911022 | 0.00483638 |
| 31 | rs6831094 | 4 | 95912731 | 0.0241445 |
| 31 | rs1545326 | 4 | 95914424 | 0.0158171 |
| 31 | rs13128586 | 4 | 95915751 | 0.0202001 |
| 31 | rs13129054 | 4 | 95915991 | 0.0202001 |
| 31 | rs4145993 | 4 | 95916279 | 0.0202001 |
| 31 | rs6855395 | 4 | 95917980 | 0.0258405 |
| 31 | rs1470409 | 4 | 95920238 | 0.0331109 |
| 31 | rs1444926 | 4 | 95920959 | 0.0424975 |
| 31 | rs4397000 | 4 | 95922630 | 0.0424975 |
| 31 | rs4331783 | 4 | 95922662 | 0.0424975 |
| 31 | rs4299590 | 4 | 95923764 | 0.00923971 |
| 31 | rs10023544 | 4 | 95927802 | 0.0327017 |
| 31 | rs6840826 | 4 | 95931754 | 0.0375039 |
| 31 | rs6532525 | 4 | 95932152 | 0.0258405 |
| 31 | rs12500473 | 4 | 95933337 | 0.0454642 |
| 31 | rs6832300 | 4 | 95935914 | 0.0532419 |
| 31 | rs1348605 | 4 | 95936263 | 0.0532419 |
| 31 | rs12647875 | 4 | 95942472 | 0.0623912 |
| 31 | rs6815044 | 4 | 95943641 | 0.0623912 |
| 31 | rs28663472 | 4 | 95948204 | 0.100796 |
| 31 | rs7679627 | 4 | 95949153 | 0.100796 |
| 31 | rs1434546 | 4 | 95954962 | 0.0331109 |
| 32 | rs7663401 | 4 | 106128954 | 0.0903045 |
| 32 | rs2007403 | 4 | 106131210 | 0.107182 |
| 32 | rs6533183 | 4 | 106133184 | 0.0663878 |
| 32 | rs2047409 | 4 | 106137033 | 0.0761319 |
| 32 | rs11735256 | 4 | 106138146 | 0.0903045 |
| 32 | rs6839705 | 4 | 106144735 | 0.0925938 |
| 32 | rs1391439 | 4 | 106151642 | 0.0903045 |
| 32 | rs7683416 | 4 | 106152984 | 0.0521374 |
| 32 | rs7670522 | 4 | 106160365 | 0.0186455 |
| 32 | rs62330911 | 4 | 106167744 | 0.015745 |
| 32 | rs2647246 | 4 | 106171635 | 0.0261995 |
| 32 | rs2647247 | 4 | 106171652 | 0.0261995 |
| 32 | rs2647248 | 4 | 106174936 | 0.0261995 |
| 32 | rs2133086 | 4 | 106178289 | 0.015745 |
| 32 | rs1032625 | 4 | 106181573 | 0.0261995 |
| 32 | rs2454205 | 4 | 106184229 | 0.0186455 |
| 32 | rs2726459 | 4 | 106184597 | 0.015745 |
| 32 | rs2726521 | 4 | 106189614 | 0.0220948 |
| 32 | rs2647250 | 4 | 106190226 | 0.0369105 |

| | | | | |
|---|---|---|---|---|
| 32 | rs2726520 | 4 | 106193160 | 0.0186455 |
| 32 | rs2726519 | 4 | 106193334 | 0.0186455 |
| 33 | rs6858755 | 4 | 124632243 | 0.0397899 |
| 33 | rs9654147 | 4 | 124638933 | 0.0239037 |
| 33 | rs1834378 | 4 | 124650005 | 0.0479018 |
| 33 | rs11737502 | 4 | 124650645 | 0.0547045 |
| 33 | rs28433008 | 4 | 124652058 | 0.0625104 |
| 33 | rs11098717 | 4 | 124652559 | 0.0479018 |
| 33 | rs12505949 | 4 | 124654206 | 0.0547045 |
| 33 | rs12509610 | 4 | 124654263 | 0.0547045 |
| 33 | rs4833289 | 4 | 124654865 | 0.0419699 |
| 33 | rs67254681 | 4 | 124655126 | 0.0479018 |
| 33 | rs28757571 | 4 | 124656268 | 0.0547045 |
| 33 | rs2060285 | 4 | 124746377 | 0.440893 |
| 34 | rs72918058 | 4 | 126906982 | 0.0897367 |
| 34 | rs12651652 | 4 | 126910178 | 0.0684773 |
| 34 | rs17823858 | 4 | 126911191 | 0.0684773 |
| 34 | rs6817714 | 4 | 126914437 | 0.0269312 |
| 34 | rs17824014 | 4 | 126917274 | 0.0783733 |
| 34 | rs17010555 | 4 | 126917337 | 0.0350873 |
| 34 | rs17010558 | 4 | 126917820 | 0.0269312 |
| 34 | rs12647708 | 4 | 126921277 | 0.0783733 |
| 34 | rs28403275 | 4 | 126924684 | 0.0783733 |
| 34 | rs17824374 | 4 | 126924999 | 0.10279 |
| 34 | rs12513259 | 4 | 126930930 | 0.0523415 |
| 34 | rs2390928 | 4 | 126931413 | 0.0457896 |
| 34 | rs28446518 | 4 | 126933901 | 0.0159453 |
| 34 | rs72920119 | 4 | 126934327 | 0.0159453 |
| 34 | rs7674065 | 4 | 126939254 | 0.0181663 |
| 34 | rs6829242 | 4 | 126951364 | 0.005024 |
| 34 | rs4373172 | 4 | 126951752 | 0.00570241 |
| 34 | rs28882098 | 4 | 126952068 | 0.00570241 |
| 34 | rs28773024 | 4 | 126952428 | 0.00570241 |
| 34 | rs28594485 | 4 | 126957129 | 0.005024 |
| 34 | rs7682992 | 4 | 126958372 | 0.00442813 |
| 34 | rs7683776 | 4 | 126958705 | 0.00442813 |
| 34 | rs7661885 | 4 | 126958758 | 0.00390457 |
| 34 | rs7689104 | 4 | 126958909 | 0.00390457 |
| 34 | rs9985873 | 4 | 126959315 | 0.00390457 |
| 34 | rs9985557 | 4 | 126959509 | 0.00390457 |
| 34 | rs9986035 | 4 | 126959516 | 0.00390457 |
| 34 | rs9985879 | 4 | 126959860 | 0.00390457 |
| 34 | rs10020123 | 4 | 126960081 | 0.00390457 |
| 34 | rs10020221 | 4 | 126960184 | 0.00390457 |
| 34 | rs6812443 | 4 | 126961254 | 0.00442813 |
| 34 | rs6813545 | 4 | 126961471 | 0.00390457 |
| 34 | rs6813583 | 4 | 126961520 | 0.00390457 |
| 34 | rs12054487 | 4 | 126962278 | 0.00390457 |
| 34 | rs7668769 | 4 | 126963073 | 0.00390457 |
| 34 | rs7669189 | 4 | 126963192 | 0.00390457 |
| 34 | rs2390957 | 4 | 126964262 | 0.00390457 |
| 34 | rs7675977 | 4 | 126964299 | 0.00390457 |

| | | | | |
|---|---|---|---|---|
| 34 | rs7676003 | 4 | 126964373 | 0.00390457 |
| 34 | rs4992078 | 4 | 126964592 | 0.00390457 |
| 34 | rs4992079 | 4 | 126964681 | 0.00390457 |
| 34 | rs4992080 | 4 | 126964691 | 0.00390457 |
| 34 | rs9307580 | 4 | 126964999 | 0.00390457 |
| 34 | rs9307581 | 4 | 126965121 | 0.00390457 |
| 34 | rs6830127 | 4 | 126965665 | 0.00390457 |
| 34 | rs6858839 | 4 | 126966165 | 0.00390457 |
| 34 | rs6858823 | 4 | 126966184 | 0.00442813 |
| 34 | rs6836332 | 4 | 126966284 | 0.00442813 |
| 34 | rs6836803 | 4 | 126966519 | 0.00570241 |
| 34 | rs6811015 | 4 | 126966525 | 0.005024 |
| 35 | rs10009710 | 4 | 145233103 | 0.00156195 |
| 35 | rs6830386 | 4 | 145235543 | 0.00180328 |
| 35 | rs13118748 | 4 | 145259126 | 0.0053941 |
| 35 | rs7661046 | 4 | 145260495 | 0.0122347 |
| 35 | rs7375701 | 4 | 145260561 | 0.00240728 |
| 35 | rs6822064 | 4 | 145263756 | 0.00672523 |
| 35 | rs6840871 | 4 | 145264014 | 0.0184031 |
| 35 | rs4493485 | 4 | 145267634 | 0.0251466 |
| 35 | rs4266245 | 4 | 145267780 | 0.0251466 |
| 35 | rs12503296 | 4 | 145269728 | 0.00135361 |
| 35 | rs987246 | 4 | 145270867 | 0.0215064 |
| 35 | rs4340757 | 4 | 145271071 | 0.0215064 |
| 35 | rs6537280 | 4 | 145280378 | 0.00431661 |
| 35 | rs7378179 | 4 | 145284208 | 0.00372737 |
| 35 | rs4465995 | 4 | 145291206 | 0.00372737 |
| 35 | rs4475093 | 4 | 145292676 | 0.00240728 |
| 35 | rs10029738 | 4 | 145297419 | 0.0157561 |
| 35 | rs10029931 | 4 | 145297618 | 0.0134971 |
| 35 | rs7681655 | 4 | 145298829 | 0.0134971 |
| 35 | rs7376541 | 4 | 145300126 | 0.0134971 |
| 35 | rs4469023 | 4 | 145300434 | 0.0134971 |
| 35 | rs4370082 | 4 | 145300629 | 0.0115683 |
| 35 | rs7665807 | 4 | 145301382 | 0.0115683 |
| 35 | rs4318599 | 4 | 145302123 | 0.0115683 |
| 35 | rs6537284 | 4 | 145303691 | 0.0115683 |
| 35 | rs4383570 | 4 | 145304604 | 0.0115683 |
| 35 | rs7693416 | 4 | 145305357 | 0.0115683 |
| 35 | rs12645910 | 4 | 145307421 | 0.0134971 |
| 35 | rs6537287 | 4 | 145312027 | 0.0134971 |
| 35 | rs11943469 | 4 | 145313847 | 0.0134971 |
| 35 | rs6812520 | 4 | 145315659 | 0.0157561 |
| 35 | rs4572828 | 4 | 145320293 | 0.0157561 |
| 35 | rs7676032 | 4 | 145321068 | 0.0157561 |
| 35 | rs4420930 | 4 | 145321237 | 0.0157561 |
| 35 | rs4376087 | 4 | 145321355 | 0.0184031 |
| 35 | rs62334726 | 4 | 145341042 | 0.00579822 |
| 35 | rs72731556 | 4 | 145345984 | 0.00672523 |
| 35 | rs17019346 | 4 | 145347117 | 0.00156195 |
| 35 | rs11726412 | 4 | 145354023 | 0.00156195 |
| 35 | rs144044336 | 4 | 145361196 | 0.00180328 |

| | | | | |
|---|---|---|---|---|
| 35 | rs62334729 | 4 | 145361908 | 0.00208297 |
| 35 | rs62334730 | 4 | 145362891 | 0.00180328 |
| 35 | rs749316 | 4 | 145371346 | 0.00180328 |
| 35 | rs749317 | 4 | 145372009 | 0.00180328 |
| 35 | rs17019365 | 4 | 145373134 | 0.00208297 |
| 35 | rs17019368 | 4 | 145375405 | 0.00208297 |
| 35 | rs17019370 | 4 | 145381766 | 0.00189619 |
| 35 | rs11733975 | 4 | 145386473 | 0.0157561 |
| 35 | rs62343635 | 4 | 145388954 | 0.0184031 |
| 35 | rs17019390 | 4 | 145417318 | 0.00240728 |
| 35 | rs11736238 | 4 | 145418294 | 0.00180328 |
| 35 | rs72731582 | 4 | 145418803 | 0.00322023 |
| 35 | rs60539540 | 4 | 145422551 | 0.00240728 |
| 35 | rs56071345 | 4 | 145423450 | 0.00208297 |
| 35 | rs13116963 | 4 | 145424767 | 0.00208297 |
| 35 | rs17019400 | 4 | 145428511 | 0.00240728 |
| 35 | rs60950736 | 4 | 145429878 | 0.00240728 |
| 35 | rs11727583 | 4 | 145433002 | 0.00372737 |
| 35 | rs17019408 | 4 | 145433452 | 0.00322023 |
| 35 | rs1032296 | 4 | 145434688 | 0.00962076 |
| 35 | rs7681384 | 4 | 145437014 | 0.00588472 |
| 35 | rs6821114 | 4 | 145452389 | 0.00139287 |
| 35 | rs6845536 | 4 | 145452783 | 0.0133933 |
| 35 | rs1489764 | 4 | 145454228 | 0.00163065 |
| 35 | rs1489761 | 4 | 145455491 | 0.00191022 |
| 35 | rs7692102 | 4 | 145457141 | 0.00191022 |
| 35 | rs7674469 | 4 | 145458001 | 0.00223912 |
| 35 | rs7663578 | 4 | 145468791 | 0.0434805 |
| 35 | rs7663740 | 4 | 145468845 | 0.00262628 |
| 35 | rs6828982 | 4 | 145470604 | 0.00500158 |
| 35 | rs12511230 | 4 | 145471245 | 0.00180962 |
| 35 | rs13113445 | 4 | 145472766 | 0.00692813 |
| 35 | rs4834988 | 4 | 145478777 | 0.00692813 |
| 35 | rs12510044 | 4 | 145484638 | 0.00816164 |
| 35 | rs10029430 | 4 | 145498412 | 0.00816164 |
| 35 | rs6537297 | 4 | 145502029 | 0.0309616 |
| 35 | rs6537298 | 4 | 145506871 | 0.0261513 |
| 35 | rs7670758 | 4 | 145511875 | 0.0366795 |
| 35 | rs1542726 | 4 | 145515769 | 0.206696 |
| 35 | rs1489765 | 4 | 145516378 | 0.0196302 |
| 36 | rs10475282 | 5 | 523995 | 0.0300861 |
| 36 | rs72704802 | 5 | 554211 | 0.00341779 |
| 36 | rs4527146 | 5 | 555236 | 0.00391917 |
| 36 | rs12522303 | 5 | 555564 | 0.00341779 |
| 36 | rs12521988 | 5 | 556421 | 0.00391917 |
| 36 | rs17497684 | 5 | 558307 | 0.00298169 |
| 36 | rs4957056 | 5 | 570105 | 0.00341779 |
| 36 | rs72703026 | 5 | 572383 | 0.0078152 |
| 36 | rs4081847 | 5 | 574072 | 0.00680233 |
| 36 | rs4957053 | 5 | 575927 | 0.00898235 |
| 36 | rs111352378 | 5 | 577079 | 0.0103278 |
| 36 | rs72703034 | 5 | 577101 | 0.0103278 |

| | | | | |
|---|---|---|---|---|
| 36 | rs7443550 | 5 | 578404 | 0.0103278 |
| 36 | rs4245972 | 5 | 578572 | 0.0118792 |
| 36 | rs72703042 | 5 | 579269 | 0.0118792 |
| 36 | rs58015612 | 5 | 580665 | 0.0118792 |
| 36 | rs56216231 | 5 | 581193 | 0.0118792 |
| 36 | rs12519469 | 5 | 581772 | 0.0103278 |
| 36 | rs56368535 | 5 | 582362 | 0.013669 |
| 36 | rs56278696 | 5 | 582669 | 0.0157346 |
| 36 | rs72703051 | 5 | 582997 | 0.015314 |
| 36 | rs56350081 | 5 | 583198 | 0.0157346 |
| 36 | rs12522140 | 5 | 587261 | 0.013669 |
| 36 | rs12518859 | 5 | 587432 | 0.013669 |
| 36 | rs72703064 | 5 | 587825 | 0.013669 |
| 36 | rs72703065 | 5 | 588123 | 0.013669 |
| 36 | rs1399381 | 5 | 589853 | 0.0118792 |
| 36 | rs72703070 | 5 | 590458 | 0.0208731 |
| 36 | rs72703072 | 5 | 590626 | 0.0181191 |
| 36 | rs12521091 | 5 | 590710 | 0.013669 |
| 36 | rs12522724 | 5 | 590742 | 0.013669 |
| 36 | rs56328416 | 5 | 591138 | 0.013669 |
| 36 | rs72703075 | 5 | 592819 | 0.0101353 |
| 36 | rs111707179 | 5 | 593308 | 0.00296548 |
| 36 | rs56282020 | 5 | 596701 | 0.0025811 |
| 36 | rs57306627 | 5 | 597643 | 0.0025811 |
| 36 | rs11750321 | 5 | 599196 | 0.00296548 |
| 36 | rs72703092 | 5 | 599269 | 0.00340845 |
| 36 | rs3749615 | 5 | 601485 | 0.0025811 |
| 36 | rs72703095 | 5 | 601789 | 0.0025811 |
| 36 | rs3749618 | 5 | 602650 | 0.00296548 |
| 36 | rs11739847 | 5 | 609661 | 0.00296548 |
| 36 | rs11739866 | 5 | 636299 | 0.0025811 |
| 36 | rs12522955 | 5 | 639231 | 0.00296548 |
| 36 | rs3792720 | 5 | 643028 | 0.00518785 |
| 36 | rs4957080 | 5 | 644552 | 0.00597233 |
| 36 | rs12517638 | 5 | 645484 | 0.00391917 |
| 36 | rs72705030 | 5 | 645562 | 0.00518785 |
| 36 | rs4957081 | 5 | 646418 | 0.00450821 |
| 36 | rs72705097 | 5 | 655067 | 0.00518785 |
| 36 | rs113896354 | 5 | 655438 | 0.00518785 |
| 36 | rs12520279 | 5 | 657291 | 0.00450821 |
| 36 | rs4957083 | 5 | 659343 | 0.00450821 |
| 36 | rs72705102 | 5 | 659437 | 0.00450821 |
| 36 | rs7558 | 5 | 660491 | 0.00450821 |
| 36 | rs7434 | 5 | 660804 | 0.00450821 |
| 36 | rs72707007 | 5 | 661856 | 0.00597233 |
| 36 | rs3762951 | 5 | 662613 | 0.00687819 |
| 36 | rs28364691 | 5 | 664084 | 0.0121487 |
| 36 | rs72707016 | 5 | 665148 | 0.00515932 |
| 36 | rs61731455 | 5 | 665295 | 0.00680233 |
| 36 | rs74553517 | 5 | 665671 | 0.005923 |
| 36 | rs72707023 | 5 | 667620 | 0.0338477 |
| 36 | rs11749927 | 5 | 668842 | 0.0266892 |

| | | | | |
|---|---|---|---|---|
| 36 | rs77015126 | 5 | 670909 | 0.0175857 |
| 36 | rs72707027 | 5 | 671294 | 0.0266892 |
| 36 | rs12515813 | 5 | 671826 | 0.0175857 |
| 36 | rs11740553 | 5 | 672593 | 0.035312 |
| 36 | rs11742854 | 5 | 674141 | 0.0130323 |
| 36 | rs12516318 | 5 | 676423 | 0.032496 |
| 36 | rs12519390 | 5 | 676546 | 0.0371971 |
| 36 | rs72707041 | 5 | 677152 | 0.00391917 |
| 36 | rs72707043 | 5 | 677200 | 0.032496 |
| 36 | rs72707044 | 5 | 677239 | 0.032496 |
| 36 | rs56060145 | 5 | 681279 | 0.0217113 |
| 36 | rs113948530 | 5 | 681976 | 0.0189934 |
| 36 | rs72707053 | 5 | 684483 | 0.0239928 |
| 36 | rs55651840 | 5 | 684848 | 0.00269876 |
| 36 | rs55660354 | 5 | 684888 | 0.0210351 |
| 36 | rs11745112 | 5 | 685849 | 0.0312455 |
| 36 | rs35696919 | 5 | 686244 | 0.00917878 |
| 37 | rs13170478 | 5 | 15602644 | 0.00290099 |
| 37 | rs12187518 | 5 | 15623646 | 0.00351305 |
| 37 | rs13185548 | 5 | 15655056 | 0.00256486 |
| 37 | rs12654662 | 5 | 15659239 | 0.00697926 |
| 37 | rs17647599 | 5 | 15659551 | 0.00256486 |
| 37 | rs12658048 | 5 | 15664613 | 0.00200833 |
| 37 | rs74655962 | 5 | 15680745 | 0.00284095 |
| 37 | rs77887104 | 5 | 15688284 | 0.0031021 |
| 37 | rs17523716 | 5 | 15688710 | 0.0023848 |
| 37 | rs17602097 | 5 | 15688927 | 0.00200419 |
| 37 | rs75872775 | 5 | 15693584 | 0.00218591 |
| 37 | rs17523929 | 5 | 15700585 | 0.00218591 |
| 37 | rs17524082 | 5 | 15722647 | 0.00198403 |
| 37 | rs17602533 | 5 | 15741441 | 0.0423664 |
| 37 | rs4379195 | 5 | 15743346 | 0.057199 |
| 37 | rs113271117 | 5 | 15744475 | 0.057199 |
| 37 | rs111608913 | 5 | 15745063 | 0.057199 |
| 37 | rs13189719 | 5 | 15749077 | 0.057199 |
| 37 | rs2173691 | 5 | 15753663 | 0.0468105 |
| 37 | rs1909723 | 5 | 15757154 | 0.0468105 |
| 37 | rs61026653 | 5 | 15757570 | 0.057199 |
| 37 | rs17602672 | 5 | 15758279 | 0.0423664 |
| 37 | rs1973477 | 5 | 15759237 | 0.0468105 |
| 37 | rs36056396 | 5 | 15765428 | 0.0517368 |
| 37 | rs36139170 | 5 | 15766089 | 0.0423664 |
| 37 | rs6870946 | 5 | 15767015 | 0.0468105 |
| 37 | rs6870947 | 5 | 15767018 | 0.0468105 |
| 37 | rs34894816 | 5 | 15768609 | 0.00592759 |
| 37 | rs10520824 | 5 | 15768708 | 0.0096138 |
| 37 | rs2402182 | 5 | 15769041 | 0.0468105 |
| 37 | rs952161 | 5 | 15769146 | 0.0096138 |
| 37 | rs13174706 | 5 | 15771042 | 0.0517368 |
| 37 | rs10462723 | 5 | 15771101 | 0.0240164 |
| 37 | rs12186993 | 5 | 15772779 | 0.00592759 |
| 37 | rs1505033 | 5 | 15773877 | 0.00531884 |

| | | | | |
|---|---|---|---|---|
| 37 | rs1505035 | 5 | 15774466 | 0.00531884 |
| 37 | rs1505036 | 5 | 15776264 | 0.0423664 |
| 37 | rs6896896 | 5 | 15776401 | 0.00592759 |
| 37 | rs2055624 | 5 | 15777817 | 0.00271135 |
| 38 | rs6449598 | 5 | 50665023 | 0.0770163 |
| 38 | rs7701852 | 5 | 50666897 | 0.0770163 |
| 38 | rs6449600 | 5 | 50675297 | 0.0770163 |
| 38 | rs6899279 | 5 | 50678414 | 0.0882768 |
| 38 | rs3811910 | 5 | 50681032 | 0.116088 |
| 38 | rs62368263 | 5 | 50726027 | 0.465367 |
| 38 | rs17824230 | 5 | 50759375 | 0.0586773 |
| 39 | rs6449598 | 5 | 50665023 | 0.0770163 |
| 39 | rs7701852 | 5 | 50666897 | 0.0770163 |
| 39 | rs6449600 | 5 | 50675297 | 0.0770163 |
| 39 | rs6899279 | 5 | 50678414 | 0.0882768 |
| 39 | rs3811910 | 5 | 50681032 | 0.116088 |
| 39 | rs62368263 | 5 | 50726027 | 0.465367 |
| 39 | rs17824230 | 5 | 50759375 | 0.0586773 |
| 40 | rs13154000 | 5 | 93027669 | 0.00192874 |
| 40 | rs67914792 | 5 | 93036584 | 0.00192874 |
| 40 | rs167570 | 5 | 93042348 | 0.00253709 |
| 40 | rs17312456 | 5 | 93043907 | 0.00192874 |
| 40 | rs10055389 | 5 | 93046997 | 0.00160887 |
| 40 | rs13154650 | 5 | 93052205 | 0.000940271 |
| 40 | rs72786647 | 5 | 93053517 | 0.00134359 |
| 40 | rs28534784 | 5 | 93063332 | 0.00134359 |
| 40 | rs17372057 | 5 | 93071916 | 0.00102759 |
| 40 | rs12153021 | 5 | 93073913 | 0.00176131 |
| 40 | rs13165400 | 5 | 93091427 | 0.00147005 |
| 40 | rs13357713 | 5 | 93145423 | 0.00086062 |
| 40 | rs10061377 | 5 | 93174513 | 0.00102759 |
| 40 | rs10075217 | 5 | 93176774 | 0.000940271 |
| 40 | rs36042718 | 5 | 93180997 | 0.00086062 |
| 40 | rs13153987 | 5 | 93190000 | 0.00102759 |
| 40 | rs10038424 | 5 | 93193437 | 0.000940271 |
| 40 | rs10066431 | 5 | 93198204 | 0.000940271 |
| 40 | rs6877030 | 5 | 93201841 | 0.00086062 |
| 40 | rs6898842 | 5 | 93213221 | 0.00086062 |
| 40 | rs6874766 | 5 | 93213807 | 0.000940271 |
| 40 | rs35496867 | 5 | 93220400 | 0.00086062 |
| 40 | rs34967135 | 5 | 93221785 | 0.00086062 |
| 40 | rs7715562 | 5 | 93222034 | 0.000940271 |
| 40 | rs13171131 | 5 | 93223028 | 0.00086062 |
| 40 | rs28705226 | 5 | 93233135 | 0.00086062 |
| 40 | rs67328989 | 5 | 93243133 | 0.00086062 |
| 40 | rs28610997 | 5 | 93248374 | 0.00086062 |
| 40 | rs28412976 | 5 | 93248377 | 0.00102759 |
| 40 | rs72786702 | 5 | 93254656 | 0.00086062 |
| 40 | rs74825443 | 5 | 93254973 | 0.00086062 |
| 40 | rs10067259 | 5 | 93290838 | 0.000940271 |
| 40 | rs17374627 | 5 | 93296306 | 0.00305025 |
| 40 | rs10055741 | 5 | 93297187 | 0.00231486 |

| | | | | |
|---|---|---|---|---|
| 40 | rs7734897 | 5 | 93305402 | 0.00253709 |
| 40 | rs7736620 | 5 | 93305762 | 0.00253709 |
| 40 | rs13186579 | 5 | 93306939 | 0.0012743 |
| 40 | rs13177800 | 5 | 93307350 | 0.00231486 |
| 40 | rs13173952 | 5 | 93307557 | 0.00253709 |
| 40 | rs10036829 | 5 | 93309897 | 0.00231486 |
| 40 | rs35747258 | 5 | 93310129 | 0.00211269 |
| 40 | rs7701974 | 5 | 93311417 | 0.00253709 |
| 40 | rs7701011 | 5 | 93311456 | 0.00231486 |
| 40 | rs34673483 | 5 | 93315079 | 0.00231486 |
| 40 | rs10055340 | 5 | 93317644 | 0.00231486 |
| 40 | rs10076052 | 5 | 93317762 | 0.00211269 |
| 40 | rs113566899 | 5 | 93322042 | 0.00943606 |
| 40 | rs9314092 | 5 | 93326362 | 0.00186551 |
| 40 | rs13169675 | 5 | 93331284 | 0.00278146 |
| 40 | rs9314093 | 5 | 93333347 | 0.00211269 |
| 40 | rs10476591 | 5 | 93333723 | 0.00186551 |
| 40 | rs13185660 | 5 | 93336500 | 0.00278146 |
| 40 | rs28575883 | 5 | 93346035 | 0.00278146 |
| 40 | rs10076192 | 5 | 93348073 | 0.00305025 |
| 40 | rs10062711 | 5 | 93348659 | 0.00305025 |
| 40 | rs10050465 | 5 | 93349963 | 0.00334596 |
| 40 | rs10066791 | 5 | 93350255 | 0.00334596 |
| 40 | rs2290937 | 5 | 93356515 | 0.00402965 |
| 40 | rs12109629 | 5 | 93359702 | 0.00402965 |
| 40 | rs12153501 | 5 | 93360866 | 0.00402965 |
| 40 | rs12153506 | 5 | 93360953 | 0.00402965 |
| 40 | rs10078194 | 5 | 93364310 | 0.0036714 |
| 40 | rs10476596 | 5 | 93367074 | 0.00334596 |
| 40 | rs10476597 | 5 | 93367172 | 0.00334596 |
| 40 | rs10476598 | 5 | 93367281 | 0.0036714 |
| 40 | rs34595581 | 5 | 93369422 | 0.00334596 |
| 40 | rs13153245 | 5 | 93374891 | 0.0036714 |
| 40 | rs13170509 | 5 | 93378564 | 0.00334596 |
| 40 | rs10065056 | 5 | 93380123 | 0.0036714 |
| 40 | rs9885329 | 5 | 93380278 | 0.00334596 |
| 40 | rs10476599 | 5 | 93383423 | 0.00334596 |
| 40 | rs35075119 | 5 | 93385457 | 0.00402965 |
| 40 | rs34138834 | 5 | 93385505 | 0.00402965 |
| 40 | rs10060160 | 5 | 93387879 | 0.0036714 |
| 40 | rs13156865 | 5 | 93390908 | 0.00402965 |
| 40 | rs10061099 | 5 | 93396715 | 0.00402965 |
| 40 | rs12716456 | 5 | 93397557 | 0.00402965 |
| 40 | rs12716457 | 5 | 93397627 | 0.0036714 |
| 40 | rs10072232 | 5 | 93398890 | 0.0036714 |
| 40 | rs10057814 | 5 | 93399092 | 0.00402965 |
| 40 | rs2085960 | 5 | 93399934 | 0.0036714 |
| 40 | rs10063612 | 5 | 93401133 | 0.00442412 |
| 40 | rs10476601 | 5 | 93402522 | 0.0036714 |
| 40 | rs7728736 | 5 | 93404277 | 0.00402965 |
| 40 | rs72788365 | 5 | 93404792 | 0.00270409 |
| 40 | rs12153788 | 5 | 93405437 | 0.0036714 |

| | | | | |
|---|---|---|---|---|
| 40 | rs12152763 | 5 | 93405509 | 0.0036714 |
| 40 | rs10072592 | 5 | 93405989 | 0.00402965 |
| 40 | rs10044139 | 5 | 93407803 | 0.00211269 |
| 40 | rs10514380 | 5 | 93409370 | 0.00402965 |
| 40 | rs71641007 | 5 | 93413044 | 0.00586481 |
| 40 | rs12716460 | 5 | 93416733 | 0.00533728 |
| 40 | rs10061080 | 5 | 93420559 | 0.00644632 |
| 40 | rs13165147 | 5 | 93422214 | 0.00586481 |
| 40 | rs13356088 | 5 | 93423479 | 0.00807197 |
| 40 | rs66696214 | 5 | 93424533 | 0.00586481 |
| 40 | rs10440717 | 5 | 93424852 | 0.00244193 |
| 40 | rs9314095 | 5 | 93425495 | 0.00533728 |
| 40 | rs9314096 | 5 | 93426005 | 0.0103866 |
| 40 | rs10064520 | 5 | 93427358 | 0.00586481 |
| 40 | rs55832164 | 5 | 93436579 | 0.00533728 |
| 40 | rs10044066 | 5 | 93437580 | 0.00586481 |
| 40 | rs17315943 | 5 | 93442830 | 0.00586481 |
| 40 | rs10057605 | 5 | 93443950 | 0.00644632 |
| 40 | rs34184666 | 5 | 93444584 | 0.00586481 |
| 40 | rs10476602 | 5 | 93445255 | 0.00708753 |
| 40 | rs17083455 | 5 | 93446380 | 0.00644632 |
| 40 | rs28633397 | 5 | 93447048 | 0.00644632 |
| 40 | rs9314099 | 5 | 93447375 | 0.00708753 |
| 40 | rs9314100 | 5 | 93447394 | 0.00586481 |
| 40 | rs10059230 | 5 | 93450347 | 0.00533728 |
| 40 | rs10070390 | 5 | 93452345 | 0.0048586 |
| 40 | rs10060462 | 5 | 93455073 | 0.00533728 |
| 40 | rs17316137 | 5 | 93456577 | 0.00533728 |
| 40 | rs10063045 | 5 | 93457584 | 0.00807197 |
| 40 | rs10059630 | 5 | 93459807 | 0.00533728 |
| 40 | rs10213956 | 5 | 93460839 | 0.00533728 |
| 40 | rs9314101 | 5 | 93461593 | 0.0048586 |
| 40 | rs10476526 | 5 | 93461916 | 0.0048586 |
| 40 | rs17083460 | 5 | 93462354 | 0.0048586 |
| 40 | rs17083462 | 5 | 93462401 | 0.00402965 |
| 40 | rs12716462 | 5 | 93466003 | 0.00533728 |
| 40 | rs10073039 | 5 | 93467709 | 0.00586481 |
| 40 | rs12153676 | 5 | 93468524 | 0.00586481 |
| 40 | rs143657929 | 5 | 93470525 | 0.00586481 |
| 40 | rs55783479 | 5 | 93473130 | 0.00644632 |
| 40 | rs34890659 | 5 | 93475424 | 0.00644632 |
| 40 | rs10052428 | 5 | 93476530 | 0.00708753 |
| 40 | rs7718842 | 5 | 93480568 | 0.00779475 |
| 40 | rs12374470 | 5 | 93481046 | 0.00708753 |
| 40 | rs12374432 | 5 | 93481348 | 0.00708753 |
| 40 | rs12374488 | 5 | 93481407 | 0.00708753 |
| 40 | rs12374463 | 5 | 93483569 | 0.00708753 |
| 40 | rs7702521 | 5 | 93484579 | 0.0036714 |
| 40 | rs10055994 | 5 | 93488425 | 0.008575 |
| 40 | rs10040648 | 5 | 93490301 | 0.00644632 |
| 40 | rs10035340 | 5 | 93490465 | 0.0066953 |
| 40 | rs56373208 | 5 | 93491276 | 0.00779475 |

| | | | | |
|---|---|---|---|---|
| 40 | rs67066109 | 5 | 93494101 | 0.00779475 |
| 40 | rs28637444 | 5 | 93495548 | 0.00779475 |
| 40 | rs9314102 | 5 | 93500251 | 0.00943606 |
| 40 | rs36026719 | 5 | 93505876 | 0.0107077 |
| 40 | rs34013242 | 5 | 93507947 | 0.00807197 |
| 40 | rs13156002 | 5 | 93508931 | 0.00281102 |
| 40 | rs9968749 | 5 | 93509559 | 0.00807197 |
| 40 | rs13185608 | 5 | 93509688 | 0.00506827 |
| 40 | rs9968691 | 5 | 93509756 | 0.00807197 |
| 40 | rs59472588 | 5 | 93511981 | 0.0066953 |
| 40 | rs12187701 | 5 | 93522290 | 0.0107077 |
| 40 | rs36091096 | 5 | 93525586 | 0.0129453 |
| 40 | rs56046246 | 5 | 93531545 | 0.0156677 |
| 40 | rs58734583 | 5 | 93532155 | 0.0172439 |
| 40 | rs13181660 | 5 | 93533658 | 0.0156677 |
| 40 | rs3940842 | 5 | 93534858 | 0.0172439 |
| 40 | rs34854140 | 5 | 93538169 | 0.0172439 |
| 40 | rs61132622 | 5 | 93539684 | 0.0209052 |
| 40 | rs17376207 | 5 | 93545524 | 0.0156677 |
| 40 | rs13163610 | 5 | 93548877 | 0.0172439 |
| 40 | rs13154249 | 5 | 93550508 | 0.0189838 |
| 40 | rs17323433 | 5 | 93551028 | 0.0172439 |
| 40 | rs66978370 | 5 | 93553633 | 0.0156677 |
| 40 | rs13174700 | 5 | 93554644 | 0.0189838 |
| 40 | rs6867042 | 5 | 93555761 | 0.0230273 |
| 40 | rs34756085 | 5 | 93556294 | 0.0189838 |
| 40 | rs17376456 | 5 | 93557702 | 0.0189838 |
| 40 | rs12188453 | 5 | 93560807 | 0.0132933 |
| 40 | rs35184505 | 5 | 93561535 | 0.0146007 |
| 40 | rs12189466 | 5 | 93562287 | 0.0121062 |
| 40 | rs34068652 | 5 | 93563319 | 0.00787905 |
| 40 | rs10514382 | 5 | 93566534 | 0.00719695 |
| 40 | rs4547912 | 5 | 93571190 | 0.00657562 |
| 41 | rs11745914 | 5 | 97654496 | 0.00268687 |
| 41 | rs17165848 | 5 | 97655053 | 0.00391717 |
| 41 | rs17165849 | 5 | 97655286 | 0.00444697 |
| 41 | rs6876270 | 5 | 97655526 | 0.00444697 |
| 41 | rs62366857 | 5 | 97655681 | 0.00391717 |
| 41 | rs12654883 | 5 | 97656313 | 0.048438 |
| 41 | rs1350022 | 5 | 97656609 | 0.00505142 |
| 41 | rs1378444 | 5 | 97656641 | 0.00574144 |
| 41 | rs4538635 | 5 | 97656704 | 0.00505142 |
| 41 | rs6898721 | 5 | 97659340 | 0.00574144 |
| 41 | rs12652566 | 5 | 97660264 | 0.00505142 |
| 41 | rs35774032 | 5 | 97660482 | 0.00743035 |
| 41 | rs60805392 | 5 | 97660706 | 0.00574144 |
| 41 | rs12513398 | 5 | 97661152 | 0.00574144 |
| 41 | rs7723838 | 5 | 97661663 | 0.00505142 |
| 41 | rs2198680 | 5 | 97662522 | 0.00743035 |
| 41 | rs1455427 | 5 | 97700050 | 0.00444697 |
| 41 | rs10067255 | 5 | 97757677 | 0.00352953 |
| 41 | rs1563319 | 5 | 97764143 | 0.0554395 |

| | | | | |
|---|---|---|---|---|
| 41 | rs7715564 | 5 | 97764522 | 0.00885493 |
| 41 | rs10043462 | 5 | 97765856 | 0.0263017 |
| 41 | rs1002268 | 5 | 97766797 | 0.0229053 |
| 41 | rs59060802 | 5 | 97768250 | 0.0302207 |
| 41 | rs1563318 | 5 | 97768368 | 0.00743035 |
| 41 | rs1563317 | 5 | 97768486 | 0.0347452 |
| 41 | rs3890433 | 5 | 97768954 | 0.0263017 |
| 41 | rs1473378 | 5 | 97769853 | 0.00743035 |
| 41 | rs1840496 | 5 | 97771540 | 0.0174041 |
| 41 | rs56410174 | 5 | 97772643 | 0.0263017 |
| 41 | rs59941249 | 5 | 97773493 | 0.0199599 |
| 41 | rs56870134 | 5 | 97773523 | 0.0199599 |
| 41 | rs12152743 | 5 | 97773634 | 0.0174041 |
| 41 | rs1840493 | 5 | 97774464 | 0.0229053 |
| 41 | rs61048056 | 5 | 97775200 | 0.0263017 |
| 41 | rs6861347 | 5 | 97775269 | 0.0199599 |
| 41 | rs11949814 | 5 | 97775678 | 0.0101236 |
| 41 | rs66986022 | 5 | 97776864 | 0.00885493 |
| 41 | rs73143889 | 5 | 97776897 | 0.0199599 |
| 41 | rs11951464 | 5 | 97777031 | 0.0199599 |
| 41 | rs11950768 | 5 | 97777300 | 0.0174041 |
| 41 | rs2085231 | 5 | 97778876 | 0.0101236 |
| 41 | rs751882 | 5 | 97779409 | 0.0174041 |
| 41 | rs6865661 | 5 | 97780336 | 0.0199599 |
| 41 | rs10063500 | 5 | 97780622 | 0.0199599 |
| 41 | rs2368553 | 5 | 97781753 | 0.0246738 |
| 41 | rs1037597 | 5 | 97783046 | 0.0284431 |
| 41 | rs1037598 | 5 | 97783125 | 0.0161704 |
| 41 | rs1037599 | 5 | 97783491 | 0.0186041 |
| 41 | rs9686816 | 5 | 97783958 | 0.0246738 |
| 41 | rs10043034 | 5 | 97784526 | 0.0328098 |
| 41 | rs11241450 | 5 | 97784711 | 0.0161704 |
| 41 | rs1072722 | 5 | 97785425 | 0.0101236 |
| 41 | rs10035465 | 5 | 97786312 | 0.0115813 |
| 41 | rs12152757 | 5 | 97786359 | 0.00678728 |
| 41 | rs1840494 | 5 | 97787110 | 0.0132572 |
| 41 | rs1840495 | 5 | 97787165 | 0.00678728 |
| 41 | rs723206 | 5 | 97789313 | 0.00885493 |
| 41 | rs723207 | 5 | 97789681 | 0.00775006 |
| 41 | rs7717448 | 5 | 97790471 | 0.0115813 |
| 41 | rs7717913 | 5 | 97790692 | 0.00885493 |
| 41 | rs6870628 | 5 | 97791720 | 0.00775006 |
| 41 | rs11241481 | 5 | 97792958 | 0.00316467 |
| 41 | rs17165974 | 5 | 97794284 | 0.00361238 |
| 41 | rs4434407 | 5 | 97795220 | 0.00401763 |
| 41 | rs12655510 | 5 | 97795382 | 0.00361238 |
| 41 | rs10463714 | 5 | 97797651 | 0.00361238 |
| 41 | rs10463716 | 5 | 97797822 | 0.00361238 |
| 41 | rs10463717 | 5 | 97797963 | 0.00361238 |
| 41 | rs11241524 | 5 | 97801699 | 0.00277427 |
| 42 | rs79121939 | 5 | 164487955 | 0.0454272 |
| 42 | rs72807969 | 5 | 164504397 | 0.0334234 |

| | | | | |
|---|---|---|---|---|
| 42 | rs72809524 | 5 | 164517704 | 0.0503486 |
| 42 | rs6556812 | 5 | 164589870 | 0.0370124 |
| 42 | rs72817597 | 5 | 164603356 | 0.0105917 |
| 42 | rs12189342 | 5 | 164604894 | 0.00680623 |
| 42 | rs72817599 | 5 | 164611054 | 0.00621441 |
| 42 | rs72817600 | 5 | 164611652 | 0.0333667 |
| 42 | rs72819561 | 5 | 164628055 | 0.00745629 |
| 42 | rs12153515 | 5 | 164631794 | 0.104273 |
| 42 | rs17392297 | 5 | 164642051 | 0.0333667 |
| 42 | rs72819565 | 5 | 164653671 | 0.00680623 |
| 42 | rs72819568 | 5 | 164669872 | 0.0156094 |
| 42 | rs2861140 | 5 | 164682127 | 0.0643547 |
| 42 | rs4291038 | 5 | 164688851 | 0.0266534 |
| 42 | rs971569 | 5 | 164690314 | 0.0499183 |
| 42 | rs973845 | 5 | 164692215 | 0.0499183 |
| 42 | rs973847 | 5 | 164692410 | 0.0439928 |
| 42 | rs6870836 | 5 | 164696206 | 0.0387875 |
| 42 | rs1160830 | 5 | 164698709 | 0.030191 |
| 42 | rs6859347 | 5 | 164708583 | 0.0080643 |
| 42 | rs1433001 | 5 | 164710430 | 0.034213 |
| 42 | rs72819586 | 5 | 164714501 | 0.00621441 |
| 42 | rs6556816 | 5 | 164731732 | 0.0266534 |
| 42 | rs13174360 | 5 | 164741327 | 0.0235405 |
| 42 | rs13166122 | 5 | 164743780 | 0.0183868 |
| 42 | rs13166291 | 5 | 164743857 | 0.0183868 |
| 42 | rs2861144 | 5 | 164744450 | 0.0183868 |
| 42 | rs2115255 | 5 | 164746324 | 0.0183868 |
| 42 | rs72821605 | 5 | 164747563 | 0.0247382 |
| 42 | rs1017790 | 5 | 164747878 | 0.0183868 |
| 42 | rs72821608 | 5 | 164749129 | 0.0247382 |
| 42 | rs11959015 | 5 | 164751760 | 0.00998915 |
| 42 | rs11956053 | 5 | 164751881 | 0.00784973 |
| 43 | rs9504383 | 6 | 589924 | 0.0285809 |
| 43 | rs9405263 | 6 | 591294 | 0.0321401 |
| 43 | rs6940427 | 6 | 591765 | 0.0285809 |
| 43 | rs2294660 | 6 | 592249 | 0.0321401 |
| 43 | rs3757121 | 6 | 593419 | 0.0113859 |
| 43 | rs3799311 | 6 | 594407 | 0.0361612 |
| 43 | rs1125815 | 6 | 595438 | 0.0321401 |
| 43 | rs4960091 | 6 | 597744 | 0.0321401 |
| 43 | rs3823137 | 6 | 604413 | 0.0361612 |
| 43 | rs9405841 | 6 | 607342 | 0.0741519 |
| 43 | rs2294666 | 6 | 617519 | 0.00283 |
| 43 | rs1150858 | 6 | 622885 | 0.0324502 |
| 43 | rs1150859 | 6 | 623284 | 0.0324502 |
| 43 | rs4960127 | 6 | 624892 | 0.135226 |
| 43 | rs4959352 | 6 | 624922 | 0.388517 |
| 43 | rs2294669 | 6 | 625673 | 0.00264485 |
| 43 | rs11753249 | 6 | 693638 | 0.012751 |
| 44 | rs2318089 | 6 | 1775436 | 0.352974 |
| 44 | rs722587 | 6 | 1775714 | 0.459289 |
| 44 | rs722585 | 6 | 1775863 | 0.160715 |

| | | | | |
|---|---|---|---|---|
| 45 | rs6917728 | 6 | 17386607 | 0.169581 |
| 45 | rs7773313 | 6 | 17386895 | 0.100339 |
| 45 | rs7759059 | 6 | 17387038 | 0.130351 |
| 45 | rs7755093 | 6 | 17387077 | 0.148652 |
| 45 | rs10807610 | 6 | 17387538 | 0.193527 |
| 45 | rs10949420 | 6 | 17387784 | 0.0416772 |
| 45 | rs10949421 | 6 | 17387932 | 0.0206872 |
| 45 | rs12190229 | 6 | 17388345 | 0.0206872 |
| 45 | rs12204235 | 6 | 17388551 | 0.0690948 |
| 45 | rs2328105 | 6 | 17457107 | 0.00807257 |
| 45 | rs1535014 | 6 | 17460161 | 0.00950983 |
| 45 | rs1535015 | 6 | 17460165 | 0.00950983 |
| 45 | rs1535016 | 6 | 17460399 | 0.0068568 |
| 45 | rs9383286 | 6 | 17461225 | 0.0068568 |
| 45 | rs10949424 | 6 | 17462113 | 0.00950983 |
| 45 | rs1320994 | 6 | 17470693 | 0.00633408 |
| 46 | rs2524005 | 6 | 29899677 | 0.0322642 |
| 46 | rs4248140 | 6 | 29935798 | 0.00299333 |
| 46 | rs6415118 | 6 | 29935850 | 0.00677859 |
| 46 | rs9260809 | 6 | 29941617 | 0.0409221 |
| 46 | rs3823376 | 6 | 29944184 | 0.00677859 |
| 46 | rs3823379 | 6 | 29944458 | 0.0217798 |
| 46 | rs3823381 | 6 | 29944558 | 0.0409221 |
| 46 | rs9261043 | 6 | 29966726 | 0.00486995 |
| 46 | rs12663724 | 6 | 30059616 | 0.00605796 |
| 46 | rs17187791 | 6 | 30060021 | 0.0126951 |
| 46 | rs7749793 | 6 | 30060364 | 0.0115598 |
| 46 | rs7754005 | 6 | 30060629 | 0.00609824 |
| 46 | rs7753935 | 6 | 30060783 | 0.0105297 |
| 46 | rs11967911 | 6 | 30060952 | 0.0072738 |
| 46 | rs28454792 | 6 | 30061010 | 0.00729812 |
| 46 | rs17194160 | 6 | 30061080 | 0.00667016 |
| 46 | rs17187805 | 6 | 30061202 | 0.0105297 |
| 46 | rs17194174 | 6 | 30061910 | 0.00358613 |
| 46 | rs12661731 | 6 | 30062345 | 0.0455012 |
| 46 | rs28780078 | 6 | 30062650 | 0.050613 |
| 46 | rs28780079 | 6 | 30062973 | 0.0455012 |
| 46 | rs28780080 | 6 | 30063086 | 0.050613 |
| 46 | rs7765810 | 6 | 30063496 | 0.0409221 |
| 46 | rs7761563 | 6 | 30063660 | 0.050613 |
| 46 | rs7741418 | 6 | 30063733 | 0.0455012 |
| 46 | rs28780082 | 6 | 30064129 | 0.0455012 |
| 46 | rs1156533 | 6 | 30065149 | 0.279255 |
| 46 | rs1264703 | 6 | 30065416 | 0.00632802 |
| 46 | rs1264702 | 6 | 30065575 | 0.00786582 |
| 46 | rs916570 | 6 | 30066031 | 0.0196029 |
| 46 | rs916569 | 6 | 30066097 | 0.00245904 |
| 46 | rs1264700 | 6 | 30066631 | 0.0027262 |
| 46 | rs1264699 | 6 | 30066659 | 0.00302381 |
| 46 | rs6457148 | 6 | 30066713 | 0.00245904 |
| 46 | rs6457149 | 6 | 30066714 | 0.00245904 |
| 46 | rs1264697 | 6 | 30067568 | 0.00245904 |

| | | | | |
|---|---|---|---|---|
| 46 | rs1116222 | 6 | 30071279 | 0.00295486 |
| 46 | rs1116221 | 6 | 30071330 | 0.00453386 |
| 47 | rs9489152 | 6 | 117734746 | 0.0269923 |
| 47 | rs3777981 | 6 | 117735255 | 0.00506666 |
| 47 | rs1853148 | 6 | 117750200 | 0.0529249 |
| 47 | rs4945584 | 6 | 117750980 | 0.0529249 |
| 47 | rs9372480 | 6 | 117760579 | 0.0903498 |
| 47 | rs9374662 | 6 | 117777804 | 0.0320171 |
| 47 | rs2180811 | 6 | 117780158 | 0.0637766 |
| 47 | rs4946256 | 6 | 117781276 | 0.0227703 |
| 47 | rs9374663 | 6 | 117782634 | 0.0536334 |
| 47 | rs6937083 | 6 | 117785308 | 0.0536334 |
| 47 | rs9387478 | 6 | 117786180 | 0.186041 |
| 47 | rs34882116 | 6 | 117789497 | 0.00704462 |
| 47 | rs2883418 | 6 | 117791751 | 0.0758855 |
| 47 | rs9387479 | 6 | 117792612 | 0.0536334 |
| 47 | rs6940922 | 6 | 117798204 | 0.0637766 |
| 47 | rs2354346 | 6 | 117798509 | 0.0637766 |
| 47 | rs9489213 | 6 | 117811706 | 0.00558233 |
| 47 | rs9320604 | 6 | 117816045 | 0.00558233 |
| 47 | rs9481728 | 6 | 117817165 | 0.00661069 |
| 47 | rs929057 | 6 | 117818911 | 0.00661069 |
| 47 | rs2057314 | 6 | 117819357 | 0.00661069 |
| 47 | rs9688361 | 6 | 117821888 | 0.00558233 |
| 47 | rs4946260 | 6 | 117822993 | 0.00661069 |
| 47 | rs10782186 | 6 | 117823508 | 0.00558233 |
| 48 | rs1535616 | 6 | 134171132 | 0.0133254 |
| 48 | rs7752775 | 6 | 134172346 | 0.0133254 |
| 48 | rs2105092 | 6 | 134184972 | 0.0219563 |
| 48 | rs10457618 | 6 | 134188626 | 0.0117766 |
| 48 | rs12207772 | 6 | 134189834 | 0.0117766 |
| 48 | rs13218281 | 6 | 134193714 | 0.0150856 |
| 48 | rs17062991 | 6 | 134193910 | 0.0170872 |
| 48 | rs12192720 | 6 | 134195719 | 0.0282712 |
| 48 | rs7769943 | 6 | 134196327 | 0.0249081 |
| 48 | rs7769954 | 6 | 134196381 | 0.032105 |
| 48 | rs12524865 | 6 | 134196674 | 0.0364774 |
| 48 | rs1967917 | 6 | 134198175 | 0.0282712 |
| 48 | rs2327426 | 6 | 134202690 | 0.262612 |
| 48 | rs12194592 | 6 | 134204247 | 0.228635 |
| 48 | rs6569912 | 6 | 134206802 | 0.0141161 |
| 48 | rs2327429 | 6 | 134209837 | 0.201041 |
| 49 | rs851981 | 6 | 152027074 | 0.0148569 |
| 49 | rs851978 | 6 | 152029556 | 0.0458466 |
| 49 | rs851977 | 6 | 152029608 | 0.0458466 |
| 49 | rs851976 | 6 | 152030008 | 0.0523599 |
| 49 | rs851975 | 6 | 152031303 | 0.0615935 |
| 49 | rs1101081 | 6 | 152032917 | 0.0401642 |
| 49 | rs1101080 | 6 | 152033643 | 0.0352042 |
| 49 | rs3020338 | 6 | 152034062 | 0.0905967 |
| 49 | rs13216081 | 6 | 152034624 | 0.0477355 |
| 49 | rs13216104 | 6 | 152034758 | 0.0542108 |

| | | | | |
|---|---|---|---|---|
| 49 | rs13201871 | 6 | 152034820 | 0.0542108 |
| 49 | rs6932703 | 6 | 152038240 | 0.0477355 |
| 49 | rs10872678 | 6 | 152039964 | 0.0401642 |
| 49 | rs12526447 | 6 | 152040125 | 0.0326842 |
| 49 | rs12525163 | 6 | 152040291 | 0.0183541 |
| 49 | rs3020339 | 6 | 152040615 | 0.0542108 |
| 49 | rs10484921 | 6 | 152042260 | 0.0370653 |
| 49 | rs11756568 | 6 | 152042413 | 0.0308727 |
| 49 | rs7772579 | 6 | 152042502 | 0.0208861 |
| 49 | rs3020340 | 6 | 152043290 | 0.0237797 |
| 49 | rs7749650 | 6 | 152044872 | 0.0401642 |
| 49 | rs7749659 | 6 | 152044884 | 0.0615935 |
| 50 | rs6941835 | 6 | 152356270 | 0.0209474 |
| 50 | rs9397472 | 6 | 152357047 | 0.0193994 |
| 50 | rs9383961 | 6 | 152361239 | 0.0193994 |
| 50 | rs4289664 | 6 | 152361383 | 0.0209474 |
| 50 | rs60693153 | 6 | 152366829 | 0.0193994 |
| 50 | rs9341004 | 6 | 152370248 | 0.0244385 |
| 50 | rs9397482 | 6 | 152379539 | 0.0179693 |
| 50 | rs3798568 | 6 | 152383383 | 0.0193994 |
| 50 | rs3778080 | 6 | 152385236 | 0.0193994 |
| 50 | rs35807076 | 6 | 152398814 | 0.0193994 |
| 50 | rs12175682 | 6 | 152403253 | 0.0244385 |
| 50 | rs3822990 | 6 | 152405965 | 0.0209474 |
| 50 | rs35494677 | 6 | 152406628 | 0.0209474 |
| 50 | rs66465244 | 6 | 152407061 | 0.0456344 |
| 50 | rs67933127 | 6 | 152407150 | 0.0456344 |
| 50 | rs13201080 | 6 | 152407782 | 0.0493835 |
| 50 | rs17082104 | 6 | 152407839 | 0.0421783 |
| 50 | rs3778090 | 6 | 152408028 | 0.0534512 |
| 50 | rs3778092 | 6 | 152408273 | 0.0493835 |
| 50 | rs9322355 | 6 | 152408649 | 0.0534512 |
| 50 | rs9322356 | 6 | 152408659 | 0.0534512 |
| 50 | rs9322357 | 6 | 152409236 | 0.0534512 |
| 50 | rs9322358 | 6 | 152409263 | 0.0534512 |
| 50 | rs9322359 | 6 | 152410022 | 0.0534512 |
| 50 | rs3778093 | 6 | 152411179 | 0.0493835 |
| 50 | rs3778094 | 6 | 152411198 | 0.0493835 |
| 50 | rs3778099 | 6 | 152418575 | 0.0338908 |
| 51 | rs6605526 | 6 | 169604100 | 0.118852 |
| 51 | rs7756742 | 6 | 169604132 | 0.118852 |
| 51 | rs7771838 | 6 | 169604158 | 0.118852 |
| 51 | rs6605527 | 6 | 169605973 | 0.139861 |
| 51 | rs9505932 | 6 | 169606389 | 0.118852 |
| 51 | rs9505933 | 6 | 169606443 | 0.118852 |
| 51 | rs3253 | 6 | 169616112 | 0.22869 |
| 52 | rs17171680 | 7 | 40364448 | 0.0121421 |
| 52 | rs10256791 | 7 | 40364726 | 0.0121421 |
| 52 | rs12701818 | 7 | 40367581 | 0.0171583 |
| 52 | rs6462972 | 7 | 40367949 | 0.0144294 |
| 52 | rs4143205 | 7 | 40368296 | 0.020416 |
| 52 | rs12701819 | 7 | 40370405 | 0.0144294 |

| | | | | |
|---|---|---|---|---|
| 52 | rs4452702 | 7 | 40371871 | 0.0144294 |
| 52 | rs11764504 | 7 | 40372958 | 0.0171583 |
| 52 | rs59533139 | 7 | 40375765 | 0.020416 |
| 52 | rs4538778 | 7 | 40378967 | 0.041742 |
| 52 | rs4598149 | 7 | 40379073 | 0.0600557 |
| 52 | rs7807217 | 7 | 40379594 | 0.0600557 |
| 52 | rs10281352 | 7 | 40383936 | 0.0243075 |
| 52 | rs11770113 | 7 | 40384494 | 0.0600557 |
| 52 | rs11772069 | 7 | 40386317 | 0.0721062 |
| 52 | rs10951634 | 7 | 40386835 | 0.0721062 |
| 52 | rs6462976 | 7 | 40396300 | 0.0866315 |
| 52 | rs10280188 | 7 | 40398139 | 0.0144294 |
| 52 | rs6944436 | 7 | 40401940 | 0.0289587 |
| 52 | rs6945101 | 7 | 40402430 | 0.0243075 |
| 52 | rs6965764 | 7 | 40409406 | 0.0243075 |
| 52 | rs17171704 | 7 | 40421488 | 0.0290893 |
| 52 | rs4421260 | 7 | 40424908 | 0.0171583 |
| 52 | rs10250287 | 7 | 40436763 | 0.0121421 |
| 52 | rs12533200 | 7 | 40437776 | 0.0144294 |
| 52 | rs16880229 | 7 | 40442741 | 0.020416 |
| 52 | rs9690724 | 7 | 40443252 | 0.0171583 |
| 52 | rs11980965 | 7 | 40460189 | 0.0289587 |
| 52 | rs3890819 | 7 | 40461332 | 0.04915 |
| 52 | rs10245007 | 7 | 40464894 | 0.0243075 |
| 52 | rs7782189 | 7 | 40472892 | 0.0289587 |
| 53 | rs11760406 | 7 | 55112410 | 0.00752432 |
| 53 | rs4947488 | 7 | 55116555 | 0.0178973 |
| 53 | rs4947965 | 7 | 55121812 | 0.0242989 |
| 53 | rs11773818 | 7 | 55123968 | 0.0178973 |
| 53 | rs11766798 | 7 | 55124319 | 0.0153712 |
| 53 | rs7795564 | 7 | 55124829 | 0.553309 |
| 53 | rs17335891 | 7 | 55131064 | 0.00976694 |
| 53 | rs12669701 | 7 | 55131670 | 0.0150043 |
| 53 | rs917879 | 7 | 55137258 | 0.0193515 |
| 53 | rs17514740 | 7 | 55146021 | 0.0763282 |
| 53 | rs12535578 | 7 | 55154586 | 0.0280383 |
| 53 | rs2302535 | 7 | 55154688 | 0.11798 |
| 53 | rs11238349 | 7 | 55156071 | 0.038447 |
| 53 | rs4947972 | 7 | 55161043 | 0.0100917 |
| 54 | rs11769827 | 7 | 73313856 | 0.56898 |
| 54 | rs11770437 | 7 | 73319809 | 0.411719 |
| 55 | rs4556017 | 7 | 100632790 | 0.961771 |
| 56 | rs6979964 | 7 | 102420427 | 0.0384524 |
| 56 | rs2430070 | 7 | 102420805 | 0.0568524 |
| 56 | rs7778418 | 7 | 102429704 | 0.315804 |
| 56 | rs10953374 | 7 | 102442173 | 0.045209 |
| 56 | rs6969054 | 7 | 102446569 | 0.0555094 |
| 56 | rs6949391 | 7 | 102446863 | 0.0555094 |
| 56 | rs10257317 | 7 | 102447056 | 0.0555094 |
| 56 | rs10279449 | 7 | 102462636 | 0.0838346 |
| 56 | rs1541519 | 7 | 102466741 | 0.0681971 |
| 56 | rs10271184 | 7 | 102477176 | 0.103119 |

| | | | | |
|---|---|---|---|---|
| 56 | rs7800548 | 7 | 102481842 | 0.0681971 |
| 56 | rs7794668 | 7 | 102510416 | 0.00894085 |
| 57 | rs806188 | 7 | 127268806 | 0.0233688 |
| 57 | rs806174 | 7 | 127291072 | 0.147321 |
| 57 | rs806172 | 7 | 127297002 | 0.0922653 |
| 57 | rs806169 | 7 | 127300668 | 0.201826 |
| 57 | rs806166 | 7 | 127309934 | 0.0580799 |
| 57 | rs712707 | 7 | 127322614 | 0.0271594 |
| 57 | rs712715 | 7 | 127333136 | 0.0498325 |
| 57 | rs712718 | 7 | 127346950 | 0.0498325 |
| 57 | rs712719 | 7 | 127357644 | 0.0233688 |
| 57 | rs56185520 | 7 | 127364310 | 0.0201187 |
| 57 | rs12667197 | 7 | 127369147 | 0.0233688 |
| 57 | rs12540484 | 7 | 127370040 | 0.0233688 |
| 57 | rs4731377 | 7 | 127381201 | 0.0149369 |
| 57 | rs17151229 | 7 | 127382155 | 0.0173304 |
| 57 | rs11772303 | 7 | 127398962 | 0.0128814 |
| 57 | rs10487497 | 7 | 127400667 | 0.0111149 |
| 57 | rs62481385 | 7 | 127410711 | 0.0111149 |
| 57 | rs765971 | 7 | 127420642 | 0.00959618 |
| 57 | rs59140894 | 7 | 127430797 | 0.00828965 |
| 57 | rs3757780 | 7 | 127435520 | 0.0111149 |
| 57 | rs2402871 | 7 | 127439519 | 0.00959618 |
| 57 | rs3757778 | 7 | 127456607 | 0.00828965 |
| 57 | rs17151377 | 7 | 127458009 | 0.00619652 |
| 57 | rs67338333 | 7 | 127461701 | 0.00828965 |
| 57 | rs12539411 | 7 | 127468928 | 0.00619652 |
| 57 | rs3824003 | 7 | 127469624 | 0.00716505 |
| 57 | rs1419967 | 7 | 127474435 | 0.00716505 |
| 57 | rs12668077 | 7 | 127476253 | 0.00959618 |
| 57 | rs62481416 | 7 | 127478125 | 0.00716505 |
| 57 | rs4728079 | 7 | 127480364 | 0.00716505 |
| 57 | rs58106688 | 7 | 127483211 | 0.00828965 |
| 57 | rs62481419 | 7 | 127488007 | 0.00828965 |
| 57 | rs2058457 | 7 | 127493657 | 0.00716505 |
| 57 | rs882694 | 7 | 127496768 | 0.00716505 |
| 57 | rs10954164 | 7 | 127533877 | 0.00716505 |
| 58 | rs2912053 | 8 | 6645793 | 0.98466 |
| 59 | rs4592028 | 8 | 22449484 | 0.00452658 |
| 59 | rs11785755 | 8 | 22453223 | 0.0214102 |
| 59 | rs11783129 | 8 | 22453426 | 0.0185371 |
| 59 | rs3735894 | 8 | 22454826 | 0.0214102 |
| 59 | rs71513892 | 8 | 22456517 | 0.0185371 |
| 59 | rs2272718 | 8 | 22457388 | 0.0214102 |
| 59 | rs746011 | 8 | 22457804 | 0.0104757 |
| 59 | rs34268501 | 8 | 22460204 | 0.0214102 |
| 59 | rs3735901 | 8 | 22462374 | 0.0443818 |
| 59 | rs11777808 | 8 | 22462755 | 0.0383175 |
| 59 | rs11778693 | 8 | 22462852 | 0.0383175 |
| 59 | rs7843828 | 8 | 22463623 | 0.0443818 |
| 59 | rs2291231 | 8 | 22463912 | 0.0443818 |
| 59 | rs2291232 | 8 | 22464064 | 0.0443818 |

| | | | | |
|---|---|---|---|---|
| 59 | rs2291234 | 8 | 22464390 | 0.0383175 |
| 59 | rs34269854 | 8 | 22465160 | 0.0331005 |
| 59 | rs34027561 | 8 | 22465385 | 0.0443818 |
| 59 | rs6999893 | 8 | 22465427 | 0.00440624 |
| 59 | rs11136092 | 8 | 22466488 | 0.0383175 |
| 59 | rs7843616 | 8 | 22466541 | 0.0514351 |
| 59 | rs13271626 | 8 | 22467760 | 0.0691999 |
| 59 | rs6986550 | 8 | 22467870 | 0.0058534 |
| 59 | rs720746 | 8 | 22471399 | 0.0443818 |
| 59 | rs3736147 | 8 | 22471824 | 0.0331005 |
| 59 | rs11781149 | 8 | 22473158 | 0.0443818 |
| 59 | rs7843128 | 8 | 22473465 | 0.00779436 |
| 59 | rs6558167 | 8 | 22473850 | 0.00675251 |
| 59 | rs57594397 | 8 | 22475657 | 0.0383175 |
| 59 | rs11136093 | 8 | 22479988 | 0.0104757 |
| 59 | rs1809452 | 8 | 22491397 | 0.0139194 |
| 59 | rs35952885 | 8 | 22491622 | 0.0139194 |
| 59 | rs11776549 | 8 | 22493124 | 0.012072 |
| 59 | rs4872527 | 8 | 22496048 | 0.00909563 |
| 59 | rs4242434 | 8 | 22501830 | 0.0160586 |
| 59 | rs13264187 | 8 | 22506098 | 0.0139194 |
| 59 | rs10099846 | 8 | 22534178 | 0.00452658 |
| 59 | rs10095121 | 8 | 22538426 | 0.00452658 |
| 60 | rs7844508 | 8 | 71602995 | 0.010335 |
| 60 | rs13252808 | 8 | 71616655 | 0.0527565 |
| 60 | rs13280922 | 8 | 71625398 | 0.0527565 |
| 60 | rs11989644 | 8 | 71641196 | 0.0527565 |
| 60 | rs13272884 | 8 | 71641648 | 0.0527565 |
| 60 | rs13281864 | 8 | 71642573 | 0.0527565 |
| 60 | rs13269690 | 8 | 71644807 | 0.0380313 |
| 60 | rs7016334 | 8 | 71654406 | 0.0380313 |
| 60 | rs28376252 | 8 | 71659887 | 0.0380313 |
| 60 | rs6991708 | 8 | 71668528 | 0.0527565 |
| 60 | rs7838421 | 8 | 71672323 | 0.0380313 |
| 60 | rs7012796 | 8 | 71676193 | 0.0380313 |
| 60 | rs2380689 | 8 | 71681983 | 0.0380313 |
| 60 | rs1838392 | 8 | 71682583 | 0.0527565 |
| 60 | rs7845516 | 8 | 71685822 | 0.0527565 |
| 60 | rs7013495 | 8 | 71693517 | 0.0380313 |
| 60 | rs10429277 | 8 | 71701391 | 0.0527565 |
| 60 | rs11994908 | 8 | 71721619 | 0.0198009 |
| 60 | rs13273979 | 8 | 71737322 | 0.0198009 |
| 60 | rs62508859 | 8 | 71740934 | 0.0143009 |
| 60 | rs13271606 | 8 | 71741723 | 0.010335 |
| 60 | rs6983845 | 8 | 71746480 | 0.0074736 |
| 60 | rs13255849 | 8 | 71752753 | 0.0198009 |
| 60 | rs6472548 | 8 | 71754299 | 0.0143009 |
| 60 | rs1156956 | 8 | 71755038 | 0.0143009 |
| 60 | rs6984056 | 8 | 71771121 | 0.010335 |
| 60 | rs4446768 | 8 | 71777589 | 0.010335 |
| 60 | rs7003794 | 8 | 71789146 | 0.0074736 |
| 60 | rs13255749 | 8 | 71801821 | 0.0074736 |

| | | | | |
|---|---|---|---|---|
| 60 | rs11985482 | 8 | 71810578 | 0.0074736 |
| 60 | rs7838671 | 8 | 71811292 | 0.0074736 |
| 60 | rs4581086 | 8 | 71814640 | 0.010335 |
| 60 | rs13248937 | 8 | 71818749 | 0.010335 |
| 60 | rs6984663 | 8 | 71827665 | 0.010335 |
| 61 | rs2581260 | 8 | 77493526 | 0.995876 |
| 62 | rs10956488 | 8 | 130717755 | 0.961845 |
| 63 | rs7461753 | 8 | 144981498 | 0.00597241 |
| 63 | rs7015048 | 8 | 144984283 | 0.00306258 |
| 63 | rs7840395 | 8 | 144984956 | 0.00306258 |
| 63 | rs4977183 | 8 | 144985200 | 0.00306258 |
| 63 | rs11777835 | 8 | 144986103 | 0.00412201 |
| 63 | rs7155 | 8 | 144989462 | 0.00294341 |
| 63 | rs1065837 | 8 | 144990335 | 0.00343816 |
| 63 | rs55836855 | 8 | 144999417 | 0.00343816 |
| 63 | rs55646585 | 8 | 144999621 | 0.0167981 |
| 63 | rs56117011 | 8 | 144999642 | 0.00343816 |
| 63 | rs35916068 | 8 | 144999684 | 0.00343816 |
| 63 | rs7002152 | 8 | 145000056 | 0.00401845 |
| 63 | rs7016860 | 8 | 145000221 | 0.00401845 |
| 63 | rs6993953 | 8 | 145000321 | 0.00401845 |
| 63 | rs7006770 | 8 | 145000602 | 0.00401845 |
| 63 | rs7017644 | 8 | 145000740 | 0.00401845 |
| 63 | rs7017789 | 8 | 145000809 | 0.00401845 |
| 63 | rs56401829 | 8 | 145000898 | 0.00469947 |
| 63 | rs55895668 | 8 | 145001031 | 0.00401845 |
| 63 | rs55984967 | 8 | 145001300 | 0.00469947 |
| 63 | rs55812715 | 8 | 145001311 | 0.00469947 |
| 63 | rs11782890 | 8 | 145001372 | 0.00469947 |
| 63 | rs11136333 | 8 | 145001509 | 0.00469947 |
| 63 | rs11136334 | 8 | 145001588 | 0.00549917 |
| 63 | rs3135109 | 8 | 145001784 | 0.00549917 |
| 63 | rs11993233 | 8 | 145002283 | 0.00549917 |
| 63 | rs62522555 | 8 | 145002431 | 0.00549917 |
| 63 | rs72693398 | 8 | 145002536 | 0.00549917 |
| 63 | rs11136335 | 8 | 145003777 | 0.00597241 |
| 63 | rs62522556 | 8 | 145005450 | 0.0129906 |
| 63 | rs11136336 | 8 | 145007187 | 0.0273508 |
| 63 | rs7003580 | 8 | 145007534 | 0.0447716 |
| 63 | rs6989119 | 8 | 145007947 | 0.0322149 |
| 63 | rs11136337 | 8 | 145007989 | 0.00950386 |
| 63 | rs11782331 | 8 | 145008342 | 0.0273508 |
| 63 | rs11783772 | 8 | 145008443 | 0.0273508 |
| 63 | rs11783799 | 8 | 145008560 | 0.0273508 |
| 63 | rs11782433 | 8 | 145008706 | 0.0273508 |
| 63 | rs6994460 | 8 | 145008931 | 0.0528279 |
| 63 | rs6983908 | 8 | 145009593 | 0.0447716 |
| 63 | rs11784762 | 8 | 145009610 | 0.0197502 |
| 63 | rs6988767 | 8 | 145010305 | 0.0447716 |
| 63 | rs56034811 | 8 | 145010514 | 0.0447716 |
| 63 | rs11784417 | 8 | 145010752 | 0.0167981 |
| 63 | rs13439519 | 8 | 145012206 | 0.0379665 |

| | | | | |
|---|---|---|---|---|
| 63 | rs11136338 | 8 | 145012466 | 0.0121733 |
| 63 | rs11784501 | 8 | 145013315 | 0.0447716 |
| 63 | rs11136339 | 8 | 145013345 | 0.023235 |
| 63 | rs4073081 | 8 | 145013893 | 0.0167981 |
| 63 | rs4073082 | 8 | 145013949 | 0.0111081 |
| 63 | rs12541377 | 8 | 145014424 | 0.00597241 |
| 63 | rs112979447 | 8 | 145014732 | 0.00597241 |
| 63 | rs56334578 | 8 | 145020025 | 0.00597241 |
| 63 | rs12543539 | 8 | 145020477 | 0.00555992 |
| 63 | rs12549853 | 8 | 145020636 | 0.00696873 |
| 63 | rs7464572 | 8 | 145021167 | 0.00597241 |
| 63 | rs7462197 | 8 | 145021428 | 0.00813587 |
| 63 | rs7832643 | 8 | 145022657 | 0.0129906 |
| 63 | rs55754956 | 8 | 145023739 | 0.00751564 |
| 63 | rs11778018 | 8 | 145026450 | 0.0208481 |
| 63 | rs62523994 | 8 | 145026582 | 0.0152007 |
| 63 | rs55786556 | 8 | 145027210 | 0.0177968 |
| 63 | rs58579887 | 8 | 145028587 | 0.0244364 |
| 63 | rs11777239 | 8 | 145031265 | 0.00597241 |
| 63 | rs10092179 | 8 | 145032302 | 0.0177968 |
| 63 | rs11780978 | 8 | 145034852 | 0.00754342 |
| 63 | rs7357417 | 8 | 145035342 | 0.00884282 |
| 63 | rs7010330 | 8 | 145036615 | 0.00549917 |
| 63 | rs11783655 | 8 | 145037573 | 0.00643878 |
| 63 | rs72695409 | 8 | 145039400 | 0.00549917 |
| 63 | rs6993022 | 8 | 145040108 | 0.00401845 |
| 63 | rs11787365 | 8 | 145041333 | 0.00377235 |
| 64 | rs10757277 | 9 | 22124450 | 0.02427 |
| 64 | rs10757278 | 9 | 22124477 | 0.02427 |
| 64 | rs1333047 | 9 | 22124504 | 0.520459 |
| 64 | rs4977575 | 9 | 22124744 | 0.392585 |
| 65 | rs10992304 | 9 | 95085365 | 0.0158626 |
| 65 | rs10116813 | 9 | 95090360 | 0.00976714 |
| 65 | rs10820968 | 9 | 95106443 | 0.0140407 |
| 65 | rs7030920 | 9 | 95122952 | 0.009434 |
| 65 | rs7045572 | 9 | 95137964 | 0.0158626 |
| 65 | rs1121978 | 9 | 95140351 | 0.0124344 |
| 65 | rs10761155 | 9 | 95153729 | 0.0110175 |
| 65 | rs2761677 | 9 | 95180558 | 0.0110175 |
| 65 | rs6479422 | 9 | 95190694 | 0.00866315 |
| 65 | rs12342019 | 9 | 95192508 | 0.00866315 |
| 65 | rs11792019 | 9 | 95195229 | 0.00866315 |
| 65 | rs12336415 | 9 | 95195458 | 0.00866315 |
| 65 | rs10115290 | 9 | 95195858 | 0.00271529 |
| 65 | rs1535754 | 9 | 95201917 | 0.00271529 |
| 65 | rs1535755 | 9 | 95202031 | 0.0106117 |
| 65 | rs1535756 | 9 | 95202032 | 0.0134462 |
| 65 | rs10117627 | 9 | 95203953 | 0.00390224 |
| 65 | rs10761158 | 9 | 95205322 | 0.00271529 |
| 65 | rs7873397 | 9 | 95205928 | 0.00664841 |
| 65 | rs10761160 | 9 | 95208701 | 0.0110175 |
| 65 | rs7850528 | 9 | 95208992 | 0.0110175 |

| | | | | |
|---|---|---|---|---|
| 65 | rs10992341 | 9 | 95211699 | 0.00682596 |
| 65 | rs10992342 | 9 | 95212068 | 0.00606379 |
| 65 | rs7022562 | 9 | 95216340 | 0.00306245 |
| 65 | rs7033979 | 9 | 95223980 | 0.00866315 |
| 65 | rs13301537 | 9 | 95229047 | 0.00866315 |
| 65 | rs10992349 | 9 | 95233887 | 0.00866315 |
| 65 | rs12341093 | 9 | 95234182 | 0.00664841 |
| 65 | rs12336884 | 9 | 95234183 | 0.00746729 |
| 65 | rs12343820 | 9 | 95236314 | 0.00271529 |
| 65 | rs3739606 | 9 | 95237222 | 0.00271529 |
| 65 | rs7868013 | 9 | 95240173 | 0.00866315 |
| 65 | rs7031567 | 9 | 95241363 | 0.00306245 |
| 65 | rs7860786 | 9 | 95245473 | 0.00271529 |
| 65 | rs11794346 | 9 | 95247288 | 0.0076879 |
| 65 | rs1980848 | 9 | 95248571 | 0.0076879 |
| 65 | rs4744135 | 9 | 95251874 | 0.00384741 |
| 65 | rs12341817 | 9 | 95252152 | 0.0076879 |
| 65 | rs10820980 | 9 | 95253342 | 0.0076879 |
| 65 | rs11788523 | 9 | 95256593 | 0.00976714 |
| 65 | rs10514815 | 9 | 95256695 | 0.00306245 |
| 65 | rs10429459 | 9 | 95258119 | 0.00345596 |
| 65 | rs927865 | 9 | 95261327 | 0.00433407 |
| 65 | rs3780347 | 9 | 95266812 | 0.00440865 |
| 65 | rs4743873 | 9 | 95270781 | 0.0124344 |
| 65 | rs968040 | 9 | 95279985 | 0.00895757 |
| 65 | rs12338938 | 9 | 95281459 | 0.00846326 |
| 65 | rs7847534 | 9 | 95282653 | 0.0101289 |
| 65 | rs9299405 | 9 | 95284873 | 0.0129719 |
| 65 | rs4743874 | 9 | 95288323 | 0.00744612 |
| 65 | rs4744137 | 9 | 95288352 | 0.0114595 |
| 65 | rs747628 | 9 | 95289532 | 0.0146919 |
| 65 | rs747629 | 9 | 95289940 | 0.00655512 |
| 65 | rs2104533 | 9 | 95290456 | 0.00962505 |
| 65 | rs2094281 | 9 | 95290654 | 0.0146919 |
| 65 | rs4744138 | 9 | 95291909 | 0.0146919 |
| 65 | rs10118939 | 9 | 95293641 | 0.0146919 |
| 65 | rs987553 | 9 | 95298694 | 0.0101289 |
| 65 | rs7872610 | 9 | 95299137 | 0.00479266 |
| 65 | rs7872644 | 9 | 95299191 | 0.00577417 |
| 65 | rs2895219 | 9 | 95300559 | 0.00744612 |
| 65 | rs4744139 | 9 | 95301397 | 0.00655512 |
| 65 | rs3927488 | 9 | 95302396 | 0.00744612 |
| 65 | rs7864575 | 9 | 95304647 | 0.00792602 |
| 65 | rs10125450 | 9 | 95306082 | 0.00655512 |
| 65 | rs1075397 | 9 | 95306844 | 0.00655512 |
| 65 | rs10114912 | 9 | 95307863 | 0.0101289 |
| 65 | rs10114932 | 9 | 95307901 | 0.00895757 |
| 65 | rs10118107 | 9 | 95308113 | 0.00655512 |
| 65 | rs10992365 | 9 | 95308895 | 0.00508928 |
| 65 | rs10117680 | 9 | 95312407 | 0.00448829 |
| 65 | rs10761163 | 9 | 95313287 | 0.00621567 |
| 65 | rs13301492 | 9 | 95315685 | 0.00508928 |

| | | | | |
|---|---|---|---|---|
| 65 | rs13301652 | 9 | 95315785 | 0.00508928 |
| 65 | rs7863049 | 9 | 95316645 | 0.00508928 |
| 65 | rs7849788 | 9 | 95316649 | 0.00508928 |
| 65 | rs7849927 | 9 | 95316768 | 0.00448829 |
| 65 | rs10992366 | 9 | 95317214 | 0.00621567 |
| 65 | rs7029939 | 9 | 95319891 | 0.00621567 |
| 65 | rs10820989 | 9 | 95322020 | 0.00448829 |
| 65 | rs10761164 | 9 | 95323548 | 0.00448829 |
| 65 | rs2026585 | 9 | 95324986 | 0.00349707 |
| 65 | rs7865019 | 9 | 95325631 | 0.00621567 |
| 65 | rs7873390 | 9 | 95329120 | 0.00448829 |
| 65 | rs10115409 | 9 | 95334627 | 0.00301252 |
| 65 | rs10992375 | 9 | 95337993 | 0.00396062 |
| 65 | rs10820994 | 9 | 95349517 | 0.00433407 |
| 65 | rs4744141 | 9 | 95352140 | 0.00384741 |
| 65 | rs724102 | 9 | 95383290 | 0.0076879 |
| 65 | rs7852698 | 9 | 95396409 | 0.00682596 |
| 65 | rs7869521 | 9 | 95411059 | 0.00606379 |
| 65 | rs7020104 | 9 | 95416524 | 0.00682596 |
| 65 | rs10992393 | 9 | 95420983 | 0.00682596 |
| 65 | rs4743875 | 9 | 95423674 | 0.00606379 |
| 65 | rs13284106 | 9 | 95433845 | 0.0124344 |
| 65 | rs4744143 | 9 | 95438627 | 0.0110175 |
| 65 | rs7865118 | 9 | 95439897 | 0.0110175 |
| 65 | rs13286566 | 9 | 95473178 | 0.00488494 |
| 65 | rs10491804 | 9 | 95487407 | 0.0202779 |
| 65 | rs10821008 | 9 | 95487497 | 0.00488494 |
| 65 | rs755209 | 9 | 95489671 | 0.0333432 |
| 65 | rs10125587 | 9 | 95499882 | 0.00488494 |
| 65 | rs4743876 | 9 | 95512216 | 0.0294222 |
| 65 | rs10992450 | 9 | 95520702 | 0.0259757 |
| 65 | rs10124058 | 9 | 95522882 | 0.00550879 |
| 65 | rs7043984 | 9 | 95529408 | 0.00550879 |
| 65 | rs10992457 | 9 | 95531261 | 0.00550879 |
| 65 | rs7046335 | 9 | 95533361 | 0.0229447 |
| 65 | rs6479431 | 9 | 95546889 | 0.0217173 |
| 65 | rs10122383 | 9 | 95554675 | 0.0294222 |
| 66 | rs7026755 | 9 | 119156088 | 0.103422 |
| 66 | rs10817884 | 9 | 119156565 | 0.162631 |
| 66 | rs1858015 | 9 | 119167220 | 0.557511 |
| 66 | rs4837514 | 9 | 119181214 | 0.126545 |
| 67 | rs687621 | 9 | 136137065 | 0.0126855 |
| 67 | rs687289 | 9 | 136137106 | 0.0126855 |
| 67 | rs514659 | 9 | 136142203 | 0.206507 |
| 67 | rs545971 | 9 | 136143372 | 0.0174704 |
| 67 | rs8176663 | 9 | 136144427 | 0.0174704 |
| 67 | rs491626 | 9 | 136144873 | 0.0174704 |
| 67 | rs492488 | 9 | 136144960 | 0.0174704 |
| 67 | rs493246 | 9 | 136144994 | 0.0240737 |
| 67 | rs495203 | 9 | 136145240 | 0.0240737 |
| 67 | rs582118 | 9 | 136145471 | 0.0174704 |
| 67 | rs582094 | 9 | 136145484 | 0.0174704 |

| | | | | |
|---|---|---|---|---|
| 67 | rs2769071 | 9 | 136145974 | 0.0174704 |
| 67 | rs677355 | 9 | 136146046 | 0.230725 |
| 67 | rs676996 | 9 | 136146077 | 0.230725 |
| 67 | rs676457 | 9 | 136146227 | 0.0126855 |
| 67 | rs527210 | 9 | 136146431 | 0.0126855 |
| 67 | rs505922 | 9 | 136149229 | 0.0632025 |
| 68 | rs6482359 | 10 | 24330805 | 0.796779 |
| 68 | rs7070917 | 10 | 24335666 | 0.0928533 |
| 68 | rs10741038 | 10 | 24366458 | 0.0862095 |
| 69 | rs2363893 | 10 | 126714200 | 0.00420696 |
| 69 | rs3012075 | 10 | 126714966 | 0.00420696 |
| 69 | rs2938006 | 10 | 126716346 | 0.00599159 |
| 69 | rs3012065 | 10 | 126737579 | 0.494938 |
| 69 | rs3012066 | 10 | 126737997 | 0.435582 |
| 69 | rs2919290 | 10 | 126738533 | 0.00701758 |
| 70 | rs2957706 | 11 | 10377175 | 0.0448313 |
| 70 | rs2957707 | 11 | 10377258 | 0.0320041 |
| 70 | rs2957708 | 11 | 10377287 | 0.0320041 |
| 70 | rs2923106 | 11 | 10377434 | 0.0378684 |
| 70 | rs2923107 | 11 | 10377498 | 0.0320041 |
| 70 | rs2957709 | 11 | 10377739 | 0.0448313 |
| 70 | rs2923108 | 11 | 10377899 | 0.0448313 |
| 70 | rs2923109 | 11 | 10378309 | 0.0193821 |
| 70 | rs2957711 | 11 | 10378330 | 0.0228964 |
| 70 | rs2957712 | 11 | 10378334 | 0.0228964 |
| 70 | rs1450272 | 11 | 10378813 | 0.0228964 |
| 70 | rs2923110 | 11 | 10378840 | 0.0270626 |
| 70 | rs2957713 | 11 | 10378914 | 0.0193821 |
| 70 | rs2957714 | 11 | 10378960 | 0.0193821 |
| 70 | rs2923111 | 11 | 10379096 | 0.0228964 |
| 70 | rs1947256 | 11 | 10379806 | 0.0228964 |
| 70 | rs1947257 | 11 | 10379952 | 0.0228964 |
| 70 | rs1375997 | 11 | 10380703 | 0.0193821 |
| 70 | rs1375998 | 11 | 10380775 | 0.0228964 |
| 70 | rs2218793 | 11 | 10380828 | 0.0831999 |
| 70 | rs2957716 | 11 | 10380858 | 0.0228964 |
| 70 | rs2923112 | 11 | 10381176 | 0.0193821 |
| 70 | rs2923078 | 11 | 10381766 | 0.0831999 |
| 70 | rs2957717 | 11 | 10382184 | 0.0320041 |
| 70 | rs2957718 | 11 | 10382282 | 0.0193821 |
| 70 | rs2923079 | 11 | 10383382 | 0.016416 |
| 70 | rs2957721 | 11 | 10383427 | 0.0139114 |
| 70 | rs2957651 | 11 | 10384468 | 0.0139114 |
| 70 | rs2957653 | 11 | 10384872 | 0.0139114 |
| 70 | rs2957654 | 11 | 10384911 | 0.0531033 |
| 70 | rs2957655 | 11 | 10385377 | 0.0270626 |
| 70 | rs2923082 | 11 | 10386083 | 0.0270626 |
| 71 | rs4442541 | 11 | 10669172 | 0.18273 |
| 71 | rs7940646 | 11 | 10669228 | 0.18273 |
| 71 | rs4909945 | 11 | 10673739 | 0.221454 |
| 71 | rs4910165 | 11 | 10674044 | 0.325812 |
| 71 | rs10840457 | 11 | 10675738 | 0.019024 |

| | | | | |
|---|---|---|---|---|
| 71 | rs1863243 | 11 | 10677373 | 0.03234 |
| 72 | rs35184771 | 11 | 47475189 | 0.0317439 |
| 72 | rs1317149 | 11 | 47486885 | 0.0274355 |
| 72 | rs11039255 | 11 | 47495746 | 0.0205302 |
| 72 | rs10838724 | 11 | 47527052 | 0.0047725 |
| 72 | rs4752845 | 11 | 47539697 | 0.0317439 |
| 72 | rs34958982 | 11 | 47547046 | 0.0317439 |
| 72 | rs66749409 | 11 | 47568074 | 0.0274355 |
| 72 | rs12798346 | 11 | 47583121 | 0.023726 |
| 72 | rs56400411 | 11 | 47586376 | 0.0274355 |
| 72 | rs7945473 | 11 | 47589707 | 0.0367507 |
| 72 | rs2030166 | 11 | 47602729 | 0.0367507 |
| 72 | rs11605348 | 11 | 47606483 | 0.0205302 |
| 72 | rs12799623 | 11 | 47622139 | 0.0317439 |
| 72 | rs12419692 | 11 | 47624714 | 0.0274355 |
| 72 | rs1064608 | 11 | 47640429 | 0.0274355 |
| 72 | rs3817335 | 11 | 47643891 | 0.0317439 |
| 72 | rs4752856 | 11 | 47648042 | 0.0317439 |
| 72 | rs4752857 | 11 | 47655752 | 0.0425725 |
| 72 | rs12794570 | 11 | 47658314 | 0.0068379 |
| 72 | rs10838738 | 11 | 47663049 | 0.0425725 |
| 72 | rs12787646 | 11 | 47669665 | 0.0274355 |
| 72 | rs11039327 | 11 | 47674084 | 0.0317439 |
| 72 | rs12787112 | 11 | 47687147 | 0.0100487 |
| 72 | rs10838747 | 11 | 47716324 | 0.0100487 |
| 72 | rs10838748 | 11 | 47716381 | 0.0100487 |
| 72 | rs11039342 | 11 | 47716975 | 0.0115784 |
| 72 | rs11604825 | 11 | 47725306 | 0.0068379 |
| 72 | rs11602395 | 11 | 47726977 | 0.0115784 |
| 72 | rs11039348 | 11 | 47728617 | 0.0115784 |
| 72 | rs7130758 | 11 | 47749960 | 0.00659223 |
| 72 | rs17788930 | 11 | 47752775 | 0.00872619 |
| 72 | rs12803191 | 11 | 47756475 | 0.00659223 |
| 72 | rs34910028 | 11 | 47758449 | 0.00758227 |
| 72 | rs11602339 | 11 | 47761471 | 0.00758227 |
| 72 | rs10838757 | 11 | 47763016 | 0.00872619 |
| 72 | rs34923397 | 11 | 47769564 | 0.00758227 |
| 72 | rs12421210 | 11 | 47779586 | 0.00659223 |
| 72 | rs7927771 | 11 | 47781306 | 0.00573486 |
| 72 | rs12361031 | 11 | 47783076 | 0.00872619 |
| 72 | rs7947730 | 11 | 47786184 | 0.00659223 |
| 72 | rs35902101 | 11 | 47795169 | 0.00659223 |
| 72 | rs11039390 | 11 | 47796295 | 0.00659223 |
| 72 | rs11039391 | 11 | 47796662 | 0.00659223 |
| 72 | rs9909 | 11 | 47799775 | 0.00758227 |
| 72 | rs35805829 | 11 | 47805478 | 0.00659223 |
| 72 | rs2290850 | 11 | 47807774 | 0.00499197 |
| 72 | rs12787330 | 11 | 47812311 | 0.00573486 |
| 72 | rs7120333 | 11 | 47817441 | 0.00573486 |
| 72 | rs11605774 | 11 | 47832793 | 0.00573486 |
| 72 | rs12785833 | 11 | 47836062 | 0.00573486 |
| 72 | rs11039412 | 11 | 47841581 | 0.00573486 |

| | | | | |
|---|---|---|---|---|
| 72 | rs11039416 | 11 | 47846711 | 0.00573486 |
| 72 | rs35985502 | 11 | 47847428 | 0.00573486 |
| 72 | rs34128973 | 11 | 47851376 | 0.00573486 |
| 72 | rs4752873 | 11 | 47857520 | 0.00573486 |
| 72 | rs11039426 | 11 | 47863119 | 0.00573486 |
| 72 | rs4752797 | 11 | 47874364 | 0.00573486 |
| 72 | rs11039433 | 11 | 47877493 | 0.00573486 |
| 72 | rs12798109 | 11 | 47889850 | 0.00758227 |
| 72 | rs4752881 | 11 | 47898535 | 0.00872619 |
| 72 | rs12364432 | 11 | 47902883 | 0.0100487 |
| 72 | rs4752801 | 11 | 47907641 | 0.013349 |
| 72 | rs12361256 | 11 | 47910823 | 0.0153995 |
| 72 | rs12802244 | 11 | 47932666 | 0.00573486 |
| 72 | rs2930191 | 11 | 47946836 | 0.0103231 |
| 73 | rs2186797 | 11 | 70007770 | 0.969962 |
| 74 | rs2212450 | 11 | 112826867 | 0.19922 |
| 74 | rs2186710 | 11 | 112827048 | 0.059884 |
| 74 | rs4937872 | 11 | 112827715 | 0.12345 |
| 74 | rs10891480 | 11 | 112830526 | 0.0067964 |
| 74 | rs10891481 | 11 | 112830562 | 0.0067964 |
| 74 | rs10789929 | 11 | 112830663 | 0.0050305 |
| 74 | rs11214436 | 11 | 112830782 | 0.0067964 |
| 74 | rs7937151 | 11 | 112835024 | 0.0067964 |
| 74 | rs10750016 | 11 | 112837740 | 0.00920515 |
| 74 | rs2155281 | 11 | 112838338 | 0.00920515 |
| 74 | rs720023 | 11 | 112838867 | 0.00790714 |
| 74 | rs7948789 | 11 | 112839532 | 0.0107229 |
| 74 | rs7126748 | 11 | 112842976 | 0.00790714 |
| 74 | rs7110863 | 11 | 112843138 | 0.00790714 |
| 74 | rs2186709 | 11 | 112843616 | 0.00790714 |
| 74 | rs11214441 | 11 | 112846713 | 0.0067964 |
| 74 | rs2186707 | 11 | 112850643 | 0.00584533 |
| 74 | rs2155284 | 11 | 112850770 | 0.00584533 |
| 74 | rs2298527 | 11 | 112851961 | 0.0145778 |
| 74 | rs2212449 | 11 | 112852032 | 0.00584533 |
| 74 | rs1940729 | 11 | 112852192 | 0.00584533 |
| 74 | rs1940728 | 11 | 112852195 | 0.00584533 |
| 74 | rs2298526 | 11 | 112852464 | 0.00584533 |
| 74 | rs1940727 | 11 | 112852588 | 0.00584533 |
| 74 | rs1940726 | 11 | 112852611 | 0.00584533 |
| 74 | rs1940725 | 11 | 112852759 | 0.00584533 |
| 74 | rs1940724 | 11 | 112852946 | 0.00584533 |
| 74 | rs1954826 | 11 | 112853012 | 0.00584533 |
| 74 | rs4439550 | 11 | 112853538 | 0.0067964 |
| 74 | rs10736463 | 11 | 112854052 | 0.00584533 |
| 74 | rs6589353 | 11 | 112854935 | 0.00584533 |
| 74 | rs7950836 | 11 | 112856137 | 0.00584533 |
| 74 | rs1940720 | 11 | 112857202 | 0.0050305 |
| 74 | rs1940718 | 11 | 112859867 | 0.00584533 |
| 74 | rs1940716 | 11 | 112860288 | 0.00584533 |
| 74 | rs2155282 | 11 | 112860348 | 0.00584533 |
| 74 | rs7106434 | 11 | 112860579 | 0.0124988 |

| | | | | |
|---|---|---|---|---|
| 74 | rs7121047 | 11 | 112860893 | 0.00584533 |
| 74 | rs4466874 | 11 | 112861434 | 0.00584533 |
| 74 | rs4479020 | 11 | 112861443 | 0.00584533 |
| 74 | rs7115165 | 11 | 112862587 | 0.0050305 |
| 74 | rs4381397 | 11 | 112866269 | 0.00584533 |
| 74 | rs4144892 | 11 | 112866456 | 0.00584533 |
| 74 | rs1940702 | 11 | 112867061 | 0.0050305 |
| 74 | rs10891487 | 11 | 112869054 | 0.0050305 |
| 74 | rs1940701 | 11 | 112869404 | 0.00584533 |
| 74 | rs723599 | 11 | 112872305 | 0.0050305 |
| 74 | rs7127930 | 11 | 112874645 | 0.0050305 |
| 74 | rs10750019 | 11 | 112875768 | 0.00584533 |
| 74 | rs1940697 | 11 | 112876886 | 0.00584533 |
| 74 | rs9919557 | 11 | 112877408 | 0.0050305 |
| 74 | rs9919558 | 11 | 112877409 | 0.0050305 |
| 74 | rs7118907 | 11 | 112878254 | 0.00584533 |
| 74 | rs6589354 | 11 | 112878547 | 0.0141992 |
| 74 | rs4589334 | 11 | 112879456 | 0.00920515 |
| 74 | rs4294596 | 11 | 112880732 | 0.00584533 |
| 74 | rs7113596 | 11 | 112883761 | 0.00584533 |
| 74 | rs10891490 | 11 | 112885527 | 0.0124988 |
| 74 | rs10732853 | 11 | 112887410 | 0.00584533 |
| 74 | rs999851 | 11 | 112889846 | 0.00584533 |
| 74 | rs1940733 | 11 | 112892274 | 0.00584533 |
| 74 | rs9919620 | 11 | 112892570 | 0.00584533 |
| 74 | rs1940734 | 11 | 112895028 | 0.00584533 |
| 74 | rs10750021 | 11 | 112896214 | 0.00584533 |
| 74 | rs1940699 | 11 | 112898428 | 0.00584533 |
| 74 | rs1892983 | 11 | 112899214 | 0.00584533 |
| 74 | rs1892981 | 11 | 112899633 | 0.00584533 |
| 74 | rs11214469 | 11 | 112900343 | 0.00584533 |
| 74 | rs1320670 | 11 | 112902671 | 0.00584533 |
| 74 | rs7945073 | 11 | 112903595 | 0.00584533 |
| 74 | rs7113099 | 11 | 112904335 | 0.00790714 |
| 74 | rs7127528 | 11 | 112904479 | 0.00584533 |
| 74 | rs7128314 | 11 | 112905039 | 0.00584533 |
| 74 | rs4480572 | 11 | 112905776 | 0.00584533 |
| 74 | rs1940712 | 11 | 112906218 | 0.00584533 |
| 74 | rs1940713 | 11 | 112906285 | 0.00584533 |
| 74 | rs1940714 | 11 | 112906391 | 0.00584533 |
| 74 | rs7935745 | 11 | 112906804 | 0.00584533 |
| 74 | rs7947502 | 11 | 112909396 | 0.0145778 |
| 74 | rs10891492 | 11 | 112909745 | 0.0067964 |
| 74 | rs7948327 | 11 | 112910077 | 0.0067964 |
| 74 | rs2155292 | 11 | 112910783 | 0.00920515 |
| 74 | rs7938812 | 11 | 112911004 | 0.0067964 |
| 74 | rs7942723 | 11 | 112911839 | 0.00584533 |
| 74 | rs7105462 | 11 | 112912048 | 0.0124988 |
| 74 | rs3802847 | 11 | 112912303 | 0.0067964 |
| 74 | rs3802848 | 11 | 112912387 | 0.00790714 |
| 75 | rs12283343 | 11 | 123008349 | 0.019087 |
| 75 | rs12283311 | 11 | 123008467 | 0.0221832 |

| | | | | |
|---|---|---|---|---|
| 75 | rs4438047 | 11 | 123009683 | 0.202916 |
| 75 | rs4345978 | 11 | 123009751 | 0.238293 |
| 75 | rs4475932 | 11 | 123009975 | 0.0319236 |
| 75 | rs4936778 | 11 | 123010654 | 0.125186 |
| 75 | rs10892971 | 11 | 123010887 | 0.125186 |
| 75 | rs7127455 | 11 | 123013822 | 0.0366096 |
| 75 | rs7111575 | 11 | 123014015 | 0.0429212 |
| 75 | rs4435010 | 11 | 123014648 | 0.0366096 |
| 75 | rs7937909 | 11 | 123015109 | 0.0781364 |
| 76 | rs7967455 | 12 | 20293797 | 0.142489 |
| 76 | rs10841452 | 12 | 20295742 | 0.129079 |
| 76 | rs10841453 | 12 | 20295896 | 0.192004 |
| 76 | rs11045079 | 12 | 20297976 | 0.532408 |
| 77 | rs502034 | 12 | 24437928 | 0.233387 |
| 77 | rs563110 | 12 | 24438591 | 0.195535 |
| 77 | rs4579999 | 12 | 24439661 | 0.570694 |
| 78 | rs6487396 | 12 | 24717422 | 0.00440936 |
| 78 | rs1487657 | 12 | 24722254 | 0.00499384 |
| 78 | rs4641567 | 12 | 24722947 | 0.0038956 |
| 78 | rs923310 | 12 | 24725637 | 0.00499384 |
| 78 | rs970290 | 12 | 24730245 | 0.0038956 |
| 78 | rs34103124 | 12 | 24744134 | 0.00435753 |
| 78 | rs11047516 | 12 | 24749226 | 0.00344374 |
| 78 | rs11047517 | 12 | 24750548 | 0.00499384 |
| 78 | rs11047520 | 12 | 24752005 | 0.00826518 |
| 78 | rs7967802 | 12 | 24752416 | 0.00728049 |
| 78 | rs11047524 | 12 | 24755531 | 0.00826518 |
| 78 | rs111235435 | 12 | 24757629 | 0.238993 |
| 78 | rs113807524 | 12 | 24757748 | 0.0519543 |
| 78 | rs4963577 | 12 | 24758533 | 0.00994598 |
| 78 | rs4963773 | 12 | 24758708 | 0.00994598 |
| 78 | rs11047526 | 12 | 24759806 | 0.0396709 |
| 78 | rs1487663 | 12 | 24759838 | 0.0110262 |
| 78 | rs10743505 | 12 | 24760571 | 0.00994598 |
| 78 | rs7969321 | 12 | 24761558 | 0.0346965 |
| 78 | rs12369950 | 12 | 24762109 | 0.011392 |
| 78 | rs7137330 | 12 | 24762161 | 0.0346965 |
| 78 | rs12423810 | 12 | 24762452 | 0.0396709 |
| 78 | rs55646115 | 12 | 24763161 | 0.00897505 |
| 78 | rs4963774 | 12 | 24763441 | 0.00731672 |
| 78 | rs12368726 | 12 | 24763534 | 0.0346965 |
| 78 | rs10842380 | 12 | 24764510 | 0.0346965 |
| 78 | rs1487655 | 12 | 24764692 | 0.00994598 |
| 78 | rs10842381 | 12 | 24765496 | 0.00897505 |
| 78 | rs10743506 | 12 | 24768229 | 0.00731672 |
| 78 | rs1386639 | 12 | 24768847 | 0.0519543 |
| 78 | rs10771103 | 12 | 24772747 | 0.00792891 |
| 78 | rs10771104 | 12 | 24773130 | 0.00810202 |
| 78 | rs7953024 | 12 | 24773658 | 0.0110262 |
| 78 | rs7298837 | 12 | 24776182 | 0.00897505 |
| 78 | rs11047537 | 12 | 24777796 | 0.00897505 |
| 78 | rs1487650 | 12 | 24778640 | 0.00597397 |

| | | | | |
|---|---|---|---|---|
| 78 | rs4963579 | 12 | 24779857 | 0.00731672 |
| 78 | rs4963580 | 12 | 24779946 | 0.00597397 |
| 78 | rs11047538 | 12 | 24780539 | 0.00810202 |
| 78 | rs55998116 | 12 | 24781017 | 0.00540115 |
| 78 | rs11047540 | 12 | 24782071 | 0.00661007 |
| 78 | rs1872874 | 12 | 24782808 | 0.00661007 |
| 78 | rs12832485 | 12 | 24786136 | 0.00938862 |
| 78 | rs1906302 | 12 | 24786587 | 0.00661007 |
| 78 | rs7974582 | 12 | 24787712 | 0.00731672 |
| 78 | rs117394206 | 12 | 24793821 | 0.0095685 |
| 78 | rs7953698 | 12 | 24798175 | 0.00501575 |
| 78 | rs10505942 | 12 | 24798946 | 0.0220854 |
| 78 | rs7960243 | 12 | 24801854 | 0.00403285 |
| 78 | rs10842387 | 12 | 24802293 | 0.0132678 |
| 78 | rs10842388 | 12 | 24802324 | 0.0116973 |
| 78 | rs11047551 | 12 | 24809882 | 0.00333735 |
| 78 | rs6487404 | 12 | 24817388 | 0.0215252 |
| 78 | rs4963583 | 12 | 24817552 | 0.00604294 |
| 78 | rs7967537 | 12 | 24819762 | 0.00304611 |
| 78 | rs1487654 | 12 | 24820731 | 0.00342363 |
| 78 | rs7961989 | 12 | 24823507 | 0.0200008 |
| 79 | rs3782906 | 12 | 48170172 | 0.0211015 |
| 79 | rs7960277 | 12 | 48173570 | 0.0762354 |
| 79 | rs2525042 | 12 | 48175093 | 0.0315479 |
| 79 | rs4760652 | 12 | 48175604 | 0.154933 |
| 79 | rs4760654 | 12 | 48175837 | 0.134319 |
| 79 | rs7418 | 12 | 48176792 | 0.0713816 |
| 79 | rs9859 | 12 | 48176900 | 0.062229 |
| 79 | rs2408874 | 12 | 48177073 | 0.0315479 |
| 79 | rs13632 | 12 | 48177238 | 0.178796 |
| 79 | rs3192737 | 12 | 48178127 | 0.062229 |
| 79 | rs3815138 | 12 | 48178465 | 0.0413419 |
| 79 | rs3815136 | 12 | 48178909 | 0.0500984 |
| 79 | rs2240109 | 12 | 48180022 | 0.0473582 |
| 80 | rs2366150 | 12 | 54353523 | 0.0356579 |
| 80 | rs4237809 | 12 | 54353799 | 0.0477081 |
| 80 | rs4759059 | 12 | 54353880 | 0.0639824 |
| 80 | rs2002472 | 12 | 54355209 | 0.0639824 |
| 80 | rs2002471 | 12 | 54355261 | 0.0139116 |
| 80 | rs2366151 | 12 | 54355708 | 0.0552328 |
| 80 | rs12312094 | 12 | 54357126 | 0.0135166 |
| 80 | rs17720428 | 12 | 54357198 | 0.0159187 |
| 80 | rs1838169 | 12 | 54357495 | 0.0178182 |
| 80 | rs7958904 | 12 | 54357552 | 0.0552328 |
| 80 | rs4759313 | 12 | 54359074 | 0.0998154 |
| 80 | rs10783616 | 12 | 54359220 | 0.0998154 |
| 80 | rs10783617 | 12 | 54359387 | 0.0998154 |
| 80 | rs920778 | 12 | 54360232 | 0.0848043 |
| 80 | rs1899663 | 12 | 54360994 | 0.0208772 |
| 80 | rs17105613 | 12 | 54362194 | 0.035905 |
| 80 | rs4512901 | 12 | 54365236 | 0.0178182 |
| 80 | rs10783618 | 12 | 54365275 | 0.035905 |

| | | | | |
|---|---|---|---|---|
| 80 | rs4511324 | 12 | 54365535 | 0.0204753 |
| 80 | rs11170775 | 12 | 54365605 | 0.0635351 |
| 81 | rs74097857 | 12 | 66393756 | 0.303231 |
| 81 | rs11175997 | 12 | 66401444 | 0.0718288 |
| 81 | rs1383304 | 12 | 66405304 | 0.0912501 |
| 81 | rs975736 | 12 | 66405372 | 0.0565572 |
| 81 | rs975917 | 12 | 66406903 | 0.0912501 |
| 81 | rs11176001 | 12 | 66409367 | 0.385883 |
| 82 | rs1920568 | 12 | 114669732 | 0.00744341 |
| 82 | rs2555015 | 12 | 114673774 | 0.149963 |
| 82 | rs1270886 | 12 | 114676470 | 0.0114399 |
| 82 | rs1265496 | 12 | 114676983 | 0.00990683 |
| 82 | rs2555013 | 12 | 114678318 | 0.149963 |
| 82 | rs2555012 | 12 | 114678725 | 0.0114399 |
| 82 | rs2252414 | 12 | 114679137 | 0.0132185 |
| 82 | rs1270885 | 12 | 114681552 | 0.0675182 |
| 82 | rs1247940 | 12 | 114682651 | 0.0274817 |
| 82 | rs1247938 | 12 | 114683568 | 0.0132185 |
| 82 | rs1269789 | 12 | 114684542 | 0.0114399 |
| 82 | rs2253207 | 12 | 114685437 | 0.0237098 |
| 82 | rs1270884 | 12 | 114685571 | 0.175849 |
| 82 | rs2555004 | 12 | 114686645 | 0.242273 |
| 82 | rs1247928 | 12 | 114686840 | 0.0274817 |
| 82 | rs1247927 | 12 | 114687056 | 0.00990683 |
| 83 | rs680705 | 13 | 51443315 | 0.37887 |
| 83 | rs7994724 | 13 | 51445560 | 0.620925 |
| 84 | rs11635984 | 15 | 33012232 | 0.619177 |
| 84 | rs7497354 | 15 | 33015402 | 0.358718 |
| 85 | rs72743461 | 15 | 67441750 | 0.229247 |
| 85 | rs17293632 | 15 | 67442596 | 0.296126 |
| 85 | rs56375023 | 15 | 67448363 | 0.229247 |
| 85 | rs17228058 | 15 | 67450305 | 0.106663 |
| 85 | rs56062135 | 15 | 67455630 | 0.137585 |
| 86 | rs7179413 | 15 | 68324839 | 0.00279599 |
| 86 | rs34399936 | 15 | 68331720 | 0.00484288 |
| 86 | rs11635759 | 15 | 68340393 | 0.00213245 |
| 86 | rs11633928 | 15 | 68351510 | 0.00320458 |
| 86 | rs11636657 | 15 | 68353444 | 0.00556447 |
| 86 | rs12905500 | 15 | 68353576 | 0.00244102 |
| 86 | rs12914713 | 15 | 68367515 | 0.00367517 |
| 86 | rs12915951 | 15 | 68367617 | 0.00639756 |
| 86 | rs72759519 | 15 | 68368817 | 0.00639756 |
| 86 | rs11633140 | 15 | 68372106 | 0.00639756 |
| 86 | rs7165099 | 15 | 68373638 | 0.00639756 |
| 86 | rs6494711 | 15 | 68374027 | 0.00279599 |
| 86 | rs11638566 | 15 | 68381608 | 0.0112483 |
| 86 | rs8029670 | 15 | 68383415 | 0.00367517 |
| 86 | rs11071978 | 15 | 68389773 | 0.00267491 |
| 86 | rs4306484 | 15 | 68396372 | 0.0112483 |
| 86 | rs8031079 | 15 | 68401582 | 0.00639756 |
| 86 | rs11856619 | 15 | 68404362 | 0.00484288 |
| 86 | rs7162430 | 15 | 68407287 | 0.00735997 |

| | | | | |
|---|---|---|---|---|
| 86 | rs62004785 | 15 | 68409265 | 0.0129728 |
| 86 | rs12912799 | 15 | 68409551 | 0.0129728 |
| 86 | rs28656984 | 15 | 68414282 | 0.00556447 |
| 86 | rs2029423 | 15 | 68420906 | 0.0129728 |
| 86 | rs2029421 | 15 | 68421623 | 0.0112483 |
| 86 | rs12148323 | 15 | 68424634 | 0.014971 |
| 86 | rs8038202 | 15 | 68425122 | 0.00639756 |
| 86 | rs8038236 | 15 | 68425265 | 0.00367517 |
| 86 | rs8038120 | 15 | 68425297 | 0.00735997 |
| 86 | rs12898743 | 15 | 68431168 | 0.00847244 |
| 86 | rs7165825 | 15 | 68431859 | 0.00735997 |
| 86 | rs7167918 | 15 | 68432140 | 0.00639756 |
| 86 | rs7402649 | 15 | 68436575 | 0.00847244 |
| 86 | rs8035890 | 15 | 68437102 | 0.0172877 |
| 86 | rs4777022 | 15 | 68440028 | 0.014971 |
| 86 | rs8030302 | 15 | 68441485 | 0.0199755 |
| 86 | rs34760852 | 15 | 68441777 | 0.0172877 |
| 86 | rs12905763 | 15 | 68449751 | 0.0172877 |
| 86 | rs35722665 | 15 | 68449779 | 0.0267195 |
| 86 | rs35595740 | 15 | 68452515 | 0.014971 |
| 86 | rs4777023 | 15 | 68453952 | 0.0172877 |
| 86 | rs62004794 | 15 | 68454523 | 0.00975916 |
| 86 | rs1878360 | 15 | 68455547 | 0.00735997 |
| 86 | rs2414990 | 15 | 68460290 | 0.0172877 |
| 86 | rs7181874 | 15 | 68461019 | 0.00320458 |
| 86 | rs2037212 | 15 | 68467760 | 0.0754269 |
| 86 | rs12438361 | 15 | 68468695 | 0.0558985 |
| 86 | rs12594232 | 15 | 68469402 | 0.0876992 |
| 86 | rs35125010 | 15 | 68471048 | 0.0754269 |
| 86 | rs2176333 | 15 | 68479259 | 0.00484288 |
| 86 | rs1049493 | 15 | 68480887 | 0.0558985 |
| 86 | rs12912276 | 15 | 68481582 | 0.0199755 |
| 86 | rs3743089 | 15 | 68490495 | 0.014971 |
| 86 | rs11855587 | 15 | 68490957 | 0.00556447 |
| 86 | rs11071986 | 15 | 68492028 | 0.0129728 |
| 86 | rs11635988 | 15 | 68492262 | 0.00484288 |
| 86 | rs938640 | 15 | 68493748 | 0.0112483 |
| 86 | rs11630382 | 15 | 68495584 | 0.00484288 |
| 86 | rs11071987 | 15 | 68495754 | 0.00639756 |
| 86 | rs11071988 | 15 | 68495852 | 0.0112483 |
| 86 | rs11071991 | 15 | 68498052 | 0.00279599 |
| 86 | rs8028241 | 15 | 68499321 | 0.00320458 |
| 86 | rs8142 | 15 | 68499661 | 0.00735997 |
| 86 | rs11632516 | 15 | 68501710 | 0.00847244 |
| 86 | rs1400432 | 15 | 68502604 | 0.00320458 |
| 86 | rs10468043 | 15 | 68503216 | 0.00975916 |
| 86 | rs10048021 | 15 | 68504806 | 0.00367517 |
| 86 | rs35873920 | 15 | 68505086 | 0.00367517 |
| 86 | rs10467939 | 15 | 68505234 | 0.00735997 |
| 86 | rs8031661 | 15 | 68505624 | 0.00320458 |
| 86 | rs10431808 | 15 | 68507272 | 0.00320458 |
| 86 | rs35627684 | 15 | 68511645 | 0.00320458 |

| | | | | |
|---|---|---|---|---|
| 86 | rs35082625 | 15 | 68513382 | 0.00367517 |
| 86 | rs55953911 | 15 | 68514011 | 0.00556447 |
| 86 | rs11638034 | 15 | 68516294 | 0.00279599 |
| 86 | rs11639222 | 15 | 68516780 | 0.00556447 |
| 86 | rs8040883 | 15 | 68517211 | 0.00244102 |
| 86 | rs8037724 | 15 | 68519855 | 0.0042175 |
| 86 | rs8037734 | 15 | 68519871 | 0.0042175 |
| 86 | rs867483 | 15 | 68525499 | 0.00213245 |
| 86 | rs7162452 | 15 | 68529760 | 0.00244102 |
| 86 | rs7163274 | 15 | 68530158 | 0.00244102 |
| 86 | rs12439517 | 15 | 68530568 | 0.00244102 |
| 86 | rs12439519 | 15 | 68530575 | 0.00244102 |
| 86 | rs11071996 | 15 | 68531108 | 0.00244102 |
| 87 | rs7183500 | 15 | 96100923 | 0.118109 |
| 87 | rs7183672 | 15 | 96101018 | 0.478735 |
| 87 | rs6496121 | 15 | 96101497 | 0.401105 |
| 88 | rs79379911 | 16 | 15863363 | 0.0256337 |
| 88 | rs58476672 | 16 | 15875922 | 0.0154471 |
| 88 | rs6498573 | 16 | 15879373 | 0.63247 |
| 88 | rs80175860 | 16 | 15881551 | 0.0798469 |
| 88 | rs72772064 | 16 | 15882857 | 0.206587 |
| 89 | rs16941554 | 16 | 86622436 | 0.049642 |
| 89 | rs4843403 | 16 | 86624652 | 0.0155635 |
| 89 | rs4843404 | 16 | 86624863 | 0.0670667 |
| 89 | rs4843405 | 16 | 86625055 | 0.0670667 |
| 89 | rs4843174 | 16 | 86625202 | 0.0514549 |
| 89 | rs11117197 | 16 | 86625694 | 0.00986286 |
| 89 | rs35900603 | 16 | 86625702 | 0.0084808 |
| 89 | rs16941567 | 16 | 86625741 | 0.0084808 |
| 89 | rs62051074 | 16 | 86625764 | 0.036817 |
| 89 | rs1424018 | 16 | 86627442 | 0.00729634 |
| 89 | rs34968499 | 16 | 86627604 | 0.00729634 |
| 89 | rs8048997 | 16 | 86627724 | 0.0084808 |
| 89 | rs12711458 | 16 | 86628058 | 0.0223706 |
| 89 | rs8043893 | 16 | 86628423 | 0.193868 |
| 89 | rs8045203 | 16 | 86628473 | 0.0223706 |
| 89 | rs4843406 | 16 | 86628948 | 0.0114763 |
| 89 | rs4843407 | 16 | 86629047 | 0.105706 |
| 89 | rs4843175 | 16 | 86629126 | 0.0084808 |
| 89 | rs4843176 | 16 | 86629141 | 0.00729634 |
| 89 | rs28483418 | 16 | 86630004 | 0.00986286 |
| 89 | rs35182705 | 16 | 86630010 | 0.0114763 |
| 89 | rs35670768 | 16 | 86630042 | 0.0114763 |
| 89 | rs28714465 | 16 | 86630070 | 0.0114763 |
| 89 | rs1364228 | 16 | 86631620 | 0.0336492 |
| 89 | rs1345873 | 16 | 86631765 | 0.0288093 |
| 89 | rs1364227 | 16 | 86631780 | 0.0142701 |
| 89 | rs28558083 | 16 | 86631959 | 0.0336492 |
| 89 | rs889599 | 16 | 86632136 | 0.0288093 |
| 89 | rs889598 | 16 | 86632293 | 0.0246788 |
| 89 | rs34599077 | 16 | 86634678 | 0.00914514 |
| 89 | rs12325438 | 16 | 86634807 | 0.00789272 |

| | | | | |
|---|---|---|---|---|
| 89 | rs9926431 | 16 | 86634866 | 0.00729634 |
| 89 | rs9936136 | 16 | 86635303 | 0.00588804 |
| 89 | rs16941621 | 16 | 86635543 | 0.00729634 |
| 90 | rs78378222 | 17 | 7571752 | 0.999952 |
| 91 | rs16946141 | 17 | 12396800 | 0.042794 |
| 91 | rs72809115 | 17 | 12397996 | 0.0271401 |
| 91 | rs720866 | 17 | 12400419 | 0.0173221 |
| 91 | rs16946148 | 17 | 12401109 | 0.152436 |
| 91 | rs62061554 | 17 | 12401309 | 0.348007 |
| 91 | rs876522 | 17 | 12401811 | 0.026645 |
| 91 | rs72809125 | 17 | 12404433 | 0.314349 |
| 91 | rs62061556 | 17 | 12406483 | 0.0219745 |
| 92 | rs8067439 | 17 | 17698254 | 0.00114294 |
| 92 | rs12946913 | 17 | 17706332 | 0.00475688 |
| 92 | rs11654081 | 17 | 17708129 | 0.00259876 |
| 92 | rs2350976 | 17 | 17708529 | 0.00101881 |
| 92 | rs12453837 | 17 | 17708846 | 0.0023069 |
| 92 | rs11652881 | 17 | 17709136 | 0.00128296 |
| 92 | rs4925114 | 17 | 17711270 | 0.00372843 |
| 92 | rs2297508 | 17 | 17715317 | 0.00537784 |
| 92 | rs4925115 | 17 | 17721457 | 0.0146615 |
| 92 | rs11656665 | 17 | 17724789 | 0.00421012 |
| 92 | rs9899634 | 17 | 17727943 | 0.0033038 |
| 92 | rs8066560 | 17 | 17728043 | 0.00608347 |
| 92 | rs11657423 | 17 | 17728574 | 0.00204903 |
| 92 | rs11078399 | 17 | 17728983 | 0.0023069 |
| 92 | rs4925116 | 17 | 17731200 | 0.0023069 |
| 92 | rs12952723 | 17 | 17731651 | 0.0023069 |
| 92 | rs9894257 | 17 | 17732319 | 0.00204903 |
| 92 | rs9902941 | 17 | 17733760 | 0.00161944 |
| 92 | rs11657130 | 17 | 17733826 | 0.00101881 |
| 92 | rs11652144 | 17 | 17733934 | 0.00128296 |
| 92 | rs1889018 | 17 | 17734740 | 0.00101881 |
| 92 | rs4925117 | 17 | 17736252 | 0.0029545 |
| 92 | rs12938501 | 17 | 17737258 | 0.00161944 |
| 92 | rs4925119 | 17 | 17742904 | 0.00259876 |
| 92 | rs9891957 | 17 | 17744439 | 0.00114294 |
| 92 | rs8078756 | 17 | 17744725 | 0.00144098 |
| 92 | rs9906673 | 17 | 17746197 | 0.00128296 |
| 92 | rs6502618 | 17 | 17746741 | 0.00161944 |
| 92 | rs3183702 | 17 | 17747289 | 0.00292928 |
| 92 | rs2236513 | 17 | 17747366 | 0.00292928 |
| 92 | rs9915248 | 17 | 17747514 | 0.00182107 |
| 92 | rs4924822 | 17 | 17748013 | 0.00292928 |
| 92 | rs3744113 | 17 | 17748645 | 0.00161944 |
| 92 | rs9915776 | 17 | 17749056 | 0.00259876 |
| 92 | rs3744115 | 17 | 17749822 | 0.00182107 |
| 92 | rs1052299 | 17 | 17750419 | 0.00204903 |
| 92 | rs1108648 | 17 | 17750558 | 0.00161944 |
| 92 | rs7501812 | 17 | 17750907 | 0.00128296 |
| 92 | rs1108646 | 17 | 17751478 | 0.00131886 |
| 92 | rs9893690 | 17 | 17753846 | 0.00475688 |

| | | | | |
|---|---|---|---|---|
| 92 | rs4925120 | 17 | 17754633 | 0.00421012 |
| 92 | rs16960744 | 17 | 17755259 | 0.00475688 |
| 92 | rs11078400 | 17 | 17757728 | 0.00372843 |
| 92 | rs8079321 | 17 | 17760789 | 0.0033038 |
| 92 | rs11658309 | 17 | 17762247 | 0.00259876 |
| 92 | rs9911281 | 17 | 17762457 | 0.0023069 |
| 92 | rs35451946 | 17 | 17764061 | 0.00259876 |
| 92 | rs4924823 | 17 | 17764502 | 0.00259876 |
| 92 | rs12941039 | 17 | 17765655 | 0.00182107 |
| 92 | rs11657074 | 17 | 17770355 | 0.00128296 |
| 92 | rs9907246 | 17 | 17770965 | 0.00182107 |
| 92 | rs9907287 | 17 | 17774118 | 0.0023069 |
| 92 | rs9908017 | 17 | 17774422 | 0.0023069 |
| 92 | rs9908299 | 17 | 17774568 | 0.00114294 |
| 92 | rs8080061 | 17 | 17776389 | 0.00204903 |
| 92 | rs12936037 | 17 | 17777245 | 0.00114294 |
| 92 | rs11871231 | 17 | 17883848 | 0.00981275 |
| 92 | rs11869491 | 17 | 17884055 | 0.00981275 |
| 92 | rs4072738 | 17 | 17884547 | 0.00680483 |
| 92 | rs4072739 | 17 | 17884660 | 0.00768356 |
| 92 | rs4459604 | 17 | 17888549 | 0.00105276 |
| 92 | rs7207821 | 17 | 17891781 | 0.00131886 |
| 92 | rs62072048 | 17 | 17894750 | 0.00147742 |
| 92 | rs4368210 | 17 | 17896090 | 0.00165597 |
| 92 | rs4584886 | 17 | 17896205 | 0.00165597 |
| 92 | rs9911850 | 17 | 17896559 | 0.00117799 |
| 92 | rs9912096 | 17 | 17896673 | 0.00131886 |
| 92 | rs4924832 | 17 | 17897739 | 0.00165597 |
| 92 | rs9899355 | 17 | 17898243 | 0.0075756 |
| 92 | rs6502631 | 17 | 17899621 | 0.00131886 |
| 92 | rs4506969 | 17 | 17899839 | 0.00165597 |
| 92 | rs8071238 | 17 | 17900990 | 0.00185715 |
| 92 | rs8075189 | 17 | 17901946 | 0.00131886 |
| 92 | rs28537385 | 17 | 17902135 | 0.00165597 |
| 92 | rs62072049 | 17 | 17906520 | 0.0023069 |
| 92 | rs62072050 | 17 | 17906564 | 0.00165597 |
| 92 | rs7209003 | 17 | 17907393 | 0.00105276 |
| 92 | rs7219324 | 17 | 17909775 | 0.00672858 |
| 92 | rs12150369 | 17 | 17911014 | 0.00117799 |
| 92 | rs9897761 | 17 | 17912011 | 0.00131886 |
| 92 | rs4365348 | 17 | 17912548 | 0.00131886 |
| 92 | rs6502632 | 17 | 17913057 | 0.00117799 |
| 92 | rs7212167 | 17 | 17913504 | 0.00117799 |
| 92 | rs7223696 | 17 | 17915791 | 0.00117799 |
| 92 | rs9913277 | 17 | 17919749 | 0.00105276 |
| 92 | rs8069811 | 17 | 17922280 | 0.00117799 |
| 92 | rs9896837 | 17 | 17924868 | 0.00147742 |
| 92 | rs7215524 | 17 | 17926605 | 0.00147742 |
| 92 | rs2955378 | 17 | 17930253 | 0.00131886 |
| 92 | rs2955377 | 17 | 17932818 | 0.00131886 |
| 92 | rs7224047 | 17 | 17932931 | 0.00131886 |
| 92 | rs2955385 | 17 | 17939247 | 0.00147742 |

| | | | | |
|---|---|---|---|---|
| 92 | rs2955384 | 17 | 17939573 | 0.00147742 |
| 92 | rs7503738 | 17 | 17940305 | 0.00147742 |
| 92 | rs6502633 | 17 | 17941037 | 0.00131886 |
| 92 | rs2955383 | 17 | 17941364 | 0.00189224 |
| 92 | rs7406982 | 17 | 17942613 | 0.00147742 |
| 92 | rs2955380 | 17 | 17944349 | 0.00147742 |
| 92 | rs2955381 | 17 | 17945500 | 0.00147742 |
| 92 | rs2955359 | 17 | 17946730 | 0.00117799 |
| 92 | rs2955354 | 17 | 17948716 | 0.00117799 |
| 92 | rs2955353 | 17 | 17948979 | 0.00117799 |
| 92 | rs7207461 | 17 | 17949789 | 0.00117799 |
| 92 | rs12948749 | 17 | 17949802 | 0.00161944 |
| 92 | rs7210400 | 17 | 17949957 | 0.000941381 |
| 92 | rs12940282 | 17 | 17950001 | 0.00117799 |
| 92 | rs11652894 | 17 | 17952439 | 0.00144098 |
| 92 | rs8070731 | 17 | 17952447 | 0.00105276 |
| 92 | rs4925135 | 17 | 17952868 | 0.00117799 |
| 92 | rs4925136 | 17 | 17953002 | 0.00117799 |
| 92 | rs2955350 | 17 | 17953548 | 0.00117799 |
| 92 | rs8080602 | 17 | 17954728 | 0.00117799 |
| 92 | rs6502634 | 17 | 17955344 | 0.00117799 |
| 92 | rs11650021 | 17 | 17960613 | 0.000941381 |
| 92 | rs2955368 | 17 | 17961349 | 0.00262849 |
| 92 | rs9894138 | 17 | 17964345 | 0.00131886 |
| 92 | rs2955356 | 17 | 17964717 | 0.000941381 |
| 92 | rs2955370 | 17 | 17966945 | 0.00117799 |
| 92 | rs2955371 | 17 | 17967397 | 0.00144098 |
| 92 | rs2955372 | 17 | 17970229 | 0.00131886 |
| 92 | rs4643387 | 17 | 17972973 | 0.00147742 |
| 92 | rs2955373 | 17 | 17973617 | 0.0037392 |
| 92 | rs8066764 | 17 | 17974668 | 0.0096185 |
| 92 | rs62073573 | 17 | 17976354 | 0.00981275 |
| 92 | rs9303144 | 17 | 17977355 | 0.0075756 |
| 92 | rs4426402 | 17 | 17977926 | 0.0125604 |
| 92 | rs4410135 | 17 | 17979039 | 0.0110987 |
| 92 | rs4561528 | 17 | 17979099 | 0.0207193 |
| 92 | rs7406744 | 17 | 17981938 | 0.00981275 |
| 92 | rs6502636 | 17 | 17983817 | 0.0142225 |
| 92 | rs59518074 | 17 | 17984162 | 0.00680483 |
| 92 | rs6502637 | 17 | 17986098 | 0.0125604 |
| 92 | rs7211714 | 17 | 17986955 | 0.00680483 |
| 92 | rs7212447 | 17 | 17987067 | 0.0110987 |
| 92 | rs4365346 | 17 | 17989373 | 0.00981275 |
| 92 | rs4349203 | 17 | 17989425 | 0.00768356 |
| 92 | rs4506967 | 17 | 17990474 | 0.0138166 |
| 92 | rs11078410 | 17 | 17990634 | 0.0110987 |
| 92 | rs9890563 | 17 | 17990671 | 0.0371687 |
| 92 | rs2974998 | 17 | 17995619 | 0.00768356 |
| 92 | rs2974999 | 17 | 17997547 | 0.00868068 |
| 92 | rs854814 | 17 | 18003648 | 0.0125604 |
| 92 | rs721669 | 17 | 18005073 | 0.00534644 |
| 92 | rs854809 | 17 | 18006539 | 0.0110987 |

| | | | | |
|---|---|---|---|---|
| 92 | rs854808 | 17 | 18006634 | 0.0110987 |
| 92 | rs2056842 | 17 | 18009028 | 0.0142225 |
| 92 | rs854763 | 17 | 18010095 | 0.00603001 |
| 92 | rs6826 | 17 | 18011140 | 0.0125604 |
| 92 | rs1101727 | 17 | 18016148 | 0.0110987 |
| 92 | rs854767 | 17 | 18018263 | 0.0110987 |
| 92 | rs854768 | 17 | 18018806 | 0.00981275 |
| 92 | rs854769 | 17 | 18019011 | 0.0125604 |
| 92 | rs854770 | 17 | 18019048 | 0.00981275 |
| 92 | rs741782 | 17 | 18019712 | 0.0110987 |
| 92 | rs712267 | 17 | 18021607 | 0.00981275 |
| 92 | rs854817 | 17 | 18021882 | 0.00981275 |
| 92 | rs854818 | 17 | 18022039 | 0.00981275 |
| 92 | rs2955365 | 17 | 18023897 | 0.0110987 |
| 92 | rs2955366 | 17 | 18024013 | 0.0110987 |
| 92 | rs712268 | 17 | 18025744 | 0.00534644 |
| 92 | rs854789 | 17 | 18026566 | 0.00680483 |
| 92 | rs1008136 | 17 | 18027142 | 0.00603001 |
| 92 | rs1008135 | 17 | 18027143 | 0.00603001 |
| 92 | rs854791 | 17 | 18029857 | 0.00421012 |
| 92 | rs854792 | 17 | 18030240 | 0.0170578 |
| 92 | rs854794 | 17 | 18030852 | 0.00142246 |
| 92 | rs854786 | 17 | 18036283 | 0.199791 |
| 92 | rs2746025 | 17 | 18151611 | 0.00534644 |
| 93 | rs2040792 | 17 | 29628549 | 0.00983965 |
| 93 | rs2214538 | 17 | 29636570 | 0.00983965 |
| 93 | rs12602834 | 17 | 29637308 | 0.0085752 |
| 93 | rs4368212 | 17 | 29639263 | 0.00380409 |
| 93 | rs4638642 | 17 | 29639325 | 0.00652451 |
| 93 | rs10512434 | 17 | 29639590 | 0.00569622 |
| 93 | rs7218930 | 17 | 29640204 | 0.00569622 |
| 93 | rs17826544 | 17 | 29640225 | 0.00569622 |
| 93 | rs17882020 | 17 | 29640679 | 0.00652451 |
| 93 | rs7503922 | 17 | 29641935 | 0.00569622 |
| 93 | rs7502556 | 17 | 29642430 | 0.00497603 |
| 93 | rs12940802 | 17 | 29643564 | 0.00747769 |
| 93 | rs7505 | 17 | 29644852 | 0.0085752 |
| 93 | rs1129506 | 17 | 29646032 | 0.00291508 |
| 93 | rs2854305 | 17 | 29647003 | 0.00652451 |
| 93 | rs3785956 | 17 | 29647707 | 0.00569622 |
| 93 | rs3785955 | 17 | 29647802 | 0.00747769 |
| 93 | rs2525563 | 17 | 29647908 | 0.00569622 |
| 93 | rs2854306 | 17 | 29648146 | 0.00747769 |
| 93 | rs35109242 | 17 | 29649223 | 0.00747769 |
| 93 | rs4794887 | 17 | 29649696 | 0.00747769 |
| 93 | rs35888506 | 17 | 29651229 | 0.00569622 |
| 93 | rs2854307 | 17 | 29652147 | 0.00747769 |
| 93 | rs2525564 | 17 | 29652531 | 0.00569622 |
| 93 | rs9894648 | 17 | 29653293 | 0.00255409 |
| 93 | rs17880873 | 17 | 29654096 | 0.00983965 |
| 93 | rs2285894 | 17 | 29654876 | 0.0085752 |
| 93 | rs3815154 | 17 | 29654974 | 0.00652451 |

| | | | | |
|---|---|---|---|---|
| 93 | rs17885030 | 17 | 29655218 | 0.00747769 |
| 93 | rs10445402 | 17 | 29661340 | 0.00652451 |
| 93 | rs2854308 | 17 | 29666539 | 0.00380409 |
| 93 | rs2525565 | 17 | 29666588 | 0.00332906 |
| 93 | rs2189525 | 17 | 29668808 | 0.00497603 |
| 93 | rs2854311 | 17 | 29671327 | 0.00497603 |
| 93 | rs2525568 | 17 | 29675878 | 0.0112973 |
| 93 | rs2525569 | 17 | 29675971 | 0.0085752 |
| 93 | rs964288 | 17 | 29679246 | 0.0295708 |
| 93 | rs12943365 | 17 | 29680526 | 0.026203 |
| 93 | rs2525570 | 17 | 29681245 | 0.331546 |
| 93 | rs7350943 | 17 | 29684553 | 0.0129785 |
| 93 | rs7502834 | 17 | 29689430 | 0.0085752 |
| 93 | rs7406783 | 17 | 29689980 | 0.0295708 |
| 93 | rs12947265 | 17 | 29690992 | 0.0295708 |
| 93 | rs10438801 | 17 | 29693785 | 0.0342322 |
| 93 | rs2854319 | 17 | 29694054 | 0.00497603 |
| 93 | rs2854320 | 17 | 29694063 | 0.0149188 |
| 93 | rs2107359 | 17 | 29697223 | 0.0221071 |
| 93 | rs1048317 | 17 | 29704002 | 0.00291508 |
| 93 | rs2525574 | 17 | 29705947 | 0.0255601 |
| 93 | rs8067440 | 17 | 29708155 | 0.00569622 |
| 93 | rs2854330 | 17 | 29710763 | 0.0171593 |
| 93 | rs2854333 | 17 | 29713300 | 0.0149188 |
| 93 | rs2854334 | 17 | 29715500 | 0.0149188 |
| 93 | rs2854303 | 17 | 29716342 | 0.0085752 |
| 93 | rs757375 | 17 | 29717879 | 0.00434949 |
| 93 | rs757376 | 17 | 29717909 | 0.0112973 |
| 93 | rs757377 | 17 | 29720426 | 0.0085752 |
| 93 | rs736598 | 17 | 29720470 | 0.00569622 |
| 93 | rs2342052 | 17 | 29721011 | 0.00497603 |
| 93 | rs2342053 | 17 | 29722196 | 0.00983965 |
| 93 | rs757378 | 17 | 29722619 | 0.00983965 |
| 93 | rs734403 | 17 | 29723346 | 0.00434949 |
| 93 | rs757379 | 17 | 29724557 | 0.0085752 |
| 93 | rs2342054 | 17 | 29724708 | 0.0129785 |
| 94 | rs10048158 | 17 | 64236318 | 0.00160365 |
| 94 | rs9895261 | 17 | 64240436 | 0.00308891 |
| 94 | rs12603947 | 17 | 64244505 | 0.00217017 |
| 94 | rs7342920 | 17 | 64244645 | 0.00217017 |
| 94 | rs56003105 | 17 | 64245535 | 0.00308891 |
| 94 | rs12601387 | 17 | 64279084 | 0.00129823 |
| 94 | rs56152251 | 17 | 64280153 | 0.0953635 |
| 94 | rs62069915 | 17 | 64280347 | 0.00160365 |
| 94 | rs16959037 | 17 | 64280416 | 0.00143374 |
| 94 | rs62069916 | 17 | 64280503 | 0.00442135 |
| 94 | rs4439797 | 17 | 64280506 | 0.109231 |
| 94 | rs4625755 | 17 | 64280518 | 0.109231 |
| 94 | rs8069670 | 17 | 64280719 | 0.125194 |
| 94 | rs4510065 | 17 | 64283183 | 0.00329565 |
| 94 | rs4641779 | 17 | 64283326 | 0.00181329 |
| 94 | rs4993336 | 17 | 64283819 | 0.00302304 |

| | | | | |
|---|---|---|---|---|
| 94 | rs4993335 | 17 | 64283843 | 0.0728227 |
| 94 | rs4993334 | 17 | 64283851 | 0.00329565 |
| 94 | rs4423457 | 17 | 64284000 | 0.164765 |
| 94 | rs76182952 | 17 | 64291695 | 0.00153329 |
| 94 | rs74514664 | 17 | 64291772 | 0.00142046 |
| 94 | rs11867898 | 17 | 64295321 | 0.00153329 |
| 94 | rs79277327 | 17 | 64300107 | 0.00153329 |
| 94 | rs67700546 | 17 | 64301081 | 0.0118254 |
| 94 | rs17633401 | 17 | 64301363 | 0.0118254 |
| 94 | rs17633437 | 17 | 64301842 | 0.0118254 |
| 94 | rs12601850 | 17 | 64302652 | 0.0118254 |
| 94 | rs12944131 | 17 | 64307611 | 0.0118254 |
| 94 | rs112520848 | 17 | 64308310 | 0.00921244 |
| 94 | rs17706845 | 17 | 64310185 | 0.0134105 |
| 94 | rs35512343 | 17 | 64310575 | 0.0118254 |
| 94 | rs76219881 | 17 | 64311216 | 0.00208742 |
| 94 | rs77703440 | 17 | 64311917 | 0.00208742 |
| 94 | rs76932180 | 17 | 64313033 | 0.00225636 |
| 94 | rs74521786 | 17 | 64315288 | 0.00225636 |
| 94 | rs35105653 | 17 | 64321521 | 0.0081388 |
| 94 | rs35687884 | 17 | 64321828 | 0.00921244 |
| 94 | rs35433169 | 17 | 64322135 | 0.00921244 |
| 94 | rs4433842 | 17 | 64322683 | 0.00921244 |
| 94 | rs12945138 | 17 | 64323610 | 0.00921244 |
| 94 | rs7210846 | 17 | 64324352 | 0.0081388 |
| 94 | rs67828405 | 17 | 64324384 | 0.0104342 |
| 94 | rs67675584 | 17 | 64324501 | 0.00921244 |
| 94 | rs79345103 | 17 | 64324732 | 0.0021413 |
| 94 | rs12938072 | 17 | 64324761 | 0.00498901 |
| 94 | rs12938407 | 17 | 64324884 | 0.00921244 |
| 94 | rs35135345 | 17 | 64325539 | 0.00921244 |
| 94 | rs4239073 | 17 | 64326292 | 0.00921244 |
| 94 | rs8074759 | 17 | 64326305 | 0.0021413 |
| 94 | rs75154965 | 17 | 64328707 | 0.00231185 |
| 94 | rs9635753 | 17 | 64329197 | 0.0081388 |
| 94 | rs78835200 | 17 | 64330278 | 0.00198387 |
| 94 | rs8081834 | 17 | 64331037 | 0.00571733 |
| 94 | rs72843895 | 17 | 64331156 | 0.00571733 |
| 95 | rs2421206 | 19 | 11262477 | 0.608719 |
| 95 | rs113783450 | 19 | 11271714 | 0.0976473 |
| 95 | rs111427795 | 19 | 11272727 | 0.0333685 |
| 95 | rs72983206 | 19 | 11274065 | 0.0627936 |
| 95 | rs7257816 | 19 | 11274445 | 0.0396917 |
| 95 | rs7188 | 19 | 11275139 | 0.0396917 |
| 95 | rs7187 | 19 | 11275258 | 0.0670286 |
| 95 | rs3185010 | 19 | 11275842 | 0.0162381 |
| 96 | rs4803662 | 19 | 44359564 | 0.0302256 |
| 96 | rs8104290 | 19 | 44367338 | 0.01977 |
| 96 | rs2191564 | 19 | 44367611 | 0.01977 |
| 96 | rs10416434 | 19 | 44367843 | 0.01977 |
| 96 | rs10417741 | 19 | 44367911 | 0.0227603 |
| 96 | rs8106090 | 19 | 44371210 | 0.0302256 |

| | | | | |
|---|---|---|---|---|
| 96 | rs12985878 | 19 | 44371393 | 0.0262201 |
| 96 | rs11673020 | 19 | 44374631 | 0.0227603 |
| 96 | rs1050054 | 19 | 44376681 | 0.01977 |
| 96 | rs12977303 | 19 | 44377669 | 0.0171838 |
| 96 | rs10406290 | 19 | 44378877 | 0.0171838 |
| 96 | rs10414702 | 19 | 44380536 | 0.01977 |
| 96 | rs10421518 | 19 | 44381286 | 0.01977 |
| 96 | rs7258418 | 19 | 44383083 | 0.0171838 |
| 96 | rs1978723 | 19 | 44383800 | 0.01977 |
| 96 | rs371875 | 19 | 44393257 | 0.0130078 |
| 96 | rs368079 | 19 | 44393356 | 0.0149458 |
| 96 | rs425217 | 19 | 44393975 | 0.00987249 |
| 96 | rs429027 | 19 | 44395489 | 0.0113285 |
| 96 | rs396874 | 19 | 44397225 | 0.0149458 |
| 96 | rs432454 | 19 | 44398831 | 0.00987249 |
| 96 | rs397913 | 19 | 44398940 | 0.0149458 |
| 96 | rs368089 | 19 | 44399084 | 0.0149458 |
| 96 | rs397346 | 19 | 44399115 | 0.0130078 |
| 96 | rs396973 | 19 | 44399306 | 0.0130078 |
| 96 | rs436249 | 19 | 44399565 | 0.0149458 |
| 96 | rs430308 | 19 | 44400375 | 0.0149458 |
| 96 | rs372491 | 19 | 44401793 | 0.0130078 |
| 96 | rs415168 | 19 | 44402574 | 0.01977 |
| 96 | rs453950 | 19 | 44403055 | 0.0130078 |
| 96 | rs440784 | 19 | 44404342 | 0.0130078 |
| 96 | rs374307 | 19 | 44404398 | 0.0149458 |
| 96 | rs441344 | 19 | 44404682 | 0.0130078 |
| 96 | rs384522 | 19 | 44405044 | 0.0130078 |
| 96 | rs398099 | 19 | 44405281 | 0.0149458 |
| 96 | rs385321 | 19 | 44405287 | 0.0130078 |
| 96 | rs108775 | 19 | 44405922 | 0.0113285 |
| 96 | rs367283 | 19 | 44406127 | 0.0130078 |
| 96 | rs448823 | 19 | 44406291 | 0.0130078 |
| 96 | rs448829 | 19 | 44406321 | 0.0130078 |
| 96 | rs376328 | 19 | 44407487 | 0.0130078 |
| 96 | rs376032 | 19 | 44407564 | 0.00987249 |
| 96 | rs365556 | 19 | 44408199 | 0.0130078 |
| 96 | rs451945 | 19 | 44408753 | 0.0130078 |
| 96 | rs454813 | 19 | 44408973 | 0.0130078 |
| 96 | rs422457 | 19 | 44409411 | 0.00987249 |
| 96 | rs417400 | 19 | 44410846 | 0.0130078 |
| 96 | rs426534 | 19 | 44412583 | 0.0113285 |
| 96 | rs376457 | 19 | 44412757 | 0.0130078 |
| 96 | rs430667 | 19 | 44413253 | 0.0113285 |
| 96 | rs398964 | 19 | 44413884 | 0.0113285 |
| 96 | rs439665 | 19 | 44414467 | 0.0113285 |
| 96 | rs421512 | 19 | 44415580 | 0.00751261 |
| 96 | rs423765 | 19 | 44416199 | 0.00987249 |
| 96 | rs376069 | 19 | 44417295 | 0.00751261 |
| 96 | rs417699 | 19 | 44417575 | 0.00573186 |
| 96 | rs407731 | 19 | 44418077 | 0.00573186 |
| 96 | rs406968 | 19 | 44418343 | 0.00573186 |

| | | | | |
|---|---|---|---|---|
| 96 | rs425221 | 19 | 44418544 | 0.00573186 |
| 96 | rs388685 | 19 | 44418680 | 0.00501159 |
| 96 | rs388706 | 19 | 44418693 | 0.00573186 |
| 96 | rs387689 | 19 | 44420111 | 0.00573186 |
| 96 | rs398193 | 19 | 44420420 | 0.00573186 |
| 96 | rs378112 | 19 | 44421929 | 0.00573186 |
| 96 | rs378109 | 19 | 44421937 | 0.00655995 |
| 96 | rs450308 | 19 | 44422358 | 0.00573186 |
| 96 | rs424729 | 19 | 44422802 | 0.00573186 |
| 96 | rs423320 | 19 | 44422982 | 0.00573186 |
| 96 | rs423752 | 19 | 44423195 | 0.00573186 |
| 96 | rs375066 | 19 | 44423570 | 0.00573186 |
| 96 | rs384329 | 19 | 44423981 | 0.00501159 |
| 96 | rs424410 | 19 | 44424794 | 0.00573186 |
| 96 | rs408549 | 19 | 44425619 | 0.00501159 |
| 96 | rs379785 | 19 | 44428101 | 0.0184948 |
| 96 | rs428505 | 19 | 44428911 | 0.00501159 |
| 97 | rs8115191 | 20 | 56019801 | 0.16629 |
| 97 | rs8115156 | 20 | 56019846 | 0.16629 |
| 97 | rs34161672 | 20 | 56020599 | 0.659985 |
| 98 | rs35968201 | 20 | 58998561 | 0.11012 |
| 98 | rs35384758 | 20 | 58998651 | 0.157675 |
| 98 | rs6100897 | 20 | 59001318 | 0.0686871 |
| 98 | rs6092939 | 20 | 59002039 | 0.0417132 |
| 98 | rs348843 | 20 | 59002646 | 0.0868882 |
| 98 | rs520838 | 20 | 59002955 | 0.0466945 |
| 98 | rs6100898 | 20 | 59003029 | 0.0977937 |
| 98 | rs6100899 | 20 | 59005430 | 0.0417132 |
| 98 | rs8126294 | 20 | 59009717 | 0.0868882 |
| 98 | rs6100902 | 20 | 59010700 | 0.0868882 |
| 98 | rs9305113 | 20 | 59011174 | 0.11012 |
| 98 | rs6100906 | 20 | 59022494 | 0.00729625 |
| 98 | rs6100916 | 20 | 59027565 | 0.00591544 |
| 98 | rs6100920 | 20 | 59030191 | 0.00656818 |
| 99 | rs2832279 | 21 | 30613466 | 0.475566 |
| 99 | rs1153277 | 21 | 30666867 | 0.0159243 |
| 99 | rs1153282 | 21 | 30681507 | 0.0252185 |
| 99 | rs1153286 | 21 | 30684383 | 0.0156894 |
| 99 | rs1153288 | 21 | 30686091 | 0.0087024 |
| 99 | rs1153289 | 21 | 30686212 | 0.00749396 |
| 99 | rs1153290 | 21 | 30687404 | 0.010112 |
| 99 | rs1153291 | 21 | 30689070 | 0.0159243 |
| 99 | rs1153292 | 21 | 30689195 | 0.00645736 |
| 99 | rs1153293 | 21 | 30691325 | 0.011671 |
| 99 | rs1312254 | 21 | 30695401 | 0.021622 |
| 99 | rs371432 | 21 | 30710876 | 0.0087024 |
| 99 | rs368322 | 21 | 30717151 | 0.0185499 |
| 99 | rs15092 | 21 | 30718201 | 0.010112 |
| 99 | rs382732 | 21 | 30719010 | 0.021622 |
| 99 | rs117213 | 21 | 30720536 | 0.021622 |
| 99 | rs407712 | 21 | 30722531 | 0.0185499 |
| 99 | rs8129076 | 21 | 30732953 | 0.0136788 |

| | | | | |
|---|---|---|---|---|
| 99 | rs2832291 | 21 | 30734388 | 0.0182072 |
| 99 | rs11088117 | 21 | 30737816 | 0.0159243 |
| 99 | rs2832293 | 21 | 30738872 | 0.0469601 |
| 99 | rs2832294 | 21 | 30739671 | 0.0136788 |
| 99 | rs2832295 | 21 | 30739843 | 0.0136788 |
| 99 | rs2832296 | 21 | 30741454 | 0.010112 |
| 99 | rs2832297 | 21 | 30742996 | 0.0087024 |
| 99 | rs2832298 | 21 | 30743863 | 0.021622 |
| 99 | rs4817285 | 21 | 30745169 | 0.0117573 |
| 99 | rs2832299 | 21 | 30749237 | 0.00749396 |
| 99 | rs2832300 | 21 | 30749315 | 0.0117573 |
| 99 | rs2832301 | 21 | 30749712 | 0.0087024 |
| 99 | rs2832302 | 21 | 30749793 | 0.0087024 |
| 99 | rs12627575 | 21 | 30750064 | 0.0182072 |
| 99 | rs8132992 | 21 | 30750561 | 0.010112 |
| 100 | rs3788410 | 22 | 29670939 | 0.00942008 |
| 100 | rs2301585 | 22 | 29702748 | 0.0177788 |
| 100 | rs6006090 | 22 | 29705927 | 0.00573134 |
| 100 | rs5763139 | 22 | 29717802 | 0.0388924 |
| 100 | rs174764 | 22 | 29721161 | 0.0444093 |
| 100 | rs174765 | 22 | 29727866 | 0.00648334 |
| 100 | rs3788411 | 22 | 29728783 | 0.229047 |
| 100 | rs98069 | 22 | 29730625 | 0.229047 |
| 100 | rs174767 | 22 | 29740728 | 0.210272 |
| 100 | rs2283849 | 22 | 29743482 | 0.160178 |
| 101 | rs35318931 | 23 | 38009121 | 0.872827 |
| 101 | rs7890623 | 23 | 38011836 | 0.0560869 |
| 101 | rs112900563 | 23 | 38017972 | 0.0694953 |
| 102 | rs2883091 | 23 | 109684139 | 0.00272715 |
| 102 | rs57639839 | 23 | 109684454 | 0.00230645 |
| 102 | rs10521528 | 23 | 109689152 | 0.00165378 |
| 102 | rs6655082 | 23 | 109689468 | 0.00195224 |
| 102 | rs767225 | 23 | 109706141 | 0.00140208 |
| 102 | rs5942634 | 23 | 109707641 | 0.00272715 |
| 102 | rs12836511 | 23 | 109709965 | 0.00140208 |
| 102 | rs12010891 | 23 | 109710796 | 0.0024131 |
| 102 | rs139566916 | 23 | 109711686 | 0.00140208 |
| 102 | rs5985476 | 23 | 109716989 | 0.00140208 |
| 102 | rs7059210 | 23 | 109718772 | 0.00140208 |
| 102 | rs12847546 | 23 | 109730477 | 0.0015057 |
| 102 | rs5942942 | 23 | 109730899 | 0.00165378 |
| 102 | rs2353114 | 23 | 109731709 | 0.00140208 |
| 102 | rs5942636 | 23 | 109733701 | 0.00140208 |
| 102 | rs7876360 | 23 | 109739964 | 0.00453029 |
| 102 | rs5942640 | 23 | 109740592 | 0.00322721 |
| 102 | rs7065452 | 23 | 109753081 | 0.00632126 |
| 102 | rs5942951 | 23 | 109757981 | 0.00453029 |
| 102 | rs12396834 | 23 | 109758608 | 0.00453029 |
| 102 | rs5942952 | 23 | 109761594 | 0.00322721 |
| 102 | rs5942953 | 23 | 109761850 | 0.00322721 |
| 102 | rs12395955 | 23 | 109762653 | 0.00322721 |
| 102 | rs5942955 | 23 | 109763603 | 0.00322721 |

| | | | | |
|---|---|---|---|---|
| 102 | rs5942642 | 23 | 109765216 | 0.00453029 |
| 102 | rs5942643 | 23 | 109774409 | 0.00453029 |
| 102 | rs12849634 | 23 | 109774654 | 0.00453029 |
| 102 | rs6567872 | 23 | 109775671 | 0.00322721 |
| 102 | rs6567873 | 23 | 109775713 | 0.00322721 |
| 102 | rs7890102 | 23 | 109775721 | 0.00453029 |
| 102 | rs5985488 | 23 | 109776088 | 0.00322721 |
| 102 | rs5985489 | 23 | 109777306 | 0.00382208 |
| 102 | rs5942957 | 23 | 109778405 | 0.00453029 |
| 102 | rs5985490 | 23 | 109778729 | 0.00272715 |
| 102 | rs7887749 | 23 | 109779387 | 0.00272715 |
| 102 | rs5985280 | 23 | 109779999 | 0.00272715 |
| 102 | rs5942644 | 23 | 109780229 | 0.00272715 |
| 102 | rs5942645 | 23 | 109780426 | 0.00272715 |
| 102 | rs5942646 | 23 | 109780612 | 0.00272715 |
| 102 | rs5985491 | 23 | 109781243 | 0.00272715 |
| 102 | rs5985281 | 23 | 109781800 | 0.00272715 |
| 102 | rs5985492 | 23 | 109782222 | 0.00230645 |
| 102 | rs7060165 | 23 | 109783258 | 0.00230645 |
| 102 | rs12860440 | 23 | 109784311 | 0.00230645 |
| 102 | rs12007540 | 23 | 109784378 | 0.00230645 |
| 102 | rs5942958 | 23 | 109784640 | 0.00230645 |
| 102 | rs12008798 | 23 | 109785471 | 0.00230645 |
| 102 | rs12007981 | 23 | 109785810 | 0.00230645 |
| 102 | rs12013156 | 23 | 109786110 | 0.00195224 |
| 102 | rs12012896 | 23 | 109786119 | 0.00195224 |
| 102 | rs7057679 | 23 | 109787954 | 0.00230645 |
| 102 | rs5942648 | 23 | 109788727 | 0.00322721 |
| 102 | rs5942960 | 23 | 109789335 | 0.00230645 |
| 102 | rs7050023 | 23 | 109789616 | 0.00322721 |
| 102 | rs2019206 | 23 | 109789900 | 0.00322721 |
| 102 | rs947578 | 23 | 109789950 | 0.00230645 |
| 102 | rs5942961 | 23 | 109790394 | 0.00170789 |
| 102 | rs12400785 | 23 | 109792100 | 0.00176076 |
| 102 | rs5985495 | 23 | 109798044 | 0.00230645 |
| 102 | rs5942965 | 23 | 109800345 | 0.00195224 |
| 102 | rs5942649 | 23 | 109801665 | 0.00322721 |
| 102 | rs5942966 | 23 | 109801862 | 0.00322721 |
| 102 | rs5985496 | 23 | 109802133 | 0.00230645 |
| 102 | rs5942967 | 23 | 109803782 | 0.00230645 |
| 102 | rs955230 | 23 | 109806482 | 0.00322721 |
| 102 | rs5942968 | 23 | 109807481 | 0.00195224 |
| 102 | rs7884019 | 23 | 109809489 | 0.00272715 |
| 102 | rs5985497 | 23 | 109810091 | 0.00230645 |
| 102 | rs12392723 | 23 | 109810458 | 0.00272715 |
| 102 | rs5942971 | 23 | 109816218 | 0.00453029 |
| 102 | rs5942972 | 23 | 109816550 | 0.0063803 |
| 102 | rs5942650 | 23 | 109818258 | 0.00453029 |
| 102 | rs1573036 | 23 | 109820068 | 0.00537411 |
| 102 | rs881090 | 23 | 109820220 | 0.00382208 |
| 102 | rs5985498 | 23 | 109822030 | 0.00453029 |
| 102 | rs12832585 | 23 | 109823282 | 0.00758106 |

| | | | | |
|---|---|---|---|---|
| 102 | rs5985499 | 23 | 109825358 | 0.00170789 |
| 102 | rs5942651 | 23 | 109827622 | 0.00901514 |
| 102 | rs5942974 | 23 | 109827725 | 0.0063803 |
| 102 | rs5942975 | 23 | 109828204 | 0.00901514 |
| 102 | rs2211379 | 23 | 109829677 | 0.00901514 |
| 102 | rs2226123 | 23 | 109829741 | 0.0063803 |
| 102 | rs5942976 | 23 | 109830181 | 0.00901514 |
| 102 | rs5942652 | 23 | 109830714 | 0.0107293 |
| 102 | rs1418334 | 23 | 109833139 | 0.00758106 |
| 102 | rs1418333 | 23 | 109833420 | 0.00537411 |
| 102 | rs5942977 | 23 | 109833905 | 0.0152344 |
| 102 | rs7063758 | 23 | 109835086 | 0.00382208 |
| 102 | rs7049750 | 23 | 109835230 | 0.00382208 |
| 102 | rs12838248 | 23 | 109836120 | 0.00382208 |
| 102 | rs12011976 | 23 | 109836588 | 0.00453029 |
| 102 | rs12863755 | 23 | 109837061 | 0.00382208 |
| 102 | rs5942978 | 23 | 109837304 | 0.00230645 |
| 102 | rs12862848 | 23 | 109837899 | 0.00230645 |
| 102 | rs5985503 | 23 | 109839225 | 0.00322721 |
| 102 | rs11152612 | 23 | 109839700 | 0.00230645 |
| 102 | rs5942979 | 23 | 109840233 | 0.00195224 |
| 102 | rs5942980 | 23 | 109840240 | 0.00165378 |
| 102 | rs5942981 | 23 | 109840253 | 0.00165378 |
| 102 | rs7887247 | 23 | 109840502 | 0.00230645 |
| 102 | rs7892619 | 23 | 109840648 | 0.00322721 |
| 102 | rs3905142 | 23 | 109841217 | 0.00230645 |
| 102 | rs7059316 | 23 | 109842750 | 0.00170789 |
| 102 | rs7059322 | 23 | 109842764 | 0.00170789 |
| 102 | rs7059343 | 23 | 109843012 | 0.00230645 |
| 102 | rs12007316 | 23 | 109843975 | 0.00272715 |
| 102 | rs7890631 | 23 | 109844332 | 0.00230645 |
| 102 | rs3905148 | 23 | 109844851 | 0.00230645 |
| 102 | rs7051081 | 23 | 109845063 | 0.00230645 |
| 102 | rs7051410 | 23 | 109845313 | 0.00230645 |
| 102 | rs7050479 | 23 | 109845353 | 0.00230645 |
| 102 | rs7056027 | 23 | 109845356 | 0.00230645 |
| 102 | rs34295465 | 23 | 109845755 | 0.00453029 |
| 102 | rs35089502 | 23 | 109845786 | 0.00322721 |
| 102 | rs12398815 | 23 | 109846556 | 0.00123945 |
| 102 | rs12012597 | 23 | 109847870 | 0.00322721 |
| 102 | rs12013506 | 23 | 109847894 | 0.00382208 |
| 102 | rs9779440 | 23 | 109848356 | 0.00453029 |
| 102 | rs5942982 | 23 | 109848491 | 0.00382208 |
| 102 | rs5942983 | 23 | 109848731 | 0.00230645 |
| 102 | rs5942984 | 23 | 109848775 | 0.00170789 |
| 102 | rs5985504 | 23 | 109848995 | 0.00230645 |
| 102 | rs5942985 | 23 | 109849018 | 0.00230645 |
| 102 | rs5985290 | 23 | 109849700 | 0.00145438 |
| 102 | rs146166586 | 23 | 109850122 | 0.00382208 |
| 102 | rs148571111 | 23 | 109851904 | 0.00537411 |
| 102 | rs148888704 | 23 | 109852170 | 0.00382208 |
| 102 | rs79392211 | 23 | 109852215 | 0.00453029 |

| | | | | |
|---|---|---|---|---|
| 102 | rs7886114 | 23 | 109852275 | 0.00170789 |
| 102 | rs7886821 | 23 | 109852380 | 0.00322721 |
| 102 | rs12391073 | 23 | 109852424 | 0.00453029 |
| 102 | rs78950079 | 23 | 109853528 | 0.00453029 |
| 102 | rs5942993 | 23 | 109855331 | 0.00453029 |
| 102 | rs5942994 | 23 | 109855465 | 0.00322721 |
| 102 | rs5942995 | 23 | 109855729 | 0.00322721 |
| 102 | rs5942996 | 23 | 109855818 | 0.00382208 |
| 102 | rs7884441 | 23 | 109855904 | 0.00322721 |
| 102 | rs4285635 | 23 | 109856397 | 0.00382208 |
| 102 | rs5985508 | 23 | 109857041 | 0.00145438 |
| 102 | rs7878983 | 23 | 109857499 | 0.00322721 |
| 102 | rs12848426 | 23 | 109858489 | 0.00382208 |
| 102 | rs5942998 | 23 | 109858647 | 0.00382208 |
| 102 | rs12216949 | 23 | 109859006 | 0.00382208 |
| 102 | rs7062685 | 23 | 109859928 | 0.00382208 |
| 102 | rs12216954 | 23 | 109860661 | 0.00453029 |
| 102 | rs4893457 | 23 | 109861620 | 0.00382208 |
| 102 | rs12012736 | 23 | 109861920 | 0.00382208 |
| 102 | rs12858834 | 23 | 109862029 | 0.00537411 |
| 102 | rs7882714 | 23 | 109865115 | 0.00382208 |
| 102 | rs7882060 | 23 | 109865707 | 0.00382208 |
| 102 | rs5943000 | 23 | 109866362 | 0.00382208 |
| 102 | rs5942656 | 23 | 109866545 | 0.00382208 |
| 102 | rs5943001 | 23 | 109866712 | 0.00537411 |
| 102 | rs5943002 | 23 | 109867110 | 0.00322721 |
| 102 | rs5943003 | 23 | 109867364 | 0.00453029 |
| 102 | rs5943004 | 23 | 109867426 | 0.00382208 |
| 102 | rs66561471 | 23 | 109867550 | 0.00537411 |
| 102 | rs5943005 | 23 | 109867987 | 0.00537411 |
| 102 | rs5942657 | 23 | 109868315 | 0.00453029 |
| 102 | rs5985514 | 23 | 109869336 | 0.00322721 |
| 102 | rs6567883 | 23 | 109869935 | 0.00382208 |
| 102 | rs5942658 | 23 | 109870316 | 0.00382208 |
| 102 | rs7055206 | 23 | 109870607 | 0.00382208 |
| 102 | rs7059732 | 23 | 109870686 | 0.00382208 |
| 102 | rs12852745 | 23 | 109871628 | 0.00453029 |
| 102 | rs5943007 | 23 | 109872292 | 0.00537411 |
| 102 | rs7066046 | 23 | 109872669 | 0.00382208 |
| 102 | rs7890266 | 23 | 109872789 | 0.00453029 |
| 102 | rs6567884 | 23 | 109873027 | 0.00123945 |
| 102 | rs5985515 | 23 | 109873137 | 0.00453029 |
| 102 | rs5943008 | 23 | 109873511 | 0.00382208 |
| 102 | rs5943010 | 23 | 109874287 | 0.00382208 |
| 102 | rs5942659 | 23 | 109874542 | 0.00236063 |
| 102 | rs7062458 | 23 | 109874849 | 0.00382208 |
| 102 | rs7062471 | 23 | 109874868 | 0.00382208 |
| 102 | rs5985517 | 23 | 109875306 | 0.00385826 |
| 102 | rs5985518 | 23 | 109875374 | 0.00382208 |
| 102 | rs4893394 | 23 | 109875463 | 0.00382208 |
| 102 | rs5943012 | 23 | 109875876 | 0.00382208 |
| 102 | rs5943013 | 23 | 109875942 | 0.00537411 |

| | | | | |
|---|---|---|---|---|
| 102 | rs5985519 | 23 | 109876267 | 0.00537411 |
| 102 | rs5985520 | 23 | 109876430 | 0.00382208 |
| 102 | rs4596812 | 23 | 109876794 | 0.00382208 |
| 102 | rs4329433 | 23 | 109876848 | 0.00382208 |
| 102 | rs5943014 | 23 | 109877047 | 0.00382208 |
| 102 | rs5943015 | 23 | 109877196 | 0.00382208 |
| 102 | rs5985522 | 23 | 109877642 | 0.00453029 |
| 102 | rs5943018 | 23 | 109878770 | 0.00322721 |
| 102 | rs5943019 | 23 | 109879017 | 0.00322721 |
| 102 | rs5942660 | 23 | 109879828 | 0.00322721 |
| 102 | rs5942661 | 23 | 109879876 | 0.00453029 |
| 102 | rs6642787 | 23 | 109880402 | 0.00322721 |
| 102 | rs12853845 | 23 | 109880410 | 0.00322721 |
| 102 | rs5985296 | 23 | 109881478 | 0.00236063 |
| 102 | rs12834261 | 23 | 109881597 | 0.00382208 |
| 102 | rs5985523 | 23 | 109881780 | 0.00200714 |
| 102 | rs5943021 | 23 | 109882597 | 0.00200714 |
| 102 | rs5942662 | 23 | 109882692 | 0.00200714 |
| 102 | rs5985297 | 23 | 109883087 | 0.00277853 |
| 102 | rs67427075 | 23 | 109883191 | 0.00277853 |
| 102 | rs7884700 | 23 | 109883343 | 0.00128852 |
| 102 | rs78458801 | 23 | 109884075 | 0.00277853 |
| 102 | rs143567589 | 23 | 109885039 | 0.00455179 |
| 102 | rs141907656 | 23 | 109885700 | 0.00744075 |
| 102 | rs9779762 | 23 | 109886050 | 0.00200714 |
| 102 | rs9779765 | 23 | 109886105 | 0.00200714 |
| 102 | rs7886076 | 23 | 109886636 | 0.00277853 |
| 102 | rs7882275 | 23 | 109888695 | 0.00176076 |
| 102 | rs7887476 | 23 | 109888719 | 0.00335889 |
| 102 | rs7884032 | 23 | 109888908 | 0.00176076 |
| 102 | rs7060071 | 23 | 109889011 | 0.00634989 |
| 102 | rs7061171 | 23 | 109889020 | 0.0018124 |
| 102 | rs7061697 | 23 | 109889021 | 0.00211246 |
| 102 | rs7061313 | 23 | 109889100 | 0.00170789 |
| 102 | rs5943022 | 23 | 109889570 | 0.00170789 |
| 102 | rs5942663 | 23 | 109889662 | 0.00170789 |
| 102 | rs5985529 | 23 | 109890023 | 0.00170789 |
| 102 | rs5985530 | 23 | 109890086 | 0.00277853 |
| 102 | rs5985531 | 23 | 109890221 | 0.00200714 |
| 102 | rs5942664 | 23 | 109890719 | 0.00170789 |
| 102 | rs5942665 | 23 | 109890903 | 0.00170789 |
| 102 | rs5942666 | 23 | 109890925 | 0.00170789 |
| 102 | rs5985532 | 23 | 109891443 | 0.00236063 |
| 102 | rs5985300 | 23 | 109891479 | 0.00453029 |
| 102 | rs12389314 | 23 | 109891673 | 0.00453029 |
| 102 | rs9698903 | 23 | 109891750 | 0.00145438 |
| 102 | rs12391613 | 23 | 109892037 | 0.00453029 |
| 102 | rs5985301 | 23 | 109892480 | 0.00453029 |
| 102 | rs5943023 | 23 | 109892754 | 0.00453029 |
| 102 | rs5985533 | 23 | 109893897 | 0.00453029 |
| 102 | rs5943024 | 23 | 109894390 | 0.00453029 |
| 102 | rs5943025 | 23 | 109894401 | 0.00236063 |

| | | | | |
|---|---|---|---|---|
| 102 | rs5985534 | 23 | 109895239 | 0.00453029 |
| 102 | rs5985535 | 23 | 109895249 | 0.00453029 |
| 102 | rs12854492 | 23 | 109895684 | 0.00453029 |
| 102 | rs5943026 | 23 | 109895746 | 0.00453029 |
| 102 | rs5942667 | 23 | 109896002 | 0.00236063 |
| 102 | rs5942668 | 23 | 109896225 | 0.00200714 |
| 102 | rs12854852 | 23 | 109896941 | 0.00145438 |
| 102 | rs5943027 | 23 | 109897121 | 0.00145438 |
| 102 | rs5943030 | 23 | 109897778 | 0.00145438 |
| 102 | rs5943031 | 23 | 109898152 | 0.00145438 |
| 102 | rs5943032 | 23 | 109898287 | 0.00145438 |
| 102 | rs5943033 | 23 | 109899250 | 0.00200714 |
| 102 | rs5943034 | 23 | 109899391 | 0.00170789 |
| 102 | rs5942669 | 23 | 109899745 | 0.00145438 |
| 102 | rs12008352 | 23 | 109900338 | 0.00453029 |
| 102 | rs60983763 | 23 | 109901389 | 0.00537411 |
| 102 | rs58665200 | 23 | 109901444 | 0.00382208 |
| 102 | rs56341516 | 23 | 109901490 | 0.00200714 |
| 102 | rs5943035 | 23 | 109901693 | 0.00382208 |
| 102 | rs5943036 | 23 | 109901872 | 0.00200714 |
| 102 | rs12007422 | 23 | 109902012 | 0.00200714 |
| 102 | rs35938117 | 23 | 109902393 | 0.00537411 |
| 102 | rs12839543 | 23 | 109903194 | 0.00382208 |
| 102 | rs5943037 | 23 | 109904365 | 0.00453029 |
| 102 | rs5943038 | 23 | 109904486 | 0.00382208 |
| 102 | rs6567897 | 23 | 109904587 | 0.00537411 |
| 102 | rs4631619 | 23 | 109904861 | 0.00200714 |
| 102 | rs7880404 | 23 | 109906577 | 0.00537411 |
| 102 | rs5943039 | 23 | 109907243 | 0.00537411 |
| 102 | rs2179764 | 23 | 109907482 | 0.00382208 |
| 102 | rs12838235 | 23 | 109908613 | 0.00322721 |
| 102 | rs2206971 | 23 | 109909035 | 0.00322721 |
| 102 | rs5942673 | 23 | 109912906 | 0.00119468 |
| 102 | rs2143760 | 23 | 109913102 | 0.00119468 |
| 102 | rs2223782 | 23 | 109918851 | 0.00119468 |
| 102 | rs5942676 | 23 | 109925412 | 0.00211246 |
| 102 | rs12857107 | 23 | 109926710 | 0.0018124 |
| 102 | rs5943047 | 23 | 109927567 | 0.0018124 |
| 102 | rs5943049 | 23 | 109928545 | 0.0018124 |
| 102 | rs5943050 | 23 | 109929063 | 0.0018124 |
| 102 | rs5943051 | 23 | 109929506 | 0.0018124 |
| 102 | rs10047014 | 23 | 109929651 | 0.0018124 |
| 102 | rs10047007 | 23 | 109929882 | 0.0018124 |
| 102 | rs5942677 | 23 | 109930730 | 0.0018124 |
| 102 | rs5942678 | 23 | 109931042 | 0.00211246 |
| 102 | rs5943052 | 23 | 109931799 | 0.0018124 |
| 102 | rs5943053 | 23 | 109931856 | 0.0018124 |
| 102 | rs2016879 | 23 | 109933937 | 0.00195224 |
| 102 | rs10482447 | 23 | 109934939 | 0.00195224 |
| 102 | rs5943055 | 23 | 109935846 | 0.00195224 |
| 102 | rs5943056 | 23 | 109935983 | 0.00195224 |
| 102 | rs5942680 | 23 | 109938188 | 0.00322721 |

| | | | | |
|---|---|---|---|---|
| 102 | rs5942681 | 23 | 109938369 | 0.00230645 |
| 102 | rs73260106 | 23 | 109938577 | 0.00455179 |
| 102 | rs5943057 | 23 | 109939205 | 0.00165378 |
| 102 | rs5985544 | 23 | 109940195 | 0.00165378 |
| 102 | rs5985545 | 23 | 109940296 | 0.00140208 |
| 102 | rs5985546 | 23 | 109940577 | 0.00140208 |
