## Supplementary Table 4 for "Genome-wide analysis of 944,133 individuals provides insights into the etiology of hemorrhoidal disease"

**Supplementary Table 4. Association of HEM risk variants with other traits and diseases.**

This table contains other reported GWAS associations of 102 lead SNPs (and/or their r2>0.8 LD proxies) of HEM for other traits, extracted from three PheWAS databases: PhenoScanner, GWAS catalog and GWAS ATLAS.

**Lead SNP (rsID)**: rs ID retrieved from NCBI's dbSNP build v150; **Lead SNP BP**: base pair position of the lead SNP (in "chromosome:base pair" format), genomic positions were retrieved from NCBI's dbSNP build v150 (genome build hg19); **Proxy SNP (rsID)**: rs ID of a SNP in LD ($r^2$>0.8) with the lead SNP and has a reported GWAS association with other traits; **Proxy SNP BP:** base pair position of the proxy SNP (in "chromosome:base pair" format); **r²**: the square of the correlation coefficient; **Associated trait:** the name of thar trait that has a reported GWAS association with the lead SNP or its proxy SNP; **Domain:** the assigned trait category of the trait (only available for traits from GWAS ATLAS); **PheWAS database:** the database where the reported association is extracted from; **Data source or PUBMEDID:** the publication reference or data source (UKBB, http://www.nealelab.is/uk-biobank) of the reported GWAS association.

| Lead SNP (rsID) | Lead SNP BP | Proxy SNP (rsID) | Proxy SNP BP | r2 | Associated trait | Domain | PheWAS database | Data source or PUBMEDID |
|---|---|---|---|---|---|---|---|---|
| rs145163454 | 1:169090748 | - | - | - | Antithrombotic agents | Environmental | GWAS_ATLAS | 31015401 |
| rs145163454 | 1:169090748 | rs2227246 | 1:169208179 | 1 | Benign lipomatous neoplasm | - | Phenoscanner | UKBB |
| rs145163454 | 1:169090748 | - | - | - | Blood clot in the leg | - | Phenoscanner | UKBB |
| rs145163454 | 1:169090748 | - | - | - | Blood clot in the lung | - | Phenoscanner | UKBB |
| rs145163454 | 1:169090748 | - | - | - | No blood clot, bronchitis, emphysema, asthma, rhinitis, eczema or allergy diagnosed by doctor | - | Phenoscanner | UKBB |
| rs145163454 | 1:169090748 | rs2227246 | 1:169208179 | 1 | Other soft tissue disorders | - | Phenoscanner | UKBB |
| rs145163454 | 1:169090748 | - | - | - | Personal history of certain other diseases | Mortality | GWAS_ATLAS | 31427789 |
| rs145163454 | 1:169090748 | rs2040445 | 1:169216412 | 0,91575 | Personal history of medical treatment | Mortality | GWAS_ATLAS | 31427789 |
| rs145163454 | 1:169090748 | - | - | - | Phlebitis and thrombophlebitis | - | Phenoscanner | UKBB |
| rs145163454 | 1:169090748 | - | - | - | Pulmonary embolism | - | Phenoscanner | UKBB |
| rs145163454 | 1:169090748 | - | - | - | Self-reported clotting disorder or excessive bleeding | - | Phenoscanner | UKBB |
| rs145163454 | 1:169090748 | - | - | - | Self-reported deep venous thrombosis | - | Phenoscanner | UKBB |
| rs145163454 | 1:169090748 | - | - | - | Self-reported hereditary or genetic haematological disorder | - | Phenoscanner | UKBB |
| rs145163454 | 1:169090748 | - | - | - | Self-reported pulmonary embolism + or - dvt | - | Phenoscanner | UKBB |
| rs145163454 | 1:169090748 | - | - | - | Treatment with warfarin | - | Phenoscanner | UKBB |
| rs145163454 | 1:169090748 | - | - | - | venous thromboembolism | - | GWAS_catalog | 31420334 |
| rs145163454 | 1:169090748 | rs2227246 | 1:169208179 | 1 | Venous thrombosis | - | Phenoscanner | 22675575 |
| rs4951080 | 1:204533284 | rs7556655 | 1:204493844 | 0,93755 | Benign neoplasm of colon, rectum, anus and anal canal | Neoplasms | GWAS_ATLAS | 31427789 |
| rs4951080 | 1:204533284 | rs2290854 | 1:204516025 | 0,93755 | breast cancer in brca1 mutation carriers | - | GWAS_catalog | 23544013 |
| rs4951080 | 1:204533284 | rs16853958 | 1:204528344 | 0,93318 | diastolic blood pressure x alcohol consumption interaction (2df test) | - | GWAS_catalog | 29912962 |
| rs4951080 | 1:204533284 | - | - | - | Forced vital capacity | - | Phenoscanner | UKBB |
| rs4951080 | 1:204533284 | - | - | - | Forced vital capacity, best measure | - | Phenoscanner | UKBB |
| rs4951080 | 1:204533284 | rs12139477 | 1:204553104 | 0,85018 | FVC | Respiratory | GWAS_ATLAS | 30804560 |
| rs4951080 | 1:204533284 | rs10494852 | 1:204457786 | 0,91995 | Hair colour: Black | Dermatological | GWAS_ATLAS | 31427789 |
| rs4951080 | 1:204533284 | rs10900594 | 1:204470129 | 0,933 | Height | Skeletal | GWAS_ATLAS | 30124842 |
| rs4951080 | 1:204533284 | rs10793765 | 1:204549375 | 0,95496 | height | - | GWAS_catalog | 30595370 |
| rs4951080 | 1:204533284 | rs11240751 | 1:204462050 | 0,9113 | Relative age of first facial hair | Reproduction | GWAS_ATLAS | 31427789 |
| rs2605097 | 1:219642109 | rs2605100 | 1:219644224 | 0,97226 | adiposity | - | GWAS_catalog | 19557161 |
| rs2605097 | 1:219642109 | - | - | - | Arm fat mass left | - | Phenoscanner | UKBB |
| rs2605097 | 1:219642109 | - | - | - | Arm fat mass right | - | Phenoscanner | UKBB |
| rs2605097 | 1:219642109 | - | - | - | Arm fat percentage left | - | Phenoscanner | UKBB |
| rs2605097 | 1:219642109 | - | - | - | Arm fat percentage right | - | Phenoscanner | UKBB |
| rs2605097 | 1:219642109 | - | - | - | Body fat percentage | - | Phenoscanner | UKBB |
| rs2605097 | 1:219642109 | rs2605091 | 1:219640305 | 0,97686 | Hip circumference | Metabolic | GWAS_ATLAS | 23754948 |
| rs2605097 | 1:219642109 | - | - | - | Hip circumference | - | Phenoscanner | 25673412 |
| rs2605097 | 1:219642109 | rs2605098 | 1:219643649 | 0,87565 | hip circumference | - | GWAS_catalog | 31453325 |
| rs2605097 | 1:219642109 | - | - | - | Hip circumference adjusted for BMI | - | Phenoscanner | 25673412 |
| rs2605097 | 1:219642109 | rs748273 | 1:219650950 | 0,95848 | hip circumference adjusted for bmi | - | GWAS_catalog | 28552196 |
| rs2605097 | 1:219642109 | - | - | - | Hip circumference in females | - | Phenoscanner | 25673412 |
| rs2605097 | 1:219642109 | rs2605098 | 1:219643649 | 0,87565 | hip circumference variance | - | GWAS_catalog | 31453325 |
| rs2605097 | 1:219642109 | rs2605091 | 1:219640305 | 0,97686 | Impedance measures - Arm fat mass (left) | Metabolic | GWAS_ATLAS | 31427789 |
| rs2605097 | 1:219642109 | rs2605091 | 1:219640305 | 0,97686 | Impedance measures - Arm fat mass (right) | Metabolic | GWAS_ATLAS | 31427789 |
| rs2605097 | 1:219642109 | rs2605091 | 1:219640305 | 0,97686 | Impedance measures - Arm fat percentage (left) | Metabolic | GWAS_ATLAS | 31427789 |
| rs2605097 | 1:219642109 | rs2605091 | 1:219640305 | 0,97686 | Impedance measures - Arm fat percentage (right) | Metabolic | GWAS_ATLAS | 31427789 |
| rs2605097 | 1:219642109 | rs2605091 | 1:219640305 | 0,97686 | Impedance measures - Body fat percentage | Metabolic | GWAS_ATLAS | 31427789 |
| rs2605097 | 1:219642109 | rs2605091 | 1:219640305 | 0,97686 | Impedance measures - Impedance of arm (left) | Metabolic | GWAS_ATLAS | 31427789 |
| rs2605097 | 1:219642109 | rs2605091 | 1:219640305 | 0,97686 | Impedance measures - Impedance of arm (right) | Metabolic | GWAS_ATLAS | 31427789 |
| rs2605097 | 1:219642109 | rs2605091 | 1:219640305 | 0,97686 | Impedance measures - Impedance of whole body | Metabolic | GWAS_ATLAS | 31427789 |
| rs2605097 | 1:219642109 | rs2605091 | 1:219640305 | 0,97686 | Impedance measures - Leg fat mass (left) | Metabolic | GWAS_ATLAS | 31427789 |
| rs2605097 | 1:219642109 | rs2605091 | 1:219640305 | 0,97686 | Impedance measures - Leg fat mass (right) | Metabolic | GWAS_ATLAS | 31427789 |
| rs2605097 | 1:219642109 | rs2605091 | 1:219640305 | 0,97686 | Impedance measures - Leg fat percentage (left) | Metabolic | GWAS_ATLAS | 31427789 |
| rs2605097 | 1:219642109 | rs2605091 | 1:219640305 | 0,97686 | Impedance measures - Leg fat percentage (right) | Metabolic | GWAS_ATLAS | 31427789 |
| rs2605097 | 1:219642109 | rs2605091 | 1:219640305 | 0,97686 | Impedance measures - Trunk fat mass | Metabolic | GWAS_ATLAS | 31427789 |
| rs2605097 | 1:219642109 | rs2605091 | 1:219640305 | 0,97686 | Impedance measures - Trunk fat percentage | Metabolic | GWAS_ATLAS | 31427789 |
| rs2605097 | 1:219642109 | rs2605091 | 1:219640305 | 0,97686 | Impedance measures - Weight | Metabolic | GWAS_ATLAS | 31427789 |
| rs2605097 | 1:219642109 | rs2605091 | 1:219640305 | 0,97686 | Impedance measures - Whole body fat mass | Metabolic | GWAS_ATLAS | 31427789 |
| rs2605097 | 1:219642109 | - | - | - | Impedance of arm left | - | Phenoscanner | UKBB |
| rs2605097 | 1:219642109 | - | - | - | Impedance of arm right | - | Phenoscanner | UKBB |
| rs2605097 | 1:219642109 | - | - | - | Impedance of whole body | - | Phenoscanner | UKBB |
| rs2605097 | 1:219642109 | rs2605091 | 1:219640305 | 0,97686 | Inguinal hernia | Gastrointestinal | GWAS_ATLAS | 31427789 |
| rs2605097 | 1:219642109 | - | - | - | Leg fat mass left | - | Phenoscanner | UKBB |
| rs2605097 | 1:219642109 | - | - | - | Leg fat mass right | - | Phenoscanner | UKBB |
| rs2605097 | 1:219642109 | - | - | - | Leg fat percentage left | - | Phenoscanner | UKBB |
| rs2605097 | 1:219642109 | - | - | - | Leg fat percentage right | - | Phenoscanner | UKBB |
| rs2605097 | 1:219642109 | rs12036836 | 1:219626676 | 0,97255 | Leg-lef fat ratio | Metabolic | GWAS_ATLAS | 30664634 |
| rs2605097 | 1:219642109 | rs2820436 | 1:219640680 | 0,88026 | osteoarthritis | - | GWAS_catalog | 30664745 |
| rs2605097 | 1:219642109 | rs2820436 | 1:219640680 | 0,88026 | osteoarthritis (hip) | - | GWAS_catalog | 30374069 |
| rs2605097 | 1:219642109 | rs2820436 | 1:219640680 | 0,88026 | osteoarthritis (hospital diagnosed) | - | GWAS_catalog | 29559693 |
| rs2605097 | 1:219642109 | rs12036836 | 1:219626676 | 0,97255 | Osteoarthritis of hip or knee | Skeletal | GWAS_ATLAS | 30664745 |
| rs2605097 | 1:219642109 | rs2820436 | 1:219640680 | 0,88026 | osteoarthritis of the hip (hospital diagnosed) | - | GWAS_catalog | 29559693 |
| rs2605097 | 1:219642109 | rs2820436 | 1:219640680 | 0,88026 | osteoarthritis of the hip or knee (hospital diagnosed) | - | GWAS_catalog | 29559693 |
| rs2605097 | 1:219642109 | - | - | - | Trunk fat mass | - | Phenoscanner | UKBB |
| rs2605097 | 1:219642109 | - | - | - | Trunk fat percentage | - | Phenoscanner | UKBB |
| rs2605097 | 1:219642109 | rs12036836 | 1:219626676 | 0,97255 | Trunk-trunk fat ratio | Metabolic | GWAS_ATLAS | 30664634 |
| rs2605097 | 1:219642109 | rs2605091 | 1:219640305 | 0,97686 | Type 2 Diabetes | Endocrine | GWAS_ATLAS | 30297969 |
| rs2605097 | 1:219642109 | rs2820436 | 1:219640680 | 0,88026 | Waist circumference | Metabolic | GWAS_ATLAS | 25673412 |
| rs2605097 | 1:219642109 | - | - | - | Waist circumference adjusted for smoking in females | - | Phenoscanner | 28443625 |
| rs2605097 | 1:219642109 | - | - | - | Waist hip ratio | - | Phenoscanner | 20935629 |
| rs2605097 | 1:219642109 | - | - | - | Waist hip ratio adjusted for BMI | - | Phenoscanner | 25673412 |
| rs2605097 | 1:219642109 | - | - | - | Waist hip ratio adjusted for BMI in females greater than 50 years of age | - | Phenoscanner | 26426971 |
| rs2605097 | 1:219642109 | - | - | - | Waist hip ratio adjusted for BMI in females less than or equal to 50 years of age | - | Phenoscanner | 26426971 |
| rs2605097 | 1:219642109 | - | - | - | Waist hip ratio adjusted for physical activity | - | Phenoscanner | 28448500 |
| rs2605097 | 1:219642109 | - | - | - | Waist hip ratio adjusted for physical activity in females | - | Phenoscanner | 28448500 |
| rs2605097 | 1:219642109 | - | - | - | Waist hip ratio adjusted for smoking | - | Phenoscanner | 28443625 |
| rs2605097 | 1:219642109 | - | - | - | Waist hip ratio in female non-smokers | - | Phenoscanner | 28443625 |
| rs2605097 | 1:219642109 | - | - | - | Waist hip ratio in females | - | Phenoscanner | 25673412 |
| rs2605097 | 1:219642109 | - | - | - | Waist hip ratio in non-smokers | - | Phenoscanner | 28443625 |
| rs2605097 | 1:219642109 | - | - | - | Waist hip ratio in physically active females | - | Phenoscanner | 28448500 |
| rs2605097 | 1:219642109 | - | - | - | Waist hip ratio in physically active indivdiuals | - | Phenoscanner | 28448500 |
| rs2605097 | 1:219642109 | rs2605091 | 1:219640305 | 0,97686 | Waist-hip ratio | Metabolic | GWAS_ATLAS | 20935629 |
| rs2605097 | 1:219642109 | rs1563355 | 1:219653101 | 0,8314 | waist-hip ratio | - | GWAS_catalog | 30239722 |
| rs2605097 | 1:219642109 | rs2605101 | 1:219644496 | 0,87636 | waist-to-hip ratio adjusted for bmi | - | GWAS_catalog | 26426971 |
| rs2605097 | 1:219642109 | rs2820436 | 1:219640680 | 0,88026 | waist-to-hip ratio adjusted for bmi (joint analysis for main effect and physical activity interaction) | - | GWAS_catalog | 28448500 |
| rs2605097 | 1:219642109 | rs2820436 | 1:219640680 | 0,88026 | waist-to-hip ratio adjusted for bmi in active individuals | - | GWAS_catalog | 28448500 |
| rs2605097 | 1:219642109 | rs2820436 | 1:219640680 | 0,88026 | waist-to-hip ratio adjusted for bmi in inactive individuals | - | GWAS_catalog | 28448500 |
| rs2605097 | 1:219642109 | rs2605101 | 1:219644496 | 0,87636 | waist-to-hip ratio adjusted for bmi x sex interaction | - | GWAS_catalog | 26426971 |
| rs2605097 | 1:219642109 | rs2605101 | 1:219644496 | 0,87636 | waist-to-hip ratio adjusted for bmi x sex x age interaction (4df test) | - | GWAS_catalog | 26426971 |
| rs2605097 | 1:219642109 | rs2820436 | 1:219640680 | 0,88026 | waist-to-hip ratio adjusted for body mass index | - | GWAS_catalog | 28448500 |
| rs2605097 | 1:219642109 | rs2605091 | 1:219640305 | 0,97686 | Weight | Metabolic | GWAS_ATLAS | 31427789 |
| rs2605097 | 1:219642109 | - | - | - | Whole body fat mass | - | Phenoscanner | UKBB |
| rs6723226 | 2:32849207 | rs17428810 | 2:32736043 | 0,81635 | cognitive performance | - | GWAS_catalog | 30038396 |
| rs6723226 | 2:32849207 | rs17428810 | 2:32736043 | 0,81635 | cognitive performance (mtag) | - | GWAS_catalog | 30038396 |
| rs6723226 | 2:32849207 | rs17428810 | 2:32736043 | 0,81635 | Height | Skeletal | GWAS_ATLAS | 30124842 |
| rs6723226 | 2:32849207 | rs17428810 | 2:32736043 | 0,81635 | Intelligence | Cognitive | GWAS_ATLAS | 30038396 |
| rs6723226 | 2:32849207 | - | - | - | intelligence (mtag) | - | GWAS_catalog | 29326435 |
| rs6723226 | 2:32849207 | - | - | - | Intelligence multi trait analysis | - | Phenoscanner | 29326435 |
| rs6723226 | 2:32849207 | rs17428810 | 2:32736043 | 0,81635 | Interleukin-18 | Immunological | GWAS_ATLAS | 27989323 |
| rs6723226 | 2:32849207 | rs17428810 | 2:32736043 | 0,81635 | Sitting height | Skeletal | GWAS_ATLAS | 31427789 |
| rs6723226 | 2:32849207 | - | - | - | Sitting height | - | Phenoscanner | UKBB |
| rs6723226 | 2:32849207 | rs17428810 | 2:32736043 | 0,81635 | Verbal-numerical reasoning | Cognitive | GWAS_ATLAS | 29844566 |
| rs4671051 | 2:63032696 | rs360807 | 2:62926859 | 0,80246 | Alcohol intake | Psychiatric | GWAS_ATLAS | 30643251 |
| rs4671051 | 2:63032696 | rs360807 | 2:62926859 | 0,80246 | First PC of the four riskly behaviours | Activities | GWAS_ATLAS | v: https://doi.org/10.1101/2 |
| rs4671051 | 2:63032696 | rs2080227 | 2:63081759 | 0,84175 | HMG CoA reductase inhibitors | Environmental | GWAS_ATLAS | 31015401 |
| rs4671051 | 2:63032696 | rs2710642 | 2:63149557 | 0,92589 | ldl cholesterol | - | GWAS_catalog | 24097068 |
| rs4671051 | 2:63032696 | rs360804 | 2:62939397 | 0,8073 | ldl cholesterol levels | - | GWAS_catalog | 30698716 |
| rs4671051 | 2:63032696 | rs360804 | 2:62939397 | 0,8073 | ldl cholesterol levels in current drinkers | - | GWAS_catalog | 30698716 |
| rs4671051 | 2:63032696 | rs360804 | 2:62939397 | 0,8073 | ldl cholesterol levels x alcohol consumption (drinkers vs non-drinkers) interaction (2df) | - | GWAS_catalog | 30698716 |
| rs4671051 | 2:63032696 | rs360804 | 2:62939397 | 0,8073 | ldl cholesterol levels x alcohol consumption (regular vs non-regular drinkers) interaction (2df) | - | GWAS_catalog | 30698716 |
| rs4671051 | 2:63032696 | rs2710642 | 2:63149557 | 0,92589 | low density lipoprotein cholesterol levels | - | GWAS_catalog | 29507422 |
| rs4671051 | 2:63032696 | rs2421816 | 2:63075033 | 0,95848 | Low-density lipoprotein cholesterol | Metabolic | GWAS_ATLAS | 24097068 |
| rs4671051 | 2:63032696 | rs2080227 | 2:63081759 | 0,84175 | medication use (hmg coa reductase inhibitors) | - | GWAS_catalog | 31015401 |
| rs4671051 | 2:63032696 | rs12713468 | 2:62983213 | 0,83348 | Total cholesterol | Metabolic | GWAS_ATLAS | 29403010 |
| rs4671051 | 2:63032696 | rs12713468 | 2:62983213 | 0,83348 | total cholesterol levels | - | GWAS_catalog | 29403010 |
| rs34532102 | 2:64905182 | rs34437502 | 2:64888426 | 0,9636 | Monocyte percentage of white cells | Immunological | GWAS_ATLAS | 27863252 |
| rs34532102 | 2:64905182 | - | - | - | Monocyte percentage of white cells | - | Phenoscanner | 27863252 |
| rs34532102 | 2:64905182 | rs34437502 | 2:64888426 | 0,9636 | Red cell distribution width | Immunological | GWAS_ATLAS | 27863252 |
| rs57116599 | 2:112770799 | rs17174870 | 2:112665201 | 0,83448 | multiple sclerosis | - | GWAS_catalog | 21833088 |
| rs7423637 | 2:145818064 | rs2252654 | 2:145801113 | 0,95611 | Bald Pattern 1 | Dermatological | GWAS_ATLAS | 31427789 |
| rs7423637 | 2:145818064 | rs34029033 | 2:145830328 | 0,84425 | Bald Pattern 2 | Dermatological | GWAS_ATLAS | 31427789 |
| rs7423637 | 2:145818064 | rs2252654 | 2:145801113 | 0,95611 | Baldness | Dermatological | GWAS_ATLAS | 30573740 |
| rs7423637 | 2:145818064 | rs7604735 | 2:145839655 | 0,93356 | coronary artery disease | - | GWAS_catalog | 29212778 |
| rs7423637 | 2:145818064 | - | - | - | Coronary artery disease | - | Phenoscanner | 29212778 |
| rs7423637 | 2:145818064 | - | - | - | Diastolic blood pressure | - | Phenoscanner | UKBB |

| | | | | | | | | |
|---|---|---|---|---|---|---|---|---|
| rs7423637 | 2:145818064 | rs2252654 | 2:145801113 | 0,95611 | Educational attainment | Environment | GWAS_ATLAS | 30038396 |
| rs7423637 | 2:145818064 | rs2252383 | 2:145803003 | 0,8569 | educational attainment (years of education) | - | GWAS_catalog | 30595370 |
| rs7423637 | 2:145818064 | rs2252654 | 2:145801113 | 0,95611 | Family (paternal) history of Heart disease | Environment | GWAS_ATLAS | 31427789 |
| rs7423637 | 2:145818064 | rs78809153 | 2:145805251 | 0,96902 | Hair or balding pattern: pattern 4 | - | Phenoscanner | UKBB |
| rs7423637 | 2:145818064 | rs2252654 | 2:145801113 | 0,95611 | Heel bone mineral density | Skeletal | GWAS_ATLAS | 30598549 |
| rs7423637 | 2:145818064 | - | - | - | Mouth or teeth dental problems: dentures | - | Phenoscanner | UKBB |
| rs7423637 | 2:145818064 | - | - | - | Peak expiratory flow | - | Phenoscanner | UKBB |
| rs7423637 | 2:145818064 | rs2252383 | 2:145803003 | 0,8569 | PEF | Respiratory | GWAS_ATLAS | 30804560 |
| rs7423637 | 2:145818064 | - | - | - | Pulse wave arterial stiffness index | - | Phenoscanner | UKBB |
| rs7423637 | 2:145818064 | - | - | - | Pulse wave peak to peak time | - | Phenoscanner | UKBB |
| rs7423637 | 2:145818064 | rs12476923 | 2:145830053 | 0,84808 | self-reported risk-taking behaviour | - | GWAS_catalog | 30271922 |
| rs7559714 | 2:148640768 | rs12992412 | 2:148792665 | 0,81772 | atrial fibrillation | - | GWAS_catalog | 29892015 |
| rs7559714 | 2:148640768 | rs12992412 | 2:148792665 | 0,81772 | Atrial Fibrillation | Cardiovascular | GWAS_ATLAS | 29892015 |
| rs7559714 | 2:148640768 | rs13022319 | 2:148534374 | 0,93768 | Basophil count | Immunological | GWAS_ATLAS | 27863252 |
| rs7559714 | 2:148640768 | - | - | - | Basophil count | - | Phenoscanner | 27863252 |
| rs7559714 | 2:148640768 | rs13022319 | 2:148534374 | 0,93768 | Basophil percentage of granulocytes | Immunological | GWAS_ATLAS | 27863252 |
| rs7559714 | 2:148640768 | - | - | - | Basophil percentage of granulocytes | - | Phenoscanner | 27863252 |
| rs7559714 | 2:148640768 | rs13022319 | 2:148534374 | 0,93768 | Basophil percentage of white cells | Immunological | GWAS_ATLAS | 27863252 |
| rs7559714 | 2:148640768 | - | - | - | Basophil percentage of white cells | - | Phenoscanner | 27863252 |
| rs7559714 | 2:148640768 | rs12990959 | 2:148572160 | 0,9336 | diastolic blood pressure | - | GWAS_catalog | 30224653 |
| rs7559714 | 2:148640768 | rs13026220 | 2:148586459 | 0,99549 | estimated glomerular filtration rate | - | GWAS_catalog | 30604766 |
| rs7559714 | 2:148640768 | rs13022319 | 2:148534374 | 0,93768 | Estimated glomerular filtration rate | Metabolic | GWAS_ATLAS | 31152163 |
| rs7559714 | 2:148640768 | rs4972270 | 2:148543331 | 0,92068 | Estimated glomerular filtration rate based on serum creatinine | Metabolic | GWAS_ATLAS | 26831199 |
| rs7559714 | 2:148640768 | rs4972356 | 2:148582309 | 0,98651 | Neutrophil percentage of granulocytes | Immunological | GWAS_ATLAS | 27863252 |
| rs7559714 | 2:148640768 | - | - | - | Neutrophil percentage of granulocytes | - | Phenoscanner | 27863252 |
| rs7559714 | 2:148640768 | rs12105411 | 2:148800730 | 0,82823 | red blood cell count | - | GWAS_catalog | 30595370 |
| rs7559714 | 2:148640768 | rs1424949 | 2:148542963 | 0,92491 | Serum Urate | Metabolic | GWAS_ATLAS | 23263486 |
| rs7559714 | 2:148640768 | rs2307394 | 2:148716428 | 0,991 | serum uric acid levels | - | GWAS_catalog | 30993211 |
| rs7559714 | 2:148640768 | rs12464617 | 2:148576646 | 0,99551 | Sum eosinophil basophil count | Immunological | GWAS_ATLAS | 27863252 |
| rs7559714 | 2:148640768 | - | - | - | Sum eosinophil basophil counts | - | Phenoscanner | 27863252 |
| rs7559714 | 2:148640768 | rs2307394 | 2:148716428 | 0,991 | urate levels | - | GWAS_catalog | 23263486 |
| rs13017210 | 2:174008623 | rs72905046 | 2:173948571 | 0,85705 | household income (mtag) | - | GWAS_catalog | 31844048 |
| rs847148 | 2:176970456 | - | - | - | Arm fat-free mass left | - | Phenoscanner | UKBB |
| rs847148 | 2:176970456 | - | - | - | Arm fat-free mass right | - | Phenoscanner | UKBB |
| rs847148 | 2:176970456 | - | - | - | Arm predicted mass left | - | Phenoscanner | UKBB |
| rs847148 | 2:176970456 | - | - | - | Arm predicted mass right | - | Phenoscanner | UKBB |
| rs847148 | 2:176970456 | rs847157 | 2:176962501 | 0,87872 | Heel bone mineral density | Skeletal | GWAS_ATLAS | 30048462 |
| rs847148 | 2:176970456 | - | - | - | Heel bone mineral density | - | Phenoscanner | UKBB |
| rs847148 | 2:176970456 | rs847153 | 2:176962989 | 0,87872 | Height | Skeletal | GWAS_ATLAS | 30124842 |
| rs847148 | 2:176970456 | rs847157 | 2:176962501 | 0,87872 | height | - | GWAS_catalog | 30595370 |
| rs847148 | 2:176970456 | rs847157 | 2:176962501 | 0,87872 | Impedance measures - Arm fat-free mass (left) | Metabolic | GWAS_ATLAS | 31427789 |
| rs847148 | 2:176970456 | rs847157 | 2:176962501 | 0,87872 | Impedance measures - Arm fat-free mass (right) | Metabolic | GWAS_ATLAS | 31427789 |
| rs847148 | 2:176970456 | rs847157 | 2:176962501 | 0,87872 | Impedance measures - Arm predicted mass (left) | Metabolic | GWAS_ATLAS | 31427789 |
| rs847148 | 2:176970456 | rs847157 | 2:176962501 | 0,87872 | Impedance measures - Arm predicted mass (right) | Metabolic | GWAS_ATLAS | 31427789 |
| rs847148 | 2:176970456 | rs847157 | 2:176962501 | 0,87872 | Impedance measures - Basal metabolic rate | Metabolic | GWAS_ATLAS | 31427789 |
| rs847148 | 2:176970456 | rs847157 | 2:176962501 | 0,87872 | Impedance measures - Impedance of arm (left) | Metabolic | GWAS_ATLAS | 31427789 |
| rs847148 | 2:176970456 | rs847157 | 2:176962501 | 0,87872 | Impedance measures - Impedance of arm (right) | Metabolic | GWAS_ATLAS | 31427789 |
| rs847148 | 2:176970456 | rs711813 | 2:176966190 | 0,97111 | Impedance measures - Impedance of whole body | Metabolic | GWAS_ATLAS | 31427789 |
| rs847148 | 2:176970456 | rs847157 | 2:176962501 | 0,87872 | Impedance measures - Trunk fat-free mass | Metabolic | GWAS_ATLAS | 31427789 |
| rs847148 | 2:176970456 | rs847157 | 2:176962501 | 0,87872 | Impedance measures - Trunk predicted mass | Metabolic | GWAS_ATLAS | 31427789 |
| rs847148 | 2:176970456 | rs847157 | 2:176962501 | 0,87872 | Impedance measures - Whole body fat-free mass | Metabolic | GWAS_ATLAS | 31427789 |
| rs847148 | 2:176970456 | rs847157 | 2:176962501 | 0,87872 | Impedance measures - Whole body water mass | Metabolic | GWAS_ATLAS | 31427789 |
| rs847148 | 2:176970456 | - | - | - | Impedance of arm left | - | Phenoscanner | UKBB |
| rs847148 | 2:176970456 | - | - | - | Impedance of arm right | - | Phenoscanner | UKBB |
| rs847148 | 2:176970456 | rs741610 | 2:176969206 | 0,97585 | Impedance of whole body | - | Phenoscanner | UKBB |
| rs847148 | 2:176970456 | rs847157 | 2:176962501 | 0,87872 | Sitting height | Skeletal | GWAS_ATLAS | 31427789 |
| rs847148 | 2:176970456 | - | - | - | Trunk fat-free mass | - | Phenoscanner | UKBB |
| rs847148 | 2:176970456 | - | - | - | Trunk predicted mass | - | Phenoscanner | UKBB |
| rs847148 | 2:176970456 | - | - | - | Whole body fat-free mass | - | Phenoscanner | UKBB |
| rs847148 | 2:176970456 | - | - | - | Whole body water mass | - | Phenoscanner | UKBB |
| rs34417560 | 2:191435716 | rs4853718 | 2:191404964 | 0,99489 | Use of sun/uv protection | Activities | GWAS_ATLAS | 31427789 |
| rs1689549 | 3:14837473 | rs1687289 | 3:14829826 | 0,94441 | Coronary artery disease | - | Phenoscanner | 29212778 |
| rs9847710 | 3:53062661 | rs9831861 | 3:53088285 | 0,98774 | colorectal cancer | - | GWAS_catalog | 31089142 |
| rs9847710 | 3:53062661 | rs11715894 | 3:53040459 | 0,97162 | disrupted circadian rhythm (low relative amplitude of rest-activity cycles) | - | GWAS_catalog | 30120083 |
| rs9847710 | 3:53062661 | rs2581778 | 3:53055311 | 0,97162 | Estimated glomerular filtration rate | Metabolic | GWAS_ATLAS | 31152163 |
| rs9847710 | 3:53062661 | rs2581817 | 3:53071797 | 0,9959 | gout | - | GWAS_catalog | 31578528 |
| rs9847710 | 3:53062661 | rs13082026 | 3:52962681 | 0,80691 | mental health study participation (completed survey) | - | GWAS_catalog | 31263887 |
| rs9847710 | 3:53062661 | rs2581824 | 3:53022408 | 0,97162 | Serum Urate | Metabolic | GWAS_ATLAS | 23263486 |
| rs9847710 | 3:53062661 | - | - | - | Serum urate | - | Phenoscanner | 23263486 |
| rs9847710 | 3:53062661 | rs6445559 | 3:53099466 | 0,99184 | serum uric acid levels | - | GWAS_catalog | 29403010 |
| rs9847710 | 3:53062661 | - | - | - | ulcerative colitis | - | GWAS_catalog | 23128233 |
| rs9847710 | 3:53062661 | - | - | - | Ulcerative colitis | - | Phenoscanner | 23128233 |
| rs9847710 | 3:53062661 | rs2244552 | 3:53055522 | 0,97162 | urate levels | - | GWAS_catalog | 31578528 |
| rs9847710 | 3:53062661 | rs2564931 | 3:53020559 | 0,97162 | Uric acid | Metabolic | GWAS_ATLAS | 29403010 |
| rs2597301 | 3:70909494 | rs2687201 | 3:70928930 | 0,89685 | barrett's esophagus | - | GWAS_catalog | 24121790 |
| rs2597301 | 3:70909494 | rs2687202 | 3:70929983 | 0,81315 | barrett's esophagus or esophageal adenocarcinoma | - | GWAS_catalog | 27527254 |
| rs2597301 | 3:70909494 | rs1522551 | 3:70902653 | 0,85326 | Diaphragmatic hernia | Gastrointestinal | GWAS_ATLAS | 31427789 |
| rs2597301 | 3:70909494 | rs2687201 | 3:70928930 | 0,89685 | digestive system disease (barrett's esophagus and esophageal adenocarcinoma combined) | - | GWAS_catalog | 24121790 |
| rs2597301 | 3:70909494 | rs2687201 | 3:70928930 | 0,89685 | esophageal adenocarcinoma | - | GWAS_catalog | 24121790 |
| rs2597301 | 3:70909494 | rs1522551 | 3:70902653 | 0,85326 | FEV1/FVC ratio | Respiratory | GWAS_ATLAS | 30804560 |
| rs2597301 | 3:70909494 | rs1522551 | 3:70902653 | 0,85326 | First PC of the four riskly behaviours | Activities | GWAS_ATLAS | v: https://doi.org/10.1101/2 |
| rs2597301 | 3:70909494 | rs1522551 | 3:70902653 | 0,85326 | Height | Skeletal | GWAS_ATLAS | 30124842 |
| rs2597301 | 3:70909494 | rs13060374 | 3:70913882 | 0,84315 | Impedance measures - Impedance of arm (left) | Metabolic | GWAS_ATLAS | 31427789 |
| rs2597301 | 3:70909494 | rs2687195 | 3:70905581 | 0,83876 | Impedance measures - Impedance of arm (right) | Metabolic | GWAS_ATLAS | 31427789 |
| rs2597301 | 3:70909494 | rs17007949 | 3:70920041 | 0,88678 | Impedance measures - Trunk fat mass | Metabolic | GWAS_ATLAS | 31427789 |
| rs2597301 | 3:70909494 | rs1522551 | 3:70902653 | 0,85326 | Impedance measures - Trunk fat percentage | Metabolic | GWAS_ATLAS | 31427789 |
| rs2597301 | 3:70909494 | rs1522551 | 3:70902653 | 0,85326 | Leg-lef fat ratio | Metabolic | GWAS_ATLAS | 30664634 |
| rs2597301 | 3:70909494 | rs1522554 | 3:70913293 | 0,95472 | lung function (fev1/fvc) | - | GWAS_catalog | 30595370 |
| rs2597301 | 3:70909494 | rs11926363 | 3:70913181 | 0,90013 | Number of sexual partners | Reproduction | GWAS_ATLAS | v: https://doi.org/10.1101/2 |
| rs2597301 | 3:70909494 | rs1522552 | 3:70907252 | 0,98149 | Trunk fat mass | - | Phenoscanner | UKBB |
| rs2597301 | 3:70909494 | - | - | - | Trunk fat percentage | - | Phenoscanner | UKBB |
| rs2597301 | 3:70909494 | rs2597302 | 3:70917189 | 0,87411 | Trunk-trunk fat ratio | Metabolic | GWAS_ATLAS | 30664634 |
| rs6792493 | 3:111477348 | rs4284954 | 3:111468099 | 0,89195 | lobe attachment (rater-scored or self-reported) | - | GWAS_catalog | 29198719 |
| rs9853475 | 3:114500255 | rs9842905 | 3:114498351 | 0,98004 | Height | Skeletal | GWAS_ATLAS | 30124842 |
| rs900400 | 3:156798775 | - | - | - | adiponectin levels in pregnancy | - | GWAS_catalog | 28317342 |
| rs900400 | 3:156798775 | - | - | - | Age at menarche | - | Phenoscanner | 25231870 |
| rs900400 | 3:156798775 | rs4680338 | 3:156794425 | 0,91631 | Age at menarche | Reproduction | GWAS_ATLAS | 31427789 |
| rs900400 | 3:156798775 | rs17451107 | 3:156797609 | 0,95455 | anthropometric traits in newborns | - | GWAS_catalog | 23575227 |
| rs900400 | 3:156798775 | rs1482853 | 3:156798473 | 0,99582 | Anthropometric traits in newborns | - | Phenoscanner | 23575227 |
| rs900400 | 3:156798775 | rs13322435 | 3:156795468 | 0,9319 | birth length (mtag) | - | GWAS_catalog | 31681408 |
| rs900400 | 3:156798775 | - | - | - | Birth weight | - | Phenoscanner | 20372150 |
| rs900400 | 3:156798775 | rs13322435 | 3:156795468 | 0,9319 | birth weight | - | GWAS_catalog | 31097437 |
| rs900400 | 3:156798775 | rs4680338 | 3:156794425 | 0,91631 | Birth weight | Metabolic | GWAS_ATLAS | 31427789 |
| rs900400 | 3:156798775 | rs13322435 | 3:156795468 | 0,9319 | birth weight (mtag) | - | GWAS_catalog | 31681408 |
| rs900400 | 3:156798775 | - | - | - | Birth weight and gestational age | - | Phenoscanner | 23202124 |
| rs900400 | 3:156798775 | rs13322435 | 3:156795468 | 0,9319 | birth weight variance | - | GWAS_catalog | 31453325 |
| rs900400 | 3:156798775 | - | - | - | Birthweight | - | Phenoscanner | 23202124 |
| rs900400 | 3:156798775 | rs900399 | 3:156798732 | 1 | body fat percentage variance | - | GWAS_catalog | 31453325 |
| rs900400 | 3:156798775 | rs4680338 | 3:156794425 | 0,91631 | Bone mineral density | Skeletal | GWAS_ATLAS | 28869591 |
| rs900400 | 3:156798775 | - | - | - | circulating leptin levels | - | GWAS_catalog | 26833098 |
| rs900400 | 3:156798775 | - | - | - | circulating leptin levels | - | Phenoscanner | 26833098 |
| rs900400 | 3:156798775 | - | - | - | circulating leptin levels adjusted for bmi | - | GWAS_catalog | 26833098 |
| rs900400 | 3:156798775 | rs9817452 | 3:156795414 | 0,94636 | hdl cholesterol | - | GWAS_catalog | 30275531 |
| rs900400 | 3:156798775 | rs17451107 | 3:156797609 | 0,95455 | hdl cholesterol levels in current drinkers | - | GWAS_catalog | 30698716 |
| rs900400 | 3:156798775 | rs17451107 | 3:156797609 | 0,95455 | hdl cholesterol levels x alcohol consumption (drinkers vs non-drinkers) interaction (2df) | - | GWAS_catalog | 30698716 |
| rs900400 | 3:156798775 | rs17451107 | 3:156797609 | 0,95455 | hdl cholesterol levels x alcohol consumption (regular vs non-regular drinkers) interaction (2df) | - | GWAS_catalog | 30698716 |
| rs900400 | 3:156798775 | rs56082403 | 3:156797225 | 0,93962 | heel bone mineral density | - | GWAS_catalog | 28869591 |
| rs900400 | 3:156798775 | rs4680338 | 3:156794425 | 0,91631 | Heel bone mineral density | Skeletal | GWAS_ATLAS | 30048462 |
| rs900400 | 3:156798775 | - | - | - | Heel bone mineral density | - | Phenoscanner | UKBB |
| rs900400 | 3:156798775 | - | - | - | Heel bone mineral density left | - | Phenoscanner | UKBB |
| rs900400 | 3:156798775 | rs67261871 | 3:156797941 | 0,99582 | Heel bone mineral density right | - | Phenoscanner | UKBB |
| rs900400 | 3:156798775 | rs900399 | 3:156798732 | 1 | high density lipoprotein cholesterol levels | - | GWAS_catalog | 29083408 |
| rs900400 | 3:156798775 | rs13322435 | 3:156795468 | 0,9319 | Impedance measures - Leg fat percentage (right) | Metabolic | GWAS_ATLAS | 31427789 |
| rs900400 | 3:156798775 | rs13322435 | 3:156795468 | 0,9319 | infant head circumference (mtag) | - | GWAS_catalog | 31681408 |
| rs900400 | 3:156798775 | rs4680338 | 3:156794425 | 0,91631 | mean corpuscular hemoglobin | - | GWAS_catalog | 30595370 |
| rs900400 | 3:156798775 | rs13322435 | 3:156795468 | 0,9319 | menarche (age at onset) | - | GWAS_catalog | 30595370 |
| rs900400 | 3:156798775 | - | - | - | Menarche age at onset | - | Phenoscanner | 25231870 |
| rs900400 | 3:156798775 | rs10049088 | 3:156797648 | 0,95868 | metabolic syndrome | - | GWAS_catalog | 31589552 |
| rs900400 | 3:156798775 | rs1482853 | 3:156798473 | 0,99582 | Newborn birth weight gm | - | Phenoscanner | 23575227 |
| rs900400 | 3:156798775 | - | - | - | Newborn fat mass | - | Phenoscanner | 23575227 |
| rs900400 | 3:156798775 | - | - | - | Newborn sum of skinfolds sqrt mm | - | Phenoscanner | 23575227 |
| rs900400 | 3:156798775 | rs1482852 | 3:156798294 | 0,9435 | offspring birth weight | - | GWAS_catalog | 31043758 |
| rs900400 | 3:156798775 | - | - | - | Ponderal index weightlength3 | - | Phenoscanner | 23202124 |
| rs900400 | 3:156798775 | - | - | - | Ponderal index z score | - | Phenoscanner | 20372150 |
| rs900400 | 3:156798775 | rs13322435 | 3:156795468 | 0,9319 | red blood cell count | - | GWAS_catalog | 30595370 |
| rs900400 | 3:156798775 | rs10049088 | 3:156797648 | 0,95868 | red cell distribution width | - | GWAS_catalog | 30595370 |
| rs900400 | 3:156798775 | rs4680338 | 3:156794425 | 0,91631 | Sitting height | Skeletal | GWAS_ATLAS | 31427789 |
| rs900400 | 3:156798775 | - | - | - | Sitting height | - | Phenoscanner | UKBB |
| rs900400 | 3:156798775 | rs900399 | 3:156798732 | 1 | triglyceride levels | - | GWAS_catalog | 29083408 |
| rs900400 | 3:156798775 | rs17451107 | 3:156797609 | 0,95455 | triglyceride levels x alcohol consumption (drinkers vs non-drinkers) interaction (2df) | - | GWAS_catalog | 30698716 |
| rs900400 | 3:156798775 | rs9817452 | 3:156795414 | 0,94636 | triglycerides | - | GWAS_catalog | 30275531 |
| rs900400 | 3:156798775 | rs4680338 | 3:156794425 | 0,91631 | Waist circumference | Metabolic | GWAS_ATLAS | 25673412 |
| rs900400 | 3:156798775 | - | - | - | Waist circumference | - | Phenoscanner | UKBB |

| | | | | | | | | |
|---|---|---|---|---|---|---|---|---|
| rs900400 | 3:156798775 | - | - | - | Waist circumference adjusted for BMI | - | Phenoscanner | 25673412 |
| rs900400 | 3:156798775 | rs17451107 | 3:156797609 | 0,95455 | waist circumference adjusted for bmi (adjusted for smoking behaviour) | - | GWAS_catalog | 28443625 |
| rs900400 | 3:156798775 | rs17451107 | 3:156797609 | 0,95455 | waist circumference adjusted for bmi (joint analysis main effects and physical activity interaction) | - | GWAS_catalog | 28448500 |
| rs900400 | 3:156798775 | rs17451107 | 3:156797609 | 0,95455 | waist circumference adjusted for bmi (joint analysis main effects and smoking interaction) | - | GWAS_catalog | 28443625 |
| rs900400 | 3:156798775 | rs17451107 | 3:156797609 | 0,95455 | waist circumference adjusted for bmi in active individuals | - | GWAS_catalog | 28448500 |
| rs900400 | 3:156798775 | rs17451107 | 3:156797609 | 0,95455 | waist circumference adjusted for body mass index | - | GWAS_catalog | 25673412 |
| rs900400 | 3:156798775 | - | - | - | Waist circumference adjusted for physical activity | - | Phenoscanner | 28448500 |
| rs900400 | 3:156798775 | - | - | - | Waist circumference adjusted for smoking | - | Phenoscanner | 28443625 |
| rs900400 | 3:156798775 | - | - | - | Waist circumference adjusted for smoking in females | - | Phenoscanner | 28443625 |
| rs900400 | 3:156798775 | rs900399 | 3:156798732 | 1 | Waist circumference in female non-smokers | - | Phenoscanner | 28443625 |
| rs900400 | 3:156798775 | - | - | - | Waist circumference in non-smokers | - | Phenoscanner | 28443625 |
| rs900400 | 3:156798775 | rs900399 | 3:156798732 | 1 | Waist circumference in physically active indivdiuals | - | Phenoscanner | 28448500 |
| rs900400 | 3:156798775 | - | - | - | Waist hip ratio | - | Phenoscanner | 25673412 |
| rs900400 | 3:156798775 | - | - | - | Waist hip ratio adjusted for BMI | - | Phenoscanner | 25673412 |
| rs900400 | 3:156798775 | rs900399 | 3:156798732 | 1 | Waist hip ratio adjusted for physical activity | - | Phenoscanner | 28448500 |
| rs900400 | 3:156798775 | rs900399 | 3:156798732 | 1 | Waist hip ratio adjusted for smoking | - | Phenoscanner | 28443625 |
| rs900400 | 3:156798775 | rs900399 | 3:156798732 | 1 | Waist hip ratio in physically active indivdiuals | - | Phenoscanner | 28448500 |
| rs900400 | 3:156798775 | rs4680338 | 3:156794425 | 0,91631 | Waist-hip ratio | Metabolic | GWAS_ATLAS | 30239722 |
| rs900400 | 3:156798775 | rs17451107 | 3:156797609 | 0,95455 | waist-hip ratio | - | GWAS_catalog | 30575882 |
| rs900400 | 3:156798775 | rs17451107 | 3:156797609 | 0,95455 | waist-to-hip ratio adjusted for bmi | - | GWAS_catalog | 30575882 |
| rs900400 | 3:156798775 | rs17451107 | 3:156797609 | 0,95455 | waist-to-hip ratio adjusted for bmi (adjusted for smoking behaviour) | - | GWAS_catalog | 28443625 |
| rs900400 | 3:156798775 | rs17451107 | 3:156797609 | 0,95455 | waist-to-hip ratio adjusted for bmi (joint analysis for main effect and physical activity interaction) | - | GWAS_catalog | 28448500 |
| rs900400 | 3:156798775 | rs17451107 | 3:156797609 | 0,95455 | waist-to-hip ratio adjusted for bmi in active individuals | - | GWAS_catalog | 28448500 |
| rs900400 | 3:156798775 | rs1482852 | 3:156798294 | 0,9435 | waist-to-hip ratio adjusted for bmi x sex x age interaction (4df test) | - | GWAS_catalog | 26426971 |
| rs900400 | 3:156798775 | rs17451107 | 3:156797609 | 0,95455 | waist-to-hip ratio adjusted for body mass index | - | GWAS_catalog | 25673412 |
| rs3851366 | 3:160191374 | rs10513551 | 3:160086055 | 0,92593 | ldl cholesterol | - | GWAS_catalog | 30275531 |
| rs3851366 | 3:160191374 | rs56394279 | 3:160171092 | 0,98812 | low density lipoprotein cholesterol levels | - | GWAS_catalog | 29507422 |
| rs3851366 | 3:160191374 | rs56394279 | 3:160171092 | 0,98812 | pulse pressure | - | GWAS_catalog | 30578418 |
| rs3851366 | 3:160191374 | rs56394279 | 3:160171092 | 0,98812 | systolic blood pressure | - | GWAS_catalog | 30578418 |
| rs3851366 | 3:160191374 | rs56394279 | 3:160171092 | 0,98812 | total cholesterol levels | - | GWAS_catalog | 29507422 |
| rs2687965 | 4:39491521 | - | - | - | Height | Skeletal | GWAS_ATLAS | 31427789 |
| rs11942410 | 4:75656079 | rs4585380 | 4:75673363 | 0,97405 | chronic obstructive pulmonary disease | - | GWAS_catalog | 30804561 |
| rs11942410 | 4:75656079 | rs35926103 | 4:75675706 | 0,97405 | diverticular disease | - | GWAS_catalog | 30177863 |
| rs11942410 | 4:75656079 | rs62316310 | 4:75676529 | 0,96875 | fev1 | - | GWAS_catalog | 30804560 |
| rs11942410 | 4:75656079 | - | - | - | FEV1/FVC ratio | Respiratory | GWAS_ATLAS | 30804560 |
| rs11942410 | 4:75656079 | - | - | - | FEV2 | Respiratory | GWAS_ATLAS | 30804560 |
| rs11942410 | 4:75656079 | rs11938276 | 4:75676256 | 0,96875 | lung function (fev1/fvc) | - | GWAS_catalog | 30595370 |
| rs28663472 | 4:95948204 | rs28676957 | 4:95903741 | 0,95668 | Angina | Cardiovascular | GWAS_ATLAS | 31427789 |
| rs6839705 | 4:106144735 | rs6533183 | 4:106133184 | 0,98277 | birth weight | - | GWAS_catalog | 31043758 |
| rs6839705 | 4:106144735 | rs1391441 | 4:106128760 | 0,82763 | Birth weight | Metabolic | GWAS_ATLAS | 31427789 |
| rs6839705 | 4:106144735 | - | - | - | Birth weight | - | Phenoscanner | UKBB |
| rs6839705 | 4:106144735 | rs1391441 | 4:106128760 | 0,82763 | Birth weight of first child | Metabolic | GWAS_ATLAS | 31427789 |
| rs6839705 | 4:106144735 | - | - | - | Birth weight of first child | - | Phenoscanner | UKBB |
| rs6839705 | 4:106144735 | rs7663401 | 4:106128954 | 0,95349 | Cheese intake | Nutritional | GWAS_ATLAS | 31427789 |
| rs6839705 | 4:106144735 | rs2047409 | 4:106137033 | 0,91744 | chronic obstructive pulmonary disease | - | GWAS_catalog | 30804561 |
| rs6839705 | 4:106144735 | rs2007403 | 4:106131210 | 0,91744 | Coffee type: Ground coffee | Nutritional | GWAS_ATLAS | 31427789 |
| rs6839705 | 4:106144735 | rs7674220 | 4:106148758 | 0,95373 | cognitive ability, years of educational attainment or schizophrenia (pleiotropy) | - | GWAS_catalog | 31374203 |
| rs6839705 | 4:106144735 | rs1391441 | 4:106128760 | 0,82763 | colorectal cancer or advanced adenoma | - | GWAS_catalog | 30510241 |
| rs6839705 | 4:106144735 | rs1391441 | 4:106128760 | 0,82763 | Educational attainment | Environment | GWAS_ATLAS | 31427789 |
| rs6839705 | 4:106144735 | - | - | - | educational attainment (years of education) | - | GWAS_catalog | 27225129 |
| rs6839705 | 4:106144735 | - | - | - | Educational attainment years of education | - | Phenoscanner | 27225129 |
| rs6839705 | 4:106144735 | rs6533183 | 4:106133184 | 0,98277 | fev1 | - | GWAS_catalog | 30804560 |
| rs6839705 | 4:106144735 | rs1391441 | 4:106128760 | 0,82763 | FEV1/FVC ratio | Respiratory | GWAS_ATLAS | 30804560 |
| rs6839705 | 4:106144735 | rs1391441 | 4:106128760 | 0,82763 | FEV2 | Respiratory | GWAS_ATLAS | 30804560 |
| rs6839705 | 4:106144735 | rs6533183 | 4:106133184 | 0,98277 | First PC of the four riskly behaviours | Activities | GWAS_ATLAS | v: https://doi.org/10.1101/2 |
| rs6839705 | 4:106144735 | - | - | - | Fluid intelligence score | - | Phenoscanner | UKBB |
| rs6839705 | 4:106144735 | - | - | - | Forced expiratory volume in 1-second | - | Phenoscanner | UKBB |
| rs6839705 | 4:106144735 | - | - | - | Forced expiratory volume in 1-second, best measure | - | Phenoscanner | UKBB |
| rs6839705 | 4:106144735 | - | - | - | Forced expiratory volume in 1-second, predicted percentage | - | Phenoscanner | UKBB |
| rs6839705 | 4:106144735 | rs7663401 | 4:106128954 | 0,95349 | Forced vital capacity, best measure | - | Phenoscanner | UKBB |
| rs6839705 | 4:106144735 | rs1391438 | 4:106151843 | 0,84434 | general cognitive ability | - | GWAS_catalog | 29844566 |
| rs6839705 | 4:106144735 | rs1391438 | 4:106151843 | 0,84434 | General risk tolerance | Activities | GWAS_ATLAS | v: https://doi.org/10.1101/2 |
| rs6839705 | 4:106144735 | - | - | - | general risk tolerance (mtag) | - | GWAS_catalog | 30643258 |
| rs6839705 | 4:106144735 | rs7663401 | 4:106128954 | 0,95349 | Granulocyte count | Immunological | GWAS_ATLAS | 27863252 |
| rs6839705 | 4:106144735 | rs6533183 | 4:106133184 | 0,98277 | Granulocyte count | - | Phenoscanner | 27863252 |
| rs6839705 | 4:106144735 | rs1391441 | 4:106128760 | 0,82763 | Heel bone mineral density | Skeletal | GWAS_ATLAS | 30048462 |
| rs6839705 | 4:106144735 | rs1391441 | 4:106128760 | 0,82763 | Height | Skeletal | GWAS_ATLAS | 31427789 |
| rs6839705 | 4:106144735 | - | - | - | Height | - | Phenoscanner | UKBB |
| rs6839705 | 4:106144735 | rs1391441 | 4:106128760 | 0,82763 | household income (mtag) | - | GWAS_catalog | 31844048 |
| rs6839705 | 4:106144735 | rs1391441 | 4:106128760 | 0,82763 | Impedance measures - Trunk fat-free mass | Metabolic | GWAS_ATLAS | 31427789 |
| rs6839705 | 4:106144735 | rs1391441 | 4:106128760 | 0,82763 | Impedance measures - Trunk predicted mass | Metabolic | GWAS_ATLAS | 31427789 |
| rs6839705 | 4:106144735 | rs1391438 | 4:106151843 | 0,84434 | intelligence | - | GWAS_catalog | 29942086 |
| rs6839705 | 4:106144735 | rs1391441 | 4:106128760 | 0,82763 | Intelligence | Cognitive | GWAS_ATLAS | 31427789 |
| rs6839705 | 4:106144735 | - | - | - | intelligence (mtag) | - | GWAS_catalog | 29326435 |
| rs6839705 | 4:106144735 | - | - | - | Intelligence multi trait analysis | - | Phenoscanner | 29326435 |
| rs6839705 | 4:106144735 | rs2007403 | 4:106131210 | 0,91744 | lung function (fev1/fvc) | - | GWAS_catalog | 30595370 |
| rs6839705 | 4:106144735 | rs2047409 | 4:106137033 | 0,91744 | lung function in never smokers (low fev1 vs average fev1) | - | GWAS_catalog | 26423011 |
| rs6839705 | 4:106144735 | rs2047409 | 4:106137033 | 0,91744 | lung function in never smokers (low fev1 vs high fev1) | - | GWAS_catalog | 26423011 |
| rs6839705 | 4:106144735 | rs1391441 | 4:106128760 | 0,82763 | Lymphocyte count | Immunological | GWAS_ATLAS | 27863252 |
| rs6839705 | 4:106144735 | rs1391441 | 4:106128760 | 0,82763 | Lymphocyte percentage of white cells | Immunological | GWAS_ATLAS | 27863252 |
| rs6839705 | 4:106144735 | - | - | - | Lymphocyte percentage of white cells | - | Phenoscanner | 27863252 |
| rs6839705 | 4:106144735 | rs7663401 | 4:106128954 | 0,95349 | Myeloid white cell count | Immunological | GWAS_ATLAS | 27863252 |
| rs6839705 | 4:106144735 | rs7663401 | 4:106128954 | 0,95349 | Myeloid white cell count | - | Phenoscanner | 27863252 |
| rs6839705 | 4:106144735 | rs7663401 | 4:106128954 | 0,95349 | Neutrophil count | Immunological | GWAS_ATLAS | 27863252 |
| rs6839705 | 4:106144735 | rs6533183 | 4:106133184 | 0,98277 | Neutrophil count | - | Phenoscanner | 27863252 |
| rs6839705 | 4:106144735 | rs1391441 | 4:106128760 | 0,82763 | Neutrophil percentage of white cells | Immunological | GWAS_ATLAS | 27863252 |
| rs6839705 | 4:106144735 | - | - | - | Neutrophil percentage of white cells | - | Phenoscanner | 27863252 |
| rs6839705 | 4:106144735 | rs1391441 | 4:106128760 | 0,82763 | Number of sexual partners | Reproduction | GWAS_ATLAS | v: https://doi.org/10.1101/2 |
| rs6839705 | 4:106144735 | rs6533183 | 4:106133184 | 0,98277 | peak expiratory flow | - | GWAS_catalog | 30804560 |
| rs6839705 | 4:106144735 | rs1391441 | 4:106128760 | 0,82763 | PEF | Respiratory | GWAS_ATLAS | 30804560 |
| rs6839705 | 4:106144735 | - | - | - | Qualifications: college or university degree | - | Phenoscanner | UKBB |
| rs6839705 | 4:106144735 | rs1391441 | 4:106128760 | 0,82763 | Sitting height | Skeletal | GWAS_ATLAS | 31427789 |
| rs6839705 | 4:106144735 | - | - | - | Sitting height | - | Phenoscanner | UKBB |
| rs6839705 | 4:106144735 | rs7663401 | 4:106128954 | 0,95349 | Sum basophil neutrophil count | Immunological | GWAS_ATLAS | 27863252 |
| rs6839705 | 4:106144735 | rs6533183 | 4:106133184 | 0,98277 | Sum basophil neutrophil counts | - | Phenoscanner | 27863252 |
| rs6839705 | 4:106144735 | rs7663401 | 4:106128954 | 0,95349 | Sum neutrophil eosinophil count | Immunological | GWAS_ATLAS | 27863252 |
| rs6839705 | 4:106144735 | rs7663401 | 4:106128954 | 0,95349 | Sum neutrophil eosinophil counts | - | Phenoscanner | 27863252 |
| rs6839705 | 4:106144735 | rs1391439 | 4:106151642 | 0,93358 | tonsillectomy | - | GWAS_catalog | 28928442 |
| rs6839705 | 4:106144735 | rs6533183 | 4:106133184 | 0,98277 | Trunk fat-free mass | - | Phenoscanner | UKBB |
| rs6839705 | 4:106144735 | rs6533183 | 4:106133184 | 0,98277 | Trunk predicted mass | - | Phenoscanner | UKBB |
| rs6839705 | 4:106144735 | - | - | - | Years of educational attainment | - | Phenoscanner | 27225129 |
| rs6839705 | 4:106144735 | - | - | - | Years of educational attainment in females | - | Phenoscanner | 27225129 |
| rs2060285 | 4:124746377 | - | - | - | Bald Pattern 4 | Dermatological | GWAS_ATLAS | 31427789 |
| rs2060285 | 4:124746377 | - | - | - | Baldness | Dermatological | GWAS_ATLAS | 30573740 |
| rs2060285 | 4:124746377 | - | - | - | FEV1/FVC ratio | Respiratory | GWAS_ATLAS | 30804560 |
| rs2060285 | 4:124746377 | - | - | - | male-pattern baldness | - | GWAS_catalog | 30573740 |
| rs17824374 | 4:126924999 | rs145365164 | 4:126954652 | 0,98266 | chin dimples | - | GWAS_catalog | 27182965 |
| rs17824374 | 4:126924999 | rs12509991 | 4:126996870 | 0,84098 | response to homoharringtonine (cytotoxicity) | - | GWAS_catalog | 25628645 |
| rs1542726 | 4:145515769 | - | - | - | Arm fat-free mass left | - | Phenoscanner | UKBB |
| rs1542726 | 4:145515769 | - | - | - | Arm fat-free mass right | - | Phenoscanner | UKBB |
| rs1542726 | 4:145515769 | - | - | - | Arm predicted mass left | - | Phenoscanner | UKBB |
| rs1542726 | 4:145515769 | - | - | - | Arm predicted mass right | - | Phenoscanner | UKBB |
| rs1542726 | 4:145515769 | - | - | - | Basal metabolic rate | - | Phenoscanner | UKBB |
| rs1542726 | 4:145515769 | rs13116999 | 4:145442364 | 0,86555 | fev1 | - | GWAS_catalog | 30804560 |
| rs1542726 | 4:145515769 | rs7681384 | 4:145437014 | 0,83657 | FEV1/FCV | Respiratory | GWAS_ATLAS | 26635072 |
| rs1542726 | 4:145515769 | rs7681384 | 4:145437014 | 0,83657 | FEV1/FVC ratio | Respiratory | GWAS_ATLAS | 30804560 |
| rs1542726 | 4:145515769 | rs7681384 | 4:145437014 | 0,83657 | FEV2 | Respiratory | GWAS_ATLAS | 30804560 |
| rs1542726 | 4:145515769 | - | - | - | Forced expiratory volume in 1-second | - | Phenoscanner | UKBB |
| rs1542726 | 4:145515769 | - | - | - | Forced expiratory volume in 1-second, best measure | - | Phenoscanner | UKBB |
| rs1542726 | 4:145515769 | - | - | - | Forced expiratory volume in 1-second, predicted percentage | - | Phenoscanner | UKBB |
| rs1542726 | 4:145515769 | - | - | - | Forced vital capacity | - | Phenoscanner | UKBB |
| rs1542726 | 4:145515769 | - | - | - | Forced vital capacity, best measure | - | Phenoscanner | UKBB |
| rs1542726 | 4:145515769 | rs7681384 | 4:145437014 | 0,83657 | FVC | Respiratory | GWAS_ATLAS | 30804560 |
| rs1542726 | 4:145515769 | rs7681384 | 4:145437014 | 0,83657 | Height | Skeletal | GWAS_ATLAS | 25282103 |
| rs1542726 | 4:145515769 | - | - | - | Height | - | Phenoscanner | 25282103 |
| rs1542726 | 4:145515769 | rs7681384 | 4:145437014 | 0,83657 | Hip circumference | Metabolic | GWAS_ATLAS | 31427789 |
| rs1542726 | 4:145515769 | - | - | - | Hip circumference | - | Phenoscanner | UKBB |
| rs1542726 | 4:145515769 | rs7681384 | 4:145437014 | 0,83657 | Impedance measures - Arm fat-free mass (left) | Metabolic | GWAS_ATLAS | 31427789 |
| rs1542726 | 4:145515769 | rs7681384 | 4:145437014 | 0,83657 | Impedance measures - Arm fat-free mass (right) | Metabolic | GWAS_ATLAS | 31427789 |
| rs1542726 | 4:145515769 | rs7681384 | 4:145437014 | 0,83657 | Impedance measures - Arm predicted mass (left) | Metabolic | GWAS_ATLAS | 31427789 |
| rs1542726 | 4:145515769 | rs7681384 | 4:145437014 | 0,83657 | Impedance measures - Arm predicted mass (right) | Metabolic | GWAS_ATLAS | 31427789 |
| rs1542726 | 4:145515769 | rs7681384 | 4:145437014 | 0,83657 | Impedance measures - Basal metabolic rate | Metabolic | GWAS_ATLAS | 31427789 |
| rs1542726 | 4:145515769 | rs7681384 | 4:145437014 | 0,83657 | Impedance measures - Impedance of whole body | Metabolic | GWAS_ATLAS | 31427789 |
| rs1542726 | 4:145515769 | rs7681384 | 4:145437014 | 0,83657 | Impedance measures - Leg fat-free mass (left) | Metabolic | GWAS_ATLAS | 31427789 |
| rs1542726 | 4:145515769 | rs7681384 | 4:145437014 | 0,83657 | Impedance measures - Leg fat-free mass (right) | Metabolic | GWAS_ATLAS | 31427789 |
| rs1542726 | 4:145515769 | rs7681384 | 4:145437014 | 0,83657 | Impedance measures - Leg predicted mass (left) | Metabolic | GWAS_ATLAS | 31427789 |
| rs1542726 | 4:145515769 | rs7681384 | 4:145437014 | 0,83657 | Impedance measures - Leg predicted mass (right) | Metabolic | GWAS_ATLAS | 31427789 |
| rs1542726 | 4:145515769 | rs7681384 | 4:145437014 | 0,83657 | Impedance measures - Trunk fat-free mass | Metabolic | GWAS_ATLAS | 31427789 |
| rs1542726 | 4:145515769 | rs7681384 | 4:145437014 | 0,83657 | Impedance measures - Trunk predicted mass | Metabolic | GWAS_ATLAS | 31427789 |
| rs1542726 | 4:145515769 | rs7681384 | 4:145437014 | 0,83657 | Impedance measures - Weight | Metabolic | GWAS_ATLAS | 31427789 |
| rs1542726 | 4:145515769 | rs7681384 | 4:145437014 | 0,83657 | Impedance measures - Whole body fat-free mass | Metabolic | GWAS_ATLAS | 31427789 |
| rs1542726 | 4:145515769 | rs7681384 | 4:145437014 | 0,83657 | Impedance measures - Whole body water mass | Metabolic | GWAS_ATLAS | 31427789 |
| rs1542726 | 4:145515769 | - | - | - | Leg fat-free mass left | - | Phenoscanner | UKBB |

| | | | | | | | | |
|---|---|---|---|---|---|---|---|---|
| rs1542726 | 4:145515769 | - | - | - | Leg fat-free mass right | - | Phenoscanner | UKBB |
| rs1542726 | 4:145515769 | - | - | - | Leg predicted mass left | - | Phenoscanner | UKBB |
| rs1542726 | 4:145515769 | - | - | - | Leg predicted mass right | - | Phenoscanner | UKBB |
| rs1542726 | 4:145515769 | rs13116999 | 4:145442364 | 0,86555 | lung function (fev1/fvc) | - | GWAS_catalog | 30804560 |
| rs1542726 | 4:145515769 | rs13116999 | 4:145442364 | 0,86555 | lung function (fvc) | - | GWAS_catalog | 30804560 |
| rs1542726 | 4:145515769 | rs6828982 | 4:145470604 | 0,93633 | lung function (low fev1 vs high fev1) | - | GWAS_catalog | 26423011 |
| rs1542726 | 4:145515769 | rs13107665 | 4:145472644 | 0,93633 | lung function in never smokers (low fev1 vs high fev1) | - | GWAS_catalog | 26423011 |
| rs1542726 | 4:145515769 | rs13116999 | 4:145442364 | 0,86555 | peak expiratory flow | - | GWAS_catalog | 30804560 |
| rs1542726 | 4:145515769 | - | - | - | Peak expiratory flow | - | Phenoscanner | UKBB |
| rs1542726 | 4:145515769 | rs7681384 | 4:145437014 | 0,83657 | PEF | Respiratory | GWAS_ATLAS | 30804560 |
| rs1542726 | 4:145515769 | rs7681384 | 4:145437014 | 0,83657 | post bronchodilator fev1 | - | GWAS_catalog | 26634245 |
| rs1542726 | 4:145515769 | rs6537298 | 4:145506871 | 0,97974 | Post bronchodilator FEV1 | - | Phenoscanner | 26634245 |
| rs1542726 | 4:145515769 | rs7681384 | 4:145437014 | 0,83657 | post bronchodilator fev1/fvc ratio | - | GWAS_catalog | 26634245 |
| rs1542726 | 4:145515769 | - | - | - | Post bronchodilator FEV1FVC ratio | - | Phenoscanner | 26634245 |
| rs1542726 | 4:145515769 | rs11407396 | 4:145516737 | 0,89115 | Right fusiform | Neurological | GWAS_ATLAS | 31676860 |
| rs1542726 | 4:145515769 | rs7681384 | 4:145437014 | 0,83657 | Sitting height | Skeletal | GWAS_ATLAS | 31427789 |
| rs1542726 | 4:145515769 | - | - | - | Sitting height | - | Phenoscanner | UKBB |
| rs1542726 | 4:145515769 | - | - | - | Trunk fat-free mass | - | Phenoscanner | UKBB |
| rs1542726 | 4:145515769 | - | - | - | Trunk predicted mass | - | Phenoscanner | UKBB |
| rs1542726 | 4:145515769 | rs7681384 | 4:145437014 | 0,83657 | Waist-hip ratio | Metabolic | GWAS_ATLAS | 30239722 |
| rs1542726 | 4:145515769 | rs7681384 | 4:145437014 | 0,83657 | Weight | Metabolic | GWAS_ATLAS | 31427789 |
| rs1542726 | 4:145515769 | rs6537297 | 4:145502029 | 0,95588 | Weight | - | Phenoscanner | UKBB |
| rs1542726 | 4:145515769 | - | - | - | Whole body fat-free mass | - | Phenoscanner | UKBB |
| rs1542726 | 4:145515769 | - | - | - | Whole body water mass | - | Phenoscanner | UKBB |
| rs72707023 | 5:667620 | rs77015126 | 5:670909 | 0,91453 | Estimated glomerular filtration rate | Metabolic | GWAS_ATLAS | 31152163 |
| rs72707023 | 5:667620 | rs11739847 | 5:609661 | 0,92974 | fev1 | - | GWAS_catalog | 30804560 |
| rs72707023 | 5:667620 | rs72704802 | 5:554211 | 0,88831 | FEV1/FVC ratio | Respiratory | GWAS_ATLAS | 30804560 |
| rs72707023 | 5:667620 | rs72704791 | 5:547896 | 0,84302 | FEV2 | Respiratory | GWAS_ATLAS | 30804560 |
| rs72707023 | 5:667620 | rs11739847 | 5:609661 | 0,92974 | lung function (fev1/fvc) | - | GWAS_catalog | 30804560 |
| rs72707023 | 5:667620 | rs11739847 | 5:609661 | 0,92974 | lung function (fvc) | - | GWAS_catalog | 30804560 |
| rs72707023 | 5:667620 | - | - | - | Potassium in urine | - | Phenoscanner | UKBB |
| rs72707023 | 5:667620 | rs74553517 | 5:665671 | 0,92373 | reaction time | - | GWAS_catalog | 29844566 |
| rs72707023 | 5:667620 | rs4957048 | 5:583442 | 0,84852 | ulcerative colitis | - | GWAS_catalog | 20228799 |
| rs72707023 | 5:667620 | rs72704791 | 5:547896 | 0,84302 | Ulcerative colitis | Gastrointestinal | GWAS_ATLAS | 28067908 |
| rs61026653 | 5:15757570 | rs13170409 | 5:15764650 | 0,98293 | Systemic Lupus Erythematosus | Skeletal | GWAS_ATLAS | 29848360 |
| rs62368263 | 5:50726027 | rs62368272 | 5:50754547 | 0,84975 | brain region volumes | - | GWAS_catalog | 31676860 |
| rs62368263 | 5:50726027 | rs113474156 | 5:50658924 | 0,94156 | Heel bone mineral density | Skeletal | GWAS_ATLAS | 30048462 |
| rs62368263 | 5:50726027 | rs62366930 | 5:50699314 | 0,85796 | Left putamen | Neurological | GWAS_ATLAS | 31676860 |
| rs4485884 | 5:51108645 | - | - | - | Body fat percentage | - | Phenoscanner | UKBB |
| rs4485884 | 5:51108645 | - | - | - | PEF | Respiratory | GWAS_ATLAS | 30804560 |
| rs4485884 | 5:51108645 | - | - | - | Trunk fat percentage | - | Phenoscanner | UKBB |
| rs6867042 | 5:93555761 | rs17376456 | 5:93557702 | 0,9915 | diabetic retinopathy | - | GWAS_catalog | 21310492 |
| rs6867042 | 5:93555761 | rs9314093 | 5:93333347 | 0,83962 | Hearing difficulty/problems | Ear, Nose, Throat | GWAS_ATLAS | 31427789 |
| rs4959352 | 6:624922 | rs4960127 | 6:624892 | 0,95119 | Hair colour: Black | Dermatological | GWAS_ATLAS | 31427789 |
| rs722587 | 6:1775714 | rs3778519 | 6:1771512 | 0,92708 | FEV1/FVC ratio | Respiratory | GWAS_ATLAS | 30804560 |
| rs722587 | 6:1775714 | rs3778519 | 6:1771512 | 0,92708 | FEV2 | Respiratory | GWAS_ATLAS | 30804560 |
| rs722587 | 6:1775714 | rs3778519 | 6:1771512 | 0,92708 | height | - | GWAS_catalog | 30595370 |
| rs722587 | 6:1775714 | rs3778519 | 6:1771512 | 0,92708 | Height | Skeletal | GWAS_ATLAS | 31427789 |
| rs722587 | 6:1775714 | - | - | - | Height | - | Phenoscanner | UKBB |
| rs722587 | 6:1775714 | rs722585 | 6:1775863 | 1 | Hip circumference | Metabolic | GWAS_ATLAS | 25673412 |
| rs722587 | 6:1775714 | rs722585 | 6:1775863 | 1 | hip circumference adjusted for bmi | - | GWAS_catalog | 25673412 |
| rs722587 | 6:1775714 | rs722585 | 6:1775863 | 1 | intraocular pressure | - | GWAS_catalog | 31959993 |
| rs722587 | 6:1775714 | rs3778519 | 6:1771512 | 0,92708 | lung function (fev1/fvc) | - | GWAS_catalog | 30595370 |
| rs722587 | 6:1775714 | - | - | - | Peak expiratory flow | - | Phenoscanner | UKBB |
| rs722587 | 6:1775714 | rs3778519 | 6:1771512 | 0,92708 | PEF | Respiratory | GWAS_ATLAS | 30804560 |
| rs1156533 | 6:30065149 | rs9280888 | 6:30064849 | 1 | Age-related Macular Degeneration | Ophthalmological | GWAS_ATLAS | 26691988 |
| rs1156533 | 6:30065149 | - | - | - | Age-related macular degeneration | - | Phenoscanner | 26691988 |
| rs1156533 | 6:30065149 | rs916570 | 6:30066031 | 0,80425 | Celiac disease | Gastrointestinal | GWAS_ATLAS | 22057235 |
| rs1156533 | 6:30065149 | - | - | - | Comparative body size at age 10 | - | Phenoscanner | UKBB |
| rs1156533 | 6:30065149 | rs9280888 | 6:30064849 | 1 | Eosinophil count | Immunological | GWAS_ATLAS | 27863252 |
| rs1156533 | 6:30065149 | - | - | - | Eosinophil count | - | Phenoscanner | 27863252 |
| rs1156533 | 6:30065149 | rs916570 | 6:30066031 | 0,80425 | eosinophil counts | - | GWAS_catalog | 31272903 |
| rs1156533 | 6:30065149 | rs9280888 | 6:30064849 | 1 | Eosinophil percentage of granulocytes | Immunological | GWAS_ATLAS | 27863252 |
| rs1156533 | 6:30065149 | - | - | - | Eosinophil percentage of granulocytes | - | Phenoscanner | 27863252 |
| rs1156533 | 6:30065149 | rs9280888 | 6:30064849 | 1 | Eosinophil percentage of white cells | Immunological | GWAS_ATLAS | 27863252 |
| rs1156533 | 6:30065149 | - | - | - | Eosinophil percentage of white cells | - | Phenoscanner | 27863252 |
| rs1156533 | 6:30065149 | - | - | - | FEV1/FVC ratio | Respiratory | GWAS_ATLAS | 30804560 |
| rs1156533 | 6:30065149 | - | - | - | FEV2 | Respiratory | GWAS_ATLAS | 30804560 |
| rs1156533 | 6:30065149 | - | - | - | Forced expiratory volume in 1-second, predicted percentage | - | Phenoscanner | UKBB |
| rs1156533 | 6:30065149 | - | - | - | FVC | Respiratory | GWAS_ATLAS | 30804560 |
| rs1156533 | 6:30065149 | rs9280888 | 6:30064849 | 1 | Granulocyte count | Immunological | GWAS_ATLAS | 27863252 |
| rs1156533 | 6:30065149 | - | - | - | Granulocyte count | - | Phenoscanner | 27863252 |
| rs1156533 | 6:30065149 | rs916570 | 6:30066031 | 0,80425 | Heel bone mineral density | Skeletal | GWAS_ATLAS | 30598549 |
| rs1156533 | 6:30065149 | rs916570 | 6:30066031 | 0,80425 | Hematocrit | Immunological | GWAS_ATLAS | 27863252 |
| rs1156533 | 6:30065149 | rs9280888 | 6:30064849 | 1 | Hemoglobin concentration | Immunological | GWAS_ATLAS | 27863252 |
| rs1156533 | 6:30065149 | rs916570 | 6:30066031 | 0,80425 | Hemoglobin concentration | - | Phenoscanner | 27863252 |
| rs1156533 | 6:30065149 | rs916570 | 6:30066031 | 0,80425 | Hip circumference | - | Phenoscanner | UKBB |
| rs1156533 | 6:30065149 | rs916570 | 6:30066031 | 0,80425 | Idiopathic membranous nephropathy | - | Phenoscanner | 21323541 |
| rs1156533 | 6:30065149 | - | - | - | IgA deficiency | - | Phenoscanner | 27723758 |
| rs1156533 | 6:30065149 | rs9280888 | 6:30064849 | 1 | Immunoglobulin A deficiency | Immunological | GWAS_ATLAS | 27723758 |
| rs1156533 | 6:30065149 | - | - | - | Intestinal malabsorption | - | Phenoscanner | UKBB |
| rs1156533 | 6:30065149 | rs916570 | 6:30066031 | 0,80425 | Leg fat mass left | - | Phenoscanner | UKBB |
| rs1156533 | 6:30065149 | rs916570 | 6:30066031 | 0,80425 | Leg fat mass right | - | Phenoscanner | UKBB |
| rs1156533 | 6:30065149 | rs9280888 | 6:30064849 | 1 | Lymphocyte count | Immunological | GWAS_ATLAS | 27863252 |
| rs1156533 | 6:30065149 | - | - | - | Lymphocyte count | - | Phenoscanner | 27863252 |
| rs1156533 | 6:30065149 | rs9280888 | 6:30064849 | 1 | Monocyte count | Immunological | GWAS_ATLAS | 27863252 |
| rs1156533 | 6:30065149 | - | - | - | Monocyte count | - | Phenoscanner | 27863252 |
| rs1156533 | 6:30065149 | - | - | - | Mouth or teeth dental problems: dentures | - | Phenoscanner | UKBB |
| rs1156533 | 6:30065149 | rs9280888 | 6:30064849 | 1 | Myeloid white cell count | Immunological | GWAS_ATLAS | 27863252 |
| rs1156533 | 6:30065149 | - | - | - | Myeloid white cell count | - | Phenoscanner | 27863252 |
| rs1156533 | 6:30065149 | rs9280888 | 6:30064849 | 1 | Neutrophil count | Immunological | GWAS_ATLAS | 27863252 |
| rs1156533 | 6:30065149 | - | - | - | Neutrophil count | - | Phenoscanner | 27863252 |
| rs1156533 | 6:30065149 | rs9280888 | 6:30064849 | 1 | Neutrophil percentage of granulocytes | Immunological | GWAS_ATLAS | 27863252 |
| rs1156533 | 6:30065149 | - | - | - | Neutrophil percentage of granulocytes | - | Phenoscanner | 27863252 |
| rs1156533 | 6:30065149 | - | - | - | Peak expiratory flow | - | Phenoscanner | UKBB |
| rs1156533 | 6:30065149 | - | - | - | PEF | Respiratory | GWAS_ATLAS | 30804560 |
| rs1156533 | 6:30065149 | - | - | - | Plateletcrit | Immunological | GWAS_ATLAS | 27863252 |
| rs1156533 | 6:30065149 | - | - | - | Plateletcrit | - | Phenoscanner | 27863252 |
| rs1156533 | 6:30065149 | rs916570 | 6:30066031 | 0,80425 | Primary biliary cholangitis | - | Phenoscanner | 23000144 |
| rs1156533 | 6:30065149 | rs9280888 | 6:30064849 | 1 | Primary sclerosing cholangitis | Gastrointestinal | GWAS_ATLAS | 27992413 |
| rs1156533 | 6:30065149 | - | - | - | Primary sclerosing cholangitis | - | Phenoscanner | 27992413 |
| rs1156533 | 6:30065149 | rs9280888 | 6:30064849 | 1 | Red cell distribution width | Immunological | GWAS_ATLAS | 27863252 |
| rs1156533 | 6:30065149 | - | - | - | Red cell distribution width | - | Phenoscanner | 27863252 |
| rs1156533 | 6:30065149 | - | - | - | Rheumatoid Arthritis | Connective Tissue | GWAS_ATLAS | 24390342 |
| rs1156533 | 6:30065149 | - | - | - | Rheumatoid arthritis | - | Phenoscanner | 24390342 |
| rs1156533 | 6:30065149 | rs9280888 | 6:30064849 | 1 | Schizophrenia | Psychiatric | GWAS_ATLAS | 29906448 |
| rs1156533 | 6:30065149 | - | - | - | Self-reported ankylosing spondylitis | - | Phenoscanner | UKBB |
| rs1156533 | 6:30065149 | - | - | - | Self-reported hyperthyroidism or thyrotoxicosis | - | Phenoscanner | UKBB |
| rs1156533 | 6:30065149 | - | - | - | Self-reported malabsorption or coeliac disease | - | Phenoscanner | UKBB |
| rs1156533 | 6:30065149 | - | - | - | Self-reported psoriasis | - | Phenoscanner | UKBB |
| rs1156533 | 6:30065149 | rs9280888 | 6:30064849 | 1 | Sum basophil neutrophil count | Immunological | GWAS_ATLAS | 27863252 |
| rs1156533 | 6:30065149 | - | - | - | Sum basophil neutrophil counts | - | Phenoscanner | 27863252 |
| rs1156533 | 6:30065149 | rs9280888 | 6:30064849 | 1 | Sum eosinophil basophil count | Immunological | GWAS_ATLAS | 27863252 |
| rs1156533 | 6:30065149 | - | - | - | Sum eosinophil basophil counts | - | Phenoscanner | 27863252 |
| rs1156533 | 6:30065149 | rs9280888 | 6:30064849 | 1 | Sum neutrophil eosinophil count | Immunological | GWAS_ATLAS | 27863252 |
| rs1156533 | 6:30065149 | - | - | - | Sum neutrophil eosinophil counts | - | Phenoscanner | 27863252 |
| rs1156533 | 6:30065149 | rs9280888 | 6:30064849 | 1 | Systemic Lupus Erythematosus | Skeletal | GWAS_ATLAS | 29848360 |
| rs1156533 | 6:30065149 | rs9280888 | 6:30064849 | 1 | Vitiligo | Dermatological | GWAS_ATLAS | 27723757 |
| rs1156533 | 6:30065149 | rs916570 | 6:30066031 | 0,80425 | Weight | - | Phenoscanner | UKBB |
| rs1156533 | 6:30065149 | rs9280888 | 6:30064849 | 1 | White blood cell count | Immunological | GWAS_ATLAS | 27863252 |
| rs1156533 | 6:30065149 | - | - | - | White blood cell count | - | Phenoscanner | 27863252 |
| rs2180811 | 6:117780158 | rs9387478 | 6:117786180 | 1 | Adenocarcinoma | Neoplasms | GWAS_ATLAS | 23143601 |
| rs2180811 | 6:117780158 | rs9387478 | 6:117786180 | 1 | lung adenocarcinoma | - | GWAS_catalog | 31326317 |
| rs2180811 | 6:117780158 | rs9387478 | 6:117786180 | 1 | lung cancer | - | GWAS_catalog | 23143601 |
| rs2180811 | 6:117780158 | rs9387478 | 6:117786180 | 1 | Lung cancer | - | Phenoscanner | 23143601 |
| rs2180811 | 6:117780158 | rs9387478 | 6:117786180 | 1 | Lung cancer adenocarcinoma in never smoking women | - | Phenoscanner | 23143601 |
| rs2180811 | 6:117780158 | rs9387478 | 6:117786180 | 1 | Lung cancer in never smoking women | - | Phenoscanner | 23143601 |
| rs2180811 | 6:117780158 | rs4946256 | 6:117781276 | 1 | lung function (fev1/fvc) | - | GWAS_catalog | 30595370 |
| rs2327426 | 6:134202690 | rs1969783 | 6:134159399 | 0,8459 | Agents acting on the renin-angiotensin system | Environmental | GWAS_ATLAS | 31015401 |
| rs2327426 | 6:134202690 | rs1969783 | 6:134159399 | 0,8459 | Angina | Cardiovascular | GWAS_ATLAS | 31427789 |
| rs2327426 | 6:134202690 | rs1969783 | 6:134159399 | 0,8459 | Beta blocking agents | Environmental | GWAS_ATLAS | 31015401 |
| rs2327426 | 6:134202690 | rs1969783 | 6:134159399 | 0,8459 | Blood pressure medication | Activities | GWAS_ATLAS | 31427789 |
| rs2327426 | 6:134202690 | rs1969783 | 6:134159399 | 0,8459 | Calcium channel blockers | Environmental | GWAS_ATLAS | 31015401 |
| rs2327426 | 6:134202690 | rs2105092 | 6:134184972 | 0,93553 | cardiovascular disease | - | GWAS_catalog | 30595370 |
| rs2327426 | 6:134202690 | rs1969783 | 6:134159399 | 0,8459 | Chronic ischemic heart disease | Cardiovascular | GWAS_ATLAS | 31427789 |
| rs2327426 | 6:134202690 | rs1969783 | 6:134159399 | 0,8459 | Coronary artery disease | Cardiovascular | GWAS_ATLAS | 26343387 |
| rs2327426 | 6:134202690 | - | - | - | Coronary artery disease | - | Phenoscanner | 26343387 |
| rs2327426 | 6:134202690 | rs1966248 | 6:134159622 | 0,85021 | coronary artery disease | - | GWAS_catalog | 29212778 |
| rs2327426 | 6:134202690 | rs12202017 | 6:134173151 | 0,9125 | coronary artery disease (myocardial infarction, percutaneous transluminal coronary angioplasty, coronary artery bypass grafting, angina or chromic ischemic heart disease) | - | GWAS_catalog | 28714975 |
| rs2327426 | 6:134202690 | rs12524865 | 6:134196674 | 0,93553 | coronary heart disease | - | GWAS_catalog | 22751097 |
| rs2327426 | 6:134202690 | rs1969783 | 6:134159399 | 0,8459 | Diuretics | Environmental | GWAS_ATLAS | 31015401 |
| rs2327426 | 6:134202690 | rs1969783 | 6:134159399 | 0,8459 | Essential (primary) hypertension | Cardiovascular | GWAS_ATLAS | 31427789 |
| rs2327426 | 6:134202690 | rs1969783 | 6:134159399 | 0,8459 | High blood pressure | Cardiovascular | GWAS_ATLAS | 31427789 |
| rs2327426 | 6:134202690 | rs1969783 | 6:134159399 | 0,8459 | Hypertension | Cardiovascular | GWAS_ATLAS | 31427789 |
| rs2327426 | 6:134202690 | rs2105092 | 6:134184972 | 0,93553 | medication use (agents acting on the renin-angiotensin system) | - | GWAS_catalog | 31015401 |
| rs2327426 | 6:134202690 | rs2105092 | 6:134184972 | 0,93553 | medication use (beta blocking agents) | - | GWAS_catalog | 31015401 |

| | | | | | | | | |
|---|---|---|---|---|---|---|---|---|
| rs2327426 | 6:134202690 | - | - | - | Myocardial infarction | - | Phenoscanner | 26343387 |
| rs2327426 | 6:134202690 | rs1969783 | 6:134159399 | 0,8459 | PEF | Respiratory | GWAS_ATLAS | 30804560 |
| rs2327426 | 6:134202690 | rs1969783 | 6:134159399 | 0,8459 | Pulse rate | Cardiovascular | GWAS_ATLAS | 31427789 |
| rs2327426 | 6:134202690 | - | - | - | Pulse rate | - | Phenoscanner | UKBB |
| rs2327426 | 6:134202690 | rs1969783 | 6:134159399 | 0,8459 | Resting heart rate | Cardiovascular | GWAS_ATLAS | 30940143 |
| rs2327426 | 6:134202690 | rs2105092 | 6:134184972 | 0,93553 | Self-reported hypertension | - | Phenoscanner | UKBB |
| rs2327426 | 6:134202690 | rs2327429 | 6:134209837 | 0,88576 | systolic blood pressure | - | GWAS_catalog | 30595370 |
| rs2327426 | 6:134202690 | rs2105092 | 6:134184972 | 0,93553 | Vascular or heart problems diagnosed by doctor: high blood pressure | - | Phenoscanner | UKBB |
| rs2327426 | 6:134202690 | - | - | - | Vascular or heart problems diagnosed by doctor: none of the above | - | Phenoscanner | UKBB |
| rs3020338 | 6:152034062 | rs851977 | 6:152029608 | 0,85511 | birth length (mtag) | - | GWAS_catalog | 31681408 |
| rs3020338 | 6:152034062 | rs1101081 | 6:152032917 | 0,86816 | birth weight | - | GWAS_catalog | 27680694 |
| rs3020338 | 6:152034062 | rs851981 | 6:152027074 | 0,94751 | Birth weight | Metabolic | GWAS_ATLAS | 31427789 |
| rs3020338 | 6:152034062 | - | - | - | Birth weight | - | Phenoscanner | UKBB |
| rs3020338 | 6:152034062 | rs10872678 | 6:152039964 | 0,86377 | birth weight (mtag) | - | GWAS_catalog | 31681408 |
| rs3020338 | 6:152034062 | rs851981 | 6:152027074 | 0,94751 | Bone mineral density | Skeletal | GWAS_ATLAS | 29304378 |
| rs3020338 | 6:152034062 | rs851980 | 6:152027955 | 0,95797 | bone mineral density (spine) | - | GWAS_catalog | 30172743 |
| rs3020338 | 6:152034062 | rs3020340 | 6:152043290 | 0,85511 | bone mineral density (ward's triangle area) | - | GWAS_catalog | 27397699 |
| rs3020338 | 6:152034062 | rs2206949 | 6:152037556 | 0,86377 | endometriosis | - | GWAS_catalog | 28537267 |
| rs3020338 | 6:152034062 | rs851981 | 6:152027074 | 0,94751 | Heel bone mineral density | Skeletal | GWAS_ATLAS | 30048462 |
| rs3020338 | 6:152034062 | - | - | - | Heel bone mineral density | - | Phenoscanner | UKBB |
| rs3020338 | 6:152034062 | - | - | - | Heel bone mineral density left | - | Phenoscanner | UKBB |
| rs3020338 | 6:152034062 | - | - | - | Heel bone mineral density right | - | Phenoscanner | UKBB |
| rs3020338 | 6:152034062 | rs7772579 | 6:152042502 | 0,85511 | offspring birth weight | - | GWAS_catalog | 31043758 |
| rs3020338 | 6:152034062 | rs851978 | 6:152029556 | 0,86377 | Waist-hip ratio | Metabolic | GWAS_ATLAS | 30239722 |
| rs9322356 | 6:152408659 | rs6941835 | 6:152356270 | 1 | Height | Skeletal | GWAS_ATLAS | 30124842 |
| rs9322356 | 6:152408659 | rs6941835 | 6:152356270 | 1 | Sitting height | Skeletal | GWAS_ATLAS | 31427789 |
| rs9322356 | 6:152408659 | - | - | - | Sitting height | - | Phenoscanner | UKBB |
| rs9322356 | 6:152408659 | rs6941835 | 6:152356270 | 1 | Systolic Blood Pressure | Cardiovascular | GWAS_ATLAS | 31427789 |
| rs3253 | 6:169616112 | rs6605526 | 6:169604100 | 0,95919 | FEV2 | Respiratory | GWAS_ATLAS | 30804560 |
| rs3253 | 6:169616112 | - | - | - | Forced expiratory volume in 1-second | - | Phenoscanner | UKBB |
| rs3253 | 6:169616112 | - | - | - | Forced expiratory volume in 1-second, best measure | - | Phenoscanner | UKBB |
| rs3253 | 6:169616112 | - | - | - | Forced vital capacity | - | Phenoscanner | UKBB |
| rs3253 | 6:169616112 | - | - | - | Forced vital capacity, best measure | - | Phenoscanner | UKBB |
| rs3253 | 6:169616112 | rs6605526 | 6:169604100 | 0,95919 | FVC | Respiratory | GWAS_ATLAS | 30804560 |
| rs3253 | 6:169616112 | - | - | - | lung function (fvc) | - | GWAS_catalog | 30595370 |
| rs6462976 | 7:40396300 | rs2329772 | 7:40476625 | 0,88802 | moyamoya disease | - | GWAS_catalog | 29273593 |
| rs6462976 | 7:40396300 | rs17171704 | 7:40421488 | 0,90762 | Pulse pressure | Cardiovascular | GWAS_ATLAS | 29403010 |
| rs4556017 | 7:100632790 | rs7797740 | 7:100618993 | 0,92379 | Diastolic Blood Pressure | Cardiovascular | GWAS_ATLAS | 31427789 |
| rs4556017 | 7:100632790 | - | - | - | Diastolic blood pressure | - | Phenoscanner | UKBB |
| rs4556017 | 7:100632790 | rs7797740 | 7:100618993 | 0,92379 | Heart rate recovery at 10 secnds | Cardiovascular | GWAS_ATLAS | 29497042 |
| rs4556017 | 7:100632790 | rs7797740 | 7:100618993 | 0,92379 | Heart rate recovery at 20 secnds | Cardiovascular | GWAS_ATLAS | 29497042 |
| rs4556017 | 7:100632790 | rs7797740 | 7:100618993 | 0,92379 | Heart rate recovery at 30 secnds | Cardiovascular | GWAS_ATLAS | 29497042 |
| rs4556017 | 7:100632790 | rs7797740 | 7:100618993 | 0,92379 | Heart rate recovery at 40 secnds | Cardiovascular | GWAS_ATLAS | 29497042 |
| rs4556017 | 7:100632790 | rs7797740 | 7:100618993 | 0,92379 | Heart rate recovery at 50 secnds | Cardiovascular | GWAS_ATLAS | 29497042 |
| rs4556017 | 7:100632790 | rs7797740 | 7:100618993 | 0,92379 | Heel bone mineral density | Skeletal | GWAS_ATLAS | 30048462 |
| rs4556017 | 7:100632790 | rs7797740 | 7:100618993 | 0,92379 | Pulse rate | Cardiovascular | GWAS_ATLAS | 31427789 |
| rs4556017 | 7:100632790 | - | - | - | Pulse rate | - | Phenoscanner | UKBB |
| rs4556017 | 7:100632790 | rs7797740 | 7:100618993 | 0,92379 | Resting heart rate | Cardiovascular | GWAS_ATLAS | 30940143 |
| rs7778418 | 7:102429704 | rs2411046 | 7:102419304 | 0,9908 | Chronotype | Psychiatric | GWAS_ATLAS | 31427789 |
| rs7778418 | 7:102429704 | rs6949391 | 7:102446863 | 0,95006 | diverticular disease | - | GWAS_catalog | 30177863 |
| rs7778418 | 7:102429704 | rs139389026 | 7:102412361 | 0,99082 | Diverticular disease | Gastrointestinal | GWAS_ATLAS | 30661054 |
| rs7778418 | 7:102429704 | rs10953374 | 7:102442173 | 0,95006 | Diverticular disease of intestine | Gastrointestinal | GWAS_ATLAS | 31427789 |
| rs7778418 | 7:102429704 | - | - | - | Morning or evening person | - | Phenoscanner | UKBB |
| rs7778418 | 7:102429704 | rs6465876 | 7:102497569 | 0,86312 | morning person | - | GWAS_catalog | 30696823 |
| rs7778418 | 7:102429704 | rs3972456 | 7:102436907 | 0,98163 | morning vs. evening chronotype | - | GWAS_catalog | 26835600 |
| rs806169 | 7:127300668 | rs806188 | 7:127268806 | 0,95207 | Anxious | Psychiatric | GWAS_ATLAS | 31427789 |
| rs806169 | 7:127300668 | rs806174 | 7:127291072 | 0,99116 | Birth weight | Metabolic | GWAS_ATLAS | 31427789 |
| rs806169 | 7:127300668 | rs806166 | 7:127309934 | 0,99116 | feeling worry | - | GWAS_catalog | 29500382 |
| rs806169 | 7:127300668 | rs712707 | 7:127322614 | 0,9824 | general risk tolerance (mtag) | - | GWAS_catalog | 30643258 |
| rs806169 | 7:127300668 | - | - | - | Worrier or anxious feelings | - | Phenoscanner | UKBB |
| rs13271626 | 8:22467760 | rs3064 | 8:22451688 | 0,90905 | Estimated glomerular filtration rate | Metabolic | GWAS_ATLAS | 31152163 |
| rs13271626 | 8:22467760 | rs11778693 | 8:22462852 | 0,96489 | exploratory eye movement dysfunction in schizophrenia (cognitive search score) | - | GWAS_catalog | 26242244 |
| rs13271626 | 8:22467760 | rs11778693 | 8:22462852 | 0,96489 | exploratory eye movement dysfunction in schizophrenia (responsive search score) | - | GWAS_catalog | 26242244 |
| rs13271626 | 8:22467760 | rs1545837 | 8:22477807 | 0,83145 | FVC | Respiratory | GWAS_ATLAS | 30804560 |
| rs13271626 | 8:22467760 | rs1545837 | 8:22477807 | 0,83145 | lung function (fvc) | - | GWAS_catalog | 30595370 |
| rs13271626 | 8:22467760 | rs2280104 | 8:22525980 | 0,81758 | parkinson's disease | - | GWAS_catalog | 28892059 |
| rs13271626 | 8:22467760 | rs2280104 | 8:22525980 | 0,81758 | parkinson's disease or first degree relation to individual with parkinson's disease | - | GWAS_catalog | 31701892 |
| rs13271626 | 8:22467760 | rs4872007 | 8:22538879 | 0,86475 | superior temporal gyrus volume | - | GWAS_catalog | 31530798 |
| rs13271626 | 8:22467760 | rs746011 | 8:22457804 | 0,95576 | triglycerides | - | GWAS_catalog | 30275531 |
| rs1838392 | 8:71682583 | rs34326193 | 8:71502376 | 0,80721 | Bone mineral density | Skeletal | GWAS_ATLAS | 28869591 |
| rs1838392 | 8:71682583 | rs7003794 | 8:71789146 | 0,99168 | heel bone mineral density | - | GWAS_catalog | 28869591 |
| rs1838392 | 8:71682583 | rs34326193 | 8:71502376 | 0,80721 | Heel bone mineral density | Skeletal | GWAS_ATLAS | 30048462 |
| rs1838392 | 8:71682583 | - | - | - | Heel bone mineral density | - | Phenoscanner | UKBB |
| rs1838392 | 8:71682583 | - | - | - | Heel bone mineral density left | - | Phenoscanner | UKBB |
| rs1838392 | 8:71682583 | rs7844508 | 8:71602995 | 0,97935 | Snoring | Psychiatric | GWAS_ATLAS | 30804565 |
| rs1838392 | 8:71682583 | rs7844508 | 8:71602995 | 0,97935 | Waist-hip ratio | Metabolic | GWAS_ATLAS | 30239722 |
| rs1838392 | 8:71682583 | rs1481801 | 8:71989719 | 0,85102 | waist-hip ratio | - | GWAS_catalog | 30595370 |
| rs2581260 | 8:77493526 | - | - | - | Heel bone mineral density | Skeletal | GWAS_ATLAS | 30048462 |
| rs2581260 | 8:77493526 | - | - | - | heel bone mineral density | - | GWAS_catalog | 30595370 |
| rs10956488 | 8:130717755 | - | - | - | Arm fat-free mass left | - | Phenoscanner | UKBB |
| rs10956488 | 8:130717755 | - | - | - | Arm fat-free mass right | - | Phenoscanner | UKBB |
| rs10956488 | 8:130717755 | - | - | - | Arm predicted mass left | - | Phenoscanner | UKBB |
| rs10956488 | 8:130717755 | - | - | - | Arm predicted mass right | - | Phenoscanner | UKBB |
| rs10956488 | 8:130717755 | - | - | - | Back pain | Neurological | GWAS_ATLAS | 31427789 |
| rs10956488 | 8:130717755 | - | - | - | Basal metabolic rate | - | Phenoscanner | UKBB |
| rs10956488 | 8:130717755 | - | - | - | Comparative height size | Skeletal | GWAS_ATLAS | 31427789 |
| rs10956488 | 8:130717755 | - | - | - | Comparative height size at age 10 | - | Phenoscanner | UKBB |
| rs10956488 | 8:130717755 | - | - | - | Forced vital capacity | - | Phenoscanner | UKBB |
| rs10956488 | 8:130717755 | - | - | - | Forced vital capacity, best measure | - | Phenoscanner | UKBB |
| rs10956488 | 8:130717755 | - | - | - | Heel bone mineral density | Skeletal | GWAS_ATLAS | 30048462 |
| rs10956488 | 8:130717755 | - | - | - | Height | Skeletal | GWAS_ATLAS | 31427789 |
| rs10956488 | 8:130717755 | - | - | - | Height | - | Phenoscanner | UKBB |
| rs10956488 | 8:130717755 | - | - | - | Impedance measures - Arm fat-free mass (left) | Metabolic | GWAS_ATLAS | 31427789 |
| rs10956488 | 8:130717755 | - | - | - | Impedance measures - Arm fat-free mass (right) | Metabolic | GWAS_ATLAS | 31427789 |
| rs10956488 | 8:130717755 | - | - | - | Impedance measures - Arm predicted mass (left) | Metabolic | GWAS_ATLAS | 31427789 |
| rs10956488 | 8:130717755 | - | - | - | Impedance measures - Arm predicted mass (right) | Metabolic | GWAS_ATLAS | 31427789 |
| rs10956488 | 8:130717755 | - | - | - | Impedance measures - Basal metabolic rate | Metabolic | GWAS_ATLAS | 31427789 |
| rs10956488 | 8:130717755 | - | - | - | Impedance measures - Leg fat-free mass (left) | Metabolic | GWAS_ATLAS | 31427789 |
| rs10956488 | 8:130717755 | - | - | - | Impedance measures - Leg fat-free mass (right) | Metabolic | GWAS_ATLAS | 31427789 |
| rs10956488 | 8:130717755 | - | - | - | Impedance measures - Leg predicted mass (left) | Metabolic | GWAS_ATLAS | 31427789 |
| rs10956488 | 8:130717755 | - | - | - | Impedance measures - Leg predicted mass (right) | Metabolic | GWAS_ATLAS | 31427789 |
| rs10956488 | 8:130717755 | - | - | - | Impedance measures - Trunk fat-free mass | Metabolic | GWAS_ATLAS | 31427789 |
| rs10956488 | 8:130717755 | - | - | - | Impedance measures - Trunk predicted mass | Metabolic | GWAS_ATLAS | 31427789 |
| rs10956488 | 8:130717755 | - | - | - | Impedance measures - Whole body fat-free mass | Metabolic | GWAS_ATLAS | 31427789 |
| rs10956488 | 8:130717755 | - | - | - | Impedance measures - Whole body water mass | Metabolic | GWAS_ATLAS | 31427789 |
| rs10956488 | 8:130717755 | - | - | - | Leg fat-free mass left | - | Phenoscanner | UKBB |
| rs10956488 | 8:130717755 | - | - | - | Leg fat-free mass right | - | Phenoscanner | UKBB |
| rs10956488 | 8:130717755 | - | - | - | Leg predicted mass left | - | Phenoscanner | UKBB |
| rs10956488 | 8:130717755 | - | - | - | Leg predicted mass right | - | Phenoscanner | UKBB |
| rs10956488 | 8:130717755 | - | - | - | Leg-lef fat ratio | Metabolic | GWAS_ATLAS | 30664634 |
| rs10956488 | 8:130717755 | - | - | - | Sitting height | Skeletal | GWAS_ATLAS | 31427789 |
| rs10956488 | 8:130717755 | - | - | - | Sitting height | - | Phenoscanner | UKBB |
| rs10956488 | 8:130717755 | - | - | - | Trunk fat-free mass | - | Phenoscanner | UKBB |
| rs10956488 | 8:130717755 | - | - | - | Trunk predicted mass | - | Phenoscanner | UKBB |
| rs10956488 | 8:130717755 | - | - | - | Trunk-trunk fat ratio | Metabolic | GWAS_ATLAS | 30664634 |
| rs10956488 | 8:130717755 | - | - | - | Whole body fat-free mass | - | Phenoscanner | UKBB |
| rs10956488 | 8:130717755 | - | - | - | Whole body water mass | - | Phenoscanner | UKBB |
| rs58579887 | 8:145028587 | rs7014582 | 8:144990528 | 0,82786 | 5-oxoproline | Metabolic | GWAS_ATLAS | 24816252 |
| rs58579887 | 8:145028587 | rs7003580 | 8:145007534 | 0,95547 | anti-saccade response | - | GWAS_catalog | 29064472 |
| rs58579887 | 8:145028587 | - | - | - | Arm fat-free mass left | - | Phenoscanner | UKBB |
| rs58579887 | 8:145028587 | - | - | - | Arm fat-free mass right | - | Phenoscanner | UKBB |
| rs58579887 | 8:145028587 | - | - | - | Arm predicted mass left | - | Phenoscanner | UKBB |
| rs58579887 | 8:145028587 | - | - | - | Arm predicted mass right | - | Phenoscanner | UKBB |
| rs58579887 | 8:145028587 | rs11777239 | 8:145031265 | 0,9716 | Basal metabolic rate | - | Phenoscanner | UKBB |
| rs58579887 | 8:145028587 | rs7819099 | 8:144992862 | 0,85686 | Body of corpus callosum fractional anisotropy | Neurological | GWAS_ATLAS | v: https://doi.org/10.1101/2 |
| rs58579887 | 8:145028587 | rs7819099 | 8:144992862 | 0,85686 | Body of corpus callosum radial diusivities | Neurological | GWAS_ATLAS | v: https://doi.org/10.1101/2 |
| rs58579887 | 8:145028587 | rs11136342 | 8:145044749 | 0,91195 | Cholesterol lowering medication | Activities | GWAS_ATLAS | 31427789 |
| rs58579887 | 8:145028587 | rs7155 | 8:144989462 | 0,82004 | Comparative height size | Skeletal | GWAS_ATLAS | 31427789 |
| rs58579887 | 8:145028587 | - | - | - | Comparative height size at age 10 | - | Phenoscanner | UKBB |
| rs58579887 | 8:145028587 | rs11136336 | 8:145007187 | 0,95566 | fat-free mass | - | GWAS_catalog | 30593698 |
| rs58579887 | 8:145028587 | rs7464572 | 8:145021167 | 0,96362 | fibrinogen | - | GWAS_catalog | 23969696 |
| rs58579887 | 8:145028587 | rs7464572 | 8:145021167 | 0,96362 | fibrinogen levels | - | GWAS_catalog | 28107422 |
| rs58579887 | 8:145028587 | rs7819099 | 8:144992862 | 0,85686 | Genu of corpus callosum mean diusivities | Neurological | GWAS_ATLAS | v: https://doi.org/10.1101/2 |
| rs58579887 | 8:145028587 | rs7155 | 8:144989462 | 0,82004 | Genu of corpus callosum radial diusivities | Neurological | GWAS_ATLAS | v: https://doi.org/10.1101/2 |
| rs58579887 | 8:145028587 | rs11783772 | 8:145008443 | 0,95566 | height | - | GWAS_catalog | 30595370 |
| rs58579887 | 8:145028587 | rs7155 | 8:144989462 | 0,82004 | Height | Skeletal | GWAS_ATLAS | 31427789 |
| rs58579887 | 8:145028587 | - | - | - | Height | - | Phenoscanner | UKBB |
| rs58579887 | 8:145028587 | rs7155 | 8:144989462 | 0,82004 | HMG CoA reductase inhibitors | Environmental | GWAS_ATLAS | 31015401 |
| rs58579887 | 8:145028587 | rs7155 | 8:144989462 | 0,82004 | Impedance measures - Arm fat-free mass (left) | Metabolic | GWAS_ATLAS | 31427789 |
| rs58579887 | 8:145028587 | rs7155 | 8:144989462 | 0,82004 | Impedance measures - Arm fat-free mass (right) | Metabolic | GWAS_ATLAS | 31427789 |
| rs58579887 | 8:145028587 | rs7155 | 8:144989462 | 0,82004 | Impedance measures - Arm predicted mass (left) | Metabolic | GWAS_ATLAS | 31427789 |
| rs58579887 | 8:145028587 | rs7155 | 8:144989462 | 0,82004 | Impedance measures - Arm predicted mass (right) | Metabolic | GWAS_ATLAS | 31427789 |
| rs58579887 | 8:145028587 | rs7014582 | 8:144990528 | 0,82786 | Impedance measures - Basal metabolic rate | Metabolic | GWAS_ATLAS | 31427789 |
| rs58579887 | 8:145028587 | rs7155 | 8:144989462 | 0,82004 | Impedance measures - Trunk fat-free mass | Metabolic | GWAS_ATLAS | 31427789 |

| | | | | | | | | |
|---|---|---|---|---|---|---|---|---|
| rs58579887 | 8:145028587 | rs7155 | 8:144989462 | 0,82004 | Impedance measures - Trunk predicted mass | Metabolic | GWAS_ATLAS | 31427789 |
| rs58579887 | 8:145028587 | rs7155 | 8:144989462 | 0,82004 | Impedance measures - Whole body fat-free mass | Metabolic | GWAS_ATLAS | 31427789 |
| rs58579887 | 8:145028587 | rs7155 | 8:144989462 | 0,82004 | Impedance measures - Whole body water mass | Metabolic | GWAS_ATLAS | 31427789 |
| rs58579887 | 8:145028587 | rs11777239 | 8:145031265 | 0,9716 | LDL cholesterol | - | Phenoscanner | 20686565 |
| rs58579887 | 8:145028587 | rs55831924 | 8:145031968 | 0,85753 | ldl cholesterol | - | GWAS_catalog | 30275531 |
| rs58579887 | 8:145028587 | rs7832643 | 8:145022657 | 0,96354 | ldl cholesterol levels | - | GWAS_catalog | 28334899 |
| rs58579887 | 8:145028587 | rs11777239 | 8:145031265 | 0,9716 | Low density lipoprotein | - | Phenoscanner | 24097068 |
| rs58579887 | 8:145028587 | rs11783655 | 8:145037573 | 0,93559 | low density lipoprotein cholesterol levels | - | GWAS_catalog | 29507422 |
| rs58579887 | 8:145028587 | rs7014582 | 8:144990528 | 0,82786 | Low-density lipoprotein cholesterol | Metabolic | GWAS_ATLAS | 24097068 |
| rs58579887 | 8:145028587 | rs7819099 | 8:144992862 | 0,85686 | Mean corpuscular volume | Immunological | GWAS_ATLAS | 27863252 |
| rs58579887 | 8:145028587 | - | - | - | Mean corpuscular volume | - | Phenoscanner | 27863252 |
| rs58579887 | 8:145028587 | rs7155 | 8:144989462 | 0,82004 | Mean platelet volume | Immunological | GWAS_ATLAS | 27863252 |
| rs58579887 | 8:145028587 | - | - | - | Mean platelet volume | - | Phenoscanner | 27863252 |
| rs58579887 | 8:145028587 | rs11787365 | 8:145041333 | 0,94749 | medication use (hmg coa reductase inhibitors) | - | GWAS_catalog | 31015401 |
| rs58579887 | 8:145028587 | rs11136335 | 8:145003777 | 0,8057 | mosquito bite size | - | GWAS_catalog | 28199695 |
| rs58579887 | 8:145028587 | rs11780978 | 8:145034852 | 0,94354 | osteoarthritis (hip) | - | GWAS_catalog | 30374069 |
| rs58579887 | 8:145028587 | rs11780978 | 8:145034852 | 0,94354 | osteoarthritis of the hip (hospital diagnosed) | - | GWAS_catalog | 29559693 |
| rs58579887 | 8:145028587 | rs7833924 | 8:144996029 | 0,86044 | platelet count | - | GWAS_catalog | 27863252 |
| rs58579887 | 8:145028587 | rs7155 | 8:144989462 | 0,82004 | Platelet count | Immunological | GWAS_ATLAS | 27863252 |
| rs58579887 | 8:145028587 | - | - | - | Platelet count | - | Phenoscanner | 27863252 |
| rs58579887 | 8:145028587 | rs7155 | 8:144989462 | 0,82004 | Platelet distribution width | Immunological | GWAS_ATLAS | 27863252 |
| rs58579887 | 8:145028587 | - | - | - | Platelet distribution width | - | Phenoscanner | 27863252 |
| rs58579887 | 8:145028587 | rs7155 | 8:144989462 | 0,82004 | Plateletcrit | Immunological | GWAS_ATLAS | 27863252 |
| rs58579887 | 8:145028587 | - | - | - | Plateletcrit | - | Phenoscanner | 27863252 |
| rs58579887 | 8:145028587 | rs7155 | 8:144989462 | 0,82004 | post bronchodilator fev1 | - | GWAS_catalog | 26634245 |
| rs58579887 | 8:145028587 | rs7819099 | 8:144992862 | 0,85686 | red blood cell count | - | GWAS_catalog | 30595370 |
| rs58579887 | 8:145028587 | rs7155 | 8:144989462 | 0,82004 | Sitting height | Skeletal | GWAS_ATLAS | 31427789 |
| rs58579887 | 8:145028587 | - | - | - | Sitting height | - | Phenoscanner | UKBB |
| rs58579887 | 8:145028587 | rs11784762 | 8:145009610 | 0,94757 | Total cholesterol | Metabolic | GWAS_ATLAS | 24097068 |
| rs58579887 | 8:145028587 | rs11777239 | 8:145031265 | 0,9716 | Total cholesterol | - | Phenoscanner | 24097068 |
| rs58579887 | 8:145028587 | rs11784762 | 8:145009610 | 0,94757 | Total cholesterol in IDL | Metabolic | GWAS_ATLAS | 27005778 |
| rs58579887 | 8:145028587 | rs62523994 | 8:145026582 | 0,9635 | Total cholesterol in LDL | Metabolic | GWAS_ATLAS | 27005778 |
| rs58579887 | 8:145028587 | rs7832643 | 8:145022657 | 0,96354 | total cholesterol levels | - | GWAS_catalog | 28334899 |
| rs58579887 | 8:145028587 | rs62523994 | 8:145026582 | 0,9635 | Total lipids in IDL | Metabolic | GWAS_ATLAS | 27005778 |
| rs58579887 | 8:145028587 | - | - | - | Trunk fat-free mass | - | Phenoscanner | UKBB |
| rs58579887 | 8:145028587 | - | - | - | Trunk predicted mass | - | Phenoscanner | UKBB |
| rs58579887 | 8:145028587 | rs11777239 | 8:145031265 | 0,9716 | white matter microstructure (radial diusivities) | - | GWAS_catalog | 31666681 |
| rs58579887 | 8:145028587 | - | - | - | Whole body fat-free mass | - | Phenoscanner | UKBB |
| rs58579887 | 8:145028587 | - | - | - | Whole body water mass | - | Phenoscanner | UKBB |
| rs1333047 | 9:22124504 | rs10757274 | 9:22096055 | 0,83651 | abdominal aortic aneurysm | - | GWAS_catalog | 27899403 |
| rs1333047 | 9:22124504 | - | - | - | Acute myocardial infarction | - | Phenoscanner | UKBB |
| rs1333047 | 9:22124504 | rs1333048 | 9:22125347 | 0,87696 | age-related diseases and mortality | - | GWAS_catalog | 27790247 |
| rs1333047 | 9:22124504 | rs1333049 | 9:22125503 | 0,93387 | age-related diseases, mortality and associated endophenotypes | - | GWAS_catalog | 27790247 |
| rs1333047 | 9:22124504 | rs10738606 | 9:22088090 | 0,85091 | Agents acting on the renin-angiotensin system | Environmental | GWAS_ATLAS | 31015401 |
| rs1333047 | 9:22124504 | rs10738606 | 9:22088090 | 0,85091 | Angina | Cardiovascular | GWAS_ATLAS | 31427789 |
| rs1333047 | 9:22124504 | - | - | - | Angina pectoris | - | Phenoscanner | UKBB |
| rs1333047 | 9:22124504 | rs10738606 | 9:22088090 | 0,85091 | Antithrombotic agents | Environmental | GWAS_ATLAS | 31015401 |
| rs1333047 | 9:22124504 | rs10738606 | 9:22088090 | 0,85091 | Aspirin | Activities | GWAS_ATLAS | 31427789 |
| rs1333047 | 9:22124504 | rs10738606 | 9:22088090 | 0,85091 | Atenolol | Activities | GWAS_ATLAS | 31427789 |
| rs1333047 | 9:22124504 | rs10738606 | 9:22088090 | 0,85091 | Atorvastatin | Activities | GWAS_ATLAS | 31427789 |
| rs1333047 | 9:22124504 | rs10738606 | 9:22088090 | 0,85091 | Beta blocking agents | Environmental | GWAS_ATLAS | 31015401 |
| rs1333047 | 9:22124504 | rs9644862 | 9:22090936 | 0,87051 | carotid plaque burden | - | GWAS_catalog | 28282560 |
| rs1333047 | 9:22124504 | rs10738606 | 9:22088090 | 0,85091 | Cholesterol lowering medication | Activities | GWAS_ATLAS | 31427789 |
| rs1333047 | 9:22124504 | - | - | - | Chronic ischaemic heart disease | - | Phenoscanner | UKBB |
| rs1333047 | 9:22124504 | rs10738606 | 9:22088090 | 0,85091 | Chronic ischemic heart disease | Cardiovascular | GWAS_ATLAS | 31427789 |
| rs1333047 | 9:22124504 | rs1412834 | 9:22110131 | 0,87178 | colorectal cancer | - | GWAS_catalog | 31089142 |
| rs1333047 | 9:22124504 | rs1333049 | 9:22125503 | 0,93387 | coronary artery calcification | - | GWAS_catalog | 22144573 |
| rs1333047 | 9:22124504 | - | - | - | Coronary artery calcification CAC | - | Phenoscanner | 22144573 |
| rs1333047 | 9:22124504 | - | - | - | Coronary artery calcification CAC excluding previous myocardial infarction cases | - | Phenoscanner | 22144573 |
| rs1333047 | 9:22124504 | - | - | - | Coronary artery disease | - | Phenoscanner | 21378990 |
| rs1333047 | 9:22124504 | rs10738606 | 9:22088090 | 0,85091 | Coronary artery disease | Cardiovascular | GWAS_ATLAS | 26343387 |
| rs1333047 | 9:22124504 | rs10738607 | 9:22088094 | 0,85091 | coronary artery disease | - | GWAS_catalog | 26708285 |
| rs1333047 | 9:22124504 | rs2891168 | 9:22098619 | 0,83651 | coronary artery disease (myocardial infarction, percutaneous transluminal coronary angioplasty, coronary artery bypass grafting, angina or chromic ischemic heart disease) | - | GWAS_catalog | 28714975 |
| rs1333047 | 9:22124504 | rs1333049 | 9:22125503 | 0,93387 | coronary artery disease or ischemic stroke | - | GWAS_catalog | 24262325 |
| rs1333047 | 9:22124504 | rs1333049 | 9:22125503 | 0,93387 | coronary artery disease or large artery stroke | - | GWAS_catalog | 24262325 |
| rs1333047 | 9:22124504 | rs10757278 | 9:22124477 | 0,96478 | Coronary heart disease | - | Phenoscanner | 21347282 |
| rs1333047 | 9:22124504 | rs10757274 | 9:22096055 | 0,83651 | coronary heart disease | - | GWAS_catalog | 22751097 |
| rs1333047 | 9:22124504 | rs10738606 | 9:22088090 | 0,85091 | Cup area | Ophthalmological | GWAS_ATLAS | 28073927 |
| rs1333047 | 9:22124504 | rs1333045 | 9:22119195 | 0,82743 | Diabetes | Endocrine | GWAS_ATLAS | 31427789 |
| rs1333047 | 9:22124504 | rs9644861 | 9:22090935 | 0,88859 | diastolic blood pressure | - | GWAS_catalog | 30578418 |
| rs1333047 | 9:22124504 | rs10738606 | 9:22088090 | 0,85091 | Disorders of lipoprotein metabolism and other lipidemias | Metabolic | GWAS_ATLAS | 31427789 |
| rs1333047 | 9:22124504 | rs10757272 | 9:22088260 | 0,84351 | endometriosis | - | GWAS_catalog | 28537267 |
| rs1333047 | 9:22124504 | rs10738606 | 9:22088090 | 0,85091 | Family (maternal) history of Heart disease | Environment | GWAS_ATLAS | 31427789 |
| rs1333047 | 9:22124504 | rs10738606 | 9:22088090 | 0,85091 | Family (paternal) history of Bowel cancer | Environment | GWAS_ATLAS | 31427789 |
| rs1333047 | 9:22124504 | rs10738606 | 9:22088090 | 0,85091 | Family (paternal) history of Heart disease | Environment | GWAS_ATLAS | 31427789 |
| rs1333047 | 9:22124504 | rs10738606 | 9:22088090 | 0,85091 | Family (siblings) history of Heart disease | Environment | GWAS_ATLAS | 31427789 |
| rs1333047 | 9:22124504 | rs10738606 | 9:22088090 | 0,85091 | Family history of certain disabilities and chronic diseases (leading to disablement) | Mortality | GWAS_ATLAS | 31427789 |
| rs1333047 | 9:22124504 | rs10738606 | 9:22088090 | 0,85091 | Father's age at death | Mortality | GWAS_ATLAS | 31427789 |
| rs1333047 | 9:22124504 | - | - | - | Fathers age at death | - | Phenoscanner | UKBB |
| rs1333047 | 9:22124504 | rs10738606 | 9:22088090 | 0,85091 | Hair colour: Black | Dermatological | GWAS_ATLAS | 31427789 |
| rs1333047 | 9:22124504 | rs10738606 | 9:22088090 | 0,85091 | Hair colour: Dark brown | Dermatological | GWAS_ATLAS | 31427789 |
| rs1333047 | 9:22124504 | rs1537371 | 9:22099568 | 0,89244 | Hair colour: Light brown | Dermatological | GWAS_ATLAS | 31427789 |
| rs1333047 | 9:22124504 | rs7859727 | 9:22102165 | 0,85827 | healthspan | - | GWAS_catalog | 30729179 |
| rs1333047 | 9:22124504 | rs1556516 | 9:22100176 | 0,89244 | heart failure | - | GWAS_catalog | 31919418 |
| rs1333047 | 9:22124504 | rs10738606 | 9:22088090 | 0,85091 | HMG CoA reductase inhibitors | Environmental | GWAS_ATLAS | 31015401 |
| rs1333047 | 9:22124504 | - | - | - | hypertension | - | GWAS_catalog | 30487518 |
| rs1333047 | 9:22124504 | - | - | - | Illnesses of father: heart disease | - | Phenoscanner | UKBB |
| rs1333047 | 9:22124504 | - | - | - | Illnesses of father: none of the above, group 1 | - | Phenoscanner | UKBB |
| rs1333047 | 9:22124504 | - | - | - | Illnesses of mother: heart disease | - | Phenoscanner | UKBB |
| rs1333047 | 9:22124504 | - | - | - | Illnesses of siblings: heart disease | - | Phenoscanner | UKBB |
| rs1333047 | 9:22124504 | rs10757272 | 9:22088260 | 0,84351 | intracranial aneurysm | - | GWAS_catalog | 22286173 |
| rs1333047 | 9:22124504 | rs7866503 | 9:22091924 | 0,86667 | intracranial, abdominal aortic or thoracic aortic aneurysm (pleiotropy) | - | GWAS_catalog | 27418160 |
| rs1333047 | 9:22124504 | rs7859727 | 9:22102165 | 0,85827 | ischemic stroke | - | GWAS_catalog | 29531354 |
| rs1333047 | 9:22124504 | - | - | - | large artery stroke | - | GWAS_catalog | 24262325 |
| rs1333047 | 9:22124504 | rs1333045 | 9:22119195 | 0,82743 | Life span | Mortality | GWAS_ATLAS | 28748955 |
| rs1333047 | 9:22124504 | rs10738606 | 9:22088090 | 0,85091 | Long-standing illness, disability or infirmity | Mortality | GWAS_ATLAS | 31427789 |
| rs1333047 | 9:22124504 | - | - | - | Medication for cholesterol, blood pressure or diabetes: cholesterol lowering medication | - | Phenoscanner | UKBB |
| rs1333047 | 9:22124504 | - | - | - | Medication for pain relief, constipation, heartburn: aspirin | - | Phenoscanner | UKBB |
| rs1333047 | 9:22124504 | rs2891168 | 9:22098619 | 0,83651 | medication use (beta blocking agents) | - | GWAS_catalog | 31015401 |
| rs1333047 | 9:22124504 | rs1537371 | 9:22099568 | 0,89244 | medication use (hmg coa reductase inhibitors) | - | GWAS_catalog | 31015401 |
| rs1333047 | 9:22124504 | - | - | - | medication use (vasodilators used in cardiac diseases) | - | GWAS_catalog | 31015401 |
| rs1333047 | 9:22124504 | - | - | - | Myocardial infarction | - | Phenoscanner | 26343387 |
| rs1333047 | 9:22124504 | rs10738607 | 9:22088094 | 0,85091 | myocardial infarction | - | GWAS_catalog | 26708285 |
| rs1333047 | 9:22124504 | rs4977574 | 9:22098574 | 0,83651 | myocardial infarction (early onset) | - | GWAS_catalog | 19198609 |
| rs1333047 | 9:22124504 | rs10738606 | 9:22088090 | 0,85091 | Number of treatments/medications taken | Activities | GWAS_ATLAS | 31427789 |
| rs1333047 | 9:22124504 | rs10738606 | 9:22088090 | 0,85091 | Open-angle glaucoma | Ophthalmological | GWAS_ATLAS | 29891935 |
| rs1333047 | 9:22124504 | rs1333042 | 9:22103813 | 0,8888 | parental longevity (both parents in top 10%) | - | GWAS_catalog | 29227965 |
| rs1333047 | 9:22124504 | rs1556516 | 9:22100176 | 0,89244 | parental longevity (combined parental age at death) | - | GWAS_catalog | 29227965 |
| rs1333047 | 9:22124504 | rs1556516 | 9:22100176 | 0,89244 | parental longevity (combined parental attained age, martingale residuals) | - | GWAS_catalog | 29227965 |
| rs1333047 | 9:22124504 | rs1556516 | 9:22100176 | 0,89244 | parental longevity (father's age at death) | - | GWAS_catalog | 29227965 |
| rs1333047 | 9:22124504 | rs1556516 | 9:22100176 | 0,89244 | parental longevity (father's attained age) | - | GWAS_catalog | 29227965 |
| rs1333047 | 9:22124504 | rs1333042 | 9:22103813 | 0,8888 | parental longevity (mother's age at death or mother's attained age) | - | GWAS_catalog | 31484785 |
| rs1333047 | 9:22124504 | rs1333045 | 9:22119195 | 0,82743 | parental longevity (mother's age at death) | - | GWAS_catalog | 29227965 |
| rs1333047 | 9:22124504 | rs10738606 | 9:22088090 | 0,85091 | Presence of cardiac and vascular implants and grafts | Mortality | GWAS_ATLAS | 31427789 |
| rs1333047 | 9:22124504 | rs1333045 | 9:22119195 | 0,82743 | pulse pressure | - | GWAS_catalog | 30578418 |
| rs1333047 | 9:22124504 | - | - | - | Self-reported angina | - | Phenoscanner | UKBB |
| rs1333047 | 9:22124504 | - | - | - | Self-reported heart attack or myocardial infarction | - | Phenoscanner | UKBB |
| rs1333047 | 9:22124504 | rs1333045 | 9:22119195 | 0,82743 | serum total cholesterol levels | - | GWAS_catalog | 31201950 |
| rs1333047 | 9:22124504 | rs10738606 | 9:22088090 | 0,85091 | Simvastatin | Activities | GWAS_ATLAS | 31427789 |
| rs1333047 | 9:22124504 | rs7859727 | 9:22102165 | 0,85827 | stroke | - | GWAS_catalog | 29531354 |
| rs1333047 | 9:22124504 | rs1333045 | 9:22119195 | 0,82743 | systolic blood pressure | - | GWAS_catalog | 30595370 |
| rs1333047 | 9:22124504 | rs10738607 | 9:22088094 | 0,85091 | total cholesterol levels | - | GWAS_catalog | 30275531 |
| rs1333047 | 9:22124504 | - | - | - | Treatment with aspirin | - | Phenoscanner | UKBB |
| rs1333047 | 9:22124504 | - | - | - | Treatment with atorvastatin | - | Phenoscanner | UKBB |
| rs1333047 | 9:22124504 | - | - | - | Treatment with bisoprolol | - | Phenoscanner | UKBB |
| rs1333047 | 9:22124504 | - | - | - | Treatment with clopidogrel | - | Phenoscanner | UKBB |
| rs1333047 | 9:22124504 | - | - | - | Treatment with nicorandil | - | Phenoscanner | UKBB |
| rs1333047 | 9:22124504 | - | - | - | Treatment with simvastatin | - | Phenoscanner | UKBB |
| rs1333047 | 9:22124504 | rs1333045 | 9:22119195 | 0,82743 | Type 2 Diabetes | Endocrine | GWAS_ATLAS | 31427789 |
| rs1333047 | 9:22124504 | - | - | - | Vascular or heart problems diagnosed by doctor: angina | - | Phenoscanner | UKBB |
| rs1333047 | 9:22124504 | - | - | - | Vascular or heart problems diagnosed by doctor: heart attack | - | Phenoscanner | UKBB |
| rs1333047 | 9:22124504 | rs10738606 | 9:22088090 | 0,85091 | Vasodilators used in cardiac diseases | Environmental | GWAS_ATLAS | 31015401 |
| rs1333047 | 9:22124504 | rs10738606 | 9:22088090 | 0,85091 | Vertical cup-disc ratio | Ophthalmological | GWAS_ATLAS | 28073927 |
| rs755209 | 9:95489671 | rs10992304 | 9:95085365 | 0,81825 | Comparative height size | Skeletal | GWAS_ATLAS | 31427789 |
| rs755209 | 9:95489671 | rs10992304 | 9:95085365 | 0,81825 | FEV2 | Respiratory | GWAS_ATLAS | 30804560 |
| rs755209 | 9:95489671 | rs10992304 | 9:95085365 | 0,81825 | Height | Skeletal | GWAS_ATLAS | 25282103 |
| rs755209 | 9:95489671 | - | - | - | Height | - | Phenoscanner | UKBB |
| rs755209 | 9:95489671 | rs10992304 | 9:95085365 | 0,81825 | Impedance measures - Impedance of arm (left) | Metabolic | GWAS_ATLAS | 31427789 |
| rs755209 | 9:95489671 | rs10992304 | 9:95085365 | 0,81825 | Impedance measures - Impedance of arm (right) | Metabolic | GWAS_ATLAS | 31427789 |
| rs755209 | 9:95489671 | rs10992304 | 9:95085365 | 0,81825 | Impedance measures - Impedance of leg (left) | Metabolic | GWAS_ATLAS | 31427789 |
| rs755209 | 9:95489671 | rs10992304 | 9:95085365 | 0,81825 | Impedance measures - Impedance of whole body | Metabolic | GWAS_ATLAS | 31427789 |
| rs755209 | 9:95489671 | - | - | - | Impedance of arm left | - | Phenoscanner | UKBB |

| | | | | | | | | |
|---|---|---|---|---|---|---|---|---|
| rs755209 | 9:95489671 | - | - | - | Impedance of arm right | - | Phenoscanner | UKBB |
| rs755209 | 9:95489671 | - | - | - | Impedance of whole body | - | Phenoscanner | UKBB |
| rs755209 | 9:95489671 | rs10992304 | 9:95085365 | 0,81825 | Sitting height | Skeletal | GWAS_ATLAS | 31427789 |
| rs755209 | 9:95489671 | rs10992304 | 9:95085365 | 0,81825 | Trunk-trunk fat ratio | Metabolic | GWAS_ATLAS | 30664634 |
| rs755209 | 9:95489671 | rs10992304 | 9:95085365 | 0,81825 | Waist-hip ratio | Metabolic | GWAS_ATLAS | 30239722 |
| rs676996 | 9:136146077 | chr9:136142203 | 9:136142203 | 0,99152 | Activated partial thromboplastin time | - | Phenoscanner | 22703881 |
| rs676996 | 9:136146077 | rs687621 | 9:136137065 | 0,98307 | Activated partial thromboplastin time | Metabolic | GWAS_ATLAS | 29403010 |
| rs676996 | 9:136146077 | rs687621 | 9:136137065 | 0,98307 | ADpSGEGDFXAEGGGVR | Metabolic | GWAS_ATLAS | 24816252 |
| rs676996 | 9:136146077 | - | - | - | Alkaline phosphatase | - | Phenoscanner | 28887542 |
| rs676996 | 9:136146077 | rs687621 | 9:136137065 | 0,98307 | Antithrombotic agents | Environmental | GWAS_ATLAS | 31015401 |
| rs676996 | 9:136146077 | - | - | - | Arm fat percentage left | - | Phenoscanner | UKBB |
| rs676996 | 9:136146077 | - | - | - | Arm fat percentage right | - | Phenoscanner | UKBB |
| rs676996 | 9:136146077 | rs687289 | 9:136137106 | 0,98728 | BCAM - Basal Cell Adhesion Molecule | Cell | GWAS_ATLAS | 28240269 |
| rs676996 | 9:136146077 | - | - | - | Blood clot in the leg | - | Phenoscanner | UKBB |
| rs676996 | 9:136146077 | - | - | - | Blood clot in the lung | - | Phenoscanner | UKBB |
| rs676996 | 9:136146077 | chr9:136143442 | 9:136143442 | 0,9873 | blood protein levels | - | GWAS_catalog | 28240269 |
| rs676996 | 9:136146077 | rs687621 | 9:136137065 | 0,98307 | Bone mineral density | Skeletal | GWAS_ATLAS | 28869591 |
| rs676996 | 9:136146077 | chr9:136142355 | 9:136142355 | 0,87713 | c-reactive protein levels | - | GWAS_catalog | 30388399 |
| rs676996 | 9:136146077 | chr9:136142203 | 9:136142203 | 0,99152 | Cardiovascular diseases | - | Phenoscanner | 21239051 |
| rs676996 | 9:136146077 | rs687289 | 9:136137106 | 0,98728 | CD209 - CD209 antigen | Cell | GWAS_ATLAS | 28240269 |
| rs676996 | 9:136146077 | rs687289 | 9:136137106 | 0,98728 | CD36 - Platelet glycoprotein 4 | Cell | GWAS_ATLAS | 28240269 |
| rs676996 | 9:136146077 | rs687621 | 9:136137065 | 0,98307 | Cholesterol lowering medication | Activities | GWAS_ATLAS | 31427789 |
| rs676996 | 9:136146077 | rs687289 | 9:136137106 | 0,98728 | CHST15 - Carbohydrate sulfotransferase 15 | Cell | GWAS_ATLAS | 28240269 |
| rs676996 | 9:136146077 | chr9:136142203 | 9:136142203 | 0,99152 | Circulating galectin 3 levels | - | Phenoscanner | 23056639 |
| rs676996 | 9:136146077 | chr9:136149229 | 9:136149229 | 0,97891 | clinical laboratory measurements | - | GWAS_catalog | 27897004 |
| rs676996 | 9:136146077 | rs687621 | 9:136137065 | 0,98307 | Coronary artery disease | Cardiovascular | GWAS_ATLAS | 28714975 |
| rs676996 | 9:136146077 | chr9:136142203 | 9:136142203 | 0,99152 | Coronary artery disease | - | Phenoscanner | 28714975 |
| rs676996 | 9:136146077 | rs687621 | 9:136137065 | 0,98307 | Creatine kinase | Metabolic | GWAS_ATLAS | 29403010 |
| rs676996 | 9:136146077 | chr9:136144873 | 9:136144873 | 0,99152 | creatine kinase levels | - | GWAS_catalog | 29403010 |
| rs676996 | 9:136146077 | rs687621 | 9:136137065 | 0,98307 | Diastolic Blood Pressure | Cardiovascular | GWAS_ATLAS | 31427789 |
| rs676996 | 9:136146077 | - | - | - | Diastolic blood pressure | - | Phenoscanner | UKBB |
| rs676996 | 9:136146077 | rs8176719 | 9:136132908 | 0,84854 | Diverticular disease | Gastrointestinal | GWAS_ATLAS | 30661054 |
| rs676996 | 9:136146077 | rs687621 | 9:136137065 | 0,98307 | Diverticular disease of intestine | Gastrointestinal | GWAS_ATLAS | 31427789 |
| rs676996 | 9:136146077 | chr9:136149229 | 9:136149229 | 0,97891 | duodenal ulcer | - | GWAS_catalog | 22387998 |
| rs676996 | 9:136146077 | chr9:136142355 | 9:136142355 | 0,87713 | end-stage coagulation | - | GWAS_catalog | 23381943 |
| rs676996 | 9:136146077 | chr9:136142203 | 9:136142203 | 0,99152 | Factor XIII antigen | - | Phenoscanner | 23381943 |
| rs676996 | 9:136146077 | - | - | - | Ferritin | - | Phenoscanner | 28887542 |
| rs676996 | 9:136146077 | rs687621 | 9:136137065 | 0,98307 | FEV2 | Respiratory | GWAS_ATLAS | 30804560 |
| rs676996 | 9:136146077 | rs687289 | 9:136137106 | 0,98728 | FLT4 - Vascular endothelial growth factor receptor 3 | Cell | GWAS_ATLAS | 28240269 |
| rs676996 | 9:136146077 | chr9:136139265 | 9:136139265 | 0,87713 | gastric parietal cell autoantibody levels in type 1 diabetes | - | GWAS_catalog | 21829393 |
| rs676996 | 9:136146077 | rs687621 | 9:136137065 | 0,98307 | Glycine | Metabolic | GWAS_ATLAS | 31070104 |
| rs676996 | 9:136146077 | chr9:136149229 | 9:136149229 | 0,97891 | graves' disease | - | GWAS_catalog | 23612905 |
| rs676996 | 9:136146077 | - | - | - | Haemorrhoids | - | Phenoscanner | UKBB |
| rs676996 | 9:136146077 | rs687621 | 9:136137065 | 0,98307 | Hayfever, allergic rhinitis or eczema | Respiratory | GWAS_ATLAS | 31427789 |
| rs676996 | 9:136146077 | rs8176719 | 9:136132908 | 0,84854 | Heel bone mineral density | Skeletal | GWAS_ATLAS | 30048462 |
| rs676996 | 9:136146077 | chr9:136142203 | 9:136142203 | 0,99152 | Heel bone mineral density | - | Phenoscanner | UKBB |
| rs676996 | 9:136146077 | rs8176719 | 9:136132908 | 0,84854 | Hematocrit | Immunological | GWAS_ATLAS | 27863252 |
| rs676996 | 9:136146077 | - | - | - | Hematocrit | - | Phenoscanner | 27863252 |
| rs676996 | 9:136146077 | rs8176719 | 9:136132908 | 0,84854 | Hemoglobin concentration | Immunological | GWAS_ATLAS | 27863252 |
| rs676996 | 9:136146077 | - | - | - | Hemoglobin concentration | - | Phenoscanner | 27863252 |
| rs676996 | 9:136146077 | rs687621 | 9:136137065 | 0,98307 | High blood pressure | Cardiovascular | GWAS_ATLAS | 31427789 |
| rs676996 | 9:136146077 | rs687621 | 9:136137065 | 0,98307 | High cholesterol | Metabolic | GWAS_ATLAS | 31427789 |
| rs676996 | 9:136146077 | - | - | - | High grade serous ovarian cancer | - | Phenoscanner | 28346442 |
| rs676996 | 9:136146077 | rs687621 | 9:136137065 | 0,98307 | HMG CoA reductase inhibitors | Environmental | GWAS_ATLAS | 31015401 |
| rs676996 | 9:136146077 | rs687621 | 9:136137065 | 0,98307 | Hypertension | Cardiovascular | GWAS_ATLAS | 31427789 |
| rs676996 | 9:136146077 | chr9:136145484 | 9:136145484 | 0,99152 | Impedance measures - Arm fat percentage (left) | Metabolic | GWAS_ATLAS | 31427789 |
| rs676996 | 9:136146077 | chr9:136143372 | 9:136143372 | 0,99152 | Impedance measures - Arm fat percentage (right) | Metabolic | GWAS_ATLAS | 31427789 |
| rs676996 | 9:136146077 | rs687621 | 9:136137065 | 0,98307 | Impedance measures - Arm fat-free mass (left) | Metabolic | GWAS_ATLAS | 31427789 |
| rs676996 | 9:136146077 | rs687621 | 9:136137065 | 0,98307 | Impedance measures - Arm fat-free mass (right) | Metabolic | GWAS_ATLAS | 31427789 |
| rs676996 | 9:136146077 | rs687621 | 9:136137065 | 0,98307 | Impedance measures - Arm predicted mass (left) | Metabolic | GWAS_ATLAS | 31427789 |
| rs676996 | 9:136146077 | rs687621 | 9:136137065 | 0,98307 | Impedance measures - Arm predicted mass (right) | Metabolic | GWAS_ATLAS | 31427789 |
| rs676996 | 9:136146077 | rs687621 | 9:136137065 | 0,98307 | Impedance measures - Impedance of arm (left) | Metabolic | GWAS_ATLAS | 31427789 |
| rs676996 | 9:136146077 | rs687621 | 9:136137065 | 0,98307 | Impedance measures - Impedance of arm (right) | Metabolic | GWAS_ATLAS | 31427789 |
| rs676996 | 9:136146077 | rs687289 | 9:136137106 | 0,98728 | Impedance measures - Trunk fat-free mass | Metabolic | GWAS_ATLAS | 31427789 |
| rs676996 | 9:136146077 | - | - | - | Impedance of arm left | - | Phenoscanner | UKBB |
| rs676996 | 9:136146077 | - | - | - | Impedance of arm right | - | Phenoscanner | UKBB |
| rs676996 | 9:136146077 | chr9:136142355 | 9:136142355 | 0,87713 | inflammatory biomarkers | - | GWAS_catalog | 22291609 |
| rs676996 | 9:136146077 | rs687289 | 9:136137106 | 0,98728 | INSR - Insulin receptor | Cell | GWAS_ATLAS | 28240269 |
| rs676996 | 9:136146077 | chr9:136149229 | 9:136149229 | 0,97891 | insulin disposition index | - | GWAS_catalog | 23263489 |
| rs676996 | 9:136146077 | chr9:136142203 | 9:136142203 | 0,99152 | Interleukin 6 IL 6 levels | - | Phenoscanner | 22291609 |
| rs676996 | 9:136146077 | rs687289 | 9:136137106 | 0,98728 | KDR - Vascular endothelial growth factor receptor 2 | Cell | GWAS_ATLAS | 28240269 |
| rs676996 | 9:136146077 | rs687289 | 9:136137106 | 0,98728 | Lactate dehydrogenase | Metabolic | GWAS_ATLAS | 29403010 |
| rs676996 | 9:136146077 | chr9:136142203 | 9:136142203 | 0,99152 | LDL cholesterol | - | Phenoscanner | 20686565 |
| rs676996 | 9:136146077 | rs8176719 | 9:136132908 | 0,84854 | Leg-lef fat ratio | Metabolic | GWAS_ATLAS | 30664634 |
| rs676996 | 9:136146077 | chr9:136139265 | 9:136139265 | 0,87713 | liver enzyme levels | - | GWAS_catalog | 18940312 |
| rs676996 | 9:136146077 | chr9:136142203 | 9:136142203 | 0,99152 | Low density lipoprotein | - | Phenoscanner | 24097068 |
| rs676996 | 9:136146077 | rs687621 | 9:136137065 | 0,98307 | Low-density lipoprotein cholesterol | Metabolic | GWAS_ATLAS | 24097068 |
| rs676996 | 9:136146077 | rs687289 | 9:136137106 | 0,98728 | MBL2 - Mannose-binding protein C | Cell | GWAS_ATLAS | 28240269 |
| rs676996 | 9:136146077 | rs687289 | 9:136137106 | 0,98728 | MET - Hepatocyte growth factor receptor | Cell | GWAS_ATLAS | 28240269 |
| rs676996 | 9:136146077 | chr9:136143442 | 9:136143442 | 0,9873 | metabolic traits | - | GWAS_catalog | 21886157 |
| rs676996 | 9:136146077 | rs8176719 | 9:136132908 | 0,84854 | Monocyte count | Immunological | GWAS_ATLAS | 27863252 |
| rs676996 | 9:136146077 | - | - | - | Monocyte count | - | Phenoscanner | 27863252 |
| rs676996 | 9:136146077 | rs8176719 | 9:136132908 | 0,84854 | Monocyte percentage of white cells | Immunological | GWAS_ATLAS | 27863252 |
| rs676996 | 9:136146077 | - | - | - | Monocyte percentage of white cells | - | Phenoscanner | 27863252 |
| rs676996 | 9:136146077 | rs687621 | 9:136137065 | 0,98307 | Myeloid white cell count | Immunological | GWAS_ATLAS | 27863252 |
| rs676996 | 9:136146077 | - | - | - | Myeloid white cell count | - | Phenoscanner | 27863252 |
| rs676996 | 9:136146077 | - | - | - | Myocardial infarction | - | Phenoscanner | 26343387 |
| rs676996 | 9:136146077 | chr9:136142203 | 9:136142203 | 0,99152 | Myocardial infarction in coronary artery disease | - | Phenoscanner | 21239051 |
| rs676996 | 9:136146077 | chr9:136142203 | 9:136142203 | 0,99152 | Myocardial infarction in patients with coronary artery disease by angiography | - | Phenoscanner | 21239051 |
| rs676996 | 9:136146077 | - | - | - | No blood clot, bronchitis, emphysema, asthma, rhinitis, eczema or allergy diagnosed by doctor | - | Phenoscanner | UKBB |
| rs676996 | 9:136146077 | rs8176645 | 9:136149098 | 0,86458 | Non-albumin protein | Metabolic | GWAS_ATLAS | 29403010 |
| rs676996 | 9:136146077 | chr9:136139265 | 9:136139265 | 0,87713 | obesity-related traits | - | GWAS_catalog | 23251661 |
| rs676996 | 9:136146077 | chr9:136149229 | 9:136149229 | 0,97891 | pancreatic cancer | - | GWAS_catalog | 19648918 |
| rs676996 | 9:136146077 | chr9:136149229 | 9:136149229 | 0,97891 | Pancreatic Cancer | Neoplasms | GWAS_ATLAS | 22523087 |
| rs676996 | 9:136146077 | - | - | - | Peak expiratory flow | - | Phenoscanner | UKBB |
| rs676996 | 9:136146077 | rs687621 | 9:136137065 | 0,98307 | PEF | Respiratory | GWAS_ATLAS | 30804560 |
| rs676996 | 9:136146077 | chr9:136149229 | 9:136149229 | 0,97891 | peripheral artery disease | - | GWAS_catalog | 31285632 |
| rs676996 | 9:136146077 | rs687621 | 9:136137065 | 0,98307 | Personal history of certain other diseases | Mortality | GWAS_ATLAS | 31427789 |
| rs676996 | 9:136146077 | rs687621 | 9:136137065 | 0,98307 | Personal history of medical treatment | Mortality | GWAS_ATLAS | 31427789 |
| rs676996 | 9:136146077 | - | - | - | Phlebitis and thrombophlebitis | - | Phenoscanner | UKBB |
| rs676996 | 9:136146077 | chr9:136139265 | 9:136139265 | 0,87713 | phytosterol levels | - | GWAS_catalog | 20529992 |
| rs676996 | 9:136146077 | chr9:136149229 | 9:136149229 | 0,97891 | protein quantitative trait loci | - | GWAS_catalog | 18464913 |
| rs676996 | 9:136146077 | - | - | - | Pulmonary embolism | - | Phenoscanner | UKBB |
| rs676996 | 9:136146077 | rs8176719 | 9:136132908 | 0,84854 | Red blood cell count | Immunological | GWAS_ATLAS | 27863252 |
| rs676996 | 9:136146077 | - | - | - | Red blood cell count | - | Phenoscanner | 27863252 |
| rs676996 | 9:136146077 | rs687621 | 9:136137065 | 0,98307 | Salicylic acid and derivatives | Environmental | GWAS_ATLAS | 31015401 |
| rs676996 | 9:136146077 | rs687289 | 9:136137106 | 0,98728 | SELE - E-Selectin | Cell | GWAS_ATLAS | 28240269 |
| rs676996 | 9:136146077 | - | - | - | Self-reported deep venous thrombosis | - | Phenoscanner | UKBB |
| rs676996 | 9:136146077 | - | - | - | Self-reported high cholesterol | - | Phenoscanner | UKBB |
| rs676996 | 9:136146077 | - | - | - | Self-reported pulmonary embolism + or - dvt | - | Phenoscanner | UKBB |
| rs676996 | 9:136146077 | - | - | - | Serous invasive ovarian cancer | - | Phenoscanner | 28346442 |
| rs676996 | 9:136146077 | chr9:136139265 | 9:136139265 | 0,87713 | serum alkaline phosphatase levels | - | GWAS_catalog | 24094242 |
| rs676996 | 9:136146077 | chr9:136149229 | 9:136149229 | 0,97891 | Serum levels of soluble E-selectin with T1D | Immunological | GWAS_ATLAS | 19729612 |
| rs676996 | 9:136146077 | chr9:136139265 | 9:136139265 | 0,87713 | serum thyroid-stimulating hormone levels | - | GWAS_catalog | 25436638 |
| rs676996 | 9:136146077 | chr9:136149500 | 9:136149500 | 0,97891 | thrombosis | - | GWAS_catalog | 26908601 |
| rs676996 | 9:136146077 | chr9:136139265 | 9:136139265 | 0,87713 | thyroid hormone levels | - | GWAS_catalog | 23408906 |
| rs676996 | 9:136146077 | rs687621 | 9:136137065 | 0,98307 | Thyroid stimulating hormone | Endocrine | GWAS_ATLAS | 30367059 |
| rs676996 | 9:136146077 | rs687289 | 9:136137106 | 0,98728 | TIE1 - Tyrosine-protein kinase receptor Tie-1, soluble | Cell | GWAS_ATLAS | 28240269 |
| rs676996 | 9:136146077 | chr9:136142203 | 9:136142203 | 0,99152 | Total cholesterol | - | Phenoscanner | 24097068 |
| rs676996 | 9:136146077 | rs687621 | 9:136137065 | 0,98307 | Total cholesterol | Metabolic | GWAS_ATLAS | 29403010 |
| rs676996 | 9:136146077 | - | - | - | Treatment with cholesterol lowering medication | - | Phenoscanner | UKBB |
| rs676996 | 9:136146077 | - | - | - | Treatment with warfarin | - | Phenoscanner | UKBB |
| rs676996 | 9:136146077 | rs8176719 | 9:136132908 | 0,84854 | Trunk-trunk fat ratio | Metabolic | GWAS_ATLAS | 30664634 |
| rs676996 | 9:136146077 | rs687621 | 9:136137065 | 0,98307 | Type 2 Diabetes | Endocrine | GWAS_ATLAS | 30054458 |
| rs676996 | 9:136146077 | chr9:136149229 | 9:136149229 | 0,97891 | type 2 diabetes | - | GWAS_catalog | 30297969 |
| rs676996 | 9:136146077 | chr9:136139265 | 9:136139265 | 0,87713 | urinary metabolites (h-nmr features) | - | GWAS_catalog | 24586186 |
| rs676996 | 9:136146077 | - | - | - | Vascular or heart problems diagnosed by doctor: high blood pressure | - | Phenoscanner | UKBB |
| rs676996 | 9:136146077 | chr9:136149229 | 9:136149229 | 0,97891 | venous thromboembolism | - | GWAS_catalog | 19278955 |
| rs676996 | 9:136146077 | chr9:136142203 | 9:136142203 | 0,99152 | Venous thromboembolism | - | Phenoscanner | 22672568 |
| rs676996 | 9:136146077 | chr9:136146046 | 9:136146046 | 1 | Venous thrombosis | - | Phenoscanner | 22675575 |
| rs676996 | 9:136146077 | chr9:136142203 | 9:136142203 | 0,99152 | von Willebrand factor vWF | - | Phenoscanner | 23381943 |
| rs676996 | 9:136146077 | rs687289 | 9:136137106 | 0,98728 | VWF - von Willebrand factor | Cell | GWAS_ATLAS | 28240269 |
| rs676996 | 9:136146077 | chr9:136149229 | 9:136149229 | 0,97891 | vwf levels in ischaemic stroke and hyperhomocysteinaemia | - | GWAS_catalog | 28495826 |
| rs676996 | 9:136146077 | rs687621 | 9:136137065 | 0,98307 | Waist-hip ratio | Metabolic | GWAS_ATLAS | 30239722 |
| rs676996 | 9:136146077 | rs8176719 | 9:136132908 | 0,84854 | White blood cell count | Immunological | GWAS_ATLAS | 27863252 |
| rs676996 | 9:136146077 | - | - | - | White blood cell count | - | Phenoscanner | 27863252 |
| rs3012065 | 10:126737579 | - | - | - | Body Mass Index | Metabolic | GWAS_ATLAS | 30239722 |
| rs4910165 | 11:10674044 | rs6484437 | 11:10667275 | 0,93997 | 3rd ventricle | Neurological | GWAS_ATLAS | 31676860 |
| rs4910165 | 11:10674044 | rs10840457 | 11:10675738 | 0,95375 | arterial stiffness index | - | GWAS_catalog | 31235810 |
| rs4910165 | 11:10674044 | rs2052690 | 11:10664033 | 0,812 | asthma | - | GWAS_catalog | 30929738 |
| rs4910165 | 11:10674044 | rs2052690 | 11:10664033 | 0,812 | Asthma | Respiratory | GWAS_ATLAS | 30929738 |
| rs4910165 | 11:10674044 | rs2052690 | 11:10664033 | 0,812 | asthma (childhood onset) | - | GWAS_catalog | 30929738 |
| rs4910165 | 11:10674044 | rs4442541 | 11:10669172 | 0,96287 | brain region volumes | - | GWAS_catalog | 31676860 |
| rs4910165 | 11:10674044 | rs2052690 | 11:10664033 | 0,812 | Diastolic Blood Pressure | Cardiovascular | GWAS_ATLAS | 31427789 |
| rs4910165 | 11:10674044 | - | - | - | Diastolic blood pressure | - | Phenoscanner | UKBB |

| | | | | | | | | |
|---|---|---|---|---|---|---|---|---|
| rs4910165 | 11:10674044 | rs6484437 | 11:10667275 | 0,93997 | Fornix (column and body of fornix) fractional anisotropy | Neurological | GWAS_ATLAS | v: https://doi.org/10.1101/2 |
| rs4910165 | 11:10674044 | rs4909945 | 11:10673739 | 0,97672 | headache | - | GWAS_catalog | 29397368 |
| rs4910165 | 11:10674044 | rs4909945 | 11:10673739 | 0,97672 | Headache | - | Phenoscanner | 29397368 |
| rs4910165 | 11:10674044 | rs7940646 | 11:10669228 | 0,95369 | Headache | Neurological | GWAS_ATLAS | 31427789 |
| rs4910165 | 11:10674044 | rs6484437 | 11:10667275 | 0,93997 | Left lateral ventricle | Neurological | GWAS_ATLAS | 31676860 |
| rs4910165 | 11:10674044 | rs2052690 | 11:10664033 | 0,812 | Mean platelet volume | Immunological | GWAS_ATLAS | 27863252 |
| rs4910165 | 11:10674044 | - | - | - | Mean platelet volume | - | Phenoscanner | 27863252 |
| rs4910165 | 11:10674044 | rs7940646 | 11:10669228 | 0,95369 | Medication for pain relief, constipation, heartburn: ibuprofen | - | Phenoscanner | UKBB |
| rs4910165 | 11:10674044 | rs4909945 | 11:10673739 | 0,97672 | migraine | - | GWAS_catalog | 23793025 |
| rs4910165 | 11:10674044 | - | - | - | Migraine | - | Phenoscanner | 27182965 |
| rs4910165 | 11:10674044 | - | - | - | Pain type experienced in last month: headache | - | Phenoscanner | UKBB |
| rs4910165 | 11:10674044 | rs7940646 | 11:10669228 | 0,95369 | platelet aggregation | - | GWAS_catalog | 20526338 |
| rs4910165 | 11:10674044 | rs7940646 | 11:10669228 | 0,95369 | Platelet aggregation in response to ADP 5um10um | - | Phenoscanner | 20526338 |
| rs4910165 | 11:10674044 | rs6484437 | 11:10667275 | 0,93997 | red blood cell count | - | GWAS_catalog | 30595370 |
| rs4910165 | 11:10674044 | - | - | - | respiratory diseases | - | GWAS_catalog | 30595370 |
| rs4910165 | 11:10674044 | rs6484437 | 11:10667275 | 0,93997 | Right lateral ventricle | Neurological | GWAS_ATLAS | 31676860 |
| rs4910165 | 11:10674044 | rs11042906 | 11:10665235 | 0,84699 | systolic blood pressure | - | GWAS_catalog | 30595370 |
| rs4910165 | 11:10674044 | rs2052691 | 11:10664421 | 0,82479 | Systolic Blood Pressure | Cardiovascular | GWAS_ATLAS | 31427789 |
| rs4910165 | 11:10674044 | - | - | - | Systolic blood pressure | - | Phenoscanner | UKBB |
| rs4910165 | 11:10674044 | rs6484437 | 11:10667275 | 0,93997 | white matter microstructure (fractional anisotropy) | - | GWAS_catalog | 31666681 |
| rs10838738 | 11:47663049 | rs1317164 | 11:47419757 | 0,80578 | Age at menarche | Reproduction | GWAS_ATLAS | 31427789 |
| rs10838738 | 11:47663049 | rs11604680 | 11:47457539 | 0,83849 | alcohol consumption (drinks per week) | - | GWAS_catalog | 30643258 |
| rs10838738 | 11:47663049 | rs11604680 | 11:47457539 | 0,83849 | alcohol consumption (drinks per week) (mtag) | - | GWAS_catalog | 30643251 |
| rs10838738 | 11:47663049 | rs1317164 | 11:47419757 | 0,80578 | Alcohol intake | Psychiatric | GWAS_ATLAS | v: https://doi.org/10.1101/2 |
| rs10838738 | 11:47663049 | rs1317164 | 11:47419757 | 0,80578 | Alcohol intake frequency | Psychiatric | GWAS_ATLAS | 31427789 |
| rs10838738 | 11:47663049 | - | - | - | Alcohol intake frequency | - | Phenoscanner | UKBB |
| rs10838738 | 11:47663049 | - | - | - | Arm fat mass left | - | Phenoscanner | UKBB |
| rs10838738 | 11:47663049 | - | - | - | Arm fat mass right | - | Phenoscanner | UKBB |
| rs10838738 | 11:47663049 | - | - | - | Arm fat percentage left | - | Phenoscanner | UKBB |
| rs10838738 | 11:47663049 | - | - | - | Arm fat percentage right | - | Phenoscanner | UKBB |
| rs10838738 | 11:47663049 | rs1317164 | 11:47419757 | 0,80578 | Arm-arm fat ratio | Metabolic | GWAS_ATLAS | 30664634 |
| rs10838738 | 11:47663049 | rs12787646 | 11:47669665 | 0,95279 | bitter beverage consumption | - | GWAS_catalog | 31046077 |
| rs10838738 | 11:47663049 | - | - | - | Body fat percentage | - | Phenoscanner | UKBB |
| rs10838738 | 11:47663049 | - | - | - | Body mass index | - | Phenoscanner | 28892062 |
| rs10838738 | 11:47663049 | rs1064608 | 11:47640429 | 1 | body mass index | - | GWAS_catalog | 29273807 |
| rs10838738 | 11:47663049 | rs1317164 | 11:47419757 | 0,80578 | Body Mass Index | Metabolic | GWAS_ATLAS | 31427789 |
| rs10838738 | 11:47663049 | rs1064608 | 11:47640429 | 1 | Body mass index adjusted for physical activity | - | Phenoscanner | 28448500 |
| rs10838738 | 11:47663049 | - | - | - | Body mass index adjusted for smoking | - | Phenoscanner | 28443625 |
| rs10838738 | 11:47663049 | - | - | - | Body mass index females | - | Phenoscanner | 28892062 |
| rs10838738 | 11:47663049 | - | - | - | Body mass index in females | - | Phenoscanner | 25673413 |
| rs10838738 | 11:47663049 | - | - | - | Body mass index in males | - | Phenoscanner | 25673413 |
| rs10838738 | 11:47663049 | - | - | - | Body mass index in males greater than 50 years of age | - | Phenoscanner | 26426971 |
| rs10838738 | 11:47663049 | rs1064608 | 11:47640429 | 1 | cognitive performance | - | GWAS_catalog | 30038396 |
| rs10838738 | 11:47663049 | rs1317164 | 11:47419757 | 0,80578 | Comparative body size | Metabolic | GWAS_ATLAS | 31427789 |
| rs10838738 | 11:47663049 | - | - | - | Comparative body size at age 10 | - | Phenoscanner | UKBB |
| rs10838738 | 11:47663049 | rs1317164 | 11:47419757 | 0,80578 | Comparative height size | Skeletal | GWAS_ATLAS | 31427789 |
| rs10838738 | 11:47663049 | - | - | - | Comparative height size at age 10 | - | Phenoscanner | UKBB |
| rs10838738 | 11:47663049 | rs34467936 | 11:47915299 | 0,85403 | depressed affect | - | GWAS_catalog | 29942085 |
| rs10838738 | 11:47663049 | rs35184771 | 11:47475189 | 0,97413 | Depressive affect | Psychiatric | GWAS_ATLAS | 29942085 |
| rs10838738 | 11:47663049 | rs2290850 | 11:47807774 | 0,93602 | depressive symptoms | - | GWAS_catalog | 30643256 |
| rs10838738 | 11:47663049 | rs10838704 | 11:47424717 | 0,82229 | Depressive symptoms | Psychiatric | GWAS_ATLAS | 30643256 |
| rs10838738 | 11:47663049 | rs1317164 | 11:47419757 | 0,80578 | Ease of skin tanning | Dermatological | GWAS_ATLAS | 31427789 |
| rs10838738 | 11:47663049 | rs4752999 | 11:47428565 | 0,80968 | Fasting proinsulin | Metabolic | GWAS_ATLAS | 21873549 |
| rs10838738 | 11:47663049 | rs35184771 | 11:47475189 | 0,97413 | feeling lonely | - | GWAS_catalog | 29500382 |
| rs10838738 | 11:47663049 | rs4752801 | 11:47907641 | 0,85036 | feeling miserable | - | GWAS_catalog | 29500382 |
| rs10838738 | 11:47663049 | rs1317164 | 11:47419757 | 0,80578 | First PC of the four riskly behaviours | Activities | GWAS_ATLAS | v: https://doi.org/10.1101/2 |
| rs10838738 | 11:47663049 | - | - | - | Forced vital capacity | - | Phenoscanner | UKBB |
| rs10838738 | 11:47663049 | - | - | - | Forced vital capacity, best measure | - | Phenoscanner | UKBB |
| rs10838738 | 11:47663049 | rs11039355 | 11:47737501 | 0,9486 | Free thyroxine | Endocrine | GWAS_ATLAS | 30367059 |
| rs10838738 | 11:47663049 | rs11039355 | 11:47737501 | 0,9486 | free thyroxine concentration | - | GWAS_catalog | 30367059 |
| rs10838738 | 11:47663049 | - | - | - | Gene expression | - | Phenoscanner | 20935630 |
| rs10838738 | 11:47663049 | rs34923397 | 11:47769564 | 0,94432 | general cognitive ability | - | GWAS_catalog | 29844566 |
| rs10838738 | 11:47663049 | rs34467936 | 11:47915299 | 0,85403 | general factor of neuroticism | - | GWAS_catalog | 30867560 |
| rs10838738 | 11:47663049 | rs60956317 | 11:47825735 | 0,86143 | Granulocyte count | Immunological | GWAS_ATLAS | 27863252 |
| rs10838738 | 11:47663049 | rs35184771 | 11:47475189 | 0,97413 | Granulocyte percentage of myeloid white cells | Immunological | GWAS_ATLAS | 27863252 |
| rs10838738 | 11:47663049 | - | - | - | Granulocyte percentage of myeloid white cells | - | Phenoscanner | 27863252 |
| rs10838738 | 11:47663049 | - | - | - | Hand grip strength left | - | Phenoscanner | UKBB |
| rs10838738 | 11:47663049 | - | - | - | HDL cholesterol | - | Phenoscanner | 20686565 |
| rs10838738 | 11:47663049 | - | - | - | Height | - | Phenoscanner | 25282103 |
| rs10838738 | 11:47663049 | rs34958982 | 11:47547046 | 0,98706 | height | - | GWAS_catalog | 30595370 |
| rs10838738 | 11:47663049 | rs1317164 | 11:47419757 | 0,80578 | Height | Skeletal | GWAS_ATLAS | 31427789 |
| rs10838738 | 11:47663049 | - | - | - | High density lipoprotein | - | Phenoscanner | 24097068 |
| rs10838738 | 11:47663049 | rs10838704 | 11:47424717 | 0,82229 | High-density lipoprotein cholesterol | Metabolic | GWAS_ATLAS | 24097068 |
| rs10838738 | 11:47663049 | rs1317164 | 11:47419757 | 0,80578 | Hip circumference | Metabolic | GWAS_ATLAS | 31427789 |
| rs10838738 | 11:47663049 | - | - | - | Hip circumference | - | Phenoscanner | UKBB |
| rs10838738 | 11:47663049 | rs1317164 | 11:47419757 | 0,80578 | Impedance measures - Arm fat mass (left) | Metabolic | GWAS_ATLAS | 31427789 |
| rs10838738 | 11:47663049 | rs1317164 | 11:47419757 | 0,80578 | Impedance measures - Arm fat mass (right) | Metabolic | GWAS_ATLAS | 31427789 |
| rs10838738 | 11:47663049 | rs1317164 | 11:47419757 | 0,80578 | Impedance measures - Arm fat percentage (left) | Metabolic | GWAS_ATLAS | 31427789 |
| rs10838738 | 11:47663049 | rs1317164 | 11:47419757 | 0,80578 | Impedance measures - Arm fat percentage (right) | Metabolic | GWAS_ATLAS | 31427789 |
| rs10838738 | 11:47663049 | rs1317164 | 11:47419757 | 0,80578 | Impedance measures - Body fat percentage | Metabolic | GWAS_ATLAS | 31427789 |
| rs10838738 | 11:47663049 | rs1317164 | 11:47419757 | 0,80578 | Impedance measures - Body mass index (BMI) | Metabolic | GWAS_ATLAS | 31427789 |
| rs10838738 | 11:47663049 | rs1317164 | 11:47419757 | 0,80578 | Impedance measures - Leg fat mass (left) | Metabolic | GWAS_ATLAS | 31427789 |
| rs10838738 | 11:47663049 | rs1317164 | 11:47419757 | 0,80578 | Impedance measures - Leg fat mass (right) | Metabolic | GWAS_ATLAS | 31427789 |
| rs10838738 | 11:47663049 | rs1317164 | 11:47419757 | 0,80578 | Impedance measures - Leg fat percentage (left) | Metabolic | GWAS_ATLAS | 31427789 |
| rs10838738 | 11:47663049 | rs1317164 | 11:47419757 | 0,80578 | Impedance measures - Leg fat percentage (right) | Metabolic | GWAS_ATLAS | 31427789 |
| rs10838738 | 11:47663049 | rs1317164 | 11:47419757 | 0,80578 | Impedance measures - Trunk fat mass | Metabolic | GWAS_ATLAS | 31427789 |
| rs10838738 | 11:47663049 | rs1317164 | 11:47419757 | 0,80578 | Impedance measures - Trunk fat percentage | Metabolic | GWAS_ATLAS | 31427789 |
| rs10838738 | 11:47663049 | rs3781625 | 11:47443080 | 0,81765 | Impedance measures - Trunk fat-free mass | Metabolic | GWAS_ATLAS | 31427789 |
| rs10838738 | 11:47663049 | rs11600581 | 11:47448497 | 0,80145 | Impedance measures - Trunk predicted mass | Metabolic | GWAS_ATLAS | 31427789 |
| rs10838738 | 11:47663049 | rs1317164 | 11:47419757 | 0,80578 | Impedance measures - Whole body fat mass | Metabolic | GWAS_ATLAS | 31427789 |
| rs10838738 | 11:47663049 | rs11605348 | 11:47606483 | 0,94852 | insomnia | - | GWAS_catalog | 30804565 |
| rs10838738 | 11:47663049 | rs11605348 | 11:47606483 | 0,94852 | intelligence | - | GWAS_catalog | 29942086 |
| rs10838738 | 11:47663049 | rs1317164 | 11:47419757 | 0,80578 | Intelligence | Cognitive | GWAS_ATLAS | 30038396 |
| rs10838738 | 11:47663049 | - | - | - | Leg fat mass left | - | Phenoscanner | UKBB |
| rs10838738 | 11:47663049 | - | - | - | Leg fat mass right | - | Phenoscanner | UKBB |
| rs10838738 | 11:47663049 | - | - | - | Leg fat percentage left | - | Phenoscanner | UKBB |
| rs10838738 | 11:47663049 | - | - | - | Leg fat percentage right | - | Phenoscanner | UKBB |
| rs10838738 | 11:47663049 | rs2290850 | 11:47807774 | 0,93602 | life satisfaction | - | GWAS_catalog | 30643256 |
| rs10838738 | 11:47663049 | rs35184771 | 11:47475189 | 0,97413 | Life satisfaction | Activities | GWAS_ATLAS | 30643256 |
| rs10838738 | 11:47663049 | - | - | - | log Proinsulin | - | Phenoscanner | 21873549 |
| rs10838738 | 11:47663049 | rs35184771 | 11:47475189 | 0,97413 | Loneliness | Psychiatric | GWAS_ATLAS | 31427789 |
| rs10838738 | 11:47663049 | rs11039265 | 11:47523214 | 0,96582 | loneliness | - | GWAS_catalog | 31518406 |
| rs10838738 | 11:47663049 | rs11605348 | 11:47606483 | 0,94852 | loneliness (mtag) | - | GWAS_catalog | 29970889 |
| rs10838738 | 11:47663049 | - | - | - | Loneliness, isolation | - | Phenoscanner | UKBB |
| rs10838738 | 11:47663049 | rs35184771 | 11:47475189 | 0,97413 | Miserableness | Psychiatric | GWAS_ATLAS | 31427789 |
| rs10838738 | 11:47663049 | - | - | - | Miserableness | - | Phenoscanner | UKBB |
| rs10838738 | 11:47663049 | rs35184771 | 11:47475189 | 0,97413 | Monocyte percentage of white cells | Immunological | GWAS_ATLAS | 27863252 |
| rs10838738 | 11:47663049 | - | - | - | Monocyte percentage of white cells | - | Phenoscanner | 27863252 |
| rs10838738 | 11:47663049 | rs11039389 | 11:47796062 | 0,92779 | neurociticism | - | GWAS_catalog | 29500382 |
| rs10838738 | 11:47663049 | rs11039389 | 11:47796062 | 0,92779 | neuroticism | - | GWAS_catalog | 29942085 |
| rs10838738 | 11:47663049 | rs1317164 | 11:47419757 | 0,80578 | Neuroticism | Psychiatric | GWAS_ATLAS | 30643256 |
| rs10838738 | 11:47663049 | - | - | - | Neuroticism score | - | Phenoscanner | UKBB |
| rs10838738 | 11:47663049 | rs12787112 | 11:47687147 | 0,96136 | Neutrophil count | Immunological | GWAS_ATLAS | 27863252 |
| rs10838738 | 11:47663049 | rs1317164 | 11:47419757 | 0,80578 | Platelet count | Immunological | GWAS_ATLAS | 27863252 |
| rs10838738 | 11:47663049 | - | - | - | Platelet count | - | Phenoscanner | 27863252 |
| rs10838738 | 11:47663049 | rs10838724 | 11:47527052 | 0,82203 | Plateletcrit | Immunological | GWAS_ATLAS | 27863252 |
| rs10838738 | 11:47663049 | rs2290850 | 11:47807774 | 0,93602 | positive affect | - | GWAS_catalog | 30643256 |
| rs10838738 | 11:47663049 | rs1317164 | 11:47419757 | 0,80578 | Possitive affect | Activities | GWAS_ATLAS | 30643256 |
| rs10838738 | 11:47663049 | rs12363232 | 11:47623890 | 1 | risk-taking tendency (4-domain principal component model) | - | GWAS_catalog | 30643258 |
| rs10838738 | 11:47663049 | rs4752845 | 11:47539697 | 0,98706 | serum metabolite levels | - | GWAS_catalog | 31636271 |
| rs10838738 | 11:47663049 | rs4752845 | 11:47539697 | 0,98706 | serum metabolite levels (cms) | - | GWAS_catalog | 31636271 |
| rs10838738 | 11:47663049 | rs1317164 | 11:47419757 | 0,80578 | Sitting height | Skeletal | GWAS_ATLAS | 31427789 |
| rs10838738 | 11:47663049 | - | - | - | Sitting height | - | Phenoscanner | UKBB |
| rs10838738 | 11:47663049 | rs11039348 | 11:47728617 | 0,9486 | smoking initiation (ever regular vs never regular) (mtag) | - | GWAS_catalog | 30643251 |
| rs10838738 | 11:47663049 | rs7927771 | 11:47781306 | 0,94432 | subjective well-being | - | GWAS_catalog | 27089181 |
| rs10838738 | 11:47663049 | rs1317164 | 11:47419757 | 0,80578 | Suffer from nerves | Psychiatric | GWAS_ATLAS | 31427789 |
| rs10838738 | 11:47663049 | - | - | - | Suffer from nerves | - | Phenoscanner | UKBB |
| rs10838738 | 11:47663049 | rs12787112 | 11:47687147 | 0,96136 | suffering from nerves | - | GWAS_catalog | 29500382 |
| rs10838738 | 11:47663049 | rs12787112 | 11:47687147 | 0,96136 | Sum basophil neutrophil count | Immunological | GWAS_ATLAS | 27863252 |
| rs10838738 | 11:47663049 | rs12787112 | 11:47687147 | 0,96136 | Sum neutrophil eosinophil count | Immunological | GWAS_ATLAS | 27863252 |
| rs10838738 | 11:47663049 | - | - | - | Trunk fat mass | - | Phenoscanner | UKBB |
| rs10838738 | 11:47663049 | - | - | - | Trunk fat percentage | - | Phenoscanner | UKBB |
| rs10838738 | 11:47663049 | - | - | - | Trunk fat-free mass | - | Phenoscanner | UKBB |
| rs10838738 | 11:47663049 | - | - | - | Trunk predicted mass | - | Phenoscanner | UKBB |
| rs10838738 | 11:47663049 | rs35184771 | 11:47475189 | 0,97413 | Usual side of head for mobile phone use | Activities | GWAS_ATLAS | 31427789 |
| rs10838738 | 11:47663049 | rs1317164 | 11:47419757 | 0,80578 | Usual walking pace | Activities | GWAS_ATLAS | 31427789 |
| rs10838738 | 11:47663049 | rs35184771 | 11:47475189 | 0,97413 | Verbal-numerical reasoning | Cognitive | GWAS_ATLAS | 29844566 |
| rs10838738 | 11:47663049 | rs1317164 | 11:47419757 | 0,80578 | Waist circumference | Metabolic | GWAS_ATLAS | 31427789 |
| rs10838738 | 11:47663049 | - | - | - | Waist circumference | - | Phenoscanner | UKBB |
| rs10838738 | 11:47663049 | rs3781625 | 11:47443080 | 0,81765 | Waist-hip ratio | Metabolic | GWAS_ATLAS | 30239722 |
| rs10838738 | 11:47663049 | rs1317164 | 11:47419757 | 0,80578 | Well-being spectrum | Psychiatric | GWAS_ATLAS | 30643256 |
| rs10838738 | 11:47663049 | rs12787646 | 11:47669665 | 0,95279 | White blood cell count | Immunological | GWAS_ATLAS | 27863252 |
| rs10838738 | 11:47663049 | rs12364432 | 11:47902883 | 0,92779 | white blood cell count | - | GWAS_catalog | 30595370 |
| rs10838738 | 11:47663049 | - | - | - | Whole body fat mass | - | Phenoscanner | UKBB |
| rs10838738 | 11:47663049 | rs4752856 | 11:47648042 | 1 | worry | - | GWAS_catalog | 29942085 |
| rs10838738 | 11:47663049 | rs35184771 | 11:47475189 | 0,97413 | Worry | Psychiatric | GWAS_ATLAS | 29942085 |
| rs2212450 | 11:112826867 | rs2212450 | 11:112826867 | 1 | Age first had sexual intercourse | Reproduction | GWAS_ATLAS | 31427789 |
| rs2212450 | 11:112826867 | rs2212450 | 11:112826867 | 1 | body mass index | - | GWAS_catalog | 30239722 |

| | | | | | | | | |
|---|---|---|---|---|---|---|---|---|
| rs2212450 | 11:112826867 | rs2212450 | 11:112826867 | 1 | Body Mass Index | Metabolic | GWAS_ATLAS | 30239722 |
| rs2212450 | 11:112826867 | rs2212450 | 11:112826867 | 1 | Depressive symptoms | Psychiatric | GWAS_ATLAS | 29942085 |
| rs2212450 | 11:112826867 | rs2212450 | 11:112826867 | 1 | Ever vs never smokers | Psychiatric | GWAS_ATLAS | 31427789 |
| rs2212450 | 11:112826867 | rs2212450 | 11:112826867 | 1 | First PC of the four riskly behaviours | Activities | GWAS_ATLAS | v: https://doi.org/10.1101/2 |
| rs2212450 | 11:112826867 | rs2212450 | 11:112826867 | 1 | Neuroticism | Psychiatric | GWAS_ATLAS | 31427789 |
| rs2212450 | 11:112826867 | rs2212450 | 11:112826867 | 1 | Number of sexual partners | Reproduction | GWAS_ATLAS | v: https://doi.org/10.1101/2 |
| rs2212450 | 11:112826867 | rs2212450 | 11:112826867 | 1 | Past tobacco smoking | Psychiatric | GWAS_ATLAS | 31427789 |
| rs2212450 | 11:112826867 | rs2212450 | 11:112826867 | 1 | Tense | Psychiatric | GWAS_ATLAS | 31427789 |
| rs2212450 | 11:112826867 | rs2212450 | 11:112826867 | 1 | Tobacco smoking | Psychiatric | GWAS_ATLAS | 31427789 |
| rs2212450 | 11:112826867 | rs2212450 | 11:112826867 | 1 | Worry | Psychiatric | GWAS_ATLAS | 29942085 |
| rs4345978 | 11:123009751 | rs113451750 | 11:123004938 | 0,8455 | Pulse rate | Cardiovascular | GWAS_ATLAS | 31427789 |
| rs4345978 | 11:123009751 | rs12283343 | 11:123008349 | 0,96727 | Pulse rate | - | Phenoscanner | UKBB |
| rs4345978 | 11:123009751 | rs113451750 | 11:123004938 | 0,8455 | Resting heart rate | Cardiovascular | GWAS_ATLAS | 30940143 |
| rs4579999 | 12:24439661 | rs10842304 | 12:24424791 | 0,80226 | FEV1/FVC ratio | Respiratory | GWAS_ATLAS | 30804560 |
| rs4579999 | 12:24439661 | rs563110 | 12:24438591 | 0,92189 | lung function (fev1/fvc) | - | GWAS_catalog | 30595370 |
| rs4579999 | 12:24439661 | - | - | - | PEF | Respiratory | GWAS_ATLAS | 30804560 |
| rs111235435 | 12:24757629 | rs11047526 | 12:24759806 | 0,98359 | calcium levels | - | GWAS_catalog | 25972035 |
| rs111235435 | 12:24757629 | rs7969321 | 12:24761558 | 0,98359 | Heart rate variability | Cardiovascular | GWAS_ATLAS | 28613276 |
| rs111235435 | 12:24757629 | rs1386639 | 12:24768847 | 0,98774 | Heart rate variability pvRSAHF | - | Phenoscanner | 28613276 |
| rs111235435 | 12:24757629 | rs113807524 | 12:24757748 | 0,9918 | Height | Skeletal | GWAS_ATLAS | 31427789 |
| rs111235435 | 12:24757629 | - | - | - | Pulse rate | Cardiovascular | GWAS_ATLAS | 31427789 |
| rs111235435 | 12:24757629 | - | - | - | Pulse rate | - | Phenoscanner | UKBB |
| rs111235435 | 12:24757629 | rs11047547 | 12:24797562 | 0,83222 | Resting heart rate | Cardiovascular | GWAS_ATLAS | 30940143 |
| rs13632 | 12:48177238 | rs4760742 | 12:48172058 | 0,9845 | Heel bone mineral density | Skeletal | GWAS_ATLAS | 30598549 |
| rs13632 | 12:48177238 | rs4760636 | 12:48173352 | 0,97401 | urate levels | - | GWAS_catalog | 23263486 |
| rs920778 | 12:54360232 | rs2366150 | 12:54353523 | 0,93526 | Heel bone mineral density | Skeletal | GWAS_ATLAS | 30048462 |
| rs920778 | 12:54360232 | rs2002472 | 12:54355209 | 0,99062 | Impedance measures - Impedance of arm (left) | Metabolic | GWAS_ATLAS | 31427789 |
| rs920778 | 12:54360232 | - | - | - | Impedance of arm left | - | Phenoscanner | UKBB |
| rs920778 | 12:54360232 | rs2366150 | 12:54353523 | 0,93526 | Waist-hip ratio | Metabolic | GWAS_ATLAS | 30239722 |
| rs11176001 | 12:66409367 | - | - | - | fev1 | - | GWAS_catalog | 30804560 |
| rs11176001 | 12:66409367 | rs74097857 | 12:66393756 | 0,97342 | FEV2 | Respiratory | GWAS_ATLAS | 30804560 |
| rs11176001 | 12:66409367 | rs74097857 | 12:66393756 | 0,97342 | FVC | Respiratory | GWAS_ATLAS | 30804560 |
| rs11176001 | 12:66409367 | - | - | - | lung function (fvc) | - | GWAS_catalog | 30804560 |
| rs11176001 | 12:66409367 | - | - | - | peak expiratory flow | - | GWAS_catalog | 30804560 |
| rs11176001 | 12:66409367 | rs74097857 | 12:66393756 | 0,97342 | PEF | Respiratory | GWAS_ATLAS | 30804560 |
| rs2555004 | 12:114686645 | rs1920568 | 12:114669732 | 0,81823 | FEV2 | Respiratory | GWAS_ATLAS | 30804560 |
| rs2555004 | 12:114686645 | rs1920568 | 12:114669732 | 0,81823 | FVC | Respiratory | GWAS_ATLAS | 30804560 |
| rs2555004 | 12:114686645 | rs1270884 | 12:114685571 | 0,93807 | Prostate cancer | - | Phenoscanner | 23535732 |
| rs2555004 | 12:114686645 | rs1270884 | 12:114685571 | 0,93807 | prostate cancer | - | GWAS_catalog | 29892016 |
| rs11635984 | 15:33012232 | rs11633862 | 15:33008359 | 0,85053 | Benign neoplasm of colon, rectum, anus and anal canal | Neoplasms | GWAS_ATLAS | 31427789 |
| rs17293632 | 15:67442596 | rs72743461 | 15:67441750 | 1 | Adrenergics, inhalants | Environmental | GWAS_ATLAS | 31015401 |
| rs17293632 | 15:67442596 | - | - | - | Allergic disease | - | Phenoscanner | 29083406 |
| rs17293632 | 15:67442596 | rs72743461 | 15:67441750 | 1 | allergic disease (asthma, hay fever or eczema) | - | GWAS_catalog | 31361310 |
| rs17293632 | 15:67442596 | rs56375023 | 15:67448363 | 0,98831 | Allergic disease asthma hay fever or eczema | - | Phenoscanner | 29083406 |
| rs17293632 | 15:67442596 | - | - | - | allergic rhinitis | - | GWAS_catalog | 31361310 |
| rs17293632 | 15:67442596 | - | - | - | Asthma | - | Phenoscanner | 29273806 |
| rs17293632 | 15:67442596 | rs72743461 | 15:67441750 | 1 | asthma | - | GWAS_catalog | 31361310 |
| rs17293632 | 15:67442596 | rs72743461 | 15:67441750 | 1 | Asthma | Respiratory | GWAS_ATLAS | 31427789 |
| rs17293632 | 15:67442596 | rs72743461 | 15:67441750 | 1 | asthma (adult onset) | - | GWAS_catalog | 30929738 |
| rs17293632 | 15:67442596 | rs72743461 | 15:67441750 | 1 | asthma (childhood onset) | - | GWAS_catalog | 30929738 |
| rs17293632 | 15:67442596 | rs72743461 | 15:67441750 | 1 | asthma (moderate or severe) | - | GWAS_catalog | 30552067 |
| rs17293632 | 15:67442596 | rs72743461 | 15:67441750 | 1 | asthma onset (childhood vs adult) | - | GWAS_catalog | 30929738 |
| rs17293632 | 15:67442596 | rs56062135 | 15:67455630 | 0,98823 | asthma or allergic disease (pleiotropy) | - | GWAS_catalog | 29785011 |
| rs17293632 | 15:67442596 | rs72743461 | 15:67441750 | 1 | Asthma, hay fever or eczema | Respiratory | GWAS_ATLAS | 29083406 |
| rs17293632 | 15:67442596 | - | - | - | chronic inflammatory diseases (ankylosing spondylitis, crohn's disease, psoriasis, primary sclerosing cholangitis, ulcerative colitis) (pleiotropy) | - | GWAS_catalog | 26974007 |
| rs17293632 | 15:67442596 | rs72743461 | 15:67441750 | 1 | Coronary artery disease | Cardiovascular | GWAS_ATLAS | 26343387 |
| rs17293632 | 15:67442596 | - | - | - | Coronary artery disease | - | Phenoscanner | 26343387 |
| rs17293632 | 15:67442596 | rs72743461 | 15:67441750 | 1 | coronary artery disease | - | GWAS_catalog | 29212778 |
| rs17293632 | 15:67442596 | rs72743461 | 15:67441750 | 1 | coronary artery disease (myocardial infarction, percutaneous transluminal coronary angioplasty, coronary artery bypass grafting, angina or chromic ischemic heart disease) | - | GWAS_catalog | 28714975 |
| rs17293632 | 15:67442596 | - | - | - | crohn's disease | - | GWAS_catalog | 21102463 |
| rs17293632 | 15:67442596 | rs72743461 | 15:67441750 | 1 | Crohn's Disease | Gastrointestinal | GWAS_ATLAS | 28067908 |
| rs17293632 | 15:67442596 | - | - | - | Crohns disease | - | Phenoscanner | 21102463 |
| rs17293632 | 15:67442596 | - | - | - | Doctor diagnosed asthma | - | Phenoscanner | UKBB |
| rs17293632 | 15:67442596 | rs72743461 | 15:67441750 | 1 | Doctor diagnosed hayfever or allergic rhinitis | Respiratory | GWAS_ATLAS | 31427789 |
| rs17293632 | 15:67442596 | rs56375023 | 15:67448363 | 0,98831 | eczema | - | GWAS_catalog | 30595370 |
| rs17293632 | 15:67442596 | rs72743461 | 15:67441750 | 1 | Eosinophil count | Immunological | GWAS_ATLAS | 27863252 |
| rs17293632 | 15:67442596 | - | - | - | Eosinophil count | - | Phenoscanner | 27863252 |
| rs17293632 | 15:67442596 | - | - | - | eosinophil counts | - | GWAS_catalog | 30595370 |
| rs17293632 | 15:67442596 | rs72743461 | 15:67441750 | 1 | Eosinophil percentage of granulocytes | Immunological | GWAS_ATLAS | 27863252 |
| rs17293632 | 15:67442596 | - | - | - | Eosinophil percentage of granulocytes | - | Phenoscanner | 27863252 |
| rs17293632 | 15:67442596 | rs72743461 | 15:67441750 | 1 | Eosinophil percentage of white cells | Immunological | GWAS_ATLAS | 27863252 |
| rs17293632 | 15:67442596 | - | - | - | Eosinophil percentage of white cells | - | Phenoscanner | 27863252 |
| rs17293632 | 15:67442596 | rs72743461 | 15:67441750 | 1 | FEV1/FVC ratio | Respiratory | GWAS_ATLAS | 30804560 |
| rs17293632 | 15:67442596 | rs72743461 | 15:67441750 | 1 | Glucocorticoids | Environmental | GWAS_ATLAS | 31015401 |
| rs17293632 | 15:67442596 | - | - | - | hay fever and/or eczema | - | GWAS_catalog | 31361310 |
| rs17293632 | 15:67442596 | rs72743461 | 15:67441750 | 1 | Hayfever, allergic rhinitis or eczema | Respiratory | GWAS_ATLAS | 31427789 |
| rs17293632 | 15:67442596 | - | - | - | Hayfever, allergic rhinitis or eczema | - | Phenoscanner | UKBB |
| rs17293632 | 15:67442596 | rs72743461 | 15:67441750 | 1 | Hayfever/Allergic Rhinitis | Respiratory | GWAS_ATLAS | 31427789 |
| rs17293632 | 15:67442596 | - | - | - | inflammatory bowel disease | - | GWAS_catalog | 23128233 |
| rs17293632 | 15:67442596 | - | - | - | Inflammatory bowel disease | - | Phenoscanner | 23128233 |
| rs17293632 | 15:67442596 | rs72743461 | 15:67441750 | 1 | Inflammatory Bowel Disease | Gastrointestinal | GWAS_ATLAS | 28067908 |
| rs17293632 | 15:67442596 | rs72743477 | 15:67464291 | 0,89579 | lung function (fev1/fvc) | - | GWAS_catalog | 30595370 |
| rs17293632 | 15:67442596 | rs72743461 | 15:67441750 | 1 | medication use (adrenergics, inhalants) | - | GWAS_catalog | 31015401 |
| rs17293632 | 15:67442596 | rs72743461 | 15:67441750 | 1 | medication use (glucocorticoids) | - | GWAS_catalog | 31015401 |
| rs17293632 | 15:67442596 | rs72743461 | 15:67441750 | 1 | myocardial infarction | - | GWAS_catalog | 26343387 |
| rs17293632 | 15:67442596 | - | - | - | No blood clot, bronchitis, emphysema, asthma, rhinitis, eczema or allergy diagnosed by doctor | - | Phenoscanner | UKBB |
| rs17293632 | 15:67442596 | rs72743477 | 15:67464291 | 0,89579 | pediatric autoimmune diseases | - | GWAS_catalog | 26301688 |
| rs17293632 | 15:67442596 | rs72743461 | 15:67441750 | 1 | respiratory diseases | - | GWAS_catalog | 30595370 |
| rs17293632 | 15:67442596 | rs17228058 | 15:67450305 | 0,98244 | Self reported allergy | - | Phenoscanner | 23817569 |
| rs17293632 | 15:67442596 | rs17228058 | 15:67450305 | 0,98244 | self-reported allergy | - | GWAS_catalog | 23817569 |
| rs17293632 | 15:67442596 | - | - | - | Self-reported asthma | - | Phenoscanner | UKBB |
| rs17293632 | 15:67442596 | - | - | - | Self-reported hayfever or allergic rhinitis | - | Phenoscanner | UKBB |
| rs17293632 | 15:67442596 | rs72743461 | 15:67441750 | 1 | Sum eosinophil basophil count | Immunological | GWAS_ATLAS | 27863252 |
| rs17293632 | 15:67442596 | - | - | - | Sum eosinophil basophil counts | - | Phenoscanner | 27863252 |
| rs17293632 | 15:67442596 | rs56062135 | 15:67455630 | 0,98823 | thyroid cancer | - | GWAS_catalog | 28195142 |
| rs17293632 | 15:67442596 | rs56062135 | 15:67455630 | 0,98823 | Thyroid cancer | - | Phenoscanner | 28195142 |
| rs17293632 | 15:67442596 | - | - | - | Treatment with ventolin 100micrograms inhaler | - | Phenoscanner | UKBB |
| rs17293632 | 15:67442596 | - | - | - | ulcerative colitis | - | GWAS_catalog | 26192919 |
| rs17293632 | 15:67442596 | rs72743461 | 15:67441750 | 1 | Ulcerative colitis | Gastrointestinal | GWAS_ATLAS | 28067908 |
| rs17293632 | 15:67442596 | - | - | - | Ulcerative colitis | - | Phenoscanner | 28067908 |
| rs17293632 | 15:67442596 | rs72743461 | 15:67441750 | 1 | Ventolin 100micrograms inhaler | Activities | GWAS_ATLAS | 31427789 |
| rs17293632 | 15:67442596 | - | - | - | Wheeze or whistling in the chest in last year | - | Phenoscanner | UKBB |
| rs17293632 | 15:67442596 | rs72743461 | 15:67441750 | 1 | Wheeze/whistling | Respiratory | GWAS_ATLAS | 31427789 |
| rs12594232 | 15:68469402 | rs8040883 | 15:68517211 | 0,87893 | Height | Skeletal | GWAS_ATLAS | 30124842 |
| rs4843407 | 16:86629047 | rs16941554 | 16:86622436 | 0,90796 | Height | Skeletal | GWAS_ATLAS | 30124842 |
| rs78378222 | 17:7571752 | - | - | - | appendicular lean mass | - | GWAS_catalog | 31761296 |
| rs78378222 | 17:7571752 | - | - | - | Arm fat-free mass left | - | Phenoscanner | UKBB |
| rs78378222 | 17:7571752 | - | - | - | Arm fat-free mass right | - | Phenoscanner | UKBB |
| rs78378222 | 17:7571752 | - | - | - | Arm predicted mass left | - | Phenoscanner | UKBB |
| rs78378222 | 17:7571752 | - | - | - | Arm predicted mass right | - | Phenoscanner | UKBB |
| rs78378222 | 17:7571752 | - | - | - | Basal cell carcinoma | - | Phenoscanner | 21946351 |
| rs78378222 | 17:7571752 | - | - | - | basal cell carcinoma | - | GWAS_catalog | 27539887 |
| rs78378222 | 17:7571752 | - | - | - | Basal cell carcinoma | Neoplasms | GWAS_ATLAS | 31427789 |
| rs78378222 | 17:7571752 | - | - | - | Basal metabolic rate | - | Phenoscanner | UKBB |
| rs78378222 | 17:7571752 | - | - | - | birth weight | - | GWAS_catalog | 31043758 |
| rs78378222 | 17:7571752 | - | - | - | Birth weight | Metabolic | GWAS_ATLAS | 31427789 |
| rs78378222 | 17:7571752 | - | - | - | Birth weight | - | Phenoscanner | UKBB |
| rs78378222 | 17:7571752 | - | - | - | cancer | - | GWAS_catalog | 28346444 |
| rs78378222 | 17:7571752 | - | - | - | Cause of death: brain, unspecified | - | Phenoscanner | UKBB |
| rs78378222 | 17:7571752 | - | - | - | Comparative height size | Skeletal | GWAS_ATLAS | 31427789 |
| rs78378222 | 17:7571752 | - | - | - | Comparative height size at age 10 | - | Phenoscanner | UKBB |
| rs78378222 | 17:7571752 | - | - | - | Cutaneous squamous cell carcinoma skin cancer | - | Phenoscanner | 21946351 |
| rs78378222 | 17:7571752 | - | - | - | diastolic blood pressure | - | GWAS_catalog | 30578418 |
| rs78378222 | 17:7571752 | - | - | - | Diastolic Blood Pressure | Cardiovascular | GWAS_ATLAS | 31427789 |
| rs78378222 | 17:7571752 | - | - | - | Diastolic blood pressure | - | Phenoscanner | UKBB |
| rs78378222 | 17:7571752 | - | - | - | Ever had hysterectomy (womb removed) (female) | Reproduction | GWAS_ATLAS | 31427789 |
| rs78378222 | 17:7571752 | - | - | - | fat-free mass | - | GWAS_catalog | 30593698 |
| rs78378222 | 17:7571752 | - | - | - | Forced expiratory volume in 1-second | - | Phenoscanner | UKBB |
| rs78378222 | 17:7571752 | - | - | - | Forced expiratory volume in 1-second, best measure | - | Phenoscanner | UKBB |
| rs78378222 | 17:7571752 | - | - | - | Forced vital capacity | - | Phenoscanner | UKBB |
| rs78378222 | 17:7571752 | - | - | - | Forced vital capacity, best measure | - | Phenoscanner | UKBB |
| rs78378222 | 17:7571752 | - | - | - | Glioblastoma | - | Phenoscanner | 28346443 |
| rs78378222 | 17:7571752 | - | - | - | glioblastoma | - | GWAS_catalog | 29743610 |
| rs78378222 | 17:7571752 | - | - | - | glioblastoma (age-stratified) | - | GWAS_catalog | 30152087 |
| rs78378222 | 17:7571752 | - | - | - | Glioma | - | Phenoscanner | 28346443 |
| rs78378222 | 17:7571752 | - | - | - | glioma | - | GWAS_catalog | 29743610 |
| rs78378222 | 17:7571752 | - | - | - | Height | Skeletal | GWAS_ATLAS | 31427789 |
| rs78378222 | 17:7571752 | - | - | - | Height | - | Phenoscanner | UKBB |
| rs78378222 | 17:7571752 | - | - | - | Hip circumference | Metabolic | GWAS_ATLAS | 31427789 |
| rs78378222 | 17:7571752 | - | - | - | Hip circumference | - | Phenoscanner | UKBB |
| rs78378222 | 17:7571752 | - | - | - | Impedance measures - Arm fat-free mass (left) | Metabolic | GWAS_ATLAS | 31427789 |
| rs78378222 | 17:7571752 | - | - | - | Impedance measures - Arm fat-free mass (right) | Metabolic | GWAS_ATLAS | 31427789 |
| rs78378222 | 17:7571752 | - | - | - | Impedance measures - Arm predicted mass (left) | Metabolic | GWAS_ATLAS | 31427789 |

| | | | | | | | | |
|---|---|---|---|---|---|---|---|---|
| rs78378222 | 17:7571752 | - | - | - | Impedance measures - Arm predicted mass (right) | Metabolic | GWAS_ATLAS | 31427789 |
| rs78378222 | 17:7571752 | - | - | - | Impedance measures - Basal metabolic rate | Metabolic | GWAS_ATLAS | 31427789 |
| rs78378222 | 17:7571752 | - | - | - | Impedance measures - Impedance of arm (left) | Metabolic | GWAS_ATLAS | 31427789 |
| rs78378222 | 17:7571752 | - | - | - | Impedance measures - Impedance of arm (right) | Metabolic | GWAS_ATLAS | 31427789 |
| rs78378222 | 17:7571752 | - | - | - | Impedance measures - Impedance of leg (left) | Metabolic | GWAS_ATLAS | 31427789 |
| rs78378222 | 17:7571752 | - | - | - | Impedance measures - Impedance of leg (right) | Metabolic | GWAS_ATLAS | 31427789 |
| rs78378222 | 17:7571752 | - | - | - | Impedance measures - Impedance of whole body | Metabolic | GWAS_ATLAS | 31427789 |
| rs78378222 | 17:7571752 | - | - | - | Impedance measures - Leg fat-free mass (left) | Metabolic | GWAS_ATLAS | 31427789 |
| rs78378222 | 17:7571752 | - | - | - | Impedance measures - Leg fat-free mass (right) | Metabolic | GWAS_ATLAS | 31427789 |
| rs78378222 | 17:7571752 | - | - | - | Impedance measures - Leg predicted mass (left) | Metabolic | GWAS_ATLAS | 31427789 |
| rs78378222 | 17:7571752 | - | - | - | Impedance measures - Leg predicted mass (right) | Metabolic | GWAS_ATLAS | 31427789 |
| rs78378222 | 17:7571752 | - | - | - | Impedance measures - Trunk fat-free mass | Metabolic | GWAS_ATLAS | 31427789 |
| rs78378222 | 17:7571752 | - | - | - | Impedance measures - Trunk predicted mass | Metabolic | GWAS_ATLAS | 31427789 |
| rs78378222 | 17:7571752 | - | - | - | Impedance measures - Weight | Metabolic | GWAS_ATLAS | 31427789 |
| rs78378222 | 17:7571752 | - | - | - | Impedance measures - Whole body fat-free mass | Metabolic | GWAS_ATLAS | 31427789 |
| rs78378222 | 17:7571752 | - | - | - | Impedance measures - Whole body water mass | Metabolic | GWAS_ATLAS | 31427789 |
| rs78378222 | 17:7571752 | - | - | - | Impedance of arm left | - | Phenoscanner | UKBB |
| rs78378222 | 17:7571752 | - | - | - | Impedance of arm right | - | Phenoscanner | UKBB |
| rs78378222 | 17:7571752 | - | - | - | Impedance of leg left | - | Phenoscanner | UKBB |
| rs78378222 | 17:7571752 | - | - | - | Impedance of leg right | - | Phenoscanner | UKBB |
| rs78378222 | 17:7571752 | - | - | - | Impedance of whole body | - | Phenoscanner | UKBB |
| rs78378222 | 17:7571752 | - | - | - | keratinocyte cancer (mtag) | - | GWAS_catalog | 31174203 |
| rs78378222 | 17:7571752 | - | - | - | Leg fat-free mass left | - | Phenoscanner | UKBB |
| rs78378222 | 17:7571752 | - | - | - | Leg fat-free mass right | - | Phenoscanner | UKBB |
| rs78378222 | 17:7571752 | - | - | - | Leg predicted mass left | - | Phenoscanner | UKBB |
| rs78378222 | 17:7571752 | - | - | - | Leg predicted mass right | - | Phenoscanner | UKBB |
| rs78378222 | 17:7571752 | - | - | - | Leiomyoma of uterus | - | Phenoscanner | UKBB |
| rs78378222 | 17:7571752 | - | - | - | Mean corpuscular hemoglobin | Immunological | GWAS_ATLAS | 27863252 |
| rs78378222 | 17:7571752 | - | - | - | Mean corpuscular hemoglobin | - | Phenoscanner | 27863252 |
| rs78378222 | 17:7571752 | - | - | - | mean corpuscular hemoglobin | - | GWAS_catalog | 30595370 |
| rs78378222 | 17:7571752 | - | - | - | Mean corpuscular volume | Immunological | GWAS_ATLAS | 27863252 |
| rs78378222 | 17:7571752 | - | - | - | Mean corpuscular volume | - | Phenoscanner | 27863252 |
| rs78378222 | 17:7571752 | - | - | - | mosaic loss of chromosome y (y chromosome dosage) | - | GWAS_catalog | 28346444 |
| rs78378222 | 17:7571752 | - | - | - | Non glioblastoma glioma | - | Phenoscanner | 28346443 |
| rs78378222 | 17:7571752 | - | - | - | non-glioblastoma glioma | - | GWAS_catalog | 29743610 |
| rs78378222 | 17:7571752 | - | - | - | non-melanoma skin cancer | - | GWAS_catalog | 29739929 |
| rs78378222 | 17:7571752 | - | - | - | Number of operations | - | Phenoscanner | UKBB |
| rs78378222 | 17:7571752 | - | - | - | Other and unspecified malignant neoplasm of skin | Neoplasms | GWAS_ATLAS | 31427789 |
| rs78378222 | 17:7571752 | - | - | - | Other malignant neoplasms of skin | - | Phenoscanner | UKBB |
| rs78378222 | 17:7571752 | - | - | - | Pulse pressure | - | Phenoscanner | 28135244 |
| rs78378222 | 17:7571752 | - | - | - | pulse pressure | - | GWAS_catalog | 30578418 |
| rs78378222 | 17:7571752 | - | - | - | red blood cell count | - | GWAS_catalog | 30595370 |
| rs78378222 | 17:7571752 | - | - | - | Sitting height | Skeletal | GWAS_ATLAS | 31427789 |
| rs78378222 | 17:7571752 | - | - | - | Sitting height | - | Phenoscanner | UKBB |
| rs78378222 | 17:7571752 | - | - | - | Trunk fat-free mass | - | Phenoscanner | UKBB |
| rs78378222 | 17:7571752 | - | - | - | Trunk predicted mass | - | Phenoscanner | UKBB |
| rs78378222 | 17:7571752 | - | - | - | uterine fibroids | - | GWAS_catalog | 31649266 |
| rs78378222 | 17:7571752 | - | - | - | uterine fibroids and heavy menstrual bleeding | - | GWAS_catalog | 31649266 |
| rs78378222 | 17:7571752 | - | - | - | Weight | Metabolic | GWAS_ATLAS | 31427789 |
| rs78378222 | 17:7571752 | - | - | - | Weight | - | Phenoscanner | UKBB |
| rs78378222 | 17:7571752 | - | - | - | white blood cell count | - | GWAS_catalog | 30595370 |
| rs78378222 | 17:7571752 | - | - | - | Whole body fat-free mass | - | Phenoscanner | UKBB |
| rs78378222 | 17:7571752 | - | - | - | Whole body water mass | - | Phenoscanner | UKBB |
| rs78378222 | 17:7571752 | - | - | - | Years since last cervical smear test | Activities | GWAS_ATLAS | 31427789 |
| rs854786 | 17:18036283 | - | - | - | Waist-hip ratio | Metabolic | GWAS_ATLAS | 30239722 |
| rs2525570 | 17:29681245 | rs2854334 | 17:29715500 | 0,94669 | alcohol consumption (drinks per week) | - | GWAS_catalog | 30643251 |
| rs2525570 | 17:29681245 | rs2525565 | 17:29666588 | 0,9347 | alcohol consumption (drinks per week) (mtag) | - | GWAS_catalog | 30643251 |
| rs2525570 | 17:29681245 | - | - | - | Alcohol intake frequency | Psychiatric | GWAS_ATLAS | 31427789 |
| rs2525570 | 17:29681245 | - | - | - | Arm fat-free mass left | - | Phenoscanner | UKBB |
| rs2525570 | 17:29681245 | - | - | - | Arm fat-free mass right | - | Phenoscanner | UKBB |
| rs2525570 | 17:29681245 | - | - | - | Arm predicted mass left | - | Phenoscanner | UKBB |
| rs2525570 | 17:29681245 | - | - | - | Arm predicted mass right | - | Phenoscanner | UKBB |
| rs2525570 | 17:29681245 | - | - | - | Basal metabolic rate | - | Phenoscanner | UKBB |
| rs2525570 | 17:29681245 | rs12943365 | 17:29680526 | 0,92698 | bitter alcoholic beverage consumption | - | GWAS_catalog | 31046077 |
| rs2525570 | 17:29681245 | rs2040792 | 17:29628549 | 0,93842 | Comparative height size | Skeletal | GWAS_ATLAS | 31427789 |
| rs2525570 | 17:29681245 | - | - | - | Comparative height size at age 10 | - | Phenoscanner | UKBB |
| rs2525570 | 17:29681245 | rs2040792 | 17:29628549 | 0,93842 | Height | Skeletal | GWAS_ATLAS | 31427789 |
| rs2525570 | 17:29681245 | - | - | - | Height | - | Phenoscanner | UKBB |
| rs2525570 | 17:29681245 | rs2040792 | 17:29628549 | 0,93842 | Hip circumference | Metabolic | GWAS_ATLAS | 31427789 |
| rs2525570 | 17:29681245 | - | - | - | Hip circumference | - | Phenoscanner | UKBB |
| rs2525570 | 17:29681245 | rs2040792 | 17:29628549 | 0,93842 | Impedance measures - Arm fat-free mass (left) | Metabolic | GWAS_ATLAS | 31427789 |
| rs2525570 | 17:29681245 | rs2040792 | 17:29628549 | 0,93842 | Impedance measures - Arm fat-free mass (right) | Metabolic | GWAS_ATLAS | 31427789 |
| rs2525570 | 17:29681245 | rs2040792 | 17:29628549 | 0,93842 | Impedance measures - Arm predicted mass (left) | Metabolic | GWAS_ATLAS | 31427789 |
| rs2525570 | 17:29681245 | rs2040792 | 17:29628549 | 0,93842 | Impedance measures - Arm predicted mass (right) | Metabolic | GWAS_ATLAS | 31427789 |
| rs2525570 | 17:29681245 | rs2040792 | 17:29628549 | 0,93842 | Impedance measures - Basal metabolic rate | Metabolic | GWAS_ATLAS | 31427789 |
| rs2525570 | 17:29681245 | rs4368212 | 17:29639263 | 0,93853 | Impedance measures - Impedance of arm (right) | Metabolic | GWAS_ATLAS | 31427789 |
| rs2525570 | 17:29681245 | rs2040792 | 17:29628549 | 0,93842 | Impedance measures - Leg fat-free mass (left) | Metabolic | GWAS_ATLAS | 31427789 |
| rs2525570 | 17:29681245 | rs2040792 | 17:29628549 | 0,93842 | Impedance measures - Leg fat-free mass (right) | Metabolic | GWAS_ATLAS | 31427789 |
| rs2525570 | 17:29681245 | rs2040792 | 17:29628549 | 0,93842 | Impedance measures - Leg predicted mass (left) | Metabolic | GWAS_ATLAS | 31427789 |
| rs2525570 | 17:29681245 | rs2040792 | 17:29628549 | 0,93842 | Impedance measures - Leg predicted mass (right) | Metabolic | GWAS_ATLAS | 31427789 |
| rs2525570 | 17:29681245 | rs2040792 | 17:29628549 | 0,93842 | Impedance measures - Trunk fat-free mass | Metabolic | GWAS_ATLAS | 31427789 |
| rs2525570 | 17:29681245 | rs2040792 | 17:29628549 | 0,93842 | Impedance measures - Trunk predicted mass | Metabolic | GWAS_ATLAS | 31427789 |
| rs2525570 | 17:29681245 | rs2040792 | 17:29628549 | 0,93842 | Impedance measures - Weight | Metabolic | GWAS_ATLAS | 31427789 |
| rs2525570 | 17:29681245 | rs2040792 | 17:29628549 | 0,93842 | Impedance measures - Whole body fat-free mass | Metabolic | GWAS_ATLAS | 31427789 |
| rs2525570 | 17:29681245 | rs2040792 | 17:29628549 | 0,93842 | Impedance measures - Whole body water mass | Metabolic | GWAS_ATLAS | 31427789 |
| rs2525570 | 17:29681245 | - | - | - | Impedance of arm right | - | Phenoscanner | UKBB |
| rs2525570 | 17:29681245 | - | - | - | Leg fat-free mass left | - | Phenoscanner | UKBB |
| rs2525570 | 17:29681245 | - | - | - | Leg fat-free mass right | - | Phenoscanner | UKBB |
| rs2525570 | 17:29681245 | - | - | - | Leg predicted mass left | - | Phenoscanner | UKBB |
| rs2525570 | 17:29681245 | - | - | - | Leg predicted mass right | - | Phenoscanner | UKBB |
| rs2525570 | 17:29681245 | - | - | - | Trunk fat-free mass | - | Phenoscanner | UKBB |
| rs2525570 | 17:29681245 | - | - | - | Trunk predicted mass | - | Phenoscanner | UKBB |
| rs2525570 | 17:29681245 | rs2040792 | 17:29628549 | 0,93842 | Weight | Metabolic | GWAS_ATLAS | 31427789 |
| rs2525570 | 17:29681245 | - | - | - | Weight | - | Phenoscanner | UKBB |
| rs2525570 | 17:29681245 | - | - | - | Whole body fat-free mass | - | Phenoscanner | UKBB |
| rs2525570 | 17:29681245 | - | - | - | Whole body water mass | - | Phenoscanner | UKBB |
| rs4423457 | 17:64284000 | rs72843895 | 17:64331156 | 0,83008 | height | - | GWAS_catalog | 30595370 |
| rs4423457 | 17:64284000 | rs56152251 | 17:64280153 | 0,93318 | Height | Skeletal | GWAS_ATLAS | 31427789 |
| rs4423457 | 17:64284000 | - | - | - | Height | - | Phenoscanner | UKBB |
| rs4423457 | 17:64284000 | rs56152251 | 17:64280153 | 0,93318 | Impedance measures - Trunk fat-free mass | Metabolic | GWAS_ATLAS | 31427789 |
| rs4423457 | 17:64284000 | rs67700546 | 17:64301081 | 0,86769 | Impedance measures - Trunk predicted mass | Metabolic | GWAS_ATLAS | 31427789 |
| rs4423457 | 17:64284000 | rs56152251 | 17:64280153 | 0,93318 | qt interval | - | GWAS_catalog | 29213071 |
| rs4423457 | 17:64284000 | rs56152251 | 17:64280153 | 0,93318 | QT interval | - | Phenoscanner | 29213071 |
| rs4423457 | 17:64284000 | rs56152251 | 17:64280153 | 0,93318 | Sitting height | Skeletal | GWAS_ATLAS | 31427789 |
| rs4423457 | 17:64284000 | - | - | - | Sitting height | - | Phenoscanner | UKBB |
| rs2421206 | 19:11262477 | - | - | - | Comparative height size at age 10 | - | Phenoscanner | UKBB |
| rs2421206 | 19:11262477 | - | - | - | Free cholesterol in large LDL | Metabolic | GWAS_ATLAS | 27005778 |
| rs2421206 | 19:11262477 | - | - | - | Height | - | Phenoscanner | UKBB |
| rs2421206 | 19:11262477 | - | - | - | Low-density lipoprotein cholesterol | Metabolic | GWAS_ATLAS | 31217584 |
| rs2421206 | 19:11262477 | - | - | - | Phospholipids in medium LDL | Metabolic | GWAS_ATLAS | 27005778 |
| rs2421206 | 19:11262477 | - | - | - | Resting heart rate | Cardiovascular | GWAS_ATLAS | 30940143 |
| rs2421206 | 19:11262477 | - | - | - | Self-reported high cholesterol | - | Phenoscanner | UKBB |
| rs2421206 | 19:11262477 | - | - | - | Sitting height | - | Phenoscanner | UKBB |
| rs2421206 | 19:11262477 | - | - | - | Total cholesterol in LDL | Metabolic | GWAS_ATLAS | 27005778 |
| rs2421206 | 19:11262477 | - | - | - | Total cholesterol in medium LDL | Metabolic | GWAS_ATLAS | 27005778 |
| rs2421206 | 19:11262477 | - | - | - | Treatment with atorvastatin | - | Phenoscanner | UKBB |
| rs8106090 | 19:44371210 | rs375066 | 19:44423570 | 0,92325 | breast cancer | - | GWAS_catalog | 29059683 |
| rs8106090 | 19:44371210 | rs4802201 | 19:44358042 | 0,8511 | Granulocyte percentage of myeloid white cells | Immunological | GWAS_ATLAS | 27863252 |
| rs8106090 | 19:44371210 | - | - | - | Granulocyte percentage of myeloid white cells | - | Phenoscanner | 27863252 |
| rs8106090 | 19:44371210 | rs4802201 | 19:44358042 | 0,8511 | Monocyte count | Immunological | GWAS_ATLAS | 27863252 |
| rs8106090 | 19:44371210 | - | - | - | Monocyte count | - | Phenoscanner | 27863252 |
| rs8106090 | 19:44371210 | rs4802201 | 19:44358042 | 0,8511 | Monocyte percentage of white cells | Immunological | GWAS_ATLAS | 27863252 |
| rs8106090 | 19:44371210 | - | - | - | Monocyte percentage of white cells | - | Phenoscanner | 27863252 |
| rs34161672 | 20:56020599 | - | - | - | Mean platelet volume | Immunological | GWAS_ATLAS | 27863252 |
| rs34161672 | 20:56020599 | - | - | - | Mean platelet volume | - | Phenoscanner | 27863252 |
| rs35384758 | 20:58998651 | - | - | - | Haemorrhoids | - | Phenoscanner | UKBB |
| rs174767 | 22:29740728 | rs3788411 | 22:29728783 | 0,98694 | Immature fraction of reticulocytes | Immunological | GWAS_ATLAS | 27863252 |
| rs174767 | 22:29740728 | - | - | - | Immature fraction of reticulocytes | - | Phenoscanner | 27863252 |
| rs174767 | 22:29740728 | rs4820804 | 22:29669693 | 0,89233 | Mean platelet volume | Immunological | GWAS_ATLAS | 27863252 |
| rs174767 | 22:29740728 | - | - | - | Mean platelet volume | - | Phenoscanner | 27863252 |
| rs35318931 | X:38009121 | - | - | - | Height | - | Phenoscanner | 28146470 |
| rs35318931 | X:38009121 | - | - | - | Nonsyndromic striae distensae stretch marks | - | Phenoscanner | 23633020 |
