## Supplementary Table 5 for "Genome-wide analysis of 944,133 individuals provides insights into the etiology of hemorrhoidal disease"

**Supplementary Table 5: Results of subset-based pleiotropy meta-analysis (SBM).**

**Lead SNP (rsID):** rs ID retrieved from NCBI's dbSNP build v150; **CHR:** chromosome; **BP:** base pair position; **start-end**: left/right association boundaries for each lead SNP defined by FUMA (see **Supplementary Note**); **Phenotype(s):** subset of phenotypes from SBM with genome-wide significance ($P_{SBM}$<5×10$^{-8}$); **P**: Final *P*-value ($P_{SBM}$) from two-sided statistic for the detection of potential effects in opposite directions (combined from two-sided statistics *P* subset1 and *P* subset 2). i.e. adjusted (disease-combined) *P*-value ($P_{SBM}$) from SBM (see **Online Methods**); **Phenotype(s) subset 1:** risk (OR>1) disease subset from SBM; **P subset 1:** *P*-value from two-sided statistic across phenotype(s) of subset 1; **OR (95% CI) subset 1:** odds ratio and corresponding 95% confidence interval of phenotype(s) from subset 1; **Phenotype(s) subset 2:** protective (OR<1) disease subset from SBM; **P subset 2:** *P*-value from two-sided statistic across phenotype(s) of subset 2; **OR (95% CI) subset 2:** odds ratio and corresponding 95% confidence interval of phenotype(s) from subset 2.

| Lead SNP (rsID) | CHR | BP | start-end | Phenotype(s) | P | Phenotypes(s) subset1 | P subset1 | OR (95%CI) subset1 | Phenotypes(s) subset2 | P subset2 | OR (95%CI) subset2 |
|---|---|---|---|---|---|---|---|---|---|---|---|
| rs11580446 | 1 | 95241697 | 95203992-95309134 | IBS,HEM | 7.2e-13 | IBS | 0.01368 | 1.05 (1.01-1.08) | HEM | 1.6e-12 | 0.97 (0.96-0.98) |
| rs145163454 | 1 | 169090748 | 169081792-169521553 | HEM,DIV | 1.6e-28 | HEM,DIV | 1.6e-28 | 1.01 (1.01-1.01) | na | 1.00000 | NA (NA-NA) |
| rs4951080 | 1 | 204533284 | 204449952-204599461 | HEM,IBS | 3.1e-08 | na | 1.00000 | NA (NA-NA) | HEM,IBS | 3.1e-08 | 0.98 (0.97-0.98) |
| rs61823192 | 1 | 219294570 | 219268532-219704939 | HEM,DIV,IBS | 1.4e-12 | HEM | 0.91549 | 1 (0.96-1.05) | DIV,IBS | 4.8e-14 | 0.99 (0.98-0.99) |
| rs79059272 | 2 | 67922634 | 67874054-67954046 | HEM,DIV | 8.2e-09 | HEM,DIV | 8.2e-09 | 1 (1-1) | na | 1.00000 | NA (NA-NA) |
| rs6747171 | 2 | 145819888 | 145756260-145904907 | IBS,HEM,DIV | 1.2e-08 | IBS | 0.06787 | 1.03 (1-1.06) | HEM,DIV | 7.9e-09 | 1 (1-1) |
| rs12987602 | 2 | 160960788 | 160899369-161340963 | DIV,HEM,IBS | 5.2e-12 | DIV | 0.01576 | 1 (1-1) | HEM,IBS | 1.1e-11 | 0.98 (0.97-0.98) |
| rs13017210 | 2 | 174008623 | 173943763-174107004 | HEM,DIV | 7.4e-11 | HEM | 7.9e-10 | 1.03 (1.02-1.03) | DIV | 0.00340 | 1 (1-1) |
| rs4853718 | 2 | 191404964 | 191259714-191478211 | HEM,IBS | 1.4e-08 | HEM | 1.2e-07 | 1.03 (1.02-1.03) | IBS | 0.00534 | 0.95 (0.92-0.99) |
| **rs3731912** | **2** | **220406722** | **220399638-220436704** | **HEM,IBS** | **1.1e-08** | **HEM** | **1.3e-07** | **1.03 (1.02-1.03)** | **IBS** | **0.00366** | **0.95 (0.92-0.98)** |
| rs2597301 | 3 | 70909494 | 70836756-71047711 | HEM,IBS | 4.6e-14 | HEM | 1.3e-13 | 1.03 (1.02-1.04) | IBS | 0.01050 | 0.96 (0.93-0.99) |
| **rs9876268** | **3** | **151064376** | **150800402-151130197** | **IBS,DIV** | **2.2e-12** | **IBS** | **0.00017** | **1.08 (1.04-1.12)** | **DIV** | **4.1e-10** | **1 (0.99-1)** |
| rs3851366 | 3 | 160191374 | 160004026-160309529 | HEM,DIV | 5.7e-10 | HEM | 6.1e-10 | 1.02 (1.02-1.03) | DIV | 0.03679 | 1 (1-1) |
| rs11942410 | 4 | 75656079 | 75656079-75686640 | DIV,IBS,HEM | 1.8e-12 | DIV,IBS | 5.3e-06 | 1 (1-1) | HEM | 1.0e-08 | 0.97 (0.96-0.98) |
| **rs9307546** | **4** | **125672407** | **125669671-125751084** | **DIV,HEM** | **3.4e-08** | **DIV** | **0.00093** | **1 (1-1)** | **HEM** | **1.7e-06** | **0.98 (0.97-0.99)** |
| rs17824374 | 4 | 126924999 | 126893588-127030561 | HEM,IBS,DIV | 4.5e-10 | HEM,IBS | 1.8e-11 | 1.03 (1.02-1.04) | DIV | 0.95425 | 1 (1-1) |
| rs1542726 | 4 | 145515769 | 145227879-145868370 | HEM,DIV | 1.0e-12 | HEM | 1.5e-12 | 1.03 (1.02-1.04) | DIV | 0.02179 | 1 (1-1) |
| **rs10471645** | **5** | **64295363** | **64128671-64330833** | **DIV,HEM** | **3.0e-11** | **DIV** | **6.4e-11** | **1 (1-1.01)** | **HEM** | **0.01656** | **0.99 (0.98-1)** |
| rs13174700 | 5 | 93554644 | 92948485-93571190 | IBS,HEM | 1.7e-08 | IBS | 0.04059 | 1.05 (1-1.09) | HEM | 1.9e-08 | 0.97 (0.95-0.98) |
| rs12654883 | 5 | 97656313 | 97652398-97700050 | HEM,IBS | 2.6e-08 | HEM,IBS | 2.6e-08 | 1.02 (1.02-1.03) | na | 1.00000 | NA (NA-NA) |
| **rs965821** | **5** | **122137708** | **122073550-122374459** | **IBS,DIV** | **1.2e-08** | **IBS** | **0.00133** | **1.05 (1.02-1.08)** | **DIV** | **3.9e-07** | **1 (1-1)** |
| rs971569 | 5 | 164690314 | 164487955-164786699 | HEM,IBS | 4.2e-09 | HEM | 3.7e-09 | 1.03 (1.02-1.04) | IBS | 0.04850 | 0.96 (0.93-1) |
| rs7755093 | 6 | 17387077 | 17386607-17479588 | HEM,IBS | 1.3e-11 | na | 1.00000 | NA (NA-NA) | HEM,IBS | 1.3e-11 | 0.96 (0.96-0.98) |
| **rs6934679** | **6** | **127761588** | **127664661-127777516** | **HEM,DIV** | **1.6e-08** | **HEM,DIV** | **1.6e-08** | **1 (1-1)** | **na** | **1.00000** | **NA (NA-NA)** |
| rs2327429 | 6 | 134209837 | 134150561-134214525 | HEM,IBS | 9.8e-09 | HEM,IBS | 9.8e-09 | 1.03 (1.02-1.03) | na | 1.00000 | NA (NA-NA) |
| rs3253 | 6 | 169616112 | 169572602-169618454 | HEM,DIV | 1.5e-11 | HEM,DIV | 1.5e-11 | 1 (1-1) | na | 1.00000 | NA (NA-NA) |
| rs4556017 | 7 | 100632790 | 100528223-100753880 | DIV,IBS,HEM | 3.9e-24 | DIV,IBS | 0.00065 | 1 (1-1) | HEM | 1.0e-22 | 0.94 (0.93-0.96) |
| **rs12672683** | **7** | **113998486** | **113970468-114004121** | **HEM,IBS,DIV** | **7.0e-09** | **HEM,IBS** | **1.8e-07** | **1.02 (1.01-1.03)** | **DIV** | **0.00168** | **1 (1-1)** |
| rs3735894 | 8 | 22454826 | 22447426-22542962 | IBS,HEM,DIV | 3.3e-11 | IBS | 0.00121 | 1.05 (1.02-1.08) | HEM,DIV | 9.7e-10 | 1 (1-1) |
| rs13252808 | 8 | 71616655 | 71042430-72277688 | DIV,HEM,IBS | 1.7e-39 | DIV | 0.00455 | 1 (1-1) | HEM,IBS | 4.1e-39 | 0.95 (0.94-0.96) |
| **rs2293889** | **8** | **116599199** | **116464988-116639474** | **HEM,IBS,DIV** | **1.4e-08** | **HEM,IBS** | **0.00276** | **1.01 (1-1.02)** | **DIV** | **2.3e-07** | **1 (1-1)** |
| rs1333047 | 9 | 22124504 | 21995044-22125503 | HEM,IBS | 4.7e-28 | HEM,IBS | 4.7e-28 | 1.04 (1.03-1.05) | na | 1.00000 | NA (NA-NA) |
| **rs12421446** | **11** | **1612992** | **1492445-1645885** | **HEM,DIV** | **4.5e-08** | **HEM** | **4.7e-05** | **1.02 (1.01-1.03)** | **DIV** | **4.6e-05** | **1 (1-1)** |
| rs4910165 | 11 | 10674044 | 10655021-10701556 | HEM,DIV | 1.8e-17 | HEM | 3.2e-16 | 1.03 (1.03-1.04) | DIV | 0.00134 | 1 (1-1) |
| rs2077561 | 11 | 69992199 | 69971277-70040925 | DIV,HEM | 2.1e-11 | DIV | 2.5e-08 | 1 (1-1) | HEM | 2.9e-05 | 0.98 (0.97-0.99) |
| rs4937872 | 11 | 112827715 | 112826867-112912811 | IBS,HEM | 1.9e-10 | IBS | 0.02136 | 1.03 (1-1.07) | HEM | 3.3e-10 | 0.97 (0.97-0.98) |
| rs12594232 | 15 | 68469402 | 68324830-68567752 | HEM,DIV | 8.7e-10 | HEM | 1.7e-09 | 1.02 (1.02-1.03) | DIV | 0.02061 | 1 (1-1) |
| **rs12441130** | **15** | **74234902** | **74220599-74243246** | **DIV,IBS,HEM** | **4.8e-10** | **DIV,IBS** | **1.6e-06** | **1 (1-1)** | **HEM** | **1.2e-05** | **0.98 (0.98-0.99)** |
| **rs112991346** | **16** | **19967668** | **19707417-19980931** | **HEM,DIV** | **6.1e-09** | **HEM** | **9.1e-07** | **1.03 (1.02-1.04)** | **DIV** | **0.00029** | **1 (1-1)** |
| rs62061554 | 17 | 12401309 | 12394980-12410649 | HEM,IBS | 1.6e-09 | HEM,IBS | 1.6e-09 | 1.04 (1.03-1.05) | na | 1.00000 | NA (NA-NA) |
| rs2525570 | 17 | 29681245 | 29400158-29735829 | IBS,HEM,DIV | 1.9e-10 | IBS | 0.62685 | 1.01 (0.98-1.04) | HEM,DIV | 1.1e-11 | 1 (1-1) |
| **rs61043970** | **18** | **52881622** | **52863279-52921673** | **IBS,DIV** | **3.0e-08** | **IBS** | **7.0e-05** | **1.06 (1.03-1.1)** | **DIV** | **2.0e-05** | **1 (1-1)** |
| rs113783450 | 19 | 11271714 | 11262477-11277074 | HEM,IBS | 2.9e-13 | HEM,IBS | 2.9e-13 | 1.03 (1.02-1.04) | na | 1.00000 | NA (NA-NA) |
| **rs2205558** | **21** | **29484711** | **29482552-29513936** | **IBS,HEM,DIV** | **3.8e-08** | **IBS** | **0.64411** | **1.01 (0.98-1.04)** | **HEM,DIV** | **2.8e-09** | **1 (1-1)** |
