## Supplementary Table 6 for "Genome-wide analysis of 944,133 individuals provides insights into the etiology of hemorrhoidal disease"

**Supplementary Table 6. Significant genetic correlations between HEM and other traits estimated by genome-wide LD Score Regression (LDSC).**

**Trait:** the name of the complex trait or disease being compared to HEM for genetic correlation; **rg:** genetic correlation between HEM and each trait, ranging from -1 to 1. rg>0 represents a po negative genetic correlation; **se:** standard error of rg; **z score:** the z-score from statistical test of genetic correlation; **P FDR:** the z-score from statistical test of genetic correlation after FDR cc at α=0.05; **N total:** the sample size being included in the genetic analysis of each trait; **Cohort:** the data source from which the summary statistic of the trait were extracted.

| Trait | rg | se | z score | P FDR | N total |
|---|---|---|---|---|---|
| Diagnoses - main ICD10: I84 Haemorrhoids | 0,809 | 0,049 | 16,4 | 4,1E-57 | 361194 |
| Diagnoses - main ICD10: K62 Other diseases of anus and rectum | 0,777 | 0,130 | 6,0 | 4,9E-08 | 361194 |
| Diagnoses - main ICD10: K60 Fissure and fistula of anal and rectal regions | 0,583 | 0,077 | 7,5 | 2,7E-12 | 361194 |
| Ever had bowel cancer screening | 0,545 | 0,040 | 13,5 | 8,5E-39 | 355353 |
| Non-cancer illness code, self-reported: irritable bowel syndrome | 0,424 | 0,074 | 5,8 | 1,9E-07 | 361141 |
| Medication for pain relief, constipation, heartburn: Laxatives (e.g. Dulcolax, Senokot) | 0,421 | 0,052 | 8,2 | 2,4E-14 | 357084 |
| Mouth/teeth dental problems: Painful gums | 0,406 | 0,072 | 5,6 | 4,0E-07 | 359841 |
| Treatment/medication code: citalopram | 0,400 | 0,078 | 5,1 | 5,0E-06 | 361141 |
| Diagnoses - main ICD10: K63 Other diseases of intestine | 0,390 | 0,108 | 3,6 | 3,0E-03 | 361194 |
| Diagnoses - main ICD10: R19 Other symptoms and signs involving the digestive system and abdomen | 0,389 | 0,127 | 3,1 | 1,6E-02 | 361194 |
| Destinations on discharge from hospital (recoded): Usual Place of residence | 0,369 | 0,035 | 10,6 | 4,2E-24 | 361058 |
| Tinnitus: Yes, now some of the time | 0,358 | 0,100 | 3,6 | 3,3E-03 | 117882 |
| Paingul gums | 0,357 | 0,059 | 6,1 | 3,1E-08 | 461031 |
| Hospital episode type: General episode | 0,356 | 0,042 | 8,5 | 2,2E-15 | 361194 |
| Treatment/medication code: omeprazole | 0,350 | 0,038 | 9,1 | 6,6E-18 | 361141 |
| Had other major operations | 0,349 | 0,052 | 6,7 | 6,9E-10 | 192470 |
| Illnesses of mother: Severe depression | 0,347 | 0,059 | 5,9 | 6,8E-08 | 330653 |
| Any ICDMAIN event in hilmo or causes of death | 0,340 | 0,033 | 10,2 | 2,2E-22 | 361194 |
| Non-cancer illness code, self-reported: gastro-oesophageal reflux (gord) / gastric reflux | 0,338 | 0,051 | 6,6 | 1,3E-09 | 361141 |
| Substances taken for anxiety: Medication prescribed to you (for at least two weeks) | 0,334 | 0,049 | 6,8 | 3,8E-10 | 98990 |
| Mental health problems ever diagnosed by a professional: Anxiety, nerves or generalized anxiety disorder | 0,334 | 0,051 | 6,5 | 2,5E-09 | 117751 |
| Medication for pain relief, constipation, heartburn: Omeprazole (e.g. Zanprol) | 0,332 | 0,035 | 9,5 | 1,9E-19 | 357084 |
| Factors influencing health status and contact with health services | 0,327 | 0,064 | 5,1 | 5,6E-06 | 361194 |
| Diagnoses - main ICD10: K29 Gastritis and duodenitis | 0,326 | 0,050 | 6,5 | 3,1E-09 | 361194 |
| Other gastritis (incl. Duodenitis) | 0,325 | 0,054 | 6,0 | 4,9E-08 | 361194 |
| Pain type(s) experienced in last month: Stomach or abdominal pain | 0,324 | 0,034 | 9,5 | 2,4E-19 | 360391 |
| Activities undertaken to treat anxiety: Talking therapies, such as psychotherapy, counselling, group therapy or CBT | 0,324 | 0,044 | 7,3 | 1,5E-11 | 98990 |
| Diagnoses - main ICD10: T81 Complications of procedures, not elsewhere classified | 0,322 | 0,096 | 3,4 | 6,1E-03 | 361194 |
| Non-cancer illness code, self-reported: anxiety/panic attacks | 0,317 | 0,061 | 5,2 | 4,2E-06 | 361141 |
| Diagnoses - main ICD10: R31 Unspecified haematuria | 0,310 | 0,071 | 4,4 | 1,9E-04 | 361194 |
| Pain type(s) experienced in last month: Facial pain | 0,304 | 0,061 | 5,0 | 1,2E-05 | 360391 |
| Treatment/medication code: fluoxetine | 0,302 | 0,085 | 3,6 | 3,5E-03 | 361141 |
| Illness, injury, bereavement, stress in last 2 years: Marital separation/divorce | 0,302 | 0,111 | 2,7 | 3,9E-02 | 358836 |
| Seen doctor (GP) for nerves, anxiety, tension or depression | 0,300 | 0,026 | 11,5 | 6,6E-28 | 358693 |
| Non-cancer illness code, self-reported: spine arthritis/spondylitis | 0,298 | 0,093 | 3,2 | 1,0E-02 | 361141 |
| Symptoms, signs and abnormal clinical and laboratory findings, not elsewhere classified | 0,292 | 0,035 | 8,3 | 6,3E-15 | 361194 |
| Diagnoses - main ICD10: R10 Abdominal and pelvic pain | 0,286 | 0,048 | 5,9 | 7,9E-08 | 361194 |
| Mental health problems ever diagnosed by a professional: Panic attacks | 0,281 | 0,059 | 4,8 | 2,7E-05 | 117722 |
| Major Depresive Disorder | 0,279 | 0,026 | 10,6 | 8,9E-24 | 480359 |
| Activities undertaken to treat depression: Talking therapies, such as psychotherapy, counselling, group therapy or CBT | 0,278 | 0,043 | 6,5 | 2,2E-09 | 117763 |
| Knee pain for 3+ months | 0,275 | 0,084 | 3,3 | 8,3E-03 | 76000 |
| Mental health problems ever diagnosed by a professional: Depression | 0,275 | 0,041 | 6,7 | 8,4E-10 | 117782 |
| Ever felt worried, tense, or anxious for most of a month or longer | 0,273 | 0,039 | 7,0 | 1,2E-10 | 110315 |
| Diseases of the circulatory system | 0,270 | 0,033 | 8,1 | 4,0E-14 | 361194 |
| Professional informed about depression | 0,267 | 0,061 | 4,4 | 1,8E-04 | 66302 |
| Multisite chronic pain | 0,267 | 0,022 | 12,4 | 1,8E-32 | 380000 |
| Illnesses of siblings: Severe depression | 0,263 | 0,044 | 5,9 | 6,8E-08 | 279858 |
| Back pain for 3+ months | 0,261 | 0,046 | 5,7 | 2,7E-07 | 90555 |
| Other eye problems | 0,261 | 0,052 | 5,0 | 9,6E-06 | 360134 |
| Diagnoses - main ICD10: K44 Diaphragmatic hernia | 0,261 | 0,068 | 3,8 | 1,5E-03 | 361194 |
| Substances taken for anxiety: Drugs or alcohol (more than once) | 0,260 | 0,075 | 3,5 | 4,6E-03 | 98990 |
| Other serious medical condition/disability diagnosed by doctor | 0,258 | 0,035 | 7,3 | 1,3E-11 | 354588 |
| Bipolar and major depression status: Probable Recurrent major depression (severe) | 0,258 | 0,074 | 3,5 | 4,6E-03 | 86895 |
| Medication for pain relief, constipation, heartburn: Ranitidine (e.g. Zantac) | 0,257 | 0,065 | 4,0 | 9,2E-04 | 357084 |
| Ever sought or received professional help for mental distress | 0,256 | 0,037 | 6,9 | 2,8E-10 | 117677 |
| Substances taken for depression: Medication prescribed to you (for at least two weeks) | 0,255 | 0,037 | 6,9 | 2,7E-10 | 117763 |
| Non-cancer illness code, self-reported: depression | 0,254 | 0,042 | 6,0 | 4,8E-08 | 361141 |
| Diseases of the genitourinary system | 0,254 | 0,040 | 6,4 | 4,1E-09 | 361194 |
| Treatment/medication code: ranitidine | 0,252 | 0,066 | 3,8 | 1,6E-03 | 361141 |
| Tinnitus: Yes, but not now, but have in the past | 0,251 | 0,071 | 3,5 | 3,9E-03 | 117882 |
| Taking other prescription medications | 0,247 | 0,027 | 9,1 | 6,6E-18 | 360027 |
| Non-cancer illness code, self-reported: back problem | 0,247 | 0,072 | 3,4 | 5,3E-03 | 361141 |
| Diagnoses - main ICD10: Z09 Follow-up examination after treatment for conditions other than malignant neoplasms | 0,246 | 0,067 | 3,7 | 2,3E-03 | 361194 |
| Neck/shoulder pain for 3+ months | 0,246 | 0,082 | 3,0 | 1,8E-02 | 81276 |
| Fibromyalgia related co-morbidities | 0,246 | 0,094 | 2,6 | 4,8E-02 | 361194 |
| Tense, sore, or aching muscles during worst period of anxiety | 0,244 | 0,060 | 4,1 | 5,4E-04 | 33301 |
| Tense / 'highly strung' | 0,243 | 0,028 | 8,8 | 1,6E-16 | 350159 |
| Activities undertaken to treat anxiety: Other therapeutic activities such as mindfulness, yoga or art classes | 0,242 | 0,063 | 3,8 | 1,5E-03 | 98990 |
| Seen a psychiatrist for nerves, anxiety, tension or depression | 0,240 | 0,034 | 7,1 | 5,7E-11 | 359535 |
| Treatment/medication code: amitriptyline | 0,237 | 0,064 | 3,7 | 2,4E-03 | 361141 |
| Ever worried more than most people would in similar situation | 0,236 | 0,038 | 6,2 | 1,5E-08 | 98990 |
| Number of things worried about during worst period of anxiety | 0,236 | 0,088 | 2,7 | 4,1E-02 | 35558 |
| Fractured bone site(s): Ankle | 0,234 | 0,072 | 3,2 | 9,1E-03 | 359241 |
| Diagnoses - main ICD10: K57 Diverticular disease of intestine | 0,233 | 0,037 | 6,3 | 6,7E-09 | 361194 |
| Ever had prolonged feelings of sadness or depression | 0,233 | 0,037 | 6,3 | 8,2E-09 | 117763 |
| Ever used hormone-replacement therapy (HRT) | 0,232 | 0,034 | 6,9 | 1,9E-10 | 193606 |
| Diagnoses - main ICD10: R07 Pain in throat and chest | 0,232 | 0,037 | 6,2 | 1,3E-08 | 361194 |
| Hair colour (natural, before greying): Other | 0,228 | 0,084 | 2,7 | 3,9E-02 | 360270 |
| Activities undertaken to treat depression: Other therapeutic activities such as mindfulness, yoga or art classes | 0,227 | 0,052 | 4,4 | 1,8E-04 | 117763 |
| Diagnoses - main ICD10: F31 Bipolar affective disorder | 0,226 | 0,065 | 3,5 | 4,1E-03 | 361194 |
| Non-cancer illness code, self-reported: mania/bipolar disorder/manic depression | 0,224 | 0,074 | 3,0 | 1,8E-02 | 361141 |
| Professional informed about anxiety | 0,222 | 0,078 | 2,9 | 2,7E-02 | 36366 |
| Diagnoses - main ICD10: D12 Benign neoplasm of colon, rectum, anus and anal canal | 0,220 | 0,045 | 4,9 | 2,0E-05 | 361194 |
| Non-cancer illness code, self-reported: cervical spondylosis | 0,220 | 0,082 | 2,7 | 4,0E-02 | 361141 |
| Ever suffered mental distress preventing usual activities | 0,219 | 0,035 | 6,3 | 7,5E-09 | 116527 |
| Manifestations of mania or irritability: I was easily distracted | 0,217 | 0,060 | 3,6 | 2,8E-03 | 114422 |
| Ever self-harmed | 0,216 | 0,060 | 3,6 | 3,0E-03 | 117733 |
| Illnesses of father: Severe depression | 0,215 | 0,067 | 3,2 | 1,0E-02 | 312437 |

| | | | | | |
|---|---|---|---|---|---|
| Severity of problems due to mania or irritability | 0,214 | 0,078 | 2,7 | 3,6E-02 | 26655 |
| Back pain | 0,212 | 0,024 | 8,8 | 1,6E-16 | 509000 |
| Bipolar and major depression status: Probable Recurrent major depression (moderate) | 0,212 | 0,063 | 3,4 | 6,1E-03 | 86895 |
| Treatment/medication code: diclofenac | 0,212 | 0,064 | 3,3 | 7,3E-03 | 361141 |
| Toothache | 0,211 | 0,053 | 4,0 | 7,3E-04 | 461031 |
| Osteoporosis | 0,211 | 0,070 | 3,0 | 1,8E-02 | 361194 |
| Pain type(s) experienced in last month: Back pain | 0,210 | 0,027 | 7,7 | 5,7E-13 | 360391 |
| Had major operations | 0,210 | 0,044 | 4,8 | 2,9E-05 | 165737 |
| Non-cancer illness code, self-reported: osteoporosis | 0,210 | 0,048 | 4,3 | 1,9E-04 | 361141 |
| Bring up phlegm/sputum/mucus on most days | 0,208 | 0,063 | 3,3 | 8,3E-03 | 91787 |
| Neuroticism score | 0,207 | 0,025 | 8,1 | 2,8E-14 | 293006 |
| Medication for pain relief, constipation, heartburn: Paracetamol | 0,207 | 0,027 | 7,7 | 8,9E-13 | 357084 |
| Chest pain or discomfort | 0,206 | 0,031 | 6,7 | 7,4E-10 | 357507 |
| Long-standing illness, disability or infirmity | 0,206 | 0,025 | 8,2 | 2,3E-14 | 352798 |
| Worrier / anxious feelings | 0,205 | 0,028 | 7,4 | 7,0E-12 | 351833 |
| Treatment/medication code: lansoprazole | 0,204 | 0,042 | 4,8 | 2,0E-05 | 361141 |
| Manifestations of mania or irritability: I was more restless than usual | 0,202 | 0,046 | 4,4 | 1,4E-04 | 114422 |
| Diagnoses - main ICD10: K50 Crohn's disease [regional enteritis] | 0,201 | 0,074 | 2,7 | 3,9E-02 | 361194 |
| Non-cancer illness code, self-reported: chronic fatigue syndrome | 0,201 | 0,077 | 2,6 | 4,9E-02 | 361141 |
| Diagnoses - main ICD10: N39 Other disorders of urinary system | 0,200 | 0,052 | 3,9 | 1,2E-03 | 361194 |
| Non-cancer illness code, self-reported: osteoarthritis | 0,199 | 0,034 | 5,8 | 1,2E-07 | 361141 |
| Pain type(s) experienced in last month: Headache | 0,198 | 0,029 | 6,8 | 4,6E-10 | 360391 |
| Detention categories: Informal, not formally detained | 0,198 | 0,066 | 3,0 | 1,8E-02 | 361194 |
| Noninfectious colitis NAS | 0,197 | 0,069 | 2,8 | 2,8E-02 | 361194 |
| Mineral and other dietary supplements: Calcium | 0,197 | 0,047 | 4,2 | 3,6E-04 | 360016 |
| Treatment/medication code: paracetamol | 0,197 | 0,029 | 6,8 | 5,3E-10 | 361141 |
| Other/unspecified dorsalgia | 0,197 | 0,074 | 2,6 | 4,6E-02 | 361194 |
| Treatment/medication code: alendronate sodium | 0,197 | 0,063 | 3,1 | 1,3E-02 | 361141 |
| Mouth/teeth dental problems: Toothache | 0,195 | 0,060 | 3,2 | 9,4E-03 | 359841 |
| Ever had period of mania / excitability | 0,194 | 0,061 | 3,2 | 1,0E-02 | 115338 |
| Breathing problems improved/stopped away from workplace or on holiday: No | 0,194 | 0,074 | 2,6 | 4,6E-02 | 91149 |
| Ever had period extreme irritability | 0,194 | 0,036 | 5,4 | 1,6E-06 | 114422 |
| Diagnoses - main ICD10: K52 Other non-infective gastro-enteritis and colitis | 0,193 | 0,069 | 2,8 | 3,1E-02 | 361194 |
| Non-cancer illness code, self-reported: hiatus hernia | 0,192 | 0,052 | 3,7 | 2,3E-03 | 361141 |
| Non-cancer illness code, self-reported: diverticular disease/diverticulitis | 0,192 | 0,047 | 4,1 | 5,8E-04 | 361141 |
| Manifestations of mania or irritability: I needed less sleep than usual | 0,189 | 0,061 | 3,1 | 1,5E-02 | 114422 |
| Ever depressed for a whole week | 0,189 | 0,039 | 4,9 | 1,8E-05 | 117705 |
| Medication for cholesterol, blood pressure, diabetes, or take exogenous hormones: Hormone replacement therapy | 0,188 | 0,053 | 3,5 | 3,8E-03 | 193148 |
| Mood swings | 0,187 | 0,026 | 7,2 | 2,6E-11 | 352604 |
| Medication for pain relief, constipation, heartburn: Ibuprofen (e.g. Nurofen) | 0,187 | 0,039 | 4,8 | 3,0E-05 | 357084 |
| Vitamin and mineral supplements: Vitamin D | 0,187 | 0,064 | 2,9 | 2,4E-02 | 359245 |
| Mouth ulcers | 0,186 | 0,028 | 6,6 | 1,2E-09 | 461031 |
| Mouth/teeth dental problems: Mouth ulcers | 0,185 | 0,032 | 5,7 | 2,5E-07 | 359841 |
| Bilateral oophorectomy (both ovaries removed) | 0,185 | 0,050 | 3,7 | 1,9E-03 | 191515 |
| Vitamin and mineral supplements: Vitamin B | 0,185 | 0,056 | 3,3 | 7,9E-03 | 359245 |
| Diagnoses - main ICD10: N32 Other disorders of bladder | 0,184 | 0,069 | 2,7 | 4,2E-02 | 361194 |
| Iron deficiency anaemia | 0,184 | 0,066 | 2,8 | 3,2E-02 | 361194 |
| Diagnoses - main ICD10: D50 Iron deficiency anaemia | 0,184 | 0,066 | 2,8 | 3,2E-02 | 361194 |
| Treatment/medication code: ibuprofen | 0,183 | 0,043 | 4,3 | 2,4E-04 | 361141 |
| Major dietary changes in the last 5 years: Yes, because of illness | 0,183 | 0,032 | 5,6 | 3,4E-07 | 360294 |
| Suffer from 'nerves' | 0,182 | 0,033 | 5,5 | 6,3E-07 | 348082 |
| Pain type(s) experienced in last month: Hip pain | 0,182 | 0,034 | 5,3 | 1,8E-06 | 360391 |
| Nervous feelings | 0,180 | 0,029 | 6,1 | 2,2E-08 | 351829 |
| Miserableness | 0,180 | 0,029 | 6,1 | 2,7E-08 | 355182 |
| Injury, poisoning and certain other consequences of external causes | 0,180 | 0,041 | 4,4 | 1,6E-04 | 361194 |
| Sensitivity / hurt feelings | 0,179 | 0,030 | 6,1 | 3,1E-08 | 350821 |
| Shoulder lesions | 0,179 | 0,052 | 3,4 | 5,4E-03 | 361194 |
| Diagnoses - main ICD10: M75 Shoulder lesions | 0,179 | 0,052 | 3,4 | 5,5E-03 | 361194 |
| Manifestations of mania or irritability: My thoughts were racing | 0,178 | 0,050 | 3,6 | 3,5E-03 | 114422 |
| Diseases of the blood and blood-forming organs and certain disorders involving the immune mechanism | 0,177 | 0,065 | 2,7 | 3,9E-02 | 361194 |
| Breathing problems during period of job: Yes | 0,177 | 0,055 | 3,2 | 1,0E-02 | 91149 |
| Manifestations of mania or irritability: I was more talkative than usual | 0,172 | 0,054 | 3,2 | 1,0E-02 | 114422 |
| Ever had prostate specific antigen (PSA) test | 0,171 | 0,044 | 3,9 | 1,2E-03 | 158108 |
| Illness, injury, bereavement, stress in last 2 years: Serious illness, injury or assault to yourself | 0,170 | 0,042 | 4,1 | 6,2E-04 | 358836 |
| Manifestations of mania or irritability: I was more active than usual | 0,170 | 0,060 | 2,8 | 2,8E-02 | 114422 |
| Diagnoses - main ICD10: N20 Calculus of kidney and ureter | 0,169 | 0,049 | 3,4 | 5,2E-03 | 361194 |
| Ulcerative colitis, NAS | 0,168 | 0,058 | 2,9 | 2,3E-02 | 361194 |
| Breathing problems responsible for leaving job: No | 0,168 | 0,053 | 3,2 | 1,1E-02 | 91149 |
| Treatment/medication code: thyroxine product | 0,167 | 0,060 | 2,8 | 3,1E-02 | 361141 |
| Doctor diagnosed hayfever or allergic rhinitis | 0,165 | 0,040 | 4,1 | 5,0E-04 | 91787 |
| Mood [affective] disorders | 0,164 | 0,062 | 2,6 | 4,7E-02 | 361194 |
| Mood disorders | 0,164 | 0,062 | 2,6 | 4,7E-02 | 361194 |
| Dorsalgia | 0,164 | 0,043 | 3,8 | 1,5E-03 | 361194 |
| Diagnoses - main ICD10: M54 Dorsalgia | 0,164 | 0,043 | 3,8 | 1,5E-03 | 361194 |
| Diagnoses - main ICD10: K51 Ulcerative colitis | 0,163 | 0,059 | 2,8 | 3,4E-02 | 361194 |
| Ever had prolonged loss of interest in normal activities | 0,161 | 0,032 | 5,0 | 1,0E-05 | 117727 |
| Pain type(s) experienced in last month: Knee pain | 0,161 | 0,029 | 5,5 | 6,7E-07 | 360391 |
| Diagnoses - main ICD10: K21 Gastro-oesophageal reflux disease | 0,159 | 0,049 | 3,2 | 9,3E-03 | 361194 |
| Ever highly irritable/argumentative for 2 days | 0,157 | 0,042 | 3,8 | 1,9E-03 | 117359 |
| Low back pain | 0,157 | 0,054 | 2,9 | 2,4E-02 | 361194 |
| Non-cancer illness code, self-reported: kidney stone/ureter stone/bladder stone | 0,156 | 0,053 | 2,9 | 2,4E-02 | 361141 |
| Hearing difficulty/problems with background noise | 0,155 | 0,027 | 5,8 | 1,3E-07 | 353983 |
| Leg pain on walking | 0,154 | 0,042 | 3,7 | 2,3E-03 | 118905 |
| Neoplasms | 0,154 | 0,044 | 3,5 | 3,7E-03 | 361194 |
| ADHD | 0,153 | 0,034 | 4,5 | 1,1E-04 | 55374 |
| Treatment/medication code: co-codamol | 0,153 | 0,048 | 3,2 | 1,1E-02 | 361141 |
| Irritability | 0,152 | 0,029 | 5,3 | 2,0E-06 | 345231 |
| Current employment status: Unable to work because of sickness or disability | 0,152 | 0,035 | 4,3 | 2,4E-04 | 359931 |
| Headaches for 3+ months | 0,150 | 0,043 | 3,5 | 4,6E-03 | 70181 |
| Milk type used: Soya | 0,149 | 0,049 | 3,0 | 1,7E-02 | 360806 |
| Illnesses of siblings: Heart disease | 0,148 | 0,042 | 3,5 | 3,9E-03 | 280784 |
| Guilty feelings | 0,148 | 0,031 | 4,8 | 2,7E-05 | 351907 |
| Hernia | 0,147 | 0,042 | 3,5 | 3,9E-03 | 361194 |
| Nerve, nerve root and plexus disorders | 0,144 | 0,042 | 3,5 | 4,7E-03 | 361194 |
| Fed-up feelings | 0,143 | 0,027 | 5,3 | 2,0E-06 | 353764 |
| Carpal tunnel syndrome | 0,142 | 0,042 | 3,4 | 5,5E-03 | 361194 |

| | | | | | |
|---|---|---|---|---|---|
| Diseases of the musculoskeletal system and connective tissue | 0,142 | 0,036 | 3,9 | 9,9E-04 | 361194 |
| Victim of sexual assault | 0,139 | 0,047 | 3,0 | 2,0E-02 | 116671 |
| Fractured/broken bones in last 5 years | 0,135 | 0,041 | 3,3 | 8,2E-03 | 359241 |
| Ever unenthusiastic/disinterested for a whole week | 0,134 | 0,037 | 3,6 | 2,7E-03 | 115145 |
| Illnesses of mother: Chronic bronchitis/emphysema | 0,133 | 0,051 | 2,6 | 4,8E-02 | 331008 |
| Diagnoses - main ICD10: G56 Mononeuropathies of upper limb | 0,129 | 0,042 | 3,0 | 1,6E-02 | 361194 |
| Diseases of the nervous system | 0,129 | 0,042 | 3,1 | 1,6E-02 | 361194 |
| Worry too long after embarrassment | 0,129 | 0,030 | 4,3 | 2,7E-04 | 346527 |
| Bipolar Disorder | 0,129 | 0,028 | 4,7 | 4,5E-05 | 51710 |
| Non-cancer illness code, self-reported: high cholesterol | 0,127 | 0,031 | 4,1 | 6,2E-04 | 361141 |
| Attendance/disability/mobility allowance: Disability living allowance | 0,120 | 0,032 | 3,8 | 1,6E-03 | 358597 |
| Attendance/disability/mobility allowance: Blue badge | 0,119 | 0,038 | 3,2 | 1,2E-02 | 358597 |
| Ever smoked | 0,118 | 0,025 | 4,8 | 2,6E-05 | 359751 |
| Hearing difficulty/problems: Yes | 0,117 | 0,032 | 3,7 | 2,6E-03 | 346635 |
| Medication for cholesterol, blood pressure or diabetes: Cholesterol lowering medication | 0,112 | 0,035 | 3,2 | 1,0E-02 | 165340 |
| #Other joint disorders | 0,112 | 0,038 | 3,0 | 2,0E-02 | 361194 |
| Qualifications: CSEs or equivalent | 0,110 | 0,035 | 3,1 | 1,3E-02 | 357549 |
| Wheeze or whistling in the chest in last year | 0,110 | 0,028 | 4,0 | 8,3E-04 | 354523 |
| Loneliness, isolation | 0,106 | 0,032 | 3,3 | 8,1E-03 | 355583 |
| Cardiac arrhytmias, COPD co-morbidities | 0,105 | 0,037 | 2,9 | 2,7E-02 | 361194 |
| Smoking status: Previous | 0,104 | 0,027 | 3,8 | 1,8E-03 | 359706 |
| Illnesses of father: Heart disease | 0,102 | 0,035 | 2,9 | 2,5E-02 | 318570 |
| Schizophrenia | 0,101 | 0,024 | 4,3 | 2,6E-04 | 150064 |
| Illness, injury, bereavement, stress in last 2 years: Financial difficulties | 0,100 | 0,032 | 3,2 | 1,2E-02 | 358836 |
| Treatment/medication code: levothyroxine sodium | 0,094 | 0,035 | 2,7 | 4,1E-02 | 361141 |
| Bone fractures | 0,093 | 0,036 | 2,6 | 4,8E-02 | 426824 |
| Medication for cholesterol, blood pressure, diabetes, or take exogenous hormones: Cholesterol lowering medication | 0,092 | 0,035 | 2,7 | 4,4E-02 | 193148 |
| Blood clot, DVT, bronchitis, emphysema, asthma, rhinitis, eczema, allergy diagnosed by doctor: Hayfever, allergic rhinitis or eczema | 0,091 | 0,026 | 3,5 | 3,6E-03 | 360527 |
| Own or rent accommodation lived in: Rent - from local authority, local council, housing association | 0,086 | 0,033 | 2,6 | 4,8E-02 | 356340 |
| Lymphocyte percentage of white cells | 0,082 | 0,028 | 3,0 | 2,0E-02 | 173496 |
| Number of incorrect matches in round | 0,080 | 0,026 | 3,1 | 1,5E-02 | 360686 |
| Lymphocyte percentage | 0,072 | 0,024 | 3,0 | 1,9E-02 | 349861 |
| Time to complete round | 0,069 | 0,025 | 2,7 | 3,9E-02 | 354739 |
| Qualifications: College or University degree | -0,057 | 0,021 | -2,8 | 3,4E-02 | 357549 |
| Neutrophill percentage | -0,068 | 0,025 | -2,7 | 3,7E-02 | 349861 |
| Snoring | -0,079 | 0,027 | -2,9 | 2,7E-02 | 336320 |
| Cognitive Performance | -0,081 | 0,022 | -3,8 | 1,8E-03 | 257841 |
| Smoking status: Never | -0,082 | 0,025 | -3,3 | 7,3E-03 | 359706 |
| Neutrophil percentage of white cells | -0,084 | 0,029 | -2,9 | 2,4E-02 | 173507 |
| Blood clot, DVT, bronchitis, emphysema, asthma, rhinitis, eczema, allergy diagnosed by doctor: None of the above | -0,087 | 0,026 | -3,4 | 6,4E-03 | 360527 |
| Medication for cholesterol, blood pressure or diabetes: None of the above | -0,092 | 0,033 | -2,8 | 3,4E-02 | 165340 |
| Fluid intelligence score | -0,095 | 0,027 | -3,5 | 3,9E-03 | 117131 |
| Mouth/teeth dental problems: None of the above | -0,100 | 0,035 | -2,9 | 2,7E-02 | 359841 |
| Types of transport used (excluding work): Public transport | -0,101 | 0,037 | -2,8 | 3,4E-02 | 359324 |
| Medication for cholesterol, blood pressure, diabetes, or take exogenous hormones: None of the above | -0,103 | 0,034 | -3,0 | 1,9E-02 | 193148 |
| Age started oral contraceptive pill | -0,107 | 0,034 | -3,1 | 1,3E-02 | 154112 |
| Major dietary changes in the last 5 years: No | -0,115 | 0,032 | -3,6 | 3,0E-03 | 360294 |
| Age at first live birth | -0,117 | 0,027 | -4,3 | 2,5E-04 | 131987 |
| Hearing difficulty/problems: No | -0,117 | 0,032 | -3,7 | 2,6E-03 | 346635 |
| Vitamin and mineral supplements: None of the above | -0,122 | 0,034 | -3,6 | 2,9E-03 | 359245 |
| Attendance/disability/mobility allowance: None of the above | -0,132 | 0,033 | -4,0 | 7,7E-04 | 358597 |
| Reason for reducing amount of alcohol drunk: Other reason | -0,145 | 0,040 | -3,6 | 3,0E-03 | 134033 |
| Age started smoking in former smokers | -0,150 | 0,053 | -2,8 | 3,1E-02 | 88898 |
| Illness, injury, bereavement, stress in last 2 years: None of the above | -0,161 | 0,035 | -4,5 | 8,7E-05 | 358836 |
| Age at first episode of depression | -0,204 | 0,069 | -3,0 | 2,0E-02 | 61033 |
| Job coding: civil engineer | -0,214 | 0,079 | -2,7 | 4,0E-02 | 89866 |
| Age started smoking in current smokers | -0,220 | 0,068 | -3,2 | 9,9E-03 | 27291 |
| Bipolar and major depression status: No Bipolar or Depression | -0,221 | 0,041 | -5,4 | 1,1E-06 | 86895 |
| Illnesses of siblings: None of the above (group 2) | -0,227 | 0,057 | -3,9 | 9,4E-04 | 284896 |
| Medication for pain relief, constipation, heartburn: None of the above | -0,233 | 0,024 | -9,7 | 3,4E-20 | 357084 |
| Job coding: heavy goods vehicle (hgv) driver, lorry or truck driver, tanker driver, haulage driver | -0,233 | 0,087 | -2,7 | 4,3E-02 | 89866 |
| Tinnitus: No, never | -0,248 | 0,039 | -6,4 | 3,7E-09 | 117882 |
| Pain type(s) experienced in last month: None of the above | -0,274 | 0,025 | -11,1 | 2,9E-26 | 360391 |
| Illnesses of mother: None of the above (group 2) | -0,279 | 0,061 | -4,6 | 6,3E-05 | 334401 |
