## Supplementary Table 7 for "Genome-wide analysis of 944,133 individuals provides insights into the etiology of hemorrhoidal disease"

**Supplementary Table 7. Summary of HEM genes mapping and prioritization.**

This table contains detailed information of all 819 HEM mapped genes and the criteria for gene prioritization (see **Results** and **Online Methods).**

**Gene symbol:** gene ID extracted from HUGO Gene Nomenclature Committee; **EntrezID:** gene ID extracted from Entrez Gene database; **EnsemblID:** gene ID extracted from ensembl database GRCh38.p13; **Gene type:** the biotype of the gene annotated by ensembl database GRCh38.p13; **Locus number:** the sequential number of the HEM HWAS loci; **Lead SNP (rsID):** rs ID retrieved from NCBI's dbSNP build v150**; Prioritized HEM genes**: a binary variable indicating whether the gene is a prioritized HEM gene (1=yes, 0=no); **Gene mapping**: specifying the gene mapping method for each gene among four mapping categories (FUMA_positional, FUMA_eQTL, MAGMA, DEPICT, 1=yes, 0=no, see **Online Methods**); **DEPICT prioritized:** indicating whether a gene is prioritized by DEPICT gene prioritization algorithm (1=yes, 0=no); **Linked to fine mapped variant (probabliity>50%):** Genes that are linked to a fine mapped variant (posterior probability > 50%) by either significant eQTL associations only (labelled "eQTL") or significant eQTL associations plus physically containing the fine mapped coding variant (labelled "coding variant & eQTL"). The rs ID of the linked fine mapped variant and its posterior probability are shown in the brackets. **Expression in HEM tissue:** Genes that were expressed (labelled "E") or not expressed (labelled "NE") from RNAseq results of HEM tissues. Differentially expressed genes were highlighted with either "E+" (for overexpressed genes) or "E-" (for underexpressed genes) according to the RNAseq analysis results comparing tissue expression profiles between HEM patients and healthy controls. Genes were labelled with "(UQ)" if were highly expressed (in upper quartile). **WGCNA module**: indicating whether a gene is included in any of the significant gene co-expression modules of HEM tissue (M1, M4 or M7, see **Online Methods**), a suffix "_hub" after the module names (e.g. M1_hub) represents a hub gene within the gene co-expression module. **Mouse phenotypes from MGI**: type of mammalian phenotype categories that are associated with the gene mutations from Mouse Genome Informatics Database (MGI, http://www.informatics.jax.org/). **OMIM Phenotypes:** a list of genetic disorders that are associated with the gene mutations, information extracted from Online Mendelian Inheritance in Man (OMIM). **"-"**: data is not available.

| | | | | | | | Gene mapping | | | | Gene prioritization | | | | | |
|---|---|---|---|---|---|---|---|---|---|---|---|---|---|---|---|---|
| **Gene symbol** | **EntrezID** | **EnsemblID** | **Gene type** | **Locus number** | **Lead SNP (rsID)** | **Prioritized HEM genes** | **FUMA_positional** | **FUMA_eQTL** | **MAGMA** | **DEPICT** | **DEPICT prioritized** | **Linked to fine mapped variant (probabliity>50%)** | **Expression in HEM tissue** | **WGCNA module** | **Mouse phenotypes from MGI** | **OMIM Phenotypes** |
| BMP8B | 656 | ENSG00000116985 | protein_coding | 1 | rs11585073 | 0 | 1 | 1 | 0 | 1 | 0 | - | E | - | cellular; endocrine/exocrine gland; growth/size/body region; embryo; reproductive system; mortality/aging; | - |
| TRIT1 | 54802 | ENSG00000043514 | protein_coding | 1 | rs11585073 | 0 | 1 | 1 | 0 | 0 | 0 | - | E (UQ) | - | growth/size/body region; cardiovascular system; mortality/aging; behavior/neurological; embryo; | Combined oxidative phosphorylation deficiency 35, 617873 (3), Autosomal recessive |
| RP1-118J21.25 | 101929536 | ENSG00000261798 | lncRNA | 1 | rs11585073 | 0 | 1 | 0 | 0 | 0 | 0 | - | E | - | - | - |
| HPCAL4 | 51440 | ENSG00000116983 | protein_coding | 1 | rs11585073 | 0 | 0 | 1 | 0 | 0 | 0 | - | E | - | behavior/neurological; integument; | - |
| MYCL | 4610 | ENSG00000116990 | protein_coding | 1 | rs11585073 | 0 | 0 | 1 | 0 | 0 | 0 | - | E | - | mortality/aging; hematopoietic system; immune system; | - |
| OXCT2 | 64064 | ENSG00000198754 | protein_coding | 1 | rs11585073 | 0 | 0 | 1 | 0 | 0 | 0 | - | E | M1 | - | - |
| PPIE | 10450 | ENSG00000084072 | protein_coding | 1 | rs11585073 | 0 | 0 | 1 | 0 | 0 | 0 | - | E (UQ) | - | - | - |
| RP1-39G22.7 | - | ENSG00000259943 | lncRNA | 1 | rs11585073 | 0 | 0 | 1 | 0 | 0 | 0 | - | E | M1 | - | - |
| SLC44A3 | 126969 | ENSG00000143036 | protein_coding | 2 | rs11578225 | 0 | 1 | 1 | 1 | 1 | 0 | - | E (UQ) | M4 | vision/eye; cardiovascular system; | - |
| LINC01057 | 101928079 | ENSG00000224081 | transcribed_processed_pseudogene | 2 | rs11578225 | 0 | 1 | 1 | 0 | 1 | 0 | - | E | M4 | - | - |
| MIR378G | 100616321 | ENSG00000263526 | miRNA | 2 | rs11578225 | 0 | 1 | 0 | 0 | 0 | 0 | - | E | - | - | - |
| RP4-639F20.1 | 729970 | ENSG00000235501 | lncRNA | 2 | rs11578225 | 0 | 0 | 1 | 0 | 0 | 0 | - | E | M1 | - | - |
| F5 | 2153 | ENSG00000198734 | protein_coding | 3 | rs145163454 | 0 | 1 | 0 | 0 | 1 | 0 | - | E | - | integument; homeostasis/metabolism; embryo; behavior/neurological; liver/biliary system; mortality/aging; cardiovascular system; | {Pregnancy loss, recurrent, susceptibility to, 1}, 614389 (3), Autosomal dominant; Thrombophilia due to activated protein C resistance, 188055 (3), Autosomal dominant; {Thrombophilia, susceptibility to, due to factor V Leiden}, 188055 (3), Autosomal dominant; Factor V deficiency, 227400 (3), Autosomal recessive; {Budd-Chiari syndrome}, 600880 (3), Autosomal recessive; {Stroke, ischemic, susceptibility to}, 601367 (3), Multifactorial |
| ATP1B1 | 481 | ENSG00000143153 | protein_coding | 3 | rs145163454 | 0 | 1 | 1 | 0 | 0 | 0 | - | E (UQ) | M4 | cardiovascular system; mortality/aging; respiratory system; homeostasis/metabolism; muscle; | [Blood pressure regulation QTL], 145500 (2), Multifactorial |
| NME7 | 29922 | ENSG00000143156 | protein_coding | 3 | rs145163454 | 0 | 1 | 1 | 0 | 0 | 0 | - | E (UQ) | M1 | nervous system; skeleton; respiratory system; mortality/aging; craniofacial; cellular; growth/size/body region; | - |
| SLC19A2 | 10560 | ENSG00000117479 | protein_coding | 3 | rs145163454 | 0 | 1 | 1 | 0 | 0 | 0 | - | E | M7 | immune system; hearing/vestibular/ear; nervous system; hematopoietic system; reproductive system; mortality/aging; behavior/neurological; homeostasis/metabolism; cellular; endocrine/exocrine gland; | Thiamine-responsive megaloblastic anemia syndrome, 249270 (3), Autosomal recessive |
| BLZF1 | 8548 | ENSG00000117475 | protein_coding | 3 | rs145163454 | 0 | 1 | 0 | 0 | 0 | 0 | - | E (UQ) | - | - | - |
| CCDC181 | 57821 | ENSG00000117477 | protein_coding | 3 | rs145163454 | 0 | 1 | 0 | 0 | 0 | 0 | - | E | M1 | - | - |
| RP4-800F24.1 | - | ENSG00000235575 | lncRNA | 3 | rs145163454 | 0 | 1 | 0 | 0 | 0 | 0 | - | NE | - | - | - |
| RP5-1018K9.1 | 101928596 | ENSG00000237707 | lncRNA | 3 | rs145163454 | 0 | 1 | 0 | 0 | 0 | 0 | - | NE | - | - | - |
| METTL18 | 92342 | ENSG00000171806 | protein_coding | 3 | rs145163454 | 0 | 0 | 1 | 0 | 0 | 0 | - | E | - | - | - |
| MDM4 | 4194 | ENSG00000198625 | protein_coding | 4 | rs4951080 | 0 | 1 | 1 | 1 | 1 | 0 | - | E (UQ) | - | muscle; cellular; homeostasis/metabolism; integument; growth/size/body region; endocrine/exocrine gland; pigmentation; limbs/digits/tail; nervous system; immune system; embryo; mortality/aging; hematopoietic system; cardiovascular system; | ?Bone marrow failure syndrome 6, 618849 (3), Autosomal dominant |
| LRRN2 | 10446 | ENSG00000170382 | protein_coding | 4 | rs4951080 | 0 | 1 | 1 | 0 | 1 | 0 | - | E | M7 | behavior/neurological; | - |
| PIK3C2B | 5287 | ENSG00000133056 | protein_coding | 4 | rs4951080 | 0 | 1 | 1 | 0 | 1 | 0 | - | E (UQ) | M4 | - | - |
| RP11-430C7.4 | - | ENSG00000240710 | lncRNA | 4 | rs4951080 | 0 | 1 | 1 | 0 | 1 | 0 | - | NE | - | - | - |
| RP11-430C7.5 | - | ENSG00000240219 | lncRNA | 4 | rs4951080 | 0 | 1 | 1 | 0 | 0 | 0 | - | E | - | - | - |
| RNA5SP74 | 100873308 | ENSG00000200408 | rRNA_pseudogene | 4 | rs4951080 | 0 | 1 | 0 | 0 | 0 | 0 | - | NE | - | - | - |
| PPP1R15B | 84919 | ENSG00000158615 | protein_coding | 4 | rs4951080 | 0 | 0 | 1 | 0 | 0 | 0 | - | E (UQ) | - | integument; growth/size/body region; mortality/aging; hematopoietic system; liver/biliary system; behavior/neurological; | Microcephaly, short stature, and impaired glucose metabolism 2, 616817 (3), Autosomal recessive |
| RP11-392O17.1 | - | ENSG00000228536 | lncRNA | 5 | rs2605097 | 0 | 0 | 1 | 0 | 1 | 0 | - | NE | - | - | - |
| GDF7 | 151449 | ENSG00000143869 | protein_coding | 6 | rs7594056 | 0 | 1 | 1 | 1 | 0 | 0 | - | E | - | endocrine/exocrine gland; mortality/aging; reproductive system; nervous system; embryo; | - |
| C2orf43 | 60526 | ENSG00000118961 | protein_coding | 6 | rs7594056 | 0 | 1 | 1 | 0 | 1 | 0 | - | E (UQ) | - | - | - |
| RP11-130L8.1 | - | ENSG00000270100 | lncRNA | 6 | rs7594056 | 0 | 1 | 1 | 0 | 0 | 0 | - | E | - | - | - |
| BIRC6 | 57448 | ENSG00000115760 | protein_coding | 7 | rs6723226 | 0 | 1 | 1 | 0 | 1 | 0 | - | E (UQ) | M4 | endocrine/exocrine gland; growth/size/body region; integument; homeostasis/metabolism; cellular; nervous system; hematopoietic system; cardiovascular system; mortality/aging; liver/biliary system; embryo; immune system; | - |
| YIPF4 | 84272 | ENSG00000119820 | protein_coding | 7 | rs6723226 | 0 | 1 | 1 | 0 | 1 | 0 | - | E (UQ) | - | - | - |
| NLRC4 | 58484 | ENSG00000091106 | protein_coding | 7 | rs6723226 | 0 | 1 | 1 | 0 | 0 | 0 | - | E | M1 | integument; cellular; homeostasis/metabolism; hematopoietic system; skeleton; immune system; | Autoinflammation with infantile enterocolitis, 616050 (3), Autosomal dominant; ?Familial cold autoinflammatory syndrome 4, 616115 (3), Autosomal dominant |
| AL133245.2 | - | ENSG00000272754 | lncRNA | 7 | rs6723226 | 0 | 1 | 0 | 0 | 0 | 0 | - | E | - | - | - |
| BIRC6-AS1 | 100874009 | ENSG00000230046 | lncRNA | 7 | rs6723226 | 0 | 1 | 0 | 0 | 0 | 0 | - | NE | - | - | - |
| MIR558 | 693143 | ENSG00000207653 | miRNA | 7 | rs6723226 | 0 | 1 | 0 | 0 | 0 | 0 | - | NE | - | - | - |
| TTC27 | 55622 | ENSG00000018699 | protein_coding | 7 | rs6723226 | 0 | 1 | 0 | 0 | 0 | 0 | - | E (UQ) | - | - | - |
| LINC00486 | 285045 | ENSG00000230876 | lncRNA | 7 | rs6723226 | 0 | 0 | 1 | 0 | 0 | 0 | - | NE | - | - | - |
| SPAST | 6683 | ENSG00000021574 | protein_coding | 7 | rs6723226 | 0 | 0 | 1 | 0 | 0 | 0 | - | E | M4 | growth/size/body region; reproductive system; behavior/neurological; nervous system; | Spastic paraplegia 4, autosomal dominant, 182601 (3), Autosomal dominant |
| XDH | 7498 | ENSG00000158125 | protein_coding | 7 | rs6723226 | 0 | 0 | 1 | 0 | 0 | 0 | - | E (UQ) | - | mortality/aging; immune system; renal/urinary system; integument; growth/size/body region; adipose tissue; endocrine/exocrine gland; cellular; homeostasis/metabolism; muscle; | Xanthinuria, type I, 278300 (3), Autosomal recessive |
| CRIM1 | 51232 | ENSG00000150938 | protein_coding | 8 | rs4670149 | 0 | 1 | 1 | 1 | 0 | 0 | - | E (UQ) | - | limbs/digits/tail; nervous system; digestive/alimentary; vision/eye; immune system; renal/urinary system; skeleton; embryo; liver/biliary system; behavior/neurological; mortality/aging; cardiovascular system; muscle; craniofacial; cellular; homeostasis/metabolism; integument; endocrine/exocrine gland; | - |
| FEZ2 | 9637 | ENSG00000171055 | protein_coding | 8 | rs4670149 | 0 | 1 | 1 | 0 | 1 | 0 | - | E (UQ) | - | - | - |
| SRBD1 | 55133 | ENSG00000068784 | protein_coding | 9 | rs728327 | 1 | 1 | 1 | 1 | 1 | 0 | eQTL (rs728327, PP=0.669) | E | - | mortality/aging; | - |
| CAMKMT | 79823 | ENSG00000143919 | protein_coding | 9 | rs728327 | 0 | 0 | 1 | 0 | 0 | 0 | - | E | - | adipose tissue; growth/size/body region; homeostasis/metabolism; cellular; muscle; hematopoietic system; behavior/neurological; skeleton; | - |
| PIGF | 5281 | ENSG00000151665 | protein_coding | 9 | rs728327 | 0 | 0 | 1 | 0 | 0 | 0 | - | E (UQ) | M4 | embryo; mortality/aging; skeleton; adipose tissue; growth/size/body region; | - |
| EHBP1 | 23301 | ENSG00000115504 | protein_coding | 10 | rs4671051 | 0 | 1 | 0 | 1 | 1 | 0 | - | E (UQ) | - | - | {Prostate cancer, hereditary, 12}, 611868 (3) |
| OTX1 | 5013 | ENSG00000115507 | protein_coding | 10 | rs4671051 | 1 | 1 | 1 | 0 | 1 | 0 | - | E | M7_hub | growth/size/body region; endocrine/exocrine gland; homeostasis/metabolism; vision/eye; craniofacial; reproductive system; mortality/aging; behavior/neurological; skeleton; nervous system; hearing/vestibular/ear; digestive/alimentary; | - |
| AC007098.1 | - | ENSG00000226605 | lncRNA | 10 | rs4671051 | 0 | 1 | 0 | 0 | 0 | 0 | - | NE | - | - | - |
| AC009501.4 | 100132215 | ENSG00000231609 | lncRNA | 10 | rs4671051 | 0 | 1 | 0 | 0 | 0 | 0 | - | E | M7 | - | - |
| MDH1 | 4190 | ENSG00000014641 | polymorphic_pseudogene | 10 | rs4671051 | 0 | 0 | 1 | 1 | 0 | 0 | - | E (UQ) | - | mortality/aging; homeostasis/metabolism; | - |
| AC092155.4 | - | ENSG00000226622 | lncRNA | 10 | rs4671051 | 0 | 0 | 1 | 0 | 1 | 0 | - | NE | - | - | - |
| DBIL5P2 | 100169989 | ENSG00000242412 | nscribed_processed_pseudoge | 10 | rs4671051 | 0 | 0 | 0 | 0 | 1 | 0 | - | E | - | - | - |
| TMEM17 | 200728 | ENSG00000186889 | protein_coding | 10 | rs4671051 | 0 | 0 | 1 | 0 | 0 | 0 | - | E | - | - | - |
| WDPCP | 51057 | ENSG00000143951 | protein_coding | 10:11 | rs4671051:rs34532102 | 0 | 0 | 1 | 1 | 1 | 0 | - | E | - | vision/eye; digestive/alimentary; limbs/digits/tail; nervous system; hearing/vestibular/ear; renal/urinary system; respiratory system; embryo; cardiovascular system; mortality/aging; craniofacial; cellular; growth/size/body region; | ?Bardet-Biedl syndrome 15, 615992 (3), Autosomal recessive; ?Congenital heart defects, hamartomas of tongue, and polysyndactyly, 217085 (3), Autosomal recessive |
| SERTAD2 | 9792 | ENSG00000179833 | protein_coding | 11 | rs34532102 | 1 | 1 | 0 | 1 | 1 | 1 | - | E (UQ) | - | immune system; liver/biliary system; hematopoietic system; adipose tissue; cellular; homeostasis/metabolism; integument; growth/size/body region; | - |
| AC010987.5 | - | ENSG00000235495 | lncRNA | 12 | rs2861709 | 0 | 1 | 0 | 0 | 0 | 0 | - | NE | - | - | - |
| ETAA1 | 54465 | ENSG00000143971 | protein_coding | 12 | rs2861709 | 0 | 0 | 0 | 1 | 0 | 0 | - | E | M1 | - | - |
| AC010987.6 | 101927701 | ENSG00000237013 | lncRNA | 12 | rs2861709 | 0 | 0 | 0 | 0 | 1 | 0 | - | E | - | - | - |
| AC015969.3 | - | ENSG00000203395 | lncRNA | 12 | rs2861709 | 0 | 0 | 1 | 0 | 0 | 0 | - | NE | - | - | - |
| PLEK | 5341 | ENSG00000115956 | protein_coding | 12 | rs2861709 | 1 | 0 | 1 | 0 | 0 | 0 | - | E- (UQ) | - | hematopoietic system; homeostasis/metabolism; | - |
| MERTK | 10461 | ENSG00000153208 | protein_coding | 13 | rs57116599 | 0 | 1 | 1 | 1 | 1 | 0 | - | E | - | cardiovascular system; hematopoietic system; mortality/aging; reproductive system; pigmentation; digestive/alimentary; nervous system; vision/eye; immune system; endocrine/exocrine gland; homeostasis/metabolism; cellular; | Retinitis pigmentosa 38, 613862 (3), Autosomal recessive |
| TMEM87B | 84910 | ENSG00000153214 | protein_coding | 13 | rs57116599 | 0 | 1 | 1 | 0 | 1 | 0 | - | E (UQ) | - | - | - |
| AC012442.5 | - | ENSG00000243389 | lncRNA | 13 | rs57116599 | 0 | 1 | 1 | 0 | 0 | 0 | - | NE | - | - | - |
| NT5DC4 | 284958 | ENSG00000144130 | protein_coding | 13 | rs57116599 | 0 | 1 | 1 | 0 | 0 | 0 | - | E | - | - | - |
| SLC20A1 | 6574 | ENSG00000144136 | protein_coding | 13 | rs57116599 | 1 | 1 | 1 | 0 | 0 | 0 | - | E+ (UQ) | - | homeostasis/metabolism; cellular; growth/size/body region; integument; immune system; skeleton; embryo; liver/biliary system; cardiovascular system; hematopoietic system; mortality/aging; | - |
| CKAP2L | 150468 | ENSG00000169607 | protein_coding | 13 | rs57116599 | 0 | 1 | 0 | 0 | 0 | 0 | - | E | - | - | Filippi syndrome, 272440 (3), Autosomal recessive |
| IL1A | 3552 | ENSG00000115008 | protein_coding | 13 | rs57116599 | 0 | 1 | 0 | 0 | 0 | 0 | - | E | - | immune system; cardiovascular system; neoplasm; homeostasis/metabolism; integument; | - |
| IL1F10 | 84639 | ENSG00000136697 | protein_coding | 13 | rs57116599 | 0 | 1 | 0 | 0 | 0 | 0 | - | E | M7 | - | - |
| IL36A | 27179 | ENSG00000136694 | protein_coding | 13 | rs57116599 | 1 | 1 | 0 | 0 | 0 | 0 | - | E+ | M7 | limbs/digits/tail; immune system; homeostasis/metabolism; growth/size/body region; integument; | - |
| IL36B | 27177 | ENSG00000136696 | protein_coding | 13 | rs57116599 | 0 | 1 | 0 | 0 | 0 | 0 | - | E | M7 | - | - |
| IL36RN | 26525 | ENSG00000136695 | protein_coding | 13 | rs57116599 | 0 | 1 | 0 | 0 | 0 | 0 | - | E (UQ) | M7 | homeostasis/metabolism; integument; immune system; hematopoietic system; | Psoriasis 14, pustular, 614204 (3), Autosomal recessive |
| IL37 | 27178 | ENSG00000125571 | protein_coding | 13 | rs57116599 | 0 | 1 | 0 | 0 | 0 | 0 | - | E | - | - | - |
| AC016683.5 | - | ENSG00000234174 | lncRNA | 13 | rs57116599 | 0 | 0 | 1 | 0 | 0 | 0 | - | NE | - | - | - |

| | | | | | | | | | | | | | | | | |
|---|---|---|---|---|---|---|---|---|---|---|---|---|---|---|---|---|
| AC079922.3 | - | ENSG00000237753 | lncRNA | 13 | rs57116599 | 0 | 0 | 1 | 0 | 0 | 0 | - | E | - | - | - |
| CHCHD5 | 84269 | ENSG00000125611 | protein_coding | 13 | rs57116599 | 0 | 0 | 1 | 0 | 0 | 0 | - | E (UQ) | - | - | - |
| FBLN7 | 129804 | ENSG00000144152 | protein_coding | 13 | rs57116599 | 0 | 0 | 1 | 0 | 0 | 0 | - | E | - | cellular; homeostasis/metabolism; renal/urinary system; | - |
| IL1RN | 3557 | ENSG00000136689 | protein_coding | 13 | rs57116599 | 0 | 0 | 1 | 0 | 0 | 0 | - | E (UQ) | M7 | skeleton; immune system; cardiovascular system; hematopoietic system; integument; mortality/aging; reproductive system; homeostasis/metabolism; cellular; growth/size/body region; | {Gastric cancer risk after H. pylori infection}, 137215 (3), Autosomal dominant; {Microvascular complications of diabetes 4}, 612628 (3); Interleukin 1 receptor antagonist deficiency, 612852 (3), Autosomal recessive |
| PAX8 | 7849 | ENSG00000125618 | protein_coding | 13 | rs57116599 | 0 | 0 | 1 | 0 | 0 | 0 | - | E | - | endocrine/exocrine gland; growth/size/body region; craniofacial; homeostasis/metabolism; cellular; embryo; cardiovascular system; reproductive system; mortality/aging; vision/eye; limbs/digits/tail; hearing/vestibular/ear; nervous system; skeleton; | Hypothyroidism, congenital, due to thyroid dysgenesis or hypoplasia, 218700 (3), Autosomal dominant |
| PAX8-AS1 | 654433 | ENSG00000189223 | lncRNA | 13 | rs57116599 | 0 | 0 | 1 | 0 | 0 | 0 | - | E (UQ) | M4 | - | - |
| POLR1B | 84172 | ENSG00000125630 | protein_coding | 13 | rs57116599 | 0 | 0 | 1 | 0 | 0 | 0 | - | E (UQ) | - | embryo; mortality/aging; cellular; | - |
| PSD4 | 23550 | ENSG00000125637 | protein_coding | 13 | rs57116599 | 0 | 0 | 1 | 0 | 0 | 0 | - | E (UQ) | M4 | - | - |
| RGPD8 | 727851 | ENSG00000169629 | protein_coding | 13 | rs57116599 | 0 | 0 | 1 | 0 | 0 | 0 | - | E | - | - | - |
| TTL | 150465 | ENSG00000114999 | protein_coding | 13 | rs57116599 | 0 | 0 | 1 | 0 | 0 | 0 | - | E (UQ) | M1 | homeostasis/metabolism; cellular; mortality/aging; nervous system; behavior/neurological; respiratory system; | - |
| TEX41 | 401014 | ENSG00000226674 | lncRNA | 14 | rs7423637 | 0 | 1 | 1 | 0 | 1 | 0 | - | E | - | - | - |
| ZEB2 | 9839 | ENSG00000169554 | protein_coding | 14 | rs7423637 | 0 | 0 | 1 | 0 | 0 | 0 | - | E | M1 | integument; endocrine/exocrine gland; craniofacial; cellular; embryo; mortality/aging; neoplasm; pigmentation; nervous system; vision/eye; | Mowat-Wilson syndrome, 235730 (3), Autosomal dominant |
| ACVR2A | 92 | ENSG00000121989 | protein_coding | 15 | rs7559714 | 0 | 1 | 1 | 1 | 0 | 0 | - | E | M4 | embryo; reproductive system; mortality/aging; vision/eye; digestive/alimentary; skeleton; cellular; homeostasis/metabolism; endocrine/exocrine gland; growth/size/body region; craniofacial; | - |
| ORC4 | 5000 | ENSG00000115947 | protein_coding | 15 | rs7559714 | 0 | 1 | 1 | 1 | 0 | 0 | - | E (UQ) | - | - | Meier-Gorlin syndrome 2, 613800 (3), Autosomal recessive |
| AC009480.3 | - | ENSG00000223911 | lncRNA | 15 | rs7559714 | 0 | 1 | 1 | 0 | 0 | 0 | - | NE | - | - | - |
| MBD5 | 55777 | ENSG00000204406 | protein_coding | 15 | rs7559714 | 0 | 1 | 1 | 0 | 0 | 0 | - | E | M1 | craniofacial; homeostasis/metabolism; integument; growth/size/body region; adipose tissue; endocrine/exocrine gland; limbs/digits/tail; nervous system; skeleton; behavior/neurological; liver/biliary system; mortality/aging; cardiovascular system; | Mental retardation, autosomal dominant 1, 156200 (3), Autosomal dominant |
| RNA5SP106 | 106478994 | ENSG00000253083 | rRNA_pseudogene | 15 | rs7559714 | 0 | 1 | 0 | 0 | 0 | 0 | - | NE | - | - | - |
| RNU6-1275P | 106480122 | ENSG00000212389 | snRNA | 15 | rs7559714 | 0 | 1 | 0 | 0 | 0 | 0 | - | NE | - | - | - |
| ITGB6 | 3694 | ENSG00000115221 | protein_coding | 16 | rs4233681 | 1 | 1 | 1 | 1 | 1 | 1 | - | E (UQ) | M4 | homeostasis/metabolism; integument; immune system; hematopoietic system; respiratory system; | Amelogenesis imperfecta, type IH, 616221 (3), Autosomal recessive |
| PLA2R1 | 22925 | ENSG00000153246 | protein_coding | 16 | rs4233681 | 0 | 1 | 1 | 1 | 1 | 0 | - | E | M7 | immune system; | - |
| RBMS1 | 5937 | ENSG00000153250 | protein_coding | 16 | rs4233681 | 0 | 1 | 1 | 1 | 0 | 0 | - | E (UQ) | - | embryo; reproductive system; mortality/aging; cardiovascular system; homeostasis/metabolism; integument; growth/size/body region; | - |
| AC009961.3 | 29994 | ENSG00000226266 | lncRNA | 16 | rs4233681 | 0 | 0 | 1 | 0 | 0 | 0 | - | E | - | - | - |
| LY75 | 4065 | ENSG00000054219 | protein_coding | 16 | rs4233681 | 0 | 0 | 1 | 0 | 0 | 0 | - | E | M4 | immune system; mortality/aging; hematopoietic system; | - |
| LY75-CD302 | 100526664 | ENSG00000248672 | protein_coding | 16 | rs4233681 | 0 | 0 | 1 | 0 | 0 | 0 | - | E | - | - | - |
| MLTK | 51776 | ENSG00000091436 | protein_coding | 17 | rs13017210 | 1 | 1 | 1 | 1 | 1 | 1 | - | E (UQ) | M1_hub | - | - |
| MLK7-AS1 | 339751 | ENSG00000238133 | lncRNA | 17 | rs13017210 | 1 | 1 | 1 | 0 | 1 | 0 | - | E+ | M7 | - | - |
| HOXD11 | 3237 | ENSG00000128713 | protein_coding | 18 | rs847148 | 1 | 1 | 0 | 1 | 1 | 1 | - | E | M4 | reproductive system; skeleton; limbs/digits/tail; | - |
| HOXD12 | 3238 | ENSG00000170178 | protein_coding | 18 | rs847148 | 0 | 1 | 0 | 1 | 1 | 0 | - | E | - | digestive/alimentary; limbs/digits/tail; skeleton; muscle; | - |
| HOXD10 | 3236 | ENSG00000128710 | protein_coding | 18 | rs847148 | 0 | 1 | 0 | 1 | 0 | 0 | - | E (UQ) | M4 | reproductive system; behavior/neurological; skeleton; nervous system; limbs/digits/tail; | Charcot-Marie-Tooth disease, foot deformity of, 192950 (3), Autosomal dominant; Vertical talus, congenital, 192950 (3), Autosomal dominant |
| HOXD13 | 3239 | ENSG00000128714 | protein_coding | 18 | rs847148 | 0 | 1 | 0 | 0 | 0 | 0 | - | E (UQ) | - | reproductive system; embryo; skeleton; renal/urinary system; limbs/digits/tail; digestive/alimentary; integument; growth/size/body region; endocrine/exocrine gland; muscle; | Brachydactyly, type D, 113200 (3), Autosomal dominant; Brachydactyly, type E, 113300 (3), Autosomal dominant; ?Brachydactyly-syndactyly syndrome, 610713 (3); Syndactyly, type V, 186300 (3), Autosomal dominant; Synpolydactyly 1, 186000 (3), Autosomal dominant |
| AC009336.24 | - | ENSG00000226363 | lncRNA | 18 | rs847148 | 0 | 0 | 1 | 0 | 0 | 0 | - | E | - | - | - |
| HOXD-AS1 | 401022 | ENSG00000224189 | lncRNA | 18 | rs847148 | 0 | 0 | 1 | 0 | 0 | 0 | - | E | - | - | - |
| HOXD1 | 3231 | ENSG00000128645 | protein_coding | 18 | rs847148 | 0 | 0 | 1 | 0 | 0 | 0 | - | E | - | craniofacial; integument; nervous system; skeleton; behavior/neurological; | - |
| HOXD8 | 3234 | ENSG00000175879 | protein_coding | 18 | rs847148 | 0 | 0 | 1 | 0 | 0 | 0 | - | E | M1 | skeleton; | - |
| KIAA1715 | 80856 | ENSG00000144320 | protein_coding | 18 | rs847148 | 0 | 0 | 1 | 0 | 0 | 0 | - | E (UQ) | M4 | - | - |
| COL5A2 | 1290 | ENSG00000204262 | protein_coding | 19 | rs16831319 | 1 | 1 | 1 | 0 | 1 | 1 | - | E (UQ) | M1 | integument; growth/size/body region; vision/eye; mortality/aging; cardiovascular system; respiratory system; behavior/neurological; skeleton; | Ehlers-Danlos syndrome, classic type, 2, 130010 (3), Autosomal dominant |
| WDR75 | 84128 | ENSG00000115368 | protein_coding | 19 | rs16831319 | 0 | 1 | 0 | 0 | 1 | 0 | - | E (UQ) | - | - | - |
| MIR3129 | 100422908 | ENSG00000264725 | miRNA | 19 | rs16831319 | 0 | 1 | 0 | 0 | 0 | 0 | - | E | - | - | - |
| TMEM194B | 100131211 | ENSG00000189362 | protein_coding | 19:20 | rs16831319:rs34417560 | 0 | 1 | 1 | 1 | 1 | 0 | - | E | - | - | - |
| AC093388.3 | - | ENSG00000233654 | lncRNA | 19:20 | rs16831319:rs34417560 | 0 | 1 | 1 | 0 | 0 | 0 | - | E | - | - | - |
| AC006460.2 | - | ENSG00000228509 | lncRNA | 19:20 | rs16831319:rs34417560 | 0 | 0 | 1 | 0 | 0 | 0 | - | E | - | - | - |
| NAB1 | 4664 | ENSG00000138386 | protein_coding | 19:20 | rs16831319:rs34417560 | 0 | 0 | 1 | 0 | 0 | 0 | - | E (UQ) | - | - | - |
| MFSD6 | 54842 | ENSG00000151690 | protein_coding | 20 | rs34417560 | 1 | 1 | 1 | 1 | 1 | 0 | - | E (UQ) | M4_hub | - | - |
| RP11-647K16.1 | - | ENSG00000272979 | lncRNA | 20 | rs34417560 | 0 | 1 | 0 | 0 | 0 | 0 | - | E | M1 | - | - |
| C2orf88 | 84281 | ENSG00000187699 | protein_coding | 20 | rs34417560 | 1 | 0 | 1 | 0 | 0 | 0 | - | E+ (UQ) | - | - | - |
| HIBCH | 26275 | ENSG00000198130 | protein_coding | 20 | rs34417560 | 0 | 0 | 1 | 0 | 0 | 0 | - | E (UQ) | M4 | mortality/aging; | 3-hydroxyisobutyryl-CoA hydrolase deficiency, 250620 (3), Autosomal recessive |
| INPP1 | 3628 | ENSG00000151689 | protein_coding | 20 | rs34417560 | 0 | 0 | 1 | 0 | 0 | 0 | - | E (UQ) | - | hematopoietic system; vision/eye; immune system; skeleton; growth/size/body region; | - |
| MYO1B | 4430 | ENSG00000128641 | protein_coding | 20 | rs34417560 | 0 | 0 | 1 | 0 | 0 | 0 | - | E (UQ) | - | - | - |
| STAT4 | 6775 | ENSG00000138378 | protein_coding | 20 | rs34417560 | 0 | 0 | 1 | 0 | 0 | 0 | - | E | - | endocrine/exocrine gland; homeostasis/metabolism; cellular; renal/urinary system; hematopoietic system; mortality/aging; immune system; | {Systemic lupus erythematosus, susceptibility to, 11}, 612253 (3) |
| FGD5 | 152273 | ENSG00000154783 | protein_coding | 21 | rs1689549 | 0 | 1 | 1 | 0 | 1 | 0 | - | E | M1 | mortality/aging; | - |
| AC090957.2 | - | ENSG00000230172 | lncRNA | 21 | rs1689549 | 0 | 1 | 0 | 0 | 0 | 0 | - | NE | - | - | - |
| C3orf20 | 84077 | ENSG00000131379 | protein_coding | 21 | rs1689549 | 0 | 1 | 0 | 0 | 0 | 0 | - | E | - | reproductive system; | - |
| RP11-95M5.1 | 152274 | ENSG00000260902 | lncRNA | 21 | rs1689549 | 0 | 1 | 0 | 0 | 0 | 0 | - | E | M4 | - | - |
| FGD5-AS1 | 100505641 | ENSG00000225733 | lncRNA | 21 | rs1689549 | 0 | 0 | 1 | 0 | 0 | 0 | - | E (UQ) | - | - | - |
| LSM3 | 27258 | ENSG00000170860 | protein_coding | 21 | rs1689549 | 0 | 0 | 1 | 0 | 0 | 0 | - | E (UQ) | - | immune system; mortality/aging; homeostasis/metabolism; | - |
| ALAS1 | 211 | ENSG00000023330 | protein_coding | 22 | rs9847710 | 0 | 1 | 1 | 1 | 1 | 0 | - | E (UQ) | - | mortality/aging; embryo; growth/size/body region; | - |
| NT5DC2 | 64943 | ENSG00000168268 | protein_coding | 22 | rs9847710 | 1 | 1 | 1 | 1 | 1 | 1 | - | E (UQ) | M7 | behavior/neurological; hearing/vestibular/ear; | - |
| SFMBT1 | 51460 | ENSG00000163935 | protein_coding | 22 | rs9847710 | 0 | 1 | 1 | 1 | 1 | 0 | - | E | - | - | - |
| STAB1 | 23166 | ENSG00000010327 | protein_coding | 22 | rs9847710 | 0 | 1 | 1 | 1 | 1 | 0 | - | E (UQ) | - | - | - |
| GLT8D1 | 55830 | ENSG00000016864 | protein_coding | 22 | rs9847710 | 0 | 1 | 1 | 1 | 0 | 0 | - | E (UQ) | - | nervous system; | - |
| GNL3 | 26354 | ENSG00000163938 | protein_coding | 22 | rs9847710 | 0 | 1 | 1 | 1 | 0 | 0 | - | E (UQ) | M4 | embryo; respiratory system; mortality/aging; nervous system; growth/size/body region; cellular; | - |
| ITIH1 | 3697 | ENSG00000055957 | protein_coding | 22 | rs9847710 | 0 | 1 | 1 | 1 | 0 | 0 | - | E | - | - | - |
| ITIH3 | 3699 | ENSG00000162267 | protein_coding | 22 | rs9847710 | 0 | 1 | 1 | 1 | 0 | 0 | - | E | M1 | - | - |
| ITIH4 | 3700 | ENSG00000055955 | protein_coding | 22 | rs9847710 | 0 | 1 | 1 | 1 | 0 | 0 | - | E | - | endocrine/exocrine gland; reproductive system; | - |
| MUSTN1 | 389125 | ENSG00000272573 | protein_coding | 22 | rs9847710 | 0 | 1 | 1 | 1 | 0 | 0 | - | E | M1 | - | - |
| NEK4 | 6787 | ENSG00000114904 | protein_coding | 22 | rs9847710 | 0 | 1 | 1 | 1 | 0 | 0 | - | E | - | - | - |
| PBRM1 | 55193 | ENSG00000163939 | protein_coding | 22 | rs9847710 | 0 | 1 | 1 | 1 | 0 | 0 | - | E (UQ) | M1 | renal/urinary system; immune system; mortality/aging; hematopoietic system; cardiovascular system; behavior/neurological; liver/biliary system; respiratory system; embryo; cellular; | - |
| RFT1 | 91869 | ENSG00000163933 | protein_coding | 22 | rs9847710 | 0 | 1 | 1 | 1 | 0 | 0 | - | E (UQ) | - | mortality/aging; hematopoietic system; immune system; | Congenital disorder of glycosylation, type In, 612015 (3), Autosomal recessive |
| RP11-894J14.5 | - | ENSG00000272305 | protein_coding | 22 | rs9847710 | 0 | 1 | 1 | 1 | 0 | 0 | - | E | M4 | - | - |
| RP5-966M1.6 | - | ENSG00000243696 | protein_coding | 22 | rs9847710 | 0 | 1 | 1 | 1 | 0 | 0 | - | E | M1 | - | - |
| SMIM4 | 440957 | ENSG00000168273 | protein_coding | 22 | rs9847710 | 0 | 1 | 1 | 1 | 0 | 0 | - | E (UQ) | - | - | - |
| SPCS1 | 28972 | ENSG00000114902 | protein_coding | 22 | rs9847710 | 0 | 1 | 1 | 1 | 0 | 0 | - | E (UQ) | M4 | - | - |
| TLR9 | 54106 | ENSG00000173366 | protein_coding | 22 | rs9847710 | 0 | 1 | 1 | 1 | 0 | 0 | - | E | - | cellular; mortality/aging; hematopoietic system; immune system; digestive/alimentary; | - |
| TMEM110 | 375346 | ENSG00000213533 | protein_coding | 22 | rs9847710 | 0 | 1 | 1 | 1 | 0 | 0 | - | E | M1 | - | - |
| TMEM110-MUSTN1 | 100526772 | ENSG00000248592 | protein_coding | 22 | rs9847710 | 0 | 1 | 1 | 1 | 0 | 0 | - | E | M1 | - | - |
| DNAH1 | 25981 | ENSG00000114841 | protein_coding | 22 | rs9847710 | 0 | 1 | 1 | 0 | 1 | 0 | - | E | - | integument; cellular; respiratory system; reproductive system; | ?Ciliary dyskinesia, primary, 37, 617577 (3), Autosomal recessive; Spermatogenic failure 18, 617576 (3), Autosomal recessive |
| GLYCTK | 132158 | ENSG00000168237 | protein_coding | 22 | rs9847710 | 0 | 1 | 1 | 0 | 1 | 0 | - | E | M4 | vision/eye; | D-glyceric aciduria, 220120 (3), Autosomal recessive |
| GLYCTK-AS1 | 100874110 | ENSG00000242797 | lncRNA | 22 | rs9847710 | 0 | 1 | 1 | 0 | 1 | 0 | - | E | - | - | - |
| NISCH | 11188 | ENSG00000010322 | protein_coding | 22 | rs9847710 | 0 | 1 | 1 | 0 | 1 | 0 | - | E (UQ) | - | growth/size/body region; adipose tissue; craniofacial; homeostasis/metabolism; behavior/neurological; respiratory system; mortality/aging; hearing/vestibular/ear; immune system; skeleton; | - |
| PHF7 | 51533 | ENSG00000010318 | protein_coding | 22 | rs9847710 | 0 | 1 | 1 | 0 | 1 | 0 | - | E | - | reproductive system; | - |
| PPM1M | 132160 | ENSG00000164088 | protein_coding | 22 | rs9847710 | 0 | 1 | 1 | 0 | 1 | 0 | - | E | - | homeostasis/metabolism; hematopoietic system; skeleton; immune system; | - |
| RP5-1157M23.2 | 101929054 | ENSG00000243224 | lncRNA | 22 | rs9847710 | 0 | 1 | 1 | 0 | 1 | 0 | - | E | - | - | - |
| SEMA3G | 56920 | ENSG00000010319 | protein_coding | 22 | rs9847710 | 0 | 1 | 1 | 0 | 1 | 0 | - | E | - | immune system; | - |
| TNNC1 | 7134 | ENSG00000114854 | protein_coding | 22 | rs9847710 | 0 | 1 | 1 | 0 | 1 | 0 | - | E | - | nervous system; mortality/aging; cardiovascular system; behavior/neurological; muscle; endocrine/exocrine gland; | Cardiomyopathy, hypertrophic, 13, 613243 (3), Autosomal dominant; Cardiomyopathy, dilated, 1Z, 611879 (3) |
| TWF2 | 11344 | ENSG00000247596 | protein_coding | 22 | rs9847710 | 0 | 1 | 1 | 0 | 1 | 0 | - | E (UQ) | - | - | - |
| WDR82 | 80335 | ENSG00000164091 | protein_coding | 22 | rs9847710 | 0 | 1 | 1 | 0 | 1 | 0 | - | E (UQ) | - | - | - |
| BAP1 | 8314 | ENSG00000163930 | protein_coding | 22 | rs9847710 | 0 | 1 | 0 | 0 | 1 | 0 | - | E (UQ) | - | cardiovascular system; hematopoietic system; reproductive system; mortality/aging; neoplasm; embryo; respiratory system; immune system; digestive/alimentary; growth/size/body region; integument; endocrine/exocrine gland; homeostasis/metabolism; cellular; muscle; | Tumor predisposition syndrome, 614327 (3), Autosomal dominant |
| ITIH4-AS1 | 100873993 | ENSG00000239799 | lncRNA | 22 | rs9847710 | 0 | 1 | 1 | 0 | 0 | 0 | - | NE | - | - | - |
| MIR135A1 | 406925 | ENSG00000207926 | miRNA | 22 | rs9847710 | 1 | 1 | 1 | 0 | 0 | 0 | - | E- | - | - | - |
| SNORD19B | 100113381 | ENSG00000238862 | snoRNA | 22 | rs9847710 | 0 | 1 | 1 | 0 | 0 | 0 | - | E | - | - | - |
| TLR9 | 54106 | ENSG00000239732 | protein_coding | 22 | rs9847710 | 0 | 1 | 1 | 0 | 0 | 0 | - | E | - | cellular; mortality/aging; hematopoietic system; immune system; digestive/alimentary; | - |
| MIRLET7G | 406890 | ENSG00000199150 | miRNA | 22 | rs9847710 | 0 | 1 | 0 | 0 | 0 | 0 | - | E (UQ) | - | - | - |
| RNU6-856P | 106481451 | ENSG00000252768 | snRNA | 22 | rs9847710 | 0 | 1 | 0 | 0 | 0 | 0 | - | NE | - | - | - |
| RNU6ATAC16P | 106479550 | ENSG00000221518 | snRNA | 22 | rs9847710 | 0 | 1 | 0 | 0 | 0 | 0 | - | NE | - | - | - |
| SNORD19 | 692089 | ENSG00000212493 | snoRNA | 22 | rs9847710 | 0 | 1 | 0 | 0 | 0 | 0 | - | NE | - | - | - |
| SNORD19 | 109616991 | ENSG00000222345 | snoRNA | 22 | rs9847710 | 0 | 1 | 0 | 0 | 0 | 0 | - | E | - | - | - |
| SNORD19B | 100113381 | ENSG00000252787 | snoRNA | 22 | rs9847710 | 0 | 1 | 0 | 0 | 0 | 0 | - | E | - | - | - |
| SNORD69 | 692109 | ENSG00000212452 | snoRNA | 22 | rs9847710 | 1 | 1 | 0 | 0 | 0 | 0 | - | E- (UQ) | - | - | - |
| ABHD14A | 25864 | ENSG00000248487 | protein_coding | 22 | rs9847710 | 0 | 0 | 1 | 0 | 0 | 0 | - | E | M1 | - | - |

| | | | | | | | | | | | | | | | | |
|---|---|---|---|---|---|---|---|---|---|---|---|---|---|---|---|---|
| DUSP7 | 1849 | ENSG00000164086 | protein_coding | 22 | rs9847710 | 1 | 0 | 1 | 0 | 0 | 0 | - | E (UQ) | M7_hub | homeostasis/metabolism; integument; pigmentation; reproductive system; | - |
| GPR62 | 118442 | ENSG00000180929 | protein_coding | 22 | rs9847710 | 0 | 0 | 1 | 0 | 0 | 0 | - | E | M7 | - | - |
| PARP3 | 10039 | ENSG00000041880 | protein_coding | 22 | rs9847710 | 0 | 0 | 1 | 0 | 0 | 0 | - | E | M4 | - | - |
| PCBP4 | 57060 | ENSG00000090097 | protein_coding | 22 | rs9847710 | 0 | 0 | 1 | 0 | 0 | 0 | - | E | - | renal/urinary system; respiratory system; neoplasm; cellular; growth/size/body region; | - |
| POC1A | 25886 | ENSG00000164087 | protein_coding | 22 | rs9847710 | 0 | 0 | 1 | 0 | 0 | 0 | - | E | - | behavior/neurological; reproductive system; mortality/aging; skeleton; homeostasis/metabolism; cellular; endocrine/exocrine gland; growth/size/body region; | Short stature, onychodysplasia, facial dysmorphism, and hypotrichosis, 614813 (3), Autosomal recessive |
| PRKCD | 5580 | ENSG00000163932 | protein_coding | 22 | rs9847710 | 0 | 0 | 1 | 0 | 0 | 0 | - | E (UQ) | M4 | liver/biliary system; cardiovascular system; hematopoietic system; mortality/aging; digestive/alimentary; nervous system; skeleton; renal/urinary system; homeostasis/metabolism; immune system; cellular; endocrine/exocrine gland; growth/size/body region; muscle; | Autoimmune lymphoproliferative syndrome, type III, 615559 (3), Autosomal recessive |
| RAD54L2 | 23132 | ENSG00000164080 | protein_coding | 22 | rs9847710 | 0 | 0 | 1 | 0 | 0 | 0 | - | E (UQ) | - | mortality/aging; embryo; nervous system; growth/size/body region; cellular; | - |
| RPL29 | 6159 | ENSG00000162244 | protein_coding | 22 | rs9847710 | 0 | 0 | 1 | 0 | 0 | 0 | - | E (UQ) | - | skeleton; embryo; mortality/aging; reproductive system; cellular; growth/size/body region; | - |
| TKT | 7086 | ENSG00000163931 | protein_coding | 22 | rs9847710 | 0 | 0 | 1 | 0 | 0 | 0 | - | E (UQ) | - | liver/biliary system; behavior/neurological; reproductive system; mortality/aging; cardiovascular system; nervous system; vision/eye; renal/urinary system; growth/size/body region; endocrine/exocrine gland; adipose tissue; | Short stature, developmental delay, and congenital heart defects, 617044 (3), Autosomal recessive |
| FOXP1 | 27086 | ENSG00000114861 | protein_coding | 23 | rs2597301 | 1 | 1 | 0 | 1 | 1 | 1 | - | E (UQ) | M1 | immune system; behavior/neurological; hematopoietic system; cardiovascular system; mortality/aging; muscle; homeostasis/metabolism; cellular; adipose tissue; growth/size/body region; | Mental retardation with language impairment and with or without autistic features, 613670 (3), Autosomal dominant |
| RNU6-281P | 106481267 | ENSG00000206939 | snRNA | 23 | rs2597301 | 0 | 1 | 0 | 0 | 0 | 0 | - | NE | - | - | - |
| GPR27 | 2850 | ENSG00000170837 | protein_coding | 23 | rs2597301 | 0 | 0 | 1 | 0 | 0 | 0 | - | E | - | - | - |
| PHLDB2 | 90102 | ENSG00000144824 | protein_coding | 24 | rs6792493 | 1 | 1 | 1 | 1 | 1 | 1 | - | E (UQ) | M1_hub | behavior/neurological; nervous system; vision/eye; | - |
| PLCXD2 | 257068 | ENSG00000240891 | protein_coding | 24 | rs6792493 | 0 | 1 | 1 | 1 | 1 | 0 | - | E | M7 | - | - |
| TAGLN3 | 29114 | ENSG00000144834 | protein_coding | 24 | rs6792493 | 0 | 0 | 0 | 1 | 0 | 0 | - | E | - | - | - |
| CD96 | 10225 | ENSG00000153283 | protein_coding | 24 | rs6792493 | 0 | 0 | 1 | 0 | 0 | 0 | - | E | - | homeostasis/metabolism; hematopoietic system; neoplasm; immune system; | C syndrome, 211750 (3), Autosomal dominant |
| ZBTB20 | 26137 | ENSG00000181722 | protein_coding | 25 | rs9853475 | 1 | 1 | 1 | 1 | 1 | 1 | eQTL (rs9853475, PP=0.599) | E | M1 | cardiovascular system; mortality/aging; liver/biliary system; behavior/neurological; vision/eye; nervous system; growth/size/body region; integument; homeostasis/metabolism; | Primrose syndrome, 259050 (3), Autosomal dominant |
| RP11-6F2.5 | 105374177 | ENSG00000241544 | lncRNA | 26 | rs900400 | 0 | 1 | 1 | 0 | 1 | 0 | - | NE | - | - | - |
| LINC00880 | 339894 | ENSG00000243629 | lncRNA | 26 | rs900400 | 0 | 1 | 0 | 0 | 1 | 0 | - | E | - | - | - |
| LINC00881 | 100498859 | ENSG00000241135 | lncRNA | 26 | rs900400 | 0 | 1 | 1 | 0 | 0 | 0 | - | E | M4 | - | - |
| CCNL1 | 57018 | ENSG00000163660 | protein_coding | 26 | rs900400 | 0 | 0 | 1 | 0 | 0 | 0 | - | E (UQ) | - | - | - |
| LINC00886 | 730091 | ENSG00000240875 | lncRNA | 26 | rs900400 | 0 | 0 | 1 | 0 | 0 | 0 | - | E | - | - | - |
| TIPARP | 25976 | ENSG00000163659 | protein_coding | 26 | rs900400 | 0 | 0 | 1 | 0 | 0 | 0 | - | E (UQ) | - | nervous system; digestive/alimentary; immune system; renal/urinary system; skeleton; mortality/aging; reproductive system; cardiovascular system; hematopoietic system; endocrine/exocrine gland; muscle; craniofacial; homeostasis/metabolism; integument; growth/size/body region; | - |
| IFT80 | 57560 | ENSG00000068885 | protein_coding | 27 | rs3851366 | 0 | 1 | 1 | 1 | 1 | 0 | - | E | - | mortality/aging; skeleton; limbs/digits/tail; growth/size/body region; cellular; homeostasis/metabolism; craniofacial; | Short-rib thoracic dysplasia 2 with or without polydactyly, 611263 (3), Autosomal recessive |
| KPNA4 | 3840 | ENSG00000186432 | protein_coding | 27 | rs3851366 | 0 | 1 | 1 | 1 | 1 | 0 | - | E (UQ) | - | - | - |
| SMC4 | 10051 | ENSG00000113810 | protein_coding | 27 | rs3851366 | 0 | 1 | 1 | 1 | 1 | 0 | - | E (UQ) | - | - | - |
| TRIM59 | 286827 | ENSG00000213186 | protein_coding | 27 | rs3851366 | 0 | 1 | 1 | 1 | 1 | 0 | - | E | - | nervous system; embryo; growth/size/body region; reproductive system; mortality/aging; | - |
| RP11-432B6.3 | 100174949 | ENSG00000248710 | protein_coding | 27 | rs3851366 | 0 | 1 | 1 | 1 | 0 | 0 | - | E | - | - | - |
| C3orf80 | 401097 | ENSG00000180044 | protein_coding | 27 | rs3851366 | 0 | 1 | 0 | 0 | 1 | 0 | - | E | - | - | - |
| IL12A-AS1 | 101928376 | ENSG00000244040 | lncRNA | 27 | rs3851366 | 0 | 1 | 0 | 0 | 0 | 0 | - | E | M7 | - | - |
| MIR15B | 406949 | ENSG00000207779 | miRNA | 27 | rs3851366 | 0 | 1 | 0 | 0 | 0 | 0 | - | E (UQ) | - | - | - |
| MIR16-2 | 406951 | ENSG00000198987 | miRNA | 27 | rs3851366 | 0 | 1 | 0 | 0 | 0 | 0 | - | E | - | - | - |
| RNU7-136P | 106479076 | ENSG00000238427 | snRNA | 27 | rs3851366 | 0 | 1 | 0 | 0 | 0 | 0 | - | NE | - | - | - |
| SCARNA7 | 677767 | ENSG00000238741 | scaRNA | 27 | rs3851366 | 0 | 1 | 0 | 0 | 0 | 0 | - | E | - | - | - |
| KRT8P12 | 90133 | ENSG00000229320 | nscribed_processed_pseudoge | 27 | rs3851366 | 1 | 0 | 0 | 0 | 1 | 1 | - | E | - | - | - |
| ARL14 | 80117 | ENSG00000179674 | protein_coding | 27 | rs3851366 | 1 | 0 | 1 | 0 | 0 | 0 | - | E+ (UQ) | - | - | - |
| LINC01100 | 730109 | ENSG00000242107 | lncRNA | 27 | rs3851366 | 0 | 0 | 1 | 0 | 0 | 0 | - | NE | - | - | - |
| ACOX3 | 8310 | ENSG00000087008 | protein_coding | 28 | rs2631752 | 0 | 1 | 1 | 1 | 1 | 0 | - | E | M7 | vision/eye; | - |
| TRMT44 | 152992 | ENSG00000155275 | protein_coding | 28 | rs2631752 | 0 | 1 | 1 | 0 | 1 | 0 | - | E | - | - | - |
| RP11-689P11.2 | - | ENSG00000205959 | lncRNA | 28 | rs2631752 | 0 | 1 | 1 | 0 | 0 | 0 | - | E | M7 | - | - |
| RP11-774O3.3 | - | ENSG00000251615 | lncRNA | 28 | rs2631752 | 0 | 1 | 1 | 0 | 0 | 0 | - | E | - | - | - |
| RNA5SP152 | 100873418 | ENSG00000202054 | rRNA_pseudogene | 28 | rs2631752 | 0 | 1 | 0 | 0 | 0 | 0 | - | NE | - | - | - |
| RP11-689P11.3 | - | ENSG00000251186 | lncRNA | 28 | rs2631752 | 0 | 1 | 0 | 0 | 0 | 0 | - | NE | - | - | - |
| AFAP1 | 60312 | ENSG00000196526 | protein_coding | 28 | rs2631752 | 0 | 0 | 1 | 0 | 0 | 0 | - | E (UQ) | - | integument; endocrine/exocrine gland; cellular; behavior/neurological; | - |
| CPZ | 8532 | ENSG00000109625 | protein_coding | 28 | rs2631752 | 0 | 0 | 1 | 0 | 0 | 0 | - | E | M1 | - | - |
| LIAS | 11019 | ENSG00000121897 | protein_coding | 29 | rs2687965 | 1 | 1 | 1 | 0 | 1 | 0 | eQTL (rs2687965, PP=0.502) | E | M4 | embryo; cardiovascular system; mortality/aging; nervous system; growth/size/body region; homeostasis/metabolism; | Hyperglycinemia, lactic acidosis, and seizures, 614462 (3), Autosomal recessive |
| UGDH | 7358 | ENSG00000109814 | protein_coding | 29 | rs2687965 | 1 | 1 | 1 | 0 | 1 | 0 | eQTL (rs2687965, PP=0.502) | E (UQ) | - | endocrine/exocrine gland; growth/size/body region; craniofacial; digestive/alimentary; mortality/aging; embryo; vision/eye; | Epileptic encephalopathy, early infantile, 84, 618792 (3), Autosomal recessive |
| KLB | 152831 | ENSG00000134962 | protein_coding | 29 | rs2687965 | 1 | 1 | 1 | 0 | 0 | 0 | eQTL (rs2687965, PP=0.502) | E | - | growth/size/body region; adipose tissue; endocrine/exocrine gland; homeostasis/metabolism; liver/biliary system; | - |
| SMIM14 | 201895 | ENSG00000163683 | protein_coding | 29 | rs2687965 | 1 | 0 | 1 | 0 | 0 | 0 | eQTL (rs2687965, PP=0.502) | E (UQ) | - | - | - |
| UGDH-AS1 | 100885776 | ENSG00000249348 | lncRNA | 29 | rs2687965 | 1 | 0 | 1 | 0 | 0 | 0 | eQTL (rs2687965, PP=0.502) | E | - | - | - |
| RPL9 | 6133 | ENSG00000163682 | protein_coding | 29 | rs2687965 | 1 | 0 | 1 | 0 | 0 | 0 | eQTL (rs2687965, PP=0.502) | E (UQ) | - | - | - |
| BTC | 685 | ENSG00000174808 | protein_coding | 30 | rs11942410 | 0 | 1 | 1 | 1 | 1 | 0 | - | E | M1 | endocrine/exocrine gland; growth/size/body region; craniofacial; muscle; homeostasis/metabolism; immune system; liver/biliary system; respiratory system; reproductive system; mortality/aging; cardiovascular system; hematopoietic system; vision/eye; skeleton; | - |
| MTHFD2L | 441024 | ENSG00000163738 | protein_coding | 30 | rs11942410 | 0 | 0 | 1 | 0 | 0 | 0 | - | E | - | integument; cardiovascular system; | - |
| RP11-44F21.3 | 100507388 | ENSG00000249717 | lncRNA | 30 | rs11942410 | 0 | 0 | 1 | 0 | 0 | 0 | - | E | - | - | - |
| RP11-542G1.3 | 441025 | ENSG00000251383 | lncRNA | 30 | rs11942410 | 0 | 0 | 1 | 0 | 0 | 0 | - | NE | - | - | - |
| RP11-567N4.3 | - | ENSG00000250735 | lncRNA | 30 | rs11942410 | 0 | 0 | 1 | 0 | 0 | 0 | - | E | - | - | - |
| BMPR1B | 658 | ENSG00000138696 | protein_coding | 31 | rs28663472 | 0 | 1 | 1 | 1 | 1 | 0 | - | E | M1 | endocrine/exocrine gland; cellular; vision/eye; reproductive system; skeleton; nervous system; limbs/digits/tail; | Brachydactyly, type A2, 112600 (3), Autosomal dominant; Brachydactyly, type A1, D, 616849 (3), Autosomal dominant; Acromesomelic dysplasia, Demirhan type, 609441 (3), Autosomal recessive |
| UNC5C | 8633 | ENSG00000182168 | protein_coding | 31 | rs28663472 | 1 | 1 | 1 | 0 | 1 | 1 | - | E | - | behavior/neurological; reproductive system; mortality/aging; nervous system; growth/size/body region; adipose tissue; cellular; | - |
| TET2 | 54790 | ENSG00000168769 | protein_coding | 32 | rs6839705 | 0 | 1 | 1 | 1 | 1 | 0 | - | E | M7 | renal/urinary system; immune system; liver/biliary system; neoplasm; mortality/aging; growth/size/body region; hematopoietic system; cardiovascular system; cellular; endocrine/exocrine gland; | Myelodysplastic syndrome, somatic, 614286 (3) |
| TET2-AS1 | 104384744 | ENSG00000251586 | lncRNA | 32 | rs6839705 | 0 | 1 | 1 | 0 | 0 | 0 | - | NE | - | - | - |
| AC004069.2 | - | ENSG00000251259 | lncRNA | 32 | rs6839705 | 0 | 1 | 0 | 0 | 0 | 0 | - | E | - | - | - |
| RP11-556I14.1 | - | ENSG00000248373 | lncRNA | 32 | rs6839705 | 0 | 1 | 0 | 0 | 0 | 0 | - | NE | - | - | - |
| PPA2 | 27068 | ENSG00000138777 | protein_coding | 32 | rs6839705 | 0 | 0 | 1 | 0 | 1 | 0 | - | E (UQ) | M4 | - | ?Sudden cardiac failure, alcohol-induced, 617223 (3), Autosomal recessive; Sudden cardiac failure, infantile, 617222 (3), Autosomal recessive |
| AC096732.2 | - | ENSG00000226655 | lncRNA | 33 | rs2060285 | 0 | 1 | 0 | 0 | 0 | 0 | - | NE | - | - | - |
| LINC01091 | 285419 | ENSG00000249464 | lncRNA | 33 | rs2060285 | 0 | 1 | 0 | 0 | 0 | 0 | - | E | - | - | - |
| FAT4 | 79633 | ENSG00000196159 | protein_coding | 33:34 | rs2060285:rs17824374 | 1 | 0 | 1 | 0 | 1 | 1 | - | E | M1 | embryo; respiratory system; mortality/aging; cardiovascular system; nervous system; hearing/vestibular/ear; limbs/digits/tail; digestive/alimentary; skeleton; renal/urinary system; growth/size/body region; cellular; | Van Maldergem syndrome 2, 615546 (3), Autosomal recessive; Hennekam lymphangiectasia-lymphedema syndrome 2, 616006 (3), Autosomal recessive |
| RP11-318I4.1 | - | ENSG00000251649 | lncRNA | 34 | rs17824374 | 0 | 1 | 0 | 0 | 0 | 0 | - | NE | - | - | - |
| RP11-361D14.2 | - | ENSG00000261129 | lncRNA | 35 | rs1542726 | 0 | 1 | 0 | 0 | 0 | 0 | - | E | - | - | - |
| ABCE1 | 6059 | ENSG00000164163 | protein_coding | 35 | rs1542726 | 0 | 0 | 0 | 1 | 0 | 0 | - | E (UQ) | - | homeostasis/metabolism; endocrine/exocrine gland; mortality/aging; vision/eye; nervous system; immune system; behavior/neurological; reproductive system; | - |
| HHIP-AS1 | 646576 | ENSG00000248890 | lncRNA | 35 | rs1542726 | 0 | 0 | 1 | 0 | 1 | 0 | - | E | - | - | - |
| FREM3 | 166752 | ENSG00000183090 | protein_coding | 35 | rs1542726 | 0 | 0 | 1 | 0 | 0 | 0 | - | E | M1 | - | - |
| HHIP | 64399 | ENSG00000164161 | protein_coding | 35 | rs1542726 | 0 | 0 | 1 | 0 | 0 | 0 | - | E | - | immune system; digestive/alimentary; nervous system; hematopoietic system; mortality/aging; embryo; respiratory system; cellular; growth/size/body region; endocrine/exocrine gland; | - |
| OTUD4 | 54726 | ENSG00000164164 | protein_coding | 35 | rs1542726 | 0 | 0 | 1 | 0 | 0 | 0 | - | E | - | mortality/aging; | - |
| RP11-673E1.1 | 101927636 | ENSG00000251600 | lncRNA | 35 | rs1542726 | 0 | 0 | 1 | 0 | 0 | 0 | - | E | - | - | - |
| TPPP | 11076 | ENSG00000171368 | protein_coding | 36 | rs72707023 | 0 | 1 | 1 | 1 | 1 | 0 | - | E | M7 | growth/size/body region; homeostasis/metabolism; skeleton; behavior/neurological; hematopoietic system; nervous system; | - |
| CEP72 | 55722 | ENSG00000112877 | protein_coding | 36 | rs72707023 | 0 | 1 | 1 | 1 | 0 | 0 | - | E | - | - | - |
| SLC9A3 | 6550 | ENSG00000066230 | protein_coding | 36 | rs72707023 | 0 | 1 | 1 | 1 | 0 | 0 | - | E | - | endocrine/exocrine gland; homeostasis/metabolism; cardiovascular system; reproductive system; digestive/alimentary; renal/urinary system; | Diarrhea 8, secretory sodium, congenital, 616868 (3), Autosomal recessive |
| CTD-2589H19.6 | - | ENSG00000271781 | lncRNA | 36 | rs72707023 | 0 | 1 | 1 | 0 | 0 | 0 | - | E | M7 | - | - |
| RP11-310P5.1 | - | ENSG00000249650 | lncRNA | 36 | rs72707023 | 0 | 1 | 1 | 0 | 0 | 0 | - | NE | - | - | - |
| MIR4456 | 100616381 | ENSG00000264233 | miRNA | 36 | rs72707023 | 0 | 1 | 0 | 0 | 0 | 0 | - | NE | - | - | - |
| RP11-310P5.2 | - | ENSG00000250385 | lncRNA | 36 | rs72707023 | 0 | 1 | 0 | 0 | 0 | 0 | - | NE | - | - | - |
| AHRR | 57491 | ENSG00000063438 | protein_coding | 36 | rs72707023 | 0 | 0 | 1 | 0 | 0 | 0 | - | E | - | homeostasis/metabolism; neoplasm; behavior/neurological; hearing/vestibular/ear; | - |
| BRD9 | 65980 | ENSG00000028310 | protein_coding | 36 | rs72707023 | 0 | 0 | 1 | 0 | 0 | 0 | - | E (UQ) | - | mortality/aging; | - |
| C5orf55 | 116349 | ENSG00000221990 | lncRNA | 36 | rs72707023 | 0 | 0 | 1 | 0 | 0 | 0 | - | E | - | - | - |
| CCDC127 | 133957 | ENSG00000164366 | protein_coding | 36 | rs72707023 | 0 | 0 | 1 | 0 | 0 | 0 | - | E (UQ) | M1 | homeostasis/metabolism; | - |
| CTD-2228K2.5 | 25845 | ENSG00000188242 | lncRNA | 36 | rs72707023 | 0 | 0 | 1 | 0 | 0 | 0 | - | E | - | - | - |
| CTD-2228K2.7 | - | ENSG00000225138 | lncRNA | 36 | rs72707023 | 0 | 0 | 1 | 0 | 0 | 0 | - | E (UQ) | M7 | - | - |
| CTD-2245E15.3 | - | ENSG00000251532 | lncRNA | 36 | rs72707023 | 0 | 0 | 1 | 0 | 0 | 0 | - | NE | - | - | - |
| EXOC3 | 11336 | ENSG00000180104 | protein_coding | 36 | rs72707023 | 0 | 0 | 1 | 0 | 0 | 0 | - | E (UQ) | - | growth/size/body region; | - |
| PDCD6 | 10016 | ENSG00000249915 | protein_coding | 36 | rs72707023 | 0 | 0 | 1 | 0 | 0 | 0 | - | E (UQ) | - | - | - |

| | | | | | | | | | | | | | | | | |
|---|---|---|---|---|---|---|---|---|---|---|---|---|---|---|---|---|
| ZDHHC11 | 79844 | ENSG00000188818 | protein_coding | 36 | rs72707023 | 0 | 0 | 1 | 0 | 0 | 0 | - | E | - | hearing/vestibular/ear; | - |
| ZDHHC11B | 653082 | ENSG00000206077 | protein_coding | 36 | rs72707023 | 0 | 0 | 1 | 0 | 0 | 0 | - | E | - | hearing/vestibular/ear; | - |
| FBXL7 | 23194 | ENSG00000183580 | protein_coding | 37 | rs61026653 | 1 | 1 | 0 | 1 | 1 | 1 | - | E | M1_hub | - | - |
| CTD-2350J17.1 | 101929472 | ENSG00000250250 | lncRNA | 37 | rs61026653 | 0 | 0 | 0 | 0 | 1 | 0 | - | NE | - | - | - |
| CTC-461F20.1 | - | ENSG00000250415 | lncRNA | 37 | rs61026653 | 0 | 0 | 1 | 0 | 0 | 0 | - | NE | - | - | - |
| ISL1 | 3670 | ENSG00000016082 | protein_coding | 38:39 | rs62368263:rs4485884 | 1 | 1 | 0 | 1 | 1 | 0 | - | E | M1_hub | endocrine/exocrine gland; growth/size/body region; craniofacial; cellular; homeostasis/metabolism; immune system; behavior/neurological; embryo; mortality/aging; cardiovascular system; vision/eye; limbs/digits/tail; hearing/vestibular/ear; nervous system; digestive/alimentary; skeleton; | - |
| CTD-2314G24.2 | - | ENSG00000259663 | lncRNA | 38:39 | rs62368263:rs4485884 | 0 | 1 | 0 | 0 | 0 | 0 | - | E | M1 | - | - |
| CTD-2335O3.3 | - | ENSG00000251125 | lncRNA | 38:39 | rs62368263:rs4485884 | 0 | 1 | 0 | 0 | 0 | 0 | - | NE | - | - | - |
| FAM172A | 83989 | ENSG00000113391 | protein_coding | 40 | rs6867042 | 0 | 1 | 1 | 1 | 1 | 0 | - | E | M1 | endocrine/exocrine gland; liver/biliary system; hematopoietic system; immune system; | - |
| KIAA0825 | 285600 | ENSG00000185261 | protein_coding | 40 | rs6867042 | 0 | 1 | 0 | 1 | 1 | 0 | - | E | M7 | reproductive system; skeleton; renal/urinary system; nervous system; adipose tissue; growth/size/body region; homeostasis/metabolism; | Polydactyly, postaxial, type A10, 618498 (3), Autosomal recessive |
| POU5F2 | 134187 | ENSG00000248483 | protein_coding | 40 | rs6867042 | 0 | 1 | 0 | 1 | 1 | 0 | - | E | - | reproductive system; | - |
| RP11-549J18.1 | - | ENSG00000251023 | lncRNA | 40 | rs6867042 | 0 | 1 | 1 | 0 | 1 | 0 | - | E | - | - | - |
| RP11-185E12.2 | - | ENSG00000272406 | lncRNA | 40 | rs6867042 | 0 | 1 | 0 | 0 | 0 | 0 | - | NE | - | - | - |
| NR2F1 | 7025 | ENSG00000175745 | protein_coding | 40 | rs6867042 | 1 | 0 | 0 | 1 | 1 | 1 | - | E | - | growth/size/body region; cellular; mortality/aging; embryo; behavior/neurological; hearing/vestibular/ear; nervous system; | Bosch-Boonstra-Schaaf optic atrophy syndrome, 615722 (3), Autosomal dominant |
| NR2F1-AS1 | 441094 | ENSG00000237187 | lncRNA | 40 | rs6867042 | 0 | 0 | 0 | 0 | 1 | 0 | - | E | M1 | - | - |
| RGMB | 285704 | ENSG00000174136 | protein_coding | 41 | rs1563319 | 0 | 0 | 1 | 0 | 1 | 0 | - | E | - | mortality/aging; | - |
| RGMB-AS1 | 503569 | ENSG00000246763 | lncRNA | 41 | rs1563319 | 0 | 0 | 1 | 0 | 0 | 0 | - | E | - | - | - |
| CTC-340A15.2 | 102546299 | ENSG00000241956 | lncRNA | 42 | rs12153515 | 0 | 1 | 1 | 0 | 1 | 0 | - | E | - | - | - |
| CTB-181F24.1 | - | ENSG00000254066 | lncRNA | 42 | rs12153515 | 0 | 1 | 0 | 0 | 0 | 0 | - | NE | - | - | - |
| CTC-535M15.2 | - | ENSG00000253693 | lncRNA | 42 | rs12153515 | 0 | 1 | 0 | 0 | 0 | 0 | - | NE | - | - | - |
| EXOC2 | 55770 | ENSG00000112685 | protein_coding | 43 | rs4959352 | 0 | 1 | 0 | 1 | 1 | 0 | - | E (UQ) | - | cellular; embryo; mortality/aging; | - |
| HUS1B | 135458 | ENSG00000188996 | protein_coding | 43 | rs4959352 | 0 | 1 | 0 | 0 | 0 | 0 | - | E | - | - | - |
| RP11-532F6.3 | - | ENSG00000272463 | lncRNA | 43 | rs4959352 | 0 | 0 | 1 | 0 | 0 | 0 | - | E | M1 | - | - |
| RP11-532F6.5 | - | ENSG00000271911 | lncRNA | 43 | rs4959352 | 0 | 0 | 1 | 0 | 0 | 0 | - | E | M1 | - | - |
| GMDS | 2762 | ENSG00000112699 | protein_coding | 43:44 | rs4959352:rs722587 | 0 | 1 | 1 | 1 | 1 | 0 | - | E (UQ) | M4 | hematopoietic system; behavior/neurological; skeleton; immune system; growth/size/body region; homeostasis/metabolism; craniofacial; | - |
| MYLK4 | 340156 | ENSG00000145949 | protein_coding | 43:44 | rs4959352:rs722587 | 0 | 0 | 1 | 0 | 0 | 0 | - | E | M7 | - | - |
| CAP2 | 10486 | ENSG00000112186 | protein_coding | 45 | rs10807610 | 0 | 1 | 1 | 1 | 1 | 0 | - | E | M1 | cardiovascular system; mortality/aging; behavior/neurological; immune system; vision/eye; muscle; growth/size/body region; homeostasis/metabolism; | - |
| TRIM31 | 11074 | ENSG00000204616 | protein_coding | 46 | rs1156533 | 0 | 1 | 1 | 0 | 0 | 0 | - | E (UQ) | M4 | immune system; digestive/alimentary; hematopoietic system; homeostasis/metabolism; growth/size/body region; | - |
| TRIM31-AS1 | 104533120 | ENSG00000231226 | lncRNA | 46 | rs1156533 | 0 | 1 | 1 | 0 | 0 | 0 | - | E | - | - | - |
| ABCF1 | 23 | ENSG00000204574 | protein_coding | 46 | rs1156533 | 0 | 0 | 1 | 0 | 0 | 0 | - | E (UQ) | - | mortality/aging; | - |
| HCG18 | 414777 | ENSG00000231074 | lncRNA | 46 | rs1156533 | 0 | 0 | 1 | 0 | 0 | 0 | - | E (UQ) | - | - | - |
| HCG22 | 285834 | ENSG00000228789 | lncRNA | 46 | rs1156533 | 0 | 0 | 1 | 0 | 0 | 0 | - | E | M7 | - | - |
| HCG9 | 10255 | ENSG00000204625 | lncRNA | 46 | rs1156533 | 0 | 0 | 1 | 0 | 0 | 0 | - | E | - | - | - |
| HLA-A | 3105 | ENSG00000206503 | protein_coding | 46 | rs1156533 | 0 | 0 | 1 | 0 | 0 | 0 | - | E (UQ) | M4 | - | {Hypersensitivity syndrome, carbamazepine-induced, susceptibility to}, 608579 (3) |
| HLA-C | 3107 | ENSG00000204525 | protein_coding | 46 | rs1156533 | 0 | 0 | 1 | 0 | 0 | 0 | - | E (UQ) | - | - | {Psoriasis susceptibility 1}, 177900 (3), Multifactorial; {HIV-1 viremia, susceptibility to}, 609423 (3) |
| HLA-E | 3133 | ENSG00000204592 | protein_coding | 46 | rs1156533 | 0 | 0 | 1 | 0 | 0 | 0 | - | E (UQ) | - | homeostasis/metabolism; hematopoietic system; immune system; | - |
| HLA-F | 3134 | ENSG00000204642 | protein_coding | 46 | rs1156533 | 0 | 0 | 1 | 0 | 0 | 0 | - | E (UQ) | M4 | - | - |
| HLA-F-AS1 | 285830 | ENSG00000214922 | lncRNA | 46 | rs1156533 | 0 | 0 | 1 | 0 | 0 | 0 | - | E | M1 | - | - |
| HLA-G | 3135 | ENSG00000204632 | protein_coding | 46 | rs1156533 | 0 | 0 | 1 | 0 | 0 | 0 | - | E (UQ) | M4 | homeostasis/metabolism; hematopoietic system; immune system; | {Asthma, susceptibility to}, 600807 (2), Autosomal dominant |
| MOG | 4340 | ENSG00000204655 | protein_coding | 46 | rs1156533 | 0 | 0 | 1 | 0 | 0 | 0 | - | E | - | immune system; hematopoietic system; cellular; | ?Narcolepsy 7, 614250 (3), Autosomal dominant |
| MRPS18B | 28973 | ENSG00000204568 | protein_coding | 46 | rs1156533 | 0 | 0 | 1 | 0 | 0 | 0 | - | E (UQ) | M4 | - | - |
| NRM | 11270 | ENSG00000137404 | protein_coding | 46 | rs1156533 | 0 | 0 | 1 | 0 | 0 | 0 | - | E | - | - | - |
| OR2H2 | 7932 | ENSG00000204657 | protein_coding | 46 | rs1156533 | 0 | 0 | 1 | 0 | 0 | 0 | - | NE | - | - | - |
| PPP1R11 | 6992 | ENSG00000204619 | protein_coding | 46 | rs1156533 | 0 | 0 | 1 | 0 | 0 | 0 | - | E (UQ) | - | - | - |
| PPP1R18 | 170954 | ENSG00000146112 | protein_coding | 46 | rs1156533 | 0 | 0 | 1 | 0 | 0 | 0 | - | E (UQ) | M1 | - | - |
| PRR3 | 80742 | ENSG00000204576 | protein_coding | 46 | rs1156533 | 0 | 0 | 1 | 0 | 0 | 0 | - | E | M1 | - | - |
| RNF39 | 80352 | ENSG00000204618 | protein_coding | 46 | rs1156533 | 0 | 0 | 1 | 0 | 0 | 0 | - | E | M7 | - | - |
| TRIM10 | 10107 | ENSG00000204613 | protein_coding | 46 | rs1156533 | 0 | 0 | 1 | 0 | 0 | 0 | - | E | - | - | - |
| TRIM26 | 7726 | ENSG00000234127 | protein_coding | 46 | rs1156533 | 0 | 0 | 1 | 0 | 0 | 0 | - | E (UQ) | M4 | - | - |
| VARS2 | 57176 | ENSG00000137411 | protein_coding | 46 | rs1156533 | 0 | 0 | 1 | 0 | 0 | 0 | - | E (UQ) | M7 | mortality/aging; growth/size/body region; | Combined oxidative phosphorylation deficiency 20, 615917 (3), Autosomal recessive |
| ZFP57 | 346171 | ENSG00000204644 | protein_coding | 46 | rs1156533 | 0 | 0 | 1 | 0 | 0 | 0 | - | E | - | mortality/aging; cellular; | Diabetes mellitus, transient neonatal, 1, 601410 (3), Autosomal dominant |
| ZNRD1 | 30834 | ENSG00000066379 | protein_coding | 46 | rs1156533 | 0 | 0 | 1 | 0 | 0 | 0 | - | E (UQ) | - | - | - |
| ZNRD1-AS1 | 80862 | ENSG00000204623 | ranscribed_unitary_pseudogen | 46 | rs1156533 | 0 | 0 | 1 | 0 | 0 | 0 | - | E | - | - | - |
| DCBLD1 | 285761 | ENSG00000164465 | protein_coding | 47 | rs2180811 | 0 | 1 | 1 | 1 | 1 | 0 | - | E | - | - | - |
| GOPC | 57120 | ENSG00000047932 | protein_coding | 47 | rs2180811 | 0 | 1 | 1 | 1 | 1 | 0 | - | E (UQ) | - | growth/size/body region; homeostasis/metabolism; cellular; vision/eye; mortality/aging; reproductive system; embryo; | - |
| ROS1 | 6098 | ENSG00000047936 | protein_coding | 47 | rs2180811 | 0 | 1 | 1 | 1 | 1 | 0 | - | E | M4 | reproductive system; cellular; | - |
| RNU6-253P | 106480697 | ENSG00000207461 | snRNA | 47 | rs2180811 | 0 | 1 | 0 | 0 | 0 | 0 | - | NE | - | - | - |
| RP1-92C8.3 | - | ENSG00000226181 | lncRNA | 47 | rs2180811 | 0 | 1 | 0 | 0 | 0 | 0 | - | NE | - | - | - |
| FAM26F | 441168 | ENSG00000188820 | protein_coding | 47 | rs2180811 | 0 | 0 | 1 | 0 | 0 | 0 | - | E | - | - | - |
| GPRC6A | 222545 | ENSG00000173612 | protein_coding | 47 | rs2180811 | 0 | 0 | 1 | 0 | 0 | 0 | - | NE | - | integument; growth/size/body region; adipose tissue; endocrine/exocrine gland; cellular; homeostasis/metabolism; reproductive system; hematopoietic system; cardiovascular system; liver/biliary system; skeleton; renal/urinary system; | - |
| RWDD1 | 51389 | ENSG00000111832 | protein_coding | 47 | rs2180811 | 1 | 0 | 1 | 0 | 0 | 0 | - | E- (UQ) | M1 | homeostasis/metabolism; growth/size/body region; skeleton; immune system; hematopoietic system; | - |
| TCF21 | 6943 | ENSG00000118526 | protein_coding | 48 | rs2327426 | 1 | 1 | 1 | 0 | 1 | 1 | - | E (UQ) | - | renal/urinary system; immune system; hematopoietic system; cardiovascular system; mortality/aging; reproductive system; respiratory system; homeostasis/metabolism; muscle; endocrine/exocrine gland; integument; | - |
| RP3-323P13.2 | 100507308 | ENSG00000227954 | lncRNA | 48 | rs2327426 | 0 | 1 | 1 | 0 | 0 | 0 | - | E | - | - | - |
| RP4-662A9.2 | - | ENSG00000223586 | lncRNA | 48 | rs2327426 | 0 | 1 | 0 | 0 | 0 | 0 | - | NE | - | - | - |
| RP1-283K11.2 | - | ENSG00000234567 | lncRNA | 48 | rs2327426 | 0 | 0 | 1 | 0 | 0 | 0 | - | NE | - | - | - |
| ESR1 | 2099 | ENSG00000091831 | protein_coding | 49:50 | rs3020338:rs9322356 | 0 | 1 | 1 | 1 | 1 | 0 | - | E | - | respiratory system; behavior/neurological; embryo; neoplasm; cardiovascular system; hematopoietic system; reproductive system; mortality/aging; nervous system; limbs/digits/tail; skeleton; renal/urinary system; homeostasis/metabolism; immune system; cellular; endocrine/exocrine gland; adipose tissue; growth/size/body region; integument; muscle; | {Myocardial infarction, susceptibility to}, 608446 (3); Estrogen resistance, 615363 (3), Autosomal recessive; Breast cancer, somatic, 114480 (3); {Migraine, susceptibility to}, 157300 (3), Autosomal dominant |
| THBS2 | 7058 | ENSG00000186340 | protein_coding | 51 | rs3253 | 1 | 1 | 1 | 1 | 1 | 1 | - | E (UQ) | - | limbs/digits/tail; skeleton; behavior/neurological; neoplasm; mortality/aging; integument; cardiovascular system; hematopoietic system; muscle; cellular; homeostasis/metabolism; | {Lumbar disc herniation, susceptibility to}, 603932 (3) |
| RP11-417E7.1 | 101929484 | ENSG00000223485 | lncRNA | 51 | rs3253 | 0 | 1 | 1 | 0 | 0 | 0 | - | E | M4 | - | - |
| RP11-417E7.2 | 101929504 | ENSG00000261039 | lncRNA | 51 | rs3253 | 0 | 1 | 1 | 0 | 0 | 0 | - | NE | - | - | - |
| XXyac-YX65C7_A.2 | 101929523 | ENSG00000226445 | lncRNA | 51 | rs3253 | 0 | 1 | 1 | 0 | 0 | 0 | - | E | - | - | - |
| LINC00242 | 401288 | ENSG00000229214 | lncRNA | 51 | rs3253 | 0 | 0 | 1 | 0 | 0 | 0 | - | E | - | - | - |
| SUGCT | 79783 | ENSG00000175600 | protein_coding | 52 | rs6462976 | 0 | 1 | 0 | 1 | 1 | 0 | - | E | M1 | - | Glutaric aciduria III, 231690 (3), Autosomal recessive |
| EGFR | 1956 | ENSG00000146648 | protein_coding | 53 | rs7795564 | 1 | 1 | 1 | 1 | 1 | 1 | eQTL (rs7795564, PP=0.553) | E (UQ) | - | endocrine/exocrine gland; integument; growth/size/body region; craniofacial; muscle; cellular; immune system; homeostasis/metabolism; liver/biliary system; respiratory system; embryo; pigmentation; reproductive system; mortality/aging; hematopoietic system; cardiovascular system; vision/eye; limbs/digits/tail; nervous system; hearing/vestibular/ear; digestive/alimentary; renal/urinary system; skeleton; | ?Inflammatory skin and bowel disease, neonatal, 2, 616069 (3), Autosomal recessive; Nonsmall cell lung cancer, response to tyrosine kinase inhibitor in, 211980 (3), Autosomal dominant, Somatic mutation; Adenocarcinoma of lung, response to tyrosine kinase inhibitor in, 211980 (3), Autosomal dominant, Somatic mutation; {Nonsmall cell lung cancer, susceptibility to}, 211980 (3), Autosomal dominant, Somatic mutation |
| ABHD11 | 83451 | ENSG00000106077 | protein_coding | 54 | rs11770437 | 1 | 1 | 1 | 0 | 0 | 0 | eQTL (rs11769827, PP=0.569) | E (UQ) | - | endocrine/exocrine gland; renal/urinary system; skeleton; vision/eye; nervous system; reproductive system; | - |
| WBSCR27 | 155368 | ENSG00000165171 | protein_coding | 54 | rs11770437 | 1 | 1 | 1 | 0 | 0 | 0 | eQTL (rs11769827, PP=0.569) | E | - | - | - |
| LIMK1 | 3984 | ENSG00000106683 | protein_coding | 54 | rs11770437 | 1 | 0 | 1 | 0 | 0 | 0 | eQTL (rs11769827, PP=0.569) | E | - | cellular; behavior/neurological; nervous system; | - |
| ELN | 2006 | ENSG00000049540 | protein_coding | 54 | rs11770437 | 1 | 1 | 0 | 1 | 1 | 1 | - | E (UQ) | M1_hub | homeostasis/metabolism; muscle; cardiovascular system; mortality/aging; respiratory system; | Cutis laxa, autosomal dominant, 123700 (3), Autosomal dominant; Supravalvar aortic stenosis, 185500 (3), Autosomal dominant |
| WBSCR28 | 135886 | ENSG00000175877 | protein_coding | 54 | rs11770437 | 0 | 1 | 0 | 0 | 1 | 0 | - | E | - | - | - |
| CLDN3 | 1365 | ENSG00000165215 | protein_coding | 54 | rs11770437 | 1 | 1 | 1 | 0 | 0 | 0 | - | E+ (UQ) | - | behavior/neurological; | - |
| CLDN4 | 1364 | ENSG00000189143 | protein_coding | 54 | rs11770437 | 0 | 1 | 1 | 0 | 0 | 0 | - | E (UQ) | - | mortality/aging; hematopoietic system; cardiovascular system; renal/urinary system; cellular; homeostasis/metabolism; growth/size/body region; | - |
| LINC00035 | 171022 | ENSG00000225969 | ranscribed_unitary_pseudogen | 54 | rs11770437 | 0 | 1 | 1 | 0 | 0 | 0 | - | E | - | - | - |
| CLIP2 | 7461 | ENSG00000106665 | protein_coding | 54 | rs11770437 | 0 | 0 | 0 | 1 | 0 | 0 | - | E (UQ) | - | skeleton; nervous system; behavior/neurological; cellular; growth/size/body region; | - |
| FZD9 | 8326 | ENSG00000188763 | protein_coding | 54 | rs11770437 | 0 | 0 | 1 | 0 | 0 | 0 | - | E | M4 | growth/size/body region; endocrine/exocrine gland; cellular; skeleton; behavior/neurological; mortality/aging; hematopoietic system; nervous system; immune system; | - |
| NSUN5 | 55695 | ENSG00000130305 | protein_coding | 54 | rs11770437 | 0 | 0 | 1 | 0 | 0 | 0 | - | E (UQ) | - | hematopoietic system; behavior/neurological; nervous system; | - |
| STX1A | 6804 | ENSG00000106089 | protein_coding | 54 | rs11770437 | 0 | 0 | 1 | 0 | 0 | 0 | - | E | M7 | embryo; behavior/neurological; mortality/aging; nervous system; growth/size/body region; adipose tissue; homeostasis/metabolism; | - |
| VPS37D | 155382 | ENSG00000176428 | protein_coding | 54 | rs11770437 | 0 | 0 | 1 | 0 | 0 | 0 | - | E | - | vision/eye; mortality/aging; cardiovascular system; behavior/neurological; growth/size/body region; | - |
| TRIM56 | 81844 | ENSG00000169871 | protein_coding | 55 | rs4556017 | 1 | 1 | 1 | 0 | 0 | 0 | eQTL (rs4556017, PP=0.962) | E (UQ) | - | hematopoietic system; immune system; homeostasis/metabolism; growth/size/body region; | - |
| ACHE | 43 | ENSG00000087085 | protein_coding | 55 | rs4556017 | 1 | 0 | 1 | 0 | 0 | 0 | eQTL (rs4556017, PP=0.962) | E (UQ) | M4 | respiratory system; behavior/neurological; reproductive system; mortality/aging; hematopoietic system; nervous system; hearing/vestibular/ear; vision/eye; growth/size/body region; adipose tissue; muscle; craniofacial; homeostasis/metabolism; | [Blood group, Yt system], 112100 (3) |

| | | | | | | | | | | | | | | | | |
|---|---|---|---|---|---|---|---|---|---|---|---|---|---|---|---|---|
| COPS6 | 10980 | ENSG00000168090 | protein_coding | 55 | rs4556017 | 1 | 0 | 1 | 0 | 0 | 0 | eQTL (rs4556017, PP=0.962) | E (UQ) | - | mortality/aging; hematopoietic system; neoplasm; immune system; endocrine/exocrine gland; cellular; homeostasis/metabolism; | - |
| SRRT | 51593 | ENSG00000087087 | protein_coding | 55 | rs4556017 | 1 | 0 | 1 | 0 | 0 | 0 | eQTL (rs4556017, PP=0.962) | E (UQ) | M7 | endocrine/exocrine gland; cellular; hematopoietic system; mortality/aging; embryo; immune system; | - |
| TRIP6 | 7205 | ENSG00000087077 | protein_coding | 55 | rs4556017 | 1 | 0 | 1 | 0 | 0 | 0 | eQTL (rs4556017, PP=0.962) | E (UQ) | - | nervous system; immune system; skeleton; hematopoietic system; craniofacial; | - |
| MUC12 | 10071 | ENSG00000205277 | protein_coding | 55 | rs4556017 | 1 | 1 | 1 | 1 | 1 | 0 | - | E+ (UQ) | - | - | - |
| MUC17 | 140453 | ENSG00000169876 | protein_coding | 55 | rs4556017 | 1 | 1 | 1 | 1 | 0 | 0 | - | E+ (UQ) | - | - | - |
| RP11-395B7.4 | 102724094 | ENSG00000227053 | lncRNA | 55 | rs4556017 | 0 | 1 | 1 | 0 | 1 | 0 | - | E | M1 | - | - |
| MUC3A | 4584 | ENSG00000169894 | protein_coding | 55 | rs4556017 | 0 | 1 | 1 | 0 | 0 | 0 | - | E (UQ) | - | integument; | - |
| AP1S1 | 1174 | ENSG00000106367 | protein_coding | 55 | rs4556017 | 0 | 0 | 1 | 0 | 0 | 0 | - | E (UQ) | M4 | mortality/aging; | MEDNIK syndrome, 609313 (3), Autosomal recessive |
| CLDN15 | 24146 | ENSG00000106404 | protein_coding | 55 | rs4556017 | 0 | 0 | 1 | 0 | 0 | 0 | - | E | - | digestive/alimentary; cellular; endocrine/exocrine gland; | - |
| EPHB4 | 2050 | ENSG00000196411 | protein_coding | 55 | rs4556017 | 0 | 0 | 1 | 0 | 0 | 0 | - | E (UQ) | - | muscle; growth/size/body region; renal/urinary system; cardiovascular system; mortality/aging; embryo; | Capillary malformation-arteriovenous malformation 2, 618196 (3), Autosomal dominant; Lymphatic malformation 7, 617300 (3), Autosomal dominant |
| GIGYF1 | 64599 | ENSG00000146830 | protein_coding | 55 | rs4556017 | 0 | 0 | 1 | 0 | 0 | 0 | - | E | - | - | - |
| MEPCE | 56257 | ENSG00000146834 | protein_coding | 55 | rs4556017 | 0 | 0 | 1 | 0 | 0 | 0 | - | E (UQ) | - | hematopoietic system; mortality/aging; | - |
| MIR25 | 407014 | ENSG00000207547 | miRNA | 55 | rs4556017 | 0 | 0 | 1 | 0 | 0 | 0 | - | E (UQ) | - | - | - |
| MOSPD3 | 64598 | ENSG00000106330 | protein_coding | 55 | rs4556017 | 0 | 0 | 1 | 0 | 0 | 0 | - | E | M7 | cardiovascular system; mortality/aging; muscle; | - |
| NAT16 | 375607 | ENSG00000167011 | protein_coding | 55 | rs4556017 | 0 | 0 | 1 | 0 | 0 | 0 | - | E | - | - | - |
| NYAP1 | 222950 | ENSG00000166924 | protein_coding | 55 | rs4556017 | 0 | 0 | 1 | 0 | 0 | 0 | - | E | - | - | - |
| PCOLCE | 5118 | ENSG00000106333 | protein_coding | 55 | rs4556017 | 0 | 0 | 1 | 0 | 0 | 0 | - | E (UQ) | - | limbs/digits/tail; muscle; skeleton; | - |
| PILRB | 29990 | ENSG00000121716 | protein_coding | 55 | rs4556017 | 0 | 0 | 1 | 0 | 0 | 0 | - | E (UQ) | - | mortality/aging; homeostasis/metabolism; immune system; | - |
| RP11-758P17.2 | - | ENSG00000241357 | lncRNA | 55 | rs4556017 | 0 | 0 | 1 | 0 | 0 | 0 | - | NE | - | - | - |
| SLC12A9 | 56996 | ENSG00000146828 | protein_coding | 55 | rs4556017 | 0 | 0 | 1 | 0 | 0 | 0 | - | E | - | - | - |
| SPDYE3 | 441272 | ENSG00000214300 | protein_coding | 55 | rs4556017 | 0 | 0 | 1 | 0 | 0 | 0 | - | E | - | - | - |
| TAG3L5P-PVRIG2P-PILR | 101752399 | ENSG00000272752 | lncRNA | 55 | rs4556017 | 0 | 0 | 1 | 0 | 0 | 0 | - | E (UQ) | - | - | - |
| UFSP1 | 402682 | ENSG00000176125 | protein_coding | 55 | rs4556017 | 0 | 0 | 1 | 0 | 0 | 0 | - | E | M4 | - | - |
| ARMC10 | 83787 | ENSG00000170632 | protein_coding | 56 | rs7778418 | 0 | 1 | 1 | 1 | 1 | 0 | - | E (UQ) | - | - | - |
| FAM185A | 222234 | ENSG00000222011 | protein_coding | 56 | rs7778418 | 0 | 1 | 1 | 1 | 1 | 0 | - | E | - | - | - |
| FBXL13 | 222235 | ENSG00000161040 | protein_coding | 56 | rs7778418 | 0 | 1 | 1 | 1 | 1 | 0 | - | E | - | skeleton; | - |
| LRRC17 | 10234 | ENSG00000128606 | protein_coding | 56 | rs7778418 | 1 | 1 | 1 | 1 | 1 | 1 | - | E | M1 | - | - |
| NAPEPLD | 222236 | ENSG00000161048 | protein_coding | 56 | rs7778418 | 0 | 1 | 1 | 1 | 1 | 0 | - | E | - | homeostasis/metabolism; nervous system; | - |
| SLC26A5 | 375611 | ENSG00000170615 | protein_coding | 56 | rs7778418 | 0 | 1 | 1 | 1 | 0 | 0 | - | E | - | growth/size/body region; hearing/vestibular/ear; nervous system; | ?Deafness, autosomal recessive 61, 613865 (3), Autosomal recessive |
| NFE4 | 58160 | ENSG00000230257 | protein_coding | 56 | rs7778418 | 0 | 1 | 1 | 0 | 1 | 0 | - | E | - | - | - |
| CTB-107G13.1 | 101927870 | ENSG00000234715 | lncRNA | 56 | rs7778418 | 0 | 1 | 1 | 0 | 0 | 0 | - | E | M7 | - | - |
| RELN | 5649 | ENSG00000189056 | protein_coding | 56 | rs7778418 | 0 | 1 | 1 | 0 | 0 | 0 | - | E | - | reproductive system; mortality/aging; behavior/neurological; skeleton; nervous system; digestive/alimentary; vision/eye; integument; growth/size/body region; adipose tissue; cellular; homeostasis/metabolism; muscle; | {Epilepsy, familial temporal lobe, 7}, 616436 (3), Autosomal dominant; Lissencephaly 2 (Norman-Roberts type), 257320 (3), Autosomal recessive |
| RP11-645N11.3 | - | ENSG00000236226 | lncRNA | 56 | rs7778418 | 0 | 1 | 1 | 0 | 0 | 0 | - | NE | - | - | - |
| RNU6-1136P | 106480066 | ENSG00000252643 | snRNA | 56 | rs7778418 | 0 | 1 | 0 | 0 | 0 | 0 | - | NE | - | - | - |
| PMPCB | 9512 | ENSG00000105819 | protein_coding | 56 | rs7778418 | 0 | 0 | 1 | 1 | 0 | 0 | - | E (UQ) | - | mortality/aging; nervous system; | Multiple mitochondrial dysfunctions syndrome 6, 617954 (3), Autosomal recessive |
| DNAJC2 | 27000 | ENSG00000105821 | protein_coding | 56 | rs7778418 | 0 | 0 | 1 | 0 | 0 | 0 | - | E (UQ) | - | - | - |
| LRWD1 | 222229 | ENSG00000161036 | protein_coding | 56 | rs7778418 | 0 | 0 | 1 | 0 | 0 | 0 | - | E | - | - | - |
| ORC5 | 5001 | ENSG00000164815 | protein_coding | 56 | rs7778418 | 0 | 0 | 1 | 0 | 0 | 0 | - | E (UQ) | - | - | - |
| POLR2J | 5439 | ENSG00000005075 | protein_coding | 56 | rs7778418 | 0 | 0 | 1 | 0 | 0 | 0 | - | E (UQ) | M4 | - | - |
| POLR2J2 | 246721 | ENSG00000228049 | protein_coding | 56 | rs7778418 | 0 | 0 | 1 | 0 | 0 | 0 | - | E | M4 | - | - |
| POLR2J2 | 246721 | ENSG00000267645 | protein_coding | 56 | rs7778418 | 1 | 0 | 1 | 0 | 0 | 0 | - | E (UQ) | M7_hub | - | - |
| POLR2J3 | 548644 | ENSG00000168255 | protein_coding | 56 | rs7778418 | 0 | 0 | 1 | 0 | 0 | 0 | - | E (UQ) | M7 | - | - |
| PRKRIP1 | 79706 | ENSG00000128563 | protein_coding | 56 | rs7778418 | 0 | 0 | 1 | 0 | 0 | 0 | - | E (UQ) | - | - | - |
| RASA4 | 10156 | ENSG00000105808 | protein_coding | 56 | rs7778418 | 0 | 0 | 1 | 0 | 0 | 0 | - | E (UQ) | - | cellular; immune system; hematopoietic system; | - |
| RASA4B | 100271927 | ENSG00000170667 | protein_coding | 56 | rs7778418 | 0 | 0 | 1 | 0 | 0 | 0 | - | E | - | - | - |
| RP11-514P8.6 | - | ENSG00000205236 | protein_coding | 56 | rs7778418 | 0 | 0 | 1 | 0 | 0 | 0 | - | E | M7 | - | - |
| SPDYE2 | 441273 | ENSG00000205238 | protein_coding | 56 | rs7778418 | 0 | 0 | 1 | 0 | 0 | 0 | - | E | - | - | - |
| SPDYE2B | 100310812 | ENSG00000173678 | protein_coding | 56 | rs7778418 | 0 | 0 | 1 | 0 | 0 | 0 | - | E | - | - | - |
| UPK3BL | 100134938 | ENSG00000267368 | protein_coding | 56 | rs7778418 | 1 | 0 | 1 | 0 | 0 | 0 | - | E (UQ) | M7_hub | - | - |
| SND1 | 27044 | ENSG00000197157 | protein_coding | 57 | rs806169 | 0 | 1 | 1 | 1 | 1 | 0 | - | E (UQ) | - | mortality/aging; | - |
| PAX4 | 5078 | ENSG00000106331 | protein_coding | 57 | rs806169 | 0 | 1 | 0 | 1 | 1 | 0 | - | E | - | homeostasis/metabolism; endocrine/exocrine gland; mortality/aging; growth/size/body region; behavior/neurological; | Maturity-onset diabetes of the young, type IX, 612225 (3); Diabetes mellitus, type 2, 125853 (3), Autosomal dominant; {Diabetes mellitus, ketosis-prone, susceptibility to}, 612227 (3), Autosomal dominant, Autosomal recessive |
| LRRC4 | 64101 | ENSG00000128594 | protein_coding | 57 | rs806169 | 0 | 1 | 1 | 0 | 1 | 0 | - | E | M7 | hearing/vestibular/ear; behavior/neurological; | - |
| AC073934.6 | - | ENSG00000240790 | lncRNA | 57 | rs806169 | 0 | 1 | 0 | 0 | 0 | 0 | - | NE | - | - | - |
| MIR593 | 693178 | ENSG00000207588 | miRNA | 57 | rs806169 | 0 | 1 | 0 | 0 | 0 | 0 | - | NE | - | - | - |
| GCC1 | 79571 | ENSG00000179562 | protein_coding | 57 | rs806169 | 0 | 0 | 1 | 1 | 1 | 0 | - | E | M4 | - | - |
| FSCN3 | 29999 | ENSG00000106328 | protein_coding | 57 | rs806169 | 0 | 0 | 0 | 1 | 1 | 0 | - | E (UQ) | - | - | - |
| CCDC136 | 64753 | ENSG00000128596 | protein_coding | 57 | rs806169 | 0 | 0 | 1 | 0 | 0 | 0 | - | E | M1 | cellular; reproductive system; | - |
| ZNF800 | 168850 | ENSG00000048405 | protein_coding | 57 | rs806169 | 0 | 0 | 1 | 0 | 0 | 0 | - | E | - | - | - |
| ANGPT2 | 285 | ENSG00000091879 | protein_coding | 58 | rs2912053 | 0 | 1 | 1 | 1 | 1 | 0 | - | E | M1 | respiratory system; cardiovascular system; mortality/aging; vision/eye; digestive/alimentary; immune system; cellular; integument; homeostasis/metabolism; | - |
| MCPH1 | 79648 | ENSG00000147316 | protein_coding | 58 | rs2912053 | 0 | 1 | 1 | 1 | 1 | 0 | - | E | - | cellular; homeostasis/metabolism; integument; growth/size/body region; endocrine/exocrine gland; pigmentation; nervous system; hearing/vestibular/ear; vision/eye; immune system; skeleton; behavior/neurological; reproductive system; mortality/aging; hematopoietic system; cardiovascular system; | Microcephaly 1, primary, autosomal recessive, 251200 (3), Autosomal recessive |
| GS1-24F4.2 | 100652791 | ENSG00000245857 | lncRNA | 58 | rs2912053 | 0 | 0 | 0 | 0 | 1 | 0 | - | E | - | - | - |
| RP11-115C21.2 | 100287015 | ENSG00000246089 | lncRNA | 58 | rs2912053 | 0 | 0 | 1 | 0 | 0 | 0 | - | E | - | - | - |
| BIN3 | 55909 | ENSG00000147439 | protein_coding | 59 | rs13271626 | 0 | 1 | 1 | 1 | 1 | 0 | - | E (UQ) | - | immune system; respiratory system; liver/biliary system; neoplasm; hematopoietic system; vision/eye; | - |
| C8orf58 | 541565 | ENSG00000241852 | protein_coding | 59 | rs13271626 | 1 | 1 | 1 | 1 | 1 | 1 | - | E | - | homeostasis/metabolism; integument; | - |
| CCAR2 | 57805 | ENSG00000158941 | protein_coding | 59 | rs13271626 | 0 | 1 | 1 | 1 | 1 | 0 | - | E (UQ) | - | adipose tissue; growth/size/body region; immune system; homeostasis/metabolism; liver/biliary system; behavior/neurological; | - |
| PDLIM2 | 64236 | ENSG00000120913 | protein_coding | 59 | rs13271626 | 0 | 1 | 1 | 1 | 1 | 0 | - | E (UQ) | M7 | immune system; hematopoietic system; | - |
| AC037459.4 | - | ENSG00000248235 | protein_coding | 59 | rs13271626 | 0 | 1 | 1 | 1 | 0 | 0 | - | E | M7 | - | - |
| RP11-582J16.5 | - | ENSG00000253200 | lncRNA | 59 | rs13271626 | 0 | 1 | 1 | 0 | 1 | 0 | - | E | - | - | - |
| CTD-3247F14.2 | - | ENSG00000261026 | lncRNA | 59 | rs13271626 | 0 | 1 | 1 | 0 | 0 | 0 | - | E | - | - | - |
| EGR3 | 1960 | ENSG00000179388 | protein_coding | 59 | rs13271626 | 0 | 1 | 1 | 0 | 0 | 0 | - | E | - | homeostasis/metabolism; muscle; behavior/neurological; skeleton; nervous system; vision/eye; mortality/aging; | - |
| RP11-459E5.1 | - | ENSG00000253125 | lncRNA | 59 | rs13271626 | 0 | 1 | 0 | 0 | 0 | 0 | - | E | M1 | - | - |
| EYA1 | 2138 | ENSG00000104313 | protein_coding | 60 | rs1838392 | 1 | 1 | 1 | 1 | 1 | 1 | - | E | - | endocrine/exocrine gland; adipose tissue; growth/size/body region; craniofacial; muscle; cellular; homeostasis/metabolism; immune system; respiratory system; behavior/neurological; embryo; reproductive system; mortality/aging; cardiovascular system; hematopoietic system; vision/eye; hearing/vestibular/ear; nervous system; digestive/alimentary; skeleton; renal/urinary system; | ?Otofaciocervical syndrome, 166780 (3), Autosomal dominant; Anterior segment anomalies with or without cataract, 602588 (3), Autosomal dominant; Branchiootorenal syndrome 1, with or without cataracts, 113650 (3), Autosomal dominant; Branchiootic syndrome 1, 602588 (3), Autosomal dominant |
| LACTB2 | 51110 | ENSG00000147592 | protein_coding | 60 | rs1838392 | 0 | 1 | 1 | 1 | 1 | 0 | - | E | - | - | - |
| RP11-382J12.1 | 286190 | ENSG00000246366 | lncRNA | 60 | rs1838392 | 0 | 1 | 1 | 1 | 1 | 0 | - | E | - | - | - |
| TRAM1 | 23471 | ENSG00000067167 | protein_coding | 60 | rs1838392 | 0 | 1 | 1 | 1 | 1 | 0 | - | E (UQ) | - | - | - |
| XKR9 | 389668 | ENSG00000221947 | protein_coding | 60 | rs1838392 | 1 | 1 | 1 | 1 | 0 | 0 | - | E+ | M4 | - | - |
| NCOA2 | 10499 | ENSG00000140396 | protein_coding | 60 | rs1838392 | 1 | 1 | 1 | 0 | 1 | 1 | - | E | - | embryo; reproductive system; integument; growth/size/body region; cellular; endocrine/exocrine gland; | - |
| BTF3P12 | 652963 | ENSG00000213003 | processed_pseudogene | 60 | rs1838392 | 0 | 0 | 0 | 0 | 1 | 0 | - | E | - | - | - |
| RP11-326E22.1 | - | ENSG00000254031 | lncRNA | 60 | rs1838392 | 0 | 0 | 1 | 0 | 0 | 0 | - | NE | - | - | - |
| RP11-333A23.4 | 101926892 | ENSG00000253967 | lncRNA | 60 | rs1838392 | 0 | 0 | 1 | 0 | 0 | 0 | - | E | - | - | - |
| ZFHX4-AS1 | 100192378 | ENSG00000253661 | lncRNA | 61 | rs2581260 | 0 | 1 | 0 | 0 | 1 | 0 | - | E | - | - | - |
| RP11-115I9.1 | - | ENSG00000253681 | lncRNA | 61 | rs2581260 | 0 | 1 | 0 | 0 | 0 | 0 | - | NE | - | - | - |
| GSDMC | 56169 | ENSG00000147697 | protein_coding | 62 | rs10956488 | 1 | 0 | 1 | 0 | 0 | 0 | eQTL (rs10956488, PP=0.962) | E | M7 | - | - |
| GRINA | 2907 | ENSG00000178719 | protein_coding | 63 | rs58579887 | 0 | 1 | 1 | 1 | 1 | 0 | - | E (UQ) | - | - | - |
| PARP10 | 84875 | ENSG00000178685 | protein_coding | 63 | rs58579887 | 0 | 1 | 1 | 1 | 1 | 0 | - | E (UQ) | - | nervous system; | - |
| PLEC | 5339 | ENSG00000178209 | protein_coding | 63 | rs58579887 | 1 | 1 | 1 | 1 | 1 | 1 | - | E (UQ) | - | integument; growth/size/body region; cellular; homeostasis/metabolism; muscle; mortality/aging; cardiovascular system; hematopoietic system; behavior/neurological; immune system; nervous system; | Muscular dystrophy, limb-girdle, autosomal recessive 17, 613723 (3), Autosomal recessive; Epidermolysis bullosa simplex with pyloric atresia, 612138 (3), Autosomal recessive; Epidermolysis bullosa simplex with muscular dystrophy, 226670 (3), Autosomal recessive; ?Epidermolysis bullosa simplex with nail dystrophy, 616487 (3), Autosomal recessive; Epidermolysis bullosa simplex, Ogna type, 131950 (3), Autosomal dominant |
| SPATC1 | 375686 | ENSG00000186583 | protein_coding | 63 | rs58579887 | 0 | 1 | 1 | 0 | 0 | 0 | - | E | - | - | - |
| MIR661 | 724031 | ENSG00000207574 | miRNA | 63 | rs58579887 | 0 | 1 | 0 | 0 | 0 | 0 | - | NE | - | - | - |
| AC084125.4 | - | ENSG00000232600 | lncRNA | 63 | rs58579887 | 0 | 0 | 1 | 0 | 0 | 0 | - | NE | - | - | - |
| CPSF1 | 29894 | ENSG00000071894 | protein_coding | 63 | rs58579887 | 0 | 0 | 1 | 0 | 0 | 0 | - | E (UQ) | - | - | Myopia 27, 618827 (3), Autosomal dominant |
| CTD-3065J16.6 | - | ENSG00000204791 | ranscribed_unitary_pseudogen | 63 | rs58579887 | 0 | 0 | 1 | 0 | 0 | 0 | - | E | - | - | - |
| FAM203A | 51236 | ENSG00000235173 | protein_coding | 63 | rs58579887 | 0 | 0 | 1 | 0 | 0 | 0 | - | E | - | - | - |
| GPAA1 | 8733 | ENSG00000197858 | protein_coding | 63 | rs58579887 | 0 | 0 | 1 | 0 | 0 | 0 | - | E (UQ) | - | nervous system; mortality/aging; embryo; growth/size/body region; | Glycosylphosphatidylinositol biosynthesis defect 15, 617810 (3), Autosomal recessive |
| KIAA1875 | 340390 | ENSG00000179698 | protein_coding | 63 | rs58579887 | 0 | 0 | 1 | 0 | 0 | 0 | - | E | - | - | - |
| MAF1 | 84232 | ENSG00000179632 | protein_coding | 63 | rs58579887 | 0 | 0 | 1 | 0 | 0 | 0 | - | E (UQ) | - | homeostasis/metabolism; adipose tissue; growth/size/body region; reproductive system; mortality/aging; integument; liver/biliary system; behavior/neurological; | - |
| MROH1 | 727957 | ENSG00000179832 | protein_coding | 63 | rs58579887 | 0 | 0 | 1 | 0 | 0 | 0 | - | E (UQ) | M4 | - | - |
| NRBP2 | 340371 | ENSG00000185189 | protein_coding | 63 | rs58579887 | 0 | 0 | 1 | 0 | 0 | 0 | - | E (UQ) | M1 | hematopoietic system; immune system; homeostasis/metabolism; | - |
| OPLAH | 26873 | ENSG00000178814 | protein_coding | 63 | rs58579887 | 0 | 0 | 1 | 0 | 0 | 0 | - | E | M4 | behavior/neurological; vision/eye; | 5-oxoprolinase deficiency, 260005 (3), Autosomal dominant, Autosomal recessive |
| PPP1R16A | 84988 | ENSG00000160972 | protein_coding | 63 | rs58579887 | 1 | 0 | 1 | 0 | 0 | 0 | - | E (UQ) | M4_hub | - | - |
| RP11-299M14.2 | - | ENSG00000255343 | lncRNA | 63 | rs58579887 | 0 | 0 | 1 | 0 | 0 | 0 | - | E | M1 | - | - |
| RP11-661A12.9 | - | ENSG00000255050 | lncRNA | 63 | rs58579887 | 0 | 0 | 1 | 0 | 0 | 0 | - | E | - | - | - |
| SHARPIN | 81858 | ENSG00000179526 | protein_coding | 63 | rs58579887 | 0 | 0 | 1 | 0 | 0 | 0 | - | E (UQ) | M4 | digestive/alimentary; vision/eye; immune system; skeleton; respiratory system; liver/biliary system; reproductive system; hematopoietic system; cardiovascular system; craniofacial; cellular; homeostasis/metabolism; integument; growth/size/body region; | - |
| ZNF517 | 340385 | ENSG00000197363 | protein_coding | 63 | rs58579887 | 0 | 0 | 1 | 0 | 0 | 0 | - | E | M1 | - | - |
| ZNF623 | 9831 | ENSG00000183309 | protein_coding | 63 | rs58579887 | 0 | 0 | 1 | 0 | 0 | 0 | - | E | - | - | - |

| | | | | | | | | | | | | | | | |
|---|---|---|---|---|---|---|---|---|---|---|---|---|---|---|---|
| CDKN2B-AS1 | 100048912 | ENSG00000240498 | lncRNA | 64 | rs1333047 | 1 | 1 | 1 | 0 | 1 | 1 | eQTL (rs1333047, PP=0.520) | E+ (UQ) | - | - | - |
| RP11-145E5.5 | - | ENSG00000264545 | protein_coding | 64 | rs1333047 | 0 | 1 | 0 | 1 | 0 | 0 | - | E | M7 | - | - |
| CDKN2A | 1029 | ENSG00000147889 | protein_coding | 64 | rs1333047 | 0 | 1 | 1 | 0 | 0 | 0 | - | E | M7 | integument; endocrine/exocrine gland; cellular; homeostasis/metabolism; renal/urinary system; skeleton; behavior/neurological; respiratory system; mortality/aging; reproductive system; hematopoietic system; neoplasm; nervous system; digestive/alimentary; vision/eye; immune system; | {Melanoma and neural system tumor syndrome}, 155755 (3), Autosomal dominant; {Melanoma, cutaneous malignant, 2}, 155601 (3), Autosomal dominant; {Melanoma-pancreatic cancer syndrome}, 606719 (3), Autosomal dominant |
| CDKN2B | 1030 | ENSG00000147883 | protein_coding | 64 | rs1333047 | 0 | 1 | 1 | 0 | 0 | 0 | - | E (UQ) | - | immune system; neoplasm; hematopoietic system; mortality/aging; cellular; | - |
| RP11-149I2.4 | - | ENSG00000266446 | lncRNA | 64 | rs1333047 | 0 | 1 | 0 | 0 | 0 | 0 | - | NE | - | - | - |
| ASPN | 54829 | ENSG00000106819 | protein_coding | 65 | rs755209 | 1 | 1 | 1 | 1 | 1 | 1 | - | E | M1 | hearing/vestibular/ear; vision/eye; | {Osteoarthritis susceptibility 3}, 607850 (3), Autosomal dominant; {Lumbar disc degeneration}, 603932 (3) |
| CENPP | 401541 | ENSG00000188312 | protein_coding | 65 | rs755209 | 0 | 1 | 1 | 1 | 1 | 0 | - | E | M7 | - | - |
| ECM2 | 1842 | ENSG00000106823 | protein_coding | 65 | rs755209 | 0 | 1 | 1 | 1 | 1 | 0 | - | E | M1 | - | - |
| BICD2 | 23299 | ENSG00000185963 | protein_coding | 65 | rs755209 | 1 | 1 | 1 | 0 | 1 | 1 | - | E (UQ) | - | mortality/aging; behavior/neurological; skeleton; nervous system; growth/size/body region; cellular; craniofacial; | Spinal muscular atrophy, lower extremity-predominant, 2B, autosomal dominant, 618291 (3), Autosomal dominant; Spinal muscular atrophy, lower extremity-predominant, 2A, autosomal dominant, 615290 (3), Autosomal dominant |
| IARS | 3376 | ENSG00000196305 | protein_coding | 65 | rs755209 | 0 | 1 | 1 | 0 | 1 | 0 | - | E (UQ) | - | - | - |
| IPPK | 64768 | ENSG00000127080 | protein_coding | 65 | rs755209 | 0 | 1 | 1 | 0 | 1 | 0 | - | E (UQ) | M7 | mortality/aging; cardiovascular system; | - |
| NOL8 | 55035 | ENSG00000198000 | protein_coding | 65 | rs755209 | 0 | 1 | 1 | 0 | 1 | 0 | - | E (UQ) | - | mortality/aging; behavior/neurological; growth/size/body region; homeostasis/metabolism; | - |
| OGN | 4969 | ENSG00000106809 | protein_coding | 65 | rs755209 | 1 | 1 | 1 | 0 | 1 | 1 | - | E (UQ) | M1 | homeostasis/metabolism; integument; hematopoietic system; vision/eye; immune system; | - |
| OMD | 4958 | ENSG00000127083 | protein_coding | 65 | rs755209 | 1 | 1 | 1 | 0 | 1 | 1 | - | E | M1 | skeleton; behavior/neurological; | - |
| MIR4670 | 100616351 | ENSG00000264158 | miRNA | 65 | rs755209 | 0 | 1 | 0 | 0 | 0 | 0 | - | NE | - | - | - |
| RP11-62C3.6 | - | ENSG00000236115 | lncRNA | 65 | rs755209 | 0 | 1 | 0 | 0 | 0 | 0 | - | NE | - | - | - |
| SNORA84 | 100124534 | ENSG00000239183 | snoRNA | 65 | rs755209 | 0 | 1 | 0 | 0 | 0 | 0 | - | E | - | - | - |
| FGD3 | 89846 | ENSG00000127084 | protein_coding | 65 | rs755209 | 0 | 0 | 1 | 0 | 0 | 0 | - | E | - | - | - |
| LINC00475 | 158314 | ENSG00000225511 | scribed_unprocessed_pseudog | 65 | rs755209 | 0 | 0 | 1 | 0 | 0 | 0 | - | E | - | - | - |
| ROR2 | 4920 | ENSG00000169071 | protein_coding | 65 | rs755209 | 0 | 0 | 1 | 0 | 0 | 0 | - | E | M1 | adipose tissue; endocrine/exocrine gland; growth/size/body region; craniofacial; cellular; homeostasis/metabolism; respiratory system; embryo; mortality/aging; reproductive system; cardiovascular system; vision/eye; hearing/vestibular/ear; nervous system; limbs/digits/tail; digestive/alimentary; skeleton; | Brachydactyly, type B1, 113000 (3), Autosomal dominant; Robinow syndrome, autosomal recessive, 268310 (3), Autosomal recessive |
| SFT2D1 | 113402 | ENSG00000198818 | protein_coding | 65 | rs755209 | 0 | 0 | 1 | 0 | 0 | 0 | - | E (UQ) | - | - | - |
| SPTLC1 | 10558 | ENSG00000090054 | protein_coding | 65 | rs755209 | 0 | 0 | 1 | 0 | 0 | 0 | - | E (UQ) | - | mortality/aging; homeostasis/metabolism; | Neuropathy, hereditary sensory and autonomic, type IA, 162400 (3), Autosomal dominant |
| ZNF484 | 83744 | ENSG00000127081 | protein_coding | 65 | rs755209 | 0 | 0 | 1 | 0 | 0 | 0 | - | E | - | - | - |
| PAPPA | 5069 | ENSG00000182752 | protein_coding | 66 | rs1858015 | 0 | 1 | 0 | 1 | 1 | 0 | - | E | M1 | growth/size/body region; homeostasis/metabolism; cardiovascular system; skeleton; | - |
| ASTN2 | 23245 | ENSG00000148219 | protein_coding | 66 | rs1858015 | 0 | 1 | 0 | 0 | 0 | 0 | - | E | - | - | - |
| PAPPA-AS1 | 493913 | ENSG00000256040 | lncRNA | 66 | rs1858015 | 0 | 1 | 0 | 0 | 0 | 0 | - | NE | - | - | - |
| PAPPA-AS2 | 103611155 | ENSG00000226604 | lncRNA | 66 | rs1858015 | 0 | 0 | 1 | 0 | 0 | 0 | - | NE | - | - | - |
| MED22 | 6837 | ENSG00000148297 | protein_coding | 67 | rs676996 | 0 | 1 | 1 | 1 | 0 | 0 | - | E | M1 | behavior/neurological; hematopoietic system; mortality/aging; skeleton; growth/size/body region; | - |
| OBP2B | 29989 | ENSG00000171102 | protein_coding | 67 | rs676996 | 0 | 1 | 1 | 1 | 0 | 0 | - | NE | - | integument; pigmentation; | - |
| ABO | 28 | ENSG00000175164 | protein_coding | 67 | rs676996 | 0 | 1 | 1 | 0 | 1 | 0 | - | E (UQ) | M4 | - | [Blood group, ABO system], 616093 (3) |
| ADAMTS13 | 11093 | ENSG00000160323 | protein_coding | 67 | rs676996 | 0 | 1 | 1 | 0 | 0 | 0 | - | E | - | homeostasis/metabolism; mortality/aging; hematopoietic system; immune system; | Thrombotic thrombocytopenic purpura, hereditary, 274150 (3), Autosomal recessive |
| CACFD1 | 11094 | ENSG00000160325 | protein_coding | 67 | rs676996 | 1 | 1 | 1 | 0 | 0 | 0 | - | E (UQ) | M4_hub | homeostasis/metabolism; cellular; behavior/neurological; neoplasm; | - |
| REXO4 | 57109 | ENSG00000148300 | protein_coding | 67 | rs676996 | 0 | 1 | 1 | 0 | 0 | 0 | - | E | - | - | - |
| SLC2A6 | 11182 | ENSG00000160326 | protein_coding | 67 | rs676996 | 0 | 1 | 1 | 0 | 0 | 0 | - | E | M1 | homeostasis/metabolism; adipose tissue; | - |
| SURF1 | 6834 | ENSG00000148290 | protein_coding | 67 | rs676996 | 0 | 1 | 1 | 0 | 0 | 0 | - | E (UQ) | - | homeostasis/metabolism; cellular; growth/size/body region; nervous system; limbs/digits/tail; embryo; behavior/neurological; reproductive system; mortality/aging; | Leigh syndrome, due to COX IV deficiency, 256000 (3), Autosomal recessive, Mitochondrial; Charcot-Marie-Tooth disease, type 4K, 616684 (3), Autosomal recessive |
| SURF2 | 6835 | ENSG00000148291 | protein_coding | 67 | rs676996 | 0 | 1 | 1 | 0 | 0 | 0 | - | E (UQ) | M7 | - | - |
| SURF6 | 6838 | ENSG00000148296 | protein_coding | 67 | rs676996 | 0 | 1 | 1 | 0 | 0 | 0 | - | E (UQ) | M4 | - | - |
| C9orf96 | 169436 | ENSG00000198870 | protein_coding | 67 | rs676996 | 0 | 1 | 0 | 0 | 0 | 0 | - | E | - | - | - |
| RPL7A | 6130 | ENSG00000148303 | protein_coding | 67 | rs676996 | 0 | 1 | 0 | 0 | 0 | 0 | - | E (UQ) | - | - | - |
| SNORD24 | 26820 | ENSG00000206611 | snoRNA | 67 | rs676996 | 0 | 1 | 0 | 0 | 0 | 0 | - | E (UQ) | - | - | - |
| SNORD36A | 26815 | ENSG00000199744 | snoRNA | 67 | rs676996 | 0 | 1 | 0 | 0 | 0 | 0 | - | E | - | - | - |
| SNORD36B | 26814 | ENSG00000200831 | snoRNA | 67 | rs676996 | 1 | 1 | 0 | 0 | 0 | 0 | - | E- | - | - | - |
| SNORD36C | 26813 | ENSG00000252542 | snoRNA | 67 | rs676996 | 1 | 1 | 0 | 0 | 0 | 0 | - | E- (UQ) | - | - | - |
| SURF4 | 6836 | ENSG00000148248 | protein_coding | 67 | rs676996 | 0 | 1 | 0 | 0 | 0 | 0 | - | E (UQ) | M4 | mortality/aging; | - |
| ADAMTSL2 | 9719 | ENSG00000197859 | protein_coding | 67 | rs676996 | 0 | 0 | 1 | 0 | 0 | 0 | - | E | - | respiratory system; mortality/aging; limbs/digits/tail; skeleton; muscle; homeostasis/metabolism; | Geleophysic dysplasia 1, 231050 (3), Autosomal recessive |
| DBH | 1621 | ENSG00000123454 | protein_coding | 67 | rs676996 | 0 | 0 | 1 | 0 | 0 | 0 | - | E | - | behavior/neurological; cardiovascular system; mortality/aging; nervous system; vision/eye; growth/size/body region; endocrine/exocrine gland; homeostasis/metabolism; | Orthostatic hypotension 1, due to DBH deficiency, 223360 (3), Autosomal recessive |
| DBH-AS1 | 138948 | ENSG00000225756 | lncRNA | 67 | rs676996 | 0 | 0 | 1 | 0 | 0 | 0 | - | E | M1 | - | - |
| GBGT1 | 26301 | ENSG00000148288 | protein_coding | 67 | rs676996 | 0 | 0 | 1 | 0 | 0 | 0 | - | E | - | endocrine/exocrine gland; behavior/neurological; reproductive system; | - |
| LL09NC01-139C3.1 | - | ENSG00000273473 | lncRNA | 67 | rs676996 | 0 | 0 | 1 | 0 | 0 | 0 | - | E | - | - | - |
| RALGDS | 5900 | ENSG00000160271 | protein_coding | 67 | rs676996 | 0 | 0 | 1 | 0 | 0 | 0 | - | E (UQ) | M7 | neoplasm; homeostasis/metabolism; | - |
| SARDH | 1757 | ENSG00000123453 | protein_coding | 67 | rs676996 | 0 | 0 | 1 | 0 | 0 | 0 | - | E | - | - | [Sarcosinemia], 268900 (3), Autosomal recessive |
| KIAA1217 | 56243 | ENSG00000120549 | protein_coding | 68 | rs6482359 | 1 | 1 | 1 | 0 | 1 | 0 | eQTL (rs6482359, PP=0.797) | E (UQ) | M4 | skeleton; limbs/digits/tail; embryo; | - |
| MSRB2 | 22921 | ENSG00000148450 | protein_coding | 68 | rs6482359 | 0 | 0 | 1 | 0 | 0 | 0 | - | E (UQ) | - | - | - |
| CTBP2 | 1488 | ENSG00000175029 | protein_coding | 69 | rs3012065 | 1 | 1 | 1 | 1 | 1 | 1 | - | E (UQ) | - | growth/size/body region; embryo; nervous system; cardiovascular system; mortality/aging; | - |
| RP11-298J20.4 | - | ENSG00000249456 | lncRNA | 69 | rs3012065 | 0 | 0 | 1 | 0 | 0 | 0 | - | E | - | - | - |
| ZRANB1 | 54764 | ENSG00000019995 | protein_coding | 69 | rs3012065 | 0 | 0 | 1 | 0 | 0 | 0 | - | E (UQ) | M1 | homeostasis/metabolism; mortality/aging; hematopoietic system; immune system; | - |
| RP11-179A10.1 | - | ENSG00000254401 | lncRNA | 70 | rs2218793 | 0 | 0 | 1 | 0 | 0 | 0 | - | E | - | - | - |
| AMPD3 | 272 | ENSG00000133805 | protein_coding | 70:71 | rs2218793:rs4910165 | 0 | 1 | 0 | 1 | 1 | 0 | - | E (UQ) | M4 | homeostasis/metabolism; respiratory system; behavior/neurological; immune system; hematopoietic system; | [AMP deaminase deficiency, erythrocytic], 612874 (3), Autosomal recessive |
| MRVI1-AS1 | 100129827 | ENSG00000177112 | lncRNA | 70:71 | rs2218793:rs4910165 | 0 | 0 | 1 | 0 | 0 | 0 | - | E | - | - | - |
| RNF141 | 50862 | ENSG00000110315 | protein_coding | 70:71 | rs2218793:rs4910165 | 0 | 0 | 1 | 0 | 0 | 0 | - | E (UQ) | M7 | reproductive system; | - |
| RP11-748C4.1 | - | ENSG00000254865 | lncRNA | 70:71 | rs2218793:rs4910165 | 0 | 0 | 1 | 0 | 0 | 0 | - | NE | - | - | - |
| MRVI1 | 10335 | ENSG00000072952 | protein_coding | 70:71 | rs2218793:rs4910165 | 1 | 1 | 1 | 1 | 1 | 1 | - | E (UQ) | M1_hub | - | - |
| RP11-685M7.3 | 101928053 | ENSG00000246308 | lncRNA | 71 | rs4910165 | 0 | 0 | 1 | 0 | 0 | 0 | - | E | - | - | - |
| FNBP4 | 23360 | ENSG00000109920 | protein_coding | 72 | rs10838738 | 0 | 1 | 1 | 1 | 1 | 0 | - | E (UQ) | - | - | - |
| MTCH2 | 23788 | ENSG00000109919 | protein_coding | 72 | rs10838738 | 0 | 1 | 1 | 1 | 1 | 0 | - | E (UQ) | M4 | skeleton; embryo; liver/biliary system; mortality/aging; growth/size/body region; | - |
| NDUFS3 | 4722 | ENSG00000213619 | protein_coding | 72 | rs10838738 | 0 | 1 | 1 | 1 | 1 | 0 | - | E (UQ) | - | hematopoietic system; cardiovascular system; behavior/neurological; homeostasis/metabolism; growth/size/body region; | Mitochondrial complex I deficiency, nuclear type 8, 618230 (3), Autosomal recessive |
| NUP160 | 23279 | ENSG00000030066 | protein_coding | 72 | rs10838738 | 0 | 1 | 1 | 1 | 1 | 0 | - | E (UQ) | - | - | ?Nephrotic syndrome, type 19, 618178 (3), Autosomal recessive |
| PSMC3 | 5702 | ENSG00000165916 | protein_coding | 72 | rs10838738 | 0 | 1 | 1 | 1 | 1 | 0 | - | E (UQ) | - | embryo; mortality/aging; reproductive system; | - |
| SLC39A13 | 91252 | ENSG00000165915 | protein_coding | 72 | rs10838738 | 0 | 1 | 1 | 1 | 1 | 0 | - | E | M1 | cellular; craniofacial; integument; growth/size/body region; skeleton; vision/eye; | Ehlers-Danlos syndrome, spondylodysplastic type, 3, 612350 (3), Autosomal recessive |
| AGBL2 | 79841 | ENSG00000165923 | protein_coding | 72 | rs10838738 | 0 | 1 | 0 | 1 | 1 | 0 | - | E | - | - | - |
| CELF1 | 10658 | ENSG00000149187 | protein_coding | 72 | rs10838738 | 0 | 1 | 0 | 1 | 1 | 0 | - | E (UQ) | M1 | endocrine/exocrine gland; growth/size/body region; embryo; reproductive system; mortality/aging; | - |
| SPI1 | 6688 | ENSG00000066336 | protein_coding | 72 | rs10838738 | 0 | 1 | 0 | 1 | 1 | 0 | - | E | - | growth/size/body region; endocrine/exocrine gland; cellular; vision/eye; craniofacial; mortality/aging; cardiovascular system; hematopoietic system; neoplasm; liver/biliary system; immune system; skeleton; nervous system; | - |
| C1QTNF4 | 114900 | ENSG00000172247 | protein_coding | 72 | rs10838738 | 0 | 1 | 1 | 0 | 1 | 0 | - | E | M1 | - | - |
| KBTBD4 | 55709 | ENSG00000123444 | protein_coding | 72 | rs10838738 | 0 | 1 | 1 | 0 | 1 | 0 | - | E (UQ) | - | - | - |
| MYBPC3 | 4607 | ENSG00000134571 | protein_coding | 72 | rs10838738 | 0 | 1 | 1 | 0 | 1 | 0 | - | E | - | homeostasis/metabolism; cellular; muscle; cardiovascular system; mortality/aging; behavior/neurological; | Cardiomyopathy, hypertrophic, 4, 115197 (3), Autosomal dominant, Autosomal recessive; Cardiomyopathy, dilated, 1MM, 615396 (3), Autosomal dominant; Left ventricular noncompaction 10, 615396 (3), Autosomal dominant |
| PTPMT1 | 114971 | ENSG00000110536 | protein_coding | 72 | rs10838738 | 0 | 1 | 1 | 0 | 1 | 0 | - | E (UQ) | - | immune system; embryo; mortality/aging; growth/size/body region; hematopoietic system; cellular; endocrine/exocrine gland; | - |
| RAPSN | 5913 | ENSG00000165917 | protein_coding | 72 | rs10838738 | 1 | 1 | 1 | 0 | 1 | 1 | - | E | - | muscle; immune system; homeostasis/metabolism; respiratory system; mortality/aging; hematopoietic system; nervous system; | Fetal akinesia deformation sequence 2, 618388 (3), Autosomal recessive; Myasthenic syndrome, congenital, 11, associated with acetylcholine receptor deficiency, 616326 (3), Autosomal recessive |
| FAM180B | 399888 | ENSG00000196666 | protein_coding | 72 | rs10838738 | 0 | 1 | 1 | 0 | 0 | 0 | - | E | - | - | - |
| RP11-750H9.5 | - | ENSG00000255197 | lncRNA | 72 | rs10838738 | 0 | 1 | 1 | 0 | 0 | 0 | - | NE | - | - | - |
| KBTBD4 | 55709 | ENSG00000231880 | lncRNA | 72 | rs10838738 | 0 | 1 | 0 | 0 | 0 | 0 | - | NE | - | - | - |
| MIR4487 | 100616222 | ENSG00000264583 | miRNA | 72 | rs10838738 | 0 | 1 | 0 | 0 | 0 | 0 | - | NE | - | - | - |
| RNA5SP340 | 100873603 | ENSG00000252941 | rRNA_pseudogene | 72 | rs10838738 | 0 | 1 | 0 | 0 | 0 | 0 | - | NE | - | - | - |
| RNU5E-10P | 100873888 | ENSG00000200376 | snRNA | 72 | rs10838738 | 0 | 1 | 0 | 0 | 0 | 0 | - | NE | - | - | - |
| RP11-750H9.7 | - | ENSG00000270072 | lncRNA | 72 | rs10838738 | 0 | 1 | 0 | 0 | 0 | 0 | - | E | - | - | - |
| MADD | 8567 | ENSG00000110514 | protein_coding | 72 | rs10838738 | 0 | 0 | 1 | 0 | 1 | 0 | - | E (UQ) | M4 | homeostasis/metabolism; endocrine/exocrine gland; integument; mortality/aging; nervous system; behavior/neurological; respiratory system; | - |
| ACP2 | 53 | ENSG00000134575 | polymorphic_pseudogene | 72 | rs10838738 | 0 | 0 | 1 | 0 | 0 | 0 | - | E (UQ) | M4 | integument; growth/size/body region; cellular; hematopoietic system; behavior/neurological; immune system; skeleton; renal/urinary system; nervous system; | ?Lysosomal acid phosphatase deficiency, 200950 (1), Autosomal recessive |
| C11orf49 | 79096 | ENSG00000149179 | protein_coding | 72 | rs10838738 | 0 | 0 | 1 | 0 | 0 | 0 | - | E (UQ) | - | - | - |
| DDB2 | 1643 | ENSG00000134574 | protein_coding | 72 | rs10838738 | 0 | 0 | 1 | 0 | 0 | 0 | - | E (UQ) | M7 | neoplasm; mortality/aging; growth/size/body region; cellular; integument; | Xeroderma pigmentosum, group E, DDB-negative subtype, 278740 (3), Autosomal recessive |
| LRP4 | 4038 | ENSG00000134569 | protein_coding | 72 | rs10838738 | 0 | 0 | 1 | 0 | 0 | 0 | - | E | M4 | embryo; behavior/neurological; respiratory system; mortality/aging; reproductive system; nervous system; limbs/digits/tail; skeleton; renal/urinary system; integument; growth/size/body region; endocrine/exocrine gland; craniofacial; homeostasis/metabolism; | ?Myasthenic syndrome, congenital, 17, 616304 (3), Autosomal recessive; Sclerosteosis 2, 614305 (3), Autosomal dominant, Autosomal recessive; Cenani-Lenz syndactyly syndrome, 212780 (3), Autosomal recessive |
| LRP4-AS1 | 100507401 | ENSG00000247675 | lncRNA | 72 | rs10838738 | 0 | 0 | 1 | 0 | 0 | 0 | - | E | - | - | - |

| | | | | | | | | | | | | | | | |
|---|---|---|---|---|---|---|---|---|---|---|---|---|---|---|---|
| NR1H3 | 10062 | ENSG00000025434 | protein_coding | 72 | rs10838738 | 0 | 0 | 1 | 0 | 0 | 0 | - | E | - | liver/biliary system; mortality/aging; reproductive system; hematopoietic system; cardiovascular system; renal/urinary system; cellular; immune system; homeostasis/metabolism; endocrine/exocrine gland; | - |
| PACSIN3 | 29763 | ENSG00000165912 | protein_coding | 72 | rs10838738 | 0 | 0 | 1 | 0 | 0 | 0 | - | E | M7 | hematopoietic system; | - |
| PTPRJ | 5795 | ENSG00000149177 | protein_coding | 72 | rs10838738 | 0 | 0 | 1 | 0 | 0 | 0 | - | E (UQ) | - | homeostasis/metabolism; muscle; growth/size/body region; embryo; immune system; neoplasm; cardiovascular system; hematopoietic system; mortality/aging; reproductive system; | Colon cancer, somatic, 114500 (3) |
| ANO1 | 55107 | ENSG00000131620 | protein_coding | 73 | rs2186797 | 1 | 1 | 1 | 0 | 1 | 1 | coding variant & eQTL (rs2186797, PP=0.970) | E (UQ) | - | homeostasis/metabolism; muscle; growth/size/body region; integument; renal/urinary system; skeleton; digestive/alimentary; nervous system; cardiovascular system; mortality/aging; behavior/neurological; respiratory system; | - |
| FADD | 8772 | ENSG00000168040 | protein_coding | 73 | rs2186797 | 1 | 0 | 1 | 0 | 0 | 0 | eQTL (rs2186797, PP=0.970) | E (UQ) | - | mortality/aging; reproductive system; hematopoietic system; cardiovascular system; embryo; immune system; skeleton; nervous system; digestive/alimentary; growth/size/body region; endocrine/exocrine gland; cellular; muscle; | Infections, recurrent, with encephalopathy, hepatic dysfunction, and cardiovascular malformations, 613759 (3), Autosomal recessive |
| ANO1-AS1 | 100873981 | ENSG00000254902 | lncRNA | 73 | rs2186797 | 0 | 1 | 0 | 0 | 0 | 0 | - | NE | - | - | - |
| RP11-805J14.3 | - | ENSG00000255143 | lncRNA | 73 | rs2186797 | 0 | 1 | 0 | 0 | 0 | 0 | - | NE | - | - | - |
| RP11-805J14.5 | - | ENSG00000254721 | lncRNA | 73 | rs2186797 | 1 | 0 | 1 | 0 | 0 | 0 | eQTL (rs2186797, PP=0.970) | NE | - | - | - |
| NCAM1 | 4684 | ENSG00000149294 | protein_coding | 74 | rs2212450 | 0 | 1 | 1 | 1 | 1 | 0 | - | E | M1 | mortality/aging; behavior/neurological; vision/eye; nervous system; cellular; | - |
| RP11-629G13.1 | 101928847 | ENSG00000247416 | lncRNA | 74 | rs2212450 | 0 | 1 | 1 | 0 | 1 | 0 | - | E | - | - | - |
| RNU7-187P | 106479095 | ENSG00000238998 | snRNA | 74 | rs2212450 | 0 | 1 | 0 | 0 | 0 | 0 | - | NE | - | - | - |
| NCAM1-AS1 | 100288346 | ENSG00000227487 | lncRNA | 74 | rs2212450 | 0 | 0 | 1 | 0 | 0 | 0 | - | NE | - | - | - |
| RP11-839D17.3 | - | ENSG00000255129 | lncRNA | 74 | rs2212450 | 0 | 0 | 1 | 0 | 0 | 0 | - | E | M1 | - | - |
| TIMM8B | 26521 | ENSG00000150779 | protein_coding | 74 | rs2212450 | 0 | 0 | 1 | 0 | 0 | 0 | - | E (UQ) | - | - | - |
| CLMP | 79827 | ENSG00000166250 | protein_coding | 75 | rs4345978 | 1 | 1 | 1 | 1 | 1 | 1 | - | E (UQ) | M1 | digestive/alimentary; renal/urinary system; behavior/neurological; reproductive system; mortality/aging; cardiovascular system; growth/size/body region; muscle; homeostasis/metabolism; | Congenital short bowel syndrome, 615237 (3), Autosomal recessive |
| SNORD14E | 85391 | ENSG00000200879 | snoRNA | 75 | rs4345978 | 0 | 0 | 1 | 0 | 0 | 0 | - | E | - | - | - |
| VWA5A | 4013 | ENSG00000110002 | protein_coding | 75 | rs4345978 | 0 | 0 | 1 | 0 | 0 | 0 | - | E (UQ) | - | integument; behavior/neurological; vision/eye; | - |
| CTC-465D4.1 | - | ENSG00000256499 | lncRNA | 76 | rs11045079 | 0 | 1 | 0 | 0 | 0 | 0 | - | NE | - | - | - |
| PDE3A | 5139 | ENSG00000172572 | protein_coding | 76 | rs11045079 | 0 | 0 | 0 | 0 | 1 | 0 | - | E | - | reproductive system; | Hypertension and brachydactyly syndrome, 112410 (3), Autosomal dominant |
| LINC00477 | 144360 | ENSG00000197503 | lncRNA | 77:78 | rs4579999:rs111235435 | 0 | 1 | 1 | 0 | 1 | 0 | - | E | - | - | - |
| LYRM5 | 144363 | ENSG00000205707 | protein_coding | 78 | rs111235435 | 0 | 0 | 1 | 0 | 0 | 0 | - | E | M1 | - | - |
| HDAC7 | 51564 | ENSG00000061273 | protein_coding | 79 | rs13632 | 0 | 1 | 1 | 1 | 1 | 0 | - | E (UQ) | - | craniofacial; muscle; homeostasis/metabolism; mortality/aging; cardiovascular system; embryo; | - |
| SLC48A1 | 55652 | ENSG00000211584 | protein_coding | 79 | rs13632 | 0 | 1 | 1 | 1 | 1 | 0 | - | E (UQ) | - | - | - |
| RAPGEF3 | 10411 | ENSG00000079337 | protein_coding | 79 | rs13632 | 0 | 1 | 1 | 0 | 1 | 0 | - | E (UQ) | - | endocrine/exocrine gland; adipose tissue; growth/size/body region; homeostasis/metabolism; cellular; behavior/neurological; nervous system; immune system; | - |
| AC004466.1 | - | ENSG00000268069 | lncRNA | 79 | rs13632 | 0 | 1 | 1 | 0 | 0 | 0 | - | E | - | - | - |
| RP1-197B17.3 | 105369748 | ENSG00000257433 | lncRNA | 79 | rs13632 | 0 | 0 | 1 | 0 | 1 | 0 | - | E | - | - | - |
| ENDOU | 8909 | ENSG00000111405 | protein_coding | 79 | rs13632 | 0 | 0 | 0 | 0 | 1 | 0 | - | E (UQ) | M7 | renal/urinary system; growth/size/body region; adipose tissue; | - |
| RPAP3 | 79657 | ENSG00000005175 | protein_coding | 79 | rs13632 | 0 | 0 | 0 | 0 | 1 | 0 | - | E (UQ) | - | mortality/aging; | - |
| PCED1B-AS1 | 100233209 | ENSG00000247774 | lncRNA | 79 | rs13632 | 0 | 0 | 1 | 0 | 0 | 0 | - | E | - | - | - |
| HOTAIR | 100124700 | ENSG00000228630 | lncRNA | 80 | rs920778 | 0 | 1 | 1 | 0 | 1 | 0 | - | E | M1 | limbs/digits/tail; skeleton; | - |
| HOXC11 | 3227 | ENSG00000123388 | protein_coding | 80 | rs920778 | 1 | 1 | 0 | 0 | 1 | 1 | - | E | - | - | - |
| HOXC-AS3 | 100874365 | ENSG00000251151 | lncRNA | 80 | rs920778 | 0 | 1 | 0 | 0 | 0 | 0 | - | E | - | - | - |
| HOXC10 | 3226 | ENSG00000180818 | protein_coding | 80 | rs920778 | 0 | 1 | 0 | 0 | 0 | 0 | - | E | M1 | growth/size/body region; muscle; behavior/neurological; renal/urinary system; skeleton; nervous system; limbs/digits/tail; | - |
| HOXC12 | 3228 | ENSG00000123407 | protein_coding | 80 | rs920778 | 0 | 1 | 0 | 0 | 0 | 0 | - | E | - | limbs/digits/tail; | - |
| HOXC6 | 3223 | ENSG00000197757 | protein_coding | 80 | rs920778 | 0 | 1 | 0 | 0 | 0 | 0 | - | E | - | endocrine/exocrine gland; integument; skeleton; | - |
| MIR196A2 | 406973 | ENSG00000207924 | miRNA | 80 | rs920778 | 0 | 1 | 0 | 0 | 0 | 0 | - | NE | - | skeleton; | - |
| RP11-834C11.12 | - | ENSG00000273049 | protein_coding | 80 | rs920778 | 0 | 1 | 0 | 0 | 0 | 0 | - | NE | - | - | - |
| CBX5 | 23468 | ENSG00000094916 | protein_coding | 80 | rs920778 | 0 | 0 | 1 | 0 | 0 | 0 | - | E (UQ) | M1 | growth/size/body region; mortality/aging; hematopoietic system; behavior/neurological; skeleton; immune system; | - |
| HOXC4 | 3221 | ENSG00000198353 | protein_coding | 80 | rs920778 | 0 | 0 | 1 | 0 | 0 | 0 | - | E | - | muscle; hematopoietic system; mortality/aging; behavior/neurological; immune system; skeleton; digestive/alimentary; | - |
| RP11-834C11.11 | - | ENSG00000260470 | lncRNA | 80 | rs920778 | 0 | 0 | 1 | 0 | 0 | 0 | - | E | - | - | - |
| RP11-834C11.6 | - | ENSG00000249388 | lncRNA | 80 | rs920778 | 0 | 0 | 1 | 0 | 0 | 0 | - | NE | - | - | - |
| HMGA2 | 8091 | ENSG00000149948 | protein_coding | 81 | rs11176001 | 1 | 1 | 1 | 1 | 1 | 1 | - | E | - | cellular; homeostasis/metabolism; craniofacial; adipose tissue; endocrine/exocrine gland; integument; growth/size/body region; nervous system; hearing/vestibular/ear; limbs/digits/tail; reproductive system; behavior/neurological; | Leiomyoma, uterine, somatic, 150699 (1) |
| RP11-366L20.3 | - | ENSG00000256083 | lncRNA | 81 | rs11176001 | 0 | 1 | 0 | 0 | 0 | 0 | - | NE | - | - | - |
| RP11-366L20.4 | 105369810 | ENSG00000256259 | lncRNA | 81 | rs11176001 | 0 | 1 | 0 | 0 | 0 | 0 | - | NE | - | - | - |
| HELB | 92797 | ENSG00000127311 | protein_coding | 81 | rs11176001 | 0 | 0 | 1 | 0 | 0 | 0 | - | E | - | cellular; homeostasis/metabolism; | - |
| RP11-139B1.1 | 105369996 | ENSG00000257476 | lncRNA | 82 | rs2555004 | 0 | 1 | 0 | 0 | 0 | 0 | - | NE | - | - | - |
| TBX5 | 6910 | ENSG00000089225 | protein_coding | 82 | rs2555004 | 0 | 0 | 1 | 0 | 1 | 0 | - | E | M1 | growth/size/body region; muscle; homeostasis/metabolism; skeleton; embryo; cardiovascular system; mortality/aging; limbs/digits/tail; | Holt-Oram syndrome, 142900 (3), Autosomal dominant |
| DLEU1 | 10301 | ENSG00000176124 | lncRNA | 83 | rs7994724 | 0 | 1 | 0 | 1 | 1 | 0 | - | E | - | - | - |
| DLEU7 | 220107 | ENSG00000186047 | protein_coding | 83 | rs7994724 | 0 | 1 | 1 | 1 | 0 | 0 | - | E | - | cardiovascular system; behavior/neurological; | - |
| RNASEH2B | 79621 | ENSG00000136104 | protein_coding | 83 | rs7994724 | 1 | 1 | 1 | 1 | 0 | 0 | eQTL (rs7994724, PP=0.621) | E (UQ) | - | immune system; embryo; liver/biliary system; hematopoietic system; cardiovascular system; mortality/aging; endocrine/exocrine gland; homeostasis/metabolism; cellular; growth/size/body region; | Aicardi-Goutieres syndrome 2, 610181 (3), Autosomal recessive |
| RNASEH2B-AS1 | 100874255 | ENSG00000233672 | lncRNA | 83 | rs7994724 | 1 | 1 | 1 | 0 | 0 | 0 | eQTL (rs7994724, PP=0.621) | E | - | - | - |
| RNA5SP28 | 100873360 | ENSG00000200711 | rRNA_pseudogene | 83 | rs7994724 | 0 | 1 | 0 | 0 | 0 | 0 | - | NE | - | - | - |
| INTS6-AS1 | 100507398 | ENSG00000236778 | lncRNA | 83 | rs7994724 | 0 | 0 | 1 | 0 | 0 | 0 | - | E | - | - | - |
| ARHGAP11A | 9824 | ENSG00000198826 | protein_coding | 84 | rs11635984 | 1 | 0 | 1 | 0 | 0 | 0 | eQTL (rs11635984, PP=0.619) | E | - | - | - |
| GREM1 | 26585 | ENSG00000166923 | protein_coding | 84 | rs11635984 | 0 | 1 | 1 | 1 | 1 | 0 | - | E (UQ) | M1 | mortality/aging; embryo; respiratory system; renal/urinary system; skeleton; limbs/digits/tail; homeostasis/metabolism; cellular; | - |
| SCG5 | 6447 | ENSG00000166922 | protein_coding | 84 | rs11635984 | 0 | 1 | 0 | 1 | 1 | 0 | - | E | - | respiratory system; liver/biliary system; behavior/neurological; reproductive system; mortality/aging; cardiovascular system; hematopoietic system; nervous system; immune system; integument; growth/size/body region; endocrine/exocrine gland; adipose tissue; cellular; homeostasis/metabolism; | - |
| RP11-758N13.1 | 100131315 | ENSG00000259721 | lncRNA | 84 | rs11635984 | 1 | 1 | 1 | 0 | 0 | 0 | eQTL (rs11635984, PP=0.619) | E | M1 | - | - |
| SMAD3 | 4088 | ENSG00000166949 | protein_coding | 85:86 | rs17293632:rs12594232 | 1 | 1 | 1 | 1 | 1 | 1 | - | E (UQ) | M7 | growth/size/body region; integument; endocrine/exocrine gland; homeostasis/metabolism; cellular; cardiovascular system; hematopoietic system; mortality/aging; reproductive system; neoplasm; behavior/neurological; immune system; renal/urinary system; skeleton; digestive/alimentary; limbs/digits/tail; | Loeys-Dietz syndrome 3, 613795 (3), Autosomal dominant |
| RP11-342M21.2 | - | ENSG00000259202 | lncRNA | 85:86 | rs17293632:rs12594232 | 0 | 1 | 0 | 1 | 0 | 0 | - | NE | - | - | - |
| AAGAB | 79719 | ENSG00000103591 | protein_coding | 85:86 | rs17293632:rs12594232 | 0 | 0 | 1 | 0 | 0 | 0 | - | E (UQ) | M4 | - | Keratoderma, palmoplantar, punctate type IA, 148600 (3), Autosomal dominant |
| CTD-3185P2.1 | - | ENSG00000261351 | lncRNA | 85:86 | rs17293632:rs12594232 | 0 | 0 | 1 | 0 | 0 | 0 | - | E | - | - | - |
| IQCH | 64799 | ENSG00000103599 | protein_coding | 85:86 | rs17293632:rs12594232 | 0 | 0 | 1 | 0 | 0 | 0 | - | E | M4 | - | - |
| MAP2K5 | 5607 | ENSG00000137764 | protein_coding | 85:86 | rs17293632:rs12594232 | 0 | 0 | 1 | 0 | 0 | 0 | - | E | - | embryo; cardiovascular system; mortality/aging; growth/size/body region; muscle; cellular; | - |
| RP11-502I4.3 | - | ENSG00000270964 | lncRNA | 85:86 | rs17293632:rs12594232 | 0 | 0 | 1 | 0 | 0 | 0 | - | E | - | - | - |
| SKOR1 | 390598 | ENSG00000188779 | protein_coding | 85:86 | rs17293632:rs12594232 | 0 | 0 | 1 | 0 | 0 | 0 | - | E | M4 | - | - |
| PIAS1 | 8554 | ENSG00000033800 | protein_coding | 85:86 | rs17293632:rs12594232 | 0 | 1 | 1 | 1 | 1 | 0 | - | E (UQ) | - | growth/size/body region; mortality/aging; immune system; | - |
| CALML4 | 91860 | ENSG00000129007 | protein_coding | 86 | rs12594232 | 1 | 1 | 1 | 1 | 1 | 0 | - | E+ (UQ) | - | - | - |
| CLN6 | 54982 | ENSG00000128973 | protein_coding | 86 | rs12594232 | 0 | 1 | 1 | 1 | 1 | 0 | - | E (UQ) | - | hematopoietic system; mortality/aging; behavior/neurological; nervous system; vision/eye; adipose tissue; | Ceroid lipofuscinosis, neuronal, Kufs type, adult onset, 204300 (3), Autosomal recessive; Ceroid lipofuscinosis, neuronal, 6, 601780 (3), Autosomal recessive |
| RP11-315D16.2 | - | ENSG00000260007 | protein_coding | 86 | rs12594232 | 0 | 1 | 1 | 1 | 0 | 0 | - | E | - | - | - |
| RP11-315D16.4 | - | ENSG00000260657 | lncRNA | 86 | rs12594232 | 0 | 0 | 1 | 0 | 0 | 0 | - | NE | - | - | - |
| LINC00924 | 145820 | ENSG00000259134 | lncRNA | 87 | rs7183672 | 0 | 0 | 0 | 0 | 1 | 0 | - | E | M1 | - | - |
| MYH11 | 4629 | ENSG00000133392 | protein_coding | 88 | rs6498573 | 1 | 1 | 1 | 1 | 1 | 1 | eQTL (rs6498573, PP=0.632) | E+ (UQ) | M1_hub | growth/size/body region; integument; homeostasis/metabolism; cellular; muscle; digestive/alimentary; cardiovascular system; mortality/aging; respiratory system; renal/urinary system; | Aortic aneurysm, familial thoracic 4, 132900 (3), Autosomal dominant |
| NDE1 | 54820 | ENSG00000072864 | protein_coding | 88 | rs6498573 | 1 | 0 | 1 | 0 | 1 | 0 | eQTL (rs6498573, PP=0.632) | E | - | cellular; nervous system; | Lissencephaly 4 (with microcephaly), 614019 (3), Autosomal recessive; ?Microhydranencephaly, 605013 (3), Autosomal recessive |
| NTAN1 | 123803 | ENSG00000157045 | protein_coding | 88 | rs6498573 | 1 | 0 | 1 | 0 | 0 | 0 | eQTL (rs6498573, PP=0.632) | E (UQ) | M1 | cellular; behavior/neurological; hematopoietic system; mortality/aging; immune system; | - |
| AF001548.5 | - | ENSG00000263335 | lncRNA | 88 | rs6498573 | 1 | 0 | 1 | 0 | 0 | 0 | eQTL (rs6498573, PP=0.632) | E | - | - | - |
| AF001548.6 | - | ENSG00000263065 | lncRNA | 88 | rs6498573 | 1 | 0 | 1 | 0 | 0 | 0 | eQTL (rs6498573, PP=0.632) | NE | - | - | - |
| RP11-1186N24.5 | 105369154 | ENSG00000270580 | lncRNA | 88 | rs6498573 | 0 | 0 | 1 | 0 | 0 | 0 | - | E | - | - | - |
| RP11-72I8.1 | 100505915 | ENSG00000260735 | nscribed_processed_pseudoge | 88 | rs6498573 | 0 | 0 | 1 | 0 | 0 | 0 | - | E | - | - | - |
| FOXL1 | 2300 | ENSG00000176678 | protein_coding | 89 | rs4843407 | 1 | 1 | 0 | 0 | 1 | 1 | - | E | - | cellular; growth/size/body region; endocrine/exocrine gland; digestive/alimentary; mortality/aging; | - |
| MTHFSD | 64779 | ENSG00000103248 | protein_coding | 89 | rs4843407 | 0 | 0 | 1 | 0 | 0 | 0 | - | E | - | - | - |
| RP11-463O9.9 | - | ENSG00000270020 | lncRNA | 89 | rs4843407 | 0 | 0 | 1 | 0 | 0 | 0 | - | E | - | - | - |
| POLR2A | 5430 | ENSG00000181222 | protein_coding | 90 | rs78378222 | 0 | 1 | 0 | 0 | 0 | 0 | - | E (UQ) | M7 | mortality/aging; growth/size/body region; | Neurodevelopmental disorder with hypotonia and variable intellectual and behavioral abnormalities, 618603 (3), Autosomal dominant |
| SENP3 | 26168 | ENSG00000161956 | protein_coding | 90 | rs78378222 | 0 | 1 | 0 | 0 | 0 | 0 | - | E (UQ) | M7 | behavior/neurological; homeostasis/metabolism; | - |
| TNFSF12 | 8742 | ENSG00000239697 | protein_coding | 90 | rs78378222 | 0 | 1 | 0 | 0 | 0 | 0 | - | E | M1 | cellular; liver/biliary system; immune system; hematopoietic system; | - |
| TNFSF12-TNFSF13 | 407977 | ENSG00000248871 | protein_coding | 90 | rs78378222 | 0 | 1 | 0 | 0 | 0 | 0 | - | E | M4 | - | - |
| TNFSF13 | 8741 | ENSG00000161955 | protein_coding | 90 | rs78378222 | 0 | 1 | 0 | 0 | 0 | 0 | - | E (UQ) | M4 | hematopoietic system; skeleton; immune system; cellular; | - |
| TP53 | 7157 | ENSG00000141510 | protein_coding | 90 | rs78378222 | 0 | 1 | 0 | 0 | 0 | 0 | - | E (UQ) | - | behavior/neurological; liver/biliary system; respiratory system; embryo; neoplasm; pigmentation; mortality/aging; reproductive system; hematopoietic system; cardiovascular system; vision/eye; limbs/digits/tail; hearing/vestibular/ear; nervous system; digestive/alimentary; renal/urinary system; skeleton; cellular; immune system; homeostasis/metabolism; adipose tissue; endocrine/exocrine gland; integument; growth/size/body region; craniofacial; muscle; | {Adrenocortical carcinoma, pediatric}, 202300 (3), Autosomal dominant; {Glioma susceptibility 1}, 137800 (3), Autosomal dominant, Somatic mutation; {Basal cell carcinoma 7}, 614740 (3), Autosomal dominant; Bone marrow failure syndrome 5, 618165 (3), Autosomal dominant; {Colorectal cancer}, 114500 (3), Autosomal dominant, Somatic mutation; Nasopharyngeal carcinoma, somatic, 607107 (3); Breast cancer, somatic, 114480 (3); {Osteosarcoma}, 259500 (3), Somatic mutation; {Choroid plexus papilloma}, 260500 (3), Autosomal dominant; Li-Fraumeni syndrome, 151623 (3), Autosomal dominant; Hepatocellular carcinoma, somatic, 114550 (3); Pancreatic cancer, somatic, 260350 (3) |

| | | | | | | | | | | | | | | | | |
|---|---|---|---|---|---|---|---|---|---|---|---|---|---|---|---|---|
| AC027763.2 | 100506713 | ENSG00000215067 | lncRNA | 90 | rs78378222 | 0 | 0 | 1 | 0 | 0 | 0 | - | E | - | - | - |
| MYOCD | 93649 | ENSG00000141052 | protein_coding | 91 | rs62061554 | 1 | 0 | 0 | 0 | 1 | 1 | - | E | M1 | muscle; homeostasis/metabolism; cellular; growth/size/body region; embryo; cardiovascular system; mortality/aging; | Megabladder, congenital, 618719 (3), Autosomal dominant |
| ATPAF2 | 91647 | ENSG00000171953 | protein_coding | 92 | rs854786 | 0 | 1 | 1 | 1 | 1 | 0 | - | E | M4 | mortality/aging; skeleton; growth/size/body region; craniofacial; | ?Mitochondrial complex V (ATP synthase) deficiency, nuclear type 1, 604273 (3), Autosomal recessive |
| DRG2 | 1819 | ENSG00000108591 | protein_coding | 92 | rs854786 | 0 | 1 | 1 | 1 | 1 | 0 | - | E (UQ) | - | behavior/neurological; skeleton; limbs/digits/tail; growth/size/body region; homeostasis/metabolism; | - |
| GID4 | 79018 | ENSG00000141034 | protein_coding | 92 | rs854786 | 0 | 1 | 1 | 1 | 1 | 0 | - | E | - | - | - |
| LRRC48 | 83450 | ENSG00000171962 | protein_coding | 92 | rs854786 | 0 | 1 | 1 | 1 | 1 | 0 | - | E | - | - | - |
| MYO15A | 51168 | ENSG00000091536 | protein_coding | 92 | rs854786 | 0 | 1 | 1 | 1 | 1 | 0 | - | E | - | vision/eye; hearing/vestibular/ear; nervous system; behavior/neurological; hematopoietic system; adipose tissue; | Deafness, autosomal recessive 3, 600316 (3), Autosomal recessive |
| TOM1L2 | 146691 | ENSG00000175662 | protein_coding | 92 | rs854786 | 0 | 1 | 1 | 1 | 1 | 0 | - | E (UQ) | M7 | vision/eye; nervous system; renal/urinary system; skeleton; immune system; respiratory system; behavior/neurological; neoplasm; reproductive system; integument; hematopoietic system; cardiovascular system; growth/size/body region; craniofacial; endocrine/exocrine gland; adipose tissue; | - |
| ALKBH5 | 54890 | ENSG00000091542 | protein_coding | 92 | rs854786 | 0 | 0 | 1 | 1 | 0 | 0 | - | E (UQ) | - | reproductive system; cellular; endocrine/exocrine gland; | - |
| SREBF1 | 6720 | ENSG00000072310 | protein_coding | 92 | rs854786 | 0 | 0 | 1 | 1 | 0 | 0 | - | E (UQ) | - | liver/biliary system; mortality/aging; homeostasis/metabolism; | - |
| RAI1 | 10743 | ENSG00000108557 | protein_coding | 92 | rs854786 | 0 | 0 | 0 | 1 | 0 | 0 | - | E | - | homeostasis/metabolism; craniofacial; adipose tissue; endocrine/exocrine gland; growth/size/body region; skeleton; limbs/digits/tail; nervous system; mortality/aging; behavior/neurological; respiratory system; | Smith-Magenis syndrome, 182290 (3), Isolated cases, Autosomal dominant |
| COPS3 | 8533 | ENSG00000141030 | protein_coding | 92 | rs854786 | 0 | 0 | 1 | 0 | 0 | 0 | - | E (UQ) | - | mortality/aging; embryo; growth/size/body region; | - |
| LLGL1 | 3996 | ENSG00000131899 | protein_coding | 92 | rs854786 | 0 | 0 | 1 | 0 | 0 | 0 | - | E | M7 | mortality/aging; cardiovascular system; embryo; skeleton; nervous system; craniofacial; cellular; | - |
| MIEF2 | 125170 | ENSG00000177427 | protein_coding | 92 | rs854786 | 0 | 0 | 1 | 0 | 0 | 0 | - | E | - | - | - |
| PEMT | 10400 | ENSG00000133027 | protein_coding | 92 | rs854786 | 0 | 0 | 1 | 0 | 0 | 0 | - | E (UQ) | - | homeostasis/metabolism; liver/biliary system; | - |
| RP11-258F1.1 | - | ENSG00000266677 | lncRNA | 92 | rs854786 | 0 | 0 | 1 | 0 | 0 | 0 | - | E | - | - | - |
| SMCR5 | 140771 | ENSG00000226746 | lncRNA | 92 | rs854786 | 0 | 0 | 1 | 0 | 0 | 0 | - | E | - | - | - |
| SMCR8 | 140775 | ENSG00000176994 | protein_coding | 92 | rs854786 | 0 | 0 | 1 | 0 | 0 | 0 | - | E | - | cellular; immune system; homeostasis/metabolism; hematopoietic system; digestive/alimentary; | - |
| TOP3A | 7156 | ENSG00000177302 | protein_coding | 92 | rs854786 | 0 | 0 | 1 | 0 | 0 | 0 | - | E | - | cellular; mortality/aging; embryo; | Microcephaly, growth restriction, and increased sister chromatid exchange 2, 618097 (3), Autosomal recessive; ?Progressive external ophthalmoplegia with mitochondrial DNA deletions, autosomal recessive 5, 618098 (3), Autosomal recessive |
| TVP23B | 51030 | ENSG00000171928 | protein_coding | 92 | rs854786 | 0 | 0 | 1 | 0 | 0 | 0 | - | E (UQ) | - | - | - |
| EVI2A | 2123 | ENSG00000126860 | protein_coding | 93 | rs2525570 | 0 | 1 | 1 | 1 | 1 | 0 | - | E | - | - | - |
| EVI2B | 2124 | ENSG00000185862 | protein_coding | 93 | rs2525570 | 0 | 1 | 1 | 1 | 1 | 0 | - | E (UQ) | - | endocrine/exocrine gland; reproductive system; cardiovascular system; renal/urinary system; | - |
| NF1 | 4763 | ENSG00000196712 | protein_coding | 93 | rs2525570 | 0 | 1 | 1 | 1 | 1 | 0 | - | E (UQ) | - | growth/size/body region; integument; endocrine/exocrine gland; muscle; craniofacial; homeostasis/metabolism; cellular; skeleton; renal/urinary system; embryo; behavior/neurological; liver/biliary system; respiratory system; cardiovascular system; hematopoietic system; mortality/aging; reproductive system; pigmentation; neoplasm; digestive/alimentary; limbs/digits/tail; hearing/vestibular/ear; nervous system; vision/eye; immune system; | Neurofibromatosis-Noonan syndrome, 601321 (3), Autosomal dominant; Leukemia, juvenile myelomonocytic, 607785 (3), Autosomal dominant, Somatic mutation; Neurofibromatosis, familial spinal, 162210 (3), Autosomal dominant; Watson syndrome, 193520 (3), Autosomal dominant; Neurofibromatosis, type 1, 162200 (3), Autosomal dominant |
| OMG | 4974 | ENSG00000126861 | protein_coding | 93 | rs2525570 | 0 | 1 | 1 | 1 | 1 | 0 | - | E | - | homeostasis/metabolism; | - |
| RAB11FIP4 | 84440 | ENSG00000131242 | protein_coding | 93 | rs2525570 | 1 | 1 | 1 | 1 | 1 | 0 | - | E (UQ) | M4_hub | behavior/neurological; | - |
| CTD-2370N5.3 | - | ENSG00000265118 | protein_coding | 93 | rs2525570 | 0 | 1 | 1 | 1 | 0 | 0 | - | E | - | - | - |
| RP11-142O6.1 | - | ENSG00000266371 | lncRNA | 93 | rs2525570 | 0 | 1 | 1 | 0 | 0 | 0 | - | E | - | - | - |
| RP11-848P1.5 | - | ENSG00000264107 | lncRNA | 93 | rs2525570 | 0 | 1 | 1 | 0 | 0 | 0 | - | E | M7 | - | - |
| MIR4733 | 100616266 | ENSG00000265444 | miRNA | 93 | rs2525570 | 0 | 1 | 0 | 0 | 0 | 0 | - | NE | - | - | - |
| AC006050.2 | - | ENSG00000231421 | lncRNA | 93 | rs2525570 | 0 | 0 | 1 | 0 | 0 | 0 | - | NE | - | - | - |
| CTD-2349P21.10 | - | ENSG00000265791 | lncRNA | 93 | rs2525570 | 0 | 0 | 1 | 0 | 0 | 0 | - | NE | - | - | - |
| PRKCA | 5578 | ENSG00000154229 | protein_coding | 94 | rs4423457 | 0 | 1 | 1 | 0 | 0 | 0 | - | E (UQ) | - | cellular; homeostasis/metabolism; muscle; adipose tissue; integument; neoplasm; cardiovascular system; | Pituitary tumor, invasive (3) |
| APOH | 350 | ENSG00000091583 | protein_coding | 94 | rs4423457 | 0 | 1 | 0 | 0 | 0 | 0 | - | E | - | mortality/aging; homeostasis/metabolism; | - |
| CACNG5 | 27091 | ENSG00000075429 | protein_coding | 94 | rs4423457 | 0 | 0 | 1 | 0 | 0 | 0 | - | NE | - | - | - |
| KANK2 | 25959 | ENSG00000197256 | protein_coding | 95 | rs2421206 | 1 | 1 | 1 | 0 | 1 | 1 | eQTL (rs2421206, PP=0.609) | E (UQ) | M1_hub | - | Palmoplantar keratoderma and woolly hair, 616099 (3), Autosomal recessive; Nephrotic syndrome, type 16, 617783 (3), Autosomal recessive |
| SPC24 | 147841 | ENSG00000161888 | protein_coding | 95 | rs2421206 | 0 | 1 | 1 | 1 | 0 | 0 | - | E | - | - | - |
| ZNF404 | 342908 | ENSG00000176222 | protein_coding | 96 | rs8106090 | 0 | 1 | 1 | 1 | 1 | 0 | - | E | M1 | - | - |
| ZNF45 | 7596 | ENSG00000124459 | protein_coding | 96 | rs8106090 | 0 | 1 | 1 | 1 | 1 | 0 | - | E | - | - | - |
| ZNF283 | 284349 | ENSG00000167637 | protein_coding | 96 | rs8106090 | 0 | 1 | 1 | 0 | 1 | 0 | - | E | - | - | - |
| RP11-15A1.2 | - | ENSG00000267191 | lncRNA | 96 | rs8106090 | 0 | 1 | 1 | 0 | 0 | 0 | - | NE | - | - | - |
| RP11-15A1.3 | 100505715 | ENSG00000267058 | lncRNA | 96 | rs8106090 | 0 | 1 | 1 | 0 | 0 | 0 | - | E | - | - | - |
| LYPD5 | 284348 | ENSG00000159871 | protein_coding | 96 | rs8106090 | 0 | 0 | 1 | 0 | 1 | 0 | - | E (UQ) | - | - | - |
| AC084219.4 | 100379224 | ENSG00000186019 | lncRNA | 96 | rs8106090 | 0 | 0 | 1 | 0 | 0 | 0 | - | E | - | - | - |
| AC115522.3 | - | ENSG00000268601 | lncRNA | 96 | rs8106090 | 0 | 0 | 1 | 0 | 0 | 0 | - | E | M7 | - | - |
| IRGC | 56269 | ENSG00000124449 | protein_coding | 96 | rs8106090 | 0 | 0 | 1 | 0 | 0 | 0 | - | NE | - | - | - |
| KCNN4 | 3783 | ENSG00000104783 | protein_coding | 96 | rs8106090 | 0 | 0 | 1 | 0 | 0 | 0 | - | E (UQ) | - | hematopoietic system; cardiovascular system; immune system; renal/urinary system; digestive/alimentary; endocrine/exocrine gland; homeostasis/metabolism; muscle; | Dehydrated hereditary stomatocytosis 2, 616689 (3), Autosomal dominant |
| RP11-15A1.7 | 101928063 | ENSG00000266921 | lncRNA | 96 | rs8106090 | 0 | 0 | 1 | 0 | 0 | 0 | - | E | M1 | - | - |
| SMG9 | 56006 | ENSG00000105771 | protein_coding | 96 | rs8106090 | 0 | 0 | 1 | 0 | 0 | 0 | - | E (UQ) | - | vision/eye; limbs/digits/tail; hearing/vestibular/ear; nervous system; skeleton; renal/urinary system; immune system; liver/biliary system; respiratory system; embryo; cardiovascular system; mortality/aging; craniofacial; muscle; homeostasis/metabolism; endocrine/exocrine gland; growth/size/body region; | Heart and brain malformation syndrome, 616920 (3), Autosomal recessive |
| ZNF112 | 7771 | ENSG00000062370 | protein_coding | 96 | rs8106090 | 0 | 0 | 1 | 0 | 0 | 0 | - | E | - | - | - |
| ZNF155 | 7711 | ENSG00000204920 | protein_coding | 96 | rs8106090 | 0 | 0 | 1 | 0 | 0 | 0 | - | E | - | - | - |
| ZNF221 | 7638 | ENSG00000159905 | protein_coding | 96 | rs8106090 | 0 | 0 | 1 | 0 | 0 | 0 | - | E | - | - | - |
| ZNF223 | 7766 | ENSG00000267022 | protein_coding | 96 | rs8106090 | 0 | 0 | 1 | 0 | 0 | 0 | - | E | M1 | - | - |
| ZNF224 | 7767 | ENSG00000267680 | protein_coding | 96 | rs8106090 | 0 | 0 | 1 | 0 | 0 | 0 | - | E | - | - | - |
| ZNF230 | 7773 | ENSG00000159882 | protein_coding | 96 | rs8106090 | 0 | 0 | 1 | 0 | 0 | 0 | - | E | - | - | - |
| ZNF284 | 342909 | ENSG00000186026 | protein_coding | 96 | rs8106090 | 0 | 0 | 1 | 0 | 0 | 0 | - | E | M1 | - | - |
| RBM38 | 55544 | ENSG00000132819 | protein_coding | 97 | rs34161672 | 1 | 0 | 1 | 0 | 1 | 0 | eQTL (rs34161672, PP=0.660) | E | - | skeleton; immune system; neoplasm; mortality/aging; hematopoietic system; liver/biliary system; homeostasis/metabolism; adipose tissue; integument; growth/size/body region; | - |
| RAB22A | 57403 | ENSG00000124209 | protein_coding | 97 | rs34161672 | 0 | 0 | 1 | 0 | 0 | 0 | - | E (UQ) | - | - | - |
| RP5-1043L13.1 | 284757 | ENSG00000228340 | lncRNA | 98 | rs35384758 | 0 | 1 | 0 | 0 | 1 | 0 | - | E | - | - | - |
| MIR4533 | 100616362 | ENSG00000266140 | miRNA | 98 | rs35384758 | 0 | 1 | 0 | 0 | 0 | 0 | - | NE | - | - | - |
| BACH1 | 571 | ENSG00000156273 | protein_coding | 99 | rs2832279 | 0 | 1 | 1 | 1 | 1 | 0 | - | E (UQ) | - | behavior/neurological; cardiovascular system; mortality/aging; muscle; homeostasis/metabolism; cellular; | - |
| MAP3K7CL | 56911 | ENSG00000156265 | protein_coding | 99 | rs2832279 | 0 | 1 | 1 | 1 | 1 | 0 | - | E | M1 | - | - |
| BACH1-IT1 | - | ENSG00000248476 | lncRNA | 99 | rs2832279 | 0 | 1 | 1 | 0 | 1 | 0 | - | E | M1 | - | - |
| CCT8 | 10694 | ENSG00000156261 | protein_coding | 99 | rs2832279 | 0 | 1 | 1 | 0 | 1 | 0 | - | E (UQ) | - | - | - |
| LINC00189 | 193629 | ENSG00000215533 | lncRNA | 99 | rs2832279 | 0 | 1 | 1 | 0 | 1 | 0 | - | E | - | - | - |
| RWDD2B | 10069 | ENSG00000156253 | protein_coding | 99 | rs2832279 | 0 | 1 | 1 | 0 | 1 | 0 | - | E (UQ) | - | - | - |
| USP16 | 10600 | ENSG00000156256 | protein_coding | 99 | rs2832279 | 0 | 1 | 0 | 0 | 1 | 0 | - | E (UQ) | - | mortality/aging; | - |
| AF124730.4 | - | ENSG00000224649 | lncRNA | 99 | rs2832279 | 0 | 1 | 1 | 0 | 0 | 0 | - | NE | - | - | - |
| BACH1-AS1 | 106478962 | ENSG00000232118 | lncRNA | 99 | rs2832279 | 0 | 1 | 1 | 0 | 0 | 0 | - | E | - | - | - |
| BACH1-IT2 | 100874322 | ENSG00000228817 | lncRNA | 99 | rs2832279 | 0 | 1 | 1 | 0 | 0 | 0 | - | E | M7 | - | - |
| LTN1 | 26046 | ENSG00000198862 | protein_coding | 99 | rs2832279 | 0 | 1 | 1 | 0 | 0 | 0 | - | E | - | nervous system; mortality/aging; behavior/neurological; muscle; growth/size/body region; | - |
| U3 | - | ENSG00000212479 | snoRNA | 99 | rs2832279 | 0 | 1 | 1 | 0 | 0 | 0 | - | NE | - | - | - |
| AF129075.5 | - | ENSG00000231125 | lncRNA | 99 | rs2832279 | 0 | 1 | 0 | 0 | 0 | 0 | - | NE | - | - | - |
| AP000240.9 | - | ENSG00000273017 | lncRNA | 99 | rs2832279 | 0 | 1 | 0 | 0 | 0 | 0 | - | E | - | - | - |
| RP1-100J12.1 | - | ENSG00000273254 | lncRNA | 99 | rs2832279 | 0 | 1 | 0 | 0 | 0 | 0 | - | E | M7 | - | - |
| AF131217.1 | - | ENSG00000232855 | lncRNA | 99 | rs2832279 | 0 | 0 | 1 | 0 | 0 | 0 | - | E | M1 | - | - |
| AL035610.2 | 101927973 | ENSG00000236532 | lncRNA | 99 | rs2832279 | 0 | 0 | 1 | 0 | 0 | 0 | - | E | - | - | - |
| GRIK1 | 2897 | ENSG00000171189 | protein_coding | 99 | rs2832279 | 0 | 0 | 1 | 0 | 0 | 0 | - | E | M1 | adipose tissue; integument; growth/size/body region; homeostasis/metabolism; behavior/neurological; hematopoietic system; nervous system; | - |
| GRIK1-AS1 | 642976 | ENSG00000174680 | lncRNA | 99 | rs2832279 | 0 | 0 | 1 | 0 | 0 | 0 | - | E | M1 | - | - |
| LINC00161 | 118421 | ENSG00000226935 | lncRNA | 99 | rs2832279 | 0 | 0 | 1 | 0 | 0 | 0 | - | E | - | - | - |
| N6AMT1 | 29104 | ENSG00000156239 | protein_coding | 99 | rs2832279 | 0 | 0 | 1 | 0 | 0 | 0 | - | E | - | growth/size/body region; embryo; mortality/aging; | - |
| AP1B1 | 162 | ENSG00000100280 | protein_coding | 100 | rs174767 | 0 | 1 | 1 | 1 | 1 | 0 | - | E (UQ) | - | growth/size/body region; cellular; skeleton; respiratory system; cardiovascular system; hematopoietic system; immune system; | Keratitis-ichthyosis-deafness syndrome, autosomal recessive, 242150 (3), Autosomal recessive |
| GAS2L1 | 10634 | ENSG00000185340 | protein_coding | 100 | rs174767 | 0 | 1 | 1 | 1 | 1 | 0 | - | E | - | reproductive system; | - |
| EWSR1 | 2130 | ENSG00000182944 | protein_coding | 100 | rs174767 | 0 | 1 | 0 | 1 | 1 | 0 | - | E (UQ) | - | endocrine/exocrine gland; growth/size/body region; immune system; homeostasis/metabolism; behavior/neurological; neoplasm; hematopoietic system; mortality/aging; limbs/digits/tail; skeleton; | Neuroepithelioma, 612219 (3); Ewing sarcoma, 612219 (3) |
| RASL10A | 10633 | ENSG00000100276 | protein_coding | 100 | rs174767 | 0 | 1 | 1 | 0 | 1 | 0 | - | E | M1 | - | - |
| RHBDD3 | 25807 | ENSG00000100263 | protein_coding | 100 | rs174767 | 0 | 1 | 1 | 0 | 0 | 0 | - | E | - | homeostasis/metabolism; cellular; hematopoietic system; mortality/aging; liver/biliary system; immune system; | - |
| SNORD125 | 100113380 | ENSG00000239127 | snoRNA | 100 | rs174767 | 0 | 1 | 1 | 0 | 0 | 0 | - | NE | - | - | - |
| AC002059.10 | - | ENSG00000273216 | lncRNA | 100 | rs174767 | 0 | 1 | 0 | 0 | 0 | 0 | - | E | - | - | - |
| CTA-984G1.5 | - | ENSG00000237015 | lncRNA | 100 | rs174767 | 0 | 0 | 1 | 0 | 0 | 0 | - | NE | - | - | - |
| EMID1 | 129080 | ENSG00000186998 | protein_coding | 100 | rs174767 | 0 | 0 | 1 | 0 | 0 | 0 | - | E | - | homeostasis/metabolism; | - |
| KREMEN1 | 83999 | ENSG00000183762 | protein_coding | 100 | rs174767 | 0 | 0 | 1 | 0 | 0 | 0 | - | E (UQ) | M7 | - | Ectodermal dysplasia 13, hair/tooth type, 617392 (3), Autosomal recessive |
| NIPSNAP1 | 8508 | ENSG00000184117 | protein_coding | 100 | rs174767 | 0 | 0 | 1 | 0 | 0 | 0 | - | E | - | hematopoietic system; behavior/neurological; homeostasis/metabolism; integument; | - |
| RFPL1S | 10740 | ENSG00000225465 | lncRNA | 100 | rs174767 | 0 | 0 | 1 | 0 | 0 | 0 | - | E | - | - | - |
| SRPX | 8406 | ENSG00000101955 | protein_coding | 101 | rs35318931 | 1 | 1 | 1 | 1 | 0 | 0 | coding variant & eQTL (rs35318931, PP=0.873) | E (UQ) | M1 | endocrine/exocrine gland; neoplasm; immune system; respiratory system; liver/biliary system; hematopoietic system; | - |
| SYTL5 | 94122 | ENSG00000147041 | protein_coding | 101 | rs35318931 | 0 | 1 | 1 | 0 | 0 | 0 | - | E | M4 | - | - |
| TM4SF2 | 7102 | ENSG00000250349 | protein_coding | 101 | rs35318931 | 0 | 1 | 0 | 0 | 0 | 0 | - | E | - | - | - |
| CHRDL1 | 91851 | ENSG00000101938 | protein_coding | 102 | rs5942977 | 1 | 1 | 1 | 1 | 0 | 0 | - | E (UQ) | M1_hub | nervous system; behavior/neurological; homeostasis/metabolism; | Megalocornea 1, X-linked, 309300 (3), X-linked recessive |

| | | | | | | | | | | | | | | | | |
|---|---|---|---|---|---|---|---|---|---|---|---|---|---|---|---|---|
| RGAG1 | 57529 | ENSG00000243978 | protein_coding | 102 | rs5942977 | 0 | 1 | 1 | 0 | 0 | 0 | - | E | M1 | - | - |
| AMMECR1 | 9949 | ENSG00000101935 | protein_coding | 102 | rs5942977 | 0 | 1 | 0 | 0 | 0 | 0 | - | E | M7 | - | Midface hypoplasia, hearing impairment, elliptocytosis, and nephrocalcinosis, 300990 (3), X-linked recessive |
| ABHD12 | - | ENSG00000100997 | protein_coding | - | - | 0 | 0 | 0 | 1 | 0 | 0 | - | E (UQ) | M4 | hearing/vestibular/ear; nervous system; immune system; behavior/neurological; hematopoietic system; muscle; homeostasis/metabolism; | Polyneuropathy, hearing loss, ataxia, retinitis pigmentosa, and cataract, 612674 (3), Autosomal recessive |
| AKT3 | - | ENSG00000117020 | protein_coding | - | - | 1 | 0 | 0 | 1 | 0 | 0 | - | E (UQ) | M1_hub | cellular; homeostasis/metabolism; behavior/neurological; nervous system; | Megalencephaly-polymicrogyria-polydactyly-hydrocephalus syndrome 2, 615937 (3), Autosomal dominant |
| ALKBH4 | - | ENSG00000160993 | protein_coding | - | - | 0 | 0 | 0 | 1 | 0 | 0 | - | E | - | cellular; mortality/aging; | - |
| ANKRA2 | - | ENSG00000164331 | protein_coding | - | - | 0 | 0 | 0 | 1 | 0 | 0 | - | E (UQ) | - | - | - |
| ANKRD44 | - | ENSG00000065413 | protein_coding | - | - | 0 | 0 | 0 | 1 | 0 | 0 | - | E | M1 | - | - |
| ARIH2 | - | ENSG00000177479 | protein_coding | - | - | 0 | 0 | 0 | 1 | 0 | 0 | - | E (UQ) | - | integument; growth/size/body region; endocrine/exocrine gland; cellular; homeostasis/metabolism; skeleton; renal/urinary system; embryo; respiratory system; liver/biliary system; behavior/neurological; reproductive system; mortality/aging; cardiovascular system; nervous system; immune system; | - |
| ASIC4 | - | ENSG00000072182 | protein_coding | - | - | 0 | 0 | 0 | 1 | 0 | 0 | - | E | - | nervous system; reproductive system; | - |
| BAG1 | - | ENSG00000107262 | protein_coding | - | - | 0 | 0 | 0 | 1 | 0 | 0 | - | E (UQ) | M4 | liver/biliary system; mortality/aging; hematopoietic system; | - |
| BCL11A | - | ENSG00000119866 | protein_coding | - | - | 0 | 0 | 0 | 1 | 0 | 0 | - | E | M4 | nervous system; immune system; mortality/aging; hematopoietic system; cellular; homeostasis/metabolism; endocrine/exocrine gland; | Dias-Logan syndrome, 617101 (3), Autosomal dominant |
| BOK | - | ENSG00000176720 | protein_coding | - | - | 0 | 0 | 0 | 1 | 0 | 0 | - | E (UQ) | - | immune system; behavior/neurological; hematopoietic system; endocrine/exocrine gland; | - |
| C14orf93 | - | ENSG00000100802 | protein_coding | - | - | 0 | 0 | 0 | 1 | 0 | 0 | - | E | - | endocrine/exocrine gland; homeostasis/metabolism; | - |
| C16orf70 | - | ENSG00000125149 | protein_coding | - | - | 0 | 0 | 0 | 1 | 0 | 0 | - | E | M7 | - | - |
| CBFB | - | ENSG00000067955 | protein_coding | - | - | 0 | 0 | 0 | 1 | 0 | 0 | - | E (UQ) | - | skeleton; immune system; vision/eye; digestive/alimentary; limbs/digits/tail; nervous system; cardiovascular system; hematopoietic system; mortality/aging; liver/biliary system; respiratory system; embryo; homeostasis/metabolism; cellular; craniofacial; endocrine/exocrine gland; growth/size/body region; integument; | Myeloid leukemia, acute, M4/M4Eo subtype, somatic, 601626 (1) |
| CCDC36 | - | ENSG00000173421 | protein_coding | - | - | 0 | 0 | 0 | 1 | 0 | 0 | - | E | - | - | - |
| CCNE1 | - | ENSG00000105173 | protein_coding | - | - | 0 | 0 | 0 | 1 | 0 | 0 | - | E | - | hematopoietic system; digestive/alimentary; immune system; skeleton; integument; endocrine/exocrine gland; cellular; | - |
| CD109 | - | ENSG00000156535 | protein_coding | - | - | 0 | 0 | 0 | 1 | 0 | 0 | - | E (UQ) | - | endocrine/exocrine gland; integument; immune system; | - |
| CDKN1A | - | ENSG00000124762 | protein_coding | - | - | 0 | 0 | 0 | 1 | 0 | 0 | - | E (UQ) | - | adipose tissue; endocrine/exocrine gland; integument; growth/size/body region; craniofacial; cellular; immune system; homeostasis/metabolism; respiratory system; liver/biliary system; neoplasm; mortality/aging; reproductive system; hematopoietic system; cardiovascular system; nervous system; digestive/alimentary; skeleton; renal/urinary system; | - |
| CHPF | - | ENSG00000123989 | protein_coding | - | - | 0 | 0 | 0 | 1 | 0 | 0 | - | E (UQ) | - | limbs/digits/tail; skeleton; behavior/neurological; growth/size/body region; homeostasis/metabolism; | - |
| CMSS1 | - | ENSG00000184220 | protein_coding | - | - | 0 | 0 | 0 | 1 | 0 | 0 | - | E (UQ) | M1 | - | - |
| CRB1 | - | ENSG00000134376 | protein_coding | - | - | 0 | 0 | 0 | 1 | 0 | 0 | - | E | - | homeostasis/metabolism; cardiovascular system; mortality/aging; pigmentation; nervous system; vision/eye; | Pigmented paravenous chorioretinal atrophy, 172870 (3), Autosomal dominant; Retinitis pigmentosa-12, 600105 (3), Autosomal recessive; Leber congenital amaurosis 8, 613835 (3), Autosomal recessive |
| CSPG4 | - | ENSG00000173546 | protein_coding | - | - | 0 | 0 | 0 | 1 | 0 | 0 | - | E | M1 | nervous system; immune system; liver/biliary system; hematopoietic system; adipose tissue; homeostasis/metabolism; cellular; growth/size/body region; | - |
| CSRP1 | - | ENSG00000159176 | protein_coding | - | - | 0 | 0 | 0 | 1 | 0 | 0 | - | E (UQ) | M1 | cardiovascular system; homeostasis/metabolism; | - |
| CUL9 | - | ENSG00000112659 | protein_coding | - | - | 0 | 0 | 0 | 1 | 0 | 0 | - | E (UQ) | - | liver/biliary system; respiratory system; neoplasm; reproductive system; mortality/aging; nervous system; cellular; homeostasis/metabolism; endocrine/exocrine gland; growth/size/body region; | - |
| DHX57 | - | ENSG00000163214 | protein_coding | - | - | 0 | 0 | 0 | 1 | 0 | 0 | - | E | - | - | - |
| ENTPD6 | - | ENSG00000197586 | protein_coding | - | - | 0 | 0 | 0 | 1 | 0 | 0 | - | E (UQ) | - | adipose tissue; growth/size/body region; skeleton; limbs/digits/tail; | - |
| ERLEC1 | - | ENSG00000068912 | protein_coding | - | - | 0 | 0 | 0 | 1 | 0 | 0 | - | E (UQ) | - | nervous system; hearing/vestibular/ear; behavior/neurological; homeostasis/metabolism; | - |
| ESRRG | - | ENSG00000196482 | protein_coding | - | - | 0 | 0 | 0 | 1 | 0 | 0 | - | E | - | growth/size/body region; cellular; homeostasis/metabolism; renal/urinary system; behavior/neurological; mortality/aging; cardiovascular system; hearing/vestibular/ear; | - |
| EVX2 | - | ENSG00000174279 | protein_coding | - | - | 0 | 0 | 0 | 1 | 0 | 0 | - | E | - | growth/size/body region; limbs/digits/tail; reproductive system; skeleton; | - |
| FILIP1L | - | ENSG00000168386 | protein_coding | - | - | 0 | 0 | 0 | 1 | 0 | 0 | - | E (UQ) | M1 | - | - |
| FOXP2 | - | ENSG00000128573 | protein_coding | - | - | 0 | 0 | 0 | 1 | 0 | 0 | - | E (UQ) | M1 | craniofacial; cellular; homeostasis/metabolism; growth/size/body region; adipose tissue; nervous system; hearing/vestibular/ear; vision/eye; immune system; behavior/neurological; respiratory system; mortality/aging; hematopoietic system; cardiovascular system; | Speech-language disorder-1, 602081 (3), Autosomal dominant |
| GABBR1 | - | ENSG00000204681 | protein_coding | - | - | 0 | 0 | 0 | 1 | 0 | 0 | - | E (UQ) | M1 | nervous system; behavior/neurological; mortality/aging; reproductive system; adipose tissue; homeostasis/metabolism; integument; growth/size/body region; | - |
| GFRA1 | - | ENSG00000151892 | protein_coding | - | - | 0 | 0 | 0 | 1 | 0 | 0 | - | E | - | integument; cellular; nervous system; muscle; digestive/alimentary; mortality/aging; embryo; renal/urinary system; immune system; | - |
| GINS1 | - | ENSG00000101003 | protein_coding | - | - | 0 | 0 | 0 | 1 | 0 | 0 | - | E | - | embryo; mortality/aging; cellular; | Immunodeficiency 55, 617827 (3), Autosomal recessive |
| GPR139 | - | ENSG00000180269 | protein_coding | - | - | 0 | 0 | 0 | 1 | 0 | 0 | - | E | - | cardiovascular system; | - |
| GPR75-ASB3 | - | ENSG00000115239 | protein_coding | - | - | 0 | 0 | 0 | 1 | 0 | 0 | - | E | M1 | - | - |
| HEY2 | - | ENSG00000135547 | protein_coding | - | - | 0 | 0 | 0 | 1 | 0 | 0 | - | E | M1 | homeostasis/metabolism; muscle; growth/size/body region; cardiovascular system; mortality/aging; respiratory system; liver/biliary system; | - |
| HLCS | - | ENSG00000159267 | protein_coding | - | - | 0 | 0 | 0 | 1 | 0 | 0 | - | E | M4 | mortality/aging; embryo; growth/size/body region; | Holocarboxylase synthetase deficiency, 253270 (3), Autosomal recessive |
| HSPG2 | - | ENSG00000142798 | protein_coding | - | - | 0 | 0 | 0 | 1 | 0 | 0 | - | E (UQ) | M1 | growth/size/body region; integument; craniofacial; muscle; homeostasis/metabolism; immune system; cellular; behavior/neurological; respiratory system; liver/biliary system; embryo; cardiovascular system; hematopoietic system; mortality/aging; vision/eye; digestive/alimentary; nervous system; hearing/vestibular/ear; limbs/digits/tail; skeleton; | Dyssegmental dysplasia, Silverman-Handmaker type, 224410 (3), Autosomal recessive; Schwartz-Jampel syndrome, type 1, 255800 (3), Autosomal recessive |
| IQCK | - | ENSG00000174628 | protein_coding | - | - | 0 | 0 | 0 | 1 | 0 | 0 | - | E | - | - | - |
| KCNK2 | - | ENSG00000082482 | protein_coding | - | - | 0 | 0 | 0 | 1 | 0 | 0 | - | E | - | behavior/neurological; mortality/aging; nervous system; homeostasis/metabolism; | - |
| KCTD19 | - | ENSG00000168676 | protein_coding | - | - | 0 | 0 | 0 | 1 | 0 | 0 | - | E | - | - | - |
| KIAA0040 | - | ENSG00000235750 | protein_coding | - | - | 0 | 0 | 0 | 1 | 0 | 0 | - | E (UQ) | - | - | - |
| KIAA0408 | - | ENSG00000189367 | protein_coding | - | - | 1 | 0 | 0 | 1 | 0 | 0 | - | E | M1_hub | - | - |
| LBH | - | ENSG00000213626 | protein_coding | - | - | 0 | 0 | 0 | 1 | 0 | 0 | - | E (UQ) | M1 | integument; endocrine/exocrine gland; reproductive system; | - |
| MAPRE2 | - | ENSG00000166974 | protein_coding | - | - | 0 | 0 | 0 | 1 | 0 | 0 | - | E (UQ) | - | - | Symmetric circumferential skin creases, congenital, 2, 616734 (3), Autosomal dominant |
| MECOM | - | ENSG00000085276 | protein_coding | - | - | 0 | 0 | 0 | 1 | 0 | 0 | - | E (UQ) | - | immune system; skeleton; hearing/vestibular/ear; nervous system; limbs/digits/tail; hematopoietic system; cardiovascular system; mortality/aging; reproductive system; embryo; behavior/neurological; respiratory system; liver/biliary system; homeostasis/metabolism; cellular; muscle; craniofacial; growth/size/body region; integument; | Radioulnar synostosis with amegakaryocytic thrombocytopenia 2, 616738 (3), Autosomal dominant |
| MOB4 | - | ENSG00000115540 | protein_coding | - | - | 0 | 0 | 0 | 1 | 0 | 0 | - | E (UQ) | - | - | - |
| MOV10 | - | ENSG00000155363 | protein_coding | - | - | 0 | 0 | 0 | 1 | 0 | 0 | - | E (UQ) | M7 | nervous system; mortality/aging; behavior/neurological; | - |
| MSRA | - | ENSG00000175806 | protein_coding | - | - | 0 | 0 | 0 | 1 | 0 | 0 | - | E (UQ) | - | mortality/aging; behavior/neurological; homeostasis/metabolism; cellular; | - |
| NDP | 4693 | ENSG00000124479 | protein_coding | - | - | 0 | 0 | 0 | 1 | 0 | 0 | - | E | M1 | cardiovascular system; pigmentation; nervous system; hearing/vestibular/ear; vision/eye; skeleton; | Exudative vitreoretinopathy 2, X-linked, 305390 (3), X-linked recessive, X-linked dominant; Norrie disease, 310600 (3), X-linked recessive |
| NEGR1 | - | ENSG00000172260 | protein_coding | - | - | 1 | 0 | 0 | 1 | 0 | 0 | - | E | M1_hub | reproductive system; hematopoietic system; cardiovascular system; behavior/neurological; liver/biliary system; immune system; skeleton; vision/eye; growth/size/body region; endocrine/exocrine gland; homeostasis/metabolism; | - |
| NFIX | - | ENSG00000008441 | protein_coding | - | - | 0 | 0 | 0 | 1 | 0 | 0 | - | E (UQ) | - | skeleton; immune system; vision/eye; hearing/vestibular/ear; nervous system; digestive/alimentary; mortality/aging; hematopoietic system; behavior/neurological; homeostasis/metabolism; craniofacial; adipose tissue; growth/size/body region; | Marshall-Smith syndrome, 602535 (3), Autosomal dominant; Sotos syndrome 2, 614753 (3), Autosomal dominant |
| NMB | - | ENSG00000197696 | protein_coding | - | - | 0 | 0 | 0 | 1 | 0 | 0 | - | E | - | behavior/neurological; nervous system; | - |
| NPC1L1 | - | ENSG00000015520 | protein_coding | - | - | 0 | 0 | 0 | 1 | 0 | 0 | - | E | M4 | liver/biliary system; digestive/alimentary; growth/size/body region; homeostasis/metabolism; | [Low density lipoprotein cholesterol level QTL 7], 617966 (3); [Ezetimibe, nonresponse to], 617966 (3) |
| NUDT5 | - | ENSG00000165609 | protein_coding | - | - | 0 | 0 | 0 | 1 | 0 | 0 | - | E (UQ) | - | - | - |
| OBSL1 | - | ENSG00000124006 | protein_coding | - | - | 0 | 0 | 0 | 1 | 0 | 0 | - | E | M1 | - | 3-M syndrome 2, 612921 (3), Autosomal recessive |
| OSBPL2 | - | ENSG00000130703 | protein_coding | - | - | 0 | 0 | 0 | 1 | 0 | 0 | - | E (UQ) | M4 | - | Deafness, autosomal dominant 67, 616340 (3), Autosomal dominant |
| PARD3 | - | ENSG00000148498 | protein_coding | - | - | 0 | 0 | 0 | 1 | 0 | 0 | - | E (UQ) | M7 | embryo; neoplasm; pigmentation; mortality/aging; hematopoietic system; cardiovascular system; vision/eye; limbs/digits/tail; nervous system; homeostasis/metabolism; integument; growth/size/body region; | - |
| PBX1 | - | ENSG00000185630 | protein_coding | - | - | 0 | 0 | 0 | 1 | 0 | 0 | - | E (UQ) | - | integument; growth/size/body region; endocrine/exocrine gland; muscle; craniofacial; cellular; homeostasis/metabolism; renal/urinary system; skeleton; embryo; behavior/neurological; respiratory system; liver/biliary system; mortality/aging; reproductive system; hematopoietic system; cardiovascular system; hearing/vestibular/ear; limbs/digits/tail; digestive/alimentary; immune system; | Congenital anomalies of kidney and urinary tract syndrome with or without hearing loss, abnormal ears, or developmental delay, 617641 (3), Autosomal dominant |
| PLEKHG4 | - | ENSG00000196155 | protein_coding | - | - | 0 | 0 | 0 | 1 | 0 | 0 | - | E | - | - | - |
| PLOD2 | - | ENSG00000152952 | protein_coding | - | - | 0 | 0 | 0 | 1 | 0 | 0 | - | E (UQ) | - | mortality/aging; | Bruck syndrome 2, 609220 (3), Autosomal recessive |
| PRDM6 | - | ENSG00000061455 | protein_coding | - | - | 1 | 0 | 0 | 1 | 0 | 0 | - | E | M1_hub | mortality/aging; cardiovascular system; respiratory system; embryo; muscle; integument; homeostasis/metabolism; | Patent ductus arteriosus 3, 617039 (3), Autosomal dominant |
| PRKAR2A | - | ENSG00000114302 | protein_coding | - | - | 0 | 0 | 0 | 1 | 0 | 0 | - | E (UQ) | M4 | - | - |

| | | | | | | | | | | | | | | | | |
|---|---|---|---|---|---|---|---|---|---|---|---|---|---|---|---|---|
| PSME4 | - | ENSG00000068878 | protein_coding | - | - | 0 | 0 | 0 | 1 | 0 | 0 | - | E (UQ) | - | cellular; reproductive system; | - |
| PSPC1 | - | ENSG00000121390 | protein_coding | - | - | 0 | 0 | 0 | 1 | 0 | 0 | - | E (UQ) | - | - | - |
| PYGB | - | ENSG00000100994 | protein_coding | - | - | 0 | 0 | 0 | 1 | 0 | 0 | - | E (UQ) | M4 | - | - |
| QARS | - | ENSG00000172053 | protein_coding | - | - | 0 | 0 | 0 | 1 | 0 | 0 | - | E (UQ) | M4 | - | - |
| QRICH1 | - | ENSG00000198218 | protein_coding | - | - | 0 | 0 | 0 | 1 | 0 | 0 | - | E (UQ) | M1 | cardiovascular system; hematopoietic system; digestive/alimentary; renal/urinary system; immune system; endocrine/exocrine gland; growth/size/body region; craniofacial; muscle; | Ververi-Brady syndrome, 617982 (3), Autosomal dominant |
| RAD51B | - | ENSG00000182185 | protein_coding | - | - | 0 | 0 | 0 | 1 | 0 | 0 | - | E | M7 | mortality/aging; | - |
| RASA2 | - | ENSG00000155903 | protein_coding | - | - | 0 | 0 | 0 | 1 | 0 | 0 | - | E | - | - | - |
| SCARA5 | - | ENSG00000168079 | protein_coding | - | - | 0 | 0 | 0 | 1 | 0 | 0 | - | E (UQ) | M1 | integument; adipose tissue; endocrine/exocrine gland; immune system; renal/urinary system; liver/biliary system; respiratory system; reproductive system; | - |
| SEC11A | - | ENSG00000140612 | protein_coding | - | - | 0 | 0 | 0 | 1 | 0 | 0 | - | E (UQ) | - | - | - |
| SEC61A2 | - | ENSG00000065665 | protein_coding | - | - | 0 | 0 | 0 | 1 | 0 | 0 | - | E | - | - | - |
| SLC25A20 | - | ENSG00000178537 | protein_coding | - | - | 0 | 0 | 0 | 1 | 0 | 0 | - | E (UQ) | - | homeostasis/metabolism; endocrine/exocrine gland; cardiovascular system; mortality/aging; nervous system; renal/urinary system; liver/biliary system; embryo; | Carnitine-acylcarnitine translocase deficiency, 212138 (3), Autosomal recessive |
| SLC35F3 | - | ENSG00000183780 | protein_coding | - | - | 0 | 0 | 0 | 1 | 0 | 0 | - | E | - | - | - |
| SLC9A5 | - | ENSG00000135740 | protein_coding | - | - | 0 | 0 | 0 | 1 | 0 | 0 | - | E | - | - | - |
| SNTG1 | - | ENSG00000147481 | protein_coding | - | - | 0 | 0 | 0 | 1 | 0 | 0 | - | E | M1 | - | - |
| SOGA3 | - | ENSG00000214338 | protein_coding | - | - | 0 | 0 | 0 | 1 | 0 | 0 | - | E | M1 | - | - |
| SOGA3 | - | ENSG00000255330 | protein_coding | - | - | 1 | 0 | 0 | 1 | 0 | 0 | - | E | M1_hub | - | - |
| TMEM198 | - | ENSG00000188760 | protein_coding | - | - | 0 | 0 | 0 | 1 | 0 | 0 | - | E | M1 | - | - |
| TTC31 | - | ENSG00000115282 | protein_coding | - | - | 0 | 0 | 0 | 1 | 0 | 0 | - | E (UQ) | M1 | - | - |
| WDR70 | - | ENSG00000082068 | protein_coding | - | - | 0 | 0 | 0 | 1 | 0 | 0 | - | E | - | - | - |
| WDR73 | - | ENSG00000177082 | protein_coding | - | - | 0 | 0 | 0 | 1 | 0 | 0 | - | E (UQ) | - | - | Galloway-Mowat syndrome 1, 251300 (3), Autosomal recessive |
| XKR6 | - | ENSG00000171044 | protein_coding | - | - | 0 | 0 | 0 | 1 | 0 | 0 | - | E | - | - | - |
| ZBTB38 | - | ENSG00000177311 | protein_coding | - | - | 0 | 0 | 0 | 1 | 0 | 0 | - | E (UQ) | - | - | - |
| ZNF521 | - | ENSG00000198795 | protein_coding | - | - | 0 | 0 | 0 | 1 | 0 | 0 | - | E | M1 | cellular; craniofacial; growth/size/body region; endocrine/exocrine gland; respiratory system; liver/biliary system; behavior/neurological; immune system; renal/urinary system; skeleton; nervous system; hematopoietic system; cardiovascular system; mortality/aging; | - |
| ZNF592 | - | ENSG00000166716 | protein_coding | - | - | 0 | 0 | 0 | 1 | 0 | 0 | - | E (UQ) | - | - | - |
| ZSCAN2 | - | ENSG00000176371 | protein_coding | - | - | 0 | 0 | 0 | 1 | 0 | 0 | - | E | - | skeleton; | - |
