## Supplementary Table 8 for "Genome-wide analysis of 944,133 individuals provides insights into the etiology of hemorrhoidal disease"

**Supplementary Table 8. Gene set and tissue enrichment analyses of HEM genes**

This table contains all tissues and pathways if they are significantly enriched in at least one out of three analytic tools (FUMA, MAGMA or DEPICT; **Online Methods**).

**Genesets**: The enriched pathway or tissue. Gene-sets were only reported if their FDR corrected *P*-values were lower than 0.05 in at least one of the three methods. **Category**: GTEx tissue: 30 GTEx general tissue types using data from GTEx release v7 database), GO_BP: gene ontology biological process, GP_MF: gene ontology molecular function and GO_CC: gene ontology cellular component from Molecular Signature Database; **P.FDR.FUMA:** the *P*-value from the statistical test of enrichment in FUMA after FDR correction for multiple comparisons at $\alpha$=0.05; **P.FDR.MAGMA:** the *P*-value from the statistical test of enrichment in MAGMA after FDR correction for multiple comparisons at $\alpha$=0.05; **P.FDR.DEPICT:** the *P*-value from the statistical test of enrichment in DEPICT after FDR correction for multiple comparisons at $\alpha$=0.05. "-": data is not available.

| Genesets | Category | P.FDR.FUMA | P.FDR.MAGMA | P.FDR.DEPICT |
|---|---|---|---|---|
| positive regulation of interleukin 6 production | GO_BP | 1,1E-02 | 5,6E-01 | - |
| tube morphogenesis | GO_BP | 1,2E-02 | 3,4E-03 | 3,6E-02 |
| negative regulation of mitotic cell cycle | GO_BP | 1,2E-02 | 3,9E-01 | 1,0E+00 |
| skeletal system development | GO_BP | 1,8E-02 | 3,5E-02 | 3,6E-02 |
| regulation of anatomical structure morphogenesis | GO_BP | 1,8E-02 | 1,1E-01 | 3,0E-01 |
| inflammatory response | GO_BP | 2,1E-02 | 1,2E-01 | 1,0E+00 |
| regulation of vasculature development | GO_BP | 2,2E-02 | 3,0E-01 | - |
| tube development | GO_BP | 2,4E-02 | 4,6E-03 | 2,4E-02 |
| embryo development | GO_BP | 5,5E-02 | 5,5E-03 | - |
| fibroblast proliferation | GO_BP | 6,1E-02 | 3,7E-03 | 7,7E-01 |
| embryonic morphogenesis | GO_BP | 6,9E-02 | 2,5E-02 | 1,3E-01 |
| regulation of response to stress | GO_BP | 6,9E-02 | 4,7E-03 | - |
| regulation of cell population proliferation | GO_BP | 7,7E-02 | 2,0E-02 | - |
| positive regulation of transcription by rna polymerase ii | GO_BP | 8,4E-02 | 4,6E-03 | - |
| positive regulation of rna biosynthetic process | GO_BP | 8,5E-02 | 1,7E-03 | - |
| positive regulation of rna polymerase ii transcriptional preinitiation complex assembl | GO_BP | 8,5E-02 | 2,3E-02 | - |
| spinal cord motor neuron cell fate specification | GO_BP | 9,8E-02 | 7,6E-03 | - |
| appendage morphogenesis | GO_BP | 9,9E-02 | 4,2E-02 | 3,2E-01 |
| ameboidal type cell migration | GO_BP | 1,0E-01 | 5,1E-03 | - |
| proximal distal pattern formation | GO_BP | 1,0E-01 | 4,3E-02 | - |
| tissue morphogenesis | GO_BP | 1,2E-01 | 1,1E-02 | 1,1E-02 |
| neuron fate specification | GO_BP | 1,2E-01 | 4,7E-02 | 7,7E-01 |
| positive regulation of biosynthetic process | GO_BP | 1,4E-01 | 4,3E-03 | - |
| tissue remodeling | GO_BP | 1,5E-01 | 7,1E-01 | 4,9E-02 |
| macrophage colony stimulating factor production | GO_BP | 1,6E-01 | 2,5E-02 | - |
| osteoblast differentiation | GO_BP | 1,6E-01 | 3,8E-02 | 1,5E-01 |
| positive regulation of signaling | GO_BP | 1,7E-01 | 4,3E-02 | - |
| appendage development | GO_BP | 1,8E-01 | 1,5E-02 | 2,4E-01 |
| positive regulation of gene expression | GO_BP | 1,8E-01 | 3,8E-03 | - |
| chromatin organization | GO_BP | 1,8E-01 | 4,5E-02 | 1,0E+00 |
| response to growth factor | GO_BP | 1,9E-01 | 4,6E-03 | - |
| animal organ morphogenesis | GO_BP | 2,4E-01 | 2,2E-03 | - |
| positive regulation of fibroblast proliferation | GO_BP | 2,6E-01 | 5,6E-03 | 8,2E-01 |
| muscle tissue development | GO_BP | 3,2E-01 | 2,2E-03 | 6,8E-02 |
| embryo development ending in birth or egg hatching | GO_BP | 3,3E-01 | 5,6E-03 | 9,6E-02 |
| mesenchymal cell differentiation | GO_BP | 3,3E-01 | 1,8E-02 | 1,1E-01 |
| muscle organ development | GO_BP | 3,3E-01 | 5,5E-03 | 1,5E-01 |
| morphogenesis of an epithelium | GO_BP | 3,6E-01 | 4,9E-02 | 1,5E-02 |
| cardiac muscle cell myoblast differentiation | GO_BP | 3,6E-01 | 3,8E-02 | - |
| negative regulation of rna biosynthetic process | GO_BP | 3,7E-01 | 2,1E-02 | - |
| mesonephros development | GO_BP | 3,7E-01 | 1,0E-02 | 7,6E-01 |
| mesonephric tubule morphogenesis | GO_BP | 3,8E-01 | 1,2E-02 | - |
| negative regulation of developmental process | GO_BP | 3,8E-01 | 2,5E-02 | 4,7E-01 |
| regulation of cellular response to stress | GO_BP | 3,8E-01 | 3,3E-02 | 8,3E-01 |
| branching morphogenesis of an epithelial tube | GO_BP | 4,1E-01 | 1,5E-02 | - |
| morphogenesis of a branching structure | GO_BP | 4,1E-01 | 6,7E-03 | 1,6E-02 |
| activin receptor signaling pathway | GO_BP | 4,6E-01 | 3,8E-02 | 8,4E-01 |
| cartilage development | GO_BP | 4,7E-01 | 6,9E-01 | 1,3E-02 |
| sensory system development | GO_BP | 4,9E-01 | 1,8E-02 | - |
| nephron morphogenesis | GO_BP | 4,9E-01 | 1,9E-02 | 3,4E-01 |
| muscle structure development | GO_BP | 5,1E-01 | 1,3E-02 | 8,7E-02 |
| epithelial tube morphogenesis | GO_BP | 5,1E-01 | 6,7E-03 | 7,6E-02 |
| sensory organ development | GO_BP | 5,2E-01 | 2,5E-02 | 4,0E-01 |
| cell proliferation involved in heart morphogenesis | GO_BP | 5,3E-01 | 3,8E-02 | - |
| mesenchyme development | GO_BP | 5,4E-01 | 5,5E-03 | 1,0E-01 |
| cardioblast differentiation | GO_BP | 5,5E-01 | 4,7E-03 | 8,9E-01 |
| skeletal system morphogenesis | GO_BP | 6,9E-01 | 1,1E-02 | 2,3E-01 |
| positive regulation of intracellular signal transduction | GO_BP | 7,0E-01 | 4,6E-02 | - |
| negative regulation of biosynthetic process | GO_BP | 7,4E-01 | 2,0E-02 | - |
| adrenal gland development | GO_BP | 7,5E-01 | 2,5E-02 | 1,0E+00 |
| stem cell differentiation | GO_BP | 7,6E-01 | 5,1E-03 | 8,7E-01 |
| reproductive system development | GO_BP | 8,1E-01 | 4,5E-02 | - |
| bone development | GO_BP | 8,7E-01 | 2,2E-01 | 4,7E-02 |
| heart development | GO_BP | 8,7E-01 | 6,5E-02 | 4,6E-02 |
| formation of primary germ layer | GO_BP | 9,0E-01 | 2,6E-02 | 1,8E-01 |
| mesoderm morphogenesis | GO_BP | 9,1E-01 | 3,9E-02 | 7,3E-02 |
| mesodermal cell differentiation | GO_BP | 9,1E-01 | 1,5E-01 | 2,4E-02 |
| positive regulation of cell population proliferation | GO_BP | 9,1E-01 | 3,2E-02 | - |
| supramolecular fiber organization | GO_BP | 9,1E-01 | 3,9E-02 | - |
| outflow tract morphogenesis | GO_BP | 9,2E-01 | 4,0E-03 | 8,0E-01 |
| mesoderm development | GO_BP | 9,4E-01 | 2,8E-02 | 4,3E-01 |
| extracellular structure organization | GO_BP | 9,6E-01 | 7,2E-02 | 2,6E-02 |
| camera type eye development | GO_BP | 1,0E+00 | 2,5E-02 | - |
| artery morphogenesis | GO_BP | - | 4,7E-03 | 4,0E-02 |
| smooth muscle tissue development | GO_BP | - | 3,0E-02 | 2,8E-02 |
| aging | GO_BP | - | 8,2E-01 | 3,9E-02 |
| artery development | GO_BP | - | 1,2E-02 | 6,8E-02 |
| atrial septum morphogenesis | GO_BP | - | 4,7E-02 | - |
| cardiac atrium development | GO_BP | - | 1,1E-02 | 1,0E+00 |
| exocrine system development | GO_BP | - | 1,1E-01 | 4,1E-02 |
| heart morphogenesis | GO_BP | - | 3,7E-02 | 2,6E-01 |
| integrin biosynthetic process | GO_BP | - | 3,0E-02 | - |
| mammary gland branching involved in pregnancy | GO_BP | - | 4,3E-02 | - |
| mesenchyme morphogenesis | GO_BP | - | 4,3E-02 | 5,7E-01 |
| muscle cell proliferation | GO_BP | - | 1,7E-02 | 2,6E-01 |
| myotube differentiation | GO_BP | - | 2,6E-01 | 4,6E-02 |
| negative regulation of chondrocyte differentiation | GO_BP | - | 6,8E-01 | 1,2E-02 |
| negative regulation of smooth muscle cell apoptotic process | GO_BP | - | 1,7E-02 | - |
| neural crest cell differentiation | GO_BP | - | 3,9E-02 | 1,2E-01 |
| paraxial mesoderm development | GO_BP | - | 1,1E-02 | 2,1E-01 |

| | | | | |
|---|---|---|---|---|
| paraxial mesoderm morphogenesis | GO_BP | - | 6,7E-03 | - |
| positive regulation of alkaline phosphatase activity | GO_BP | - | 2,8E-02 | - |
| positive regulation of cardiac muscle tissue development | GO_BP | - | 2,5E-02 | - |
| positive regulation of cellular component organization | GO_BP | - | 4,1E-02 | 4,0E-01 |
| regulation of alkaline phosphatase activity | GO_BP | - | 3,7E-02 | - |
| regulation of cardiocyte differentiation | GO_BP | - | 3,8E-02 | - |
| regulation of cell fate specification | GO_BP | - | 4,5E-02 | 1,0E+00 |
| regulation of epithelial cell proliferation involved in lung morphogenesis | GO_BP | - | 2,9E-02 | - |
| regulation of lung blood pressure | GO_BP | - | 6,8E-03 | - |
| regulation of mesoderm formation | GO_BP | - | 4,9E-02 | - |
| regulation of mesodermal cell differentiation | GO_BP | - | 4,9E-02 | - |
| regulation of mesodermal cell fate specification | GO_BP | - | 4,9E-02 | - |
| regulation of muscle organ development | GO_BP | - | 4,6E-03 | 5,2E-01 |
| regulation of phosphorus metabolic process | GO_BP | - | 3,6E-02 | - |
| skeletal muscle cell differentiation | GO_BP | - | 6,5E-01 | 4,6E-02 |
| smooth muscle contraction | GO_BP | - | 7,9E-01 | 4,1E-02 |
| smoothened signaling pathway | GO_BP | - | 8,3E-01 | 3,9E-02 |
| tendon development | GO_BP | - | 4,5E-02 | - |
| tricuspid valve development | GO_BP | - | 4,3E-02 | - |
| tricuspid valve morphogenesis | GO_BP | - | 2,5E-02 | - |
| extracellular matrix | GO_CC | 1,4E-01 | 4,9E-02 | 4,1E-01 |
| collagen containing extracellular matrix | GO_CC | 2,4E-01 | 4,5E-02 | - |
| transcription factor complex | GO_CC | - | 4,3E-01 | 4,6E-02 |
| interleukin 1 receptor binding | GO_MF | 5,1E-07 | 7,5E-01 | - |
| growth factor receptor binding | GO_MF | 7,6E-03 | 3,8E-01 | 7,2E-01 |
| cytokine receptor binding | GO_MF | 1,0E-02 | 8,4E-01 | 9,0E-01 |
| cytokine activity | GO_MF | 1,1E-02 | 8,2E-01 | 6,5E-01 |
| signaling receptor binding | GO_MF | 3,1E-02 | 1,3E-01 | - |
| nuclear hormone receptor binding | GO_MF | 3,9E-02 | 6,0E-03 | 6,7E-01 |
| hormone receptor binding | GO_MF | 8,5E-02 | 3,6E-02 | 1,0E+00 |
| nuclear receptor binding | GO_MF | 8,5E-02 | 6,7E-03 | - |
| rna polymerase ii specific dna binding transcription factor binding | GO_MF | 8,5E-02 | 5,7E-03 | - |
| transmembrane receptor protein kinase activity | GO_MF | 8,5E-02 | 3,7E-03 | 2,8E-01 |
| dna binding transcription factor binding | GO_MF | 1,5E-01 | 1,8E-02 | - |
| smad binding | GO_MF | 3,0E-01 | 5,0E-02 | 3,6E-02 |
| transcription factor binding | GO_MF | 7,4E-01 | 5,4E-02 | 3,8E-02 |
| transmembrane receptor protein serine threonine kinase activity | GO_MF | 7,6E-01 | 6,7E-03 | - |
| activin activated receptor activity | GO_MF | - | 4,3E-02 | - |
| bmp receptor activity | GO_MF | - | 4,3E-02 | - |
| chromatin dna binding | GO_MF | - | 4,4E-01 | 1,4E-02 |
| regulatory region nucleic acid binding | GO_MF | - | 1,3E-02 | 7,7E-02 |
| transforming growth factor beta receptor activity type i | GO_MF | - | 1,2E-02 | - |
| colon | GTEx tissue | 7,5E-03 | 7,6E-04 | 2,6E-02 |
| blood vessel | GTEx tissue | 9,8E-03 | 3,7E-03 | 1,0E-01 |
| esophagus | GTEx tissue | 5,1E-01 | 4,4E-03 | 4,8E-03 |
| vagina | GTEx tissue | 5,3E-01 | 9,9E-03 | 1,0E-01 |
| bladder | GTEx tissue | 7,1E-01 | 3,8E-04 | 1,0E-01 |
| cervix uteri | GTEx tissue | 7,1E-01 | 2,2E-06 | 1,3E-01 |
| prostate | GTEx tissue | 9,4E-01 | 1,3E-03 | 7,5E-02 |
| stomach | GTEx tissue | 1,0E+00 | 9,3E-03 | 1,9E-02 |
| uterus | GTEx tissue | 1,0E+00 | 5,0E-05 | 1,0E-01 |
