## Supplementary Table 9 for "Genome-wide analysis of 944,133 individuals provides insights into the etiology of hemorrhoidal disease"

**Supplementary Table 9. HEM gene overrepresentation analysis in gene co-expression network modules of hemorrhoidal tissue.**

The overrepresentation analysis was performed using Fisher's exact test with the alternative hypothesis that the true odds ratio is greater than one.

**Module:** network module of co-expressed genes in hemorrhoids tissue; **N HEM genes:** number of HEM candidate genes in module; **N genes:** total number of genes in module; **HEM gene frac, %:** percentage of HEM candidate genes in a given module; **OR:** odds ratio; **FDR:** *P*-value after FDR correction.

| Module | N HEM genes | N genes | HEM gene frac, % | OR | P FDR |
|---|---|---|---|---|---|
| M1 | 121 | 3975 | 3,0 | 1,65 | 6,4E-05 |
| M4 | 75 | 2547 | 2,9 | 1,55 | 1,0E-02 |
| M7 | 64 | 2171 | 2,9 | 1,54 | 1,7E-02 |
| M2 | 73 | 2956 | 2,5 | 1,27 | 3,6E-01 |
| M29 | 7 | 212 | 3,3 | 1,68 | 9,1E-01 |
| M3 | 63 | 2721 | 2,3 | 1,18 | 9,1E-01 |
| M10 | 19 | 1617 | 1,2 | 0,57 | 1,0E+00 |
| M11 | 28 | 1254 | 2,2 | 1,13 | 1,0E+00 |
| M12 | 11 | 998 | 1,1 | 0,54 | 1,0E+00 |
| M13 | 15 | 914 | 1,6 | 0,82 | 1,0E+00 |
| M14 | 9 | 863 | 1,0 | 0,51 | 1,0E+00 |
| M15 | 13 | 777 | 1,7 | 0,83 | 1,0E+00 |
| M16 | 8 | 670 | 1,2 | 0,59 | 1,0E+00 |
| M17 | 16 | 622 | 2,6 | 1,30 | 1,0E+00 |
| M18 | 10 | 595 | 1,7 | 0,84 | 1,0E+00 |
| M19 | 4 | 456 | 0,9 | 0,43 | 1,0E+00 |
| M20 | 9 | 424 | 2,1 | 1,07 | 1,0E+00 |
| M21 | 5 | 424 | 1,2 | 0,58 | 1,0E+00 |
| M22 | 6 | 364 | 1,6 | 0,82 | 1,0E+00 |
| M23 | 9 | 361 | 2,5 | 1,26 | 1,0E+00 |
| M24 | 7 | 335 | 2,1 | 1,05 | 1,0E+00 |
| M25 | 4 | 323 | 1,2 | 0,61 | 1,0E+00 |
| M26 | 4 | 295 | 1,4 | 0,67 | 1,0E+00 |
| M27 | 3 | 242 | 1,2 | 0,62 | 1,0E+00 |
| M28 | 3 | 233 | 1,3 | 0,64 | 1,0E+00 |
| M30 | 3 | 190 | 1,6 | 0,79 | 1,0E+00 |
| M31 | 3 | 175 | 1,7 | 0,86 | 1,0E+00 |
| M32 | 1 | 153 | 0,7 | 0,32 | 1,0E+00 |
| M33 | 2 | 142 | 1,4 | 0,70 | 1,0E+00 |
| M34 | 3 | 128 | 2,3 | 1,18 | 1,0E+00 |
| M35 | 1 | 123 | 0,8 | 0,40 | 1,0E+00 |
| M36 | 0 | 121 | 0,0 | 0,00 | 1,0E+00 |
| M37 | 1 | 96 | 1,0 | 0,52 | 1,0E+00 |
| M38 | 1 | 89 | 1,1 | 0,56 | 1,0E+00 |
| M39 | 2 | 72 | 2,8 | 1,40 | 1,0E+00 |
| M40 | 2 | 64 | 3,1 | 1,59 | 1,0E+00 |
| M41 | 0 | 62 | 0,0 | 0,00 | 1,0E+00 |
| M5 | 44 | 2430 | 1,8 | 0,90 | 1,0E+00 |
| M6 | 22 | 2210 | 1,0 | 0,48 | 1,0E+00 |
| M8 | 18 | 2135 | 0,8 | 0,40 | 1,0E+00 |
| M9 | 36 | 1803 | 2,0 | 1,00 | 1,0E+00 |
