## Supplementary Table 10 for "Genome-wide analysis of 944,133 individuals provides insights into the etiology of hemorrhoidal disease"

**Supplementary Table 10. Significant eQTL associations of fine-mapped variants and HEM genes.**

This table contains a list of significant eQTL associations (FDR<0.05) with details for fine-mapped variants (posterior probability>50%, see **Online Methods** and **Supplementary_Table 3**).

**Fine mapped SNP (rsID):** rs ID of the fine-mapped variants retrieved from NCBI's dbSNP build v150; **Position:** base pair position of the lead SNP (in "chromosome:base pair" format), genomic positions were retrieved from NCBI's dbSNP build v150 (genome build hg19); **Posterior probability:** posterior probability of causality to each SNP variant; **SNP Type:** functional consequence of the SNP obtained from ANNOVAR; **eQTL database:** Data source of eQTLs from the 10 repositories (see **Online Methods**); **tissue:** tissue type of the eQTL association; **EnsemblID:** gene ID extracted from ensembl database GRCh38.p13 (www.ensembl.org/); **Gene symbol:** gene ID extracted from HUGO Gene Nomenclature Committee; **P.FDR:** *P*-value of eQTLs after FDR correction for multiple comparisons at α=0.05.

| Fine mapped SNP (rsID) | Position | Posterior probability | SNP Type | eQTL database | tissue | EnsemblID | Gene symbol | P.FDR |
|---|---|---|---|---|---|---|---|---|
| rs2186797 | 11:70007770 | 0,969962 | exonic | GTEx/v7 | Adipose_Subcutaneous | ENSG00000131620 | ANO1 | 1,5E-03 |
| rs2186797 | 11:70007770 | 0,969962 | exonic | BIOSQTL | BIOS_eQTL_geneLevel | ENSG00000168040 | FADD | 8,2E-03 |
| rs2186797 | 11:70007770 | 0,969962 | exonic | BIOSQTL | BIOS_eQTL_geneLevel | ENSG00000168040 | FADD | 7,9E-03 |
| rs10956488 | 8:130717755 | 0,961845 | intergenic | GTEx/v7 | Esophagus_Mucosa | ENSG00000147697 | GSDMC | 2,0E-66 |
| rs10956488 | 8:130717755 | 0,961845 | intergenic | GTEx/v7 | Skin_Sun_Exposed_Lower_le | ENSG00000147697 | GSDMC | 2,5E-39 |
| rs4556017 | 7:100632790 | 0,961771 | intronic | BIOSQTL | BIOS_eQTL_geneLevel | ENSG00000087085 | ACHE | 1,9E-02 |
| rs4556017 | 7:100632790 | 0,961771 | intronic | GTEx/v7 | Artery_Aorta | ENSG00000087085 | ACHE | 1,6E-21 |
| rs4556017 | 7:100632790 | 0,961771 | intronic | GTEx/v7 | Artery_Coronary | ENSG00000087085 | ACHE | 5,9E-07 |
| rs4556017 | 7:100632790 | 0,961771 | intronic | GTEx/v7 | Artery_Tibial | ENSG00000087085 | ACHE | 6,5E-22 |
| rs4556017 | 7:100632790 | 0,961771 | intronic | GTEx/v7 | Esophagus_Muscularis | ENSG00000087085 | ACHE | 1,0E-06 |
| rs4556017 | 7:100632790 | 0,961771 | intronic | GTEx/v7 | Nerve_Tibial | ENSG00000087085 | ACHE | 1,1E-24 |
| rs4556017 | 7:100632790 | 0,961771 | intronic | GTEx/v7 | Thyroid | ENSG00000087085 | ACHE | 6,0E-29 |
| rs4556017 | 7:100632790 | 0,961771 | intronic | CMC | CMC_SVA_cis | ENSG00000168090 | COPS6 | 9,0E-03 |
| rs4556017 | 7:100632790 | 0,961771 | intronic | GTEx/v7 | Nerve_Tibial | ENSG00000087087 | SRRT | 4,2E-12 |
| rs4556017 | 7:100632790 | 0,961771 | intronic | BIOSQTL | BIOS_eQTL_geneLevel | ENSG00000169871 | TRIM56 | 1,7E-03 |
| rs4556017 | 7:100632790 | 0,961771 | intronic | BIOSQTL | BIOS_eQTL_geneLevel | ENSG00000087077 | TRIP6 | 0,0E+00 |
| rs35318931 | 23:38009121 | 0,872827 | exonic | GTEx/v7 | Esophagus_Muscularis | ENSG00000101955 | SRPX | 9,4E-08 |
| rs6482359 | 10:24330805 | 0,796779 | intronic | GTEx/v7 | Thyroid | ENSG00000120549 | KIAA1217 | 1,5E-37 |
| rs728327 | 2:45775995 | 0,668817 | intronic | BIOSQTL | BIOS_eQTL_geneLevel | ENSG00000068784 | SRBD1 | 0,0E+00 |
| rs728327 | 2:45775995 | 0,668817 | intronic | GTEx/v7 | Colon_Transverse | ENSG00000068784 | SRBD1 | 1,7E-04 |
| rs728327 | 2:45775995 | 0,668817 | intronic | GTEx/v7 | Cells_Transformed_fibroblasts | ENSG00000068784 | SRBD1 | 7,1E-22 |
| rs728327 | 2:45775995 | 0,668817 | intronic | GTEx/v7 | Testis | ENSG00000068784 | SRBD1 | 2,9E-15 |
| rs6498573 | 16:15879373 | 0,63247 | intronic | BIOSQTL | BIOS_eQTL_geneLevel | ENSG00000133392 | MYH11 | 0,0E+00 |
| rs6498573 | 16:15879373 | 0,63247 | intronic | BIOSQTL | BIOS_eQTL_geneLevel | ENSG00000133392 | MYH11 | 0,0E+00 |
| rs6498573 | 16:15879373 | 0,63247 | intronic | BIOSQTL | BIOS_eQTL_geneLevel | ENSG00000133392 | MYH11 | 0,0E+00 |
| rs6498573 | 16:15879373 | 0,63247 | intronic | BIOSQTL | BIOS_eQTL_geneLevel | ENSG00000133392 | MYH11 | 0,0E+00 |
| rs6498573 | 16:15879373 | 0,63247 | intronic | BIOSQTL | BIOS_eQTL_geneLevel | ENSG00000072864 | NDE1 | 0,0E+00 |
| rs6498573 | 16:15879373 | 0,63247 | intronic | BIOSQTL | BIOS_eQTL_geneLevel | ENSG00000072864 | NDE1 | 0,0E+00 |
| rs6498573 | 16:15879373 | 0,63247 | intronic | GTEx/v7 | Colon_Sigmoid | ENSG00000157045 | NTAN1 | 1,4E-02 |
| rs11635984 | 15:33012232 | 0,619177 | intronic | BIOSQTL | BIOS_eQTL_geneLevel | ENSG00000198826 | ARHGAP11A | 0,0E+00 |
| rs2421206 | 19:11262477 | 0,608719 | intronic | PsychENCODE | PsychENCODE_eQTLs | ENSG00000197256 | KANK2 | 2,4E-08 |
| rs2421206 | 19:11262477 | 0,608719 | intronic | CMC | CMC_SVA_cis | ENSG00000197256 | KANK2 | 9,0E-03 |
| rs2421206 | 19:11262477 | 0,608719 | intronic | GTEx/v7 | Adipose_Subcutaneous | ENSG00000197256 | KANK2 | 6,5E-05 |
| rs2421206 | 19:11262477 | 0,608719 | intronic | GTEx/v7 | Brain_Cerebellum | ENSG00000197256 | KANK2 | 3,6E-05 |
| rs9853475 | 3:114500255 | 0,598875 | intronic | BRAINEAC | OCTX | ENSG00000181722 | ZBTB20 | 1,6E-02 |
| rs11769827 | 7:73313856 | 0,56898 | intergenic | BIOSQTL | BIOS_eQTL_geneLevel | ENSG00000106077 | ABHD11 | 0,0E+00 |
| rs11769827 | 7:73313856 | 0,56898 | intergenic | BIOSQTL | BIOS_eQTL_geneLevel | ENSG00000106683 | LIMK1 | 2,9E-03 |
| rs11769827 | 7:73313856 | 0,56898 | intergenic | BIOSQTL | BIOS_eQTL_geneLevel | ENSG00000106683 | LIMK1 | 8,0E-04 |
| rs11769827 | 7:73313856 | 0,56898 | intergenic | BIOSQTL | BIOS_eQTL_geneLevel | ENSG00000165171 | WBSCR27 | 0,0E+00 |

| | | | | | | | | |
|---|---|---|---|---|---|---|---|---|
| rs11769827 | 7:73313856 | 0,56898 | intergenic | GTEx/v7 | Adipose_Subcutaneous | ENSG00000165171 | WBSCR27 | 8,8E-90 |
| rs11769827 | 7:73313856 | 0,56898 | intergenic | GTEx/v7 | Adipose_Visceral_Omentum | ENSG00000165171 | WBSCR27 | 3,2E-71 |
| rs11769827 | 7:73313856 | 0,56898 | intergenic | GTEx/v7 | Adrenal_Gland | ENSG00000165171 | WBSCR27 | 2,5E-29 |
| rs11769827 | 7:73313856 | 0,56898 | intergenic | GTEx/v7 | Whole_Blood | ENSG00000165171 | WBSCR27 | 1,0E-37 |
| rs11769827 | 7:73313856 | 0,56898 | intergenic | GTEx/v7 | Artery_Aorta | ENSG00000165171 | WBSCR27 | 1,2E-44 |
| rs11769827 | 7:73313856 | 0,56898 | intergenic | GTEx/v7 | Artery_Coronary | ENSG00000165171 | WBSCR27 | 1,1E-23 |
| rs11769827 | 7:73313856 | 0,56898 | intergenic | GTEx/v7 | Artery_Tibial | ENSG00000165171 | WBSCR27 | 8,0E-71 |
| rs11769827 | 7:73313856 | 0,56898 | intergenic | GTEx/v7 | Brain_Cerebellum | ENSG00000165171 | WBSCR27 | 8,3E-33 |
| rs11769827 | 7:73313856 | 0,56898 | intergenic | GTEx/v7 | Breast_Mammary_Tissue | ENSG00000165171 | WBSCR27 | 3,1E-59 |
| rs11769827 | 7:73313856 | 0,56898 | intergenic | GTEx/v7 | Colon_Sigmoid | ENSG00000165171 | WBSCR27 | 1,1E-38 |
| rs11769827 | 7:73313856 | 0,56898 | intergenic | GTEx/v7 | Colon_Transverse | ENSG00000165171 | WBSCR27 | 3,1E-53 |
| rs11769827 | 7:73313856 | 0,56898 | intergenic | GTEx/v7 | Esophagus_Gastroesophagea | ENSG00000165171 | WBSCR27 | 2,8E-43 |
| rs11769827 | 7:73313856 | 0,56898 | intergenic | GTEx/v7 | Esophagus_Mucosa | ENSG00000165171 | WBSCR27 | 8,1E-58 |
| rs11769827 | 7:73313856 | 0,56898 | intergenic | GTEx/v7 | Esophagus_Muscularis | ENSG00000165171 | WBSCR27 | 1,9E-71 |
| rs11769827 | 7:73313856 | 0,56898 | intergenic | GTEx/v7 | Heart_Atrial_Appendage | ENSG00000165171 | WBSCR27 | 5,6E-42 |
| rs11769827 | 7:73313856 | 0,56898 | intergenic | GTEx/v7 | Heart_Left_Ventricle | ENSG00000165171 | WBSCR27 | 1,1E-36 |
| rs11769827 | 7:73313856 | 0,56898 | intergenic | GTEx/v7 | Lung | ENSG00000165171 | WBSCR27 | 9,2E-81 |
| rs11769827 | 7:73313856 | 0,56898 | intergenic | GTEx/v7 | Muscle_Skeletal | ENSG00000165171 | WBSCR27 | 6,5E-42 |
| rs11769827 | 7:73313856 | 0,56898 | intergenic | GTEx/v7 | Nerve_Tibial | ENSG00000165171 | WBSCR27 | 4,4E-80 |
| rs11769827 | 7:73313856 | 0,56898 | intergenic | GTEx/v7 | Pancreas | ENSG00000165171 | WBSCR27 | 2,1E-56 |
| rs11769827 | 7:73313856 | 0,56898 | intergenic | GTEx/v7 | Cells_Transformed_fibroblasts | ENSG00000165171 | WBSCR27 | 3,2E-79 |
| rs11769827 | 7:73313856 | 0,56898 | intergenic | GTEx/v7 | Skin_Not_Sun_Exposed_Supr | ENSG00000165171 | WBSCR27 | 4,8E-63 |
| rs11769827 | 7:73313856 | 0,56898 | intergenic | GTEx/v7 | Skin_Sun_Exposed_Lower_le | ENSG00000165171 | WBSCR27 | 7,6E-84 |
| rs11769827 | 7:73313856 | 0,56898 | intergenic | GTEx/v7 | Small_Intestine_Terminal_Ileu | ENSG00000165171 | WBSCR27 | 5,1E-24 |
| rs11769827 | 7:73313856 | 0,56898 | intergenic | GTEx/v7 | Stomach | ENSG00000165171 | WBSCR27 | 2,8E-32 |
| rs11769827 | 7:73313856 | 0,56898 | intergenic | GTEx/v7 | Testis | ENSG00000165171 | WBSCR27 | 3,1E-27 |
| rs11769827 | 7:73313856 | 0,56898 | intergenic | GTEx/v7 | Thyroid | ENSG00000165171 | WBSCR27 | 9,6E-102 |
| rs7795564 | 7:55124829 | 0,553309 | intronic | PsychENCODE | PsychENCODE_eQTLs | ENSG00000146648 | EGFR | 2,4E-03 |
| rs1333047 | 9:22124504 | 0,520459 | intergenic | CMC | CMC_SVA_cis | ENSG00000240498 | CDKN2B-AS1 | 9,0E-03 |
| rs1333047 | 9:22124504 | 0,520459 | intergenic | CMC | CMC_NoSVA_cis | ENSG00000240498 | CDKN2B-AS1 | 9,0E-03 |
| rs1333047 | 9:22124504 | 0,520459 | intergenic | CMC | CMC_NoSVA_cis | ENSG00000240498 | CDKN2B-AS1 | 9,0E-03 |
| rs2687965 | 4:39491521 | 0,50179 | intergenic | CMC | CMC_SVA_cis | ENSG00000134962 | KLB | 9,0E-03 |
| rs2687965 | 4:39491521 | 0,50179 | intergenic | CMC | CMC_NoSVA_cis | ENSG00000134962 | KLB | 9,0E-03 |
| rs2687965 | 4:39491521 | 0,50179 | intergenic | CMC | CMC_NoSVA_cis | ENSG00000134962 | KLB | 9,0E-03 |
| rs2687965 | 4:39491521 | 0,50179 | intergenic | BIOSQTL | BIOS_eQTL_geneLevel | ENSG00000121897 | LIAS | 1,9E-02 |
| rs2687965 | 4:39491521 | 0,50179 | intergenic | BIOSQTL | BIOS_eQTL_geneLevel | ENSG00000163683 | SMIM14 | 1,4E-02 |
| rs2687965 | 4:39491521 | 0,50179 | intergenic | BIOSQTL | BIOS_eQTL_geneLevel | ENSG00000109814 | UGDH | 0,0E+00 |
| rs2687965 | 4:39491521 | 0,50179 | intergenic | CMC | CMC_SVA_cis | ENSG00000109814 | UGDH | 9,0E-03 |
| rs2687965 | 4:39491521 | 0,50179 | intergenic | CMC | CMC_NoSVA_cis | ENSG00000109814 | UGDH | 4,9E-02 |
| rs2687965 | 4:39491521 | 0,50179 | intergenic | CMC | CMC_NoSVA_cis | ENSG00000109814 | UGDH | 4,9E-02 |
| rs2687965 | 4:39491521 | 0,50179 | intergenic | GTEx/v7 | Artery_Tibial | ENSG00000109814 | UGDH | 4,7E-06 |
| rs2687965 | 4:39491521 | 0,50179 | intergenic | GTEx/v7 | Muscle_Skeletal | ENSG00000109814 | UGDH | 4,3E-09 |
| rs2687965 | 4:39491521 | 0,50179 | intergenic | GTEx/v7 | Cells_Transformed_fibroblasts | ENSG00000109814 | UGDH | 1,7E-14 |
| rs2687965 | 4:39491521 | 0,50179 | intergenic | GTEx/v7 | Thyroid | ENSG00000109814 | UGDH | 9,9E-03 |
| rs2687965 | 4:39491521 | 0,50179 | intergenic | GTEx/v7 | Cells_Transformed_fibroblasts | ENSG00000249348 | UGDH-AS1 | 6,4E-09 |
| rs2186797 | 11:70007770 | 0,969962 | exonic | BIOSQTL | BIOS_eQTL_geneLevel | ENSG00000254721 | RP11-805J14.5 | 8,2E-03 |
| rs34161672 | 20:56020599 | 0,659985 | intergenic | GTEx/v7 | Pancreas | ENSG00000132819 | RBM38 | 5,1E-26 |
| rs6498573 | 16:15879373 | 0,63247 | intronic | BIOSQTL | BIOS_eQTL_geneLevel | ENSG00000263335 | AF001548.5 | 0,0E+00 |
| rs6498573 | 16:15879373 | 0,63247 | intronic | GTEx/v7 | Whole_Blood | ENSG00000263335 | AF001548.5 | 1,1E-54 |

| | | | | | | | | |
|---|---|---|---|---|---|---|---|---|
| rs6498573 | 16:15879373 | 0,63247 | intronic | GTEx/v7 | Esophagus_Mucosa | ENSG00000263335 | AF001548.5 | 9,2E-58 |
| rs6498573 | 16:15879373 | 0,63247 | intronic | GTEx/v7 | Nerve_Tibial | ENSG00000263335 | AF001548.5 | 2,7E-73 |
| rs6498573 | 16:15879373 | 0,63247 | intronic | GTEx/v7 | Skin_Sun_Exposed_Lower_le | ENSG00000263335 | AF001548.5 | 3,2E-87 |
| rs6498573 | 16:15879373 | 0,63247 | intronic | GTEx/v7 | Thyroid | ENSG00000263335 | AF001548.5 | 3,8E-63 |
| rs6498573 | 16:15879373 | 0,63247 | intronic | BIOSQTL | BIOS_eQTL_geneLevel | ENSG00000263065 | AF001548.6 | 0,0E+00 |
| rs7994724 | 13:51445560 | 0,620925 | intergenic | BIOSQTL | BIOS_eQTL_geneLevel | ENSG00000136104 | RNASEH2B | 6,2E-06 |
| rs7994724 | 13:51445560 | 0,620925 | intergenic | xQTLServer | xQTLServer_eQTLs | ENSG00000136104 | RNASEH2B | 2,7E-17 |
| rs7994724 | 13:51445560 | 0,620925 | intergenic | CMC | CMC_SVA_cis | ENSG00000136104 | RNASEH2B | 9,0E-03 |
| rs7994724 | 13:51445560 | 0,620925 | intergenic | CMC | CMC_NoSVA_cis | ENSG00000136104 | RNASEH2B | 9,0E-03 |
| rs7994724 | 13:51445560 | 0,620925 | intergenic | CMC | CMC_NoSVA_cis | ENSG00000136104 | RNASEH2B | 9,0E-03 |
| rs7994724 | 13:51445560 | 0,620925 | intergenic | GTEx/v7 | Adipose_Subcutaneous | ENSG00000136104 | RNASEH2B | 1,9E-25 |
| rs7994724 | 13:51445560 | 0,620925 | intergenic | GTEx/v7 | Adipose_Visceral_Omentum | ENSG00000136104 | RNASEH2B | 2,5E-20 |
| rs7994724 | 13:51445560 | 0,620925 | intergenic | GTEx/v7 | Artery_Aorta | ENSG00000136104 | RNASEH2B | 5,7E-17 |
| rs7994724 | 13:51445560 | 0,620925 | intergenic | GTEx/v7 | Artery_Tibial | ENSG00000136104 | RNASEH2B | 2,3E-30 |
| rs7994724 | 13:51445560 | 0,620925 | intergenic | GTEx/v7 | Brain_Anterior_cingulate_corte | ENSG00000136104 | RNASEH2B | 4,6E-16 |
| rs7994724 | 13:51445560 | 0,620925 | intergenic | GTEx/v7 | Brain_Caudate_basal_ganglia | ENSG00000136104 | RNASEH2B | 5,0E-11 |
| rs7994724 | 13:51445560 | 0,620925 | intergenic | GTEx/v7 | Brain_Cortex | ENSG00000136104 | RNASEH2B | 2,9E-21 |
| rs7994724 | 13:51445560 | 0,620925 | intergenic | GTEx/v7 | Brain_Putamen_basal_ganglia | ENSG00000136104 | RNASEH2B | 4,2E-08 |
| rs7994724 | 13:51445560 | 0,620925 | intergenic | GTEx/v7 | Colon_Transverse | ENSG00000136104 | RNASEH2B | 2,2E-18 |
| rs7994724 | 13:51445560 | 0,620925 | intergenic | GTEx/v7 | Esophagus_Gastroesophagea | ENSG00000136104 | RNASEH2B | 1,3E-20 |
| rs7994724 | 13:51445560 | 0,620925 | intergenic | GTEx/v7 | Esophagus_Muscularis | ENSG00000136104 | RNASEH2B | 1,2E-46 |
| rs7994724 | 13:51445560 | 0,620925 | intergenic | GTEx/v7 | Heart_Atrial_Appendage | ENSG00000136104 | RNASEH2B | 8,4E-22 |
| rs7994724 | 13:51445560 | 0,620925 | intergenic | GTEx/v7 | Muscle_Skeletal | ENSG00000136104 | RNASEH2B | 1,1E-82 |
| rs7994724 | 13:51445560 | 0,620925 | intergenic | GTEx/v7 | Ovary | ENSG00000136104 | RNASEH2B | 1,5E-07 |
| rs7994724 | 13:51445560 | 0,620925 | intergenic | GTEx/v7 | Pancreas | ENSG00000136104 | RNASEH2B | 1,1E-23 |
| rs7994724 | 13:51445560 | 0,620925 | intergenic | GTEx/v7 | Pituitary | ENSG00000136104 | RNASEH2B | 2,1E-16 |
| rs7994724 | 13:51445560 | 0,620925 | intergenic | GTEx/v7 | Thyroid | ENSG00000136104 | RNASEH2B | 3,8E-23 |
| rs7994724 | 13:51445560 | 0,620925 | intergenic | GTEx/v7 | Adipose_Subcutaneous | ENSG00000233672 | RNASEH2B-AS1 | 9,6E-16 |
| rs7994724 | 13:51445560 | 0,620925 | intergenic | GTEx/v7 | Artery_Aorta | ENSG00000233672 | RNASEH2B-AS1 | 1,2E-08 |
| rs7994724 | 13:51445560 | 0,620925 | intergenic | GTEx/v7 | Brain_Cerebellar_Hemisphere | ENSG00000233672 | RNASEH2B-AS1 | 3,7E-19 |
| rs7994724 | 13:51445560 | 0,620925 | intergenic | GTEx/v7 | Brain_Cerebellum | ENSG00000233672 | RNASEH2B-AS1 | 1,3E-19 |
| rs7994724 | 13:51445560 | 0,620925 | intergenic | GTEx/v7 | Muscle_Skeletal | ENSG00000233672 | RNASEH2B-AS1 | 2,2E-15 |
| rs7994724 | 13:51445560 | 0,620925 | intergenic | GTEx/v7 | Nerve_Tibial | ENSG00000233672 | RNASEH2B-AS1 | 1,2E-13 |
| rs7994724 | 13:51445560 | 0,620925 | intergenic | GTEx/v7 | Thyroid | ENSG00000233672 | RNASEH2B-AS1 | 1,2E-28 |
| rs11635984 | 15:33012232 | 0,619177 | intronic | PsychENCODE | PsychENCODE_eQTLs | ENSG00000259721 | RP11-758N13.1 | 9,9E-03 |
| rs2687965 | 4:39491521 | 0,50179 | intergenic | CMC | CMC_SVA_cis | ENSG00000163682 | RPL9 | 9,0E-03 |
| rs2687965 | 4:39491521 | 0,50179 | intergenic | GTEx/v7 | Artery_Tibial | ENSG00000163682 | RPL9 | 3,1E-88 |
